# Large-scale modelling of β-thalassaemia carrier prevalence: geostatistical predictions and population estimates from a global evidence base

**DOI:** 10.64898/2026.10.01.26363283

**Authors:** Mikaella Kontopoulou, Chigozie Edson Utazi, Somnath Chaudhuri, Maria Xenophontos, Charlie Leboff, Sotiroula Chatzimatthaiou, Kalia Orphanou, Carsten Werner Lederer, Coralea Stephanou, Petros Kountouris

## Abstract

**Background:** Evidence on β-thalassaemia carrier prevalence is fragmented across heterogeneous surveys and national summaries, leaving within-country variation and geographic concentration of carriers poorly resolved. This study characterises β-thalassaemia carrier prevalence and carrier populations at high spatial resolution and identifies where additional data would most improve geographic coverage.

**Methods:** Using carrier-prevalence records curated through ITHANET, a global haemoglobinopathy resource, Bayesian geostatistical modelling generated high-resolution predictions across 25 countries. Predictions were integrated with population data to estimate national and subnational carrier numbers. Prediction uncertainty and gaps in existing survey coverage informed prioritisation of healthcare sites for additional surveys.

**Findings:** Predicted prevalence varied markedly within countries, and highest-prevalence areas did not coincide with the largest carrier populations. National mean prevalence was highest in Cyprus (12·99%), whereas India, China, and Pakistan had the largest carrier populations, together accounting for 81·21% of estimated carriers. Random five-fold cross-validation favoured a model capturing broad geographic patterns and location-specific variation. Predictive performance deteriorated when geographically clustered records were withheld together, indicating reduced reliability across larger gaps in geographic sampling and the need for additional data from poorly characterised areas. Survey prioritisation identified 183 healthcare sites across 17 countries for future carrier-prevalence surveys.

**Interpretation:** By distinguishing areas of high carrier prevalence, large carrier populations, and evidence gaps, this updatable framework provides a subnational evidence base to inform β-thalassaemia prevention planning and guide future data collection, screening, and genetic counselling.

**Funding:** COST Action HELIOS, Horizon Europe HemaFAIR, Horizon 2020 GenoMed4ALL, and the Cyprus Institute of Neurology & Genetics.

**Research in context:** 

**Evidence before this study:** Carrier identification is central to β-thalassaemia prevention because the geographic distribution of carriers is a fundamental determinant of where at-risk reproductive pairings can occur. Population surveys and screening programmes have established the broad distribution of β-thalassaemia and informed prevention strategies in several high-prevalence settings, but the underlying evidence varies substantially in sampling frame, diagnostic method, period of observation, and geographic coverage. Carrier prevalence reflects the distribution of inherited *HBB* variants across populations and is shaped by population history and structure, including endogamy, admixture, and migration; substantial variation can therefore occur within national boundaries. National estimates are useful for broad epidemiological assessment but cannot resolve such local heterogeneity, whereas individual surveys provide direct evidence only for the populations and locations sampled. Geostatistical modelling can integrate geographically dispersed observations to estimate carrier prevalence in unsampled intervening locations while quantifying uncertainty. This approach has enabled high-resolution characterisation of the spatial distribution of other haemoglobinopathies, including global mapping of sickle haemoglobin allele frequency with estimates of neonates with sickle cell anaemia or sickle cell trait, mapping of haemoglobin C allele frequency across Africa, and estimation of α-thalassaemia variant frequencies in Thailand. Comparable large-scale spatial modelling of β-thalassaemia remains limited and geographically restricted. Consequently, the fine-scale distribution of β-thalassaemia carrier prevalence within countries, and the extent to which these patterns translate into the geographic distribution of carrier populations, remain insufficiently characterised.

**Added value of this study:** To our knowledge, this study provides the first high-resolution characterisation of β-thalassaemia carrier prevalence across 25 countries with sufficient curated carrier-prevalence evidence for spatial modelling. Quality-appraised carrier-prevalence records were integrated within a Bayesian geostatistical framework to generate predictions at approximately 5 km × 5 km resolution, with uncertainty quantified across the modelled domain. Alternative model formulations were compared, and ecological, demographic, and healthcare service availability covariates were evaluated for their contribution to predictive performance. Carrier-prevalence predictions were combined with count-preserving population surfaces to estimate the number and spatial distribution of carriers. Prediction uncertainty and existing sampling coverage were then used to identify healthcare sites where additional epidemiological surveys could most improve the geographic coverage of the empirical evidence. This framework therefore distinguishes areas where β-thalassaemia carrier prevalence is highest, where carrier populations are largest, and where additional data are most needed to improve spatial inference, while allowing estimates and survey priorities to be updated as new carrier prevalence data become available.

**Implications of all the available evidence:** High-resolution β-thalassaemia carrier-prevalence adds a missing epidemiological layer to prevention planning by showing where screening is likely to have the highest yield, while carrier numbers indicate where downstream requirements for confirmatory diagnosis and genetic counselling could be greatest. Used alongside existing programme data, these measures can help distinguish settings in which established prevention strategies remain appropriately configured from those in which their geographic reach, intensity, or capacity may warrant reassessment, without implying that new programmes are necessarily required. The priority site analysis extends this framework by identifying healthcare sites where additional surveys could strengthen geographic sampling coverage the most. The resulting maps and data have been made publicly available through the ITHANET portal for interactive exploration and download, broadening access to fine-scale evidence to support policy and programme decisions.

## Introduction

Haemoglobinopathies, including sickle cell disease and the thalassaemia syndromes, are among the most common monogenic disorders worldwide.^1^ Caused by inherited variants affecting haemoglobin structure or globin-chain synthesis, they contribute substantially to early-life morbidity, mortality and lifelong healthcare burden.^1,2^ Their historical distribution reflects malaria-driven balancing selection, whereby heterozygosity for globin gene variants conferred a survival advantage under sustained malaria transmission.^1^ Migration, displacement, and population admixture have extended haemoglobinopathies beyond areas of historical endemicity, while population structure, consanguinity, endogamy, and reproductive patterns continue to shape their distribution locally.^3,4^ Epidemiology is therefore highly heterogeneous, with substantial variation within countries and between neighbouring populations.

Successive efforts to characterise this heterogeneity have established the broad global distribution and public-health importance of haemoglobinopathies. Early compilations by Frank B. Livingstone catalogued haemoglobin variants across human populations and were later extended to thalassaemia-causing variants.^5^ Modell’s *Haemoglobinopathologist’s Almanac* synthesised evidence into national estimates and service indicators that remain influential in international assessment and prevention planning.^6^ The Global Burden of Disease study later situated haemoglobinopathies within a comparative disease framework, enabling national assessment of mortality, morbidity, and disability.^2^ Population surveys, screening programmes, regional studies, and molecular investigations have refined estimates of local prevalence and variant spectra, supporting prevention strategies in many high-prevalence settings.^4,7^ These sources remain indispensable, but address different epidemiological questions and spatial scales. National estimates are appropriate for broad assessment, whereas individual surveys describe sampled populations and cannot by themselves resolve prevalence across intervening or unsampled areas.

This distinction is important for carrier prevalence. Across autosomal recessive haemoglobinopathies, the geographic distribution of carriers defines where reproductive pairings capable of producing severe disease can occur and, therefore, provides information that is complementary to estimates of diagnosed disease. For β-thalassaemia, carrier prevalence is consequently relevant to organising and evaluating established prevention activities, including carrier detection, confirmatory diagnosis, and genetic counselling.^4^ High-prevalence areas do not necessarily have the largest carrier populations, while densely populated areas with moderate prevalence can contain a substantial number of carriers. Characterising both dimensions is necessary to understand how inherited risk intersects with population size and existing health-service provision.

Geostatistical modelling provides a means of extending geographically dispersed observations into continuous estimates while preserving uncertainty, particularly in locations without direct measurements.^8^ Its value has been demonstrated across haemoglobinopathies. Spatial modelling characterised fine-scale haemoglobin S (HbS) allele frequency and, combined with demographic data, the distribution of neonates with sickle cell trait and sickle cell anaemia.^8,9^ Haemoglobin C (HbC) allele frequency was mapped across Africa,^10^ while geostatistical analysis of α-thalassaemia in Thailand translated heterogeneous prevalence evidence into continuous estimates and subnational burden measures.^11^ These studies demonstrate how spatial modelling can add an epidemiological layer between individual surveys and national summaries by identifying geographic structure that neither source can resolve alone.

β-thalassaemia remains a conspicuous exception. Existing evidence has established its broad geographic distribution and informed successful prevention programmes,^4,6^ yet carrier prevalence has not been characterised within a comparable multi-country spatial framework. Previous geostatistical work has been confined to transfusion-dependent β-thalassaemia incidence within Fars Province, Iran.^12^ A gap therefore persists between the substantial body of local prevalence evidence and the geographically resolved estimates needed to understand how carrier prevalence varies within countries. Spatially explicit estimates can distinguish three questions that conventional summaries cannot fully resolve, namely where carrier prevalence is highest, where carrier populations are largest, and where evidence remains insufficient for reliable characterisation.

Bridging this gap requires the underlying evidence to be organised in a form that preserves source provenance and epidemiological context while allowing new information to be incorporated over time. IthaMaps, the epidemiological database of the ITHANET community portal, was redeveloped within this study for that purpose.^13^ The revised resource provides an open, source-linked and updatable foundation for haemoglobinopathy epidemiology, genetic diversity, and service information, while supporting the spatial analyses presented here.

This study aimed to characterise β-thalassaemia carrier prevalence at high spatial resolution across 25 countries with sufficient curated carrier-prevalence evidence and estimate the geographic distribution of carrier populations. The analysis further examined whether spatially varying ecological, demographic, population-structure, and service-related information improved prediction, and used prediction uncertainty and sampling coverage to identify existing healthcare sites where additional surveys could improve geographic coverage. Linking observed prevalence, spatial inference, carrier-population estimates, and future data-collection priorities within an updatable evidence framework addresses the gap between fragmented local evidence and the geographic information needed for contemporary β-thalassaemia surveillance and prevention planning.

## Methods

### Data collection

Sources were identified through three complementary streams. Published sources were retrieved using the established ITHANET PubMed strategy^13^ and additional PubMed and Google Scholar searches through Publish or Perish (Supplementary section 1.2).^14^ A third stream comprised sources independently curated by the Institute for Health Metrics and Evaluation and harmonised to IthaMaps haemoglobinopathy, epidemiological, and geographic definitions (Figure 1; Supplementary section 1.7). Sources entered a prespecified prioritisation queue based on β-thalassaemia relevance and geographic evidence sparsity; prioritisation determined screening order, not eligibility (Supplementary sections 1.3–1.4). Eligible sources contained extractable human haemoglobinopathy data on population frequency, genetic variation, or prevention and management service provision. Data were extracted using a standardised schema; a single source could contribute multiple data records (Supplementary section 1.5). Records were georeferenced at the finest supported resolution, using recruitment-site coordinates where identifiable and otherwise the centroid of the reported administrative level 0 (ADM0; country), administrative level 1 (ADM1), or administrative level 2 (ADM2) unit. For records containing pooled estimates spanning several recruitment locations, all identifiable recruitment sites were georeferenced (Supplementary sections 1.1 and 1.6). Carrier prevalence, incidence, allele frequency, and relative allele frequency records were quality appraised using measure-specific criteria adapted from the Joanna Briggs Institute (JBI) prevalence tool,^15^ covering sampling frame, sample-size adequacy, geographic and haemoglobinopathy specificity, diagnostic validity, and numerical consistency. Records satisfying over 70% of applicable criteria were classified as low risk of bias, consistent with published applications of the JBI prevalence appraisal framework.^16^ For carrier-prevalence records, this threshold required at least five of six criteria. Records with insufficient information remained unappraised but were retained during curation, with their eligibility for modelling evaluated in sensitivity analyses (Supplementary sections 1.10 and 2.10). Curated records were integrated into IthaMaps, retaining provenance and enabling dynamic geospatial and tabular visualisation, user-defined filtering, and data download (Supplementary sections 1.12 and 1.13).

**Figure 1.**
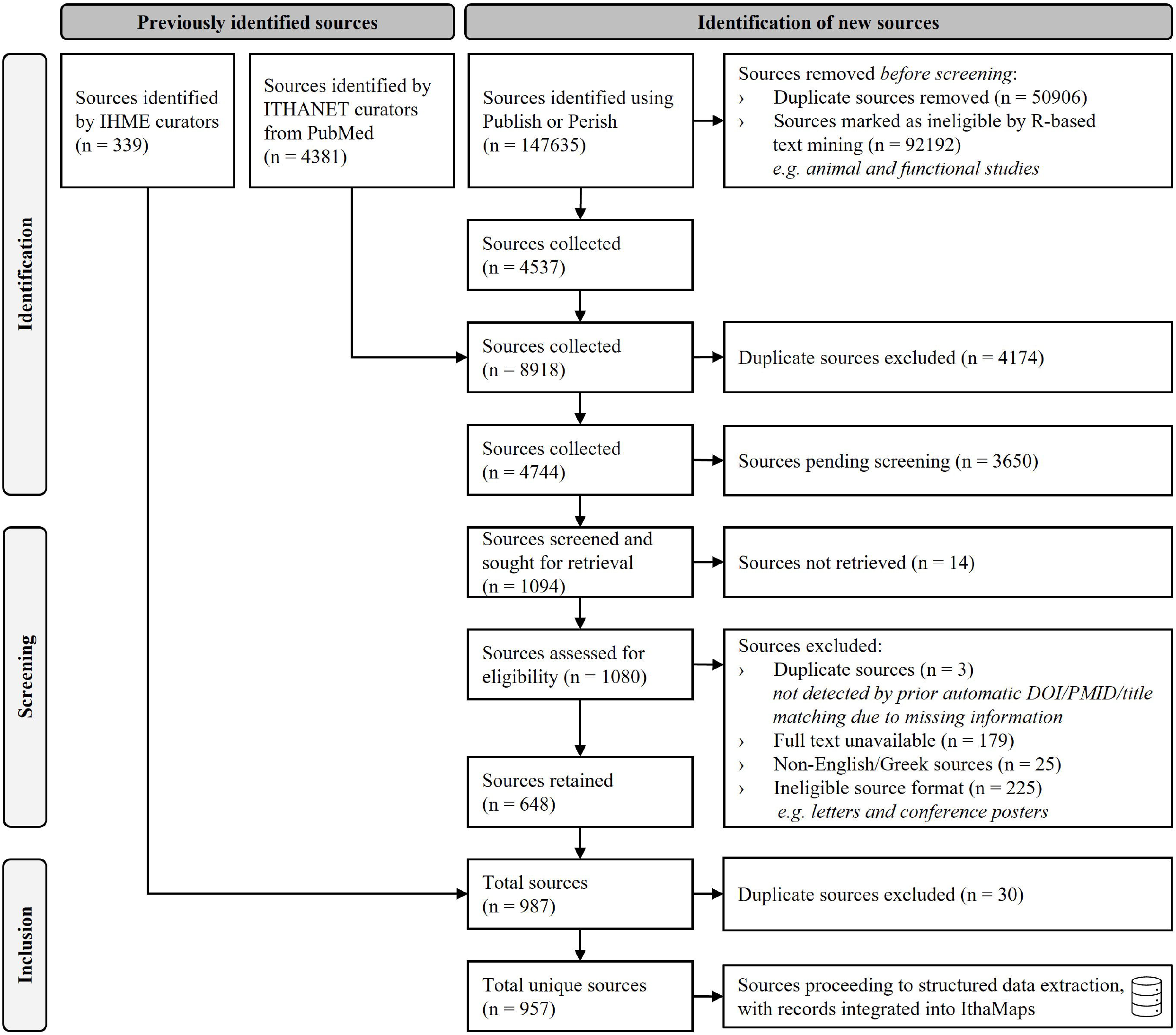
Flow diagram of source identification, prioritisation, screening, and curation. IHME: Institute for Health Metrics and Evaluation

### Data preparation for geostatistical modelling

Records were eligible for use in geostatistical analysis if they provided or permitted estimation of β-thalassaemia carrier prevalence in the general population. Where only β-thalassaemia disease prevalence was reported, carrier prevalence was derived under Hardy-Weinberg equilibrium when genotype interpretation was unambiguous; clinically selected populations were excluded from derivation (Supplementary section 1.8). Potential outliers were assessed as described in Supplementary section 1.11.

Coarse area-level records were spatially disaggregated using a previously established population-informed procedure combining k-means clustering (Supplementary section 2.4).^17^ Records georeferenced to recruitment sites, or ADM2 units retained their original coordinates. For coarser (ADM0 and ADM1) records, inhabited cells from the 2026 WorldPop 1 km × 1 km population surface^18^ were clustered to identify representative population concentrations and corresponding finer administrative units. ADM0 records were retained only for countries smaller than 20 000 km^2^ and assigned to one representative ADM1 proxy; ADM0 records from larger countries were excluded from modelling. ADM1 records were assigned to up to three ADM2 proxies where clustering identified distinct within-unit concentrations. ADM0-derived ADM1 proxies were not further disaggregated. Point-on-surface coordinates of the selected units defined proxy locations, each retaining the original record’s carrier count and sample size.

One record was retained per modelling coordinate to avoid disproportionate influence from repeatedly sampled locations (Supplementary section 2.5). A prespecified hierarchy prioritised records at original over proxy coordinates, records representing a single recruitment location over those containing pooled estimates, original records over those with Hardy-Weinberg-derived carrier prevalence, and records with lower risk of bias. General population or multiethnic records were preferred to ethnicity-specific records unless the latter had at least twice the number tested; remaining overlaps were resolved by sample size and recruitment recency. Nearest-neighbour distance identified spatially isolated records for exclusion.^19^ Moreover, records from geographically isolated countries separated from the main body of evidence were excluded to avoid unsupported long-range spatial inference (Supplementary section 2.6).^20^

Prespecified sensitivity analyses compared alternative record-inclusion and spatial-processing specifications differing in the treatment of pooled multi-location estimates, Hardy-Weinberg-derived carrier-prevalence records, risk-of-bias restrictions, population-informed spatial disaggregation, and spatial-noise records (Supplementary section 2.10).

Eight candidate covariates were specified *a priori*, comprising *Plasmodium falciparum* incidence, consanguinity, ethnic fractionalisation, religious diversity, population count, degree of urbanisation, haemoglobinopathy genetic counselling availability, and haemoglobinopathy prevention service availability. Covariates were selected for model inclusion following a previously established approach using univariable non-spatial binomial models, multicollinearity assessment, and multivariable backward selection by Akaike information criterion.^21,22^

Because selected covariate data were incomplete for some records, model development proceeded in two stages (Supplementary sections 2.1–2.3 and 2.11). Covariate-inclusive and covariate-free formulations were first compared in the covariate-restricted dataset, which comprised only records with complete data for all selected covariates. Because covariates did not improve held-out prediction, subsequent model development used covariate-free formulations in the covariate-unrestricted dataset, which retained eligible records irrespective of covariate completeness.

### Geostatistical model selection and validation

Carrier prevalence was modelled from the observed carrier count and number of individuals tested at each modelling coordinate using Bayesian binomial models. Model formulations considered three complementary components of carrier-prevalence variation: the selected covariates as fixed-effects; an independent and identically distributed (IID) location-level random-effect for non-spatial heterogeneity, and a Matérn spatial random-effect for residual correlation between nearby locations. The Matérn field was approximated by the stochastic partial differential equation approach on a spherical mesh to account for Earth curvature, with integrated nested Laplace approximation for posterior inference.^23,24^ Six formulations combining fixed, IID, and spatial components were fitted to the covariate-restricted dataset; three covariate-free formulations were subsequently fitted to the covariate-unrestricted dataset. Model evaluation combined the Watanabe-Akaike information criterion (WAIC) and conditional predictive ordinates (CPO) from full-data fits with random five-fold cross-validation (CV). WAIC assessed predictive fit relative to model complexity, whereas mean negative log CPO summarised single-record leave-one-out predictive support. WAIC and CPO were compared only among models fitted to the same set of records. Model specifications and evaluation procedures are provided in Supplementary sections 2.7–2.9 and 2.11. Figure 2 summarises the analytical workflow.

**Figure 2.**
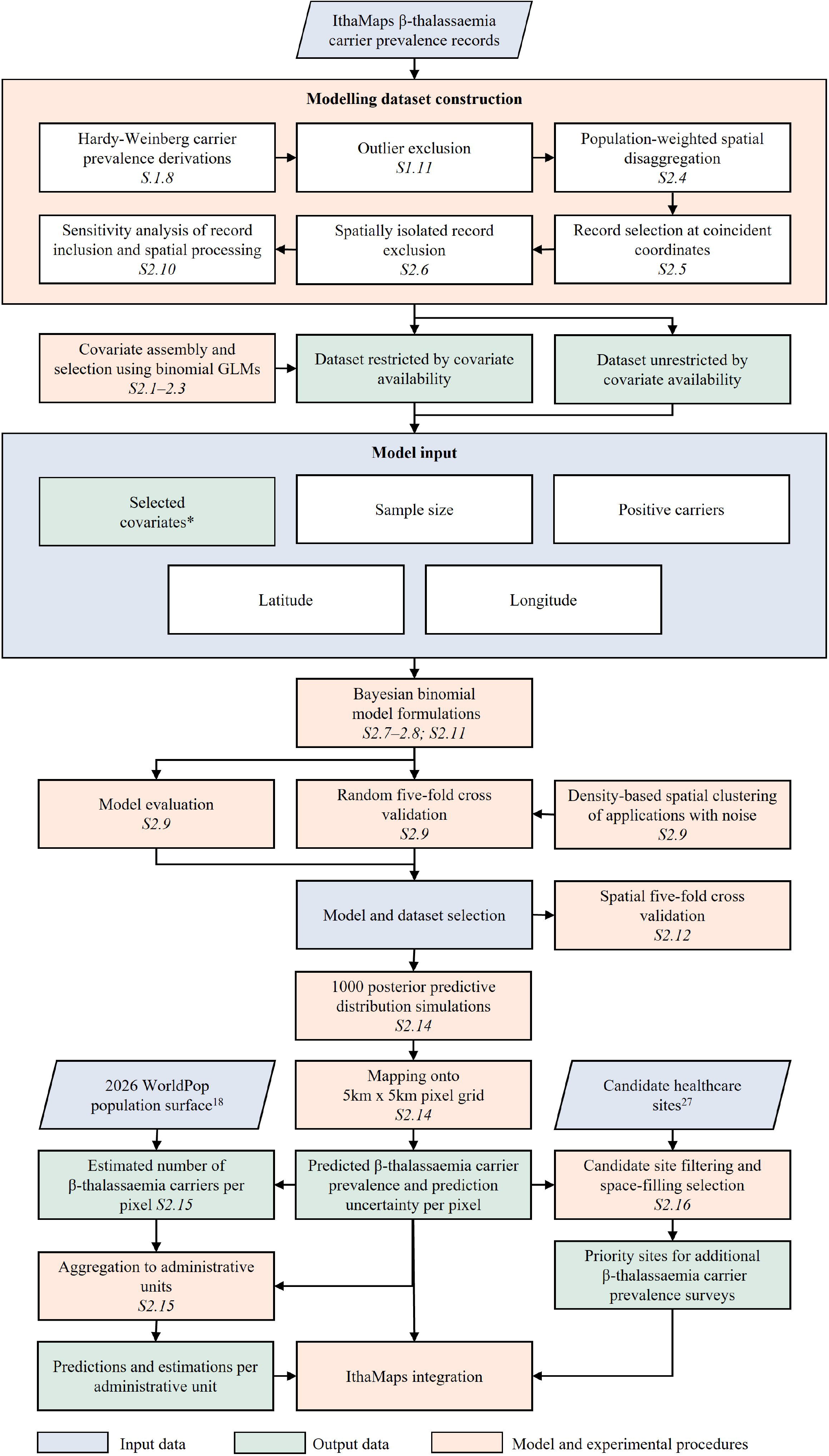
Schematic overview of the geostatistical modelling framework. GLMs: Generalised linear models. S: Supplementary section. *Selected covariates were used only in model formulations fitted to the covariate-restricted dataset.

Random five-fold CV assessed held-out predictive performance within the geographic support of the observed data, with record allocation structured by density-based clustering (Supplementary section 2.9).^25^ Within each cluster, records were randomly allocated approximately evenly across the folds; records in smaller clusters and those classified as spatial noise remained in every training set. Each fold was withheld in turn, and the model was refitted before prediction. Predictive performance was quantified using mean absolute error (MAE), root mean square error (RMSE), and Pearson correlation between observed and predicted carrier prevalence. Within each dataset, WAIC, CPO, MAE, RMSE, and Pearson correlation were interpreted jointly, with direct model comparisons restricted to formulations fitted to the same records.^9,22^ After model selection, spatial five-fold CV was applied to all three covariate-free formulations in the unrestricted dataset to contextualise the selected formulation’s performance under geographic withholding relative to alternative residual structures (Supplementary section 2.12).^26^ Clusters were consolidated into five geographically contiguous, approximately balanced folds, withheld in turn while each model was refitted and used for prediction. Performance was assessed using the same metrics as in random CV.

### Spatial prediction, carrier-number estimation, and survey prioritisation

The selected model was refitted to the unrestricted dataset and used to predict carrier prevalence across inhabited cells at approximately 5 km × 5 km resolution. At each cell, 1000 joint posterior draws were generated, with a new IID effect in each draw representing spatially unstructured heterogeneity at a new location. Carrier prevalence was summarised by the posterior mean, with prediction uncertainty quantified using the 95% credible interval, defined by the 2·5^th^ and 97·5^th^ percentiles of the posterior predictive distribution. Both were mapped across the prediction domain. To estimate carrier numbers, predicted carrier prevalence in each cell was multiplied by the corresponding 2026 WorldPop population count.^18^ Carrier-number surfaces using the cell-level 25^th^- and 75^th^-percentile carrier-prevalence estimates were calculated analogously and aggregated within ADM0, ADM1, and ADM2 units (Supplementary sections 2.14–2.15).^22^

Prediction outputs also informed priorities for additional carrier-prevalence surveys. Georeferenced healthcare sites from Healthsites.io^27^ provided candidate sites. Sites within 50 km of an existing modelling coordinate or in unpopulated cells were excluded. Remaining sites were prioritised by prediction uncertainty, population count, and predicted carrier prevalence. In countries with more than 100 candidates, sites were retained above the country-specific 75^th^ percentile of uncertainty, at or above the 25^th^ percentile of population, and at or above the median predicted carrier prevalence; otherwise, all candidates were retained. A previously established space-filling design then treated the modelling coordinates as fixed design points and selected up to 30 priority sites per country to extend geographic coverage while maintaining at least 50 km separation from existing and newly selected sites (Supplementary section 2.16).^28^ Spatial prediction, carrier estimation, and survey-prioritisation outputs were integrated into IthaMaps for interactive visualisation and download.

## Results

Among 1954 candidate β-thalassaemia carrier-prevalence records post outlier removal, sensitivity analyses supported restriction to quality-appraised records with low risk of bias, due to lower absolute prediction error under random five-fold CV. Exclusion of Hardy-Weinberg-derived or pooled multi-location records produced no consistent advantage, while population-informed spatial disaggregation produced small improvements across metrics and spatial-noise retention had little effect. After geographic processing and exclusion of records separated from the main evidence-supported domain, the final unrestricted dataset comprised 354 records from 110 sources across 25 countries (Figure 3A; Supplementary sections 2.10–2.11, tables S9–S11).

**Figure 3.**
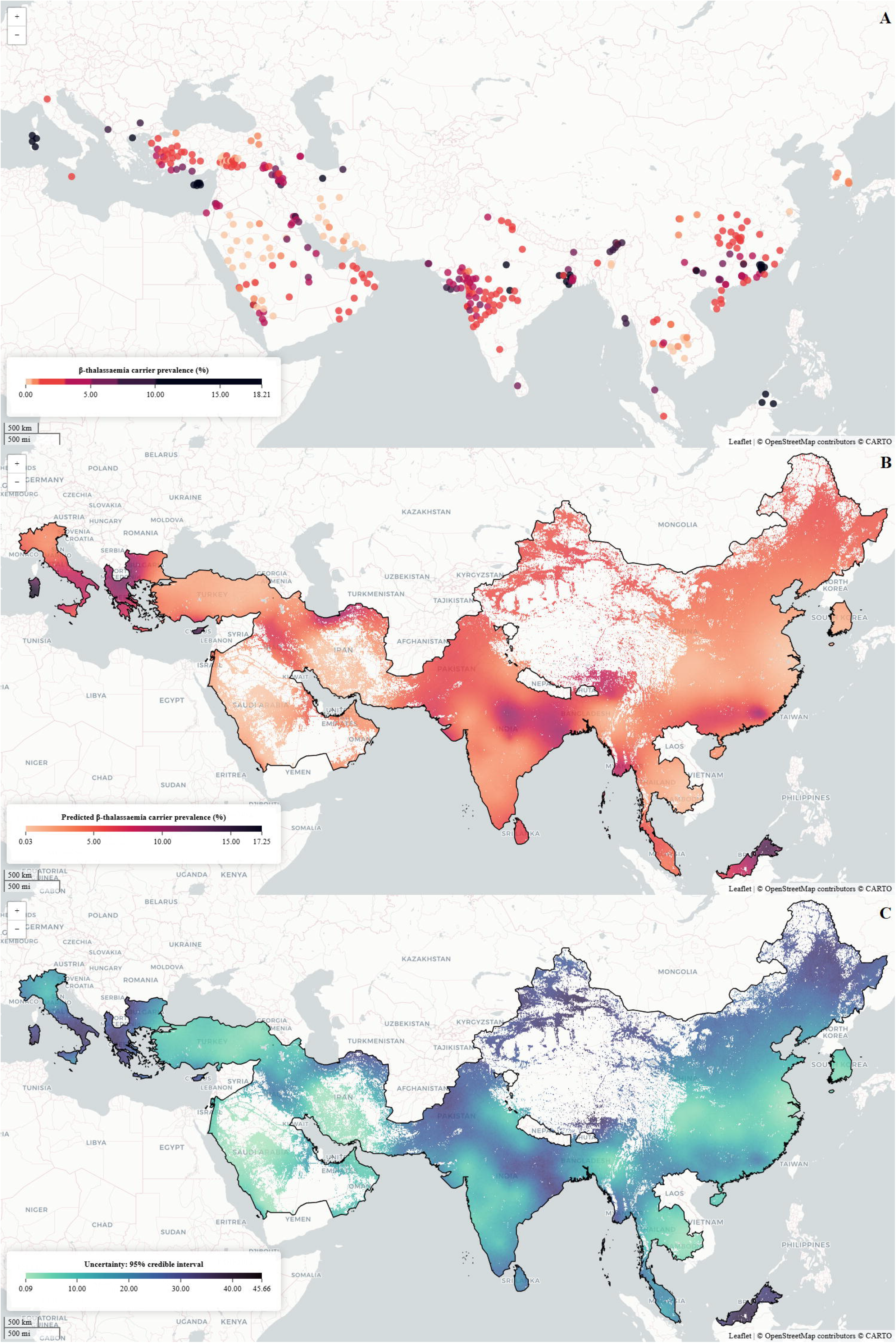
Geographic distribution of β-thalassaemia carrier-prevalence records, predicted β-thalassaemia carrier prevalence, and prediction uncertainty. A. Georeferenced β-thalassaemia carrier-prevalence records included in the unrestricted dataset; point colour indicates observed carrier prevalence, expressed as a percentage. B. Posterior mean predicted carrier prevalence across the prediction domain, expressed as a percentage. C. Prediction uncertainty, quantified as the width of the 95% credible interval. Darker shading indicates higher observed or predicted carrier prevalence and greater prediction uncertainty.

The covariate-restricted dataset comprised 290 records from 83 sources across 11 countries. Adding the selected covariates to the formulation containing spatial and IID random-effects did not improve held-out prediction: mean MAE and RMSE remained 0·016 and 0·024, respectively, while pooled Pearson correlation decreased from 0·588 to 0·571. Model development therefore proceeded with covariate-free formulations in the covariate-unrestricted dataset. The formulation containing both spatial and IID random-effects showed the strongest overall random five-fold CV performance and was retained for prediction. Mean MAE was 0·015 (SD 0·002), mean RMSE 0·023 (SD 0·005), and pooled Pearson correlation 0·688 (Table 1).

**Table 1.** Model evaluation and random five-fold cross-validation performance of candidate model formulations across alternativeβ-thalassaemia carrier-prevalence datasets.

| Dataset | Model | Evaluation metrics |  | Random cross-validation metrics |  |  |
| --- | --- | --- | --- | --- | --- | --- |
|  |  | WAIC | Mean negative log CPO (SD) | MAE (SD) | RMSE (SD) | Pearson correlation coefficient |
| Covariate-restricted | Covariate fixed-effects | 163100<br>2 | 160·226<br>(251·297) | 0·027<br>(0·004) | 0·037<br>(0·006) | 0·098 |
|  | IID random-effect | 2365 | 10·754<br>(5·617) | 0·023<br>(0·003) | 0·030<br>(0·004) | -0·017 |
|  | Covariate fixed-effects and IID random-effect | 2370 | 10·645<br>(5·543) | 0·024<br>(0·002) | 0·030<br>(0·004) | 0·185 |
|  | Spatial random-effect | 4274 | 12·113<br>(17·958) | 0·063<br>(0·041) | 0·085<br>(0·051) | 0·102 |
|  | Spatial and IID random-effects | 2339 | 8·864<br>(3·898) | 0·016<br>(0·001) | 0·024<br>(0·005) | 0·588 |
|  | Covariate fixed-effects, spatial random-effect, and IID random-effect | 2338 | 8·911<br>(3·915) | 0·016<br>(0·001) | 0·024<br>(0·005) | 0·571 |
| Covariate-unrestricted | IID random-effect | 2872 | 10·903<br>(5·787) | 0·026<br>(0·003) | 0·033<br>(0·005) | -0·027 |
|  | Spatial random-effect | 4821 | 11·609<br>(16·446) | 0·041<br>(0·011) | 0·057<br>(0·014) | 0·297 |
|  | Spatial and IID random-effects | 2846 | 9·082<br>(4·060) | 0·015<br>(0·002) | 0·023<br>(0·005) | 0·688 |

IID: independent and identically distributed. WAIC: Watanabe-Akaike information criterion. CPO: conditional predictive ordinate. MAE: mean absolute error. RMSE: root mean square error. SD: standard deviation. Covariate fixed-effect formulations included covariates retained after covariate selection. MAE and RMSE are reported as the mean with its standard deviation across the five folds, whereas a pooled Pearson correlation coefficient was calculated once from observed and out-of-fold posterior mean predicted carrier prevalence across all validation records. WAIC and mean negative log CPO were calculated from models fitted to the full dataset. Lower WAIC, mean negative log CPO, MAE, and RMSE indicate better fit or smaller prediction error; Pearson correlation coefficient values approaching 1 indicate stronger positive linear association between observed and predicted values in held-out data, whereas negative values indicate inverse association and poorer predictive performance than positive values of the same magnitude.

Performance deteriorated substantially under spatial five-fold CV. Mean MAE increased to 0·042 (SD 0·010), mean RMSE to 0·046 (SD 0·010), and pooled Pearson correlation was -0·209, with negative correlations in all five folds. Although the IID-only formulation produced smaller absolute errors, it similarly failed to reproduce the geographic pattern of carrier prevalence in withheld folds (pooled Pearson correlation -0·053). No evaluated residual formulations reliably recovered the geographic carrier-prevalence pattern when nearby records were withheld (Supplementary section 2.12, table S12).

Posterior mean predicted carrier prevalence ranged from approximately 0·03% to 17·25% across inhabited cells and showed marked within-country variation (Figure 3B). Prediction uncertainty was geographically heterogeneous and showed a spatial pattern distinct from predicted prevalence (Figure 3C). Carrier-number rankings differed from prevalence rankings; India, China, and Pakistan were estimated to contain 78·49 million, 40·84 million, and 14·38 million carriers, respectively, together accounting for 81·21% of the approximately 164·66 million carriers estimated across the prediction domain (Figure 4A). Within India, Uttar Pradesh contained an estimated 13·98 million carriers compared with 3·74 million in higher-prevalence Jharkhand. Survey prioritisation identified 183 healthcare sites across 17 countries where additional carrier-prevalence surveys could extend geographic sampling coverage beyond the existing evidence network (Figure 4B; Supplementary figure S28, table S38).

**Figure 4.**
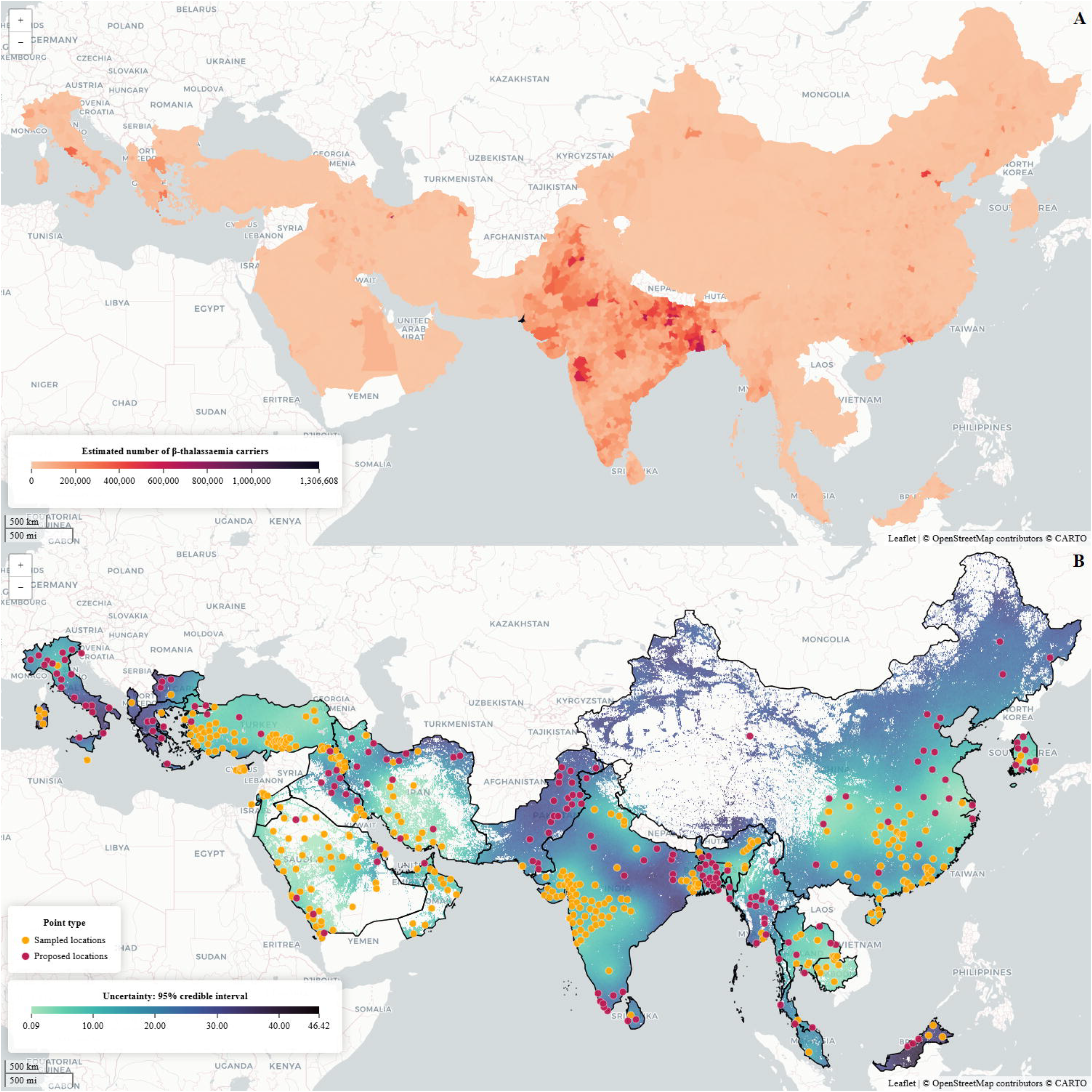
Estimated number of β-thalassaemia carriers and priority sites for future carrier-prevalence surveys. A. Estimated number of β-thalassaemia carriers across the prediction domain, derived from predicted carrier prevalence and gridded population estimates. B. Prediction uncertainty, quantified as the width of the 95% credible interval, with locations used to inform survey prioritisation. Orange points indicate sampled locations represented in the modelling dataset, and pink points indicate proposed healthcare sites prioritised for additional carrier-prevalence surveys. Darker shading indicates larger estimated carrier numbers in panel A and greater prediction uncertainty in panel B.

## Discussion

This study is the first to model β-thalassaemia carrier prevalence at high spatial resolution across 25 countries using a geospatial framework. Integrating geographically dispersed carrier-prevalence records revealed marked subnational variation and shows that the distribution of carrier frequency does not coincide with the distribution of carrier numbers. India, China, and Pakistan contained over 80% of estimated carriers despite not having the highest predicted prevalences, while within India, Uttar Pradesh contained more carriers than higher-prevalence Jharkhand. This divergence has implications for prevention. Carrier prevalence informs the expected yield of carrier detection, whereas carrier numbers indicate the population scale against which confirmatory diagnosis, partner assessment, counselling, and reproductive services can be considered.^4,6^ Neither directly measures service demand, but together they provide a more informative basis for assessing whether prevention capacity is aligned with the populations and places in which carrier risk is concentrated.

The divergence also reframes historical β-thalassaemia estimates. Modell and Darlison established the importance of relating carrier frequency to population size,^6^ while later geostatistical studies showed that haemoglobinopathy population estimates can change materially when spatial heterogeneity and contemporary population distributions are incorporated.^9,11^ HbC newborn estimates were approximately twice earlier figures,^10^ with higher estimates also reported for HbS and α-thalassaemia.^9,11^ In the present analysis, predicted prevalence remained broadly consistent with published estimates in several settings, including 12·99% versus 15·00% in Cyprus, 8·42% versus 8·00% in Greece, 5·56% versus 5·50% in Pakistan, and 3·17% versus 3·61% in Jiangxi, China.^7,29^ In India, predicted prevalence was 5·13%, compared with a pooled estimate of 3·74% from a recent systematic review that also documented substantial population heterogeneity.^30^ Although the approximately 164·66 million carriers across the 25-country prediction domain exceeds the historical worldwide estimate of 80-90 million carriers,^6^ the estimates differ in geographic coverage, population reference periods, evidence base, and analytical methods, which precludes direct comparison and interpretation of this discrepancy as temporal growth in carrier frequency. Its significance lies instead in showing that broadly familiar prevalence patterns, when resolved within countries and combined with contemporary population distributions, can translate into a substantially larger absolute carrier population than long-standing totals imply. This study therefore constitutes a contemporary reassessment across 25 countries and establishes a framework for broader global reassessment as sufficiently characterised evidence becomes available.

Resolving geographic variation within countries depends fundamentally on the quality and interpretability of the underlying evidence. Restriction to quality-appraised records at low risk of bias reduced absolute prediction error under random five-fold CV and improved CPO-based predictive support, while broad ecological, demographic, population-structure, and service-related covariates added no predictive value beyond the spatial and IID random-effects. These findings indicate that neither restrictive data processing nor generic covariate proxies can compensate for inadequately characterised primary observations. Recruitment geography, sampling frame, study period, diagnostic method, and relevant population characteristics are therefore essential for robust spatial inference and for developing more informative population-specific predictors. Future surveys should report these features with sufficient precision for independent appraisal and georeferencing, thereby determining whether their data can inform only the sampled population or contribute reliably to wider spatial inference.

Random CV showed useful predictive performance within represented geography, where held-out records remained supported by nearby prevalence records. By contrast, spatial CV deteriorated markedly when contiguous regions were withheld, and no evaluated residual formulation reliably recovered the missing spatial pattern. These results indicate that the model is most informative for interpolation within the geographic support of existing observations, while extrapolation across major unsampled gaps remains limited primarily by the absence of local carrier-prevalence records rather than residual model specification. Improving prediction in poorly represented regions will require surveys that extend geographic support. Prospective sampling guided by the presented prioritisation analysis could address this limitation with focus on 183 identified healthcare sites across 17 countries where new carrier-prevalence surveys could extend geographic coverage rather than reinforce already dense sampling.^28^ These sites provide pragmatic recruitment and diagnosis locations, although feasibility and population representativeness require local assessment.^27^ This converts a demonstrated limit of spatial prediction into a reproducible strategy for strengthening subsequent inference.

The evidence base, including records retained for and excluded from modelling, is openly accessible through IthaMaps alongside model predictions, uncertainty estimates, carrier-number estimates, and prioritised survey sites. The interface retains source provenance and supports filtering and data download, providing a transparent resource for independent appraisal, future survey design, and iterative updating as new evidence becomes available.

Several limitations warrant consideration. Geographic coverage remains uneven, and some coarse area-level records required population-informed proxy coordinates, while pooled estimates from multiple recruitment sites could contribute at more than one modelling coordinate. These procedures could increase the influence of individual estimates, although sensitivity analyses showed little effect on predictive performance (Supplementary section 2.10). The evidence also spans several decades. More recent records were prioritised at coincident locations, but migration, admixture, and changes in sampling or diagnostic practice may still introduce temporal heterogeneity.^3,4^ Temporal coverage was insufficient for spatiotemporal modelling, so the surfaces should not be interpreted as year-specific estimates. Inference is limited to the 25-country domain, and survey priorities depend on sites represented in Healthsites.io.^27^ These limitations are most consequential where evidence is sparse, coarse, or older, which are also the settings where contemporary surveys could most improve subsequent inference.

Taken together, this study provides a spatially resolved reassessment of β-thalassaemia carrier prevalence and carrier populations across 25 countries. It reveals substantial subnational heterogeneity, shows that high prevalence does not necessarily coincide with the largest carrier populations, and identifies where additional surveys could most strengthen future inference. More broadly, it provides a reproducible basis for updating spatial estimates as new evidence accumulates and for extending the framework to other haemoglobinopathies.

## Data Availability

The data curated for this study are publicly and freely available through the ITHANET portal at https://www.ithanet.eu/db/ithamaps. The sources of the covariate data are listed in the Supplementary material. The R script used for geostatistical modelling is publicly available on GitHub at https://github.com/2238154/Multicountry-geostatistical-modelling-framework-for-haemoglobinopathies.git.

https://github.com/2238154/Multicountry-geostatistical-modelling-framework-for-haemoglobinopathies.git

## Contributors

Coralea Stephanou, Mikaella Kontopoulou and Petros Kountouris were responsible for conceptualization. Chigozie Edson Utazi, Coralea Stephanou, Mikaella Kontopoulou, Petros Kountouris, and Somnath Chaudhuri were responsible for study design. Coralea Stephanou, Mikaella Kontopoulou and Petros Kountouris were responsible for source collection. Charlie Leboff, Coralea Stephanou, Mikaella Kontopoulou, and Sotiroula Chatzimatthaiou were responsible for data curation and quality appraisal. Mikaella Kontopoulou and Maria Xenophontos were responsible for updating and enhancing the infrastructure of the IthaMaps database, including data integration and interactive visualisation functionality. Chigozie Edson Utazi, Charlie Leboff, Coralea Stephanou, Kalia Orphanou, Mikaella Kontopoulou, Maria Xenophontos, Petros Kountouris and Somnath Chaudhuri developed the methodology. Mikaella Kontopoulou performed the formal analysis. Mikaella Kontopoulou prepared the visualisations. Chigozie Edson Utazi, Coralea Stephanou, Petros Kountouris and Somnath Chaudhuri supervised the work. Mikaella Kontopoulou wrote the original draft. Chigozie Edson Utazi, Coralea Stephanou, Mikaella Kontopoulou, Maria Xenophontos, Petros Kountouris and Somnath Chaudhuri had full access to all the data in this study. Chigozie Edson Utazi, Charlie Leboff, Coralea Stephanou, Carsten Werner Lederer, Kalia Orphanou, Mikaella Kontopoulou, Maria Xenophontos, Petros Kountouris, Somnath Chaudhuri and Sotiroula Chatzimatthaiou reviewed and edited the manuscript and had final responsibility for the decision to submit for publication.

## Declaration of interests

The authors declare no conflict of interest.

## Data sharing

### Acknowledgments

This work was co-funded by the European Cooperation in Science and Technology (COST) Action CA22119, Haemoglobinopathies in European Liaison of Medicine and Science (HELIOS), the European Union’s Horizon Europe programme under grant agreement No. 101159589 (HemaFAIR), the European Union’s Horizon 2020 research and innovation programme under grant agreement No. 101017549 (GenoMed4ALL), and the Cyprus Institute of Neurology and Genetics. The authors thank the Institute for Health Metrics and Evaluation for providing curated epidemiological data on haemoglobinopathies. The authors also acknowledge the ARISE initiative, through which Charlie Leboff contributed to data curation, quality appraisal, and testing of R package workflows used to derive weighted summary estimates for the interactive visualisations available through the ITHANET portal.

## Declaration of generative AI and AI-assisted technologies in the manuscript preparation process

During the preparation of this work the authors used the latest version of ChatGPT (OpenAI) available at the time of use to improve the clarity and readability of the manuscript. After using this tool, the authors reviewed and edited the content as necessary and take full responsibility for the content of the published article.

## Notes

### Competing Interest Statement

The authors have declared no competing interest.

