## Supplementary material for "Large-scale modelling of β-thalassaemia carrier prevalence: geostatistical predictions and population estimates from a global evidence base"

**List of supplementary material contents**

### 1. IthaMaps redevelopment

#### 1.1. Administrative boundaries and spatial harmonisation

Before curation and analysis, a harmonised global hierarchy of administrative boundaries was constructed. Administrative level 0 (ADM0; country or equivalent), administrative level 1 (ADM1; first-order subnational unit), and administrative level 2 (ADM2; second-order subnational unit) geometries were obtained from the geoBoundaries Comprehensive Global Administrative Zones dataset.[1] Country-specific boundary files supplemented the global layers where geographic entities were absent or inconsistently represented. Each lower-level administrative unit was assigned to its corresponding higher-level unit, such as ADM2 to ADM1, based on spatial overlap. Where a lower-level unit overlapped more than one higher-level unit, it was assigned to the unit containing the largest proportion of its area. Centroid coordinates were calculated for each administrative unit. Inconsistent names, transliterations, and special characters were reconciled manually.

The resulting boundary framework was incorporated into an R Shiny curation interface. For each country, the interface displayed synchronised ADM0, ADM1, and ADM2 layers and allowed administrative unit filtering and the overlay of reported coordinates. Pop-up fields showed the administrative unit's name, centroid coordinates, and unique geographic identifier, enabling spatial assignments to be verified during data extraction.

#### 1.2. Source identification

Potentially relevant sources were identified through two complementary search streams. First, the established ITHANET PubMed strategy developed for haemoglobinopathy curation was applied, with weekly automated alerts used to identify newly indexed sources.[2] Second, Google Scholar and PubMed were searched through Publish or Perish, version 8, on September 1, 2023.[3] This search was restricted to sources published from 1950 onwards and combined haemoglobinopathy terms with terms relating to epidemiology, genetic diversity, prevention, management, diagnostic services, and policy. Newly published sources matching the second stream were captured through automated alerts from September 2, 2023, until the literature search was closed on December 31, 2024. Complete search strategies are provided in Supplementary Figure S1.

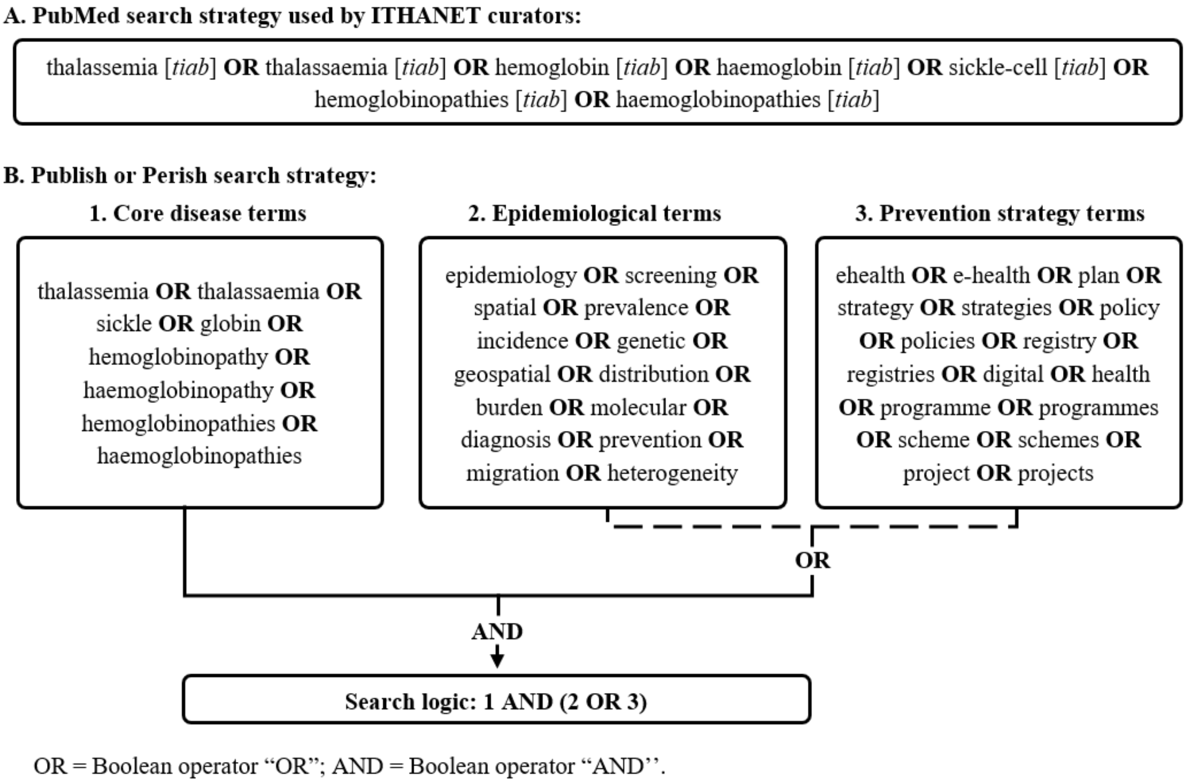

**Supplementary Figure S1. Search queries employed to identify pertinent scientific literature.**
A. PubMed search strategy used by ITHANET curators.[2] B. Publish or Perish search strategy combining core haemoglobinopathy terms with either epidemiological terms or healthcare service terms.[3]

Titles were normalised by converting characters to lower case, removing punctuation, collapsing repeated whitespace, and transliterating Greek symbols. Duplicate sources were removed using digital object identifiers or PubMed identifiers when available, followed by normalised title and publication-year matching. Potential duplicate matches that could not be resolved automatically were reviewed manually.

Prespecified title-based rules were subsequently applied to remove sources unlikely to contain information relevant to IthaMaps curation. Automated screening was restricted to titles because haemoglobinopathy terminology in abstracts frequently described contextual information rather than data reported by the study itself. Exclusion and retention expressions, their order of application, and the rules used to resolve conflicting matches are provided in Supplementary Table S1.

**Supplementary Table S1. Title-mining dictionary used for automated source exclusion and retention.**

<table>
<tr><th>Category</th><th>Purpose of rule</th><th>Exclusion terms</th><th>Retention terms</th></tr>
<tr><td>Agricultural</td><td>Exclude sources unlikely to contain human haemoglobinopathy data.</td><td>\\b blade \\b</td><td>NA</td></tr>
<tr><td>Non-human</td><td>Exclude non-human biological sources.</td><td>\\b mouse \\b | \\b mice \\b | \\b murine \\b | \\b animal | \\b rat \\b | \\b zebrafish \\b | \\b fish | \\b drosophila | \\b bovine | \\b bacterium \\b | \\b bacteria \\b | \\b microorganism | \\b organism | \\b pathogen \\b | \\b pathogens \\b | \\b mammal | \\b primate | \\b vertebrae | \\b frog \\b | \\b frogs \\b | \\b toad | \\b leaf | \\b root | \\b gill | \\b monkey | \\b bird | \\b chick | \\b laevis | \\b fung | \\b snake | \\b elegan | \\b plasmid | worm | \\b plant \\b | \\b plants \\b | \\b fungus \\b | \\b fungi \\b | \\b virus \\b | \\b parasites \\b | \\b parasite \\b | \\b bacterial \\b | \\b mullet \\b | \\b plasmodia \\b | \\b plasmodium \\b</td><td>NA</td></tr>
<tr><td>Family</td><td>Exclude sources focused on individual families rather than population or healthcare service evidence.</td><td>\\b family | \\b familial | \\b families \\b</td><td>NA</td></tr>
<tr><td>Cases</td><td>Exclude single-case sources unlikely to provide population-level evidence.</td><td>\\b case \\b | \\b a patient \\b | \\b a pediatric patient \\b | \\b a novel \\b | \\b a new \\b | \\b a mutation \\b | \\b a variant \\b | \\b an allele \\b | \\b a de novo \\b | \\b a deletion \\b | \\b boy \\b | \\b girl \\b | \\b male \\b | \\b female \\b</td><td>case study</td></tr>
<tr><td>Donor-focused</td><td>Exclude sources with blood donor cohorts which can underrepresent haemoglobinopathy carriers and affected individuals owing to anaemia-related blood donation ineligibility.</td><td>\\b donor | \\b donate</td><td>NA</td></tr>
<tr><td>Functional or mechanistic</td><td>Exclude molecular, cellular, protein, or mechanistic sources.</td><td>\\b mechanism \\b | \\b cross talkzzb | \\b cross-talk \\b | \\b evolution \\b | \\b evolv | \\b lineage \\b | \\b transposon | \\b protein stabil | \\b membrane \\b | \\b cell culture \\b | \\b protein profile \\b | \\b protein sequence \\b | \\b myoglobin \\b | \\b receptor \\b | \\b receptors \\b | \\b binding \\b | \\b vesicle | \\b transcript | \\b translat | \\b kinase | \\b stabl | \\b stabil | \\b phylogen | \\b functional | \\b glycated | \\b fiber | \\b nitric oxide \\b | \\b propert | \\b cellular | \\b acceptor | \\b ligand | \\b ribosom | \\b engineer | \\b affinity | \\b cataly | \\b proliferat | \\b proteome | \\b fold | \\b sense | \\b signal | \\b nucleosome | \\b isoform | \\b protein | \\b antibodies \\b | \\b antibody \\b | \\b pathway</td><td>status</td></tr>
<tr><td>Other disease or complication</td><td>Exclude sources focused primarily on diseases, complications, symptoms, biomarkers, or comorbidities.</td><td>\\b hip \\b | \\b hiv \\b | \\b hpv \\b | \\b covid \\b | \\b meningitis \\b | \\b rheuma | \\b osteo | \\b kidney disease \\b | \\b cardio | \\b renal | \\b onto | \\b lung | \\b diabetic | \\b cancer | \\b hepatitis | \\b papillo | \\b retino | \\b stroke | \\b cerebro | \\b vascular | \\b infection | \\b oxidati | \\b hyphema | \\b arterio | \\b arterial | \\b dental | \\b caries | \\b artery | \\b depression | \\b diabetes | \\b metabolic | \\b metabolism | \\b psycho | \\b anxiety | \\b infectious | \\b bladder | \\b fever | \\b liver disease | \\b gallstone | \\b diet | \\b ocular | \\b thrombo | \\b pulmonary | \\b bone | \\b vitamin | \\b nutrition | \\b dwarf | \\b athero</td><td>\\b simultaneous | \\b reproductive behaviour \\b | \\b reproductive age \\b | \\b prevention of beta | \\b Sickle-cell disease in febrile | \\b , thalass | \\b and thalass | \\b , sickle cell | \\b and sickle cell | \\b , sickle-cell | \\b and sickle-cell | \\b , hemoglobin \\b | \\b and hemoglobin \\b | \\b , haemoglobin \\b | \\b and</td></tr>
</table>

<table>
<tr><th>Category</th><th>Purpose of rule</th><th>Exclusion terms</th><th>Retention terms</th></tr>
<tr><td></td><td></td><td>| \\b athro | \\b arthro | \\b hormone | \\b hypertension | \\b heart | \\b cardia | \\b sleep | \\b cholelithiasis | \\b inflamma | \\b microbi | \\b priapism | \\b neuro | \\b endo | \\b ovar | \\b maculo | \\b hepatopathy | \\b cough | \\b pneumonia | \\b glioma | \\b retina | \\b wound | \\b gonorrhoeae | \\b eyelid | \\b tumor | \\b craniofacial | \\b cognitive | \\b sclerosis | \\b war | \\b pneumococcal | \\b penile | \\b metabolopathies | \\b nephro | \\b femur | \\b spondylodiscitis | \\b short stature | \\b white matter | \\b carcinoma | \\b limb | \\b growth | \\b toxic | \\b oral | \\b salmonella | \\b testic | \\b tuberculosis | \\b endocrine | \\b leukaemia | \\b puberty | \\b exposure to | \\b hazard | \\b cavities | \\b ischemia | \\b thyroid | \\b malign | \\b clamydia | \\b dyspnea | \\b gout | \\b moyamoya | \\b leukemia | \\b lympho | \\b brain \\b | \\b influenza | \\b flu | \\b hyperplasia | \\b seizure | \\b myocardia | \\b pica | \\b cholesterol | \\b lipid | \\b hypogonad | \\b iron | \\b ferritin | \\b pain \\b | \\b symptom \\b | \\b complication | \\b hemorrhage \\b | \\b fatigue \\b | \\b pigment \\b | \\b pulse \\b | \\b hernia \\b | \\b quality of life \\b | \\b splen | \\b spleen | \\b emergency \\b | \\b blood pressure \\b | \\b kidney \\b | \\b liver \\b | \\b blood count \\b | \\b load | \\b zinc | \\b copper | \\b calcium | \\b oxygen | \\b pathophysio | \\b obstruct | \\b emotion | \\b behavi | \\b psycholog | \\b suffer | \\b serum | \\b plasma | \\psychiatric | \\b abdominal | \\b gallbladder | skeletal | virus | \\b hypothyroidism | \\b glucose \\b | \\b glucose- | \\b g6pd \\b | \\b g-6-pd \\b | \\b malaria \\b | \\b menstrual \\b | \\b bacteriuria \\b | \\b haematuria \\b | \\b microalbuminuria \\b | \\b folate \\b | \\b bacteremia \\b | \\b crisis \\b | \\b height \\b | \\b cormic \\b | \\b malnutrition \\b | \\b micronutrient | \\b body composition \\b | \\b glutathione\\b | cardio | \\b manifestation | respiratory | ulcer | \\b proteinuria \\b | \\b femoral \\b | \\b cephalohaematoma \\b | \\b anthropometric \\b | \\b filtration \\b | \\b electrolytes \\b | \\b hepatobiliary \\b | \\b stress \\b | \\b cephalus \\b | \\b crises \\b | \\b enuresis \\b | \\b folic acid \\b | \\b hepatosplenomegaly \\b | \\b necrosis \\b | \\b auditory \\b | \\b renal \\b | \\b urinary \\b | \\b stunting \\b | \\b wasting \\b | \\b autosplenectomy \\b | \\b velocity \\b | \\b menstruation \\b | \\b morbidities \\b</td><td>haemoglobin \\b | \\b , hemoglobinopath | \\b , haemoglobinopath | \\b and haemoglobinopath | \\b and hemoglobinopath | \\b and other haemoglobinopath | \\b and other hemoglobinopath | trait and \\b | \\b hemoglobinopathies and \\b | \\b haemoglobinopathies and\\b | -thalassemia and\\b | -thalassaemia and\\b | -thalassemia, \\b | -thalassaemia, \\b | thalassemia and\\b | thalassaemia and\\b | thalassemia, \\b | thalassaemia, \\b | \\b hemoglobinopathies, \\b | \\b haemoglobinopathies, \\b | \\b cell anemia, \\b | \\b cell anaemia, \\b | \\b cell anemia and \\b | \\b cell anaemia and\\b | cell-anemia, \\b | cell-anaemia, \\b | cell-anemia and\\b | cell-anaemia and \\b</td></tr>
<tr><td>Anaemia not specific to sickle cell disease</td><td>Exclude general anaemia sources not clearly focused on haemoglobinopathies.</td><td>\\b anemia \\b | \\b anemic \\b | \\b anaemia \\b | \\b anaemic \\b</td><td>cell-anemia\\b | cell-anaemia\\b | cell anemia\\b | cell anaemia \\b</td></tr>
<tr><td>Awareness</td><td>Exclude awareness-centered sources.</td><td>\\b education | \\b aware | \\b percept | \\b teach | \\b attitude | \\b knowledge | \\b of information | \\b perception | \\b attitude | \\b belief</td><td>screening | prevention</td></tr>
<tr><td>Quality of life</td><td>Exclude sources focused on patient quality of life.</td><td>\\b cost of | \\b economic burden | \\b access | \\b satisfaction | \\b needs of | \\b insurance | \\b distance to \\b | \\b quality of care | \\b hospital admission | \\b experiences \\b | \\b financial burden</td><td>NA</td></tr>
<tr><td>Mortality</td><td>Exclude sources focused on survival, mortality, morbidity, or clinical outcomes.</td><td>\\b mortality | \\b morbidity | \\b longitudinal | \\b survival | \\b life expectancy | \\b long term \\b | \\b long-term \\b | \\b matérnal outcome | \\b perinatal outcome | \\b pregnancy outcome | \\b death \\b | \\b die \\b | \\b dying \\b</td><td>NA</td></tr>
<tr><td>Modifiers</td><td>Exclude sources focused on phenotype modifiers.</td><td>\\b modifier | \\b xmn</td><td>NA</td></tr>
<tr><td>Predictive</td><td>Exclude modelling or algorithmic sources not providing curatable source-level epidemiological data.</td><td>\\b machine | \\b predict | \\b model | \\b algorithm | \\b equation | \\b math | \\b simulat</td><td>NA</td></tr>
<tr><td>Treatment</td><td>Exclude therapeutic, pharmacological, gene-editing, or clinical-trial sources without relevant epidemiological or</td><td>\\b drug | \\b therapy | \\b therapeutic | \\b treatment | \\b clinical trial | \\b pharma | \\b vaccination | \\b immunization | \\b immunity | \\b antibody | \\b stem cell | \\b crispr | \\b hydroxyurea | \\b splenectomy | \\b immune system | \\b surgery | \\b ketamine | \\b</td><td>\\b epidemiolo | \\b treatment center \\b | \\b herbal | \\b herbs | \\b biomarker</td></tr>
</table>

| Category | Purpose of rule | Exclusion terms | Retention terms |
|---|---|---|---|
| | healthcare service availability data. | steroids \| \\b modification \| \\b target \\b \| \\b supplement \\b \| \\b supplements \\b \| \\b treat \| \\b chelat \| \\b deferiprone \| \\b antioxidant \| \\b enhance \| \\b marker \| \\b surgical \| \\b opioid \| \\b edit \| \\b agent \| \\b glutamine \| \\b luspatercept \| \\b emoxypine \| \\b therapies \| \\b xanthine \| \\b gene correction \| \\b correct \| \\b inhibit \| \\b meisoindigo \| \\b transfect \| \\b mrna \| \\b silence \| \\b anaesthetic \| \\b vector \| \\b thalidomide \| \\b transplant \| \\b repress \| \\b repair \| \\b alloimmuniz \| \\b reverse \| \\b interleukin | |
| Epidemiological and healthcare service availability | Retain sources likely to contain curatable IthaMaps evidence. | NA | prevalence \| incidence \| distribution \| epidemiology \| epidemiological \| frequency \| frequencies \| frequent \| spectrum \| screening \| screen \| diagnosis \| diagnosing \| diagnozing \| analysis \| analysing \| analyzing \| characterization \| characterisation \| characterizing \| characterisign \| identifying \| identification \| detect \| detection \| detecting \| evaluating \| evaluation \| profile of \| status \| spread \| variation \| variations \| pattern \| patterns \| proportion \| proportions \| phenotypic expression \| genotypic expression \| strategy \| strategies \| services \| service \| management \| manage \| managing \| prevention \| prevent \| preventing \| control \| controlling \| intervention \| intervening \| interventions \| intervent \| plan \| plans \| program \| programs \| project \| projects \| counsel \| counselling \| welfare \| bpublic health \| terminate \| termination \| e-health \| ehealth \| e health \| digital \| electronic \| policy \| policies \| registries \| registy \| biobank \| blood bank \| banking \| biorepository \| repository \| biorepositories \| repositories \| guide \| guideline \| strategy \| strategies |

The symbol | denotes “or” and \\b denotes a word boundary used to match a term as a standalone word rather than as part of a longer word. NA: not applicable as no exception or retention term was applied.

**1.3. Source prioritisation**

Geographic entities were extracted from source titles by named-entity recognition and geocoding. Place names were mapped to countries and continents, with manual review of historical, regional, and ambiguous geographic labels. Sources without a confidently assigned country or continent were retained in the source archive but were not included in geographic prioritisation.

Prioritisation determined the order of screening rather than eligibility. The highest priority was assigned to β-thalassaemia sources containing epidemiological terminology and originating from countries with sparse existing β-thalassaemia epidemiological evidence. Lower priority was assigned sequentially to sources with less specific β-thalassaemia terminology, without epidemiological terminology, or from countries with more existing β-thalassaemia epidemiological evidence. Within each priority stratum, newer sources were screened first, and countries were alternated to prevent high-volume settings from dominating curation. By the analytical freeze date, 1094 (23%) of 4744 sources in the prioritisation queue had been screened.

#### 1.4. Source screening

Sources were screened when reached in the prioritisation queue. Initial screening established whether the source could be retrieved and whether its full text was available. Retrieved sources were then assessed for interpretable language, source type, and relevance to haemoglobinopathy epidemiology, genetic diversity, or prevention and management services.

Sources were excluded when their language could not be interpreted by the curation team, when their content fell outside the IthaMaps scope, or when their publication format was ineligible. Ineligible formats comprised student theses, letters to the editor, conference abstracts, and posters. Sources satisfying all screening criteria proceeded to structured data extraction.

#### 1.5. Curation schema and terminology

Data extracted from each source were recorded in interconnected Google Sheets workbooks using controlled fields and prespecified terminology. Each distinct data entry constituted a record, and a single source could contribute multiple records. The curation schema captured source provenance, extraction status, epidemiological and genetic outcomes, population characteristics, recruitment period and setting, diagnostic methods, quality appraisal variables, and healthcare service characteristics. Records were additionally grouped into three broad content areas according to the information they contained. Epidemiology encompassed population-frequency measures, genetic diversity encompassed information on haemoglobinopathy variants and their distribution, and service availability encompassed information on prevention, diagnosis, management, and other haemoglobinopathy-related healthcare services. Field definitions and permissible values are provided in Supplementary Tables S2–S6.

**Supplementary Table S2. IthaMaps curation field dictionary.**

| Domain | Field | Data type | Definition | Options | Additional information |
|---|---|---|---|---|---|
| Provenance | doi | Text | Digital object identifier of the source. | NA | NA |
| Provenance | pmid | Numeric | PubMed identifier of the source. | NA | NA |
| Provenance | report | Text | Alternative source identifier used when doi and pmid were unavailable. | NA | NA |
| Provenance | curated_by | Text | Curator responsible for source screening and data extraction. | NA | NA |
| Provenance | expert | Text | External individual or team responsible for source screening, where applicable. | NA | NA |
| Epidemiological; Healthcare service | measure_name | Categorical | Epidemiological measure applicable to the record. | Controlled measure terms; see Supplementary Table S3. | NA |
| Epidemiological; Healthcare service | cause_name | Categorical | Haemoglobinopathy applicable to the record. | Controlled cause terms; see Supplementary Table S4. | NA |
| Epidemiological | ithaID | Numeric | IthaGenes identifier for the specific variant. | NA | Used for relative allele frequency records. |
| Epidemiological | locus | Categorical | Gene locus assigned from IthaGenes variant annotations. | α-locus; β-locus. | Used for relative allele frequency records. |
| Epidemiological | globin_phenotype | Categorical | Allele phenotype used for variant grouping, assigned from IthaGenes variant annotations. | α0; α+; β0; β+; α-thalassaemia modifier; non-deletional α+. | Used for allele frequency and relative allele frequency records. |
| Epidemiological | metric_name | Categorical | Unit of the value field, as reported for record consistency. | number; percent; rate. | NA |
| Epidemiological | sample_size | Numeric | Number of participants, births, embryos, fetuses, or alleles tested for the specified outcome. | NA | Retained as reported when compatible. Recomputed when needed for metric conversion, derivation |

| Domain | Field | Data type | Definition | Options | Additional information |
|---|---|---|---|---|---|
| | | | | | from numerator and outcome value, aggregation or disaggregation across spatial or demographic strata, Hardy–Weinberg derivation, or alignment with Supplementary Table S5 definitions. |
| Epidemiological | count | Numeric | Number of individuals, births, embryos, fetuses, or alleles positive for the specified outcome. | NA | Retained as reported when compatible. Recomputed when needed for metric conversion, derivation from denominator and outcome value, aggregation or disaggregation across spatial or demographic strata, Hardy–Weinberg derivation, or alignment with Supplementary Table S5 definitions. |
| Epidemiological | value | Numeric | Measurement value for the specified outcome. | NA | Retained as reported when compatible. Recomputed when needed for metric conversion, derivation from numerator and denominator, aggregation or disaggregation across spatial or demographic strata, Hardy–Weinberg derivation, or alignment with Supplementary Table S5 definitions. |
| Epidemiological; Healthcare service | diagnostic_method | Categorical | Diagnostic approach used to identify the cause. | molecular diagnosis; biochemical or haematological diagnosis with haemoglobin A2 measurement; biochemical or haematological diagnosis without haemoglobin A2 measurement; self-reported or other; unspecified. | NA |
| Demographic | status_group | Categorical | Participant status at recruitment. | patients; carriers; both; screening population; unspecified. | NA |
| Demographic | nationality | Categorical | Participant nationality or country of residence. | Country names. | Recorded as reported; inferred from recruitment country only when no separate nationality or residence information was available. |
| Demographic | race | Categorical | Race of recruited participants, as reported. | Caucasian; Asian; Latino; African descent; Native American; Middle Eastern; Indigenous; Pacific Islander; multiple or mixed; unspecified. | NA |

| Domain | Field | Data type | Definition | Options | Additional information |
|---|---|---|---|---|---|
| Demographic | ethnicity_name | Text | Ethnicity of recruited participants, as reported. | NA | Retained as source-reported text to avoid unsupported harmonisation. |
| Demographic | religion_name | Text | Religion of recruited participants, as reported. | NA | Retained as source-reported text to avoid unsupported harmonisation. |
| Demographic | age_group_name | Categorical | General age group of recruited participants. | adults; adults and children; all ages; children; embryos; fetuses; individuals of reproductive age; neonates; neonates and children; neonates and individuals of reproductive age; other; unspecified. | NA |
| Demographic | min_age | Text | Youngest recruited age, as reported. | NA | Units were recorded as y for years or m for months. |
| Demographic | max_age | Text | Oldest recruited age, as reported. | NA | Units were recorded as y for years or m for months. |
| Demographic | sex_group_name | Categorical | Sex composition of recruited participants. | both; female; male; unspecified. | NA |
| Demographic | n_female | Numeric | Number of female participants recruited. | NA | NA |
| Demographic | n_male | Numeric | Number of male participants recruited. | NA | NA |
| Demographic | consanguinity | Numeric | Percentage of participants related by blood up to the third cousin level. | NA | Recorded only when reported or directly calculable. |
| Spatial | ADM0 | Categorical | Administrative level 0 unit for the recruitment or healthcare service applicability location | Administrative level 0 unit names. | NA |
| Spatial | geo_admin0 | Categorical | geoBoundaries-derived identifier for the administrative level 0 unit. | geoBoundaries identifiers. | NA |
| Spatial | ADM1 | Categorical | Administrative level 1 unit for the recruitment or healthcare service applicability location. | Administrative level 1 unit names. | When a higher resolution locality was reported but no matching boundary was available, the nearest available administrative level was used as the location proxy. |
| Spatial | geo_admin1 | Categorical | geoBoundaries-derived identifier for the administrative level 1 unit. | geoBoundaries identifiers. | NA |
| Spatial | ADM2 | Categorical | Administrative level 2 unit for the recruitment or healthcare service applicability location | Administrative level 2 unit names. | When a higher resolution locality was reported but no matching boundary was available, the nearest available administrative level was used as the location proxy. |
| Spatial | geo_admin2 | Categorical | geoBoundaries-derived identifier for | geoBoundaries identifiers. | NA |

| Domain | Field | Data type | Definition | Options | Additional information |
|---|---|---|---|---|---|
| | | | the administrative level 2 unit. | | |
| Spatial | recruitment_site | Text | Institute or organisation reported for the recruitment location or healthcare service applicability location | NA | When no specific recruitment location was reported, the author institution or ethics approval institution was used as the location proxy. |
| Spatial | latitude | Numeric | Latitude coordinate for the recruitment location or healthcare service applicability location | NA | Site coordinates were used when identifiable using Google Maps; otherwise, the centroid of the highest matched administrative unit was used. |
| Spatial | longitude | Numeric | Longitude coordinate for the recruitment location or healthcare service applicability location | NA | Site coordinates were used when identifiable using Google Maps; otherwise, the centroid of the highest matched administrative unit was used. |
| Spatial | MaxADM | Categorical | Highest spatial resolution assigned to the record. | ADM0; ADM1; ADM2; site. | NA |
| Spatial | Multilocation | Categorical | Indicates whether the record was associated with more than one recruitment or healthcare service location. | yes; no. | Multi-location records were duplicated across reported locations when location-specific information was unavailable. |
| Temporal | start_year | Numeric | Year participant recruitment started or healthcare service implementation began. | NA | If recruitment timing was unavailable for an epidemiological record, publication year was used as a temporal proxy. |
| Temporal | end_year | Numeric | Year participant recruitment ended or healthcare service implementation ended. | NA | NA |
| Quality appraisal | score | Numeric | Proportion of applicable quality appraisal criteria satisfied. | NA | Calculated only for records eligible for quality appraisal. |
| Quality appraisal | bias_flag | Categorical | Risk of bias category assigned from the quality appraisal score. | low; medium; high; quality not appraised. | Records outside the appraisal framework were labelled with quality not appraised. |
| Healthcare service | hcp_name_ancesto r | Categorical | Broad type of healthcare service. | Controlled type terms; see Supplementary Table S6. | NA |
| Healthcare service | hcp_name | Categorical | Specific subtype of healthcare service. | Controlled subtype terms; see Supplementary Table S6. | NA |
| Healthcare service | implementation | Categorical | Implementation state of healthcare service. | policy; service; pilot. | NA |
| Healthcare service | coverage | Categorical | Geographic coverage of the healthcare service. | regional; universal; unclear. | NA |
| Healthcare service | eligibility | Categorical | Eligibility status for healthcare service utilization or access. | on request; targeted; universal; unclear; illegal; unavailable. | NA |

| Domain | Field | Data type | Definition | Options | Additional information |
|---|---|---|---|---|---|
| Healthcare service | application | Categorical | Whether participation or application was voluntary or mandatory. | mandatory; voluntary; unspecified. | NA |
| Healthcare service | compensation | Categorical | Funding source for healthcare service. | government; private insurance; public insurance; insurance unspecified; out of pocket; mixed government and insurance; mixed government and out of pocket; mixed insurance and out of pocket; mixed government, insurance, and out of pocket; mixed government and non-governmental organisation; non-governmental organisation; unspecified. | NA |
| Healthcare service | uptake | Numeric | Percentage of the eligible population with service access who used or participated in the healthcare service. | NA | NA |

IthaGenes is the haemoglobinopathy variant database within the ITHANET community portal. NA: not applicable.

**Supplementary Table S3. Epidemiological measure terms.**

| Parameter | Numerator | Denominator | Metric | Notes |
|---|---|---|---|---|
| Disease prevalence | Affected individuals | Screened population | Percent | NA |
| Carrier prevalence | Individuals with trait | Screened population | Percent | Individuals carrying more than one haemoglobinopathy trait, including co-inherited α- and β-thalassaemia, were included where reported. |
| Incidence | Affected live births | Screened live birth population | Affected births per 100,000 live births | NA |
| Allele frequency | Specific variant alleles | All screened alleles at the relevant locus | Percent | Includes wild-type and variant alleles in denominator |
| Relative allele frequency | Specific variant alleles | All detected haemoglobinopathy-related variant alleles at the relevant locus, as reported in each study | Percent | Includes only variant alleles in denominator |
| Prenatal disease prevalence | Affected foetuses | Screened foetus population | Percent | Foetus refers to human development in utero until birth. |
| Prenatal carrier prevalence | Foetuses with trait | Screened foetus population | Percent | Foetus refers to human development in utero until birth. |
| Preimplantation disease prevalence | Affected embryos | Screened embryo population | Percent | Embryo refers to human development up to the 8 cell stage before implantation in the uterine wall. |
| Preimplantation carrier prevalence | Embryos with trait | Screened embryo population | Percent | Embryo refers to human development up to the 8 cell stage before implantation in the uterine wall. |

NA: not applicable.

**Supplementary Table S4. Haemoglobinopathy cause terms.**[4–6]

| Cause term | Curation group | Ontology identifier |
|---|---|---|
| Thalassaemia | Broad haemoglobinopathy group | MONDO:0000984 |
| Sickle cell disease | Broad haemoglobinopathy group | MONDO:0017146; SCDO:1000106 |
| Haemoglobinopathy | Broad haemoglobinopathy group | SCDO:0000552 |
| Alpha thalassaemia | Quantitative haemoglobinopathy | MONDO:0011399 |
| Beta thalassaemia | Quantitative haemoglobinopathy | MONDO:0019402 |
| Delta thalassaemia | Quantitative haemoglobinopathy | SCDO:0000344 |
| Sickle cell disease-SS | Qualitative haemoglobinopathy | NCIT:C131682 |
| Haemoglobin E disease | Qualitative haemoglobinopathy | MONDO:0016243 |
| Haemoglobin C disease | Qualitative haemoglobinopathy | MONDO:0016242 |
| Sickle cell disease-SC | Compound haemoglobinopathy | MONDO:0016669 |
| Sickle cell disease-SE | Compound haemoglobinopathy | MONDO:0016671 |
| Sickle beta thalassaemia | Compound haemoglobinopathy | MONDO:0016668 |
| Haemoglobin C/beta thalassaemia disease | Compound haemoglobinopathy | MONDO:0016490 |
| Haemoglobin E/beta thalassaemia disease | Compound haemoglobinopathy | MONDO:0016491 |
| Delta beta thalassaemia | Compound haemoglobinopathy | MONDO:0016489 |

**Supplementary Table S5. Thalassaemia genotype-to-status interpretation rules.**

| Haemoglobinopathy | Genotype | Functional α-globin genes | Contributing α-globin variants | Status interpretation |
|---|---|---|---|---|
| α-thalassaemia | αα/αα | 4 | NA | Normal |
| α-thalassaemia | -α/αα;<br>$\alpha\alpha^{ND}/\alpha\alpha$ | 3 | 1 $\alpha^{+}$ | Trait |
| α-thalassaemia | --/αα | 2 | 1 $\alpha^{0}$ | Trait |
| α-thalassaemia | -α/-α;<br>$\alpha\alpha^{ND}/\alpha\alpha^{ND}$;<br>$–\alpha/\alpha\alpha^{ND}$ | 2 | 2 $\alpha^{+}$ | Trait |
| α-thalassaemia | --/-α;<br>$--/\alpha\alpha^{ND}$ | 1 | 1 $\alpha^{0}$ with 1 $\alpha^{+}$ | Haemoglobin H disease |
| α-thalassaemia | --/-- | 0 | 2 $\alpha^{0}$ | Haemoglobin Bart's hydrops fetalis |
| β-thalassaemia | β/β | NA | NA | Normal |
| β-thalassaemia | $\beta/\beta^{++}$;<br>$\beta/\beta^{+}$;<br>$\beta/\beta^{0}$ | NA | NA | Thalassaemia minor or Trait (asymptomatic or carrier state) |
| β-thalassaemia | $\beta^{++}/\beta^{++}$;<br>$\beta^{++}/\beta^{+}$;<br>$\beta^{++}/\beta^{0}$;<br>$\beta^{+}/\beta^{+}$;<br>$\beta^{+}/\beta^{0}$;<br>$\beta^{0}/\beta^{0}$ | NA | NA | Thalassaemia intermedia to major (variable transfusion dependence) |

$\alpha^{+}$ variants may be deletional (-α) or non-deletional ($\alpha^{ND}$). Superscript + indicates reduced globin-chain synthesis, and superscript 0 indicates absent globin-chain synthesis. NA: not applicable.

**Supplementary Table S6. Healthcare service types and subtypes.**

| Type | Subtypes |
|---|---|
| Newborn screening | Neonatal screening |
| Prevention strategy | Antenatal screening;<br>Cascade screening;<br>Educational campaign;<br>Preconception screening;<br>Premarital screening;<br>Prenatal or antenatal screening where distinction was unclear;<br>School-based screening;<br>Targeted screening;<br>Unspecified screening |

| Type | Subtypes |
|---|---|
| Prenatal or preimplantation genetic diagnosis | Non-invasive prenatal screening;<br>Preimplantation genetic diagnosis;<br>Prenatal genetic diagnosis by chorionic villus sampling, amniocentesis, or cordocentesis |
| Ethical, legal, and social issues | Carrier status disclosure;<br>Data protection policy;<br>Follow-up for neonates screening positive;<br>Reporting of unintentional findings;<br>Storage of blood samples or spots;<br>Termination of pregnancy act;<br>Translators for diagnosis reporting and counselling |
| Counselling services | Counselling by certified medical geneticist;<br>Counselling by other health professional;<br>Counselling by unspecified personnel |
| Social welfare | Social assistance services, including financial or employment assistance;<br>Psychological support services |
| Therapeutic treatment | Bone marrow transplantation;<br>Gene therapy |
| Iron chelation | Deferasirox;<br>Deferiprone;<br>Deferoxamine |
| Other treatments | Anti-pneumococcal immunisation;<br>Crizanlizumab;<br>Fever investigation or empirical antibiotic treatment protocols;<br>Folic acid;<br>Generic drugs;<br>Hepatitis C antiviral therapy;<br>Hydroxyurea;<br>L-glutamine;<br>Luspatercept;<br>Opioids;<br>Oral penicillin V;<br>Other immunisations; |
| Specialist services | Blood transfusion;<br>Dual x-ray absorptiometry monitoring;<br>FibroScan monitoring; electrocardiography monitoring;<br>Magnetic resonance imaging monitoring;<br>Paediatric to adult care transition services;<br>Transcranial Doppler monitoring |
| Diagnostic services | Haematological or biochemical diagnosis;<br>Molecular diagnosis |
| Quality management | Antigen-matched transfusion;<br>Erythrocytapheresis;<br>Pathogen-screened transfusion blood;<br>Trained diagnostic personnel;<br>Trained personnel for neurological complications;<br>Trained personnel for pain and vaso-occlusive crisis care |
| Healthcare infrastructure | Biobank;<br>Patient association;<br>Patient registry;<br>Specialised centre |

#### 1.6. Georeferencing

Each record was assigned to the most precise spatial unit supported by the information reported for that record. When a recruitment site was identifiable, coordinates were obtained from digital gazetteers and checked against the corresponding ADM0, ADM1, and ADM2 boundaries. When only an area-level location was reported, the record was linked to the corresponding administrative unit, and its centroid was used as the representative coordinate. Records for which the recruitment location was inferred from an author institution or ethics review institution were explicitly flagged as having an inferred recruitment location. For records reporting one pooled estimate across several recruitment locations, all identifiable recruitment sites were georeferenced.

#### 1.7. Harmonisation with curated data from the Institute for Health Metrics and Evaluation

In addition to sources manually curated for IthaMaps, records shared by the Institute for Health Metrics and Evaluation (IHME) were harmonised through a separate workflow. Harmonisation was restricted to records reporting prevalence or incidence of thalassaemia or sickle cell disorders. Disease terms were reviewed and mapped to the standard IthaMaps haemoglobinopathy categories listed in Supplementary Table S4. Records that could not be mapped to a recognised category were excluded. Records mapped only to broad, non-specific haemoglobinopathy categories were retained and flagged as having limited phenotype specificity. Recruitment years and age ranges identified by IHME as assumed were set to missing. Geographic fields were matched to the harmonised boundary framework at ADM0, ADM1, or ADM2 where possible, and the corresponding identifiers and centroid coordinates were assigned.

Quality appraisal criteria were reconstructed only when sufficient source information was available. Criteria that could not be reconstructed were recorded as unavailable. IHME curated records were not included in the quality appraisal scores pending manual supplementation. Cross-database duplicates were identified using bibliographic identifiers and normalised title and publication-year matching. When the same source was represented in both data collections, the manually curated IthaMaps records were retained.

#### 1.8. Hardy-Weinberg derivation of complementary prevalence measures

Complementary carrier prevalence and disease prevalence records were derived under Hardy-Weinberg equilibrium for both manually curated IthaMaps records and harmonised IHME records, where applicable.[7] Derivation was restricted to records from general populations and was undertaken only when the complementary measure was not already available for the corresponding record and haemoglobinopathy category. Derived records were flagged as recalculated, and the direction of derivation was recorded. Derivations were not applied to records from cohorts of suspected cases or known carriers, mixed cohorts that included known patients, patient-only cohorts, transfusion-dependent cohorts, or other clinically selected populations.

Derivations assumed the standard two-allele Hardy-Weinberg model, in which $p + q = 1$ and $p^2 + 2pq + q^2 = 1$. Here, $p$ denotes the frequency of non-disease-associated alleles and $q$ the aggregate frequency of disease-associated alleles within the relevant haemoglobinopathy category. Accordingly, $p^2$ represents individuals carrying no disease-associated allele, $2pq$ the carrier state, and $q^2$ the biallelic affected genotype state. For categories such as β-thalassaemia, $q$ therefore represents the aggregate frequency of β-thalassaemia-associated alleles, and $q^2$ can include both homozygous and compound-heterozygous affected genotypes.

Broad haemoglobinopathy terms, including thalassaemia, sickle cell disease, and haemoglobinopathy, were not used for Hardy-Weinberg derivation because they lacked sufficient genotype specificity. Derivations were applied only when the reported cause term could be linked unambiguously to a complementary carrier or biallelic affected state. Compound haemoglobinopathies were not used for Hardy-Weinberg derivation because no single complementary carrier state could be inferred. For α-thalassaemia and β-thalassaemia, general carrier-prevalence records were preferred; subtype-specific records were used only when they were the only available representation of the carrier state or when compatible denominators allowed valid aggregation.

For disease prevalence records, disease prevalence was converted from a percentage to a proportion, denoted as $D$. Allele frequency was estimated as $q = \sqrt{D}$, with $p = 1 - q$, and carrier prevalence was calculated as $2pq$. The derived carrier prevalence was stored as a percentage, and the derived carrier count was calculated as $2pq \times N$, rounded to the nearest integer, where $N$ was the reported denominator.

For carrier-prevalence records, carrier prevalence was converted from a percentage to a proportion, denoted as $C$. Allele frequency was estimated from $C = 2q(1 - q)$ as $q = (1 - \sqrt{(1 - 2C)}) / 2$, assuming $q \leq 0{\cdot}5$. Disease prevalence was then calculated as $q^2$. The derived disease prevalence was stored as a percentage, and the derived disease count was calculated as $q^2 \times N$, rounded to the nearest integer.

#### 1.9. Curated evidence overview

By the analytical freeze date, IthaMaps contained 31 509 curated records. Following curation, harmonisation, and derivation of complementary prevalence measures, IthaMaps records were summarised by publication year, country, content area (epidemiology, genetic diversity, and service availability), and continent. These summaries characterise the temporal and geographic coverage of the records and are shown for Oceania, Europe, Asia, the Americas, and Africa in Supplementary Figures S2–S6.

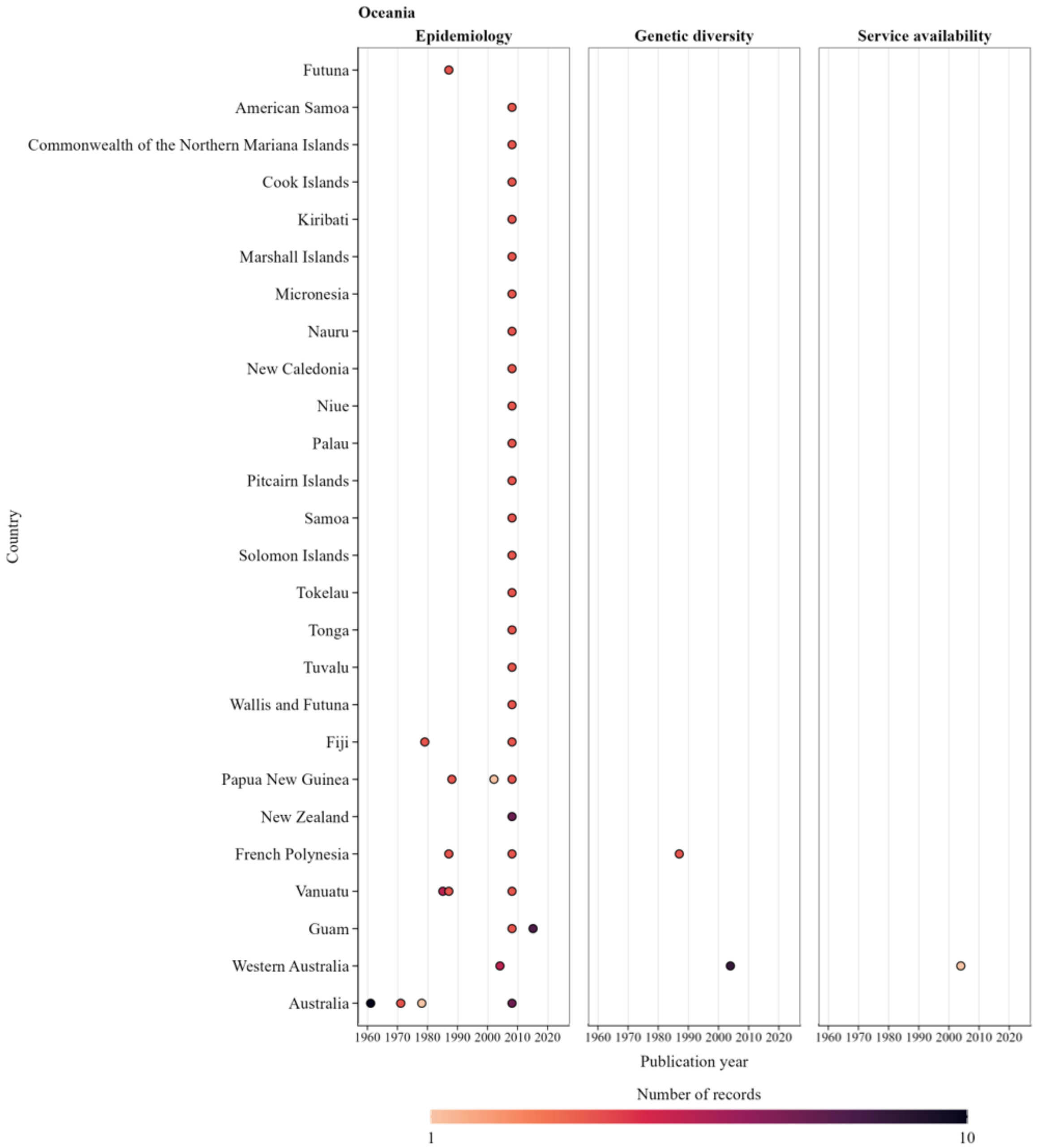

**Supplementary Figure S2. Publication-year distribution of curated IthaMaps records in Oceania, by content area.**

Each point represents records for a country and publication year within the indicated content area; colour indicates the number of records. Absence of a point indicates that no curated records were available for that country and publication year within the corresponding data domain.

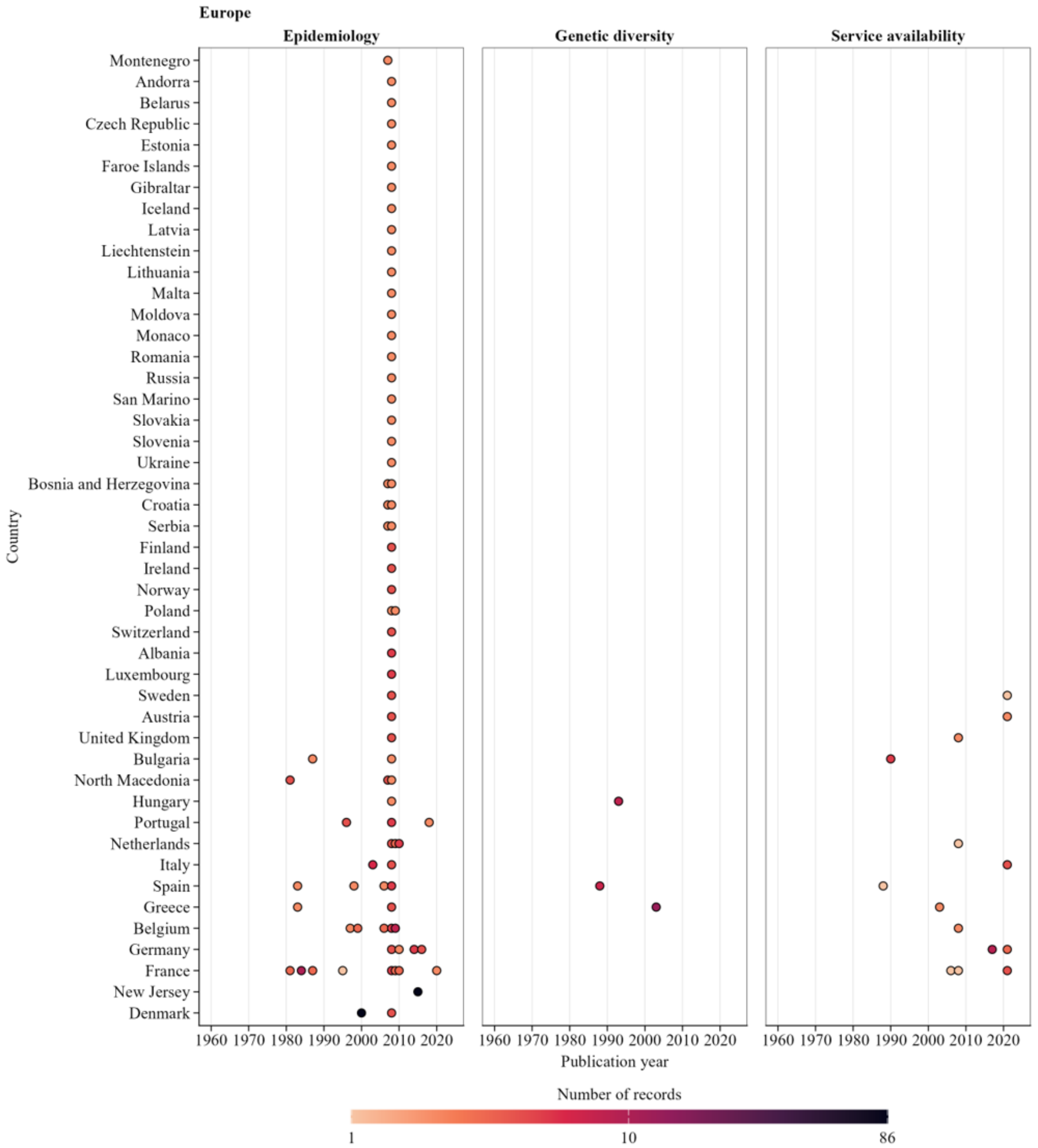

**Supplementary Figure S3. Publication-year distribution of curated IthaMaps records in Europe, by content area.**

Each point represents records for a country and publication year within the indicated content area; colour indicates the number of records. Absence of a point indicates that no curated records were available for that country and publication year within the corresponding data domain.

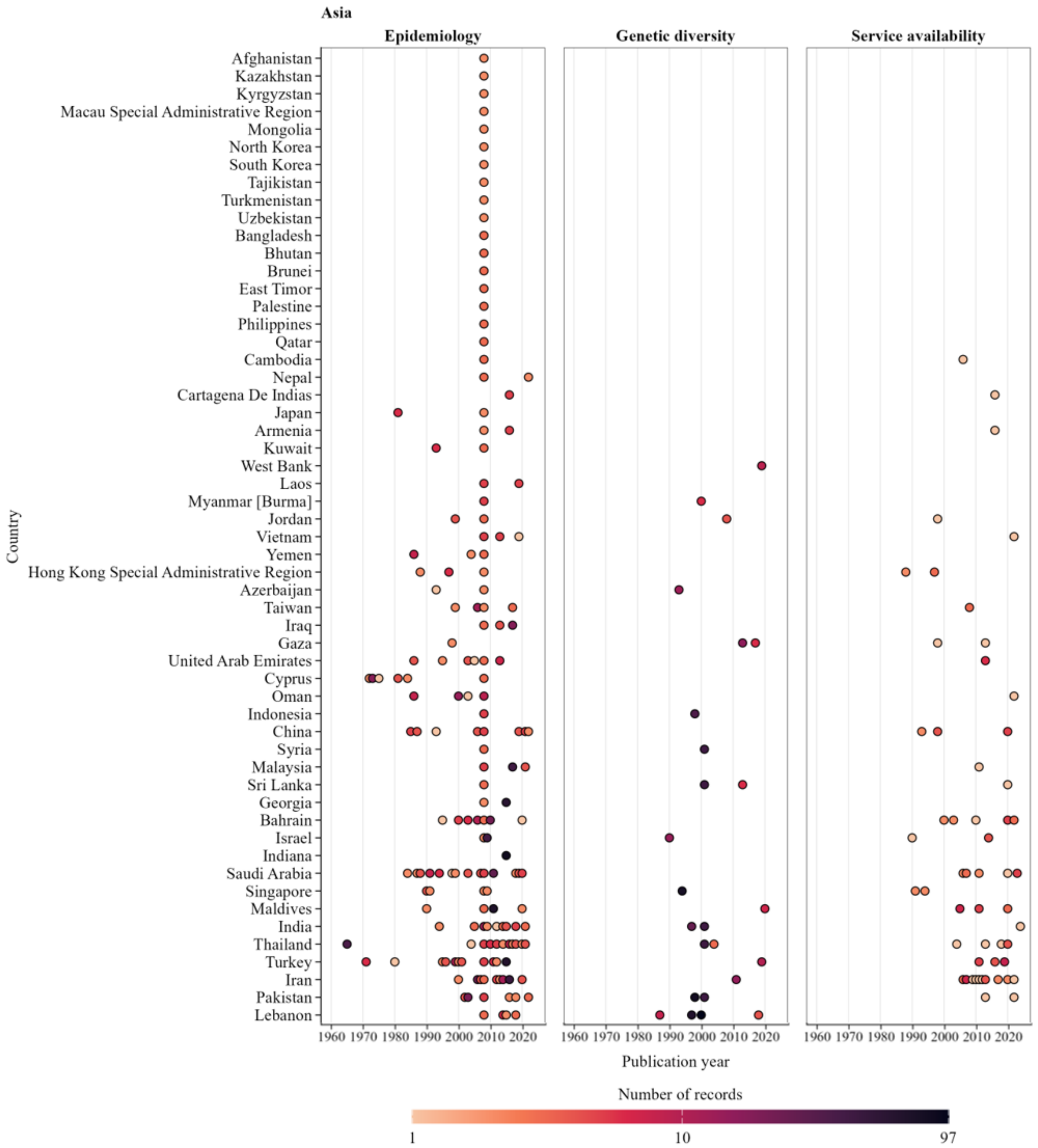

**Supplementary Figure S4. Publication-year distribution of curated IthaMaps records in Asia, by content area.**

Each point represents records for a country and publication year within the indicated content area; colour indicates the number of records. Absence of a point indicates that no curated records were available for that country and publication year within the corresponding data domain.

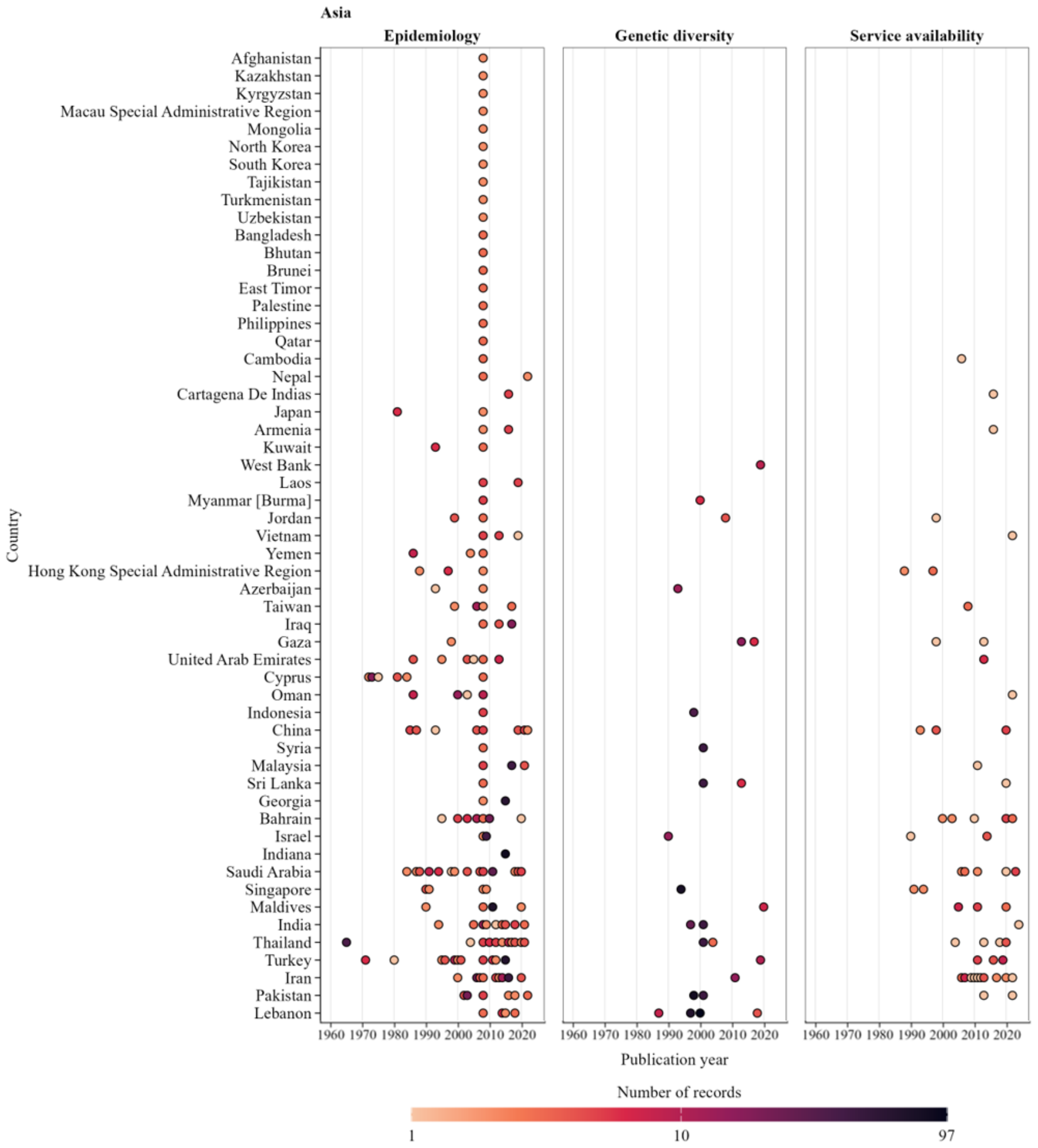

**Supplementary Figure S5. Publication-year distribution of curated IthaMaps records in the Americas, by content area.**
Each point represents records for a country and publication year within the indicated content area; colour indicates the number of records. Absence of a point indicates that no curated records were available for that country and publication year within the corresponding data domain.

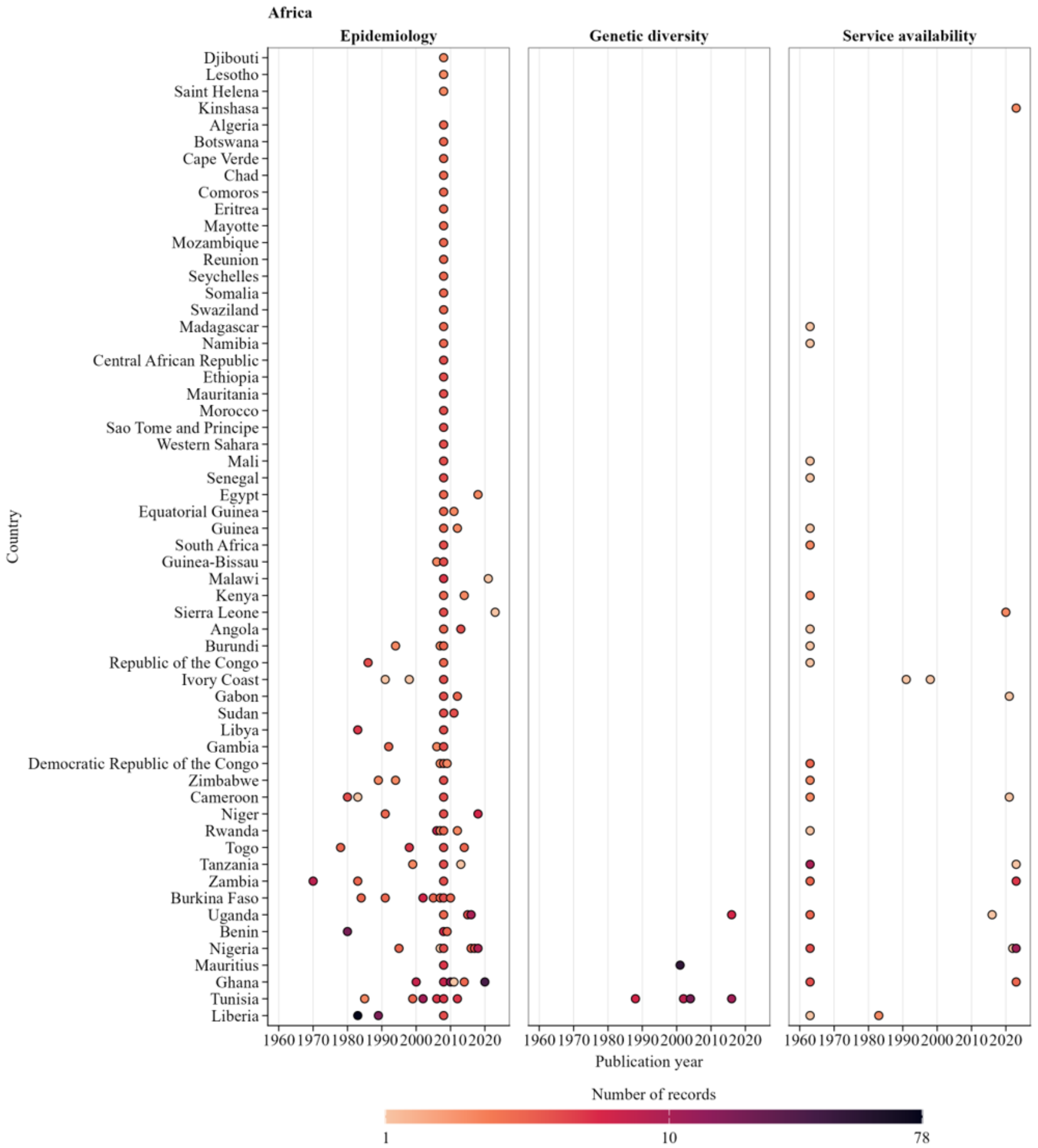

**Supplementary Figure S6. Publication-year distribution of curated IthaMaps records in Africa, by content area.**
Each point represents records for a country and publication year within the indicated content area; colour indicates the number of records. Absence of a point indicates that no curated records were available for that country and publication year within the corresponding data domain.

#### 1.10. Quality appraisal

Quality appraisal was restricted to manually curated IthaMaps records. Harmonised IHME curated records and records from compiled secondary sources, including ministry datasets and Modell's almanac, were excluded because the source information required for consistent assessment was unavailable.[8] Appraisal was restricted to epidemiological record types within the IthaMaps scope before redevelopment, including carrier prevalence, incidence, allele frequency, and relative allele frequency records.

Seven criteria adapted from the Joanna Briggs Institute prevalence critical appraisal tool were operationalised as record-level checks within the IthaMaps schema (Supplementary Table S7).[9] The retained criteria assessed sampling-frame appropriateness, reporting of recruitment and eligibility, sample-size adequacy, geographic and haemoglobinopathy specificity, diagnostic validity, and internal consistency of statistical reporting. Relative allele frequency records were additionally assessed for selective evaluation or reporting of structural and non-structural variants.

**Supplementary Table S7. Quality appraisal criteria adapted from the Joanna Briggs Institute prevalence tool for IthaMaps.[9]**

| | Appraisal criterion | Operational definition in IthaMaps | Applied to epidemiological records | Applied to relative allele frequency records | Applied to modelling framework records |
|---|---|---|---|---|---|
| 1 | Was the sample frame appropriate to address the target population? | Recruitment restricted to related individuals, known carriers, or affected individuals was treated as potentially biased. If such restriction was not reported, absence of this bias was assumed. | Yes | Yes | Yes |
| 2 | Were study participants sampled in an appropriate way? | Study rationale, recruitment setting, and inclusion and exclusion criteria were sufficiently described. | Yes | Yes | No |
| 3 | Was the sample size adequate? | The minimum denominator threshold was met; at least 76 participants for carrier prevalence, incidence or allele frequency records, or at least 19 tested alleles for relative allele frequency records. | Yes | Yes | Yes |
| 4 | Were the study subjects and the setting described in detail? | At least administrative level 0 location was reported, and the haemoglobinopathy outcome was sufficiently specified to permit georeferencing and disease mapping. | Yes | Yes | Yes |
| 5 | Was the data analysis conducted with sufficient coverage of the identified sample? | For relative allele frequency records, potential bias from selective allele evaluation was assessed. | No | Yes | No |
| 6 | Were valid methods used for the identification of the condition? | The diagnostic method used was appropriate for the haemoglobinopathy outcome evaluated. | Yes | Yes | Yes |
| 7 | Was the condition measured in a standard, reliable way for all participants? | The same diagnostic or analytical approach appeared to be applied consistently across participants. | No | No | No |
| 8 | Was there appropriate statistical analysis? | Numerators, denominators, reported values, and derived quantities were internally concordant. | Yes | Yes | Yes |
| 9 | Was the response rate adequate, and if not, was the low response rate managed appropriately? | Data were reported by demographic or geographic subgroup where relevant. | No | No | No |

Modelling framework records refer to the β-thalassaemia carrier-prevalence data utilised in the geostatistical modelling framework.

Sample-size adequacy for criterion 3 was implemented as a minimum denominator rule calibrated during a curation stage sensitivity analysis. Candidate thresholds were derived using the single proportion formula $n = Z^2P(1 - P) / d^2$, where n is the minimum denominator, P the expected proportion, d the tolerated absolute error, and Z the standard normal value for the selected confidence level.[9] The expected proportion was set at 0·052, based on Modell's estimate of global carriage of clinically significant haemoglobin variants, and (Z) was set at 1·96 for 95% confidence.[8] For carrier prevalence, incidence, and allele frequency, an absolute error margin of 5 percentage points produced a minimum denominator of 75·8, which was rounded upwards to 76 participants. For relative allele frequency, an absolute error margin of 10 percentage points produced a minimum denominator of 18·9, which was rounded upwards to 19 tested alleles. These thresholds were used as pragmatic record-screening criteria intended to remove records with extremely small denominators while limiting disproportionate loss of curated records.

Two adapted criteria were not retained in the final scoring. The criterion assessing whether the condition was measured in a standard and reliable way for all participants was excluded because its operational interpretation was ambiguous for haemoglobinopathy curation, since it could refer either to use of the same diagnostic test for all participants or to use of a consistent diagnostic protocol comprising multiple tests. Criterion 7 was therefore not retained in quality appraisal scoring. The criterion assessing response rate or stratified reporting was excluded because absence of stratification primarily reflected reporting granularity and did not consistently indicate bias for the curated haemoglobinopathy measures. Criterion 9 was therefore not retained in quality appraisal scoring.

Carrier prevalence, incidence, and allele frequency records were assessed using criteria 1, 2, 3, 4, 6, and 8. Relative allele frequency records were assessed using criteria 1, 2, 3, 4, 5, 6, and 8. Scores were expressed as percentages after excluding non-applicable criteria. Records satisfying over 70% of applicable criteria were classified as having low risk of bias, records satisfying less than 50% were classified as having high risk of bias, and all remaining records were classified as having moderate risk of bias. Records outside the appraisal framework or with insufficient information remained unappraised.

A separate qualification flag was assigned when all core requirements for sampling-frame appropriateness, sample size, geographic and haemoglobinopathy specificity, diagnostic validity, and statistical consistency were met. These requirements corresponded to criteria 1, 3, 4, 6, and 8 and defined the minimum evidential requirements for inclusion in the principal β-thalassaemia geostatistical analysis.

**1.11. Outlier detection**

Records were screened for potentially outlying values after stratification by country, haemoglobinopathy, epidemiological measure, metric, and administrative level. Additional stratification by variant identifier and globin phenotype was applied where relevant.

Screening was conducted separately at ADM0, ADM1, and ADM2. Records representing pooled estimates across more than one geographic location were excluded from the calculation of stratum-specific outlier thresholds to prevent repeated use of the same pooled estimate. Strata containing fewer than five eligible records were not screened.

Within each eligible stratum, records were evaluated using three complementary criteria; median absolute deviation (MAD), one standard deviation, and interquartile range criteria. For record $i$ with carrier prevalence $x_i$, the median absolute deviation (MAD) was defined using $\mathrm{MAD} = 1.4826\ \mathrm{median}_j|x_j - \tilde{x}|$, where $\tilde{x}$ denotes the median carrier prevalence among records in the corresponding stratum. The standardised MAD deviation was $Z_i^{\mathrm{MAD}} = \frac{x_i - \tilde{x}}{\mathrm{MAD}}$ *and* records with $|Z_i^{\mathrm{MAD}}| > 3.5$ were flagged. For the standard deviation criterion, records were flagged when $x_i < \bar{x} - s$ or $x_i > \bar{x} + s$, where $\bar{x}$ and $s$ are the mean and standard deviation of carrier prevalence among records in the corresponding stratum. For the interquartile range criterion, $IQR = Q_3 - Q_1$, and records were flagged when $x_i < Q_1 - 1.5\ IQR$ or $x_i > Q_3 + 1.5\ IQR$.

A record was classified as an outlier at a given administrative level only when all three criteria identified it as outlying. Flagged records were omitted from IthaMaps visualisations for the corresponding administrative level and stratum. Records were excluded from geostatistical modelling only when flagged consistently across all administrative levels available for assessment. This conservative procedure was intended to exclude records with values consistently discordant with comparable records while retaining those with plausible extreme values.

Application of this procedure to β-thalassaemia carrier-prevalence records identified six records as outliers across the relevant administrative level assessments; their exclusion left 1954 candidate records for subsequent modelling dataset construction. Among these records, 212 contained carrier prevalence derived under Hardy-Weinberg equilibrium and 346 records with pooled estimates across multiple recruitment locations. These characteristics were retained at this stage and evaluated explicitly in the sensitivity analyses rather than treated as additional exclusion criteria *a priori*.

**1.12. Summary metric computations**

Descriptive summaries were calculated after outlier screening to characterise the distribution of curated epidemiological records within IthaMaps. Records were stratified by country, haemoglobinopathy, epidemiological measure, metric, and administrative level, with additional stratification by variant identifier and globin phenotype where applicable.

For each stratum, calculated summaries comprised the latest reported estimate, the estimate with the largest denominator, minimum, maximum, median, arithmetic mean, and random-effects weighted mean.[10] When a stratum contained only one record, the reported estimate was retained for all summary measures.

For random-effects pooling, estimates were expressed as numerator-denominator proportions and transformed using the Freeman-Tukey double arcsine transformation.[11] Sampling variances were calculated from the corresponding numerators and denominators, allowing records with larger denominators and lower sampling variance to receive greater weight. An intercept-only random-effects model was fitted by restricted maximum likelihood to estimate a stratum-specific pooled estimate while allowing for between-record heterogeneity.[12] Pooled estimates were back-transformed to the original scale and expressed in the corresponding metric for reporting and visualisation.

#### 1.13. Integration into the ITHANET portal

Curated records were standardised to the ITHANET database schema and uploaded through structured query language operations. Upload completeness was assessed by querying the database after import and confirming permanent identifiers for all uploaded records.

An R Shiny application was developed as the interactive IthaMaps interface within ITHANET and is available at https://www.ithanet.eu/db/ithamaps.[13] The application queried records according to user-selected filters, including geography, haemoglobinopathy, epidemiological measure, summary metric, healthcare service category, and administrative level.

Epidemiological records were displayed as recruitment-site markers and administrative unit choropleths. Linked tables retained record-level estimates, denominators, appraisal classifications, and source provenance. Healthcare service availability records were reconciled within combinations of country, haemoglobinopathy, service type, and service subtype. When records conflicted, evidence of availability was prioritised over evidence of absence, and evidence of national availability was prioritised over evidence of regional availability. Reconciled classifications were used for country-level maps, whereas linked tables retained the underlying records.

Filtered outputs were made available in CSV, GeoJSON, GeoPackage, GeoTIFF, and PNG formats.

### 2. Bayesian geostatistical framework

#### 2.1. Covariate data

Eight candidate covariates were selected a priori to represent plausible historical, demographic, population-structure, and health system correlates of spatial variation in β-thalassaemia carrier prevalence. Candidate covariates comprised *Plasmodium falciparum* incidence, consanguinity, ethnic fractionalisation, religious diversity, population count, degree of urbanisation, haemoglobinopathy genetic counselling availability, and haemoglobinopathy prevention service availability. Covariate datasets, reference periods, and native spatial resolutions are reported in Supplementary Table S8.

**Supplementary Table S8. Geospatial covariates assembled for evaluation in the modelling framework.**

| Covariate | Data version | Native resolution | Sources |
|---|---|---|---|
| Consanguinity (%) | Most recent per country | Administrative level 0 and administrative level 1 where available | Consang.net (https://consang.net/) |
| Degree of urbanization | 2026 | 1 x 1 km | WorldPop (https://www.worldpop.org/) |
| Ethnic fractionalization index | Most recent per country | Administrative level 0 and 5 x 5 km where available | DHS (https://dhsprogram.com/); Cadmus - European University Institute Research Repository (https://hdl.handle.net/1814/64606) |
| Genetic counselling availability for haemoglobinopathies | Most recent per country | Administrative level 0 and administrative level 1 or 2 where available | ITHANET (https://www.ithanet.eu) |
| Religious diversity index | Most recent per country | Administrative level 0 and 5 x 5 km where available | DHS (https://dhsprogram.com/); Pew Research Center (http://pewresearch.org/) |
| *Plasmodium falciparum* incidence rate (per 1,000 population) | 2020 | 5 x 5 km | Malaria Atlas Project (https://malariaatlas.org/) |
| Population counts | 2026 | 1 x 1 km | WorldPop (https://www.worldpop.org/) |
| Prevention service availability for haemoglobinopathies | Most recent per country | Administrative level 0 and administrative level 1 or 2 where available | ITHANET (https://www.ithanet.eu) |

DHS: Demographic and Health Survey

*Plasmodium falciparum* incidence was included as a proxy for historical malaria exposure because malaria endemicity has been associated with increased frequencies of haemoglobinopathy-causing variants (Supplementary Figure S7).[14,15]

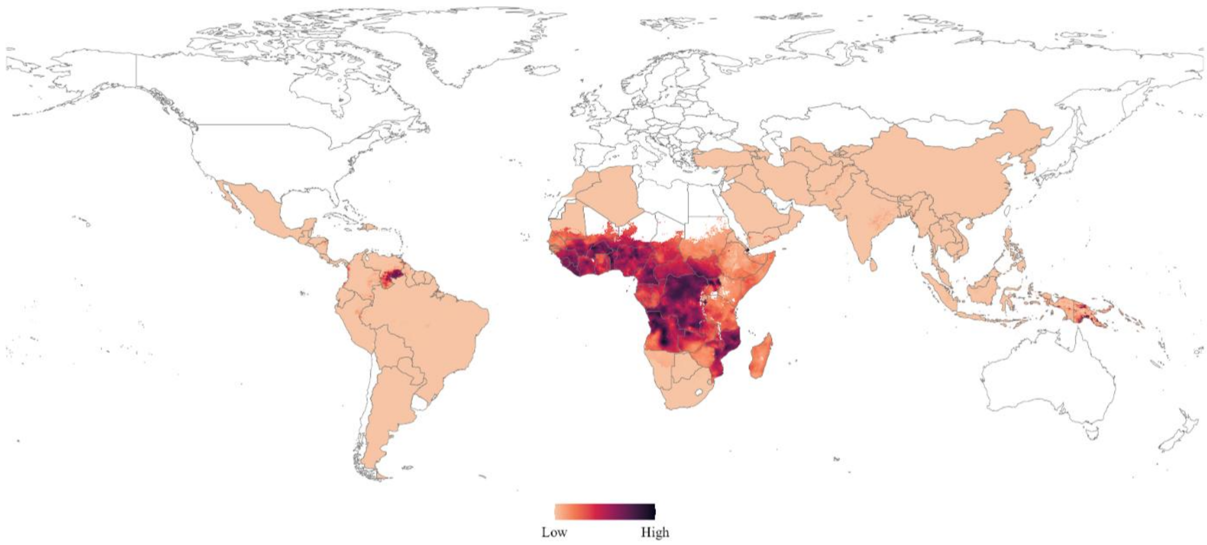

**Supplementary Figure S7: *Plasmodium falciparum* incidence covariate map.**
The *Plasmodium falciparum* incidence covariate was assembled from the 2020 Malaria Atlas Project incidence rate surface at 5 km × 5 km native resolution and resampled to the common raster grid.[16] Values are expressed per 1000 population. Darker shading indicates higher incidence. Areas shown in white indicate that no covariate value was available.

Consanguinity was included as an indicator of mating between biologically related individuals and, more broadly, endogamy and restricted mating networks, which can contribute to the local persistence and familial clustering of haemoglobinopathy-causing variants (Supplementary Figure S8).[17,18]

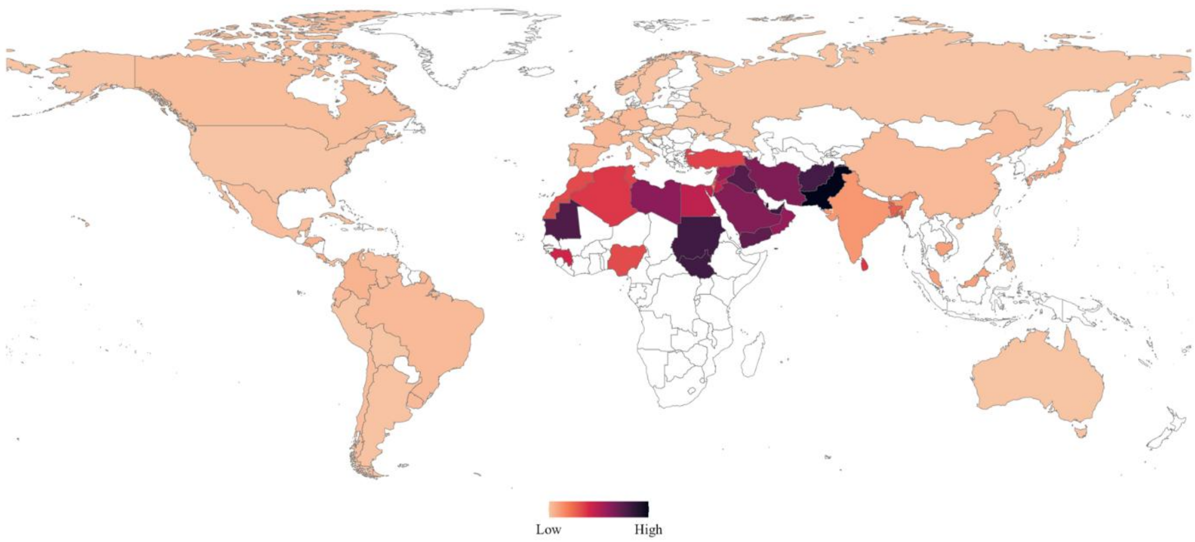

**Supplementary Figure S8: Consanguinity covariate map.**
The consanguinity covariate was assembled from the most recent available country-specific estimates from Consang.net and rasterised to the common raster grid using administrative boundaries.[18] Darker shading indicates higher reported consanguinity. Areas shown in white indicate that no covariate value was available.

Ethnic fractionalisation and religious diversity were included as measures of population heterogeneity and social structure. These variables were considered potential proxies for spatial differences in ancestry, migration history, founder effects, gene flow, reproductive practices, and engagement with healthcare services (Supplementary Figures S9–S10).[19,20]

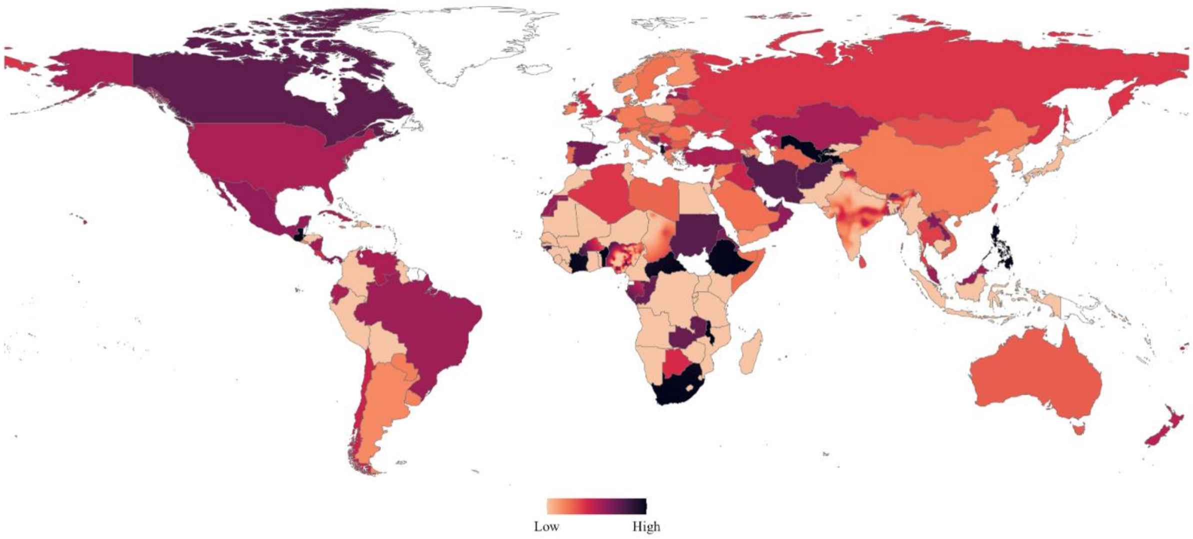

**Supplementary Figure S9: Ethnic fractionalisation covariate map.**
The ethnic fractionalisation covariate combined country-level estimates with Demographic and Health Surveys cluster-level data where available.[21–23] Country-level estimates were assigned across each country, and in countries with cluster-level data, these were replaced by values interpolated from the survey clusters at approximately 5 km × 5 km resolution. Darker shading indicates greater ethnic fractionalisation. Areas shown in white indicate that no covariate value was available.

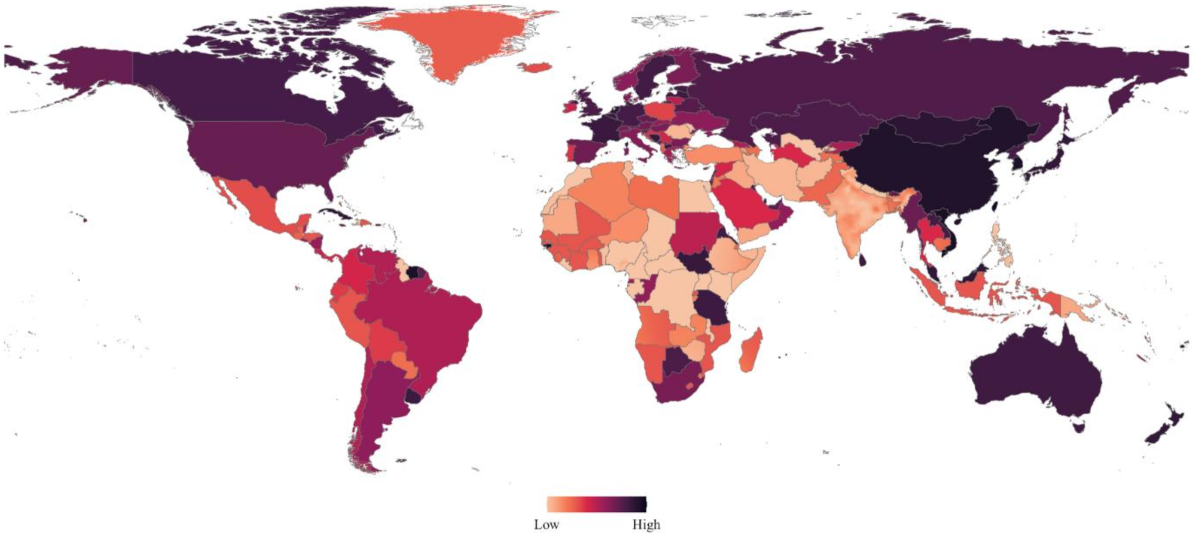

**Supplementary Figure S10: Religious diversity covariate map.**
The religious diversity covariate combined country-level estimates with Demographic and Health Surveys cluster-level data where available.[22–24] Country-level estimates were assigned across each country, and in countries with cluster-level data, these were replaced by values interpolated from the survey clusters at approximately 5 km × 5 km resolution. Darker shading indicates greater religious diversity. Areas shown in white indicate that no covariate value was available.

Population count was included to represent differences in local population concentration. Founder effects, genetic drift, and endogamy might exert greater effects in sparsely populated settings, whereas population mobility and mixing might be greater in densely populated settings (Supplementary Figure S11). [25]

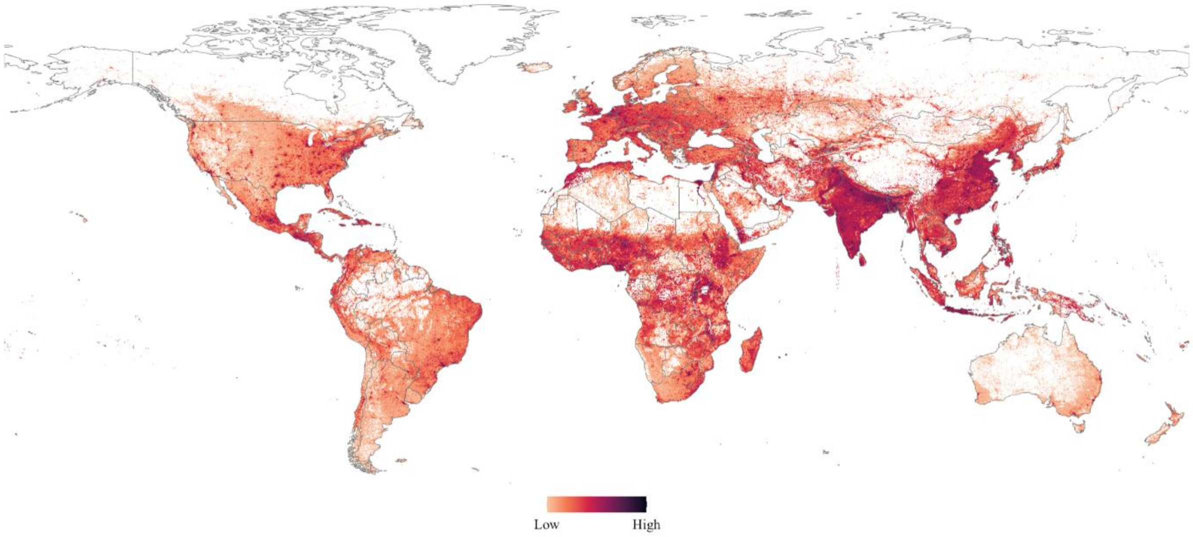

**Supplementary Figure S11: Population count covariate map.**
The population count covariate was assembled from the 2026 WorldPop global population surface at 1 km × 1 km native resolution and resampled to the common raster grid.[26] Darker shading indicates higher population count. Areas shown in white indicate that no covariate value was available.

Degree of urbanisation was included to capture rural-urban differences in population mixing and access to haemoglobinopathy services (Supplementary Figure S12). Urban populations might be more heterogeneous because of internal and international migration and might have greater access to prevention and management services, potentially influencing the ascertainment of carrier prevalence.[27,28]

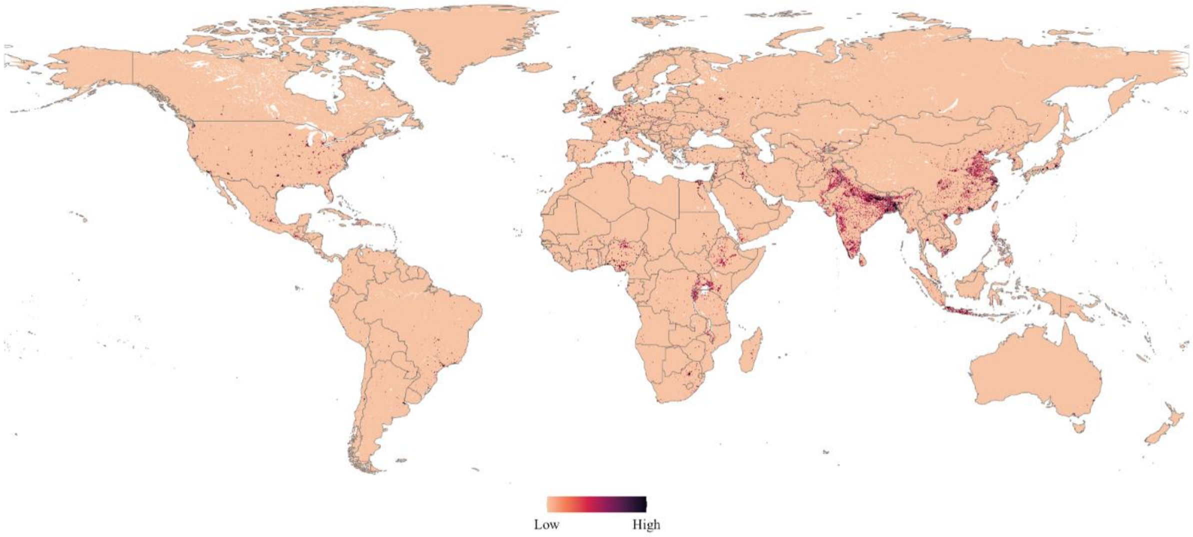

**Supplementary Figure S12: Degree of urbanisation covariate map.**
The degree of urbanisation covariate was assembled by mosaicking country-specific 2026 WorldPop gridded surfaces at 1 km × 1 km native resolution, then projecting and resampling the surface to the common raster grid.[29] Darker shading indicates higher urbanisation. Areas shown in white indicate that no covariate value was available.

Genetic counselling and prevention service availability were included as indicators of settings in which haemoglobinopathies had been recognised as a health-service priority and organised screening or counselling had been implemented (Supplementary Figures S13–S14).[30,31] These associations were regarded as potentially bidirectional and time-lagged. Service availability might reflect a response to recognised disease burden, increase carrier ascertainment, or influence reproductive behaviour when sustained over time.[32,33]

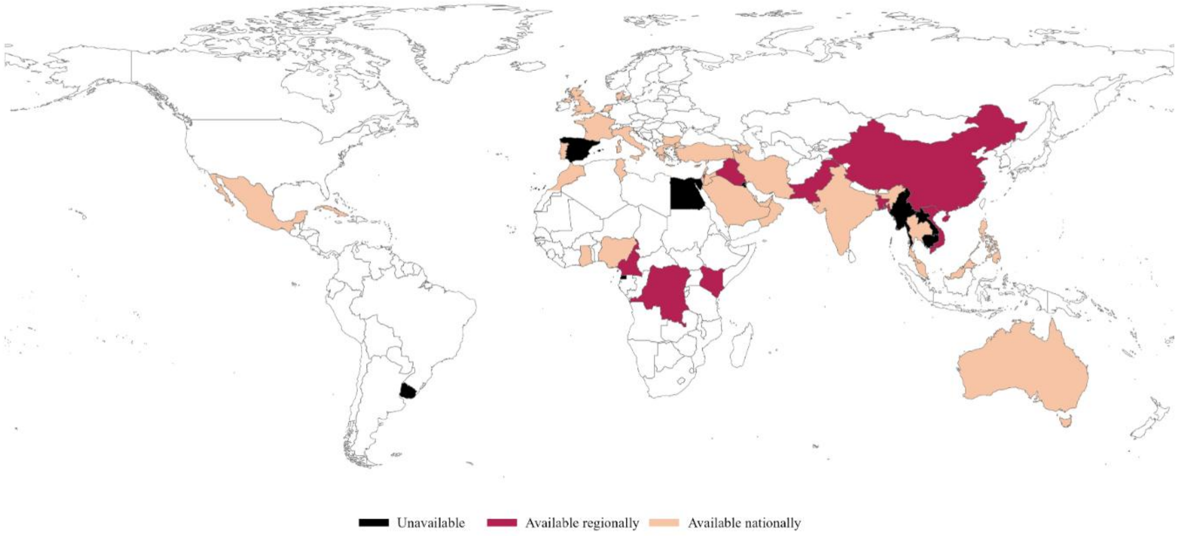

**Supplementary Figure S13: Genetic counselling availability covariate map.**
The genetic counselling availability covariate was derived from curated IthaMaps haemoglobinopathy service records.[2] Country-level availability was rasterised and updated with higher resolution service availability surfaces where available. Colours indicate whether genetic counselling was unavailable, available regionally, or available nationally. Areas shown in white indicate that no curated service record was available.

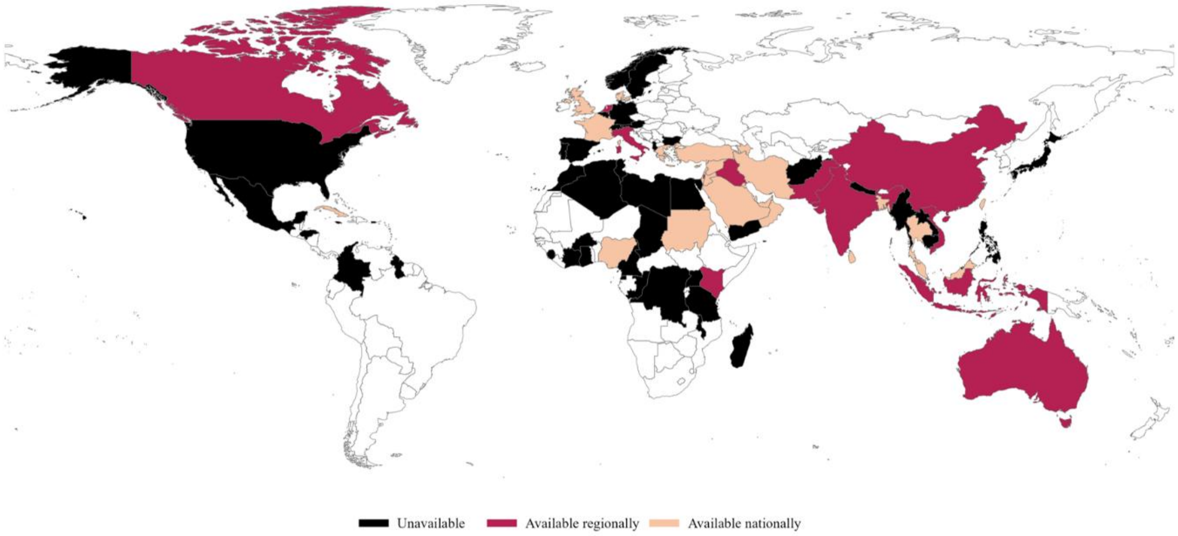

**Supplementary Figure S14: Prevention service availability covariate map.**
The prevention service availability covariate was derived from curated IthaMaps haemoglobinopathy service records.[2] Country-level availability was rasterised and updated with higher resolution service availability surfaces where available. Colours indicate whether prevention services were unavailable, available regionally, or available nationally. Areas shown in white indicate that no curated service record was available.

#### 2.2. Covariate data preparation

All covariates were aligned to a common raster grid with an approximate resolution of 5 km × 5 km. Continuous raster surfaces were resampled by bilinear interpolation, whereas categorical raster surfaces were resampled by nearest-neighbour interpolation to preserve category membership. Values reported for administrative units were linked to the corresponding boundaries and assigned to all grid cells within those units. When the same covariate was available at multiple administrative resolutions, the coarser surface was used as the baseline and replaced by finer-resolution values where available. All covariates were checked for consistency in geographic extent, coordinate reference system, resolution, and cell alignment.

Prevention service availability was derived from records of neonatal, cascade, prenatal or antenatal, premarital, and school-based screening. Records specific to sickle cell disease were excluded from the prevention service and genetic counselling availability covariates used in the β-thalassaemia analysis. Availability was coded as 1 for national coverage, 0·5 for subnational (regional) coverage, and 0 for explicitly reported unavailability. Cells with insufficient information were retained as missing rather than assumed to lack service provision.

Georeferenced Demographic and Health Survey data were used to produce spatially resolved measures of ethnic fractionalisation and religious diversity.[22,23] When survey data were available for women and men, responses were pooled and linked to the published coordinates of the corresponding survey clusters. Within each country, ethnicity labels were harmonised by correcting spelling variants and consolidating equivalent group names.

Religious affiliation was mapped to eight categories comprising Christianity, Islam, Hinduism, Buddhism, Judaism, folk or traditional religions, other religions, and religiously unaffiliated, from which the religious diversity index was calculated using a previously published approach.[24] The ethnic fractionalisation index was calculated from the proportions of respondents belonging to each reported ethnic group following a previously published approach.[21] Both indices equalled zero when all respondents belonged to one reported group and increased as respondents were distributed across more groups. Country-specific kriging was used to interpolate values for each index between survey clusters and generate continuous surfaces at an approximately 5 km x 5 km spatial resolution, following a previously published approach.[34]

#### 2.3. Covariate selection

Covariate selection was undertaken within a non-spatial frequentist framework before fitting the Bayesian geostatistical model.[35] For β-thalassaemia carrier-prevalence records georeferenced to recruitment sites, covariate values were extracted at the corresponding record coordinates. For records georeferenced only to an administrative unit, the mean covariate value across the corresponding unit was assigned. Analyses were restricted to records with complete data for all eight candidate covariates.

Covariate distributions and their relationships with the empirical logit of β-thalassaemia carrier prevalence were examined graphically and highly skewed covariates were log-transformed.[35] Potential covariate interactions were examined using pairwise binomial logistic generalised linear models, and interaction terms with $p<0·05$ were retained for subsequent multivariable assessment. A univariable binomial generalised linear model was fitted for each candidate covariate, with the number of carriers and non-carriers specified as the binomial outcome.[35] Five random subsamples, each containing 80% of the records with complete data for all eight candidate covariates, were drawn. Models were fitted and evaluated within the same subsample, and covariates were ranked by the mean of the squared Pearson correlations between fitted and observed carrier proportions across the five subsamples. Pairwise Pearson correlation coefficients were subsequently calculated to identify covariates that could introduce multicollinearity in the multivariable model.[35] For covariate pairs with an absolute correlation coefficient of 0·80 or greater, the lower-ranked covariate was excluded. Variance inflation factors (VIFs) were then calculated for the remaining covariates, which were removed iteratively in descending order of VIF until all values were 5 or lower.[36]

Covariates retained after multicollinearity screening were entered into a multivariable binomial generalised linear model.[35] Backward elimination based on the Akaike information criterion was applied, and covariates with $p\leq0·05$ were carried forward to the Bayesian geostatistical analysis.[37] Interaction terms were retained only when both corresponding main effects remained in the model.

#### 2.4. Spatial disaggregation

To provide a finer spatial representation of coarse area-level records, records georeferenced to ADM0 or ADM1 centroids were assigned to more granular administrative proxy locations using the 2026 WorldPop population count covariate surface[26] (Supplementary Table S8) and an adaptation of a previously described population-informed method.[38,39] Population weighting and k-means clustering were used to identify representative population concentrations within inhabited areas. Because administrative centroids denote the geographic unit associated with an estimate rather than the location of the sampled population, the resulting coordinates were treated as analytical proxies, not as inferred recruitment locations. This approach was used because the underlying sources did not provide sufficient information to identify the true within-unit recruitment locations, precluding direct point-level georeferencing.

Two separate, non-sequential disaggregation pathways were applied. Eligible records originally georeferenced to ADM0 were reassigned to ADM1, whereas records originally georeferenced to ADM1 were reassigned to ADM2. ADM1 records generated through the first pathway were not subsequently processed through the second.

ADM0 records were eligible for reassignment only for countries or territories smaller than 20 000 km², for which localisation to a single ADM1 unit was considered a reasonable approximation of the broader area represented by the estimate. Within each eligible ADM0 unit, populated grid cells were partitioned into a maximum of three clusters. Population counts were summed within each k-means cluster, and the cluster with the largest total population was selected. The mean location of the cells in that cluster was used to identify the corresponding ADM1 unit, after which the record was assigned to the point-on-surface coordinate of that unit. Each eligible ADM0 record was therefore represented by one ADM1-level analytical proxy.

For each record originally georeferenced to ADM1, let $N_i$ denote the number of populated grid cells within the corresponding unit. The number of cells sampled without replacement was $m_i = \min\{10,\ 000\lfloor 0.80 N_i \rfloor\}$, with selection probabilities proportional to population. The number of k-means clusters was determined as $k_i = \max\left\{1, \left\lfloor \frac{m_i}{\min(3000, m_i)} \right\rfloor\right\}$. Given the maximum sample of 10 000 cells, this procedure generated one to three clusters. Each cluster centre was used to identify the corresponding ADM2 unit, after which the record was assigned to the point-on-surface coordinate of that unit. Each original ADM1 record could therefore be represented by one to three ADM2-level analytical proxies.

Each proxy retained the parent record's carrier prevalence, numerator, and denominator unchanged. Records originally georeferenced to ADM2 or to a more precise recruitment location were retained at their original spatial resolution. The effect of this procedure on the records retained for final model development is reported in Supplementary section 2.11.

**2.5. Record selection at coincident coordinates**

Records taken forward for geostatistical modelling were restricted to those reporting β-thalassaemia carrier prevalence, with values reported as rates converted to percentages. Longitude and latitude were rounded to four decimal places, and records sharing the same rounded coordinate pair were considered spatially coincident. This retained coordinate precision at approximately 11 m in latitude and no more than approximately 11 m in longitude. To prevent multiple records at the same modelling coordinate from contributing independently to the model, one record was retained at each coordinate pair according to a prespecified sequential hierarchy.

Where records at original coordinates coincided with analytical proxy coordinates, records at the original coordinates were retained. At successive stages of the hierarchy, records from a single recruitment location were prioritised over records pooled across multiple locations; directly reported carrier-prevalence records were prioritised over those derived under Hardy-Weinberg equilibrium; and records assessed as having a low risk of bias were prioritised over records with moderate or high risk of bias or records that were unappraised.

The hierarchy then considered population scope. At coordinates containing both broad or mixed-population records and ethnicity-specific records, the summed denominator across ethnicity-specific records was compared with the summed denominator across broad or mixed-population records. Ethnicity-specific records remained eligible when the former was at least twice the latter.

Remaining overlaps were resolved by sample size and recency. When the largest sample size was at least twice that of the second largest distinct sample, the record with the largest sample was retained; otherwise, the most recent record was selected. Residual ties were resolved by retaining the record with the largest sample. Because the records entering this hierarchy differed among sensitivity and model development datasets, dataset-specific reductions were determined after the corresponding inclusion and spatial-processing decisions had been applied and are reported for the final model development sequence in Supplementary section 2.11.

**2.6. Exclusion of spatially isolated records**

Spatial isolation was assessed using nearest-neighbour distance in longitude-latitude coordinate space. For each record, Euclidean distances to all other records were calculated from longitude and latitude in decimal degrees, and the minimum was taken as the nearest-neighbour distance. A threshold of 25 coordinate degrees was selected pragmatically after visual assessment of the spatial distribution of records, with the specific aim of identifying records that were clearly separated from the broader spatial distribution of carrier-prevalence records. Records with a nearest-neighbour distance exceeding this threshold were excluded before geostatistical modelling to limit the influence of extremely isolated records on estimation of the spatial field.[40] Threshold selection was based solely on the spatial configuration of the records and did not incorporate carrier prevalence values or model performance measures. Records from countries separated from the broader spatial distribution of carrier-prevalence records by large unsampled areas were additionally excluded to avoid imposing spatial continuity across those intervening areas.[41]

These geographic exclusions were reapplied whenever record restoration or other upstream processing changed the composition or spatial configuration of a modelling dataset. Their realised effect on the final modelling dataset is reported in Supplementary section 2.11.

#### 2.7. Bayesian binomial geostatistical model

For record $i$, represented at modelling coordinate $s_i$, let $Y_i$ denote the observed number of β-thalassaemia carriers among $n_i$ individuals tested. Carrier counts were modelled directly as $Y_i \mid p_i \sim Binomial(n_i, p_i)$, where $p_i$ is the latent carrier probability represented by the record. Modelling the numerator and denominator directly retained the binomial sampling information, such that records with larger denominators contributed greater information about carrier prevalence without requiring transformation of the observed proportion or estimation of its sampling variance.[42]

Candidate linear predictors took the general form $logit(p_i) = \beta_0 + x_i^T\beta + S(s_i) + u_i$, where $\beta_0$ is the intercept; $x_i$ is the vector of selected covariate values associated with record $i$; and $\beta$ is the corresponding vector of regression coefficients. Thus, $x_i^T\beta$ is the scalar sum of each included covariate multiplied by its estimated coefficient. $S(s_i)$ is a spatially structured random-effect at coordinate; $s_i$ and $u_i$ is an independent and identically distributed (IID) location-level random-effect. The two random components represented distinct residual processes. $S(s_i)$ allowed unexplained carrier prevalence at geographically proximate locations to be correlated and therefore represented residual geographic structure not captured by the included fixed-effects. In contrast, $u_i$ allowed additional local variation without imposing greater similarity between nearby than distant coordinates.[43]

The IID effect was specified as $u_i \mid \sigma_u \sim N(0, \sigma_u^2)$, meaning that, conditional on the marginal standard deviation $\sigma_u$, the effect $u_i$ at modelling coordinate $i$ was assumed to follow a normal distribution with mean 0 and variance $\sigma_u^2$. The zero mean centres this component on the carrier prevalence predicted by the remaining model terms, whereas $\sigma_u$, expressed on the logit scale, determines the magnitude of residual variation not represented by the spatial field or fixed-effects. Conditional on $\sigma_u$, the $u_i$ were identically distributed and mutually independent, so their similarity was not assumed to depend on geographic distance.

Because one record was retained at each modelling coordinate, location-level heterogeneity could not be distinguished from unmeasured record-specific differences in the sampled population, recruitment, diagnostic practice, ascertainment, or other record characteristics. The IID term therefore represented residual location-level heterogeneity in carrier prevalence not explained by the covariates or spatial field but could absorb both genuine local heterogeneity and these unmeasured record-specific differences. It was not specified as a study-level random-effect and should not be interpreted as isolating between-study heterogeneity. Conditional on the latent effects and model parameters, the observed carrier counts were assumed independent.

The intercept was retained in all formulations. Fixed, spatial, and IID components were included in prespecified combinations so that their contribution to model fit and held-out prediction could be evaluated rather than assumed.

#### 2.8. Spatial field, mesh, priors, and posterior inference

The spatial effect $S(s_i)$ was represented as a stationary, zero-mean Matérn Gaussian random field. The field was characterised by its marginal standard deviation, $\sigma_s$, which quantifies the magnitude of spatially structured residual variation on the logit scale, and its practical range, $\rho$, which describes the distance over which appreciable spatial correlation persists.[44]

The continuous Matérn field was approximated using the stochastic partial differential equation approach.[44] This representation expresses the Gaussian field as a Gaussian Markov random field on a triangular mesh, producing a sparse precision matrix and permitting computationally efficient inference across multiple countries. Values of the spatial field at modelling and prediction coordinates were obtained by projection from the surrounding mesh vertices.

Longitude and latitude were transformed to three-dimensional Cartesian coordinates on the unit sphere before mesh construction and spatial projection. The resulting triangular mesh was defined on the $S^2$ spherical manifold. This representation accounted for Earth curvature across the multicounty domain rather than treating longitude and latitude as planar Cartesian coordinates. Mesh dimensions and spatial range parameters specified in kilometres were converted to the unit-sphere scale by dividing by an Earth radius of 6371 km.[45]

For each dataset, the mesh boundary was derived from the union of ADM0 units represented in that dataset. A cutoff of approximately 50 km was used to avoid excessive mesh refinement around closely spaced input locations. Maximum triangle edge lengths were approximately 100 km in the interior mesh domain and 1000 km in the outer mesh extension. An outer mesh extension of approximately 1000 km was specified beyond the supplied boundary to reduce boundary effects while limiting unnecessary mesh density outside the area of interest. The stochastic partial differential equation parameter was fixed at $\alpha = 2$. On the two-dimensional $S^2$ manifold, this corresponds to Matérn smoothness $\nu = \alpha - \frac{d}{2} = 1$, where $d = 2$ is the spatial dimension.

Penalised complexity priors were assigned to the practical spatial range and to the marginal standard deviations of the spatial and IID effects.[46] For each dataset, the reference range $r_0$ was defined as 5% of the great-circle distance across the diagonal of the bounding extent of the represented ADM0 units, with $P(\rho < r_0) = 0.01$. Thus, 1% prior probability was assigned to a spatial range shorter than $r_0$, regularising against highly localised spatial structure unless supported by the data. For the spatial field, $P(\sigma_s > 3) = 0.01$; the IID effect was assigned the corresponding prior $P(\sigma_u > 3) = 0.01$. Both standard deviations were defined on the logit scale. The IID prior was implemented through the equivalent penalised complexity prior on its precision, $\tau_u = \frac{1}{\sigma_u^2}$. No user-specified priors were assigned to the intercept or covariate coefficients.

Models were fitted by integrated nested Laplace approximation (INLA) with automatic numerical integration over the hyperparameters.[47] The INLA configuration required to obtain joint posterior draws was retained. For posterior prediction, 1000 draws were obtained from the approximated joint posterior distribution rather than independently from separate marginal distributions[48]. Each draw therefore contained a mutually consistent realisation of the latent model components and hyperparameters, including the intercept, any included fixed-effects, spatial field, and variance or precision parameters. Preserving this joint dependence allowed uncertainty in these components to propagate jointly into predicted carrier prevalence.

For models containing an IID component, each posterior draw also provided a draw of its precision, $\tau_u$. At prediction locations not represented among the modelling coordinates of the fitted model, a new IID effect was generated conditional on the corresponding posterior draw of the IID precision. Fitted IID effects at modelling coordinates were not transferred to these prediction locations.

#### 2.9. Model evaluation and random five-fold cross-validation

Model evaluation combined the Watanabe-Akaike information criterion (WAIC) and conditional predictive ordinate (CPO) with random five-fold cross-validation. WAIC and CPO provided measures of model fit and single-record leave-one-out predictive support, respectively, whereas random five-fold cross-validation assessed predictive performance with one of five folds withheld at each iteration.[49,50] Random cross-validation was designed to assess interpolation within the geographic support of the observed data; spatial cross-validation, described in Supplementary section 2.12, separately assessed prediction across geographic gaps.

WAIC was calculated from each full-data fit, with lower values indicating a more favourable balance between predictive fit and model complexity.[49] Direct comparisons were restricted to model formulations fitted to the same records. When record inclusion differed between sensitivity datasets, WAIC was reported descriptively but was not used to rank datasets. For record $i$, CPO was defined as $\text{CPO}_i = p(y_i \mid y_{-i})$, where $y_i$ is the observed carrier count and $y_{-i}$ denotes the carrier count data from all other records.[50] CPO values were transformed to the negative log scale and summarised as $-\frac{1}{n}\sum_{i=1}^{n} \log(\text{CPO}_i)$, where lower values indicate greater average single-record leave-one-out predictive support.

Density-Based Spatial Clustering of Applications with Noise (DBSCAN) was used to characterise the geographic configuration of each dataset and to structure random cross-validation.[51] Density-based clustering was performed on longitude and latitude in decimal degree coordinate space. For each record, the distance to its second nearest neighbour was calculated. The initial DBSCAN neighbourhood radius, $\varepsilon$, was the 95th percentile of this distance distribution and was constrained between 2% of the maximum coordinate extent and the 98th percentile of second nearest-neighbour distances to avoid an exceptionally small or large radius. The minimum number of records required to form a cluster was $\text{minPts} = \max\{\text{round},[0.75\log(n)]4\}$, where $n$ is the number of records in the corresponding dataset. Records not satisfying the local-density criterion were classified as spatial noise.

Random five-fold cross-validation was designed principally to assess interpolation within the geographic support of the observed data.[52] Within each density cluster containing at least ten records, records were distributed approximately evenly and at random among five folds. Records in smaller density clusters and those classified as spatial noise remained in every training set and were not assigned to validation folds. Each validation fold was withheld in turn, and the model was refitted to the remaining records before prediction at the withheld locations. This design retained surrounding observational support for most withheld records.

Held-out predictive performance was quantified using mean absolute error (MAE), root mean square error (RMSE), and Pearson correlation between observed and posterior mean predicted carrier prevalence.[53] For held-out record $i$, observed carrier prevalence was $p_i = \frac{y_i}{n_i}$, where $y_i$ is the observed carrier count and $n_i$ is the number of individuals tested, and $\hat{p}_i$ denoted the posterior mean predicted carrier prevalence obtained from the model fitted without the validation fold containing that record. For a validation fold containing $m$ records, $\text{MAE} = \frac{1}{m}\sum_{i=1}^{m} | \hat{p}_i - p_i |$ and $\text{RMSE} = \sqrt{\frac{1}{m}\sum_{i=1}^{m}(\hat{p}_i - p_i)^2}$.

MAE represents the average absolute prediction error, whereas RMSE gives greater weight to larger errors. MAE and RMSE were calculated separately for each validation fold and summarised by their mean and standard deviation across the five folds. Pearson correlation quantified correspondence in relative variation between observed and predicted carrier prevalence. Fold-specific signed correlations were retained to characterise variation in performance among validation partitions. Overall correlation was calculated from all out-of-fold observed and predicted values pooled across the five folds, such that each validation record contributed one observed-predicted pair.

**2.10. Sensitivity analyses of evidence inclusion and spatial processing**

Sensitivity analyses examined the robustness of predictive performance to prespecified decisions concerning multi-location pooled estimates, Hardy-Weinberg-derived carrier prevalence, risk of bias, spatial disaggregation, and DBSCAN-classified spatial noise. Before the sensitivity datasets were constructed, records with fewer than 50 individuals tested were excluded. Each sensitivity dataset was analysed using the same Bayesian binomial geostatistical model defined in Supplementary sections 2.7–2.8. Covariate selection was applied to Dataset 1 and identified population count, *Plasmodium falciparum* incidence, consanguinity, genetic counselling availability, and prevention service availability. This fixed-effect set was held constant across sensitivity datasets, which were restricted to records with complete information for these covariates. Each model additionally included Matérn spatial and IID location-level random-effects.

Seven datasets were evaluated (Supplementary Table S9). Dataset 1 was the reference dataset and applied the record-selection hierarchy and spatial-isolation criterion described in Supplementary sections 2.5–2.6, with DBSCAN-classified spatial noise records excluded. Dataset 2 additionally excluded pooled estimates spanning multiple recruitment locations, and Dataset 3 excluded Hardy-Weinberg-derived carrier prevalence. Dataset 4 retained quality appraised records classified as having low risk of bias that also met the prespecified modelling qualification criteria described in Supplementary section 1.10. Dataset 5 extended Dataset 4 by retaining records from prespecified sources considered sufficiently authoritative for inclusion despite the absence of formal quality appraisal.[54] Dataset 6 applied population-informed spatial disaggregation to Dataset 4 before coincident-coordinate selection, with spatial noise records excluded. Dataset 7 retained DBSCAN-classified spatial noise records in Dataset 4. Datasets 6 and 7 therefore assessed sensitivity to spatial disaggregation and spatial noise exclusion, respectively.

**Supplementary Table S9. Sensitivity analyses of record inclusion and spatial processing decisions within the geostatistical modelling framework.**

| Dataset | Sensitivity condition | Comparator | Records | Mean negative log CPO (SD) | Mean MAE (SD) | Mean RMSE (SD) | Pooled *r* | Decision for subsequent analysis |
|---|---|---|---|---|---|---|---|---|
| 1 | Reference sensitivity dataset | NA | 282 | 10·493 (5·341) | 0·035 (0·003) | 0·060 (0·012) | 0·750 | Reference |
| 2 | Excluded records containing pooled estimates spanning multiple recruitment locations | Dataset 1 | 236 | 10·609 (5·158) | 0·032 (0·005) | 0·053 (0·014) | 0·355 | No additional exclusion based on multi-location pooling |
| 3 | Excluded Hardy-Weinberg-derived carrier-prevalence records | Dataset 1 | 270 | 10·206 (5·043) | 0·034 (0·003) | 0·061 (0·009) | 0·755 | No additional exclusion based on Hardy-Weinberg derivation |
| 4 | Restricted to quality appraised records with low risk of bias | Dataset 1 | 209 | 8·888 (3·861) | 0·018 (0·005) | 0·026 (0·007) | 0·569 | Low risk of bias restriction retained |
| 5 | Dataset 4 with inclusion of prespecified non-appraised trusted-source records | Dataset 4 | 223 | 9·112 (3·814) | 0·019 (0·004) | 0·028 (0·008) | 0·481 | Requirement for completed quality appraisal retained |
| 6 | Dataset 4 with population-informed spatial disaggregation of eligible coarse area-level records | Dataset 4 | 274 | 8·853 (3·810) | 0·016 (0·001) | 0·025 (0·004) | 0·573 | Spatial disaggregation retained |
| 7 | Dataset 4 with DBSCAN-classified spatial noise records retained | Dataset 4 | 222 | 8·994 (4·040) | 0·018 (0·004) | 0·026 (0·006) | 0·560 | Spatial noise records retained; DBSCAN used only to structure cross-validation |

CPO: conditional predictive ordinate. MAE: mean absolute error. RMSE: root mean square error. *r*: Pearson correlation coefficient. SD: standard deviation. DBSCAN: Density-Based Spatial Clustering of Applications with Noise. NA: not applicable. All sensitivity datasets were fitted using the same Bayesian binomial geostatistical model with a common fixed-effect covariate set. MAE and RMSE are reported as the mean (SD) across five random cross-validation folds. Pooled Pearson *r* was calculated from observed and out-of-fold posterior mean predicted carrier prevalence across all validation records and is therefore reported without an SD. Mean negative log CPO and its SD were calculated across records. Lower mean negative log CPO indicates greater single-record leave-one-out predictive support, and lower MAE and RMSE indicate smaller prediction error; Pearson correlation coefficient values approaching 1 indicate stronger positive linear association between observed and predicted values, whereas negative values indicate inverse association and poorer predictive performance than positive values of the same magnitude. Density clusters and fold assignments were reconstructed separately for each sensitivity dataset. Because datasets differed in record composition and spatial distribution, performance measures were interpreted as robustness diagnostics rather than formal paired comparisons.

Each dataset was evaluated using the procedures defined in Supplementary section 2.9. DBSCAN clusters and random cross-validation folds were reconstructed separately because changes in record inclusion or location could alter the spatial configuration of the data. MAE and RMSE were summarised across the five folds, pooled Pearson correlation was calculated from all out-of-fold predictions and mean negative log CPO summarised single-record leave-one-out predictive support. Because the datasets differed in record composition and spatial configuration, comparisons were interpreted as robustness analyses rather than paired evaluations.

Excluding pooled multi-location estimates reduced mean MAE from 0·035 (SD 0·003) to 0·032 (SD 0·005) and mean RMSE from 0·060 (SD 0·012) to 0·053 (SD 0·014) but mean negative log CPO increased from 10·493 (SD 5·341) to 10·609 (SD 5·158) and pooled Pearson r decreased from 0·750 to 0·355. The discordant results provided no consistent basis for excluding otherwise eligible pooled multi-location estimates.

Excluding Hardy-Weinberg-derived carrier prevalence produced little difference across the metrics. Mean MAE was 0·034 (SD 0·003) versus 0·035 (SD 0·003), mean RMSE was 0·061 (SD 0·009) versus 0·060 (SD 0·012), mean negative log CPO was 10·206 (SD 5·043) versus 10·493 (SD 5·341), and pooled Pearson r was 0·755 versus 0·750. No additional exclusion criterion based on Hardy-Weinberg derivation was therefore applied.

Restriction to quality-appraised records with low risk of bias reduced mean MAE to 0·018 (SD 0·005), mean RMSE to 0·026 (SD 0·007), and mean negative log CPO to 8·888 (SD 3·861), whereas pooled Pearson r decreased to 0·569. Records derived from data reported in Modell's *Haemoglobinopathologist's* Almanac and reports of the Kingdom of Bahrain Ministry of Health were prespecified for inclusion despite the absence of formal quality appraisal because these sources were considered sufficiently authoritative. However, reintroducing these records conferred no compensating advantage. Mean MAE increased to 0·019 (SD 0·004), mean RMSE increased to 0·028 (SD 0·008), mean negative log CPO increased to 9·112 (SD 3·814), and pooled Pearson r decreased to 0·481. The requirement for completed quality appraisal and a low risk of bias classification was therefore retained for subsequent model development.

Population-informed spatial disaggregation gave mean MAE 0·016 (SD 0·001), mean RMSE 0·025 (SD 0·004), mean negative log CPO 8·853 (SD 3·810) and pooled Pearson r 0·573, compared with 0·018 (SD 0·005), 0·026 (SD 0·007), 8·888 (SD 3·861) and 0·569 respectively, without spatial disaggregation. These results indicated little sensitivity to spatial disaggregation and supported its use to improve the spatial representation of eligible coarse area-level records.

Similarly, retaining DBSCAN-classified spatial noise records yielded corresponding values of 0·018 (SD 0·004), 0·026 (SD 0·006), 8·994 (SD 4·040), and 0·560. Spatial noise records remained in the training sets but were not assigned to validation folds, so this comparison assessed their influence on prediction at clustered held-out records rather than prediction at the spatial noise records themselves. No material influence was observed to justify discarding the additional geographic information. Thus, DBSCAN classification was retained for structuring cross-validation rather than determining record eligibility.

Subsequent analyses therefore retained records with low risk of bias, applied population-informed spatial disaggregation to eligible coarse area-level records, retained DBSCAN-classified spatial noise records, and imposed no additional exclusion based solely on multi-location pooling or Hardy-Weinberg derivation. Subsequent model development is described in Supplementary section 2.11, and full sensitivity analysis results are reported in Supplementary Table S9.

**2.11. Model comparison and sequential model development**

Model development proceeded after the record inclusion and spatial-processing decisions established by the sensitivity analyses (Supplementary section 2.10). Applying the supported decisions yielded the baseline β-thalassaemia carrier prevalence modelling dataset from which the subsequent model development datasets were derived. Restriction to quality-appraised records at low risk of bias reduced the dataset to 821 records, and exclusion of ADM0 records representing geographic units larger than 20 000 km² further reduced it to 791 records. Population-informed spatial disaggregation increased the number of records to 1019. Coincident coordinate selection reduced these to 383 records at distinct modelling coordinates, and exclusion of three spatially isolated records reduced the dataset to 380 records. Among records entering spatial disaggregation, 14 ADM0 records yielded 14 ADM1 analytical proxies, and 212 ADM1 records yielded 465 ADM2 proxies; of the latter, 66 parent records generated one proxy, 39 generated two proxies, and 107 generated three proxies. Records that did not yield a valid proxy were not carried forward.

Covariate values were extracted for these 380 records, and covariate selection was repeated using the procedure described in Supplementary section 2.3, independently of the fixed-effect set used for the sensitivity analyses. Selection was undertaken among records with complete information for the candidate covariates and retained six covariates: population count, *Plasmodium falciparum* incidence, degree of urbanisation, consanguinity, ethnic fractionalisation, and genetic counselling availability. Records with complete information for these six covariates formed the covariate-restricted dataset, comprising 290 records from 83 sources across 11 countries. Density-based spatial clustering and random cross-validation folds were then constructed for this dataset.

Six candidate formulations were compared, all containing an intercept; $M_1$: $x_i^T\beta$, $M_2$: $u_i$, $M_3$: $x_i^T\beta + u_i$, $M_4$: $S(s_i)$, $M_5$: $S(s_i) + u_i$ and $M_6$: $x_i^T\beta + S(s_i) + u_i$. Here $x_i^T\beta$ denotes the selected covariate fixed-effects, $S(s_i)$ the Matérn spatial random-effect, and $u_i$ the IID location-level random-effect. The binomial likelihood, mesh construction rules, priors, numerical integration, posterior-draw procedure, density-based spatial clustering, and random cross-validation design were held constant across formulations. The comparison therefore assessed, conditional on the selected covariate set, whether covariates added predictive information beyond residual spatially structured and unstructured variation, and whether either random-effect component could be omitted without materially reducing model adequacy. Covariate selection was performed using all records in the covariate-restricted dataset and was not repeated within individual cross-validation training folds.

For the covariate-restricted dataset, the $M_5\colon S(s_i) + u_i$ formulation provided the strongest overall balance of full-data fit and random cross-validation performance. Inclusion of the selected covariates in the otherwise equivalent $M_6\colon x_i^T \beta + S(s_i) + u_i$ formulation did not improve held-out prediction (Supplementary Table S10).

**Supplementary Table S10. Model evaluation and random five-fold cross-validation performance of candidate model formulations across alternative β-thalassaemia carrier-prevalence datasets.**

| | | Evaluation metrics | | Random cross-validation metrics | | | | | |
|---|---|---|---|---|---|---|---|---|---|
| **Dataset** | **Model** | **WAIC** | **Mean negative log CPO (SD)** | **Fold** | **Test records** | **Training records** | **MAE** | **RMSE** | **Pearson correlation coefficient** |
| Covariate-restricted | Covariate fixed-effects | NA | NA | 1 | 54 | 236 | 0·029 | 0·041 | 0·095 |
| | | NA | NA | 2 | 54 | 236 | 0·026 | 0·035 | 0·150 |
| | | NA | NA | 3 | 52 | 238 | 0·025 | 0·036 | 0·156 |
| | | NA | NA | 4 | 50 | 240 | 0·022 | 0·029 | 0·249 |
| | | NA | NA | 5 | 49 | 241 | 0·032 | 0·045 | -0·069 |
| | | 1631002 | 160·226 (251·297) | Summary | NA | NA | 0·027 (0·004) | 0·037 (0·006) | 0·098 |
| | IID random-effect | NA | NA | 1 | 54 | 236 | 0·025 | 0·033 | 0·010 |
| | | NA | NA | 2 | 54 | 236 | 0·023 | 0·028 | 0·007 |
| | | NA | NA | 3 | 52 | 238 | 0·023 | 0·031 | 0·061 |
| | | NA | NA | 4 | 50 | 240 | 0·019 | 0·024 | -0·223 |
| | | NA | NA | 5 | 49 | 241 | 0·025 | 0·033 | 0·038 |
| | | 2365 | 10·754 (5·617) | Summary | NA | NA | 0·023 (0·003) | 0·030 (0·004) | -0·017 |
| | Covariate fixed-effects and IID random-effect | | | 1 | 54 | 236 | 0·025 | 0·032 | 0·289 |
| | | NA | NA | 2 | 54 | 236 | 0·025 | 0·029 | 0·120 |
| | | NA | NA | 3 | 52 | 238 | 0·026 | 0·034 | 0·026 |
| | | NA | NA | 4 | 50 | 240 | 0·020 | 0·024 | 0·220 |
| | | NA | NA | 5 | 49 | 241 | 0·025 | 0·032 | 0·284 |
| | | 2370 | 10·645 (5·543) | Summary | NA | NA | 0·024 (0·002) | 0·030 (0·004) | 0·185 |
| | Spatial random-effect | NA | NA | 1 | 54 | 236 | 0·063 | 0·075 | 0·188 |
| | | NA | NA | 2 | 54 | 236 | 0·085 | 0·096 | 0·297 |
| | | NA | NA | 3 | 52 | 238 | 0·021 | 0·033 | 0·276 |
| | | NA | NA | 4 | 50 | 240 | 0·120 | 0·166 | -0·105 |
| | | NA | NA | 5 | 49 | 241 | 0·029 | 0·056 | 0·409 |
| | | 4274 | 12·113 (17·958) | Summary | NA | NA | 0·063 (0·041) | 0·085 (0·051) | 0·102 |
| | Spatial and IID random-effects | NA | NA | 1 | 54 | 236 | 0·015 | 0·025 | 0·664 |
| | | NA | NA | 2 | 54 | 236 | 0·014 | 0·018 | 0·745 |
| | | NA | NA | 3 | 52 | 238 | 0·017 | 0·031 | 0·400 |
| | | NA | NA | 4 | 50 | 240 | 0·014 | 0·020 | 0·597 |
| | | NA | NA | 5 | 49 | 241 | 0·017 | 0·026 | 0·611 |
| | | 2339 | 8·864 (3·898) | Summary | NA | NA | 0·016 (0·001) | 0·024 (0·005) | 0·588 |
| | Covariate fixed-effects, spatial random-effect, and IID random-effect | NA | NA | 1 | 54 | 236 | 0·016 | 0·025 | 0·656 |
| | | NA | NA | 2 | 54 | 236 | 0·015 | 0·019 | 0·722 |
| | | NA | NA | 3 | 52 | 238 | 0·018 | 0·032 | 0·365 |
| | | NA | NA | 4 | 50 | 240 | 0·015 | 0·020 | 0·623 |
| | | NA | NA | 5 | 49 | 241 | 0·017 | 0·026 | 0·616 |
| | | 2338 | 8·911 (3·915) | Summary | NA | NA | 0·016 (0·001) | 0·024 (0·005) | 0·571 |
| Covariate-unrestricted | IID random-effect | NA | NA | 1 | 65 | 289 | 0·025 | 0·030 | -0·018 |
| | | NA | NA | 2 | 65 | 289 | 0·025 | 0·032 | -0·264 |
| | | NA | NA | 3 | 63 | 291 | 0·023 | 0·030 | 0·292 |
| | | NA | NA | 4 | 61 | 293 | 0·026 | 0·032 | 0·007 |
| | | NA | NA | 5 | 61 | 293 | 0·032 | 0·041 | -0·028 |
| | | 2872 | 10·903 (5·787) | Summary | NA | NA | 0·026 (0·003) | 0·033 (0·005) | -0·027 |
| | Spatial random-effect | NA | NA | 1 | 65 | 289 | 0·051 | 0·066 | 0·359 |
| | | NA | NA | 2 | 65 | 289 | 0·029 | 0·039 | 0·280 |
| | | NA | NA | 3 | 63 | 291 | 0·051 | 0·075 | 0·156 |

| Dataset | Model | Evaluation metrics<br>WAIC | Mean negative log CPO (SD) | Random cross-validation metrics<br>Fold | Test records | Training records | MAE | RMSE | Pearson correlation coefficient |
|---|---|---|---|---|---|---|---|---|---|
| | | NA | NA | 4 | 61 | 293 | 0·046 | 0·056 | 0·275 |
| | | NA | NA | 5 | 61 | 293 | 0·029 | 0·050 | 0·518 |
| | | 4821 | 11·609 (16·446) | Summary | NA | NA | 0·041 (0·011) | 0·057 (0·014) | 0·297 |
| | Spatial and IID random-effects | NA | NA | 1 | 65 | 289 | 0·012 | 0·017 | 0·803 |
| | | NA | NA | 2 | 65 | 289 | 0·015 | 0·024 | 0·604 |
| | | NA | NA | 3 | 63 | 291 | 0·014 | 0·021 | 0·736 |
| | | NA | NA | 4 | 61 | 293 | 0·016 | 0·024 | 0·636 |
| | | NA | NA | 5 | 61 | 293 | 0·019 | 0·030 | 0·698 |
| | | 2846 | 9·082 (4·060) | Summary | NA | NA | 0·015 (0·002) | 0·023 (0·005) | 0·688 |

IID: independent and identically distributed. WAIC: Watanabe-Akaike information criterion. CPO: conditional predictive ordinate. MAE: mean absolute error. RMSE: root mean square error. SD: standard deviation. NA: not applicable. Covariate fixed-effect formulations included covariates retained after covariate selection. Fold-specific MAE, RMSE, and Pearson correlation coefficients are shown for each random validation fold per dataset. In the summary rows, MAE and RMSE are reported as the mean with its standard deviation across the five folds, whereas a pooled Pearson correlation coefficient was calculated once from observed and out-of-fold posterior mean predicted carrier prevalence across all validation records. WAIC and mean negative log CPO were obtained from full-data model fits; CPO represents single-record leave-one-out predictive support and is distinct from the five-fold cross-validation results. Lower WAIC, mean negative log CPO, MAE, and RMSE indicate better fit or smaller prediction error; Pearson correlation coefficient values approaching 1 indicate stronger positive linear association between observed and predicted values in held-out data, whereas negative values indicate inverse association and poorer predictive performance than positive values of the same magnitude.

Because the preferred formulation did not require covariates, subsequent model development returned to the 380-record baseline modelling dataset, without imposing the covariate availability restriction. Spatial disaggregation, coincident-coordinate selection, and nearest-neighbour isolation had already been applied in constructing this dataset and were therefore not repeated. Density-based spatial clustering and random cross-validation folds were reconstructed because both depend on the composition and spatial configuration of the dataset being analysed.

For final model development, 26 records from Liberia and Vanuatu were additionally excluded because both countries were geographically disconnected from the broader evidence-supported domain by large unsampled areas. The resulting final covariate-unrestricted dataset comprised 354 records from 110 sources across 25 countries, defining the prediction domain. Of these records, 20 were classified as spatial noise and 150 were spatially disaggregated analytical proxies, comprising two generated through ADM0 to ADM1 disaggregation and 148 through ADM1 to ADM2 disaggregation; of the latter, 24 parent records generated one proxy, 14 generated two proxies, and 32 generated three proxies (Supplementary Table S11). The temporal distribution of sources contributing to this dataset is shown in Supplementary Figure S15, the geographic distribution of records by sample size in Supplementary Figure S16, and the temporal distribution of records by country, together with their sample sizes and record density, in Supplementary Figure S17. Density-cluster assignments for the covariate-unrestricted dataset are shown in Supplementary Figure S18.

**Supplementary Table S11. Characteristics of alternative β-thalassaemia carrier prevalence modelling datasets.**

| Characteristic | Restricted by covariate availability | Unrestricted by covariate availability |
|---|---|---|
| **Overall dataset** | | |
| Sources | 83 | 110 |
| Records | 290 | 354 |
| Countries | 11 | 25 |
| β-thalassaemia carrier prevalence range (%) | 0-16·8 | 0-18·2 |
| Population-informed disaggregated records | 122 | 150 |
| Records from multiple recruitment areas | 53 | 72 |
| Records classified as spatial noise by density-based clustering | 15 | 20 |
| **Highest spatial resolution** | | |
| Administrative level 0 | 0 | 0 |
| Administrative level 1 | 0 | 2 |

| Characteristic | Restricted by covariate availability | Unrestricted by covariate availability |
|---|---|---|
| Administrative level 2 | 256 | 302 |
| Site-level institution or organisation coordinates | 34 | 50 |
| **Recruited population sample size** | | |
| 77-100 | 9 | 10 |
| 101-1,000 | 83 | 109 |
| 1,001-10,000 | 81 | 99 |
| 10,001-100,000 | 80 | 93 |
| 100,001-1,000,000 | 29 | 32 |
| 1,000,001-2,076,742 | 8 | 11 |
| **Recruitment period** | | |
| 1964-1970 | 0 | 1 |
| 1971-1980 | 0 | 6 |
| 1981-1990 | 0 | 2 |
| 1991-2000 | 29 | 37 |
| 2001-2010 | 145 | 172 |
| 2011-2020 | 89 | 104 |
| 2021-2023 | 27 | 32 |

Values are numbers of records unless otherwise stated.

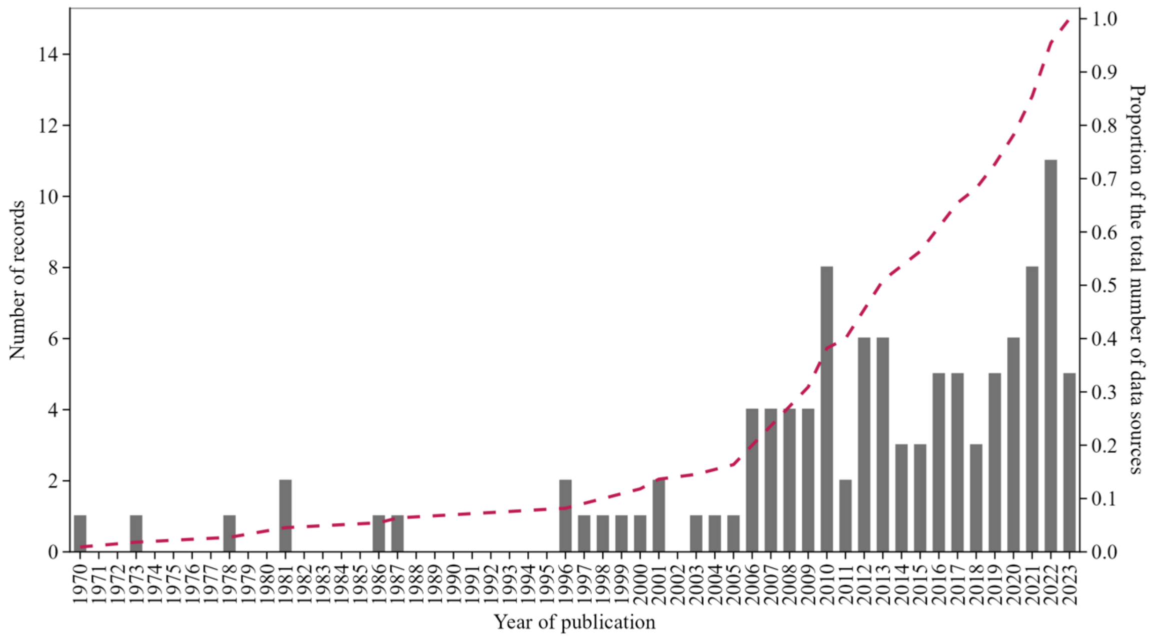

**Supplementary Figure S15. Temporal distribution of sources included in the modelling dataset.** Grey bars show the number of sources published each year. The dashed line shows the cumulative proportion of all included sources over time.

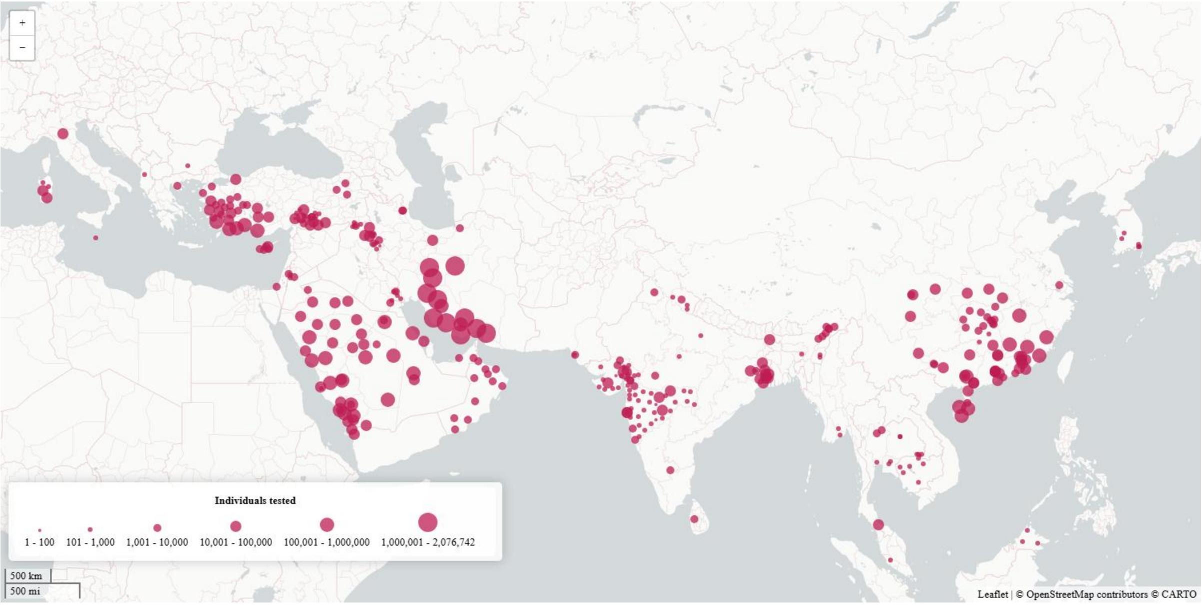

**Supplementary Figure S16. Sample size distribution of β-thalassaemia carrier-prevalence records included in the modelling dataset.**
Circle size is proportional to the number of individuals tested.

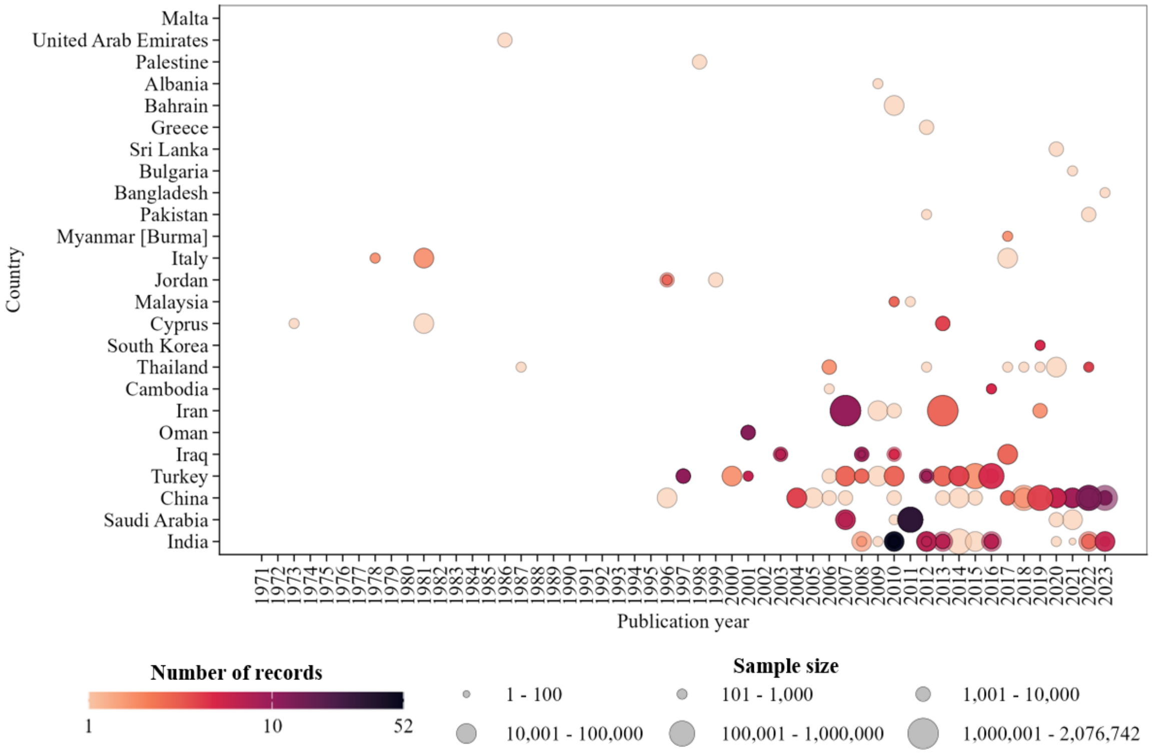

**Supplementary Figure S17. Temporal distribution of β-thalassaemia carrier-prevalence records included in the modelling dataset, by country.**
Each circle represents an individual record, positioned by country and publication year. Circle size indicates the number of individuals tested in that record, and colour indicates the number of records for the corresponding country-year combination. Multiple circles are superimposed where more than one record shares the same country and publication year. Larger circles therefore represent records with larger sample sizes, whereas darker colours indicate country-year combinations with more records.

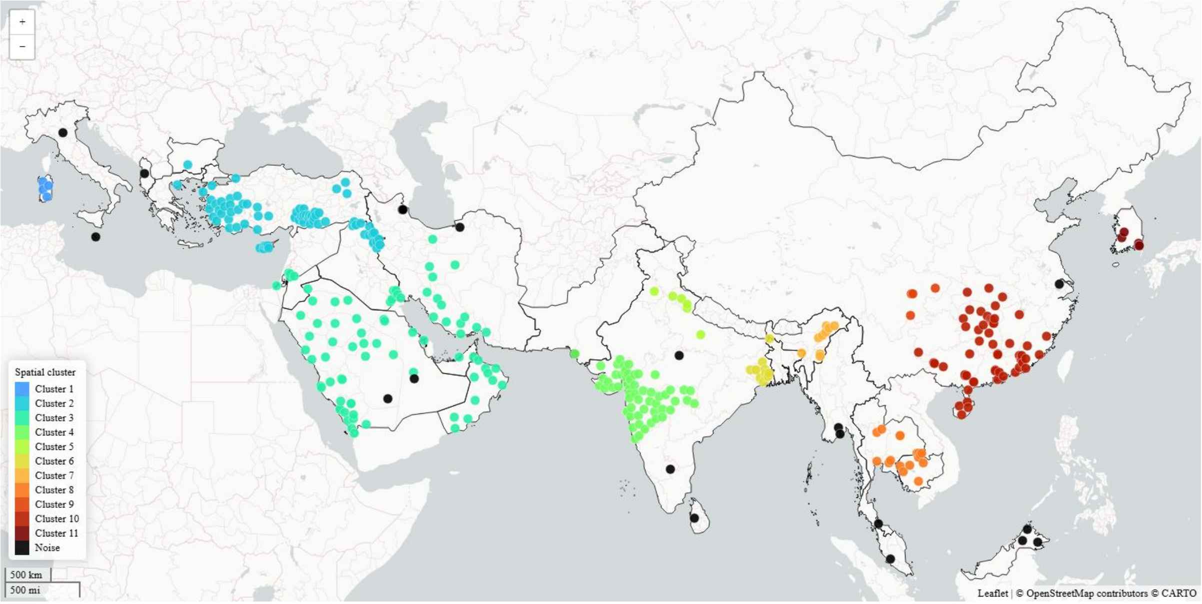

**Supplementary Figure S18: Spatial clusters of β-thalassaemia carrier-prevalence records in the modelling dataset.**

Coloured points indicate the 11 spatial clusters identified using density-based spatial clustering of applications with noise. Black points indicate spatial-noise records that were not assigned to a cluster.

Three covariate-free formulations were then compared in the covariate-unrestricted dataset, $M_2$, $M_4$ and $M_5$, using the same model fit and predictive performance criteria. The $M_5$: $S(s_i) + u_i$ formulation again provided the strongest overall random cross-validation performance while representing both residual geographic dependence and spatially unstructured residual heterogeneity (Table 1). It was therefore retained as the final model.

**2.12. Spatial five-fold cross-validation**

Spatial five-fold cross-validation was undertaken after model selection to examine performance when nearby records were deliberately withheld. This validation design addressed a different prediction problem from random cross-validation; rather than interpolation within regions surrounding observational support, it assessed prediction across larger gaps in the empirical evidence.

Records classified as spatial noise were assigned, for construction of the spatial folds only, to the nearest density cluster. Density cluster centroids and reassignment distances were calculated in a projected coordinate system. Density clusters containing ten or fewer records were then merged in their entirety with the nearest larger density cluster.

Voronoi territories were constructed around the resulting clusters, and clusters whose territories shared a boundary were treated as geographically adjacent. Whole density clusters were consolidated into five geographically contiguous folds. Partitioning targeted approximately one fifth of the records per fold; partitions within 25% of this target were preferred. When no geographically contiguous partition met this tolerance, the solution minimising the maximum proportional deviation from the target and overall imbalance between spatial folds was retained.

Each spatial fold was withheld in turn. The covariate-free formulations; $M_2$: $u_i$, $M_4$: $S(s_i)$ and $M_5$: $S(s_i) + u_i$, were refitted to the remaining four spatial folds, with the mesh and spatial projection matrices reconstructed for each training set. Predictions for the withheld records were generated from 1000 joint posterior draws and evaluated using the same MAE, RMSE, and Pearson correlation measures as random cross-validation. Spatial cross-validation was interpreted as a secondary stress test of prediction across geographic gaps rather than as a second model selection procedure. Results are reported in Supplementary Table S12.

Geographic withholding produced a substantially different performance profile from random cross-validation. For the selected formulation $M_5$, mean MAE was 0·042 (SD 0·010), mean RMSE 0·046 (SD 0·010), and pooled Pearson r -0·209; fold-specific correlations were negative in all five folds and ranged from -0·450 to -0·054. The $M_2$ formulation had lower absolute error under the same withholding design (mean MAE 0·028 [SD 0·007]; mean RMSE 0·035 [SD 0·007]) but also failed to reproduce relative geographic variation (pooled Pearson r -0·053).

The $M_4$ formulation yielded mean MAE 0·065 (SD 0·022), mean RMSE 0·070 (SD 0·022), and pooled Pearson r 0·095. Thus, the smaller absolute errors of the $M_2$ formulation did not indicate successful geographic discrimination, and none of the candidate residual structures reliably reproduced relative carrier-prevalence variation when local observational support was removed.

**Supplementary Table S12. Spatial five-fold cross-validation performance of candidate model formulations in the final covariate-unrestricted β-thalassaemia carrier-prevalence dataset.**

| Model formulation | Fold | Test records | Training records | MAE | RMSE | Pearson correlation coefficient |
|---|---|---|---|---|---|---|
| IID random-effect | 1 | 87 | 267 | 0·030 | 0·042 | 0·074 |
| | 2 | 83 | 271 | 0·029 | 0·034 | 0·047 |
| | 3 | 70 | 284 | 0·019 | 0·027 | -0·133 |
| | 4 | 47 | 307 | 0·037 | 0·043 | -0·062 |
| | 5 | 67 | 287 | 0·025 | 0·031 | 0·176 |
| | Summary | NA | NA | 0·028 (0·007) | 0·035 (0·007) | -0·053 |
| Spatial random-effect | 1 | 87 | 267 | 0·070 | 0·075 | -0·250 |
| | 2 | 83 | 271 | 0·038 | 0·042 | 0·037 |
| | 3 | 70 | 284 | 0·099 | 0·102 | -0·049 |
| | 4 | 47 | 307 | 0·058 | 0·068 | -0·114 |
| | 5 | 67 | 287 | 0·059 | 0·064 | -0·024 |
| | Summary | NA | NA | 0·065 (0·022) | 0·070 (0·022) | 0·095 |
| Spatial and IID random-effects | 1 | 87 | 267 | 0·044 | 0·050 | -0·163 |
| | 2 | 83 | 271 | 0·057 | 0·062 | -0·107 |
| | 3 | 70 | 284 | 0·032 | 0·037 | -0·054 |
| | 4 | 47 | 307 | 0·040 | 0·044 | -0·450 |
| | 5 | 67 | 287 | 0·035 | 0·039 | -0·228 |
| | Summary | NA | NA | 0·042 (0·010) | 0·046 (0·010) | -0·209 |

IID: independent and identically distributed. MAE: mean absolute error. RMSE: root mean square error. NA: not applicable. Fold-specific MAE, RMSE, and Pearson correlation coefficients are shown for each spatial validation fold. In the summary rows, MAE and RMSE are reported as the mean with its standard deviation across the five folds, whereas a pooled Pearson correlation coefficient was calculated once from observed and out-of-fold posterior mean predicted carrier prevalence across all validation records. Lower MAE, and RMSE indicate better fit or smaller prediction error; Pearson correlation coefficient values approaching 1 indicate stronger positive linear association between observed and predicted values in held-out data, whereas negative values indicate inverse association and poorer predictive performance than positive values of the same magnitude.

#### 2.13. Post-selection assessment of residual spatial dependence

Residual spatial dependence was examined after model selection in the covariate-unrestricted dataset using a baseline binomial model containing an intercept and IID location-level effects.[43] An empirical semivariogram was calculated from the posterior mean fitted IID effects using Euclidean separation in longitude-latitude coordinate space. Although these effects were modelled as geographically exchangeable, a systematic increase in semivariance with coordinate separation would indicate greater similarity between fitted effects at nearby modelling coordinates than between those farther apart.

The fitted IID effects were then randomly permuted among modelling coordinates and the empirical semivariogram recalculated using the same coordinate-space metric. Permutation preserved the fitted values and their marginal distribution while removing their geographic association. A stronger increase in semivariance with coordinate separation in the unpermuted than permuted configuration was interpreted as being consistent with residual geographic autocorrelation not represented by the IID-only formulation, $M_2$: $u_i$. The unpermuted and permuted semivariograms are shown in Supplementary Figures S19 and S20.

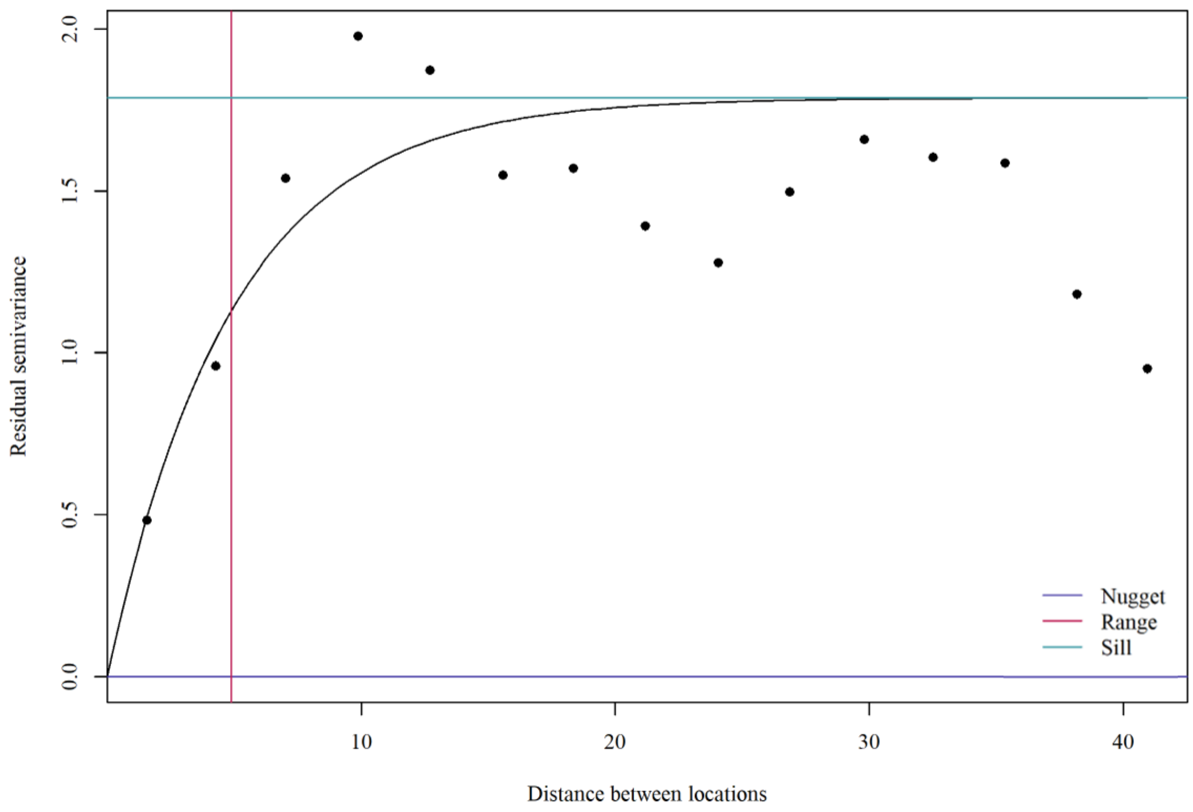

**Supplementary Figure S19. Empirical semivariogram of fitted IID effects from the baseline binomial model.**

The baseline model contained an intercept and an independent and identically distributed (IID) location-level random-effect. Points show the empirical semivariance of the posterior mean fitted IID effects according to Euclidean separation between modelling coordinates in longitude-latitude space. The solid curve shows the fitted Matérn variogram model; reference lines indicate the fitted nugget, sill, and Matérn range parameter. The increase in semivariance with coordinate separation is consistent with residual geographic autocorrelation not represented by the IID-only formulation.

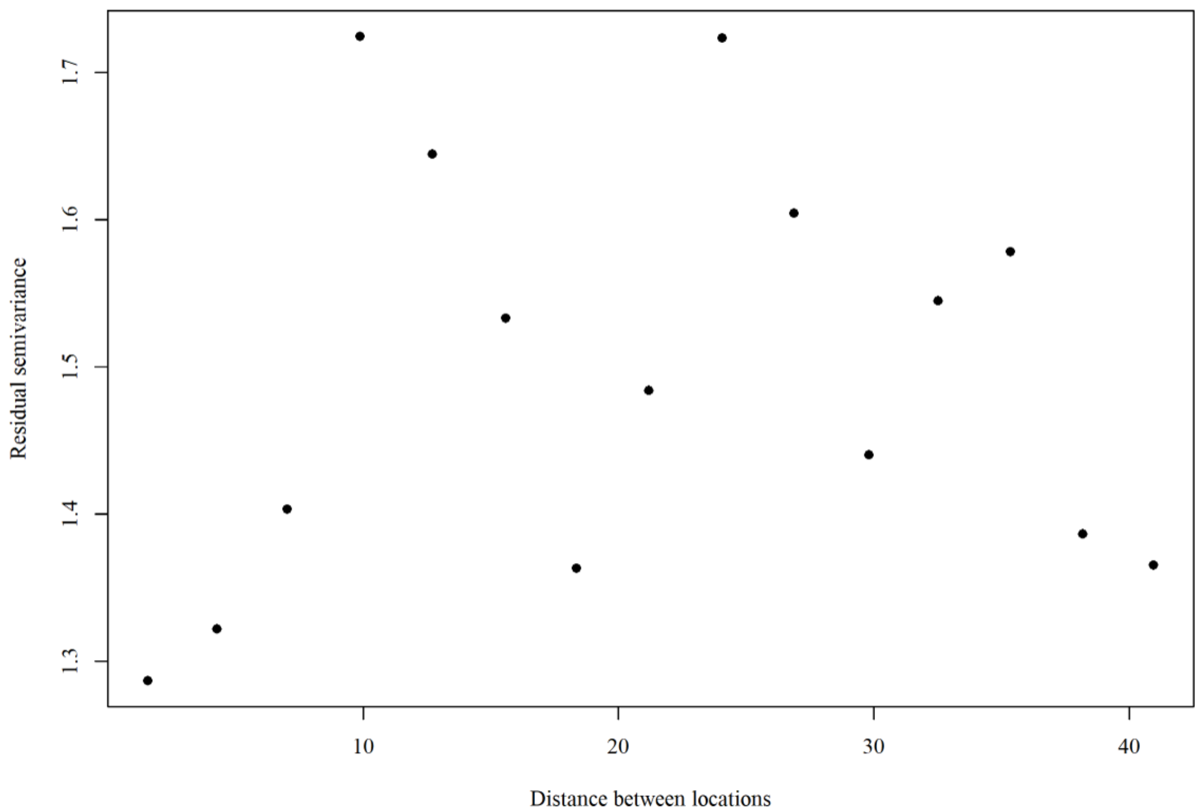

**Supplementary Figure S20. Empirical semivariogram after spatial permutation of fitted IID effects from the baseline binomial model.**
Posterior mean fitted IID effects were randomly permuted among modelling coordinates, preserving their values and marginal distribution while removing their geographic association. Points show the empirical semivariance after permutation using the same coordinate-space distance measure as in Supplementary Figure S19. The weaker distance-related pattern after permutation provides a qualitative spatial-null comparison and is consistent with the pattern in Supplementary Figure S19 reflecting geographic dependence rather than the marginal distribution of the fitted IID effects alone.

**2.14. Final model and posterior prediction**
The selected model, $logit(p_i) = \beta_0 + S(s_i) + u_i$, was fitted to the covariate-unrestricted dataset. The prediction domain comprised inhabited cells within ADM0 units represented in that dataset. The prediction grid had an approximate spatial resolution of 5 km × 5 km. Cells with zero or missing population in the 2026 WorldPop surface[26] were excluded before posterior prediction. Predictions represented carrier prevalence at a new location, including unresolved local heterogeneity not captured by the spatial field.

Posterior prediction used the joint-draw procedure described in Supplementary section 2.8. A total of 1000 joint posterior draws were obtained.

For posterior draw $j$ and prediction cell $g$, a new IID effect was generated conditional on the IID precision from the same posterior draw, $u_g^{(j)} \mid \tau_u^{(j)} \sim N\left(0, \left[\tau_u^{(j)}\right]^{-1}\right)$. The spatial field from posterior draw $j$was projected from the mesh to prediction cell $g$, giving $S_g^{(j)}$. The corresponding linear predictor was $\eta_g^{(j)} = \beta_0^{(j)} + S_g^{(j)} + u_g^{(j)}$, and predicted carrier prevalence was $p_g^{(j)} = \text{logit}^{-1}\left(\eta_g^{(j)}\right)$, $j = 1, \ldots, 1000$. Fitted IID effects at modelling coordinates were neither transferred nor spatially interpolated to prediction cells.

For prediction cell $g$, posterior quantile $q$was defined as $p_{g,q} = Q_q\left\{p_g^{(1)}, \ldots, p_g^{(1000)}\right\}$. The 1000 draws were summarised by the mean, standard deviation, median, and 2·5$^{\text{th}}$, 25$^{\text{th}}$, 75$^{\text{th}}$, and 97·5$^{\text{th}}$ percentiles. The posterior mean was used as the principal carrier-prevalence prediction. Supplementary Figure S21 overlays records from the covariate-unrestricted dataset on the posterior mean predicted carrier prevalence surface, and Supplementary Figures S22 and S23 show the 25$^{\text{th}}$- and 75$^{\text{th}}$-percentile surfaces.

For prediction cell $g$, the 95% credible interval for carrier prevalence at a new location was defined by $p_{g,0.025}$ and $p_{g,0.975}$. Prediction uncertainty was quantified as the width of this interval, $U_g = p_{g,0.975} - p_{g,0.025}$. Larger values of $U_g$ indicated less precise local prediction. The posterior standard deviation of predicted carrier prevalence was calculated as an additional measure of prediction uncertainty and is shown in Supplementary Figure S24.

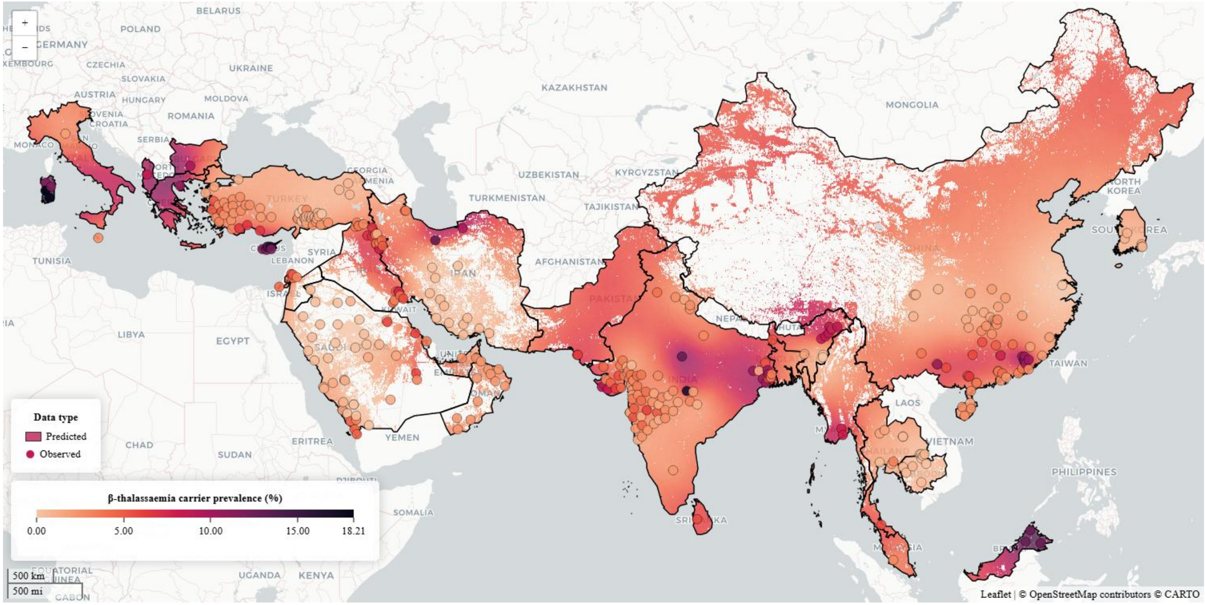

**Supplementary Figure S21. Predicted β-thalassaemia carrier prevalence with modelling coordinates.**
Map shows posterior mean predicted β-thalassaemia carrier prevalence across inhabited cells in the prediction domain. Overlaid points show the modelling coordinates of carrier-prevalence records included in the covariate-unrestricted dataset, with point colour indicating observed carrier prevalence. Darker shading indicates higher carrier prevalence.

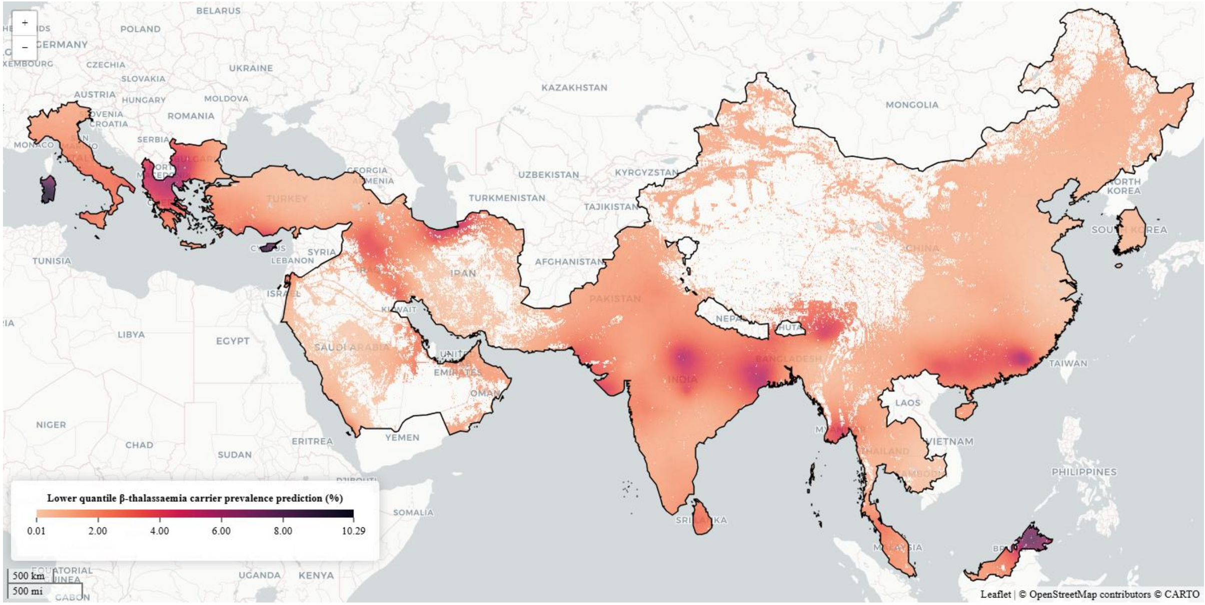

**Supplementary Figure S22. Lower quartile of predicted β-thalassaemia carrier prevalence.**
Map shows the 25th percentile of posterior predicted β-thalassaemia carrier prevalence across inhabited cells in the prediction domain. Values are expressed as percentages; Darker shading indicates higher predicted prevalence.

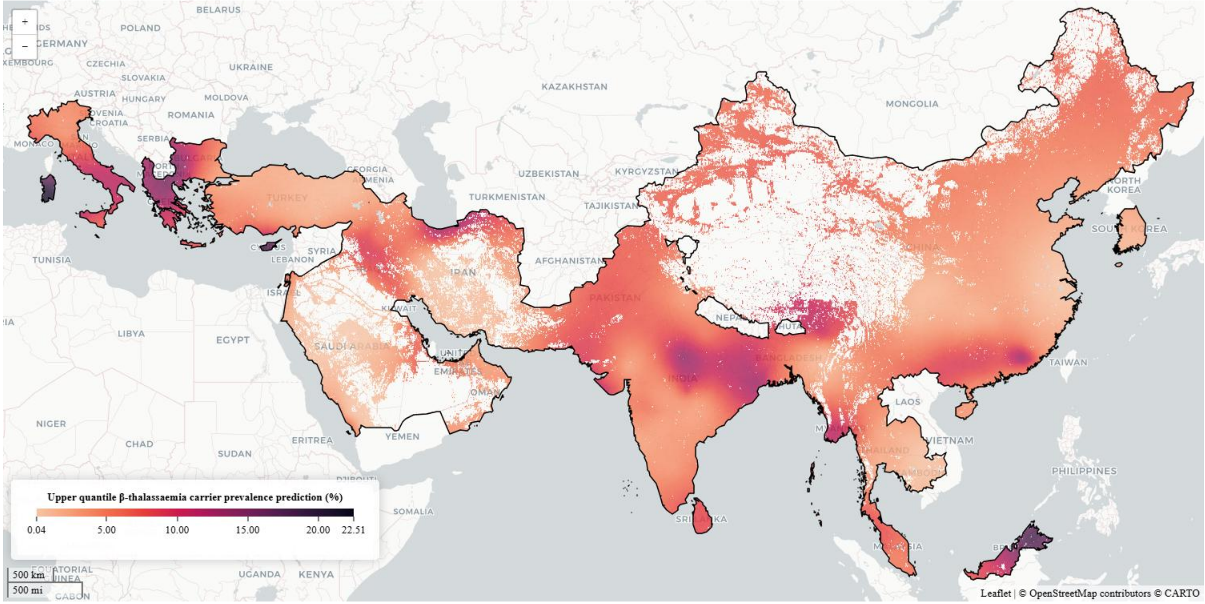

**Supplementary Figure S23. Upper quartile of predicted β-thalassaemia carrier prevalence.**
Map shows the 75th percentile of posterior predicted β-thalassaemia carrier prevalence across inhabited cells in the prediction domain. Values are expressed as percentages; Darker shading indicates higher predicted prevalence.

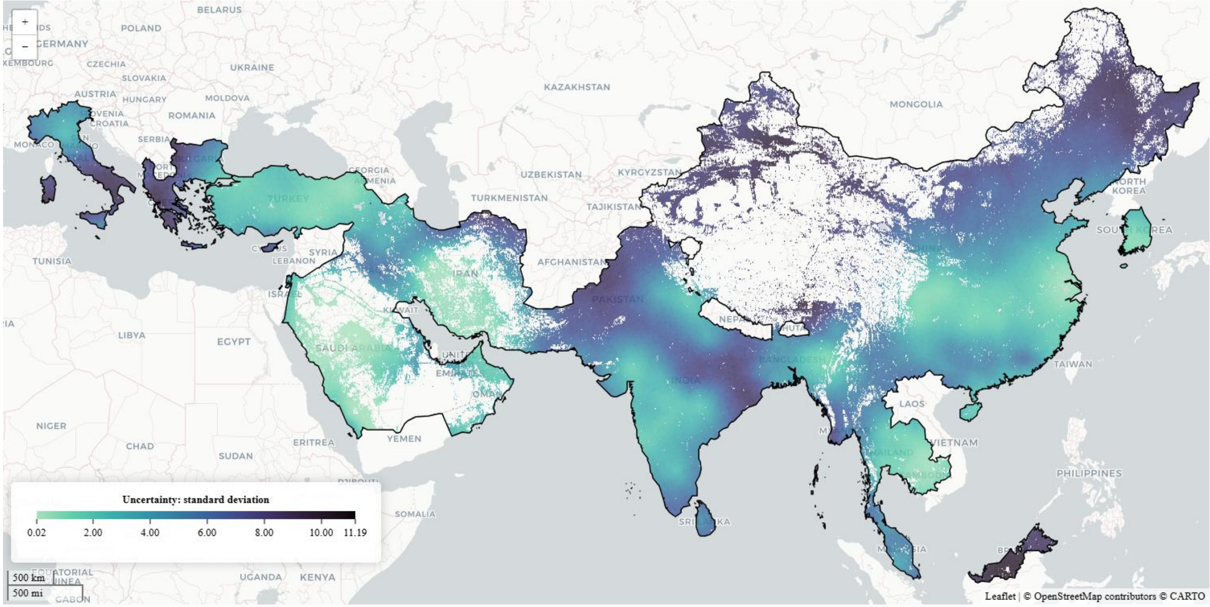

**Supplementary Figure S24. Prediction uncertainty for β-thalassaemia carrier prevalence.**
Map shows the posterior standard deviation of predicted β-thalassaemia carrier prevalence across inhabited cells in the prediction domain. Darker shading indicates greater prediction uncertainty.

#### 2.15. Carrier-number estimation and administrative aggregation

The 2026 WorldPop population surface[26] was aligned to the prediction grid using a count-preserving aggregation, conserving population totals when moving from the native population raster to the coarser prediction grid.[55,56]

For prediction cell $g$, let $N_g$ denote the corresponding population count and $\bar{p}_g$ the posterior mean predicted carrier prevalence expressed as a proportion. The estimated number of β-thalassaemia carriers was $C_g = N_g\bar{p}_g$. Carrier numbers derived from the cellwise 25th- and 75th-percentile carrier prevalence surfaces were calculated analogously as $C_g^L = N_g p_{g,0\cdot25}$ and $C_g^U = N_g p_{g,0\cdot75}$.

Carrier numbers based on the posterior mean carrier prevalence surface were summed across inhabited constituent cells within ADM0, ADM1, and ADM2 boundaries. Totals derived from the cellwise 25th- and 75th-percentile carrier prevalence surfaces were summed analogously. Cell-level posterior mean carrier prevalence, 25th- and 75th-percentile carrier prevalence, and prediction uncertainty were summarised within each administrative unit as arithmetic means across inhabited cells. National outputs are reported in Supplementary Table S13, ADM1 outputs in Supplementary Table S14, and country-specific ADM2 outputs in Supplementary Tables S15–S37.

**Supplementary Table S13. National areal predictions of β-thalassaemia carrier prevalence and estimated carrier counts.**

| | β-thalassaemia carrier prevalence | | | | | Estimated carrier counts | | |
|---|---|---|---|---|---|---|---|---|
| **ADM0** | **Mean** | **SD** | **CI95%** | **Q25** | **Q75** | **Mean** | **Q25** | **Q75** |
| Albania | 10·024 | 7·905 | 29·711 | 4·503 | 13·099 | 260487 | 120971 | 338836 |
| Bahrain | 4·061 | 3·130 | 11·394 | 1·995 | 5·151 | 61238 | 30676 | 77201 |
| Bangladesh | 4·268 | 3·735 | 13·593 | 1·854 | 5·405 | 7376193 | 3197386 | 9366395 |
| Bulgaria | 7·002 | 6·155 | 22·843 | 2·911 | 9·000 | 495629 | 211792 | 637660 |
| Cambodia | 0·730 | 0·728 | 2·553 | 0·292 | 0·903 | 132681 | 53676 | 164358 |
| China | 3·891 | 5·119 | 18·056 | 1·046 | 4·578 | 40841327 | 13979623 | 48972921 |
| Cyprus | 12·986 | 7·176 | 27·364 | 7·793 | 16·535 | 166824 | 100710 | 211730 |
| Greece | 8·416 | 7·235 | 27·032 | 3·533 | 10·877 | 726051 | 293343 | 941923 |
| India | 5·125 | 4·698 | 17·248 | 2·089 | 6·511 | 78486826 | 32582566 | 99775743 |
| Iran | 3·001 | 2·978 | 10·724 | 1·154 | 3·761 | 3815378 | 1661124 | 4853801 |
| Iraq | 5·114 | 4·169 | 15·347 | 2·366 | 6·499 | 2581200 | 1175631 | 3280701 |
| Italy | 6·226 | 6·042 | 22·022 | 2·380 | 7·892 | 3212452 | 1151508 | 4067139 |
| Jordan | 2·038 | 1·687 | 6·085 | 0·964 | 2·558 | 343535 | 180161 | 433860 |
| Malaysia | 8·047 | 7·603 | 28·146 | 3·104 | 10·278 | 1975145 | 790181 | 2516294 |
| Malta | 4·138 | 3·448 | 12·747 | 1·895 | 5·278 | 19862 | 9070 | 25248 |
| Myanmar [Burma] | 4·242 | 4·654 | 16·700 | 1·419 | 5·281 | 2762975 | 1089125 | 3473066 |
| Oman | 2·772 | 2·172 | 7·963 | 1·345 | 3·503 | 132274 | 66792 | 167639 |
| Pakistan | 5·559 | 6·898 | 25·087 | 1·468 | 6·791 | 14381588 | 4183432 | 17630255 |
| Palestine | 3·997 | 2·869 | 10·635 | 2·070 | 5·050 | 223198 | 113935 | 283642 |
| Saudi Arabia | 1·247 | 1·050 | 3·802 | 0·573 | 1·572 | 633645 | 309367 | 801673 |
| South Korea | 1·440 | 1·483 | 5·144 | 0·562 | 1·779 | 736256 | 278852 | 902942 |
| Sri Lanka | 6·318 | 5·730 | 21·144 | 2·526 | 8·142 | 1392761 | 575451 | 1795331 |
| Thailand | 2·707 | 2·685 | 9·613 | 1·043 | 3·394 | 1629228 | 671884 | 2048221 |
| Turkey | 2·303 | 1·851 | 6·699 | 1·109 | 2·897 | 2035475 | 995439 | 2563276 |
| United Arab Emirates | 3·094 | 2·591 | 9·522 | 1·399 | 3·928 | 232989 | 111993 | 294255 |

ADM0: administrative level 0. SD: standard deviation. CI95%: 95% credible interval. Q25: 25th percentile. Q75: 75th percentile. SD and CI95% are measures of prediction uncertainty.

**Supplementary Table S14. Subnational areal predictions of β-thalassaemia carrier prevalence and estimated carrier count at administrative level 1.**

| | | β-thalassaemia carrier prevalence | | | | | Estimated carrier counts | | |
|---|---|---|---|---|---|---|---|---|---|
| **ADM0** | **ADM1** | **Mean** | **SD** | **CI95%** | **Q25** | **Q75** | **Mean** | **Q25** | **Q75** |
| Albania | Berat | 10·169 | 7·948 | 29·659 | 4·627 | 13·303 | 9235 | 4212 | 12184 |
| Albania | Diber | 10·138 | 7·539 | 28·123 | 4·808 | 13·265 | 9574 | 4545 | 12490 |
| Albania | Durres | 9·672 | 7·374 | 28·071 | 4·503 | 12·563 | 22255 | 10251 | 28894 |
| Albania | Elbasan | 10·219 | 7·510 | 28·376 | 4·914 | 13·346 | 23788 | 11543 | 30908 |
| Albania | Fier | 9·825 | 7·843 | 29·438 | 4·401 | 12·787 | 26613 | 12013 | 34549 |
| Albania | Gjirokaster | 10·057 | 8·210 | 30·872 | 4·350 | 13·125 | 4748 | 2028 | 6205 |
| Albania | Korce | 10·589 | 8·074 | 30·528 | 4·903 | 13·873 | 17654 | 8231 | 23279 |
| Albania | Kukes | 10·128 | 8·369 | 31·594 | 4·288 | 13·277 | 6945 | 3003 | 9126 |
| Albania | Lezhe | 9·801 | 7·628 | 28·574 | 4·463 | 12·822 | 8937 | 4045 | 11737 |
| Albania | Shkoder | 9·601 | 8·039 | 30·160 | 4·036 | 12·533 | 17876 | 7493 | 23196 |
| Albania | Tirane | 9·802 | 7·256 | 27·204 | 4·700 | 12·759 | 96392 | 46364 | 124980 |
| Albania | Vlore | 9·839 | 8·308 | 31·078 | 4·111 | 12·888 | 10056 | 4217 | 13216 |
| Bahrain | Capital Governorate | 3·833 | 2·946 | 10·649 | 1·959 | 4·847 | 12099 | 6117 | 15310 |
| Bahrain | Muharraq Governorate | 3·799 | 3·060 | 10·552 | 1·887 | 4·765 | 9775 | 4844 | 12249 |
| Bahrain | Northern Governorate | 4·040 | 3·014 | 11·003 | 2·030 | 5·109 | 17291 | 8794 | 21643 |
| Bahrain | Southern Governorate | 4·144 | 3·211 | 11·755 | 2·007 | 5·269 | 16741 | 8209 | 21242 |
| Bangladesh | Barisal | 4·139 | 4·010 | 14·438 | 1·613 | 5·238 | 334401 | 130599 | 423103 |
| Bangladesh | Chittagong | 2·756 | 3·077 | 10·712 | 0·948 | 3·394 | 955452 | 341707 | 1183868 |
| Bangladesh | Dhaka | 3·982 | 3·552 | 12·872 | 1·689 | 5·064 | 1884915 | 790967 | 2401579 |
| Bangladesh | Khulna | 5·182 | 4·058 | 15·054 | 2·467 | 6·616 | 941493 | 454828 | 1203066 |

| | | β-thalassaemia carrier prevalence | | | | | Estimated carrier counts | | |
|---|---|---|---|---|---|---|---|---|---|
| **ADM0** | **ADM1** | **Mean** | **SD** | **CI95%** | **Q25** | **Q75** | **Mean** | **Q25** | **Q75** |
| Bangladesh | Mymensingh | 4·145 | 3·696 | 13·456 | 1·776 | 5·238 | 532488 | 227943 | 673543 |
| Bangladesh | Rajshani | 5·697 | 4·581 | 16·958 | 2·628 | 7·290 | 1210522 | 558595 | 1549571 |
| Bangladesh | Rangpur | 5·988 | 4·727 | 17·568 | 2·805 | 7·659 | 1113298 | 519799 | 1424300 |
| Bangladesh | Sylhet | 2·601 | 2·248 | 8·182 | 1·161 | 3·277 | 282546 | 127042 | 355987 |
| Bulgaria | Blagoevgrad | 10·486 | 7·549 | 28·473 | 5·163 | 13·629 | 31957 | 15704 | 41592 |
| Bulgaria | Burgas | 3·666 | 3·683 | 13·334 | 1·342 | 4·642 | 12209 | 4361 | 15291 |
| Bulgaria | Dobrich | 3·917 | 4·721 | 16·685 | 1·125 | 4·842 | 5592 | 1632 | 6940 |
| Bulgaria | Gabrovo | 7·150 | 6·340 | 23·532 | 2·884 | 9·257 | 7114 | 2832 | 9261 |
| Bulgaria | Haskovo | 5·266 | 4·295 | 16·000 | 2·380 | 6·754 | 12275 | 5633 | 15850 |
| Bulgaria | Kardzhali | 6·093 | 4·682 | 17·388 | 2·917 | 7·809 | 8983 | 4270 | 11529 |
| Bulgaria | Kyustendil | 10·787 | 8·436 | 31·811 | 4·926 | 14·051 | 12270 | 5601 | 15996 |
| Bulgaria | Lovech | 8·214 | 7·187 | 26·563 | 3·378 | 10·641 | 9788 | 4045 | 12806 |
| Bulgaria | Montana | 9·233 | 8·767 | 33·136 | 3·434 | 11·845 | 11164 | 4198 | 14323 |
| Bulgaria | Pazardzhik | 9·302 | 6·759 | 25·364 | 4·605 | 12·014 | 21077 | 10483 | 27231 |
| Bulgaria | Pernik | 10·336 | 8·465 | 32·097 | 4·515 | 13·411 | 12285 | 5502 | 16005 |
| Bulgaria | Pleven | 7·847 | 7·564 | 28·028 | 2·888 | 10·116 | 18420 | 6802 | 23843 |
| Bulgaria | Plovdiv | 7·921 | 5·931 | 22·181 | 3·829 | 10·233 | 52431 | 26055 | 67729 |
| Bulgaria | Razgrad | 5·134 | 5·559 | 20·101 | 1·627 | 6·551 | 5281 | 1698 | 6757 |
| Bulgaria | Ruse | 5·874 | 6·168 | 22·271 | 1·946 | 7·531 | 11648 | 3680 | 15035 |
| Bulgaria | Shumen | 4·546 | 4·873 | 17·754 | 1·479 | 5·772 | 7323 | 2381 | 9273 |
| Bulgaria | Silistra | 4·800 | 5·576 | 19·937 | 1·389 | 6·037 | 4808 | 1389 | 5976 |
| Bulgaria | Sliven | 5·372 | 5·030 | 18·453 | 2·090 | 6·873 | 9315 | 3642 | 11925 |
| Bulgaria | Smolyan | 8·043 | 5·695 | 21·404 | 4·072 | 10·359 | 7576 | 3793 | 9749 |
| Bulgaria | Sofia | 9·675 | 7·920 | 29·885 | 4·276 | 12·498 | 22472 | 10057 | 28967 |
| Bulgaria | Sofia City | 10·030 | 8·240 | 31·087 | 4·405 | 12·922 | 133133 | 58618 | 171772 |
| Bulgaria | Stara Zagora | 6·423 | 5·331 | 19·781 | 2·821 | 8·294 | 19286 | 8437 | 24748 |
| Bulgaria | Targovishte | 5·364 | 5·455 | 20·009 | 1·881 | 6·829 | 5221 | 1802 | 6618 |
| Bulgaria | Varna | 3·844 | 4·265 | 15·381 | 1·238 | 4·813 | 14162 | 4458 | 17399 |
| Bulgaria | Veliko Tarnovo | 6·411 | 6·120 | 22·579 | 2·402 | 8·233 | 12504 | 4673 | 16045 |
| Bulgaria | Vidin | 9·062 | 9·082 | 34·115 | 3·132 | 11·570 | 6168 | 2063 | 7883 |
| Bulgaria | Vratsa | 8·906 | 8·270 | 30·997 | 3·417 | 11·444 | 13951 | 5307 | 17916 |
| Bulgaria | Yambol | 4·424 | 4·107 | 15·005 | 1·747 | 5·654 | 5334 | 2083 | 6874 |
| Cambodia | Bantey Meanchey | 0·974 | 0·770 | 2·808 | 0·474 | 1·226 | 9193 | 4497 | 11568 |
| Cambodia | Battambang | 0·887 | 0·706 | 2·568 | 0·428 | 1·119 | 10257 | 5036 | 12947 |
| Cambodia | Kampong Cham | 0·580 | 0·579 | 1·998 | 0·236 | 0·719 | 5980 | 2437 | 7387 |
| Cambodia | Kampong Chhnang | 0·592 | 0·541 | 1·916 | 0·258 | 0·738 | 3666 | 1587 | 4568 |
| Cambodia | Kampong Speu | 0·761 | 0·746 | 2·655 | 0·305 | 0·946 | 7494 | 3010 | 9309 |
| Cambodia | Kampong Thom | 0·503 | 0·459 | 1·632 | 0·220 | 0·627 | 4104 | 1795 | 5126 |
| Cambodia | Kampot | 1·048 | 1·133 | 4·021 | 0·367 | 1·296 | 7161 | 2481 | 8843 |
| Cambodia | Kandal | 0·712 | 0·731 | 2·528 | 0·280 | 0·880 | 10037 | 3992 | 12387 |
| Cambodia | Kep | 1·179 | 1·345 | 4·546 | 0·381 | 1·468 | 515 | 166 | 638 |
| Cambodia | Koh Kong | 1·026 | 1·046 | 3·692 | 0·390 | 1·281 | 1438 | 541 | 1815 |
| Cambodia | Kratie | 0·638 | 0·720 | 2·468 | 0·223 | 0·781 | 3069 | 1064 | 3744 |
| Cambodia | Mondulkiri | 0·856 | 1·082 | 3·687 | 0·256 | 1·025 | 981 | 276 | 1168 |
| Cambodia | Oddar Meanchey | 0·714 | 0·594 | 2·140 | 0·335 | 0·896 | 2256 | 1056 | 2826 |
| Cambodia | Pailin | 1·069 | 0·872 | 3·119 | 0·505 | 1·352 | 983 | 472 | 1243 |
| Cambodia | Phnom Penh | 0·669 | 0·676 | 2·306 | 0·271 | 0·825 | 16867 | 6767 | 20897 |
| Cambodia | Preah Sihanouk | 1·150 | 1·271 | 4·472 | 0·394 | 1·425 | 2982 | 972 | 3669 |
| Cambodia | Preah Vihear | 0·451 | 0·376 | 1·355 | 0·211 | 0·565 | 1288 | 581 | 1605 |
| Cambodia | Prey Veng | 0·760 | 0·860 | 2·877 | 0·275 | 0·927 | 9693 | 3512 | 11819 |
| Cambodia | Pursat | 0·739 | 0·639 | 2·297 | 0·333 | 0·928 | 3290 | 1510 | 4138 |
| Cambodia | Ratanakiri Province | 0·828 | 0·995 | 3·424 | 0·260 | 1·008 | 2072 | 647 | 2532 |
| Cambodia | Siem Reap | 0·605 | 0·487 | 1·761 | 0·292 | 0·761 | 7267 | 3496 | 9139 |
| Cambodia | Stung Treng | 0·559 | 0·581 | 2·018 | 0·214 | 0·690 | 969 | 380 | 1215 |
| Cambodia | Svay Rieng | 0·933 | 1·207 | 3·911 | 0·294 | 1·105 | 5782 | 1822 | 6852 |
| Cambodia | Takeo | 0·904 | 0·978 | 3·426 | 0·326 | 1·117 | 9460 | 3484 | 11734 |
| Cambodia | Tbong Khmum | 0·697 | 0·787 | 2·693 | 0·247 | 0·850 | 5924 | 2132 | 7251 |

| ADM0 | ADM1 | β-thalassaemia carrier prevalence Mean | SD | CI95% | Q25 | Q75 | Estimated carrier counts Mean | Q25 | Q75 |
|---|---|---|---|---|---|---|---|---|---|
| China | Anhui Province | 1·143 | 1·447 | 4·886 | 0·341 | 1·373 | 725197 | 206653 | 867717 |
| China | Beijing Municipality | 3·643 | 5·817 | 19·884 | 0·632 | 4·092 | 790793 | 138066 | 893877 |
| China | Chongqing Municipality | 1·566 | 1·490 | 5·300 | 0·650 | 1·955 | 434016 | 177959 | 539838 |
| China | Fujian Province | 3·139 | 2·438 | 8·947 | 1·536 | 3·965 | 1156390 | 559350 | 1458755 |
| China | Gansu Province | 3·869 | 5·917 | 20·524 | 0·762 | 4·369 | 876589 | 181024 | 990206 |
| China | Guangxi Zhuang Autonomous Region | 6·254 | 4·600 | 17·203 | 3·104 | 8·013 | 3067487 | 1548624 | 3924797 |
| China | Guangzhou Province | 5·869 | 3·885 | 14·595 | 3·184 | 7·459 | 5874950 | 3236522 | 7436334 |
| China | Guizhou Province | 3·087 | 2·884 | 10·433 | 1·259 | 3·903 | 1196426 | 481085 | 1511803 |
| China | Hainan Province | 2·878 | 2·133 | 7·868 | 1·458 | 3·641 | 216332 | 109199 | 272804 |
| China | Hebei Province | 3·512 | 5·645 | 19·382 | 0·620 | 3·878 | 2322116 | 420183 | 2580136 |
| China | Heilongjiang Province | 4·816 | 7·297 | 26·033 | 0·849 | 5·502 | 1445289 | 252680 | 1660423 |
| China | Henan Province | 2·703 | 3·688 | 12·833 | 0·684 | 3·165 | 2748777 | 666940 | 3188730 |
| China | Hong Kong Special Administrative Region | 3·455 | 2·455 | 9·017 | 1·819 | 4·357 | 81455 | 42387 | 102821 |
| China | Hubei Province | 2·204 | 1·981 | 7·126 | 0·954 | 2·770 | 1207799 | 543843 | 1521090 |
| China | Hunan Province | 2·932 | 2·093 | 7·765 | 1·522 | 3·711 | 1919871 | 1013417 | 2425271 |
| China | Inner Mongolia Autonomous Region | 4·576 | 7·184 | 25·138 | 0·788 | 5·140 | 1081160 | 185226 | 1212128 |
| China | Jiangsu Province | 0·665 | 0·949 | 3·076 | 0·177 | 0·769 | 480396 | 135879 | 561346 |
| China | Jiangxi Province | 3·173 | 2·486 | 9·136 | 1·534 | 4·018 | 1358467 | 653847 | 1718665 |
| China | Jilin Province | 4·237 | 6·680 | 22·775 | 0·749 | 4·794 | 998724 | 175335 | 1131075 |
| China | Liaoning Province | 3·636 | 5·851 | 19·995 | 0·642 | 4·030 | 1350856 | 236663 | 1483644 |
| China | Macau Special Administrative Region | 3·413 | 2·407 | 9·801 | 1·799 | 4·301 | 4554 | 2400 | 5739 |
| China | Ningxia Ningxia Hui Autonomous Region | 3·845 | 5·855 | 20·211 | 0·747 | 4·385 | 268394 | 50992 | 303814 |
| China | Qinghai Province | 4·821 | 7·118 | 25·606 | 0·932 | 5·529 | 251317 | 48947 | 285647 |
| China | Shaanxi Province | 3·205 | 4·473 | 15·393 | 0·740 | 3·793 | 1208728 | 296375 | 1443942 |
| China | Shandong Province | 2·086 | 3·411 | 11·312 | 0·398 | 2·287 | 1959621 | 375440 | 2150244 |
| China | Shanghai Municipality | 0·426 | 0·532 | 1·749 | 0·139 | 0·504 | 32108 | 10541 | 37847 |
| China | Shanxi Province | 3·768 | 5·980 | 20·687 | 0·693 | 4·183 | 1268181 | 237100 | 1415122 |
| China | Sichuan Province | 1·779 | 1·898 | 6·681 | 0·649 | 2·193 | 1178991 | 505506 | 1471659 |
| China | Tianjin Municipality | 3·134 | 4·885 | 16·571 | 0·571 | 3·585 | 423781 | 77877 | 480282 |
| China | Tibet Autonomous Region | 6·822 | 7·498 | 27·696 | 2·102 | 8·617 | 251066 | 71520 | 316856 |
| China | Xinjiang Uyghur Autonomous Region | 5·302 | 8·220 | 28·914 | 0·946 | 5·976 | 1389326 | 247160 | 1571213 |
| China | Yunnan Province | 4·246 | 5·056 | 17·891 | 1·260 | 5·227 | 1894166 | 591270 | 2344472 |
| China | Zhejiang Province | 0·967 | 1·166 | 3·916 | 0·315 | 1·172 | 526250 | 165790 | 633993 |
| Cyprus | Famagusta | 11·356 | 6·830 | 26·053 | 6·465 | 14·515 | 20318 | 11756 | 25990 |
| Cyprus | Kyrenia | 12·577 | 6·953 | 26·652 | 7·577 | 15·969 | 14567 | 8734 | 18505 |
| Cyprus | Larnaca | 13·313 | 7·184 | 27·384 | 8·069 | 16·996 | 18942 | 11353 | 24043 |
| Cyprus | Limassol | 14·142 | 7·573 | 28·946 | 8·641 | 18·037 | 33489 | 20358 | 42667 |
| Cyprus | Nicosia | 13·321 | 7·105 | 27·038 | 8·164 | 16·928 | 67129 | 41129 | 84808 |
| Cyprus | Paphos | 13·648 | 7·607 | 28·971 | 8·133 | 17·336 | 12069 | 7196 | 15326 |
| Greece | Agion Oros | 7·799 | 5·556 | 20·786 | 3·945 | 9·994 | 93 | 47 | 118 |
| Greece | Attica | 7·187 | 7·213 | 26·864 | 2·471 | 9·271 | 247886 | 86159 | 323721 |
| Greece | Crete | 6·085 | 6·983 | 25·389 | 1·741 | 7·684 | 33158 | 9513 | 41495 |
| Greece | Egean | 4·958 | 4·064 | 15·037 | 2·261 | 6·323 | 16639 | 7718 | 21295 |
| Greece | Epirus-Western Macedonia | 10·430 | 8·386 | 31·434 | 4·592 | 13·634 | 52244 | 22559 | 68411 |
| Greece | Macedonia-Thrace | 8·787 | 6·194 | 23·342 | 4·429 | 11·378 | 186905 | 95335 | 243070 |
| Greece | Peloponisos-W· Greece & Ionian | 8·006 | 8·343 | 31·104 | 2·616 | 10·230 | 90673 | 30365 | 115952 |
| Greece | Thessalia-Central Greece | 8·974 | 7·691 | 28·766 | 3·733 | 11·663 | 89989 | 38148 | 117026 |
| India | Andaman and Nicobar Islands | 6·301 | 8·113 | 29·380 | 1·617 | 7·554 | 17180 | 4467 | 20369 |
| India | Andhra Pradesh | 3·617 | 4·382 | 15·686 | 1·044 | 4·414 | 1970614 | 545768 | 2395899 |
| India | Arunachal Pradesh | 7·525 | 6·048 | 22·637 | 3·417 | 9·695 | 127863 | 63949 | 164646 |
| India | Assam | 5·370 | 4·061 | 15·123 | 2·612 | 6·868 | 2115587 | 1013811 | 2709871 |
| India | Bihar | 7·638 | 7·168 | 26·658 | 2·905 | 9·842 | 10241737 | 3861812 | 13210766 |
| India | Chandigarh | 3·594 | 2·813 | 10·241 | 1·768 | 4·502 | 49609 | 24415 | 62241 |

| ADM0 | ADM1 | β-thalassaemia carrier prevalence Mean | SD | CI95% | Q25 | Q75 | Estimated carrier counts Mean | Q25 | Q75 |
|---|---|---|---|---|---|---|---|---|---|
| India | Chhattisgarh | 5·912 | 5·912 | 21·659 | 2·127 | 7·541 | 1992940 | 718189 | 2548573 |
| India | Dadra and Nagar Haveli and Daman and Diu | 3·953 | 2·689 | 10·090 | 2·121 | 5·003 | 39462 | 21062 | 49531 |
| India | Delhi | 4·516 | 4·559 | 16·380 | 1·701 | 5·641 | 1032993 | 387847 | 1288049 |
| India | Goa | 2·144 | 2·006 | 7·066 | 0·919 | 2·664 | 30203 | 13133 | 37617 |
| India | Gujarat | 4·590 | 3·149 | 11·751 | 2·436 | 5·826 | 3212387 | 1735109 | 4070217 |
| India | Haryana | 4·531 | 4·519 | 16·414 | 1·716 | 5·679 | 1464440 | 557838 | 1835229 |
| India | Himachal Pradesh | 3·634 | 3·473 | 12·532 | 1·439 | 4·586 | 291307 | 121995 | 369582 |
| India | Jammu and Kashmir | 4·724 | 5·864 | 21·126 | 1·258 | 5·794 | 681000 | 184036 | 835350 |
| India | Jharkhand | 8·917 | 7·266 | 27·255 | 3·964 | 11·531 | 3741824 | 1717929 | 4835085 |
| India | Karnataka | 2·708 | 2·668 | 9·502 | 1·064 | 3·389 | 1953745 | 786463 | 2446996 |
| India | Kerala | 4·686 | 5·643 | 20·209 | 1·310 | 5·807 | 1721967 | 473259 | 2124354 |
| India | Ladakh | 4·577 | 6·200 | 21·899 | 1·095 | 5·432 | 18460 | 4390 | 22013 |
| India | Madhya Pradesh | 7·158 | 5·856 | 21·877 | 3·176 | 9·252 | 6342960 | 2798641 | 8183940 |
| India | Maharashtra | 3·477 | 2·430 | 9·039 | 1·832 | 4·403 | 4784888 | 2557519 | 6057321 |
| India | Manipur | 1·490 | 1·310 | 4·697 | 0·663 | 1·869 | 44094 | 19872 | 55481 |
| India | Meghalaya | 3·985 | 3·445 | 12·619 | 1·739 | 5·069 | 150139 | 65698 | 191548 |
| India | Mizoram | 1·733 | 1·962 | 6·666 | 0·614 | 2·117 | 22154 | 8004 | 27103 |
| India | Nagaland | 4·373 | 3·112 | 11·555 | 2·263 | 5·549 | 89216 | 46130 | 113175 |
| India | Odisha | 7·414 | 7·460 | 27·424 | 2·594 | 9·505 | 3879991 | 1395582 | 4993345 |
| India | Puducherry | 4·968 | 6·060 | 21·361 | 1·483 | 5·996 | 59020 | 17511 | 70511 |
| India | Punjab | 4·270 | 3·922 | 14·270 | 1·768 | 5·402 | 1369670 | 579196 | 1732007 |
| India | Rajasthan | 4·991 | 5·349 | 19·431 | 1·645 | 6·305 | 4223586 | 1427754 | 5328178 |
| India | Sikkim | 6·854 | 6·347 | 23·506 | 2·678 | 8·807 | 47378 | 19879 | 60965 |
| India | Tamil Nadu | 4·698 | 5·269 | 19·059 | 1·469 | 5·877 | 3698986 | 1152777 | 4612902 |
| India | Telangana | 2·798 | 2·864 | 10·191 | 1·045 | 3·498 | 1204746 | 448064 | 1508722 |
| India | Tripura | 2·293 | 2·345 | 8·186 | 0·892 | 2·842 | 111846 | 44026 | 139384 |
| India | Uttar Pradesh | 5·599 | 5·079 | 18·725 | 2·294 | 7·129 | 13978291 | 5565553 | 17770983 |
| India | Uttarakhand | 2·957 | 2·357 | 8·640 | 1·410 | 3·738 | 367173 | 182738 | 464869 |
| India | West Bengal | 7·377 | 4·946 | 18·599 | 3·941 | 9·415 | 7378546 | 4004577 | 9399981 |
| Iran | Alborz | 7·288 | 5·671 | 21·088 | 3·414 | 9·410 | 218419 | 104630 | 280222 |
| Iran | Ardabil | 2·947 | 2·331 | 8·576 | 1·411 | 3·738 | 42194 | 20615 | 53521 |
| Iran | Bushehr | 1·081 | 0·865 | 3·139 | 0·521 | 1·361 | 14081 | 6685 | 17727 |
| Iran | Chaharmahal and Bakhtiari | 0·589 | 0·496 | 1·792 | 0·275 | 0·738 | 5997 | 2844 | 7476 |
| Iran | East Azerbaijan | 3·022 | 2·534 | 9·241 | 1·376 | 3·825 | 139743 | 62673 | 176252 |
| Iran | Fars | 0·619 | 0·494 | 1·790 | 0·301 | 0·777 | 34021 | 16754 | 42666 |
| Iran | Gilan | 5·432 | 4·631 | 17·072 | 2·370 | 6·961 | 156496 | 67261 | 200853 |
| Iran | Golestan | 8·758 | 7·362 | 27·580 | 3·758 | 11·313 | 183301 | 84319 | 237088 |
| Iran | Hamadan | 4·243 | 3·942 | 14·434 | 1·683 | 5·417 | 84152 | 32836 | 107742 |
| Iran | Hormozgan | 0·844 | 0·705 | 2·545 | 0·395 | 1·058 | 13765 | 6619 | 17304 |
| Iran | Ilam | 5·502 | 4·892 | 18·042 | 2·264 | 7·073 | 38457 | 15510 | 49936 |
| Iran | Isfahan | 0·587 | 0·515 | 1·837 | 0·265 | 0·732 | 26307 | 12661 | 32970 |
| Iran | Kerman | 0·803 | 0·939 | 3·149 | 0·277 | 0·973 | 27151 | 9123 | 33009 |
| Iran | Kermanshah | 5·559 | 4·589 | 17·000 | 2·491 | 7·123 | 121728 | 53777 | 155706 |
| Iran | Khuzestan | 2·726 | 2·269 | 8·276 | 1·250 | 3·453 | 155610 | 71439 | 197586 |
| Iran | Kohgiluyeh and Boyer-Ahmad | 0·794 | 0·630 | 2·293 | 0·387 | 0·995 | 6077 | 2962 | 7655 |
| Iran | Kurdistan | 4·771 | 3·931 | 14·487 | 2·172 | 6·078 | 90648 | 42281 | 115932 |
| Iran | Lorestan | 3·373 | 3·220 | 11·714 | 1·302 | 4·296 | 69116 | 26408 | 88205 |
| Iran | Markazi | 3·040 | 2·774 | 10·093 | 1·264 | 3·849 | 45495 | 18935 | 57324 |
| Iran | North Khorasan | 7·093 | 7·773 | 28·727 | 2·156 | 9·018 | 70233 | 20794 | 89347 |
| Iran | Qazvin | 6·030 | 5·173 | 19·078 | 2·581 | 7·762 | 94340 | 40910 | 122030 |
| Iran | Qom | 2·818 | 2·453 | 8·908 | 1·236 | 3·568 | 39736 | 17695 | 50448 |
| Iran | Razavi Khorasan | 4·667 | 6·127 | 21·899 | 1·132 | 5·636 | 370188 | 88233 | 450009 |
| Iran | Semnan | 5·038 | 4·475 | 16·401 | 2·129 | 6·411 | 47053 | 21035 | 60092 |
| Iran | Sistan and Baluchestan | 2·518 | 3·298 | 11·358 | 0·677 | 3·018 | 78175 | 19965 | 92831 |
| Iran | South Khorasan | 2·275 | 3·469 | 11·408 | 0·507 | 2·585 | 21400 | 4623 | 24413 |
| Iran | Tehran | 6·110 | 4·661 | 17·280 | 2·954 | 7·832 | 1088097 | 540881 | 1399973 |

| ADM0 | ADM1 | β-thalassaemia carrier prevalence Mean | SD | CI95% | Q25 | Q75 | Estimated carrier counts Mean | Q25 | Q75 |
|---|---|---|---|---|---|---|---|---|---|
| Iran | West Azerbaijan | 3·393 | 2·742 | 10·064 | 1·592 | 4·303 | 135863 | 65160 | 171715 |
| Iran | Yazd | 0·440 | 0·490 | 1·675 | 0·158 | 0·537 | 4945 | 1795 | 6162 |
| Iran | Zanjan | 4·676 | 4·086 | 14·942 | 1·983 | 6·016 | 59114 | 25099 | 76398 |
| Iraq | Al-Anbar | 3·997 | 4·169 | 14·920 | 1·433 | 5·011 | 144879 | 54762 | 184243 |
| Iraq | Al-Basrah | 4·776 | 3·129 | 11·743 | 2·618 | 6·059 | 179067 | 98880 | 226202 |
| Iraq | Al-Muthanna | 3·120 | 2·891 | 10·448 | 1·298 | 3·912 | 39963 | 16066 | 49634 |
| Iraq | Al-Qadisiyah | 4·629 | 4·591 | 16·590 | 1·768 | 5·807 | 76164 | 28731 | 94826 |
| Iraq | Al-Sulaimaniyah | 5·395 | 3·422 | 12·863 | 3·025 | 6·823 | 124449 | 70749 | 156510 |
| Iraq | An-Najaf | 3·285 | 3·486 | 12·449 | 1·195 | 4·058 | 74642 | 26922 | 91507 |
| Iraq | Babil | 5·342 | 5·298 | 19·194 | 2·019 | 6·732 | 237064 | 89322 | 298584 |
| Iraq | Baghdad | 6·009 | 5·637 | 20·342 | 2·387 | 7·713 | 408875 | 163945 | 527395 |
| Iraq | Dhi Qar | 5·076 | 4·089 | 15·120 | 2·359 | 6·474 | 132156 | 60971 | 168215 |
| Iraq | Diyala | 6·452 | 5·117 | 19·058 | 2·995 | 8·271 | 206359 | 91259 | 264791 |
| Iraq | Dohuk | 3·990 | 2·620 | 9·819 | 2·188 | 5·043 | 48511 | 26773 | 61416 |
| Iraq | Erbil | 5·528 | 3·562 | 13·383 | 3·061 | 7·009 | 146068 | 81849 | 184893 |
| Iraq | Karbala | 4·468 | 4·606 | 16·468 | 1·631 | 5·591 | 52604 | 19217 | 65738 |
| Iraq | Kirkuk | 6·432 | 4·398 | 16·516 | 3·393 | 8·191 | 130280 | 69792 | 165454 |
| Iraq | Maysan | 5·434 | 4·371 | 16·166 | 2·497 | 6·971 | 74410 | 35473 | 95421 |
| Iraq | Ninawa | 5·071 | 3·735 | 13·877 | 2·534 | 6·471 | 248309 | 130201 | 316086 |
| Iraq | Salah al-Din | 6·346 | 5·138 | 19·033 | 2·895 | 8·147 | 153774 | 69591 | 197919 |
| Iraq | Wasit | 5·955 | 5·435 | 19·961 | 2·393 | 7·640 | 103562 | 41099 | 131785 |
| Italy | Abruzzo | 6·959 | 7·400 | 27·224 | 2·206 | 8·914 | 82273 | 25489 | 104961 |
| Italy | Basilicata | 8·368 | 8·948 | 33·094 | 2·704 | 10·581 | 44398 | 14407 | 56069 |
| Italy | Calabria | 7·920 | 8·586 | 32·110 | 2·506 | 9·977 | 125230 | 39482 | 157335 |
| Italy | Campania | 8·255 | 8·866 | 32·769 | 2·620 | 10·496 | 444523 | 140975 | 566708 |
| Italy | Emilia-Romagna | 3·405 | 3·137 | 11·295 | 1·429 | 4·283 | 145009 | 61564 | 182309 |
| Italy | Friuli Venezia Giulia | 3·915 | 4·973 | 16·700 | 1·112 | 4·754 | 44239 | 12750 | 53288 |
| Italy | Lazio | 7·385 | 7·481 | 27·955 | 2·502 | 9·482 | 424713 | 145966 | 547476 |
| Italy | Liguria | 4·483 | 4·657 | 16·559 | 1·610 | 5·636 | 45122 | 16353 | 57015 |
| Italy | Lombardia | 3·512 | 3·825 | 13·563 | 1·214 | 4·347 | 357688 | 122129 | 441284 |
| Italy | Marche | 5·230 | 5·698 | 20·548 | 1·716 | 6·586 | 64636 | 20922 | 81344 |
| Italy | Molise | 7·580 | 8·092 | 29·438 | 2·442 | 9·657 | 21567 | 6908 | 27512 |
| Italy | Piemonte | 4·559 | 5·395 | 18·856 | 1·397 | 5·598 | 192081 | 58681 | 236070 |
| Italy | Puglia | 8·328 | 8·521 | 31·171 | 2·845 | 10·670 | 292857 | 101639 | 374895 |
| Italy | Sardegna | 14·761 | 8·596 | 32·679 | 8·484 | 19·044 | 218157 | 125348 | 281419 |
| Italy | Sicilia | 6·764 | 6·713 | 24·760 | 2·412 | 8·659 | 298507 | 105660 | 381602 |
| Italy | Toscana | 4·624 | 4·339 | 15·760 | 1·853 | 5·867 | 146413 | 60260 | 185870 |
| Italy | Trentino-Alto Adige | 3·513 | 4·183 | 14·586 | 1·061 | 4·330 | 37337 | 11717 | 45948 |
| Italy | Umbria | 5·636 | 5·847 | 21·253 | 1·941 | 7·146 | 48498 | 16724 | 61417 |
| Italy | Valle D'Aosta | 4·578 | 5·855 | 20·465 | 1·236 | 5·495 | 5561 | 1514 | 6647 |
| Italy | Veneto | 3·364 | 3·662 | 12·718 | 1·178 | 4·198 | 156759 | 57025 | 196225 |
| Jordan | Ajloun | 3·534 | 2·400 | 8·870 | 1·928 | 4·481 | 5655 | 3105 | 7136 |
| Jordan | Amman | 2·286 | 1·688 | 6·220 | 1·163 | 2·889 | 146611 | 76382 | 186347 |
| Jordan | Aqaba | 1·669 | 1·633 | 5·784 | 0·671 | 2·083 | 3368 | 1290 | 4138 |
| Jordan | Balqa | 3·323 | 2·292 | 8·544 | 1·782 | 4·198 | 14206 | 7615 | 17767 |
| Jordan | Irbid | 3·708 | 2·536 | 9·491 | 1·999 | 4·655 | 82961 | 44645 | 104240 |
| Jordan | Jerash | 3·259 | 2·203 | 8·230 | 1·762 | 4·117 | 7894 | 4314 | 9933 |
| Jordan | Karak | 2·195 | 1·793 | 6·502 | 1·038 | 2·769 | 8761 | 4146 | 11039 |
| Jordan | Ma'an | 1·310 | 1·225 | 4·323 | 0·556 | 1·636 | 2738 | 1158 | 3445 |
| Jordan | Madaba | 2·771 | 2·034 | 7·469 | 1·406 | 3·524 | 5946 | 3052 | 7624 |
| Jordan | Mafraq | 2·195 | 1·890 | 6·743 | 1·022 | 2·734 | 19811 | 10646 | 24966 |
| Jordan | Tafilah | 1·994 | 1·741 | 6·287 | 0·892 | 2·508 | 2400 | 1079 | 2980 |
| Jordan | Zarqa | 2·036 | 1·505 | 5·494 | 1·047 | 2·560 | 42694 | 22499 | 53635 |
| Malaysia | Johor | 4·860 | 5·949 | 20·164 | 1·424 | 6·016 | 208183 | 58793 | 257590 |
| Malaysia | Kedah | 5·519 | 4·870 | 17·988 | 2·307 | 7·077 | 127594 | 52729 | 163422 |
| Malaysia | Kelantan | 5·027 | 5·057 | 18·469 | 1·816 | 6·378 | 97782 | 34427 | 124625 |
| Malaysia | Kuala Lumpur | 4·102 | 3·441 | 12·415 | 1·843 | 5·207 | 94664 | 42591 | 120182 |

| | | β-thalassaemia carrier prevalence | | | | | Estimated carrier counts | | |
|---|---|---|---|---|---|---|---|---|---|
| **ADM0** | **ADM1** | **Mean** | **SD** | **CI95%** | **Q25** | **Q75** | **Mean** | **Q25** | **Q75** |
| Malaysia | Labuan | 11·722 | 9·159 | 33·989 | 5·279 | 15·418 | 5410 | 2437 | 7085 |
| Malaysia | Malacca | 4·299 | 4·339 | 15·306 | 1·624 | 5·412 | 43607 | 16351 | 54961 |
| Malaysia | Negeri Sembilan | 4·181 | 3·887 | 13·965 | 1·690 | 5·324 | 56387 | 23385 | 71516 |
| Malaysia | Pahang | 4·537 | 4·510 | 16·361 | 1·672 | 5·754 | 77035 | 27610 | 97926 |
| Malaysia | Penang | 5·575 | 5·307 | 19·307 | 2·149 | 7·111 | 94473 | 36280 | 120474 |
| Malaysia | Perak | 5·039 | 4·813 | 17·691 | 1·954 | 6·401 | 132880 | 50841 | 169058 |
| Malaysia | Perlis | 5·711 | 4·814 | 17·705 | 2·509 | 7·293 | 18210 | 7928 | 23406 |
| Malaysia | Putrajaya | 3·942 | 3·162 | 11·539 | 1·805 | 5·094 | 3432 | 1572 | 4435 |
| Malaysia | Sabah | 12·928 | 8·792 | 33·276 | 6·626 | 16·842 | 451446 | 228277 | 589040 |
| Malaysia | Sarawak | 8·532 | 9·794 | 36·348 | 2·403 | 10·753 | 192997 | 49982 | 240547 |
| Malaysia | Selangor | 4·241 | 3·692 | 13·537 | 1·826 | 5·402 | 314201 | 139093 | 399761 |
| Malaysia | Terengganu | 4·828 | 5·223 | 18·882 | 1·552 | 6·123 | 52423 | 16323 | 66678 |
| Malta | Attard | 4·055 | 3·380 | 12·262 | 1·852 | 5·160 | 10471 | 4737 | 13279 |
| Malta | Balzan | 4·091 | 3·508 | 12·661 | 1·838 | 5·141 | 7810 | 3502 | 9858 |
| Malta | Birgu | 3·961 | 3·325 | 12·405 | 1·874 | 4·915 | 3434 | 1625 | 4261 |
| Malta | Birkirkara | 4·116 | 3·486 | 12·859 | 1·855 | 5·257 | 10664 | 4797 | 13620 |
| Malta | Birzebbuga | 3·961 | 3·325 | 12·405 | 1·874 | 4·915 | 3434 | 1625 | 4261 |
| Malta | Bormla | 3·961 | 3·325 | 12·405 | 1·874 | 4·915 | 3434 | 1625 | 4261 |
| Malta | Dingli | 4·010 | 3·305 | 12·309 | 1·787 | 5·107 | 590 | 263 | 751 |
| Malta | Fgura | 3·961 | 3·325 | 12·405 | 1·874 | 4·915 | 3434 | 1625 | 4261 |
| Malta | Floriana | 4·053 | 3·362 | 12·525 | 1·855 | 5·189 | 11104 | 5069 | 14189 |
| Malta | Fontana | 4·276 | 3·514 | 13·671 | 1·959 | 5·391 | 1179 | 539 | 1477 |
| Malta | Ghajnsielem | 4·228 | 3·536 | 13·213 | 1·936 | 5·401 | 1693 | 775 | 2145 |
| Malta | Gharb | 4·261 | 3·360 | 13·131 | 1·961 | 5·431 | 260 | 120 | 331 |
| Malta | Gharghur | 4·091 | 3·508 | 12·661 | 1·838 | 5·141 | 7810 | 3502 | 9858 |
| Malta | Ghasri | 4·276 | 3·514 | 13·671 | 1·959 | 5·391 | 1179 | 539 | 1477 |
| Malta | Ghaxaq | 3·961 | 3·325 | 12·405 | 1·874 | 4·915 | 3434 | 1625 | 4261 |
| Malta | Gudja | 4·062 | 3·383 | 12·831 | 1·882 | 5·202 | 6288 | 2920 | 8023 |
| Malta | Gzira | 4·033 | 3·320 | 11·913 | 1·800 | 5·164 | 4816 | 2149 | 6166 |
| Malta | Hamrun | 4·099 | 3·380 | 12·585 | 1·845 | 5·326 | 7670 | 3444 | 9928 |
| Malta | Iklin | 4·091 | 3·508 | 12·661 | 1·838 | 5·141 | 7810 | 3502 | 9858 |
| Malta | Isla | 3·961 | 3·325 | 12·405 | 1·874 | 4·915 | 3434 | 1625 | 4261 |
| Malta | Kalkara | 3·961 | 3·325 | 12·405 | 1·874 | 4·915 | 3434 | 1625 | 4261 |
| Malta | Kercem | 4·261 | 3·360 | 13·131 | 1·961 | 5·431 | 260 | 120 | 331 |
| Malta | Kirkop | 4·164 | 3·441 | 13·256 | 1·890 | 5·489 | 2854 | 1295 | 3762 |
| Malta | Lija | 4·149 | 3·697 | 13·408 | 1·876 | 5·117 | 2994 | 1353 | 3692 |
| Malta | Luqa | 4·164 | 3·441 | 13·256 | 1·890 | 5·489 | 2854 | 1295 | 3762 |
| Malta | Marsa | 4·164 | 3·441 | 13·256 | 1·890 | 5·489 | 2854 | 1295 | 3762 |
| Malta | Marsaskala | 4·231 | 3·645 | 12·525 | 1·891 | 5·417 | 915 | 409 | 1171 |
| Malta | Marsaxlokk | 4·096 | 3·485 | 12·465 | 1·882 | 5·166 | 4349 | 2034 | 5432 |
| Malta | Mdina | 4·010 | 3·305 | 12·309 | 1·787 | 5·107 | 590 | 263 | 751 |
| Malta | Mellieha | 4·158 | 3·324 | 11·974 | 1·876 | 5·427 | 334 | 151 | 436 |
| Malta | Mgarr | 4·211 | 3·549 | 13·443 | 1·952 | 5·365 | 235 | 109 | 299 |
| Malta | Mosta | 4·111 | 3·488 | 12·746 | 1·908 | 5·169 | 3943 | 1805 | 4909 |
| Malta | Mqabba | 4·087 | 3·369 | 12·426 | 1·875 | 5·341 | 3976 | 1815 | 5215 |
| Malta | Msida | 4·099 | 3·380 | 12·585 | 1·845 | 5·326 | 7670 | 3444 | 9928 |
| Malta | Mtarfa | 4·042 | 3·292 | 12·197 | 1·863 | 5·163 | 1539 | 715 | 1968 |
| Malta | Munxar | 4·276 | 3·514 | 13·671 | 1·959 | 5·391 | 1179 | 539 | 1477 |
| Malta | Nadur | 4·234 | 3·617 | 13·455 | 1·975 | 5·424 | 440 | 205 | 563 |
| Malta | Naxxar | 4·149 | 3·697 | 13·408 | 1·876 | 5·117 | 2994 | 1353 | 3692 |
| Malta | Paola | 4·062 | 3·383 | 12·831 | 1·882 | 5·202 | 6288 | 2920 | 8023 |
| Malta | Pembroke | 4·033 | 3·320 | 11·913 | 1·800 | 5·164 | 4816 | 2149 | 6166 |
| Malta | Pieta | 4·099 | 3·380 | 12·585 | 1·845 | 5·326 | 7670 | 3444 | 9928 |
| Malta | Qala | 4·234 | 3·617 | 13·455 | 1·975 | 5·424 | 440 | 205 | 563 |
| Malta | Qormi | 4·069 | 3·353 | 12·255 | 1·850 | 5·282 | 8792 | 3964 | 11381 |
| Malta | Qrendi | 4·087 | 3·369 | 12·426 | 1·875 | 5·341 | 3976 | 1815 | 5215 |
| Malta | Rabat Gozo | 4·276 | 3·514 | 13·671 | 1·959 | 5·391 | 1179 | 539 | 1477 |

| | | β-thalassaemia carrier prevalence | | | | | Estimated carrier counts | | |
|---|---|---|---|---|---|---|---|---|---|
| ADM0 | ADM1 | Mean | SD | CI95% | Q25 | Q75 | Mean | Q25 | Q75 |
| Malta | Rabat Malta | 4·091 | 3·426 | 12·568 | 1·883 | 5·200 | 5890 | 2697 | 7412 |
| Malta | Safi | 4·062 | 3·383 | 12·831 | 1·882 | 5·202 | 6288 | 2920 | 8023 |
| Malta | Saint John | 4·033 | 3·320 | 11·913 | 1·800 | 5·164 | 4816 | 2149 | 6166 |
| Malta | Saint Julian's | 4·033 | 3·320 | 11·913 | 1·800 | 5·164 | 4816 | 2149 | 6166 |
| Malta | Saint Lawerence | 4·261 | 3·360 | 13·131 | 1·961 | 5·431 | 260 | 120 | 331 |
| Malta | Saint Lucia's | 4·062 | 3·383 | 12·831 | 1·882 | 5·202 | 6288 | 2920 | 8023 |
| Malta | Saint Paul's Bay | 4·073 | 3·280 | 12·085 | 1·940 | 5·220 | 949 | 452 | 1217 |
| Malta | Sannat | 4·276 | 3·514 | 13·671 | 1·959 | 5·391 | 1179 | 539 | 1477 |
| Malta | Santa Venera | 4·099 | 3·380 | 12·585 | 1·845 | 5·326 | 7670 | 3444 | 9928 |
| Malta | Siggiewi | 4·061 | 3·348 | 12·387 | 1·845 | 5·263 | 4566 | 2078 | 5966 |
| Malta | Sliema | 4·033 | 3·320 | 11·913 | 1·800 | 5·164 | 4816 | 2149 | 6166 |
| Malta | Swieqi | 4·033 | 3·320 | 11·913 | 1·800 | 5·164 | 4816 | 2149 | 6166 |
| Malta | Ta' Xbiex | 4·033 | 3·320 | 11·913 | 1·800 | 5·164 | 4816 | 2149 | 6166 |
| Malta | Tarxien | 4·062 | 3·383 | 12·831 | 1·882 | 5·202 | 6288 | 2920 | 8023 |
| Malta | Xaghra | 4·096 | 3·485 | 12·465 | 1·882 | 5·166 | 4349 | 2034 | 5432 |
| Malta | Xewkija | 4·291 | 3·667 | 14·210 | 1·957 | 5·351 | 919 | 419 | 1146 |
| Malta | Xghajra | 4·263 | 3·642 | 13·833 | 1·966 | 5·387 | 1359 | 624 | 1709 |
| Malta | Zabbar | 4·096 | 3·485 | 12·465 | 1·882 | 5·166 | 4349 | 2034 | 5432 |
| Malta | Zebbug Gozo | 4·276 | 3·514 | 13·671 | 1·959 | 5·391 | 1179 | 539 | 1477 |
| Malta | Zebbug Malta | 4·009 | 3·298 | 11·596 | 1·860 | 5·192 | 1122 | 520 | 1453 |
| Malta | Zejtun | 3·961 | 3·325 | 12·405 | 1·874 | 4·915 | 3434 | 1625 | 4261 |
| Malta | Zurrieq | 4·062 | 3·383 | 12·831 | 1·882 | 5·202 | 6288 | 2920 | 8023 |
| Myanmar [Burma] | Ayeyarwady | 7·936 | 6·760 | 25·183 | 3·362 | 10·275 | 479551 | 209505 | 620927 |
| Myanmar [Burma] | Bago | 6·607 | 5·881 | 21·822 | 2·726 | 8·460 | 331207 | 140073 | 423717 |
| Myanmar [Burma] | Chin | 2·203 | 2·751 | 9·430 | 0·664 | 2·651 | 10998 | 3323 | 13337 |
| Myanmar [Burma] | Kachin | 3·932 | 4·566 | 16·210 | 1·210 | 4·868 | 70943 | 21579 | 87980 |
| Myanmar [Burma] | Kayah | 5·366 | 5·896 | 21·383 | 1·765 | 6·693 | 17846 | 5655 | 21943 |
| Myanmar [Burma] | Kayin | 4·917 | 4·595 | 16·848 | 1·974 | 6·234 | 74938 | 31439 | 94911 |
| Myanmar [Burma] | Magway | 4·136 | 4·919 | 17·657 | 1·252 | 5·050 | 158504 | 47544 | 192581 |
| Myanmar [Burma] | Mandalay | 3·669 | 4·607 | 16·243 | 1·018 | 4·447 | 283245 | 77226 | 343434 |
| Myanmar [Burma] | Mon | 5·310 | 4·694 | 17·261 | 2·246 | 6·763 | 92742 | 39539 | 118413 |
| Myanmar [Burma] | Rakhine | 5·017 | 5·819 | 21·069 | 1·502 | 6·227 | 143206 | 41627 | 177251 |
| Myanmar [Burma] | Saigang | 2·364 | 2·659 | 9·210 | 0·822 | 2·895 | 120467 | 35838 | 144617 |
| Myanmar [Burma] | Shan | 3·964 | 5·109 | 18·134 | 0·988 | 4·841 | 269762 | 67563 | 328418 |
| Myanmar [Burma] | Tanitharyi | 3·313 | 3·836 | 13·630 | 1·020 | 4·096 | 39670 | 12333 | 49236 |
| Myanmar [Burma] | Yangon | 7·572 | 5·306 | 19·889 | 3·877 | 9·736 | 655622 | 347331 | 837461 |
| Oman | Ad Dakhiliyah | 2·742 | 1·998 | 7·389 | 1·407 | 3·467 | 15324 | 8260 | 19304 |
| Oman | Al Batinah | 2·475 | 1·747 | 6·479 | 1·302 | 3·127 | 32080 | 16595 | 40640 |
| Oman | Al Wusta | 3·057 | 2·658 | 9·681 | 1·352 | 3·862 | 1595 | 656 | 2014 |
| Oman | Ash Sharqiyah | 2·789 | 2·129 | 7·828 | 1·376 | 3·526 | 19232 | 9427 | 24340 |
| Oman | Az Zahirah | 2·711 | 2·036 | 7·482 | 1·363 | 3·424 | 13149 | 6722 | 16635 |
| Oman | Dhofar | 2·780 | 2·271 | 8·282 | 1·302 | 3·515 | 12653 | 6045 | 16145 |
| Oman | Muscat | 2·593 | 1·978 | 7·252 | 1·292 | 3·274 | 38067 | 19011 | 48340 |
| Pakistan | Azad Kashmir | 5·155 | 6·801 | 24·513 | 1·202 | 6·229 | 264802 | 63065 | 320339 |
| Pakistan | Balochistan | 5·370 | 6·763 | 24·544 | 1·384 | 6·535 | 811302 | 197360 | 981842 |
| Pakistan | Gilgit-Baltistan | 5·424 | 7·687 | 27·550 | 1·088 | 6·421 | 61144 | 12253 | 72211 |
| Pakistan | Islamabad Capital Territory | 5·318 | 7·165 | 26·158 | 1·202 | 6·350 | 191114 | 43087 | 229947 |
| Pakistan | Khyber Pakhtunkhwa | 5·618 | 7·817 | 28·439 | 1·138 | 6·678 | 2410735 | 484519 | 2827740 |
| Pakistan | Punjab | 5·486 | 6·968 | 25·467 | 1·370 | 6·673 | 6936268 | 1864625 | 8460740 |

| | | β-thalassaemia carrier prevalence | | | | | Estimated carrier counts | | |
|---|---|---|---|---|---|---|---|---|---|
| ADM0 | ADM1 | Mean | SD | CI95% | Q25 | Q75 | Mean | Q25 | Q75 |
| Pakistan | Sindh | 6·195 | 5·989 | 21·939 | 2·309 | 7·934 | 3675721 | 1511131 | 4701795 |
| Palestine | Gaza | 4·644 | 3·431 | 12·734 | 2·324 | 5·925 | 103682 | 51865 | 133026 |
| Palestine | West Bank | 3·951 | 2·829 | 10·486 | 2·052 | 4·987 | 119516 | 62070 | 150616 |
| Saudi Arabia | Al Bahah Region | 1·180 | 0·856 | 3·140 | 0·614 | 1·486 | 3478 | 1824 | 4403 |
| Saudi Arabia | Al Jawf Region | 0·383 | 0·312 | 1·125 | 0·183 | 0·479 | 3147 | 1511 | 3944 |
| Saudi Arabia | Al Madinah Region | 0·405 | 0·338 | 1·211 | 0·191 | 0·506 | 8306 | 4005 | 10463 |
| Saudi Arabia | Al-Qassim Region | 0·225 | 0·176 | 0·636 | 0·112 | 0·282 | 3294 | 1647 | 4165 |
| Saudi Arabia | 'Asir Region | 0·703 | 0·516 | 1·886 | 0·364 | 0·883 | 23025 | 12302 | 28863 |
| Saudi Arabia | Eastern Region | 3·772 | 3·111 | 11·448 | 1·730 | 4·790 | 223360 | 108284 | 284635 |
| Saudi Arabia | Hayel Region | 0·051 | 0·045 | 0·158 | 0·023 | 0·063 | 260 | 94 | 342 |
| Saudi Arabia | Jazan Region | 3·132 | 2·222 | 8·253 | 1·625 | 3·974 | 46294 | 24152 | 58613 |
| Saudi Arabia | Makkah Region | 1·281 | 0·991 | 3·615 | 0·634 | 1·613 | 131299 | 65843 | 165505 |
| Saudi Arabia | Najran Region | 0·388 | 0·404 | 1·355 | 0·156 | 0·475 | 2661 | 1038 | 3243 |
| Saudi Arabia | Northern Borders Region | 0·603 | 0·568 | 2·009 | 0·255 | 0·750 | 2631 | 1076 | 3254 |
| Saudi Arabia | Riyadh Region | 1·414 | 1·279 | 4·559 | 0·614 | 1·773 | 177609 | 83970 | 223871 |
| Saudi Arabia | Tabuk Region | 0·730 | 0·731 | 2·526 | 0·293 | 0·908 | 5603 | 2350 | 6994 |
| South Korea | Busan | 1·168 | 0·858 | 3·207 | 0·604 | 1·467 | 24740 | 12784 | 31072 |
| South Korea | Daegu | 1·263 | 1·013 | 3·675 | 0·606 | 1·586 | 25886 | 12570 | 32674 |
| South Korea | Daejeon | 1·231 | 1·042 | 3·700 | 0·574 | 1·539 | 17485 | 8159 | 21712 |
| South Korea | Gangwon | 1·937 | 2·363 | 8·037 | 0·608 | 2·348 | 29746 | 9321 | 35621 |
| South Korea | Gwangju | 1·109 | 1·024 | 3·643 | 0·475 | 1·391 | 3120 | 1336 | 3884 |
| South Korea | Gyeonggi | 1·729 | 2·104 | 6·932 | 0·569 | 2·093 | 247763 | 83157 | 301029 |
| South Korea | Incheon | 1·689 | 2·187 | 6·776 | 0·532 | 2·022 | 39676 | 12579 | 48135 |
| South Korea | Jeju | 1·155 | 1·417 | 4·849 | 0·366 | 1·387 | 6208 | 2011 | 7502 |
| South Korea | North Chungcheong | 1·405 | 1·312 | 4·644 | 0·593 | 1·761 | 24918 | 10579 | 31206 |
| South Korea | North Gyeongsang | 1·391 | 1·236 | 4·456 | 0·609 | 1·749 | 40121 | 18433 | 50662 |
| South Korea | North Jeolla | 1·123 | 0·945 | 3·421 | 0·521 | 1·410 | 19755 | 9329 | 24903 |
| South Korea | Sejong | 1·263 | 1·087 | 3·921 | 0·573 | 1·584 | 3193 | 1458 | 4031 |
| South Korea | Seoul | 1·710 | 2·090 | 6·902 | 0·559 | 2·071 | 140031 | 45504 | 168920 |
| South Korea | South Chungcheong | 1·272 | 1·198 | 4·206 | 0·540 | 1·584 | 27051 | 11406 | 33783 |
| South Korea | South Gyeongsang | 1·152 | 0·906 | 3·264 | 0·567 | 1·448 | 40461 | 20450 | 50917 |
| South Korea | South Jeolla | 1·095 | 1·063 | 3·736 | 0·452 | 1·359 | 28999 | 12105 | 35760 |
| South Korea | Ulsan | 1·248 | 0·968 | 3·529 | 0·616 | 1·572 | 11012 | 5376 | 13763 |
| Sri Lanka | Central Province | 6·298 | 5·201 | 19·202 | 2·772 | 8·156 | 176743 | 78555 | 228987 |
| Sri Lanka | Eastern Province | 6·379 | 6·050 | 22·281 | 2·391 | 8·272 | 98496 | 36351 | 127757 |
| Sri Lanka | North Central Province | 6·328 | 5·557 | 20·580 | 2·619 | 8·160 | 100388 | 42305 | 129475 |
| Sri Lanka | North Western Province | 6·238 | 4·918 | 18·255 | 2·877 | 8·059 | 170523 | 79704 | 220592 |
| Sri Lanka | Northern Province | 6·224 | 6·101 | 22·552 | 2·253 | 7·989 | 65767 | 22991 | 83601 |
| Sri Lanka | Sabaragamuwa Province | 6·360 | 5·640 | 20·773 | 2·620 | 8·206 | 141419 | 59131 | 182381 |
| Sri Lanka | Southern Province | 6·380 | 6·419 | 23·480 | 2·286 | 8·125 | 150843 | 54505 | 192854 |
| Sri Lanka | Uva Province | 6·350 | 5·977 | 22·038 | 2·418 | 8·171 | 101511 | 39622 | 131028 |
| Sri Lanka | Western Province | 6·333 | 5·541 | 20·420 | 2·639 | 8·167 | 369203 | 155658 | 475578 |
| Thailand | Amnat Charoen Province | 0·958 | 0·949 | 3·322 | 0·383 | 1·196 | 2064 | 831 | 2585 |
| Thailand | Ang Thong Province | 1·822 | 1·574 | 5·619 | 0·812 | 2·305 | 4193 | 1879 | 5314 |
| Thailand | Bangkok | 1·647 | 1·331 | 4·849 | 0·786 | 2·075 | 179315 | 85188 | 225770 |
| Thailand | Bueng Kan Province | 2·498 | 2·943 | 10·201 | 0·800 | 3·045 | 7935 | 2545 | 9651 |
| Thailand | Buri Ram Province | 1·187 | 1·035 | 3·736 | 0·530 | 1·493 | 12149 | 5409 | 15305 |
| Thailand | Chachoengsao Province | 1·698 | 1·305 | 4·780 | 0·842 | 2·137 | 13593 | 6657 | 17117 |
| Thailand | Chai Nat Province | 2·095 | 1·896 | 6·842 | 0·894 | 2·637 | 5092 | 2174 | 6398 |
| Thailand | Chaiyaphum Province | 1·811 | 1·560 | 5·639 | 0·817 | 2·275 | 14499 | 6538 | 18165 |
| Thailand | Chanthaburi Province | 1·335 | 1·107 | 3·997 | 0·622 | 1·685 | 5753 | 2640 | 7273 |
| Thailand | Chiang Mai Province | 4·514 | 5·051 | 17·769 | 1·498 | 5·599 | 82914 | 27097 | 102756 |
| Thailand | Chiang Rai Province | 4·159 | 5·218 | 17·714 | 1·197 | 5·068 | 46892 | 13369 | 57047 |
| Thailand | Chon Buri Province | 1·682 | 1·382 | 5·065 | 0·784 | 2·123 | 33665 | 15397 | 42757 |
| Thailand | Chumphon Province | 3·916 | 4·682 | 17·036 | 1·088 | 4·849 | 16101 | 4475 | 19950 |
| Thailand | Kalasin | 1·441 | 1·329 | 4·777 | 0·617 | 1·796 | 9662 | 4156 | 12031 |
| Thailand | Kamphaeng Phet Province | 2·752 | 2·449 | 8·890 | 1·203 | 3·453 | 22945 | 10136 | 28871 |

| ADM0 | ADM1 | β-thalassaemia carrier prevalence Mean | SD | CI95% | Q25 | Q75 | Estimated carrier counts Mean | Q25 | Q75 |
|---|---|---|---|---|---|---|---|---|---|
| Thailand | Kanchanaburi Province | 2·591 | 2·548 | 9·154 | 0·998 | 3·261 | 17492 | 6885 | 22037 |
| Thailand | Khon Kaen Province | 1·613 | 1·355 | 4·848 | 0·758 | 2·021 | 26909 | 12787 | 33761 |
| Thailand | Krabi Province | 5·517 | 5·931 | 21·643 | 1·802 | 6·970 | 17166 | 5570 | 21610 |
| Thailand | Lampang Province | 3·699 | 3·993 | 13·853 | 1·324 | 4·586 | 24350 | 8821 | 30221 |
| Thailand | Lamphun Province | 4·060 | 4·291 | 15·137 | 1·481 | 5·035 | 15481 | 5462 | 19254 |
| Thailand | Loei Province | 2·161 | 1·920 | 6·871 | 0·949 | 2·715 | 9584 | 4205 | 12002 |
| Thailand | Lopburi Province | 1·911 | 1·680 | 6·069 | 0·837 | 2·407 | 13935 | 6178 | 17542 |
| Thailand | Mae Hong Son Province | 5·067 | 5·446 | 19·696 | 1·709 | 6·356 | 10037 | 3372 | 12590 |
| Thailand | Maha Sarakham Province | 1·308 | 1·113 | 4·020 | 0·599 | 1·642 | 9004 | 4126 | 11307 |
| Thailand | Mukdahan Province | 1·322 | 1·417 | 4·834 | 0·491 | 1·632 | 4768 | 1756 | 5882 |
| Thailand | Nakhon Nayok Province | 1·801 | 1·454 | 5·346 | 0·855 | 2·270 | 4682 | 2217 | 5863 |
| Thailand | Nakhon Pathom Province | 1·706 | 1·474 | 5·321 | 0·759 | 2·151 | 18796 | 8548 | 23717 |
| Thailand | Nakhon Phanom Province | 1·905 | 2·216 | 7·727 | 0·631 | 2·316 | 9464 | 3098 | 11457 |
| Thailand | Nakhon Ratchasima Province | 1·617 | 1·398 | 5·042 | 0·726 | 2·034 | 40538 | 18090 | 50919 |
| Thailand | Nakhon Sawan Province | 2·239 | 1·985 | 7·194 | 0·979 | 2·812 | 19777 | 8732 | 24819 |
| Thailand | Nakhon Si Thammarat Province | 5·045 | 5·108 | 18·761 | 1·771 | 6·439 | 66130 | 23111 | 84716 |
| Thailand | Nan Province | 3·198 | 3·597 | 12·355 | 1·097 | 3·946 | 13353 | 4557 | 16441 |
| Thailand | Narathiwat Province | 5·168 | 4·901 | 17·919 | 1·975 | 6·615 | 31043 | 11850 | 39600 |
| Thailand | Nong Bua Lam Phu Province | 1·983 | 1·860 | 6·489 | 0·858 | 2·460 | 8848 | 3821 | 10961 |
| Thailand | Nong Khai Province | 2·373 | 2·572 | 9·005 | 0·851 | 2·926 | 9386 | 3363 | 11555 |
| Thailand | Nonthaburi Province | 1·642 | 1·359 | 4·965 | 0·764 | 2·062 | 33833 | 15823 | 42170 |
| Thailand | Pathum Thani Province | 1·683 | 1·361 | 4·973 | 0·800 | 2·113 | 40422 | 19165 | 50634 |
| Thailand | Pattani Province | 5·152 | 4·556 | 16·910 | 2·164 | 6·593 | 27650 | 11630 | 35295 |
| Thailand | Phangnga Province | 5·339 | 6·133 | 22·317 | 1·552 | 6·702 | 12881 | 3766 | 16224 |
| Thailand | Phatthalung Province | 5·421 | 4·580 | 16·888 | 2·350 | 6·999 | 22908 | 9941 | 29550 |
| Thailand | Phayao Province | 3·747 | 4·528 | 15·308 | 1·179 | 4·548 | 12764 | 3987 | 15477 |
| Thailand | Phetchabun Province | 1·977 | 1·640 | 5·961 | 0·921 | 2·489 | 16562 | 7721 | 20837 |
| Thailand | Phetchaburi Province | 2·103 | 2·123 | 7·644 | 0·771 | 2·657 | 8658 | 3381 | 10938 |
| Thailand | Phichit Province | 2·202 | 1·824 | 6·608 | 1·028 | 2·763 | 10644 | 4997 | 13311 |
| Thailand | Phitsanulok Province | 2·231 | 1·724 | 6·293 | 1·105 | 2·803 | 22442 | 11189 | 28102 |
| Thailand | Phra Nakhon Si Ayutthaya Province | 1·729 | 1·474 | 5·335 | 0·787 | 2·171 | 17690 | 8146 | 22295 |
| Thailand | Phrae Province | 3·181 | 3·233 | 11·325 | 1·218 | 3·980 | 10984 | 4286 | 13773 |
| Thailand | Phuket Province | 5·716 | 6·440 | 23·483 | 1·730 | 7·145 | 42574 | 12961 | 53393 |
| Thailand | Prachin Buri Province | 1·718 | 1·320 | 4·854 | 0·851 | 2·162 | 12668 | 6301 | 15982 |
| Thailand | Prachuap Khiri Khan Province | 2·599 | 2·899 | 10·431 | 0·833 | 3·231 | 10634 | 3490 | 13294 |
| Thailand | Ranong Province | 4·289 | 5·169 | 18·855 | 1·173 | 5·275 | 14406 | 3932 | 17344 |
| Thailand | Ratchaburi Province | 1·988 | 1·923 | 6·986 | 0·776 | 2·502 | 14476 | 5960 | 18272 |
| Thailand | Rayong Province | 1·618 | 1·388 | 4·997 | 0·728 | 2·049 | 18851 | 8298 | 23939 |
| Thailand | Roi Et Province | 1·099 | 0·979 | 3·526 | 0·478 | 1·385 | 9814 | 4291 | 12399 |
| Thailand | Sa Kaeo Province | 1·359 | 1·076 | 3·940 | 0·659 | 1·708 | 7990 | 3915 | 10049 |
| Thailand | Sakon Nakhon Province | 1·909 | 2·065 | 7·214 | 0·694 | 2·346 | 15488 | 5608 | 19055 |
| Thailand | Samut Prakan Province | 1·649 | 1·340 | 4·884 | 0·776 | 2·086 | 50912 | 24305 | 64438 |
| Thailand | Samut Sakhon Province | 1·648 | 1·409 | 5·238 | 0·730 | 2·091 | 24255 | 10915 | 30353 |
| Thailand | Samut Songkhram Province | 1·759 | 1·583 | 5·774 | 0·726 | 2·228 | 2696 | 1122 | 3411 |
| Thailand | Saraburi Province | 1·824 | 1·562 | 5·631 | 0·829 | 2·291 | 15116 | 6885 | 18933 |
| Thailand | Satun Province | 5·762 | 4·992 | 18·499 | 2·464 | 7·340 | 15690 | 6719 | 19947 |
| Thailand | Si Sa Ket Province | 0·633 | 0·533 | 1·923 | 0·292 | 0·797 | 4879 | 2243 | 6151 |
| Thailand | Sing Buri Province | 1·927 | 1·709 | 6·174 | 0·843 | 2·419 | 3431 | 1499 | 4284 |
| Thailand | Songkhla Province | 5·439 | 4·345 | 16·175 | 2·504 | 6·987 | 82098 | 38579 | 105488 |
| Thailand | Sukhothai Province | 2·879 | 2·495 | 8·987 | 1·288 | 3·632 | 17474 | 8002 | 22047 |
| Thailand | Suphan Buri Province | 1·974 | 1·789 | 6·416 | 0·838 | 2·494 | 14686 | 6296 | 18533 |
| Thailand | Surat Thani Province | 4·859 | 5·461 | 19·822 | 1·459 | 6·142 | 49353 | 14911 | 62184 |
| Thailand | Surin Province | 0·867 | 0·754 | 2·707 | 0·391 | 1·088 | 7892 | 3566 | 9886 |
| Thailand | Tak Province | 3·668 | 3·462 | 12·490 | 1·500 | 4·606 | 20607 | 8486 | 25960 |

| ADM0 | ADM1 | β-thalassaemia carrier prevalence Mean | SD | CI95% | Q25 | Q75 | Estimated carrier counts Mean | Q25 | Q75 |
|---|---|---|---|---|---|---|---|---|---|
| Thailand | Trang Province | 5·581 | 5·164 | 19·188 | 2·201 | 7·133 | 30407 | 11959 | 38889 |
| Thailand | Trat Province | 1·154 | 1·031 | 3·663 | 0·503 | 1·457 | 2333 | 1015 | 2960 |
| Thailand | Ubon Ratchathani Province | 0·691 | 0·654 | 2·313 | 0·287 | 0·867 | 11549 | 4824 | 14545 |
| Thailand | Udon Thani Province | 2·006 | 1·980 | 6·966 | 0·812 | 2·483 | 20923 | 8500 | 25934 |
| Thailand | Uthai Thani Province | 2·470 | 2·295 | 8·297 | 1·021 | 3·111 | 5939 | 2497 | 7491 |
| Thailand | Uttaradit Province | 2·622 | 2·337 | 8·409 | 1·141 | 3·308 | 10282 | 4526 | 12964 |
| Thailand | Yala Province | 5·272 | 4·661 | 17·165 | 2·192 | 6·766 | 21381 | 9017 | 27346 |
| Thailand | Yasothon Province | 0·993 | 0·935 | 3·323 | 0·410 | 1·245 | 4017 | 1650 | 5053 |
| Turkey | Adana | 2·648 | 2·080 | 7·645 | 1·278 | 3·349 | 74381 | 35456 | 94338 |
| Turkey | Adiyaman | 1·506 | 1·005 | 3·716 | 0·832 | 1·887 | 9780 | 5457 | 12192 |
| Turkey | Afyonkarahisar | 3·021 | 2·021 | 7·528 | 1·644 | 3·813 | 23486 | 12795 | 29435 |
| Turkey | Agri | 1·485 | 1·337 | 4·805 | 0·641 | 1·872 | 7784 | 3373 | 9887 |
| Turkey | Aksaray | 2·132 | 1·761 | 6·438 | 0·979 | 2·702 | 9489 | 4291 | 12004 |
| Turkey | Amasya | 1·456 | 1·834 | 6·028 | 0·443 | 1·766 | 5103 | 1534 | 6237 |
| Turkey | Ankara | 1·934 | 1·864 | 6·515 | 0·797 | 2·417 | 112777 | 44522 | 141005 |
| Turkey | Antalya | 5·407 | 3·593 | 13·466 | 2·928 | 6·868 | 136381 | 74352 | 172719 |
| Turkey | Ardahan | 1·216 | 1·265 | 4·259 | 0·481 | 1·506 | 1115 | 452 | 1395 |
| Turkey | Artvin | 1·043 | 0·970 | 3·421 | 0·444 | 1·302 | 1430 | 591 | 1780 |
| Turkey | Aydin | 3·838 | 2·481 | 9·259 | 2·142 | 4·835 | 44255 | 24845 | 55800 |
| Turkey | Balikesir | 2·754 | 1·960 | 7·224 | 1·451 | 3·464 | 31135 | 16425 | 39076 |
| Turkey | Bartin | 1·793 | 2·316 | 7·319 | 0·550 | 2·181 | 3497 | 1065 | 4275 |
| Turkey | Batman | 2·167 | 1·822 | 6·596 | 0·996 | 2·733 | 14241 | 6538 | 17943 |
| Turkey | Bayburt | 0·962 | 0·886 | 3·129 | 0·411 | 1·205 | 816 | 334 | 1009 |
| Turkey | Bilecik | 1·986 | 1·513 | 5·541 | 0·989 | 2·499 | 4792 | 2429 | 6048 |
| Turkey | Bingol | 1·218 | 1·055 | 3·808 | 0·550 | 1·527 | 3641 | 1627 | 4582 |
| Turkey | Bitlis | 1·884 | 1·601 | 5·780 | 0·864 | 2·369 | 6173 | 2830 | 7776 |
| Turkey | Bolu | 1·756 | 1·745 | 5·952 | 0·706 | 2·204 | 5683 | 2196 | 7069 |
| Turkey | Burdur | 4·148 | 2·743 | 10·229 | 2·269 | 5·249 | 11394 | 6206 | 14482 |
| Turkey | Bursa | 2·161 | 1·624 | 5·966 | 1·090 | 2·722 | 67175 | 33735 | 84888 |
| Turkey | Canakkale | 2·890 | 2·068 | 7·660 | 1·510 | 3·637 | 14719 | 7696 | 18432 |
| Turkey | Cankiri | 1·752 | 2·039 | 6·894 | 0·568 | 2·179 | 3557 | 1146 | 4420 |
| Turkey | Corum | 1·606 | 1·926 | 6·581 | 0·495 | 1·977 | 8490 | 2612 | 10514 |
| Turkey | Denizli | 3·610 | 2·321 | 8·666 | 2·020 | 4·549 | 38599 | 21867 | 48434 |
| Turkey | Diyarbakir | 1·738 | 1·426 | 5·142 | 0·820 | 2·189 | 33891 | 16215 | 42837 |
| Turkey | Duzce | 1·644 | 1·594 | 5·442 | 0·675 | 2·064 | 6303 | 2622 | 7805 |
| Turkey | Edirne | 3·484 | 2·924 | 10·708 | 1·574 | 4·425 | 14953 | 6604 | 19028 |
| Turkey | Elazig | 1·219 | 1·009 | 3·643 | 0·572 | 1·534 | 7227 | 3287 | 9009 |
| Turkey | Erzincan | 1·028 | 0·962 | 3·436 | 0·431 | 1·285 | 2459 | 991 | 3091 |
| Turkey | Erzurum | 0·995 | 0·799 | 2·883 | 0·481 | 1·249 | 7481 | 3679 | 9408 |
| Turkey | Eskisehir | 2·245 | 1·746 | 6·387 | 1·101 | 2·832 | 20495 | 10193 | 25925 |
| Turkey | Gaziantep | 2·251 | 1·539 | 5·690 | 1·215 | 2·834 | 50281 | 27100 | 63512 |
| Turkey | Giresun | 1·142 | 1·347 | 4·453 | 0·386 | 1·397 | 3643 | 1169 | 4432 |
| Turkey | Gumushane | 1·032 | 1·058 | 3·679 | 0·393 | 1·289 | 1566 | 570 | 1968 |
| Turkey | Hakkari | 3·719 | 2·670 | 9·933 | 1·913 | 4·718 | 10543 | 5354 | 13356 |
| Turkey | Hatay | 3·803 | 3·048 | 11·234 | 1·786 | 4·833 | 65345 | 30579 | 82978 |
| Turkey | Igdir | 1·639 | 1·656 | 5·784 | 0·634 | 2·052 | 3301 | 1275 | 4108 |
| Turkey | Isparta | 3·647 | 2·487 | 9·290 | 1·951 | 4·621 | 16675 | 8774 | 21195 |
| Turkey | Istanbul | 1·885 | 1·616 | 5·893 | 0·847 | 2·373 | 261413 | 121588 | 329149 |
| Turkey | Izmir | 4·068 | 2·647 | 9·909 | 2·248 | 5·138 | 169205 | 94025 | 213005 |
| Turkey | Kahramanmaras | 1·630 | 1·116 | 4·139 | 0·882 | 2·051 | 20869 | 11437 | 26084 |
| Turkey | Karabuk | 1·777 | 2·156 | 6·926 | 0·580 | 2·195 | 4652 | 1493 | 5689 |
| Turkey | Karaman | 4·404 | 3·119 | 11·614 | 2·284 | 5·594 | 11670 | 6117 | 14864 |
| Turkey | Kars | 1·246 | 1·184 | 4·158 | 0·522 | 1·555 | 3509 | 1450 | 4394 |
| Turkey | Kastamonu | 1·771 | 2·259 | 7·679 | 0·505 | 2·138 | 6587 | 1836 | 7932 |
| Turkey | Kayseri | 1·554 | 1·342 | 4·861 | 0·697 | 1·955 | 23117 | 10139 | 29464 |
| Turkey | Kilis | 2·673 | 1·933 | 7·146 | 1·383 | 3·385 | 4376 | 2244 | 5510 |
| Turkey | Kirikkale | 1·770 | 1·849 | 6·492 | 0·643 | 2·222 | 5080 | 1833 | 6400 |

| | | β-thalassaemia carrier prevalence | | | | | Estimated carrier counts | | |
|---|---|---|---|---|---|---|---|---|---|
| **ADM0** | **ADM1** | **Mean** | **SD** | **CI95%** | **Q25** | **Q75** | **Mean** | **Q25** | **Q75** |
| Turkey | Kirklareli | 2·890 | 2·691 | 9·697 | 1·195 | 3·641 | 10508 | 4487 | 13232 |
| Turkey | Kirsehir | 1·785 | 1·766 | 6·259 | 0·684 | 2·254 | 4544 | 1760 | 5689 |
| Turkey | Kocaeli | 1·613 | 1·292 | 4·710 | 0·778 | 2·025 | 26338 | 12762 | 33042 |
| Turkey | Konya | 2·974 | 2·125 | 7·857 | 1·548 | 3·760 | 69936 | 37075 | 88572 |
| Turkey | Kutahya | 2·741 | 1·861 | 6·914 | 1·482 | 3·457 | 16048 | 8623 | 20147 |
| Turkey | Malatya | 1·235 | 0·927 | 3·388 | 0·629 | 1·551 | 10329 | 5401 | 13086 |
| Turkey | Manisa | 3·452 | 2·217 | 8·289 | 1·935 | 4·345 | 54931 | 30939 | 69013 |
| Turkey | Mardin | 2·707 | 2·114 | 7·761 | 1·318 | 3·420 | 26115 | 12757 | 32873 |
| Turkey | Mersin | 5·509 | 4·003 | 14·954 | 2·780 | 7·036 | 83847 | 40636 | 107177 |
| Turkey | Mugla | 4·176 | 2·815 | 10·495 | 2·252 | 5·289 | 43546 | 23183 | 55261 |
| Turkey | Mus | 1·386 | 1·159 | 4·175 | 0·648 | 1·744 | 5926 | 2746 | 7464 |
| Turkey | Nevsehir | 1·774 | 1·615 | 5·849 | 0·740 | 2·253 | 5768 | 2455 | 7350 |
| Turkey | Nigde | 2·344 | 1·921 | 6·966 | 1·093 | 2·966 | 8451 | 3885 | 10792 |
| Turkey | Ordu | 1·292 | 1·665 | 5·516 | 0·390 | 1·535 | 8119 | 2402 | 9753 |
| Turkey | Osmaniye | 2·307 | 1·676 | 6·161 | 1·198 | 2·903 | 14163 | 7236 | 17677 |
| Turkey | Rize | 0·978 | 0·906 | 3·216 | 0·419 | 1·223 | 1770 | 719 | 2213 |
| Turkey | Sakarya | 1·618 | 1·360 | 4·907 | 0·753 | 2·033 | 17307 | 8179 | 21644 |
| Turkey | Samsun | 1·497 | 1·948 | 6·639 | 0·426 | 1·781 | 18298 | 5063 | 21555 |
| Turkey | Sanliurfa | 2·330 | 1·618 | 5·988 | 1·245 | 2·934 | 55362 | 29982 | 69614 |
| Turkey | Siirt | 2·416 | 1·922 | 6·992 | 1·154 | 3·063 | 7944 | 3726 | 10155 |
| Turkey | Sinop | 1·700 | 2·254 | 7·790 | 0·467 | 2·005 | 2897 | 760 | 3421 |
| Turkey | Sirnak | 3·103 | 2·255 | 8·383 | 1·588 | 3·933 | 18332 | 9483 | 23143 |
| Turkey | Sivas | 1·215 | 1·249 | 4·275 | 0·480 | 1·500 | 8012 | 2965 | 9849 |
| Turkey | Tekirdag | 2·563 | 2·144 | 7·793 | 1·178 | 3·225 | 26852 | 12057 | 34038 |
| Turkey | Tokat | 1·335 | 1·652 | 5·421 | 0·429 | 1·611 | 8178 | 2625 | 9807 |
| Turkey | Trabzon | 1·040 | 1·064 | 3·759 | 0·396 | 1·293 | 6873 | 2563 | 8537 |
| Turkey | Tunceli | 1·084 | 0·960 | 3·466 | 0·478 | 1·361 | 952 | 422 | 1197 |
| Turkey | Usak | 3·217 | 2·061 | 7·705 | 1·812 | 4·046 | 12160 | 6873 | 15346 |
| Turkey | Van | 2·399 | 2·021 | 7·360 | 1·084 | 3·046 | 24722 | 10973 | 31238 |
| Turkey | Yalova | 1·816 | 1·450 | 5·274 | 0·876 | 2·287 | 4084 | 1958 | 5186 |
| Turkey | Yozgat | 1·517 | 1·625 | 5·692 | 0·541 | 1·888 | 6534 | 2297 | 8208 |
| Turkey | Zonguldak | 1·703 | 1·950 | 6·243 | 0·606 | 2·113 | 7768 | 2730 | 9678 |
| United Arab Emirates | Abu Dhabi | 3·442 | 2·899 | 10·664 | 1·543 | 4·376 | 101558 | 49147 | 128936 |
| United Arab Emirates | Ajman | 1·881 | 1·558 | 5·424 | 0·898 | 2·362 | 8419 | 3906 | 10534 |
| United Arab Emirates | Dubai | 2·105 | 1·656 | 6·045 | 1·026 | 2·649 | 70649 | 34353 | 89098 |
| United Arab Emirates | Fujairah | 1·715 | 1·406 | 5·188 | 0·803 | 2·160 | 6257 | 2915 | 7901 |
| United Arab Emirates | Ras al-Khaimah | 1·648 | 1·354 | 4·936 | 0·774 | 2·073 | 8461 | 3894 | 10561 |
| United Arab Emirates | Sharjah | 1·881 | 1·519 | 5·548 | 0·895 | 2·380 | 34807 | 16481 | 43683 |
| United Arab Emirates | Umm al-Quwain | 1·651 | 1·429 | 5·079 | 0·762 | 2·057 | 2510 | 1140 | 3124 |

ADM0: administrative level 0. ADM1: administrative level 1. SD: standard deviation. CI95%: 95% credible interval. Q25: 25th percentile. Q75: 75th percentile. SD and CI95% are measures of prediction uncertainty.

**Supplementary Table S15. Subnational areal predictions of β-thalassaemia carrier prevalence and estimated carrier count at administrative level 2 of Albania.**

| | | β-thalassaemia carrier prevalence | | | | | Estimated carrier counts | | |
|---|---|---|---|---|---|---|---|---|---|
| **ADM1** | **ADM2** | **Mean** | **SD** | **CI95%** | **Q25** | **Q75** | **Mean** | **Q25** | **Q75** |
| Berat | Berat | 10·088 | 7·862 | 29·329 | 4·625 | 13·184 | 6875 | 3152 | 9002 |
| Berat | Kucove | 9·993 | 7·572 | 29·289 | 4·553 | 13·296 | 2540 | 1160 | 3415 |
| Berat | Skrapar | 10·270 | 8·053 | 29·987 | 4·638 | 13·444 | 755 | 337 | 995 |
| Diber | Bulqize | 10·264 | 7·491 | 27·890 | 4·951 | 13·428 | 1986 | 959 | 2593 |
| Diber | Diber | 10·276 | 7·772 | 29·202 | 4·783 | 13·502 | 4467 | 2094 | 5847 |
| Diber | Mat | 9·895 | 7·375 | 27·468 | 4·699 | 12·886 | 3377 | 1609 | 4384 |
| Durres | Durres | 9·684 | 7·492 | 28·606 | 4·477 | 12·569 | 23839 | 11008 | 30695 |

| | | β-thalassaemia carrier prevalence | | | | | Estimated carrier counts | | |
|---|---|---|---|---|---|---|---|---|---|
| ADM1 | ADM2 | Mean | SD | CI95% | Q25 | Q75 | Mean | Q25 | Q75 |
| Durres | Kruje | 9·706 | 7·293 | 27·654 | 4·590 | 12·540 | 5906 | 2810 | 7639 |
| Elbasan | Elbasan | 10·049 | 7·353 | 27·685 | 4·871 | 13·103 | 16936 | 8261 | 21977 |
| Elbasan | Gramsh | 10·321 | 7·709 | 29·002 | 4·864 | 13·528 | 1492 | 705 | 1952 |
| Elbasan | Librazhd | 10·427 | 7·570 | 28·851 | 5·053 | 13·622 | 3086 | 1504 | 4021 |
| Elbasan | Peqin | 9·829 | 7·412 | 27·755 | 4·680 | 12·790 | 2090 | 990 | 2721 |
| Fier | Fier | 9·790 | 7·965 | 29·613 | 4·321 | 12·731 | 14851 | 6566 | 19270 |
| Fier | Lushnje | 9·767 | 7·602 | 28·852 | 4·480 | 12·674 | 9847 | 4588 | 12760 |
| Fier | Mallakaster | 9·995 | 8·027 | 29·994 | 4·380 | 13·179 | 1635 | 720 | 2147 |
| Gjirokaster | Gjirokaster | 9·996 | 8·276 | 31·048 | 4·252 | 13·068 | 2993 | 1261 | 3921 |
| Gjirokaster | Permet | 10·192 | 8·271 | 31·094 | 4·433 | 13·306 | 792 | 346 | 1027 |
| Gjirokaster | Tepelene | 9·986 | 8·076 | 30·575 | 4·396 | 12·960 | 936 | 410 | 1220 |
| Korce | Devoll | 10·903 | 8·163 | 30·698 | 5·099 | 14·369 | 2857 | 1326 | 3775 |
| Korce | Kolonje | 10·426 | 8·420 | 31·892 | 4·557 | 13·614 | 933 | 404 | 1220 |
| Korce | Kucove | 10·602 | 8·034 | 30·346 | 4·942 | 13·885 | 11323 | 5280 | 14913 |
| Korce | Pogradec | 10·562 | 7·755 | 29·303 | 5·049 | 13·884 | 2359 | 1133 | 3130 |
| Kukes | Has | 10·284 | 8·480 | 32·086 | 4·365 | 13·537 | 2619 | 1162 | 3429 |
| Kukes | Kukes | 10·294 | 8·177 | 30·732 | 4·540 | 13·444 | 2191 | 973 | 2877 |
| Kukes | Tropoje | 9·901 | 8·499 | 32·159 | 4·014 | 12·985 | 2088 | 845 | 2757 |
| Lezhe | Kurbin | 9·699 | 7·443 | 28·023 | 4·483 | 12·772 | 4327 | 1984 | 5689 |
| Lezhe | Lezhe | 9·658 | 7·652 | 28·774 | 4·314 | 12·644 | 5335 | 2379 | 7006 |
| Lezhe | Mirdite | 9·836 | 7·621 | 28·543 | 4·498 | 12·869 | 1413 | 646 | 1862 |
| Shkoder | Malesi e Madhe | 9·478 | 8·373 | 31·578 | 3·763 | 12·315 | 3381 | 1387 | 4407 |
| Shkoder | Puke | 9·858 | 7·950 | 29·729 | 4·290 | 12·903 | 924 | 403 | 1215 |
| Shkoder | Shkoder | 9·517 | 7·904 | 29·535 | 4·031 | 12·442 | 12783 | 5359 | 16536 |
| Tirane | Kavaje | 9·640 | 7·416 | 28·116 | 4·461 | 12·491 | 10984 | 5073 | 14240 |
| Tirane | Tirane | 9·855 | 7·189 | 26·872 | 4·793 | 12·839 | 82557 | 39939 | 107033 |
| Vlore | Delvine | 9·870 | 8·288 | 30·707 | 4·120 | 12·940 | 901 | 377 | 1186 |
| Vlore | Sarande | 9·875 | 8·477 | 31·539 | 4·073 | 12·911 | 3262 | 1373 | 4271 |
| Vlore | Vlore | 9·818 | 8·242 | 30·957 | 4·121 | 12·866 | 5766 | 2411 | 7595 |

ADM1: administrative level 1. ADM2: administrative level 2. SD: standard deviation. CI95%: 95% credible interval. Q25: 25th percentile. Q75: 75th percentile. SD and CI95% are measures of prediction uncertainty.

**Supplementary Table S16. Subnational areal predictions of β-thalassaemia carrier prevalence and estimated carrier count at administrative level 2 of Bangladesh.**

| | | β-thalassaemia carrier prevalence | | | | | Estimated carrier counts | | |
|---|---|---|---|---|---|---|---|---|---|
| ADM1 | ADM2 | Mean | SD | CI95% | Q25 | Q75 | Mean | Q25 | Q75 |
| Barisal | Barguna | 4·492 | 4·390 | 15·922 | 1·726 | 5·680 | 42510 | 16380 | 53781 |
| Barisal | Barisal | 3·924 | 3·706 | 13·123 | 1·587 | 4·965 | 97404 | 39600 | 123090 |
| Barisal | Bhola | 3·790 | 3·954 | 14·170 | 1·356 | 4·783 | 66960 | 23970 | 84465 |
| Barisal | Jhalokati | 4·181 | 3·888 | 14·112 | 1·700 | 5·287 | 29297 | 11874 | 36979 |
| Barisal | Patuakhali | 4·217 | 4·257 | 15·312 | 1·557 | 5·333 | 74687 | 27957 | 94567 |
| Barisal | Pirojpur | 4·416 | 4·004 | 14·576 | 1·837 | 5·612 | 47690 | 19753 | 60680 |
| Chittagong | Bandarban | 2·937 | 3·560 | 12·449 | 0·872 | 3·595 | 21227 | 6279 | 25941 |
| Chittagong | Brahamanbaria | 2·892 | 2·728 | 9·862 | 1·177 | 3·639 | 101937 | 41471 | 128044 |
| Chittagong | Chandpur | 3·266 | 3·118 | 11·142 | 1·297 | 4·129 | 85349 | 33929 | 107942 |
| Chittagong | Chittagong | 2·742 | 3·250 | 11·202 | 0·890 | 3·326 | 259024 | 83587 | 314182 |
| Chittagong | Comilla | 2·983 | 2·984 | 10·565 | 1·147 | 3·740 | 192185 | 73873 | 240915 |
| Chittagong | Cox's Bazar | 3·446 | 4·199 | 14·724 | 0·994 | 4·205 | 94663 | 27304 | 115547 |
| Chittagong | Feni | 2·726 | 2·920 | 10·264 | 0·989 | 3·395 | 51490 | 18704 | 64134 |
| Chittagong | Khagrachhari | 2·233 | 2·515 | 8·504 | 0·799 | 2·725 | 18891 | 6654 | 23074 |
| Chittagong | Lakshmipur | 3·314 | 3·322 | 11·904 | 1·262 | 4·149 | 65643 | 25107 | 82418 |
| Chittagong | Noakhali | 3·148 | 3·307 | 11·684 | 1·139 | 3·928 | 115615 | 42405 | 144491 |
| Chittagong | Rangamati | 2·205 | 2·615 | 8·819 | 0·737 | 2·669 | 16211 | 5412 | 19546 |
| Dhaka | Dhaka | 3·930 | 3·526 | 12·759 | 1·639 | 5·030 | 591164 | 246260 | 757138 |
| Dhaka | Faridpur | 4·456 | 3·834 | 14·025 | 1·943 | 5·683 | 100678 | 43759 | 128308 |
| Dhaka | Gazipur | 3·723 | 3·418 | 12·195 | 1·549 | 4·728 | 239022 | 99376 | 303844 |
| Dhaka | Gopalganj | 4·401 | 3·871 | 14·058 | 1·879 | 5·644 | 63381 | 27064 | 81319 |
| Dhaka | Kishoreganj | 3·190 | 2·936 | 10·599 | 1·332 | 4·030 | 117709 | 48537 | 148709 |

| ADM1 | ADM2 | β-thalassaemia carrier prevalence<br>Mean | SD | CI95% | Q25 | Q75 | Estimated carrier counts<br>Mean | Q25 | Q75 |
|---|---|---|---|---|---|---|---|---|---|
| Dhaka | Madaripur | 4·046 | 3·666 | 13·216 | 1·696 | 5·142 | 57963 | 24367 | 73931 |
| Dhaka | Manikganj | 4·344 | 3·772 | 13·790 | 1·865 | 5·567 | 71270 | 30374 | 91336 |
| Dhaka | Munshiganj | 3·690 | 3·372 | 12·185 | 1·518 | 4·703 | 65514 | 27001 | 83460 |
| Dhaka | Narayanganj | 3·475 | 3·190 | 11·499 | 1·439 | 4·389 | 165416 | 68721 | 209854 |
| Dhaka | Narsingdi | 3·280 | 3·073 | 10·972 | 1·353 | 4·147 | 96209 | 39588 | 121910 |
| Dhaka | Rajbari | 4·823 | 4·031 | 14·771 | 2·178 | 6·114 | 60296 | 27126 | 76574 |
| Dhaka | Shariatpur | 3·750 | 3·492 | 12·583 | 1·519 | 4·748 | 47815 | 19389 | 60617 |
| Dhaka | Tangail | 4·416 | 3·856 | 14·149 | 1·902 | 5·618 | 194883 | 83711 | 247702 |
| Khulna | Bagerhat | 4·725 | 4·149 | 15·246 | 2·028 | 6·011 | 79912 | 34918 | 101876 |
| Khulna | Chuadanga | 5·826 | 4·219 | 15·741 | 2·936 | 7·479 | 76779 | 38655 | 98569 |
| Khulna | Jessore | 5·205 | 3·905 | 14·630 | 2·573 | 6·633 | 174084 | 86231 | 222017 |
| Khulna | Jhenaidah | 5·418 | 4·109 | 15·254 | 2·659 | 6·916 | 113121 | 55491 | 144496 |
| Khulna | Khulna | 4·935 | 3·930 | 14·501 | 2·311 | 6·306 | 129582 | 61071 | 165272 |
| Khulna | Kushtia | 5·630 | 4·300 | 15·967 | 2·721 | 7·230 | 128293 | 61960 | 164523 |
| Khulna | Magura | 5·003 | 4·144 | 15·265 | 2·288 | 6·369 | 54767 | 24982 | 69647 |
| Khulna | Meherpur | 6·084 | 4·373 | 16·331 | 3·093 | 7·796 | 44304 | 22584 | 56832 |
| Khulna | Narail | 4·718 | 3·904 | 14·404 | 2·136 | 6·008 | 41900 | 18941 | 53457 |
| Khulna | Satkhira | 5·293 | 3·904 | 14·551 | 2·640 | 6·763 | 118719 | 59953 | 151722 |
| Mymensingh | Jamalpur | 4·924 | 4·247 | 15·641 | 2·144 | 6·265 | 126034 | 54838 | 160460 |
| Mymensingh | Mymensingh | 4·042 | 3·661 | 13·208 | 1·716 | 5·103 | 257276 | 109113 | 324881 |
| Mymensingh | Netrakona | 3·630 | 3·268 | 11·873 | 1·548 | 4·577 | 87343 | 37160 | 110263 |
| Mymensingh | Sherpur | 4·704 | 4·118 | 15·191 | 2·041 | 5·957 | 74235 | 32308 | 93896 |
| Rajshani | Bogra | 5·441 | 4·529 | 16·625 | 2·433 | 6·956 | 222744 | 99765 | 285025 |
| Rajshani | Joypurhat | 5·835 | 4·789 | 17·723 | 2·644 | 7·462 | 60523 | 27428 | 77470 |
| Rajshani | Naogaon | 6·172 | 4·960 | 18·382 | 2·839 | 7·913 | 172037 | 79085 | 220729 |
| Rajshani | Natore | 5·664 | 4·511 | 16·712 | 2·625 | 7·255 | 108446 | 50251 | 138747 |
| Rajshani | Nawabganj | 6·623 | 4·974 | 18·667 | 3·228 | 8·492 | 126581 | 62232 | 161966 |
| Rajshani | Pabna | 5·147 | 4·177 | 15·416 | 2·365 | 6·562 | 155579 | 71617 | 198819 |
| Rajshani | Rajshahi | 6·185 | 4·748 | 17·663 | 2·961 | 7·932 | 190033 | 90517 | 243838 |
| Rajshani | Sirajganj | 4·987 | 4·205 | 15·493 | 2·217 | 6·367 | 173452 | 77239 | 221873 |
| Rangpur | Dinajpur | 6·180 | 4·800 | 17·827 | 2·950 | 7·872 | 208816 | 99693 | 266196 |
| Rangpur | Gaibandha | 5·543 | 4·720 | 17·319 | 2·425 | 7·073 | 142932 | 62547 | 182358 |
| Rangpur | Kurigram | 5·632 | 4·791 | 17·695 | 2·471 | 7·196 | 143085 | 62846 | 182860 |
| Rangpur | Lalmonirhat | 5·977 | 4·720 | 17·591 | 2·787 | 7·694 | 85659 | 39938 | 110092 |
| Rangpur | Nilphamari | 6·153 | 4·697 | 17·562 | 2·942 | 7·905 | 135471 | 64654 | 173924 |
| Rangpur | Panchagarh | 6·350 | 4·628 | 17·354 | 3·159 | 8·128 | 94883 | 47139 | 121558 |
| Rangpur | Rangpur | 5·866 | 4·747 | 17·640 | 2·687 | 7·516 | 200289 | 91921 | 256391 |
| Rangpur | Thakurgaon | 6·452 | 4·668 | 17·520 | 3·237 | 8·272 | 101472 | 50917 | 130100 |
| Sylhet | Habiganj | 2·629 | 2·406 | 8·689 | 1·115 | 3·301 | 65977 | 27982 | 83015 |
| Sylhet | Maulvibazar | 2·204 | 1·942 | 7·055 | 0·974 | 2·758 | 50579 | 22383 | 63341 |
| Sylhet | Sunamganj | 3·071 | 2·654 | 9·697 | 1·361 | 3·888 | 82262 | 36936 | 104084 |
| Sylhet | Sylhet | 2·401 | 1·964 | 7·150 | 1·127 | 3·020 | 95538 | 44781 | 120337 |

ADM1: administrative level 1. ADM2: administrative level 2. SD: standard deviation. CI95%: 95% credible interval. Q25: 25th percentile. Q75: 75th percentile. SD and CI95% are measures of prediction uncertainty.

**Supplementary Table S17. Subnational areal predictions of β-thalassaemia carrier prevalence and estimated carrier count at administrative level 2 of Bulgaria.**

| ADM1 | ADM2 | β-thalassaemia carrier prevalence<br>Mean | SD | CI95% | Q25 | Q75 | Estimated carrier counts<br>Mean | Q25 | Q75 |
|---|---|---|---|---|---|---|---|---|---|
| Blagoevgrad | Bansko | 10·392 | 7·541 | 28·096 | 5·100 | 13·532 | 849 | 409 | 1107 |
| Blagoevgrad | Belitsa | 10·212 | 7·543 | 28·375 | 4·972 | 13·187 | 1235 | 605 | 1588 |
| Blagoevgrad | Blagoevgrad | 10·843 | 8·105 | 30·434 | 5·133 | 14·152 | 7895 | 3720 | 10286 |
| Blagoevgrad | Garmen | 9·970 | 7·131 | 26·802 | 4·944 | 12·965 | 1350 | 672 | 1763 |
| Blagoevgrad | Gotse Delchev | 10·299 | 7·272 | 27·760 | 5·133 | 13·345 | 3388 | 1694 | 4408 |
| Blagoevgrad | Hadzhidimovo | 9·870 | 6·814 | 25·404 | 5·078 | 12·809 | 728 | 371 | 945 |
| Blagoevgrad | Kresna | 10·876 | 7·857 | 29·406 | 5·329 | 14·168 | 518 | 253 | 683 |
| Blagoevgrad | Petrich | 11·037 | 7·802 | 29·532 | 5·468 | 14·450 | 5312 | 2680 | 7016 |

| | | β-thalassaemia carrier prevalence | | | | | Estimated carrier counts | | |
|---|---|---|---|---|---|---|---|---|---|
| ADM1 | ADM2 | Mean | SD | CI95% | Q25 | Q75 | Mean | Q25 | Q75 |
| Blagoevgrad | Razlog | 10·492 | 7·633 | 29·026 | 5·144 | 13·623 | 2519 | 1238 | 3282 |
| Blagoevgrad | Sandanski | 10·530 | 7·404 | 28·228 | 5·270 | 13·688 | 3924 | 1957 | 5056 |
| Blagoevgrad | Satovcha | 9·521 | 6·645 | 25·026 | 4·846 | 12·262 | 1456 | 743 | 1880 |
| Blagoevgrad | Simitli | 10·760 | 7·893 | 29·610 | 5·227 | 13·946 | 1411 | 689 | 1819 |
| Blagoevgrad | Strumyarni | 11·172 | 8·117 | 30·319 | 5·422 | 14·649 | 490 | 241 | 627 |
| Blagoevgrad | Yakoruda | 10·024 | 7·365 | 27·995 | 4·887 | 12·935 | 883 | 433 | 1134 |
| Burgas | Aytos | 3·902 | 3·985 | 14·605 | 1·368 | 4·957 | 1001 | 355 | 1278 |
| Burgas | Burgas | 3·537 | 3·616 | 13·235 | 1·281 | 4·452 | 6415 | 2290 | 7986 |
| Burgas | Kameno | 3·766 | 3·663 | 13·341 | 1·419 | 4·799 | 300 | 112 | 377 |
| Burgas | Karnobat | 4·171 | 4·147 | 15·379 | 1·515 | 5·269 | 910 | 329 | 1149 |
| Burgas | Malko Tarnovo | 3·043 | 3·002 | 10·614 | 1·196 | 3·816 | 85 | 32 | 105 |
| Burgas | Nesebar | 3·418 | 3·656 | 12·962 | 1·164 | 4·310 | 347 | 120 | 434 |
| Burgas | Pomorie | 3·569 | 3·748 | 13·268 | 1·238 | 4·488 | 639 | 223 | 797 |
| Burgas | Primorsko | 3·086 | 3·127 | 11·171 | 1·154 | 3·891 | 85 | 31 | 110 |
| Burgas | Ruen | 3·988 | 4·178 | 15·123 | 1·353 | 5·034 | 1026 | 350 | 1286 |
| Burgas | Sozopol | 3·281 | 3·266 | 11·753 | 1·228 | 4·141 | 251 | 90 | 309 |
| Burgas | Sredets | 3·698 | 3·583 | 12·913 | 1·425 | 4·695 | 466 | 180 | 592 |
| Burgas | Sungurlare | 4·507 | 4·469 | 16·274 | 1·632 | 5·787 | 504 | 181 | 643 |
| Burgas | Tsarevo | 2·801 | 2·764 | 9·993 | 1·066 | 3·544 | 160 | 62 | 201 |
| Dobrich | Balchik | 3·651 | 4·407 | 15·661 | 1·061 | 4·503 | 365 | 106 | 446 |
| Dobrich | Dobrich | 3·922 | 4·577 | 15·792 | 1·146 | 4·825 | 1980 | 588 | 2448 |
| Dobrich | Dobrichka | 3·980 | 4·687 | 16·444 | 1·178 | 4·948 | 1604 | 471 | 2003 |
| Dobrich | General Toshevo | 3·849 | 4·695 | 16·789 | 1·083 | 4·764 | 445 | 129 | 560 |
| Dobrich | Kavarna | 3·539 | 4·472 | 15·719 | 0·982 | 4·282 | 417 | 116 | 517 |
| Dobrich | Krushari | 4·203 | 5·135 | 17·947 | 1·168 | 5·190 | 102 | 30 | 130 |
| Dobrich | Shabla | 3·546 | 4·550 | 16·091 | 0·955 | 4·256 | 129 | 33 | 156 |
| Dobrich | Tervel | 4·387 | 5·140 | 18·292 | 1·284 | 5·486 | 548 | 158 | 677 |
| Gabrovo | Dryanovo | 6·830 | 6·280 | 23·364 | 2·683 | 8·759 | 537 | 212 | 705 |
| Gabrovo | Gabrovo | 7·145 | 6·313 | 23·366 | 2·892 | 9·282 | 3732 | 1476 | 4877 |
| Gabrovo | Sevlievo | 7·388 | 6·522 | 24·221 | 2·978 | 9·588 | 2160 | 868 | 2812 |
| Gabrovo | Tryavna | 6·530 | 5·719 | 21·405 | 2·697 | 8·407 | 621 | 250 | 785 |
| Haskovo | Dimitrovgrad | 6·277 | 4·970 | 18·483 | 2·908 | 8·084 | 2788 | 1307 | 3559 |
| Haskovo | Harmanli | 5·238 | 4·319 | 16·222 | 2·336 | 6·714 | 1172 | 504 | 1522 |
| Haskovo | Haskovo | 6·002 | 4·696 | 17·363 | 2·807 | 7·753 | 4991 | 2333 | 6534 |
| Haskovo | Ivaylovgrad | 4·734 | 3·801 | 13·970 | 2·212 | 6·017 | 232 | 105 | 290 |
| Haskovo | Lyubimets | 4·932 | 4·084 | 15·097 | 2·191 | 6·316 | 468 | 212 | 592 |
| Haskovo | Madzharovo | 5·168 | 4·143 | 15·536 | 2·373 | 6·606 | 110 | 53 | 140 |
| Haskovo | Mineralni Bani | 6·574 | 4·942 | 19·032 | 3·200 | 8·425 | 312 | 150 | 398 |
| Haskovo | Simeonovgrad | 5·406 | 4·487 | 16·504 | 2·379 | 7·000 | 439 | 194 | 568 |
| Haskovo | Stambolovo | 5·669 | 4·446 | 16·856 | 2·636 | 7·277 | 386 | 178 | 498 |
| Haskovo | Svilengrad | 4·550 | 3·911 | 14·579 | 1·961 | 5·815 | 975 | 430 | 1242 |
| Haskovo | Topolovgrad | 4·699 | 4·147 | 15·430 | 1·935 | 6·034 | 414 | 172 | 523 |
| Kardzhali | Ardino | 6·822 | 5·081 | 18·853 | 3·338 | 8·778 | 822 | 405 | 1060 |
| Kardzhali | Chernoochene | 6·730 | 5·095 | 19·061 | 3·298 | 8·576 | 605 | 291 | 776 |
| Kardzhali | Dzhebel | 6·488 | 4·964 | 18·516 | 3·073 | 8·416 | 583 | 275 | 756 |
| Kardzhali | Kardzhali | 6·325 | 4·911 | 18·137 | 2·977 | 8·094 | 3980 | 1866 | 5102 |
| Kardzhali | Kirkovo | 6·047 | 4·524 | 16·866 | 2·948 | 7·734 | 1222 | 600 | 1568 |
| Kardzhali | Krumovgrad | 5·345 | 4·229 | 15·596 | 2·525 | 6·853 | 927 | 440 | 1194 |
| Kardzhali | Momchilgrad | 5·979 | 4·611 | 17·307 | 2·840 | 7·651 | 991 | 467 | 1261 |
| Kyustendil | Boboshevo | 10·870 | 8·359 | 30·925 | 5·118 | 14·195 | 202 | 96 | 261 |
| Kyustendil | Bobov Dol | 10·633 | 8·306 | 31·651 | 4·829 | 13·951 | 762 | 342 | 1003 |
| Kyustendil | Dupnitsa | 10·668 | 8·222 | 31·115 | 4·966 | 13·905 | 4236 | 1953 | 5521 |
| Kyustendil | Kocherinovo | 10·892 | 8·462 | 32·049 | 4·989 | 14·234 | 538 | 247 | 712 |
| Kyustendil | Kyustendil | 10·934 | 8·727 | 32·670 | 4·894 | 14·288 | 5499 | 2485 | 7159 |
| Kyustendil | Nevestino | 11·072 | 8·653 | 32·725 | 5·029 | 14·356 | 204 | 95 | 267 |
| Kyustendil | Rila | 10·414 | 7·701 | 29·031 | 5·045 | 13·468 | 111 | 51 | 143 |
| Kyustendil | Sapareva Banya | 10·536 | 8·089 | 30·242 | 4·901 | 13·713 | 636 | 297 | 825 |

| | | β-thalassaemia carrier prevalence | | | | | Estimated carrier counts | | |
|---|---|---|---|---|---|---|---|---|---|
| **ADM1** | **ADM2** | **Mean** | **SD** | **CI95%** | **Q25** | **Q75** | **Mean** | **Q25** | **Q75** |
| Kyustendil | Treklyano | 10·610 | 8·652 | 33·397 | 4·614 | 13·745 | 50 | 21 | 63 |
| Lovech | Apriltsi | 7·686 | 6·382 | 23·112 | 3·317 | 10·012 | 184 | 78 | 240 |
| Lovech | Letnitsa | 7·309 | 6·910 | 25·666 | 2·744 | 9·474 | 315 | 117 | 412 |
| Lovech | Lovech | 7·723 | 6·973 | 25·498 | 3·094 | 10·030 | 3064 | 1255 | 4069 |
| Lovech | Lukovit | 8·520 | 7·614 | 28·512 | 3·393 | 10·984 | 1401 | 561 | 1823 |
| Lovech | Teteven | 8·840 | 7·505 | 28·020 | 3·733 | 11·444 | 1415 | 587 | 1826 |
| Lovech | Troyan | 8·130 | 6·905 | 25·717 | 3·438 | 10·527 | 2194 | 943 | 2855 |
| Lovech | Ugarchin | 8·360 | 7·406 | 27·081 | 3·420 | 10·815 | 760 | 311 | 983 |
| Lovech | Yablanitsa | 8·790 | 7·545 | 27·613 | 3·652 | 11·471 | 502 | 212 | 660 |
| Montana | Berkovitsa | 9·612 | 8·811 | 33·520 | 3·734 | 12·297 | 1444 | 559 | 1862 |
| Montana | Boychinovtsi | 9·106 | 8·672 | 32·701 | 3·371 | 11·669 | 582 | 218 | 748 |
| Montana | Brusartsi | 9·072 | 8·775 | 33·391 | 3·294 | 11·659 | 374 | 137 | 482 |
| Montana | Chiprovtsi | 9·370 | 8·770 | 32·974 | 3·520 | 12·007 | 247 | 94 | 317 |
| Montana | Georgi Bamyanovo | 9·648 | 8·978 | 33·949 | 3·667 | 12·432 | 202 | 78 | 259 |
| Montana | Lom | 8·937 | 8·899 | 33·712 | 3·115 | 11·330 | 1815 | 615 | 2305 |
| Montana | Medkovets | 9·018 | 8·770 | 32·888 | 3·229 | 11·617 | 288 | 101 | 378 |
| Montana | Montana | 9·251 | 8·731 | 32·934 | 3·475 | 11·870 | 4372 | 1718 | 5614 |
| Montana | Valchedram | 8·827 | 8·741 | 32·269 | 3·146 | 11·323 | 686 | 243 | 871 |
| Montana | Varshets | 9·444 | 8·470 | 32·484 | 3·710 | 12·204 | 778 | 302 | 997 |
| Montana | Yakimovo | 9·026 | 8·871 | 34·236 | 3·245 | 11·595 | 386 | 137 | 503 |
| Pazardzhik | Batak | 9·115 | 6·322 | 23·451 | 4·700 | 11·769 | 420 | 213 | 554 |
| Pazardzhik | Belovo | 9·751 | 7·145 | 26·788 | 4·780 | 12·592 | 699 | 343 | 907 |
| Pazardzhik | Bratsigovo | 8·862 | 6·187 | 23·210 | 4·558 | 11·372 | 682 | 355 | 868 |
| Pazardzhik | Lesichovo | 9·413 | 7·215 | 26·606 | 4·480 | 12·166 | 265 | 125 | 343 |
| Pazardzhik | Panagyurishte | 9·203 | 7·130 | 26·853 | 4·289 | 11·975 | 1914 | 896 | 2469 |
| Pazardzhik | Pazardzhik | 8·926 | 6·464 | 24·348 | 4·438 | 11·459 | 8764 | 4411 | 11277 |
| Pazardzhik | Peshtera | 8·988 | 6·251 | 23·912 | 4·590 | 11·621 | 1249 | 637 | 1612 |
| Pazardzhik | Rakitovo | 9·381 | 6·582 | 24·441 | 4·822 | 12·102 | 1322 | 695 | 1675 |
| Pazardzhik | Septemvri | 9·393 | 6·852 | 26·038 | 4·648 | 12·131 | 1932 | 972 | 2491 |
| Pazardzhik | Strelcha | 8·949 | 6·848 | 25·916 | 4·220 | 11·510 | 524 | 246 | 673 |
| Pazardzhik | Velingrad | 9·790 | 7·112 | 26·654 | 4·829 | 12·674 | 3611 | 1746 | 4749 |
| Pernik | Breznik | 10·247 | 8·504 | 32·357 | 4·415 | 13·325 | 618 | 269 | 796 |
| Pernik | Kovachevtsi | 10·432 | 8·459 | 31·790 | 4·554 | 13·515 | 119 | 53 | 156 |
| Pernik | Pernik | 10·190 | 8·242 | 31·435 | 4·512 | 13·262 | 9151 | 4122 | 11912 |
| Pernik | Radomir | 10·439 | 8·268 | 31·156 | 4·714 | 13·539 | 1854 | 831 | 2432 |
| Pernik | Tran | 10·326 | 8·843 | 33·491 | 4·296 | 13·369 | 356 | 144 | 468 |
| Pernik | Zemen | 10·475 | 8·541 | 32·479 | 4·616 | 13·568 | 218 | 96 | 281 |
| Pleven | Belene | 7·068 | 7·073 | 26·268 | 2·470 | 9·054 | 465 | 159 | 604 |
| Pleven | Cherven Bryag | 8·510 | 7·740 | 28·808 | 3·348 | 10·931 | 1989 | 789 | 2582 |
| Pleven | Dolna Mitropoliya | 8·038 | 7·924 | 29·313 | 2·880 | 10·340 | 1136 | 404 | 1466 |
| Pleven | Dolni Dabnik | 8·237 | 7·683 | 28·616 | 3·162 | 10·694 | 788 | 302 | 1026 |
| Pleven | Gulyantsi | 7·793 | 7·914 | 29·051 | 2·665 | 10·004 | 1112 | 376 | 1433 |
| Pleven | Iskar | 8·381 | 7·934 | 29·330 | 3·125 | 10·938 | 485 | 181 | 643 |
| Pleven | Knezha | 8·452 | 8·046 | 29·713 | 3·199 | 10·742 | 1111 | 426 | 1364 |
| Pleven | Levski | 7·077 | 6·752 | 25·317 | 2·619 | 9·154 | 1130 | 423 | 1457 |
| Pleven | Nikopol | 7·329 | 7·246 | 27·215 | 2·581 | 9·511 | 551 | 197 | 721 |
| Pleven | Pleven | 7·829 | 7·450 | 27·531 | 2·947 | 10·128 | 9333 | 3423 | 12136 |
| Pleven | Pordim | 7·405 | 7·025 | 25·756 | 2·786 | 9·521 | 319 | 122 | 410 |
| Plovdiv | Asenovgrad | 7·497 | 5·407 | 20·286 | 3·758 | 9·628 | 4750 | 2380 | 6091 |
| Plovdiv | Brezovo | 7·452 | 5·758 | 21·359 | 3·503 | 9·668 | 353 | 169 | 453 |
| Plovdiv | Hisarya | 8·350 | 6·403 | 23·796 | 3·961 | 10·758 | 818 | 397 | 1047 |
| Plovdiv | Kaloyanovo | 7·921 | 5·935 | 22·015 | 3·812 | 10·259 | 774 | 377 | 1003 |
| Plovdiv | Karlovo | 8·070 | 6·476 | 24·022 | 3·640 | 10·474 | 4475 | 2033 | 5739 |
| Plovdiv | Krichim | 8·768 | 6·338 | 23·796 | 4·264 | 11·261 | 59 | 29 | 75 |
| Plovdiv | Kuklen | 8·163 | 5·740 | 21·642 | 4·167 | 10·501 | 681 | 350 | 882 |
| Plovdiv | Laki | 7·686 | 5·486 | 20·434 | 3·899 | 9·878 | 176 | 89 | 226 |
| Plovdiv | Maritsa | 8·141 | 5·938 | 22·532 | 4·040 | 10·507 | 2455 | 1224 | 3154 |

| | | β-thalassaemia carrier prevalence | | | | | Estimated carrier counts | | |
|---|---|---|---|---|---|---|---|---|---|
| **ADM1** | **ADM2** | **Mean** | **SD** | **CI95%** | **Q25** | **Q75** | **Mean** | **Q25** | **Q75** |
| Plovdiv | Parvomay | 6·945 | 5·156 | 19·352 | 3·381 | 8·985 | 1524 | 727 | 1990 |
| Plovdiv | Perushtitsa | 8·469 | 5·744 | 21·254 | 4·357 | 11·091 | 598 | 304 | 782 |
| Plovdiv | Plovdiv | 8·294 | 5·692 | 21·773 | 4·204 | 10·765 | 16280 | 8230 | 21101 |
| Plovdiv | Rakovski | 7·549 | 5·633 | 20·930 | 3·651 | 9·755 | 1982 | 962 | 2540 |
| Plovdiv | Rodopi | 8·463 | 6·006 | 22·734 | 4·263 | 10·929 | 14921 | 7502 | 19339 |
| Plovdiv | Sadovo | 7·552 | 5·594 | 21·366 | 3·698 | 9·703 | 1187 | 582 | 1521 |
| Plovdiv | Saedinenie | 8·431 | 6·245 | 23·158 | 4·112 | 10·918 | 824 | 412 | 1065 |
| Plovdiv | Sopot | 7·996 | 6·479 | 24·189 | 3·557 | 10·405 | 49 | 22 | 64 |
| Plovdiv | Stamboliyski | 9·045 | 6·487 | 24·692 | 4·506 | 11·298 | 292 | 146 | 365 |
| Razgrad | Isperih | 4·886 | 5·437 | 19·659 | 1·504 | 6·219 | 899 | 279 | 1145 |
| Razgrad | Kubrat | 5·349 | 5·893 | 21·679 | 1·614 | 6·840 | 679 | 204 | 872 |
| Razgrad | Loznitsa | 5·002 | 5·311 | 18·822 | 1·640 | 6·436 | 347 | 113 | 448 |
| Razgrad | Razgrad | 5·157 | 5·505 | 19·772 | 1·690 | 6·563 | 2351 | 785 | 3010 |
| Razgrad | Samuil | 4·817 | 5·209 | 19·209 | 1·529 | 6·166 | 378 | 122 | 483 |
| Razgrad | Tsar Kaloyan | 5·569 | 5·828 | 20·929 | 1·848 | 7·142 | 239 | 79 | 305 |
| Razgrad | Zavet | 5·094 | 5·631 | 20·289 | 1·555 | 6·441 | 449 | 137 | 569 |
| Ruse | Borovo | 6·130 | 6·283 | 22·971 | 2·100 | 7·926 | 142 | 48 | 184 |
| Ruse | Byala | 6·206 | 6·266 | 22·714 | 2·203 | 7·905 | 688 | 249 | 879 |
| Ruse | Dve Mogili | 5·951 | 6·127 | 21·724 | 2·028 | 7·658 | 489 | 163 | 626 |
| Ruse | Ivanovo | 5·954 | 6·282 | 22·990 | 1·937 | 7·649 | 488 | 161 | 628 |
| Ruse | Ruse | 5·690 | 6·197 | 22·079 | 1·804 | 7·265 | 8219 | 2545 | 10640 |
| Ruse | Slivo Pole | 5·637 | 6·197 | 22·429 | 1·721 | 7·266 | 699 | 214 | 890 |
| Ruse | Tsenovo | 6·318 | 6·457 | 23·327 | 2·175 | 8·045 | 277 | 94 | 352 |
| Ruse | Vetovo | 5·362 | 5·683 | 20·601 | 1·742 | 6·888 | 625 | 198 | 809 |
| Shumen | Hitrino | 4·711 | 5·107 | 18·836 | 1·483 | 5·965 | 333 | 105 | 422 |
| Shumen | Kaolinovo | 4·680 | 5·282 | 19·002 | 1·420 | 5·890 | 545 | 165 | 686 |
| Shumen | Kaspichan | 4·320 | 4·723 | 17·038 | 1·388 | 5·445 | 835 | 264 | 1076 |
| Shumen | Nikola Kozlevo | 4·466 | 5·069 | 18·647 | 1·323 | 5·693 | 236 | 71 | 300 |
| Shumen | Novi Pazar | 4·384 | 4·810 | 17·510 | 1·355 | 5·582 | 249 | 76 | 319 |
| Shumen | Shumen | 4·524 | 4·793 | 17·491 | 1·480 | 5·770 | 3574 | 1170 | 4503 |
| Shumen | Smyadovo | 4·323 | 4·558 | 16·730 | 1·463 | 5·464 | 238 | 80 | 302 |
| Shumen | Varbitsa | 4·745 | 4·806 | 17·335 | 1·678 | 6·021 | 558 | 196 | 707 |
| Shumen | Veliki Preslav | 4·599 | 4·774 | 17·360 | 1·560 | 5·909 | 516 | 177 | 657 |
| Shumen | Venets | 4·717 | 5·138 | 18·797 | 1·468 | 5·954 | 198 | 61 | 250 |
| Silistra | Alfatar | 4·537 | 5·403 | 18·644 | 1·302 | 5·648 | 105 | 30 | 131 |
| Silistra | Dulovo | 4·683 | 5·422 | 19·330 | 1·362 | 5·855 | 1246 | 365 | 1553 |
| Silistra | Glavinitsa | 5·009 | 5·687 | 20·606 | 1·465 | 6·399 | 376 | 110 | 474 |
| Silistra | Kaynardzha | 4·386 | 5·236 | 18·534 | 1·230 | 5·464 | 169 | 49 | 212 |
| Silistra | Silistra | 4·706 | 5·592 | 19·929 | 1·337 | 5·880 | 2026 | 574 | 2476 |
| Silistra | Sitovo | 4·896 | 5·703 | 20·527 | 1·413 | 6·145 | 203 | 58 | 253 |
| Silistra | Tutrakan | 5·245 | 5·899 | 21·435 | 1·561 | 6·691 | 685 | 204 | 880 |
| Sliven | Kotel | 5·135 | 4·952 | 18·234 | 1·916 | 6·556 | 666 | 247 | 849 |
| Sliven | Nova Zagora | 5·558 | 4·978 | 18·449 | 2·281 | 7·092 | 1785 | 746 | 2279 |
| Sliven | Sliven | 5·261 | 5·011 | 18·159 | 2·019 | 6·742 | 6039 | 2325 | 7733 |
| Sliven | Tvarditsa | 5·805 | 5·344 | 19·829 | 2·266 | 7·454 | 831 | 327 | 1072 |
| Smolyan | Banite | 7·270 | 5·220 | 19·514 | 3·657 | 9·319 | 247 | 121 | 314 |
| Smolyan | Borino | 8·881 | 6·139 | 23·429 | 4·528 | 11·530 | 217 | 113 | 278 |
| Smolyan | Chepelare | 8·125 | 5·662 | 21·247 | 4·181 | 10·455 | 468 | 241 | 605 |
| Smolyan | Devin | 8·675 | 5·991 | 22·722 | 4·487 | 11·180 | 846 | 437 | 1088 |
| Smolyan | Dospat | 9·222 | 6·460 | 24·213 | 4·707 | 11·842 | 607 | 313 | 777 |
| Smolyan | Madan | 7·263 | 5·361 | 20·043 | 3·501 | 9·415 | 711 | 341 | 920 |
| Smolyan | Nedelino | 6·956 | 5·296 | 19·989 | 3·350 | 8·868 | 475 | 233 | 599 |
| Smolyan | Rudozem | 7·513 | 5·505 | 20·721 | 3·683 | 9·757 | 602 | 292 | 783 |
| Smolyan | Smolyan | 8·037 | 5·724 | 21·436 | 4·048 | 10·355 | 2731 | 1364 | 3529 |
| Smolyan | Zlatograd | 6·796 | 4·891 | 18·141 | 3·418 | 8·731 | 524 | 263 | 666 |
| Sofia | Anton | 8·979 | 7·230 | 26·414 | 4·065 | 11·591 | 140 | 64 | 181 |
| Sofia | Botevgrad | 9·384 | 7·922 | 29·707 | 4·023 | 12·105 | 3016 | 1319 | 3917 |

| | | β-thalassaemia carrier prevalence | | | | | Estimated carrier counts | | |
|---|---|---|---|---|---|---|---|---|---|
| ADM1 | ADM2 | Mean | SD | CI95% | Q25 | Q75 | Mean | Q25 | Q75 |
| Sofia | Bozhurishte | 10·055 | 8·443 | 31·493 | 4·347 | 13·081 | 223 | 95 | 292 |
| Sofia | Chavdar | 9·177 | 7·308 | 27·030 | 4·245 | 11·698 | 35 | 16 | 45 |
| Sofia | Chelopech | 9·056 | 7·130 | 26·417 | 4·150 | 11·532 | 49 | 22 | 60 |
| Sofia | Dolna Banya | 9·985 | 7·488 | 28·330 | 4·801 | 13·058 | 471 | 222 | 611 |
| Sofia | Dragoman | 10·011 | 8·611 | 32·699 | 4·137 | 12·991 | 447 | 185 | 581 |
| Sofia | Elin Pelin | 9·725 | 7·835 | 29·888 | 4·301 | 12·661 | 2305 | 1027 | 2982 |
| Sofia | Etropole | 9·146 | 7·697 | 28·809 | 3·936 | 11·826 | 1095 | 477 | 1405 |
| Sofia | Godech | 9·941 | 8·811 | 33·507 | 3·976 | 12·760 | 530 | 208 | 688 |
| Sofia | Gorna Malina | 9·436 | 7·648 | 29·472 | 4·192 | 12·261 | 630 | 278 | 821 |
| Sofia | Ihtiman | 9·752 | 7·609 | 28·692 | 4·507 | 12·600 | 1796 | 837 | 2299 |
| Sofia | Koprivshtitsa | 8·748 | 6·857 | 25·523 | 4·014 | 11·369 | 181 | 82 | 240 |
| Sofia | Kostenets | 9·747 | 7·439 | 28·144 | 4·692 | 12·577 | 1164 | 559 | 1503 |
| Sofia | Kostinbrod | 9·933 | 8·524 | 32·536 | 4·225 | 12·816 | 1033 | 440 | 1341 |
| Sofia | Mirkovo | 9·466 | 7·607 | 28·572 | 4·259 | 12·216 | 269 | 125 | 346 |
| Sofia | Pirdop | 9·085 | 7·381 | 27·365 | 4·091 | 11·818 | 831 | 370 | 1040 |
| Sofia | Pravets | 9·063 | 7·667 | 28·792 | 3·820 | 11·756 | 480 | 205 | 620 |
| Sofia | Samokov | 10·217 | 7·881 | 29·806 | 4·795 | 13·189 | 3509 | 1676 | 4564 |
| Sofia | Slivnitsa | 10·003 | 8·493 | 31·733 | 4·207 | 12·899 | 906 | 381 | 1158 |
| Sofia | Svoge | 9·623 | 8·285 | 31·225 | 4·053 | 12·353 | 1961 | 837 | 2496 |
| Sofia | Zlatitsa | 9·144 | 7·383 | 27·901 | 4·097 | 11·878 | 504 | 224 | 655 |
| Sofia | Zlatitsa | 9·400 | 7·131 | 27·378 | 4·359 | 12·433 | 14 | 6 | 18 |
| Sofia City | Stolichna | 10·049 | 8·260 | 31·173 | 4·416 | 12·927 | 133977 | 59000 | 172825 |
| Stara Zagora | Bratya Daskalovi | 7·024 | 5·510 | 20·613 | 3·266 | 9·025 | 637 | 297 | 810 |
| Stara Zagora | Chirpan | 6·713 | 5·358 | 20·119 | 3·078 | 8·673 | 1178 | 541 | 1500 |
| Stara Zagora | Galabovo | 5·474 | 4·664 | 17·244 | 2·351 | 7·087 | 597 | 259 | 772 |
| Stara Zagora | Gurkovo | 6·122 | 5·371 | 19·794 | 2·546 | 7·885 | 214 | 92 | 272 |
| Stara Zagora | Kazanlak | 6·783 | 5·686 | 21·060 | 2·953 | 8·757 | 4459 | 1945 | 5714 |
| Stara Zagora | Maglizh | 6·339 | 5·454 | 20·258 | 2·667 | 8·195 | 587 | 250 | 753 |
| Stara Zagora | Nikolaevo | 6·033 | 5·353 | 19·223 | 2·501 | 7·816 | 322 | 134 | 417 |
| Stara Zagora | Opan | 6·099 | 5·074 | 18·825 | 2·693 | 7·829 | 168 | 72 | 216 |
| Stara Zagora | Pavel Banya | 7·292 | 5·955 | 22·220 | 3·194 | 9·523 | 931 | 409 | 1220 |
| Stara Zagora | Radnevo | 5·574 | 4·729 | 17·443 | 2·417 | 7·190 | 894 | 383 | 1139 |
| Stara Zagora | Stara Zagora | 6·293 | 5·242 | 19·417 | 2·753 | 8·090 | 9264 | 4038 | 11886 |
| Targovishte | Antonovo | 5·556 | 5·520 | 19·984 | 2·028 | 7·063 | 242 | 90 | 308 |
| Targovishte | Omurtag | 5·178 | 5·147 | 19·154 | 1·858 | 6·593 | 843 | 301 | 1068 |
| Targovishte | Opaka | 5·625 | 5·765 | 21·530 | 1·972 | 7·129 | 326 | 113 | 409 |
| Targovishte | Popovo | 5·548 | 5·681 | 20·823 | 1·931 | 7·074 | 1306 | 455 | 1666 |
| Targovishte | Targovishte | 5·095 | 5·270 | 19·319 | 1·731 | 6·499 | 2534 | 853 | 3204 |
| Varna | Aksakovo | 3·711 | 4·353 | 15·325 | 1·133 | 4·587 | 1450 | 440 | 1842 |
| Varna | Avren | 3·640 | 3·964 | 14·431 | 1·184 | 4·586 | 347 | 111 | 435 |
| Varna | Beloslav | 3·794 | 4·298 | 15·487 | 1·182 | 4·665 | 455 | 144 | 561 |
| Varna | Byala | 3·414 | 3·795 | 13·380 | 1·144 | 4·209 | 63 | 22 | 78 |
| Varna | Dalgopol | 3·978 | 4·268 | 15·178 | 1·358 | 5·019 | 551 | 189 | 694 |
| Varna | Devnya | 3·867 | 4·294 | 15·669 | 1·257 | 4·766 | 279 | 91 | 343 |
| Varna | Dolni Chiflik | 3·665 | 3·954 | 14·415 | 1·231 | 4·612 | 613 | 203 | 775 |
| Varna | Provadiya | 3·994 | 4·242 | 15·416 | 1·322 | 5·058 | 704 | 227 | 891 |
| Varna | Suvorovo | 3·862 | 4·452 | 15·963 | 1·217 | 4·769 | 250 | 78 | 316 |
| Varna | Valchi Dol | 4·110 | 4·691 | 16·881 | 1·264 | 5·133 | 313 | 96 | 386 |
| Varna | Varna | 3·628 | 4·204 | 15·157 | 1·128 | 4·509 | 8902 | 2784 | 10780 |
| Varna | Vetrino | 4·085 | 4·487 | 16·543 | 1·301 | 5·179 | 195 | 62 | 249 |
| Veliko Tarnovo | Elena | 5·938 | 5·529 | 20·568 | 2·307 | 7·619 | 424 | 163 | 532 |
| Veliko Tarnovo | Gorna Oryahovitsa | 6·270 | 6·009 | 22·315 | 2·325 | 8·022 | 2035 | 754 | 2603 |
| Veliko Tarnovo | Lyaskovets | 6·137 | 5·801 | 21·034 | 2·300 | 8·104 | 1151 | 432 | 1538 |
| Veliko Tarnovo | Pavlikeni | 6·919 | 6·560 | 24·028 | 2·610 | 8·884 | 1273 | 476 | 1642 |
| Veliko Tarnovo | Polski Trambesh | 6·444 | 6·262 | 22·872 | 2·354 | 8·274 | 823 | 293 | 1048 |
| Veliko Tarnovo | Strazhitsa | 6·011 | 5·968 | 22·084 | 2·167 | 7·623 | 622 | 221 | 788 |
| Veliko Tarnovo | Suhindol | 7·199 | 6·670 | 24·679 | 2·749 | 9·243 | 121 | 45 | 157 |

| | | β-thalassaemia carrier prevalence | | | | | Estimated carrier counts | | |
|---|---|---|---|---|---|---|---|---|---|
| ADM1 | ADM2 | Mean | SD | CI95% | Q25 | Q75 | Mean | Q25 | Q75 |
| Veliko Tarnovo | Svishtov | 6·798 | 6·766 | 24·755 | 2·354 | 8·765 | 1713 | 586 | 2183 |
| Veliko Tarnovo | Veliko Tarnovo | 6·514 | 6·018 | 22·417 | 2·533 | 8·382 | 5005 | 1913 | 6405 |
| Veliko Tarnovo | Zlataritsa | 5·933 | 5·802 | 20·987 | 2·244 | 7·584 | 270 | 102 | 338 |
| Vidin | Belogradchik | 9·324 | 9·089 | 34·294 | 3·219 | 12·130 | 472 | 158 | 615 |
| Vidin | Boynitsa | 8·797 | 9·167 | 34·938 | 3·032 | 11·105 | 68 | 23 | 85 |
| Vidin | Bregovo | 8·628 | 9·038 | 34·250 | 2·871 | 10·819 | 276 | 92 | 342 |
| Vidin | Chuprene | 9·477 | 9·143 | 34·052 | 3·419 | 12·252 | 173 | 63 | 226 |
| Vidin | Dimovo | 9·135 | 9·054 | 33·929 | 3·223 | 11·610 | 459 | 164 | 585 |
| Vidin | Gramada | 9·086 | 9·274 | 35·049 | 3·138 | 11·480 | 134 | 46 | 167 |
| Vidin | Kula | 8·926 | 9·151 | 34·096 | 3·011 | 11·231 | 307 | 102 | 387 |
| Vidin | Makresh | 9·009 | 8·865 | 32·394 | 3·136 | 11·562 | 64 | 22 | 84 |
| Vidin | Novo Selo | 8·494 | 8·930 | 32·237 | 2·761 | 10·882 | 181 | 59 | 233 |
| Vidin | Ruzhinsi | 9·145 | 8·822 | 33·113 | 3·320 | 11·812 | 254 | 92 | 327 |
| Vidin | Vidin | 8·872 | 9·108 | 34·848 | 2·957 | 11·265 | 3794 | 1247 | 4849 |
| Vratsa | Borovan | 8·918 | 8·293 | 31·313 | 3·447 | 11·450 | 485 | 184 | 622 |
| Vratsa | Byala Slatina | 8·784 | 8·180 | 30·623 | 3·363 | 11·327 | 1916 | 719 | 2501 |
| Vratsa | Hayredin | 8·841 | 8·585 | 32·937 | 3·226 | 11·316 | 279 | 101 | 356 |
| Vratsa | Kozloduy | 8·650 | 8·543 | 31·782 | 3·110 | 11·053 | 814 | 292 | 1040 |
| Vratsa | Krivodol | 9·041 | 8·434 | 31·703 | 3·441 | 11·648 | 580 | 224 | 753 |
| Vratsa | Mezdra | 9·261 | 8·209 | 30·830 | 3·783 | 11·899 | 1709 | 669 | 2211 |
| Vratsa | Miziya | 8·555 | 8·393 | 31·884 | 3·070 | 10·814 | 637 | 224 | 805 |
| Vratsa | Oryahovo | 8·381 | 8·287 | 30·837 | 2·931 | 10·715 | 1163 | 398 | 1454 |
| Vratsa | Roman | 8·974 | 7·924 | 29·487 | 3·628 | 11·584 | 456 | 185 | 584 |
| Vratsa | Vratsa | 9·155 | 8·260 | 30·869 | 3·608 | 11·803 | 5914 | 2312 | 7593 |
| Yambol | Bolyarovo | 3·986 | 3·774 | 13·697 | 1·546 | 5·080 | 134 | 51 | 169 |
| Yambol | Elhovo | 4·287 | 3·926 | 14·491 | 1·749 | 5·437 | 626 | 254 | 801 |
| Yambol | Straldzha | 4·363 | 4·149 | 15·109 | 1·661 | 5·606 | 511 | 192 | 651 |
| Yambol | Tundzha | 4·763 | 4·350 | 15·911 | 1·901 | 6·099 | 1139 | 450 | 1439 |
| Yambol | Yambol | 4·827 | 4·532 | 16·319 | 1·878 | 6·252 | 2924 | 1136 | 3814 |

ADM1: administrative level 1. ADM2: administrative level 2. SD: standard deviation. CI95%: 95% credible interval. Q25: 25th percentile. Q75: 75th percentile. SD and CI95% are measures of prediction uncertainty.

**Supplementary Table S18. Subnational areal predictions of β-thalassaemia carrier prevalence and estimated carrier count at administrative level 2 of Cambodia.**

| | | β-thalassaemia carrier prevalence | | | | | Estimated carrier counts | | |
|---|---|---|---|---|---|---|---|---|---|
| ADM1 | ADM2 | Mean | SD | CI95% | Q25 | Q75 | Mean | Q25 | Q75 |
| Bantey Meanchey | Malai | 1·192 | 0·914 | 3·339 | 0·599 | 1·505 | 696 | 347 | 885 |
| Bantey Meanchey | Mongkol Borei | 0·934 | 0·732 | 2·660 | 0·460 | 1·172 | 1991 | 973 | 2522 |
| Bantey Meanchey | Ou Chrov | 1·056 | 0·828 | 2·972 | 0·518 | 1·322 | 846 | 415 | 1056 |
| Bantey Meanchey | Paoy Paet | 1·133 | 0·864 | 3·201 | 0·565 | 1·430 | 689 | 343 | 867 |
| Bantey Meanchey | Phnum Srok | 0·849 | 0·676 | 2·513 | 0·404 | 1·081 | 598 | 290 | 758 |
| Bantey Meanchey | Preah Netr Preah | 0·840 | 0·659 | 2·428 | 0·409 | 1·060 | 1303 | 628 | 1630 |
| Bantey Meanchey | Serei Saophoan | 0·963 | 0·738 | 2·720 | 0·473 | 1·210 | 1146 | 570 | 1432 |
| Bantey Meanchey | Svay Chek | 1·012 | 0·793 | 2·901 | 0·493 | 1·272 | 987 | 483 | 1236 |
| Bantey Meanchey | Thma Puok | 1·019 | 0·847 | 3·042 | 0·480 | 1·278 | 921 | 440 | 1161 |
| Battambang | Aek Phnum | 0·756 | 0·582 | 2·123 | 0·373 | 0·951 | 678 | 335 | 850 |
| Battambang | Banan | 0·874 | 0·682 | 2·468 | 0·426 | 1·107 | 879 | 429 | 1109 |
| Battambang | Battambang | 0·823 | 0·630 | 2·251 | 0·413 | 1·043 | 1404 | 705 | 1784 |
| Battambang | Bavel | 0·978 | 0·771 | 2·821 | 0·475 | 1·237 | 1087 | 528 | 1376 |
| Battambang | Kamrieng | 1·117 | 0·875 | 3·189 | 0·546 | 1·409 | 599 | 294 | 748 |
| Battambang | Koas Krala | 0·821 | 0·659 | 2·408 | 0·391 | 1·035 | 326 | 153 | 417 |
| Battambang | Moung Ruessei | 0·723 | 0·589 | 2·126 | 0·344 | 0·908 | 864 | 419 | 1080 |
| Battambang | Phnum Proek | 1·154 | 0·909 | 3·331 | 0·562 | 1·463 | 532 | 262 | 677 |
| Battambang | Rotonak Mondol | 0·941 | 0·751 | 2·734 | 0·452 | 1·191 | 414 | 199 | 524 |
| Battambang | Rukh Kiri | 0·748 | 0·612 | 2·203 | 0·353 | 0·946 | 341 | 163 | 433 |
| Battambang | Samlout | 0·962 | 0·803 | 2·901 | 0·446 | 1·212 | 511 | 242 | 648 |
| Battambang | Sampov Lun | 1·166 | 0·914 | 3·272 | 0·576 | 1·474 | 696 | 342 | 864 |

| ADM1 | ADM2 | β-thalassaemia carrier prevalence Mean | SD | CI95% | Q25 | Q75 | Estimated carrier counts Mean | Q25 | Q75 |
|---|---|---|---|---|---|---|---|---|---|
| Battambang | Sangkae | 0·756 | 0·594 | 2·166 | 0·369 | 0·953 | 778 | 385 | 983 |
| Battambang | Thma Koul | 0·866 | 0·674 | 2·484 | 0·430 | 1·087 | 1141 | 577 | 1445 |
| Kampong Cham | Batheay | 0·577 | 0·550 | 1·913 | 0·246 | 0·720 | 762 | 320 | 945 |
| Kampong Cham | Chamkar Leu | 0·550 | 0·530 | 1·899 | 0·227 | 0·678 | 660 | 271 | 810 |
| Kampong Cham | Cheung Prey | 0·570 | 0·541 | 1·884 | 0·242 | 0·712 | 515 | 220 | 652 |
| Kampong Cham | Kampong Cham | 0·604 | 0·611 | 2·287 | 0·238 | 0·742 | 151 | 59 | 185 |
| Kampong Cham | Kampong Siem | 0·597 | 0·625 | 2·070 | 0·237 | 0·735 | 534 | 214 | 653 |
| Kampong Cham | Kang Meas | 0·602 | 0·591 | 2·086 | 0·248 | 0·750 | 596 | 244 | 736 |
| Kampong Cham | Kaoh Soutin | 0·628 | 0·663 | 2·277 | 0·242 | 0·764 | 522 | 200 | 633 |
| Kampong Cham | Prey Chhor | 0·575 | 0·572 | 1·951 | 0·238 | 0·713 | 1002 | 414 | 1243 |
| Kampong Cham | Srei Santhor | 0·619 | 0·644 | 2·138 | 0·247 | 0·765 | 650 | 260 | 804 |
| Kampong Cham | Stueng Trang | 0·565 | 0·575 | 1·989 | 0·223 | 0·700 | 588 | 235 | 726 |
| Kampong Chhnang | Baribour | 0·576 | 0·518 | 1·810 | 0·258 | 0·721 | 379 | 169 | 470 |
| Kampong Chhnang | Chol Kiri | 0·568 | 0·527 | 1·852 | 0·241 | 0·711 | 161 | 69 | 199 |
| Kampong Chhnang | Kampong Chhnang | 0·579 | 0·580 | 1·955 | 0·243 | 0·691 | 255 | 107 | 305 |
| Kampong Chhnang | Kampong Leaeng | 0·548 | 0·500 | 1·757 | 0·243 | 0·679 | 321 | 140 | 401 |
| Kampong Chhnang | Kampong Tralach | 0·591 | 0·555 | 1·933 | 0·250 | 0·735 | 700 | 296 | 871 |
| Kampong Chhnang | Rolea B'ier | 0·576 | 0·525 | 1·883 | 0·251 | 0·718 | 753 | 331 | 941 |
| Kampong Chhnang | Sameakki Mean Chey | 0·625 | 0·562 | 2·030 | 0·274 | 0·785 | 606 | 266 | 765 |
| Kampong Chhnang | Tuek Phos | 0·623 | 0·571 | 2·026 | 0·269 | 0·780 | 490 | 209 | 615 |
| Kampong Speu | Aoral | 0·752 | 0·728 | 2·581 | 0·306 | 0·939 | 322 | 130 | 406 |
| Kampong Speu | Basedth | 0·823 | 0·840 | 2·983 | 0·314 | 1·032 | 1228 | 470 | 1546 |
| Kampong Speu | Chbar Mon | 0·722 | 0·680 | 2·586 | 0·299 | 0·902 | 462 | 188 | 575 |
| Kampong Speu | Kong Pisei | 0·763 | 0·777 | 2·641 | 0·301 | 0·943 | 1200 | 471 | 1476 |
| Kampong Speu | Odongk | 0·663 | 0·614 | 2·215 | 0·284 | 0·824 | 1069 | 455 | 1326 |
| Kampong Speu | Phnum Sruoch | 0·835 | 0·839 | 2·992 | 0·323 | 1·034 | 1043 | 401 | 1283 |
| Kampong Speu | Samraong Tong | 0·730 | 0·721 | 2·593 | 0·296 | 0·905 | 1525 | 621 | 1903 |
| Kampong Speu | Thpong | 0·689 | 0·641 | 2·294 | 0·288 | 0·854 | 465 | 194 | 576 |
| Kampong Thom | Baray | 0·534 | 0·511 | 1·797 | 0·225 | 0·665 | 1054 | 444 | 1319 |
| Kampong Thom | Kampong Svay | 0·537 | 0·468 | 1·673 | 0·245 | 0·669 | 700 | 314 | 875 |
| Kampong Thom | Prasat Ballangk | 0·480 | 0·418 | 1·496 | 0·219 | 0·600 | 304 | 138 | 381 |
| Kampong Thom | Prasat Sambour | 0·486 | 0·453 | 1·616 | 0·213 | 0·602 | 253 | 112 | 313 |
| Kampong Thom | Sandan | 0·469 | 0·434 | 1·547 | 0·201 | 0·586 | 338 | 145 | 425 |
| Kampong Thom | Santuk | 0·520 | 0·498 | 1·756 | 0·217 | 0·645 | 675 | 281 | 834 |
| Kampong Thom | Stoung | 0·520 | 0·438 | 1·577 | 0·242 | 0·650 | 595 | 277 | 751 |
| Kampong Thom | Stueng Saen | 0·519 | 0·479 | 1·668 | 0·230 | 0·647 | 211 | 95 | 262 |
| Kampot | Angkor Chey | 0·965 | 1·048 | 3·694 | 0·346 | 1·196 | 753 | 269 | 928 |
| Kampot | Banteay Meas | 1·054 | 1·140 | 4·105 | 0·362 | 1·301 | 1083 | 376 | 1336 |
| Kampot | Chhuk | 0·976 | 1·011 | 3·578 | 0·356 | 1·225 | 1232 | 440 | 1533 |
| Kampot | Chum Kiri | 0·920 | 0·966 | 3·355 | 0·342 | 1·139 | 524 | 193 | 650 |
| Kampot | Dang Tong | 1·062 | 1·172 | 4·047 | 0·366 | 1·327 | 522 | 180 | 651 |
| Kampot | Kampong Trach | 1·144 | 1·296 | 4·546 | 0·384 | 1·391 | 1282 | 432 | 1556 |
| Kampot | Kampot | 1·192 | 1·347 | 4·581 | 0·391 | 1·506 | 540 | 178 | 688 |
| Kampot | Tuek Chhou | 1·133 | 1·250 | 4·517 | 0·387 | 1·390 | 1225 | 413 | 1501 |
| Kandal | Angk Snuol | 0·682 | 0·665 | 2·279 | 0·286 | 0·831 | 788 | 328 | 959 |
| Kandal | Kandal Stueng | 0·706 | 0·712 | 2·460 | 0·282 | 0·869 | 787 | 315 | 971 |
| Kandal | Kaoh Thum | 0·812 | 0·846 | 3·011 | 0·304 | 1·010 | 1369 | 509 | 1687 |
| Kandal | Khsach Kandal | 0·633 | 0·635 | 2·122 | 0·263 | 0·793 | 938 | 389 | 1177 |
| Kandal | Kien Svay | 0·708 | 0·742 | 2·516 | 0·274 | 0·877 | 831 | 318 | 1025 |
| Kandal | Leuk Daek | 0·810 | 0·898 | 3·056 | 0·293 | 0·996 | 226 | 80 | 280 |
| Kandal | Lvea Aem | 0·681 | 0·746 | 2·516 | 0·270 | 0·823 | 794 | 315 | 954 |
| Kandal | Mukh Kampul | 0·616 | 0·585 | 2·062 | 0·260 | 0·775 | 653 | 274 | 821 |
| Kandal | Ponhea Lueu | 0·622 | 0·593 | 2·086 | 0·264 | 0·769 | 1146 | 488 | 1410 |
| Kandal | S'ang | 0·727 | 0·741 | 2·557 | 0·286 | 0·896 | 2011 | 799 | 2470 |
| Kandal | Ta Khmau | 0·682 | 0·658 | 2·538 | 0·268 | 0·865 | 434 | 171 | 550 |
| Kep | Damnak Chang'aeur | 1·239 | 1·475 | 4·949 | 0·386 | 1·502 | 155 | 48 | 188 |
| Kep | Kaeb | 1·164 | 1·313 | 4·445 | 0·380 | 1·459 | 360 | 118 | 450 |

| | | β-thalassaemia carrier prevalence | | | | | Estimated carrier counts | | |
|---|---|---|---|---|---|---|---|---|---|
| ADM1 | ADM2 | Mean | SD | CI95% | Q25 | Q75 | Mean | Q25 | Q75 |
| Koh Kong | Botum Sakor | 1·160 | 1·260 | 4·406 | 0·404 | 1·446 | 234 | 79 | 293 |
| Koh Kong | Kaoh Kong | 1·088 | 1·146 | 3·920 | 0·409 | 1·356 | 51 | 17 | 65 |
| Koh Kong | Khemara Phoumin | 1·080 | 1·066 | 3·651 | 0·426 | 1·347 | 372 | 148 | 461 |
| Koh Kong | Kiri Sakor | 1·213 | 1·275 | 4·608 | 0·434 | 1·513 | 46 | 17 | 59 |
| Koh Kong | Mondol Seima | 1·044 | 0·981 | 3·523 | 0·430 | 1·317 | 203 | 81 | 258 |
| Koh Kong | Srae Ambel | 0·956 | 0·980 | 3·485 | 0·360 | 1·192 | 460 | 170 | 581 |
| Koh Kong | Thma Bang | 0·899 | 0·875 | 3·108 | 0·362 | 1·122 | 72 | 29 | 98 |
| Kratie | Chetr Borei | 0·631 | 0·703 | 2·387 | 0·221 | 0·781 | 437 | 155 | 542 |
| Kratie | Chhloung | 0·686 | 0·775 | 2·685 | 0·240 | 0·837 | 289 | 98 | 356 |
| Kratie | Kracheh | 0·615 | 0·657 | 2·252 | 0·220 | 0·766 | 205 | 73 | 252 |
| Kratie | Prek Prasab | 0·592 | 0·635 | 2·207 | 0·220 | 0·732 | 779 | 291 | 957 |
| Kratie | Sambour | 0·562 | 0·598 | 2·080 | 0·208 | 0·694 | 440 | 157 | 544 |
| Kratie | Snuol | 0·778 | 0·964 | 3·240 | 0·246 | 0·935 | 919 | 290 | 1093 |
| Mondulkiri | Kaev Seima | 0·801 | 1·008 | 3·382 | 0·250 | 0·957 | 287 | 86 | 338 |
| Mondulkiri | Kaoh Nheaek | 0·780 | 0·944 | 3·217 | 0·245 | 0·950 | 211 | 61 | 255 |
| Mondulkiri | Ou Reang | 0·991 | 1·304 | 4·487 | 0·279 | 1·164 | 103 | 27 | 121 |
| Mondulkiri | Pech Chreada | 0·937 | 1·221 | 4·163 | 0·267 | 1·114 | 229 | 61 | 275 |
| Mondulkiri | Saen Monourom | 0·956 | 1·248 | 4·385 | 0·273 | 1·133 | 151 | 41 | 179 |
| Oddar Meanchey | Anlong Veaeng | 0·635 | 0·527 | 1·905 | 0·299 | 0·794 | 425 | 200 | 528 |
| Oddar Meanchey | Banteay Ampil | 0·925 | 0·777 | 2·787 | 0·427 | 1·164 | 572 | 265 | 720 |
| Oddar Meanchey | Chong Kal | 0·753 | 0·625 | 2·273 | 0·353 | 0·941 | 320 | 151 | 403 |
| Oddar Meanchey | Samraong | 0·776 | 0·649 | 2·331 | 0·359 | 0·977 | 531 | 243 | 666 |
| Oddar Meanchey | Trapeang Prasat | 0·545 | 0·444 | 1·603 | 0·262 | 0·682 | 405 | 196 | 504 |
| Pailin | Pailin | 1·070 | 0·902 | 3·200 | 0·494 | 1·353 | 457 | 216 | 579 |
| Pailin | Sala Krau | 1·068 | 0·843 | 3·037 | 0·517 | 1·352 | 526 | 256 | 664 |
| Phnom Penh | Chamkar Mon | 0·660 | 0·651 | 2·212 | 0·266 | 0·822 | 7806 | 3138 | 9720 |
| Phnom Penh | Chbar Ampov | 0·682 | 0·685 | 2·436 | 0·276 | 0·825 | 572 | 230 | 688 |
| Phnom Penh | Chraoy Chongvar | 0·654 | 0·718 | 2·232 | 0·263 | 0·792 | 1298 | 525 | 1582 |
| Phnom Penh | Dangkao | 0·690 | 0·713 | 2·384 | 0·276 | 0·860 | 1977 | 788 | 2473 |
| Phnom Penh | Doun Penh | 0·644 | 0·644 | 2·169 | 0·263 | 0·812 | 3037 | 1241 | 3827 |
| Phnom Penh | Mean Chey | 0·682 | 0·689 | 2·086 | 0·280 | 0·853 | 1608 | 660 | 2010 |
| Phnom Penh | Praek Pnov | 0·648 | 0·610 | 2·234 | 0·275 | 0·792 | 816 | 349 | 998 |
| Phnom Penh | Prampir Meakkakra | 0·649 | 0·632 | 2·276 | 0·259 | 0·807 | 6198 | 2478 | 7710 |
| Phnom Penh | Pur SenChey | 0·677 | 0·667 | 2·241 | 0·274 | 0·848 | 1860 | 746 | 2338 |
| Phnom Penh | Russey Keo | 0·681 | 0·809 | 2·361 | 0·258 | 0·822 | 1123 | 425 | 1355 |
| Phnom Penh | Saensokh | 0·653 | 0·635 | 2·423 | 0·261 | 0·798 | 4576 | 1803 | 5626 |
| Phnom Penh | Tuol Kouk | 0·653 | 0·620 | 2·382 | 0·255 | 0·802 | 3161 | 1237 | 3883 |
| Preah Sihanouk | Kampong Seila | 0·974 | 1·028 | 3·626 | 0·359 | 1·212 | 193 | 71 | 242 |
| Preah Sihanouk | Preah Sihanouk | 1·381 | 1·580 | 5·579 | 0·447 | 1·696 | 914 | 288 | 1111 |
| Preah Sihanouk | Prey Nob | 1·255 | 1·416 | 4·999 | 0·414 | 1·556 | 1590 | 519 | 1959 |
| Preah Sihanouk | Stueng Hav | 1·220 | 1·379 | 4·750 | 0·408 | 1·503 | 285 | 94 | 357 |
| Preah Vihear | Chey Saen | 0·430 | 0·374 | 1·335 | 0·192 | 0·541 | 123 | 51 | 153 |
| Preah Vihear | Chhaeb | 0·441 | 0·382 | 1·373 | 0·199 | 0·553 | 114 | 47 | 139 |
| Preah Vihear | Choam Ksant | 0·458 | 0·369 | 1·342 | 0·221 | 0·576 | 293 | 138 | 363 |
| Preah Vihear | Kuleaen | 0·471 | 0·383 | 1·376 | 0·226 | 0·588 | 191 | 89 | 240 |
| Preah Vihear | Preah Vihear | 0·437 | 0·355 | 1·326 | 0·206 | 0·552 | 116 | 55 | 149 |
| Preah Vihear | Rovieng | 0·443 | 0·383 | 1·366 | 0·203 | 0·553 | 255 | 108 | 314 |
| Preah Vihear | Sangkum Thmei | 0·453 | 0·375 | 1·359 | 0·213 | 0·568 | 118 | 56 | 148 |
| Preah Vihear | Tbaeng Mean Chey | 0·431 | 0·352 | 1·309 | 0·204 | 0·543 | 78 | 37 | 99 |
| Prey Veng | Ba Phnum | 0·784 | 0·927 | 3·097 | 0·279 | 0·955 | 745 | 265 | 909 |
| Prey Veng | Kamchay Mear | 0·721 | 0·805 | 2·639 | 0·263 | 0·877 | 657 | 240 | 799 |
| Prey Veng | Kampong Trabaek | 0·880 | 1·050 | 3·498 | 0·300 | 1·070 | 1259 | 430 | 1532 |
| Prey Veng | Kanhchriech | 0·680 | 0·772 | 2·479 | 0·258 | 0·829 | 567 | 214 | 689 |
| Prey Veng | Me Sang | 0·793 | 0·926 | 3·178 | 0·272 | 0·958 | 1055 | 361 | 1274 |
| Prey Veng | Pea Reang | 0·650 | 0·683 | 2·277 | 0·256 | 0·799 | 855 | 338 | 1053 |
| Prey Veng | Peam Chor | 0·863 | 0·962 | 3·256 | 0·302 | 1·062 | 992 | 346 | 1218 |
| Prey Veng | Peam Ro | 0·752 | 0·839 | 2·807 | 0·270 | 0·915 | 514 | 192 | 621 |

| ADM1 | ADM2 | β-thalassaemia carrier prevalence Mean | SD | CI95% | Q25 | Q75 | Estimated carrier counts Mean | Q25 | Q75 |
|---|---|---|---|---|---|---|---|---|---|
| Prey Veng | Preah Sdach | 0·854 | 0·971 | 3·329 | 0·304 | 1·041 | 1262 | 452 | 1544 |
| Prey Veng | Prey Veng | 0·707 | 0·778 | 2·505 | 0·257 | 0·877 | 309 | 114 | 378 |
| Prey Veng | Pur Rieng | 0·679 | 0·698 | 2·428 | 0·262 | 0·837 | 323 | 124 | 399 |
| Prey Veng | Sithor Kandal | 0·642 | 0·727 | 2·343 | 0·246 | 0·774 | 403 | 154 | 487 |
| Prey Veng | Svay Antor | 0·713 | 0·811 | 2·662 | 0·261 | 0·874 | 979 | 360 | 1196 |
| Pursat | Bakan | 0·676 | 0·558 | 2·004 | 0·319 | 0·849 | 1023 | 479 | 1289 |
| Pursat | Kandieng | 0·628 | 0·532 | 1·907 | 0·292 | 0·786 | 508 | 234 | 636 |
| Pursat | Krakor | 0·614 | 0·540 | 1·939 | 0·275 | 0·766 | 625 | 279 | 781 |
| Pursat | Phnum Kravanh | 0·704 | 0·626 | 2·227 | 0·310 | 0·882 | 521 | 233 | 657 |
| Pursat | Pursat | 0·652 | 0·547 | 2·020 | 0·299 | 0·819 | 324 | 152 | 409 |
| Pursat | Veal Veaeng | 0·904 | 0·783 | 2·823 | 0·406 | 1·140 | 269 | 124 | 341 |
| Ratanakiri Province | Andoung Meas | 0·916 | 1·100 | 3·858 | 0·276 | 1·127 | 198 | 59 | 244 |
| Ratanakiri Province | Ban Lung | 0·786 | 0·927 | 3·224 | 0·253 | 0·955 | 303 | 100 | 379 |
| Ratanakiri Province | Bar Kaev | 0·850 | 1·010 | 3·515 | 0·267 | 1·033 | 284 | 87 | 343 |
| Ratanakiri Province | Koun Mom | 0·700 | 0·835 | 2·825 | 0·231 | 0·848 | 242 | 78 | 297 |
| Ratanakiri Province | Lumphat | 0·811 | 0·993 | 3·332 | 0·255 | 0·979 | 278 | 85 | 337 |
| Ratanakiri Province | Ou Chum | 0·790 | 0·939 | 3·204 | 0·258 | 0·965 | 217 | 71 | 267 |
| Ratanakiri Province | Ou Ya Dav | 0·964 | 1·196 | 4·186 | 0·281 | 1·170 | 266 | 77 | 325 |
| Ratanakiri Province | Ta Veaeng | 0·912 | 1·090 | 3·745 | 0·286 | 1·115 | 77 | 24 | 90 |
| Ratanakiri Province | Veun Sai | 0·739 | 0·850 | 2·960 | 0·246 | 0·902 | 207 | 66 | 250 |
| Siem Reap | Angkor Chum | 0·692 | 0·557 | 2·016 | 0·333 | 0·871 | 432 | 207 | 542 |
| Siem Reap | Angkor Thum | 0·644 | 0·518 | 1·904 | 0·312 | 0·801 | 220 | 105 | 274 |
| Siem Reap | Banteay Srei | 0·596 | 0·475 | 1·722 | 0·291 | 0·743 | 321 | 155 | 398 |
| Siem Reap | Chi Kraeng | 0·527 | 0·430 | 1·548 | 0·251 | 0·663 | 780 | 372 | 983 |
| Siem Reap | Kralanh | 0·750 | 0·578 | 2·107 | 0·370 | 0·951 | 406 | 203 | 515 |
| Siem Reap | Prasat Bakong | 0·604 | 0·484 | 1·782 | 0·286 | 0·766 | 427 | 199 | 545 |
| Siem Reap | Puok | 0·702 | 0·564 | 2·006 | 0·338 | 0·886 | 1082 | 516 | 1358 |
| Siem Reap | Siem Reap | 0·641 | 0·525 | 1·894 | 0·306 | 0·803 | 1901 | 914 | 2398 |
| Siem Reap | Soutr Nikom | 0·570 | 0·459 | 1·686 | 0·275 | 0·708 | 809 | 391 | 1005 |
| Siem Reap | Srei Snam | 0·754 | 0·618 | 2·254 | 0·358 | 0·948 | 319 | 151 | 399 |
| Siem Reap | Svay Leu | 0·534 | 0·426 | 1·535 | 0·259 | 0·674 | 267 | 133 | 338 |
| Siem Reap | Varin | 0·631 | 0·504 | 1·825 | 0·307 | 0·792 | 301 | 149 | 380 |
| Stung Treng | Sesan | 0·590 | 0·642 | 2·219 | 0·217 | 0·723 | 158 | 56 | 195 |
| Stung Treng | Siem Bouk | 0·490 | 0·482 | 1·677 | 0·198 | 0·605 | 172 | 66 | 214 |
| Stung Treng | Siem Pang | 0·628 | 0·686 | 2·339 | 0·229 | 0·771 | 184 | 69 | 228 |
| Stung Treng | Stueng Traeng | 0·523 | 0·513 | 1·832 | 0·206 | 0·651 | 188 | 77 | 239 |
| Stung Treng | Thala Barivat | 0·473 | 0·434 | 1·569 | 0·202 | 0·594 | 267 | 112 | 339 |
| Svay Rieng | Bavet | 1·011 | 1·315 | 4·304 | 0·304 | 1·201 | 566 | 171 | 668 |
| Svay Rieng | Chantrea | 1·098 | 1·504 | 5·061 | 0·313 | 1·269 | 378 | 108 | 441 |
| Svay Rieng | Kampong Rou | 1·044 | 1·410 | 4·545 | 0·309 | 1·209 | 870 | 260 | 1006 |
| Svay Rieng | Romeas Haek | 0·815 | 1·004 | 3·257 | 0·276 | 0·979 | 988 | 334 | 1186 |
| Svay Rieng | Rumduol | 0·899 | 1·168 | 3·693 | 0·288 | 1·064 | 562 | 180 | 664 |
| Svay Rieng | Svay Chrum | 0·906 | 1·086 | 3·671 | 0·300 | 1·104 | 1135 | 376 | 1384 |
| Svay Rieng | Svay Rieng | 0·927 | 1·251 | 3·933 | 0·279 | 1·110 | 364 | 109 | 436 |
| Svay Rieng | Svay Teab | 0·965 | 1·311 | 4·071 | 0·299 | 1·124 | 796 | 247 | 923 |
| Takeo | Angkor Borei | 0·881 | 0·953 | 3·343 | 0·321 | 1·083 | 434 | 159 | 538 |
| Takeo | Bati | 0·762 | 0·786 | 2·673 | 0·300 | 0·944 | 1744 | 687 | 2159 |
| Takeo | Borei Cholsar | 0·977 | 1·102 | 3·977 | 0·331 | 1·195 | 200 | 70 | 244 |
| Takeo | Doun Kaev | 0·876 | 0·973 | 3·220 | 0·323 | 1·077 | 478 | 178 | 591 |
| Takeo | Kaoh Andaet | 0·984 | 1·083 | 3·870 | 0·336 | 1·209 | 497 | 170 | 614 |
| Takeo | Kiri Vong | 1·045 | 1·185 | 4·104 | 0·356 | 1·289 | 1120 | 385 | 1394 |
| Takeo | Prey Kabbas | 0·808 | 0·860 | 3·088 | 0·306 | 0·991 | 878 | 333 | 1083 |
| Takeo | Samraong | 0·811 | 0·835 | 2·917 | 0·306 | 1·013 | 1142 | 431 | 1424 |
| Takeo | Tram Kak | 0·888 | 0·920 | 3·262 | 0·326 | 1·108 | 1742 | 633 | 2170 |
| Takeo | Treang | 0·931 | 1·019 | 3·526 | 0·332 | 1·152 | 1225 | 438 | 1517 |
| Tbong Khmum | Dambae | 0·666 | 0·732 | 2·520 | 0·242 | 0·820 | 664 | 243 | 825 |
| Tbong Khmum | Krouch Chhmar | 0·610 | 0·645 | 2·229 | 0·233 | 0·755 | 533 | 206 | 663 |

| ADM1 | ADM2 | β-thalassaemia carrier prevalence Mean | SD | CI95% | Q25 | Q75 | Estimated carrier counts Mean | Q25 | Q75 |
|---|---|---|---|---|---|---|---|---|---|
| Tbong Khmum | Memot | 0·776 | 0·923 | 3·179 | 0·256 | 0·937 | 1313 | 434 | 1583 |
| Tbong Khmum | Ou Reang Ov | 0·660 | 0·716 | 2·393 | 0·250 | 0·808 | 579 | 221 | 715 |
| Tbong Khmum | Ponhea Kraek | 0·729 | 0·846 | 2·867 | 0·253 | 0·884 | 1330 | 467 | 1616 |
| Tbong Khmum | Suong | 0·653 | 0·717 | 2·482 | 0·240 | 0·798 | 333 | 119 | 409 |
| Tbong Khmum | Tboung Khmum | 0·635 | 0·678 | 2·301 | 0·239 | 0·783 | 1146 | 433 | 1408 |

ADM1: administrative level 1. ADM2: administrative level 2. SD: standard deviation. CI95%: 95% credible interval. Q25: 25th percentile. Q75: 75th percentile. SD and CI95% are measures of prediction uncertainty.

**Supplementary Table S19. Subnational areal predictions of β-thalassaemia carrier prevalence and estimated carrier count at administrative level 2 of China.**

| ADM1 | ADM2 | β-thalassaemia carrier prevalence Mean | SD | CI95% | Q25 | Q75 | Estimated carrier counts Mean | Q25 | Q75 |
|---|---|---|---|---|---|---|---|---|---|
| Anhui Province | Anqingshi | 1·168 | 1·353 | 4·519 | 0·409 | 1·427 | 8503 | 2965 | 10303 |
| Anhui Province | Bangbushi | 1·019 | 1·369 | 4·583 | 0·267 | 1·220 | 11920 | 3131 | 14330 |
| Anhui Province | Bozhoushi | 1·830 | 2·493 | 8·456 | 0·452 | 2·198 | 30559 | 7559 | 36636 |
| Anhui Province | Changfeng xian | 1·055 | 1·420 | 4·672 | 0·297 | 1·247 | 11287 | 3210 | 13498 |
| Anhui Province | Chaohushi | 0·921 | 1·169 | 3·695 | 0·289 | 1·113 | 224 | 71 | 273 |
| Anhui Province | Chaohushi | 0·832 | 1·060 | 3·480 | 0·256 | 0·997 | 4430 | 1381 | 5204 |
| Anhui Province | Chuzhoushi | 0·721 | 0·949 | 3·216 | 0·210 | 0·841 | 5032 | 1483 | 5899 |
| Anhui Province | Dafengshi | 1·004 | 1·442 | 4·780 | 0·252 | 1·164 | 8164 | 2040 | 9410 |
| Anhui Province | Dangshan xian | 1·744 | 2·595 | 8·202 | 0·386 | 2·046 | 13506 | 2965 | 15842 |
| Anhui Province | Dangtu xian | 0·577 | 0·696 | 2·301 | 0·192 | 0·698 | 4615 | 1539 | 5599 |
| Anhui Province | Dingyuan xian | 0·895 | 1·194 | 4·014 | 0·251 | 1·055 | 6131 | 1732 | 7232 |
| Anhui Province | Dongzhi xian | 1·340 | 1·439 | 5·067 | 0·490 | 1·643 | 4958 | 1788 | 6034 |
| Anhui Province | Fanchang xian | 0·719 | 0·840 | 2·910 | 0·238 | 0·879 | 2763 | 920 | 3364 |
| Anhui Province | Feidong xian | 0·893 | 1·151 | 3·848 | 0·264 | 1·061 | 12983 | 3868 | 15264 |
| Anhui Province | Feixi xian | 1·097 | 1·414 | 4·677 | 0·333 | 1·304 | 18393 | 5579 | 21731 |
| Anhui Province | Feng xian | 1·748 | 2·622 | 8·493 | 0·383 | 2·018 | 17131 | 3736 | 19800 |
| Anhui Province | Fengtai xian | 1·335 | 1·743 | 5·988 | 0·359 | 1·603 | 7738 | 2085 | 9278 |
| Anhui Province | Fengyang xian | 0·949 | 1·302 | 4·402 | 0·251 | 1·110 | 6607 | 1721 | 7652 |
| Anhui Province | Funan xian | 1·699 | 2·111 | 7·310 | 0·498 | 2·049 | 24308 | 7027 | 29279 |
| Anhui Province | Fuyangshi | 1·663 | 2·122 | 7·399 | 0·458 | 2·047 | 7896 | 2170 | 9696 |
| Anhui Province | Fuyangshi | 1·648 | 2·092 | 7·293 | 0·457 | 1·991 | 25288 | 7075 | 30708 |
| Anhui Province | Gaochun xian | 0·506 | 0·593 | 1·969 | 0·174 | 0·610 | 2202 | 749 | 2675 |
| Anhui Province | Guangde xian | 0·497 | 0·566 | 1·949 | 0·172 | 0·601 | 2326 | 815 | 2823 |
| Anhui Province | Guichishi | 1·038 | 1·202 | 4·087 | 0·358 | 1·255 | 6348 | 2157 | 7769 |
| Anhui Province | Guzhen xian | 1·107 | 1·590 | 5·139 | 0·277 | 1·301 | 5482 | 1388 | 6407 |
| Anhui Province | Hanshan xian | 0·740 | 0·900 | 3·120 | 0·230 | 0·892 | 3406 | 1065 | 4102 |
| Anhui Province | He xian | 0·669 | 0·824 | 2·797 | 0·209 | 0·803 | 2934 | 925 | 3522 |
| Anhui Province | Hefeishi | 1·001 | 1·331 | 4·350 | 0·295 | 1·190 | 38391 | 11372 | 45922 |
| Anhui Province | Huaibeishi | 1·421 | 2·034 | 6·885 | 0·339 | 1·641 | 3124 | 743 | 3611 |

| ADM1 | ADM2 | β-thalassaemia carrier prevalence Mean | SD | CI95% | Q25 | Q75 | Estimated carrier counts Mean | Q25 | Q75 |
|---|---|---|---|---|---|---|---|---|---|
| Anhui Province | Huainanshi | 1·185 | 1·555 | 5·252 | 0·316 | 1·426 | 15701 | 4226 | 19036 |
| Anhui Province | Huaining xian | 1·278 | 1·407 | 4·885 | 0·455 | 1·567 | 7255 | 2553 | 8933 |
| Anhui Province | Huaiyuan xian | 1·183 | 1·629 | 5·392 | 0·304 | 1·403 | 11598 | 2975 | 13647 |
| Anhui Province | Huangshanshi | 0·904 | 1·067 | 3·581 | 0·306 | 1·095 | 1272 | 427 | 1527 |
| Anhui Province | Huangshanshi | 1·045 | 1·309 | 4·195 | 0·344 | 1·248 | 1982 | 660 | 2342 |
| Anhui Province | Huoqiu xian | 1·448 | 1·785 | 6·066 | 0·443 | 1·756 | 16633 | 5138 | 20190 |
| Anhui Province | Huoshan xian | 1·393 | 1·567 | 5·424 | 0·480 | 1·714 | 3892 | 1322 | 4792 |
| Anhui Province | Jiangpu xian | 0·613 | 0·759 | 2·652 | 0·190 | 0·730 | 3335 | 1048 | 3931 |
| Anhui Province | Jiashan xian | 0·779 | 1·035 | 3·522 | 0·215 | 0·919 | 3975 | 1087 | 4664 |
| Anhui Province | Jing xian | 0·752 | 0·893 | 3·004 | 0·251 | 0·912 | 2018 | 686 | 2504 |
| Anhui Province | Jingde xian | 0·796 | 0·950 | 3·247 | 0·263 | 0·958 | 847 | 279 | 1030 |
| Anhui Province | Jinzai xian | 1·565 | 1·697 | 5·963 | 0·548 | 1·942 | 8025 | 2792 | 9959 |
| Anhui Province | Jixi xian | 0·796 | 0·966 | 3·177 | 0·265 | 0·958 | 1126 | 373 | 1356 |
| Anhui Province | Laian xian | 0·632 | 0·800 | 2·782 | 0·188 | 0·750 | 3061 | 914 | 3612 |
| Anhui Province | Langxi xian | 0·490 | 0·560 | 1·888 | 0·173 | 0·594 | 1406 | 491 | 1704 |
| Anhui Province | Lingbi xian | 1·135 | 1·649 | 5·419 | 0·278 | 1·315 | 10264 | 2528 | 11854 |
| Anhui Province | Liuanshi | 1·312 | 1·627 | 5·422 | 0·414 | 1·581 | 22690 | 7156 | 27381 |
| Anhui Province | Lixing xian | 1·559 | 2·052 | 7·066 | 0·405 | 1·883 | 18555 | 4799 | 22282 |
| Anhui Province | Lujiang xian | 0·990 | 1·211 | 4·020 | 0·318 | 1·197 | 9131 | 2896 | 11016 |
| Anhui Province | Maanshanshi | 0·600 | 0·736 | 2·470 | 0·197 | 0·707 | 3893 | 1275 | 4558 |
| Anhui Province | Mengcheng xian | 1·378 | 1·887 | 6·280 | 0·357 | 1·638 | 15333 | 3954 | 18042 |
| Anhui Province | Nanling xian | 0·727 | 0·848 | 2·931 | 0·242 | 0·887 | 3102 | 1008 | 3781 |
| Anhui Province | Ningguoshi | 0·637 | 0·744 | 2·495 | 0·217 | 0·777 | 2297 | 787 | 2810 |
| Anhui Province | Qianshan xian | 1·333 | 1·489 | 5·166 | 0·463 | 1·631 | 5677 | 1994 | 6968 |
| Anhui Province | Qimen xian | 1·176 | 1·306 | 4·522 | 0·418 | 1·437 | 1651 | 586 | 2039 |
| Anhui Province | Qingyang xian | 0·894 | 1·042 | 3·508 | 0·303 | 1·087 | 2113 | 710 | 2604 |
| Anhui Province | Quanjiao xian | 0·729 | 0·944 | 3·169 | 0·221 | 0·851 | 2825 | 867 | 3252 |
| Anhui Province | She xian | 0·914 | 1·112 | 3·634 | 0·306 | 1·104 | 5760 | 1931 | 6949 |
| Anhui Province | Shitai xian | 1·096 | 1·246 | 4·277 | 0·388 | 1·330 | 938 | 331 | 1131 |
| Anhui Province | Shou xian | 1·198 | 1·561 | 5·221 | 0·348 | 1·431 | 10198 | 2922 | 12149 |
| Anhui Province | Shucheng xian | 1·182 | 1·446 | 4·826 | 0·383 | 1·428 | 8257 | 2541 | 9777 |
| Anhui Province | Shuining xian | 1·113 | 1·649 | 5·503 | 0·267 | 1·269 | 11344 | 2737 | 13027 |
| Anhui Province | Shuzhoushi | 1·270 | 1·838 | 6·059 | 0·307 | 1·480 | 13742 | 3324 | 16036 |
| Anhui Province | Suixi xian | 1·415 | 2·016 | 6·748 | 0·344 | 1·661 | 23899 | 5711 | 27895 |
| Anhui Province | Susong xian | 1·571 | 1·564 | 5·573 | 0·613 | 1·957 | 10309 | 4046 | 12864 |
| Anhui Province | Suzhoushi | 1·325 | 1·987 | 6·444 | 0·321 | 1·517 | 8102 | 1981 | 9385 |

| | | β-thalassaemia carrier prevalence | | | | | Estimated carrier counts | | |
|---|---|---|---|---|---|---|---|---|---|
| ADM1 | ADM2 | Mean | SD | CI95% | Q25 | Q75 | Mean | Q25 | Q75 |
| Anhui Province | Taihe xian | 1·766 | 2·300 | 7·888 | 0·469 | 2·150 | 22512 | 6020 | 27866 |
| Anhui Province | Taihu xian | 1·521 | 1·572 | 5·549 | 0·564 | 1·893 | 6820 | 2532 | 8384 |
| Anhui Province | Tongcheng xian | 1·161 | 1·373 | 4·600 | 0·390 | 1·409 | 6735 | 2264 | 8105 |
| Anhui Province | Tongling xian | 0·830 | 0·995 | 3·300 | 0·277 | 1·000 | 5879 | 1953 | 7077 |
| Anhui Province | Tonglingshi | 0·867 | 0·969 | 3·263 | 0·293 | 1·060 | 904 | 319 | 1099 |
| Anhui Province | Wangjiang xian | 1·393 | 1·524 | 5·272 | 0·512 | 1·702 | 5925 | 2167 | 7242 |
| Anhui Province | Woyang xian | 1·605 | 2·203 | 7·519 | 0·397 | 1·917 | 19003 | 4708 | 22901 |
| Anhui Province | Wuhe xian | 0·958 | 1·338 | 4·478 | 0·247 | 1·113 | 4791 | 1234 | 5612 |
| Anhui Province | Wuhu xian | 0·616 | 0·714 | 2·417 | 0·210 | 0·756 | 6326 | 2178 | 7831 |
| Anhui Province | Wuhushi | 0·642 | 0·723 | 2·532 | 0·212 | 0·796 | 4073 | 1349 | 5036 |
| Anhui Province | Wuwei xian | 0·814 | 0·987 | 3·326 | 0·263 | 0·991 | 6979 | 2226 | 8416 |
| Anhui Province | Xiao xian | 1·490 | 2·164 | 7·110 | 0·350 | 1·739 | 15913 | 3718 | 18544 |
| Anhui Province | Xiayi xian | 1·800 | 2·599 | 8·484 | 0·406 | 2·128 | 16865 | 3828 | 19916 |
| Anhui Province | Xiuning xian | 1·101 | 1·280 | 4·369 | 0·376 | 1·335 | 1731 | 576 | 2070 |
| Anhui Province | Xuanzhoushi | 0·593 | 0·685 | 2·333 | 0·202 | 0·728 | 4608 | 1584 | 5652 |
| Anhui Province | Yi xian | 1·025 | 1·192 | 4·010 | 0·353 | 1·237 | 728 | 244 | 890 |
| Anhui Province | Yingshang xian | 1·421 | 1·804 | 6·127 | 0·406 | 1·721 | 17088 | 4928 | 20882 |
| Anhui Province | Yongcheng xian | 1·654 | 2·350 | 7·795 | 0·392 | 1·949 | 19778 | 4746 | 23060 |
| Anhui Province | Yuexi xian | 1·436 | 1·545 | 5·381 | 0·514 | 1·779 | 4503 | 1592 | 5545 |
| Anhui Province | Zongyang xian | 1·022 | 1·207 | 4·064 | 0·342 | 1·237 | 5278 | 1784 | 6391 |
| Beijing Municipality | Beijingshi | 3·582 | 5·751 | 19·313 | 0·630 | 4·034 | 443049 | 77456 | 502803 |
| Beijing Municipality | Changpingxin | 3·682 | 5·851 | 20·271 | 0·640 | 4·129 | 91664 | 15976 | 101856 |
| Beijing Municipality | Dachanghui zuzizhi xian | 3·344 | 5·129 | 18·411 | 0·612 | 3·851 | 3850 | 703 | 4433 |
| Beijing Municipality | Huairou xian | 3·808 | 6·105 | 21·094 | 0·652 | 4·241 | 16742 | 2884 | 18765 |
| Beijing Municipality | Miyun xian | 3·691 | 5·917 | 20·235 | 0·643 | 4·131 | 19310 | 3323 | 21551 |
| Beijing Municipality | Pinggu xian | 3·448 | 5·373 | 18·440 | 0·607 | 3·914 | 15284 | 2653 | 17289 |
| Beijing Municipality | Shanheshi | 3·365 | 5·283 | 17·868 | 0·588 | 3·867 | 34978 | 5980 | 40150 |
| Beijing Municipality | Shunyi xian | 3·556 | 5·711 | 19·447 | 0·616 | 4·017 | 61057 | 10573 | 68545 |
| Beijing Municipality | Tong xian | 3·394 | 5·380 | 18·202 | 0·595 | 3·860 | 74240 | 12995 | 84041 |
| Beijing Municipality | Yanqing xian | 3·808 | 6·107 | 21·101 | 0·651 | 4·221 | 13796 | 2378 | 15587 |
| Chongqing Municipality | Ba xian | 1·387 | 1·416 | 4·985 | 0·545 | 1·708 | 30140 | 11850 | 37195 |
| Chongqing Municipality | Bishan xian | 1·282 | 1·280 | 4·453 | 0·521 | 1·587 | 10011 | 4084 | 12417 |
| Chongqing Municipality | Changshou xian | 1·325 | 1·286 | 4·652 | 0·541 | 1·645 | 8838 | 3608 | 10938 |
| Chongqing Municipality | Congqingshi | 1·309 | 1·306 | 4·597 | 0·522 | 1·624 | 85294 | 33535 | 105367 |
| Chongqing Municipality | Dazhu xian | 1·193 | 1·127 | 3·945 | 0·505 | 1·478 | 9712 | 4102 | 12103 |
| Chongqing Municipality | Dianjiang xian | 1·332 | 1·239 | 4·413 | 0·559 | 1·667 | 8615 | 3674 | 10764 |

| | | β-thalassaemia carrier prevalence | | | | | Estimated carrier counts | | |
|---|---|---|---|---|---|---|---|---|---|
| ADM1 | ADM2 | Mean | SD | CI95% | Q25 | Q75 | Mean | Q25 | Q75 |
| Chongqing Municipality | Fengdu xian | 1·486 | 1·472 | 5·217 | 0·595 | 1·834 | 7996 | 3194 | 9827 |
| Chongqing Municipality | Fengjie xian | 1·872 | 1·661 | 5·975 | 0·817 | 2·360 | 12913 | 5732 | 16356 |
| Chongqing Municipality | Fulingshi | 1·418 | 1·410 | 5·049 | 0·563 | 1·752 | 15768 | 6350 | 19479 |
| Chongqing Municipality | Hechuanshi | 1·186 | 1·101 | 3·916 | 0·510 | 1·474 | 14873 | 6285 | 18474 |
| Chongqing Municipality | Jiangjinshi | 1·447 | 1·518 | 5·185 | 0·562 | 1·773 | 17449 | 6895 | 21477 |
| Chongqing Municipality | Jingbei xian | 1·281 | 1·226 | 4·401 | 0·529 | 1·594 | 25867 | 10418 | 32491 |
| Chongqing Municipality | Kai xian | 1·631 | 1·552 | 5·472 | 0·675 | 2·039 | 19203 | 8047 | 24050 |
| Chongqing Municipality | Laifeng xian | 1·612 | 1·348 | 4·892 | 0·736 | 2·049 | 4763 | 2139 | 6032 |
| Chongqing Municipality | Liangping xian | 1·376 | 1·273 | 4·512 | 0·588 | 1·720 | 9150 | 3903 | 11437 |
| Chongqing Municipality | Nanchuan xian | 1·552 | 1·587 | 5·546 | 0·597 | 1·930 | 8551 | 3375 | 10743 |
| Chongqing Municipality | Pengshuimiaozutujia zuzizhi xian | 1·594 | 1·527 | 5·501 | 0·645 | 1·991 | 8542 | 3430 | 10618 |
| Chongqing Municipality | Qianjiangtujiazumiao zuzizhi xian | 1·594 | 1·411 | 5·134 | 0·687 | 2·022 | 7595 | 3267 | 9559 |
| Chongqing Municipality | Qiuyangtujiazumiao zuzizhi xian | 1·665 | 1·473 | 5·340 | 0·722 | 2·105 | 13160 | 5644 | 16484 |
| Chongqing Municipality | Shizhutujia zuzizhi xian | 1·549 | 1·470 | 5·255 | 0·638 | 1·937 | 5966 | 2428 | 7489 |
| Chongqing Municipality | Tongliang xian | 1·225 | 1·196 | 4·115 | 0·515 | 1·519 | 8417 | 3551 | 10422 |
| Chongqing Municipality | Tongnan xian | 1·142 | 1·040 | 3·656 | 0·505 | 1·421 | 7757 | 3496 | 9648 |
| Chongqing Municipality | Wanshengqu | 1·598 | 1·667 | 5·957 | 0·600 | 1·985 | 4221 | 1618 | 5279 |
| Chongqing Municipality | Wanxianshi | 1·584 | 1·468 | 5·289 | 0·665 | 1·992 | 25118 | 10589 | 31707 |
| Chongqing Municipality | Wulong xian | 1·545 | 1·558 | 5·527 | 0·602 | 1·920 | 5454 | 2142 | 6852 |
| Chongqing Municipality | Wushan xian | 2·025 | 1·682 | 6·131 | 0·940 | 2·554 | 8927 | 4172 | 11182 |
| Chongqing Municipality | Wusheng xian | 1·139 | 1·004 | 3·651 | 0·507 | 1·416 | 6113 | 2724 | 7604 |
| Chongqing Municipality | Wuxi xian | 2·033 | 1·978 | 6·962 | 0·836 | 2·524 | 7784 | 3300 | 9719 |
| Chongqing Municipality | Xianfeng xian | 1·608 | 1·400 | 5·088 | 0·708 | 2·033 | 5344 | 2360 | 6683 |
| Chongqing Municipality | Yongchuanshi | 1·288 | 1·278 | 4·486 | 0·521 | 1·595 | 13589 | 5515 | 16791 |
| Chongqing Municipality | Yunyang xian | 1·751 | 1·626 | 5·805 | 0·736 | 2·197 | 16197 | 6813 | 20098 |
| Chongqing Municipality | Zhenping xian | 2·175 | 2·243 | 7·690 | 0·834 | 2·711 | 983 | 384 | 1215 |
| Chongqing Municipality | Zhong xian | 1·460 | 1·383 | 4·964 | 0·608 | 1·821 | 9722 | 4037 | 12084 |
| Chongqing Municipality | Zhuanjiang xian | 1·568 | 1·649 | 5·751 | 0·594 | 1·935 | 11466 | 4337 | 14037 |
| Fujian Province | Anxi xian | 3·437 | 2·535 | 9·402 | 1·746 | 4·352 | 31168 | 15861 | 39384 |
| Fujian Province | Changding xian | 4·728 | 3·239 | 12·109 | 2·502 | 6·003 | 17738 | 9269 | 22410 |
| Fujian Province | Changleshi | 2·366 | 2·105 | 7·505 | 1·017 | 2·995 | 19051 | 8124 | 23935 |
| Fujian Province | Changtai xian | 3·889 | 2·745 | 10·176 | 2·015 | 4·950 | 8234 | 4239 | 10408 |
| Fujian Province | Datian xian | 3·039 | 2·323 | 8·488 | 1·498 | 3·843 | 9036 | 4464 | 11500 |
| Fujian Province | Dehua xian | 2·796 | 2·205 | 8·006 | 1·353 | 3·518 | 8806 | 4221 | 11141 |
| Fujian Province | Dongshan xian | 4·871 | 3·784 | 13·641 | 2·376 | 6·151 | 1218 | 594 | 1538 |
| Fujian Province | Fuanshi | 1·846 | 1·741 | 6·207 | 0·765 | 2·311 | 10354 | 4327 | 12986 |

| | | β-thalassaemia carrier prevalence | | | | | Estimated carrier counts | | |
|---|---|---|---|---|---|---|---|---|---|
| ADM1 | ADM2 | Mean | SD | CI95% | Q25 | Q75 | Mean | Q25 | Q75 |
| Fujian Province | Fudingshi | 1·741 | 1·850 | 6·539 | 0·638 | 2·147 | 8809 | 3174 | 10714 |
| Fujian Province | Fuqingshi | 2·449 | 2·096 | 7·648 | 1·089 | 3·104 | 29596 | 13075 | 37342 |
| Fujian Province | Fuzhoushi | 2·314 | 1·986 | 7·179 | 1·052 | 2·898 | 79858 | 36053 | 99112 |
| Fujian Province | Guangze xian | 2·225 | 2·089 | 7·508 | 0·914 | 2·796 | 3162 | 1280 | 4017 |
| Fujian Province | Gutian xian | 2·193 | 1·777 | 6·437 | 1·046 | 2·763 | 7412 | 3657 | 9322 |
| Fujian Province | Huaan xian | 4·302 | 3·025 | 11·264 | 2·248 | 5·478 | 5439 | 2853 | 6944 |
| Fujian Province | Huian xian | 2·780 | 2·266 | 8·292 | 1·282 | 3·530 | 27358 | 12493 | 34751 |
| Fujian Province | Jaioushi | 2·117 | 1·857 | 6·710 | 0·945 | 2·650 | 9301 | 4139 | 11646 |
| Fujian Province | Jiangle xian | 2·698 | 2·316 | 8·474 | 1·201 | 3·399 | 3922 | 1732 | 4870 |
| Fujian Province | Jianyangshi | 2·109 | 2·020 | 7·262 | 0·868 | 2·621 | 7085 | 2941 | 8874 |
| Fujian Province | Jinjiangshi | 3·006 | 2·424 | 8·771 | 1·439 | 3·805 | 65556 | 31351 | 83332 |
| Fujian Province | Liancheng xian | 4·174 | 2·865 | 10·749 | 2·228 | 5·275 | 10029 | 5320 | 12662 |
| Fujian Province | Lianjiang xian | 2·192 | 1·903 | 6·960 | 0·974 | 2·749 | 11531 | 5181 | 14641 |
| Fujian Province | Longhaishi | 4·199 | 3·014 | 11·294 | 2·156 | 5·365 | 71278 | 36940 | 90996 |
| Fujian Province | Longyanshi | 4·677 | 3·169 | 11·825 | 2·521 | 5·920 | 43958 | 23367 | 55150 |
| Fujian Province | Luoyuan xian | 2·118 | 1·830 | 6·542 | 0·968 | 2·639 | 4711 | 2113 | 5828 |
| Fujian Province | Mingxi xian | 2·981 | 2·375 | 8·769 | 1·406 | 3·802 | 3270 | 1572 | 4127 |
| Fujian Province | Minhou xian | 2·307 | 1·906 | 6·948 | 1·082 | 2·893 | 17583 | 8123 | 21878 |
| Fujian Province | Minqing xian | 2·386 | 1·922 | 7·013 | 1·140 | 2·999 | 6118 | 2902 | 7685 |
| Fujian Province | Nananshi | 2·975 | 2·291 | 8·387 | 1·460 | 3·761 | 40408 | 19866 | 51146 |
| Fujian Province | Nanjing xian | 5·345 | 3·641 | 13·615 | 2·846 | 6·806 | 15824 | 8356 | 20390 |
| Fujian Province | Nanpingshi | 2·384 | 1·967 | 7·129 | 1·113 | 2·999 | 10861 | 4938 | 13762 |
| Fujian Province | Ningdeshi | 2·003 | 1·760 | 6·419 | 0·898 | 2·493 | 11386 | 4861 | 14144 |
| Fujian Province | Ninghua xian | 3·499 | 2·674 | 9·923 | 1·703 | 4·466 | 8767 | 4197 | 11212 |
| Fujian Province | Pinghe xian | 6·002 | 3·964 | 14·971 | 3·247 | 7·617 | 24681 | 13075 | 31227 |
| Fujian Province | Pingnan xian | 2·013 | 1·686 | 6·082 | 0·930 | 2·535 | 2486 | 1141 | 3088 |
| Fujian Province | Pucheng xian | 1·749 | 1·829 | 6·405 | 0·653 | 2·166 | 5054 | 1873 | 6245 |
| Fujian Province | Putian xian | 2·595 | 2·208 | 8·022 | 1·165 | 3·297 | 35305 | 15404 | 44613 |
| Fujian Province | Putianshi | 2·614 | 2·260 | 8·316 | 1·178 | 3·294 | 16251 | 7261 | 20469 |
| Fujian Province | Qingliu xian | 3·411 | 2·525 | 9·382 | 1·714 | 4·322 | 4222 | 2108 | 5375 |
| Fujian Province | Qingyuan xian | 1·710 | 1·706 | 6·033 | 0·672 | 2·136 | 2463 | 992 | 3056 |
| Fujian Province | Quanzhoushi | 2·857 | 2·302 | 8·363 | 1·349 | 3·634 | 34495 | 16278 | 44208 |
| Fujian Province | Sanmingshi | 2·852 | 2·241 | 8·183 | 1·360 | 3·631 | 11397 | 5278 | 14260 |
| Fujian Province | Sha xian | 2·651 | 2·201 | 8·046 | 1·221 | 3·360 | 6752 | 3188 | 8548 |
| Fujian Province | Shanghang xian | 6·319 | 4·057 | 15·245 | 3·508 | 8·010 | 27315 | 15278 | 34730 |
| Fujian Province | Shishishi | 3·019 | 2·643 | 9·305 | 1·354 | 3·772 | 10702 | 4796 | 13376 |

| | | β-thalassaemia carrier prevalence | | | | | Estimated carrier counts | | |
|---|---|---|---|---|---|---|---|---|---|
| ADM1 | ADM2 | Mean | SD | CI95% | Q25 | Q75 | Mean | Q25 | Q75 |
| Fujian Province | Shouning xian | 1·739 | 1·765 | 6·107 | 0·686 | 2·153 | 2912 | 1165 | 3575 |
| Fujian Province | Shuchang xian | 2·404 | 2·111 | 7·603 | 1·055 | 3·022 | 4375 | 1946 | 5581 |
| Fujian Province | Songxi xian | 1·789 | 1·742 | 6·234 | 0·719 | 2·232 | 2222 | 913 | 2794 |
| Fujian Province | Taining xian | 2·775 | 2·373 | 8·629 | 1·227 | 3·519 | 2632 | 1168 | 3279 |
| Fujian Province | Taishun xian | 1·657 | 1·784 | 6·221 | 0·605 | 2·042 | 4313 | 1597 | 5327 |
| Fujian Province | Tongan xian | 3·318 | 2·505 | 9·185 | 1·666 | 4·188 | 46218 | 22994 | 58214 |
| Fujian Province | Tuorong xian | 1·750 | 1·827 | 6·238 | 0·675 | 2·176 | 1755 | 679 | 2171 |
| Fujian Province | Wuping xian | 7·345 | 4·618 | 17·449 | 4·120 | 9·369 | 19944 | 11387 | 25253 |
| Fujian Province | Wuyishanshi | 1·973 | 1·977 | 7·136 | 0·768 | 2·442 | 5245 | 2036 | 6510 |
| Fujian Province | Xiamenshi | 3·658 | 2·732 | 10·161 | 1·833 | 4·617 | 92914 | 46115 | 116604 |
| Fujian Province | Xianyou xian | 2·696 | 2·179 | 7·921 | 1·277 | 3·406 | 19241 | 9089 | 24213 |
| Fujian Province | Xiapu xian | 1·895 | 1·873 | 6·716 | 0·750 | 2·363 | 7990 | 3112 | 9864 |
| Fujian Province | Yonganshi | 3·224 | 2·418 | 8·963 | 1·610 | 4·095 | 11158 | 5270 | 14161 |
| Fujian Province | Yongchun xian | 3·065 | 2·360 | 8·688 | 1·505 | 3·894 | 12302 | 5993 | 15496 |
| Fujian Province | Yongding xian | 6·772 | 4·253 | 16·070 | 3·801 | 8·566 | 21447 | 11910 | 27017 |
| Fujian Province | Yongtai xian | 2·530 | 2·084 | 7·479 | 1·190 | 3·189 | 7222 | 3346 | 9050 |
| Fujian Province | Youxi xian | 2·603 | 2·088 | 7·602 | 1·245 | 3·279 | 8736 | 4219 | 11048 |
| Fujian Province | Yunxiao xian | 5·358 | 3·760 | 14·085 | 2·789 | 6·804 | 24420 | 12781 | 31032 |
| Fujian Province | Zhangping xian | 3·795 | 2·697 | 10·071 | 1·973 | 4·805 | 9899 | 5187 | 12540 |
| Fujian Province | Zhangpu xian | 4·556 | 3·359 | 12·478 | 2·281 | 5·818 | 35766 | 17783 | 45548 |
| Fujian Province | Zhangzhoushi | 4·509 | 3·206 | 12·115 | 2·354 | 5·685 | 5773 | 3010 | 7268 |
| Fujian Province | Zhaowushi | 2·408 | 2·224 | 8·026 | 1·005 | 3·028 | 6613 | 2669 | 8176 |
| Fujian Province | Zhenghe xian | 1·861 | 1·739 | 6·263 | 0·777 | 2·329 | 3298 | 1398 | 4142 |
| Fujian Province | Zhouning xian | 1·890 | 1·712 | 6·129 | 0·817 | 2·367 | 2699 | 1134 | 3364 |
| Gansu Province | Akesaihashake zuzizhi xian | 5·557 | 8·437 | 29·870 | 0·957 | 6·397 | 569 | 94 | 631 |
| Gansu Province | Anxi xian | 5·083 | 7·678 | 27·177 | 0·931 | 5·799 | 6591 | 1179 | 7533 |
| Gansu Province | Baiyinshi | 3·789 | 6·057 | 20·566 | 0·714 | 4·147 | 13370 | 2537 | 14810 |
| Gansu Province | Cheng xian | 2·163 | 2·838 | 9·412 | 0·607 | 2·595 | 5534 | 1585 | 6631 |
| Gansu Province | Chongxin xian | 3·030 | 4·187 | 15·005 | 0·703 | 3·549 | 2268 | 527 | 2678 |
| Gansu Province | Diebu xian | 2·757 | 4·117 | 13·815 | 0·648 | 3·124 | 1490 | 323 | 1702 |
| Gansu Province | Dingxi xian | 3·350 | 5·231 | 17·482 | 0·697 | 3·752 | 14311 | 3051 | 15672 |
| Gansu Province | Dongxiang zuzizhi xian | 3·514 | 5·510 | 18·605 | 0·705 | 3·892 | 10814 | 2190 | 12143 |
| Gansu Province | Dunhuangshi | 5·257 | 7·974 | 28·478 | 0·952 | 6·019 | 9815 | 1737 | 11367 |
| Gansu Province | Gangu xian | 2·829 | 4·255 | 14·365 | 0·661 | 3·209 | 12789 | 3025 | 14811 |
| Gansu Province | Gaolan xian | 3·765 | 5·997 | 20·674 | 0·716 | 4·136 | 6819 | 1298 | 7436 |
| Gansu Province | Gaotai xian | 4·940 | 8·016 | 28·050 | 0·841 | 5·371 | 6253 | 1060 | 6648 |

| | | β-thalassaemia carrier prevalence | | | | | Estimated carrier counts | | |
|---|---|---|---|---|---|---|---|---|---|
| ADM1 | ADM2 | Mean | SD | CI95% | Q25 | Q75 | Mean | Q25 | Q75 |
| Gansu Province | Guanghe xian | 3·451 | 5·484 | 18·122 | 0·684 | 3·841 | 8467 | 1686 | 9422 |
| Gansu Province | Gulang xian | 4·125 | 6·548 | 23·312 | 0·747 | 4·544 | 9649 | 1710 | 10568 |
| Gansu Province | Heshui xian | 3·363 | 4·609 | 16·168 | 0·753 | 4·036 | 4296 | 941 | 5198 |
| Gansu Province | Hezheng xian | 3·410 | 5·248 | 18·051 | 0·680 | 3·790 | 7061 | 1443 | 7865 |
| Gansu Province | Huachi xian | 3·498 | 4·841 | 17·251 | 0·764 | 4·144 | 4145 | 914 | 4868 |
| Gansu Province | Huan xian | 3·536 | 5·117 | 17·694 | 0·756 | 4·120 | 10373 | 2213 | 12608 |
| Gansu Province | Huating xian | 2·995 | 4·209 | 14·879 | 0·676 | 3·513 | 5609 | 1271 | 6557 |
| Gansu Province | Hui xian | 2·281 | 3·133 | 10·456 | 0·605 | 2·693 | 4073 | 1081 | 4870 |
| Gansu Province | Huining xian | 3·452 | 5·479 | 17·808 | 0·706 | 3·867 | 13773 | 2753 | 15310 |
| Gansu Province | Jinchangshi | 4·531 | 7·459 | 25·762 | 0·793 | 4·870 | 12038 | 2105 | 12458 |
| Gansu Province | Jingchuan xian | 3·108 | 4·267 | 15·105 | 0·722 | 3·652 | 6884 | 1598 | 8051 |
| Gansu Province | Jingning xian | 3·134 | 4·671 | 16·097 | 0·689 | 3·584 | 10862 | 2362 | 12401 |
| Gansu Province | Jingtai xian | 3·957 | 6·295 | 21·804 | 0·738 | 4·371 | 8067 | 1480 | 8886 |
| Gansu Province | Jingyuan xian | 3·125 | 4·596 | 15·598 | 0·695 | 3·639 | 2022 | 439 | 2339 |
| Gansu Province | Jingyuan xian | 3·752 | 5·941 | 19·725 | 0·719 | 4·206 | 21309 | 4120 | 23808 |
| Gansu Province | Jinta xian | 5·075 | 8·170 | 29·274 | 0·859 | 5·502 | 6180 | 1040 | 6849 |
| Gansu Province | Jishishanbaoanzudongxiangzusala zuzizhi xian | 3·621 | 5·594 | 19·970 | 0·696 | 3·969 | 7944 | 1509 | 8662 |
| Gansu Province | Jiuquanshi | 4·985 | 7·917 | 27·579 | 0·870 | 5·567 | 22596 | 3914 | 25345 |
| Gansu Province | Jiyuguanshi | 5·053 | 7·871 | 27·521 | 0·890 | 5·721 | 15909 | 2854 | 17949 |
| Gansu Province | Kang xian | 1·935 | 2·332 | 8·205 | 0·582 | 2·363 | 3212 | 928 | 3945 |
| Gansu Province | Kangle xian | 3·294 | 5·077 | 17·336 | 0·668 | 3·664 | 7635 | 1561 | 8541 |
| Gansu Province | Lanzhoushi | 3·703 | 5·849 | 20·198 | 0·706 | 4·065 | 119810 | 23403 | 133207 |
| Gansu Province | Li xian | 2·484 | 3·626 | 11·827 | 0·625 | 2·861 | 11019 | 2771 | 12606 |
| Gansu Province | Liangdang xian | 2·289 | 3·053 | 10·410 | 0·613 | 2·712 | 952 | 255 | 1110 |
| Gansu Province | Lingtai xian | 2·988 | 4·048 | 14·248 | 0·704 | 3·552 | 4409 | 1031 | 5316 |
| Gansu Province | Lintan xian | 3·135 | 4·884 | 16·590 | 0·662 | 3·434 | 1428 | 300 | 1577 |
| Gansu Province | Lintao xian | 3·404 | 5·356 | 18·038 | 0·691 | 3·766 | 17980 | 3551 | 19633 |
| Gansu Province | Linxia xian | 3·525 | 5·416 | 19·135 | 0·694 | 3·910 | 10702 | 2114 | 11850 |
| Gansu Province | Linxiashi | 3·559 | 5·517 | 20·023 | 0·697 | 3·958 | 13457 | 2618 | 14921 |
| Gansu Province | Linzhe xian | 4·866 | 7·900 | 27·847 | 0·841 | 5·265 | 5557 | 972 | 6135 |
| Gansu Province | Long xian | 2·834 | 3·851 | 13·703 | 0·673 | 3·362 | 5950 | 1431 | 7070 |
| Gansu Province | Longde xian | 3·198 | 4·729 | 15·871 | 0·702 | 3·688 | 3815 | 836 | 4396 |
| Gansu Province | Longxi xian | 3·099 | 4·862 | 16·011 | 0·669 | 3·469 | 13137 | 2915 | 14848 |
| Gansu Province | Lueyang xian | 1·984 | 2·436 | 8·582 | 0·580 | 2·411 | 2750 | 800 | 3367 |
| Gansu Province | Luqu xian | 3·188 | 4·784 | 16·991 | 0·694 | 3·561 | 1138 | 253 | 1288 |
| Gansu Province | Min xian | 2·866 | 4·440 | 14·649 | 0·643 | 3·211 | 12403 | 2724 | 14045 |

| | | β-thalassaemia carrier prevalence | | | | | Estimated carrier counts | | |
|---|---|---|---|---|---|---|---|---|---|
| ADM1 | ADM2 | Mean | SD | CI95% | Q25 | Q75 | Mean | Q25 | Q75 |
| Gansu Province | Minle xian | 4·641 | 7·550 | 25·912 | 0·835 | 5·067 | 8954 | 1627 | 9777 |
| Gansu Province | Minqin xian | 4·423 | 7·179 | 24·643 | 0·785 | 4·862 | 7514 | 1309 | 8228 |
| Gansu Province | Ning xian | 3·222 | 4·403 | 15·525 | 0·737 | 3·864 | 12683 | 2914 | 15152 |
| Gansu Province | Pengyang xian | 3·331 | 4·893 | 16·807 | 0·726 | 3·826 | 5225 | 1143 | 6103 |
| Gansu Province | Pingliangshi | 3·145 | 4·479 | 15·472 | 0·705 | 3·677 | 15602 | 3566 | 18500 |
| Gansu Province | Qinan xian | 2·908 | 4·290 | 14·784 | 0·658 | 3·303 | 12520 | 2901 | 14180 |
| Gansu Province | Qingshi xian | 2·739 | 3·890 | 13·444 | 0·650 | 3·189 | 6056 | 1471 | 7010 |
| Gansu Province | Qingyang xian | 3·322 | 4·604 | 16·074 | 0·746 | 3·949 | 8002 | 1776 | 9623 |
| Gansu Province | Shandan xian | 4·627 | 7·553 | 26·086 | 0·820 | 5·046 | 6972 | 1257 | 7639 |
| Gansu Province | Shubeimenggu zuzizhi xian | 5·306 | 7·928 | 28·149 | 0·940 | 6·133 | 530 | 93 | 628 |
| Gansu Province | Shubeimenggu zuzizhi xian | 5·067 | 7·848 | 27·180 | 0·915 | 5·764 | 259 | 29 | 289 |
| Gansu Province | Shunanyugu zuzizhi xian | 4·923 | 7·901 | 27·779 | 0·851 | 5·387 | 44 | 7 | 51 |
| Gansu Province | Shunanyugu zuzizhi xian | 4·383 | 7·162 | 24·751 | 0·799 | 4·794 | 270 | 46 | 290 |
| Gansu Province | Shunanyugu zuzizhi xian | 4·844 | 7·687 | 26·429 | 0·861 | 5·406 | 1120 | 189 | 1272 |
| Gansu Province | Tianshuishi | 2·555 | 3·646 | 12·296 | 0·635 | 2·963 | 32177 | 7899 | 37181 |
| Gansu Province | Tianzhuzhangzhuzizhi xian | 4·080 | 6·474 | 22·727 | 0·747 | 4·484 | 6182 | 1101 | 6734 |
| Gansu Province | Tongwei xian | 3·058 | 4·675 | 15·547 | 0·679 | 3·470 | 9989 | 2246 | 11258 |
| Gansu Province | Tuochang xian | 2·586 | 3·879 | 12·413 | 0·633 | 2·946 | 6156 | 1544 | 7039 |
| Gansu Province | Weiyuan xian | 3·196 | 4·966 | 16·760 | 0·673 | 3·556 | 8039 | 1736 | 9167 |
| Gansu Province | Wen xian | 1·881 | 2·274 | 7·835 | 0·573 | 2·289 | 3500 | 1087 | 4307 |
| Gansu Province | Wudu xian | 2·016 | 2·509 | 8·736 | 0·586 | 2·437 | 11289 | 3201 | 13380 |
| Gansu Province | Wushan xian | 2·847 | 4·499 | 14·519 | 0·647 | 3·204 | 11972 | 2745 | 13488 |
| Gansu Province | Wuweishi | 4·310 | 6·925 | 24·104 | 0·776 | 4·726 | 39406 | 7039 | 44189 |
| Gansu Province | Xiahe xian | 3·423 | 5·195 | 18·728 | 0·682 | 3·764 | 6886 | 1362 | 7476 |
| Gansu Province | Xifengshi | 3·213 | 4·359 | 15·414 | 0·728 | 3·868 | 16780 | 3666 | 20320 |
| Gansu Province | Xihei xian | 2·312 | 3·206 | 10·405 | 0·617 | 2·731 | 7880 | 2076 | 9429 |
| Gansu Province | Yongchang xian | 4·494 | 7·363 | 25·355 | 0·792 | 4·890 | 7606 | 1323 | 8267 |
| Gansu Province | Yongdeng xian | 3·911 | 6·220 | 21·809 | 0·730 | 4·289 | 21055 | 3877 | 23045 |
| Gansu Province | Yongjing xian | 3·670 | 5·702 | 19·983 | 0·715 | 4·044 | 6496 | 1250 | 7046 |
| Gansu Province | Yumenshi | 5·147 | 7·907 | 28·227 | 0·912 | 5·829 | 7169 | 1248 | 8197 |
| Gansu Province | Yuzhong xian | 3·575 | 5·699 | 19·177 | 0·702 | 3·940 | 14725 | 2925 | 16173 |
| Gansu Province | Zhang xian | 2·971 | 4·628 | 15·334 | 0·656 | 3·307 | 5089 | 1104 | 5583 |
| Gansu Province | Zhangjiachuanhui zuzizhi xian | 2·878 | 4·078 | 14·536 | 0·661 | 3·346 | 6722 | 1512 | 7706 |
| Gansu Province | Zhangyeshi | 4·766 | 7·736 | 26·451 | 0·842 | 5·233 | 24229 | 4434 | 26653 |
| Gansu Province | Zhenyuan xian | 3·241 | 4·558 | 15·988 | 0·728 | 3·806 | 11549 | 2616 | 13614 |
| Gansu Province | Zhouqu xian | 2·418 | 3·446 | 11·405 | 0·628 | 2·794 | 3126 | 814 | 3628 |

| | | β-thalassaemia carrier prevalence | | | | | Estimated carrier counts | | |
|---|---|---|---|---|---|---|---|---|---|
| ADM1 | ADM2 | Mean | SD | CI95% | Q25 | Q75 | Mean | Q25 | Q75 |
| Gansu Province | Zhuanglang xian | 3·002 | 4·357 | 15·083 | 0·683 | 3·433 | 10069 | 2278 | 11588 |
| Gansu Province | Zuoni xian | 3·163 | 4·851 | 16·793 | 0·665 | 3·502 | 5906 | 1281 | 6562 |
| Guangxi Zhuang Autonomous Region | Baiseshi | 6·492 | 4·222 | 15·894 | 3·545 | 8·280 | 30756 | 16870 | 39013 |
| Guangxi Zhuang Autonomous Region | Bamayao zuzizhi xian | 6·677 | 4·575 | 17·265 | 3·515 | 8·536 | 17991 | 9626 | 22918 |
| Guangxi Zhuang Autonomous Region | Beiliushi | 5·395 | 3·473 | 13·001 | 2·988 | 6·850 | 70107 | 39027 | 88905 |
| Guangxi Zhuang Autonomous Region | Binyang xian | 6·796 | 4·888 | 18·301 | 3·415 | 8·733 | 56261 | 28379 | 72195 |
| Guangxi Zhuang Autonomous Region | Bobai xian | 4·990 | 3·418 | 12·720 | 2·673 | 6·324 | 74093 | 39891 | 93329 |
| Guangxi Zhuang Autonomous Region | Changwu xian | 6·593 | 4·802 | 17·967 | 3·313 | 8·431 | 43948 | 21960 | 55934 |
| Guangxi Zhuang Autonomous Region | Chenxi xian | 5·804 | 3·988 | 14·987 | 3·039 | 7·406 | 42206 | 22199 | 53767 |
| Guangxi Zhuang Autonomous Region | Chongzuo xian | 6·635 | 5·187 | 19·377 | 3·108 | 8·549 | 29194 | 13167 | 38068 |
| Guangxi Zhuang Autonomous Region | Dahuayao zuzizhi xian | 6·849 | 4·850 | 18·151 | 3·522 | 8·718 | 6150 | 3193 | 7820 |
| Guangxi Zhuang Autonomous Region | Daxin xian | 6·806 | 5·107 | 19·128 | 3·314 | 8·760 | 18984 | 9262 | 24346 |
| Guangxi Zhuang Autonomous Region | Debao xian | 6·843 | 4·761 | 17·829 | 3·538 | 8·757 | 18533 | 9598 | 23447 |
| Guangxi Zhuang Autonomous Region | Donglan xian | 6·425 | 4·850 | 18·111 | 3·108 | 8·237 | 13609 | 6559 | 17452 |
| Guangxi Zhuang Autonomous Region | Duanyao zuzizhi xian | 6·852 | 5·065 | 18·969 | 3·375 | 8·791 | 52640 | 26611 | 66948 |
| Guangxi Zhuang Autonomous Region | Fangchenggangshi | 5·788 | 5·078 | 18·664 | 2·472 | 7·390 | 27991 | 12064 | 35609 |
| Guangxi Zhuang Autonomous Region | Fengshan xian | 6·121 | 4·481 | 16·752 | 3·057 | 7·821 | 10712 | 5384 | 13729 |
| Guangxi Zhuang Autonomous Region | Fuchuanyao zuzizhi xian | 6·619 | 4·820 | 18·320 | 3·329 | 8·466 | 17386 | 8794 | 22123 |
| Guangxi Zhuang Autonomous Region | Fusui xian | 6·593 | 5·114 | 19·074 | 3·109 | 8·467 | 28945 | 13719 | 37302 |

| ADM1 | ADM2 | β-thalassaemia carrier prevalence Mean | SD | CI95% | Q25 | Q75 | Estimated carrier counts Mean | Q25 | Q75 |
|---|---|---|---|---|---|---|---|---|---|
| Guangxi Zhuang Autonomous Region | Gongchengyao zuzizhi xian | 6·444 | 4·505 | 16·836 | 3·340 | 8·240 | 15288 | 7983 | 19647 |
| Guangxi Zhuang Autonomous Region | Guanyang xian | 5·988 | 4·304 | 16·102 | 3·031 | 7·660 | 12203 | 6157 | 15586 |
| Guangxi Zhuang Autonomous Region | Guigangshi | 6·552 | 4·473 | 16·775 | 3·442 | 8·372 | 110932 | 58731 | 142153 |
| Guangxi Zhuang Autonomous Region | Guilinshi | 6·078 | 4·270 | 15·981 | 3·126 | 7·791 | 79811 | 40552 | 102286 |
| Guangxi Zhuang Autonomous Region | Guipingshi | 6·679 | 4·545 | 17·026 | 3·521 | 8·573 | 101139 | 52937 | 130051 |
| Guangxi Zhuang Autonomous Region | Hechishi | 6·375 | 4·949 | 18·535 | 2·979 | 8·213 | 25306 | 11401 | 32303 |
| Guangxi Zhuang Autonomous Region | Heng xian | 6·413 | 4·581 | 17·170 | 3·252 | 8·223 | 55722 | 28108 | 71173 |
| Guangxi Zhuang Autonomous Region | Hepu xian | 4·798 | 3·645 | 13·529 | 2·352 | 6·112 | 46077 | 22274 | 58567 |
| Guangxi Zhuang Autonomous Region | Heshanshi | 7·008 | 5·137 | 18·833 | 3·532 | 8·954 | 7538 | 3829 | 9713 |
| Guangxi Zhuang Autonomous Region | Hezhoushi | 6·823 | 4·995 | 18·745 | 3·370 | 8·766 | 72712 | 35443 | 93299 |
| Guangxi Zhuang Autonomous Region | Huanjiangmaonan zuzizhi xian | 5·836 | 4·756 | 17·659 | 2·579 | 7·593 | 15964 | 7135 | 20842 |
| Guangxi Zhuang Autonomous Region | Jianghuayao zuzizhi xian | 6·530 | 4·721 | 17·731 | 3·265 | 8·389 | 27808 | 13731 | 35627 |
| Guangxi Zhuang Autonomous Region | Jiangyong xian | 6·295 | 4·504 | 16·974 | 3·181 | 8·065 | 15723 | 7970 | 20176 |
| Guangxi Zhuang Autonomous Region | Jingxi xian | 6·703 | 5·064 | 18·981 | 3·230 | 8·603 | 32957 | 16109 | 42644 |
| Guangxi Zhuang Autonomous Region | Jinxiuyao zuzizhi xian | 6·993 | 4·976 | 18·705 | 3·536 | 8·995 | 5633 | 2859 | 7263 |
| Guangxi Zhuang Autonomous Region | Laibin xian | 7·053 | 5·124 | 19·254 | 3·501 | 9·069 | 65073 | 32156 | 82631 |
| Guangxi Zhuang Autonomous Region | Leye xian | 5·534 | 4·220 | 15·523 | 2·692 | 7·056 | 8134 | 3958 | 10496 |
| Guangxi Zhuang Autonomous Region | Lingchuan xian | 5·805 | 4·186 | 15·594 | 2·921 | 7·438 | 17597 | 8943 | 22667 |

| ADM1 | ADM2 | β-thalassaemia carrier prevalence Mean | SD | CI95% | Q25 | Q75 | Estimated carrier counts Mean | Q25 | Q75 |
|---|---|---|---|---|---|---|---|---|---|
| Guangxi Zhuang Autonomous Region | Lingshan xian | 5·797 | 4·219 | 15·747 | 2·906 | 7·421 | 73493 | 37466 | 93669 |
| Guangxi Zhuang Autonomous Region | Lingui xian | 5·789 | 4·303 | 15·821 | 2·845 | 7·429 | 30412 | 15519 | 38895 |
| Guangxi Zhuang Autonomous Region | Lingyun xian | 6·154 | 4·277 | 16·050 | 3·196 | 7·891 | 11186 | 5846 | 14254 |
| Guangxi Zhuang Autonomous Region | Lipu xian | 6·713 | 4·661 | 17·445 | 3·482 | 8·557 | 22079 | 11442 | 28057 |
| Guangxi Zhuang Autonomous Region | Liucheng xian | 6·482 | 4·962 | 18·476 | 3·091 | 8·345 | 22215 | 10724 | 28564 |
| Guangxi Zhuang Autonomous Region | Liujiang xian | 6·871 | 5·116 | 19·385 | 3·353 | 8·816 | 28878 | 14132 | 37181 |
| Guangxi Zhuang Autonomous Region | Liuzhoushi | 6·764 | 5·004 | 18·985 | 3·286 | 8·715 | 126574 | 61667 | 164157 |
| Guangxi Zhuang Autonomous Region | Longan xian | 6·913 | 4·856 | 18·281 | 3·547 | 8·844 | 22494 | 11549 | 28887 |
| Guangxi Zhuang Autonomous Region | Longlinge zuzizhi xian | 4·953 | 3·718 | 13·787 | 2·446 | 6·300 | 16886 | 8287 | 21650 |
| Guangxi Zhuang Autonomous Region | Longshenge zuzizhi xian | 5·059 | 4·038 | 14·929 | 2·353 | 6·464 | 7456 | 3457 | 9553 |
| Guangxi Zhuang Autonomous Region | Longzhou xian | 6·573 | 5·464 | 20·423 | 2·891 | 8·455 | 15151 | 6627 | 19278 |
| Guangxi Zhuang Autonomous Region | Luchuan xian | 5·049 | 3·290 | 12·259 | 2·773 | 6·398 | 37533 | 20729 | 47283 |
| Guangxi Zhuang Autonomous Region | Luochengyaolao zuzizhi xian | 6·138 | 4·843 | 17·967 | 2·822 | 7·956 | 16766 | 7750 | 21620 |
| Guangxi Zhuang Autonomous Region | Luzai xian | 6·669 | 4·890 | 18·417 | 3·305 | 8·557 | 40056 | 19779 | 51443 |
| Guangxi Zhuang Autonomous Region | Mashan xian | 6·962 | 4·886 | 18·340 | 3·580 | 8·904 | 29134 | 15062 | 37158 |
| Guangxi Zhuang Autonomous Region | Mengshan xian | 6·910 | 4·882 | 18·342 | 3·544 | 8·839 | 11662 | 6060 | 14858 |
| Guangxi Zhuang Autonomous Region | Nandan xian | 5·660 | 4·719 | 17·552 | 2·497 | 7·250 | 14465 | 6451 | 18672 |
| Guangxi Zhuang Autonomous Region | Nanningshi | 6·772 | 5·049 | 18·815 | 3·325 | 8·687 | 267420 | 132134 | 341421 |

| ADM1 | ADM2 | β-thalassaemia carrier prevalence Mean | SD | CI95% | Q25 | Q75 | Estimated carrier counts Mean | Q25 | Q75 |
|---|---|---|---|---|---|---|---|---|---|
| Guangxi Zhuang Autonomous Region | Napo xian | 6·481 | 5·243 | 19·260 | 2·951 | 8·341 | 10477 | 4797 | 13438 |
| Guangxi Zhuang Autonomous Region | Ningming xian | 6·264 | 5·589 | 20·747 | 2·607 | 8·013 | 19532 | 8218 | 25134 |
| Guangxi Zhuang Autonomous Region | Pingguo xian | 6·930 | 4·667 | 17·535 | 3·710 | 8·815 | 31224 | 16488 | 39775 |
| Guangxi Zhuang Autonomous Region | Pingle xian | 6·730 | 4·674 | 17·696 | 3·509 | 8·592 | 22567 | 11823 | 28753 |
| Guangxi Zhuang Autonomous Region | Pingnan xian | 6·805 | 4·765 | 17·954 | 3·496 | 8·731 | 78465 | 40531 | 100986 |
| Guangxi Zhuang Autonomous Region | Pingxiangshi | 6·387 | 5·827 | 21·651 | 2·608 | 8·127 | 7882 | 3191 | 10061 |
| Guangxi Zhuang Autonomous Region | Pubei xian | 5·498 | 3·817 | 14·296 | 2·889 | 6·990 | 29174 | 15329 | 37218 |
| Guangxi Zhuang Autonomous Region | Qinzhoushi | 5·766 | 4·527 | 16·828 | 2·677 | 7·424 | 69259 | 32884 | 88503 |
| Guangxi Zhuang Autonomous Region | Quanzhou xian | 5·214 | 3·684 | 13·747 | 2·700 | 6·652 | 28648 | 14951 | 36650 |
| Guangxi Zhuang Autonomous Region | Rong xian | 5·833 | 3·852 | 14·441 | 3·163 | 7·414 | 38669 | 21190 | 49091 |
| Guangxi Zhuang Autonomous Region | Rongan xian | 5·914 | 4·658 | 17·440 | 2·769 | 7·571 | 14936 | 6901 | 19301 |
| Guangxi Zhuang Autonomous Region | Rongshuimiao zuzizhi xian | 5·608 | 4·631 | 17·155 | 2·485 | 7·232 | 24019 | 10621 | 30693 |
| Guangxi Zhuang Autonomous Region | Shanglin xian | 6·985 | 5·108 | 19·211 | 3·478 | 8·951 | 23619 | 11726 | 30219 |
| Guangxi Zhuang Autonomous Region | Shangsi xian | 6·221 | 5·216 | 19·282 | 2·722 | 8·032 | 12662 | 5537 | 16485 |
| Guangxi Zhuang Autonomous Region | Shanjiangdong zuzizhi xian | 5·061 | 4·151 | 15·247 | 2·272 | 6·525 | 15201 | 6808 | 19513 |
| Guangxi Zhuang Autonomous Region | Tiandeng xian | 6·901 | 4·828 | 18·199 | 3·564 | 8·832 | 19139 | 9845 | 24658 |
| Guangxi Zhuang Autonomous Region | Tiandong xian | 6·970 | 4·589 | 17·305 | 3·783 | 8·868 | 23546 | 12887 | 29637 |
| Guangxi Zhuang Autonomous Region | Tianer xian | 5·531 | 4·467 | 16·600 | 2·550 | 7·048 | 7307 | 3458 | 9265 |

| | | β-thalassaemia carrier prevalence | | | | | Estimated carrier counts | | |
|---|---|---|---|---|---|---|---|---|---|
| ADM1 | ADM2 | Mean | SD | CI95% | Q25 | Q75 | Mean | Q25 | Q75 |
| Guangxi Zhuang Autonomous Region | Tianlin xian | 5·778 | 4·136 | 15·473 | 2·956 | 7·355 | 13296 | 6908 | 16916 |
| Guangxi Zhuang Autonomous Region | Tianyang xian | 6·868 | 4·477 | 16·815 | 3·743 | 8·778 | 24962 | 13647 | 31565 |
| Guangxi Zhuang Autonomous Region | Wuming xian | 6·889 | 4·904 | 18·331 | 3·504 | 8·834 | 47114 | 23738 | 60221 |
| Guangxi Zhuang Autonomous Region | Wuxuan xian | 7·070 | 5·090 | 19·056 | 3·563 | 9·060 | 23804 | 12039 | 30606 |
| Guangxi Zhuang Autonomous Region | Wuzhoushi | 6·546 | 4·814 | 17·978 | 3·268 | 8·378 | 32242 | 16004 | 41390 |
| Guangxi Zhuang Autonomous Region | Xiangzhou xian | 7·041 | 5·154 | 19·328 | 3·493 | 9·072 | 21824 | 10844 | 28330 |
| Guangxi Zhuang Autonomous Region | Xilin xian | 5·159 | 3·968 | 14·788 | 2·477 | 6·587 | 7376 | 3603 | 9369 |
| Guangxi Zhuang Autonomous Region | Xincheng xian | 6·931 | 5·155 | 19·357 | 3·380 | 8·909 | 20884 | 10246 | 26860 |
| Guangxi Zhuang Autonomous Region | Xingan xian | 5·603 | 4·095 | 15·268 | 2·797 | 7·191 | 17336 | 8686 | 22214 |
| Guangxi Zhuang Autonomous Region | Yangsuo xian | 6·522 | 4·549 | 17·027 | 3·377 | 8·315 | 18173 | 9364 | 23256 |
| Guangxi Zhuang Autonomous Region | Yizhoushi | 6·526 | 4·950 | 18·523 | 3·103 | 8·439 | 35102 | 16720 | 45618 |
| Guangxi Zhuang Autonomous Region | Yongfu xian | 6·184 | 4·500 | 16·837 | 3·093 | 7·927 | 14664 | 7428 | 18764 |
| Guangxi Zhuang Autonomous Region | Yongning xian | 6·464 | 4·831 | 18·140 | 3·142 | 8·332 | 90577 | 44538 | 116411 |
| Guangxi Zhuang Autonomous Region | Yulinshi | 5·846 | 3·686 | 13·940 | 3·276 | 7·398 | 102439 | 57711 | 129442 |
| Guangxi Zhuang Autonomous Region | Zhaoping xian | 6·699 | 4·765 | 17·891 | 3·412 | 8·576 | 41902 | 21228 | 53735 |
| Guangxi Zhuang Autonomous Region | Zhaoping xian | 6·903 | 4·954 | 18·588 | 3·486 | 8·834 | 21140 | 10579 | 27276 |
| Guangxi Zhuang Autonomous Region | Zhongshan xian | 6·814 | 4·927 | 18·499 | 3·433 | 8·726 | 27844 | 13904 | 35517 |
| Guangxi Zhuang Autonomous Region | Ziyuan xian | 4·828 | 3·621 | 13·298 | 2·397 | 6·131 | 7069 | 3537 | 8897 |

| | | β-thalassaemia carrier prevalence | | | | | Estimated carrier counts | | |
|---|---|---|---|---|---|---|---|---|---|
| **ADM1** | **ADM2** | **Mean** | **SD** | **CI95%** | **Q25** | **Q75** | **Mean** | **Q25** | **Q75** |
| Guangzhou Province | Boluo xian | 5·225 | 3·461 | 12·983 | 2·836 | 6·646 | 97997 | 53834 | 123791 |
| Guangzhou Province | Chaoyang xian | 4·757 | 3·185 | 11·967 | 2·573 | 6·079 | 134358 | 72459 | 171604 |
| Guangzhou Province | Chaozhoushi | 6·175 | 3·846 | 14·416 | 3·503 | 7·811 | 87188 | 48813 | 110569 |
| Guangzhou Province | Chenghaishi | 5·093 | 3·424 | 12·935 | 2·736 | 6·421 | 64578 | 34313 | 82133 |
| Guangzhou Province | Chonghuashi | 5·941 | 3·798 | 14·371 | 3·303 | 7·496 | 41500 | 23553 | 52026 |
| Guangzhou Province | Dapu xian | 8·078 | 4·820 | 18·288 | 4·680 | 10·262 | 26514 | 15298 | 33996 |
| Guangzhou Province | Deqing xian | 5·980 | 4·314 | 16·151 | 3·038 | 7·652 | 23668 | 12001 | 30706 |
| Guangzhou Province | Dianba xian | 3·783 | 2·635 | 9·791 | 2·002 | 4·784 | 48844 | 26001 | 61935 |
| Guangzhou Province | Dongguanshi | 4·420 | 2·806 | 10·544 | 2·482 | 5·583 | 432565 | 243102 | 546304 |
| Guangzhou Province | Doumen xian | 3·762 | 2·643 | 9·660 | 1·977 | 4·795 | 15702 | 8222 | 20086 |
| Guangzhou Province | Enpingshi | 4·275 | 3·126 | 11·681 | 2·165 | 5·436 | 18566 | 9323 | 23396 |
| Guangzhou Province | Fengkai xian | 6·399 | 4·642 | 17·525 | 3·224 | 8·181 | 27724 | 13994 | 35537 |
| Guangzhou Province | Fengshun xian | 7·758 | 4·590 | 17·456 | 4·513 | 9·822 | 33978 | 19673 | 42921 |
| Guangzhou Province | Fogang xian | 6·346 | 4·186 | 15·736 | 3·463 | 8·050 | 20405 | 11377 | 25749 |
| Guangzhou Province | Foshanshi | 5·142 | 3·258 | 11·877 | 2·886 | 6·573 | 52713 | 29786 | 67278 |
| Guangzhou Province | Gaomingshi | 5·059 | 3·481 | 13·009 | 2·683 | 6·431 | 13760 | 7327 | 17466 |
| Guangzhou Province | Gaoyaoshi | 5·623 | 3·937 | 14·749 | 2·927 | 7·153 | 40517 | 21277 | 51238 |
| Guangzhou Province | Gaozhoushi | 4·284 | 2·814 | 10·518 | 2·352 | 5·430 | 61211 | 33745 | 77593 |
| Guangzhou Province | Guangning xian | 6·306 | 4·511 | 16·824 | 3·243 | 8·025 | 24697 | 12675 | 31707 |
| Guangzhou Province | Guangzhoushi | 5·290 | 3·228 | 12·265 | 3·037 | 6·646 | 602588 | 346779 | 756998 |
| Guangzhou Province | Haifeng xian | 4·370 | 3·056 | 11·313 | 2·310 | 5·523 | 53847 | 28051 | 68545 |
| Guangzhou Province | Heping xian | 8·390 | 5·371 | 20·258 | 4·603 | 10·745 | 28250 | 15276 | 36547 |
| Guangzhou Province | Heshanshi | 4·701 | 3·201 | 11·994 | 2·508 | 5·980 | 22272 | 12206 | 28432 |
| Guangzhou Province | Heyuanshi | 7·222 | 4·743 | 17·916 | 3·920 | 9·200 | 72142 | 38872 | 91921 |
| Guangzhou Province | Huadushi | 5·797 | 3·634 | 13·780 | 3·278 | 7·338 | 87068 | 49415 | 109968 |
| Guangzhou Province | Huaiji xian | 6·595 | 4·755 | 17·859 | 3·329 | 8·460 | 54409 | 27660 | 69708 |
| Guangzhou Province | Huazhoushi | 4·191 | 2·803 | 10·492 | 2·283 | 5·284 | 48887 | 26512 | 61489 |
| Guangzhou Province | Huidong xian | 4·633 | 3·150 | 11·770 | 2·484 | 5·860 | 37231 | 19979 | 47098 |
| Guangzhou Province | Huilai xian | 4·461 | 3·114 | 11·466 | 2·353 | 5·637 | 44582 | 23284 | 56457 |
| Guangzhou Province | Huiyangshi | 4·470 | 3·032 | 11·253 | 2·402 | 5·658 | 85212 | 45761 | 107451 |
| Guangzhou Province | Huizhoushi | 4·428 | 2·923 | 10·728 | 2·411 | 5·620 | 48480 | 26090 | 62009 |
| Guangzhou Province | Jiangmenshi | 4·361 | 2·921 | 11·156 | 2·377 | 5·496 | 36088 | 19782 | 45770 |
| Guangzhou Province | Jiaoling xian | 8·976 | 5·175 | 19·602 | 5·286 | 11·433 | 15458 | 9126 | 19694 |
| Guangzhou Province | Jiexi xian | 5·979 | 3·793 | 14·348 | 3·353 | 7·580 | 41932 | 23286 | 53491 |
| Guangzhou Province | Jieyangshi | 6·052 | 3·732 | 14·091 | 3·448 | 7·663 | 113376 | 64207 | 143731 |
| Guangzhou Province | Kaipingshi | 4·410 | 3·211 | 11·903 | 2·238 | 5·590 | 29265 | 14942 | 36805 |

| | | β-thalassaemia carrier prevalence | | | | | Estimated carrier counts | | |
|---|---|---|---|---|---|---|---|---|---|
| **ADM1** | **ADM2** | **Mean** | **SD** | **CI95%** | **Q25** | **Q75** | **Mean** | **Q25** | **Q75** |
| Guangzhou Province | Lechang xian | 5·857 | 3·851 | 14·538 | 3·186 | 7·423 | 20131 | 10999 | 25331 |
| Guangzhou Province | Lianjiang xian | 4·100 | 2·830 | 10·542 | 2·183 | 5·177 | 64533 | 34587 | 81275 |
| Guangzhou Province | Liannanyao zuzizhi xian | 6·636 | 4·733 | 17·689 | 3·365 | 8·542 | 9628 | 4870 | 12435 |
| Guangzhou Province | Lianping xian | 7·458 | 4·983 | 18·653 | 4·016 | 9·502 | 20736 | 11089 | 26377 |
| Guangzhou Province | Lianshanzhuang zuzizhi xian | 6·656 | 4·761 | 17·883 | 3·327 | 8·533 | 2893 | 1459 | 3709 |
| Guangzhou Province | Lianzhoushi | 6·366 | 4·475 | 16·914 | 3·282 | 8·132 | 24371 | 12299 | 31072 |
| Guangzhou Province | Longchuan xian | 8·889 | 5·527 | 20·987 | 4·981 | 11·339 | 51893 | 29007 | 66080 |
| Guangzhou Province | Longmen xian | 6·056 | 4·055 | 15·145 | 3·256 | 7·699 | 19536 | 10469 | 24960 |
| Guangzhou Province | Lufeng xian | 4·784 | 3·225 | 12·078 | 2·579 | 6·040 | 58510 | 31235 | 74092 |
| Guangzhou Province | Luodingshi | 5·235 | 3·654 | 13·731 | 2·723 | 6·667 | 47926 | 24728 | 60945 |
| Guangzhou Province | Maomingshi | 3·761 | 2·498 | 9·493 | 2·043 | 4·771 | 31681 | 17295 | 39882 |
| Guangzhou Province | Meizhoushi | 9·078 | 5·197 | 19·737 | 5·370 | 11·518 | 91025 | 54238 | 116223 |
| Guangzhou Province | Nanhaishi | 5·231 | 3·315 | 12·424 | 2·926 | 6·621 | 201543 | 113011 | 254856 |
| Guangzhou Province | Panyushi | 4·716 | 3·006 | 11·343 | 2·663 | 5·928 | 124113 | 69930 | 156460 |
| Guangzhou Province | Pingyuan xian | 9·656 | 5·638 | 21·405 | 5·659 | 12·233 | 19313 | 11405 | 24635 |
| Guangzhou Province | Puningshi | 5·147 | 3·303 | 12·523 | 2·859 | 6·540 | 106848 | 59639 | 135636 |
| Guangzhou Province | Qingxin xian | 6·400 | 4·385 | 16·432 | 3·372 | 8·186 | 13627 | 7143 | 17517 |
| Guangzhou Province | Qingyuanshi | 6·053 | 3·942 | 14·666 | 3·303 | 7·717 | 88553 | 48214 | 113064 |
| Guangzhou Province | Qujiang xian | 6·247 | 4·009 | 15·033 | 3·467 | 7·919 | 42260 | 23529 | 53608 |
| Guangzhou Province | Raoping xian | 6·099 | 3·909 | 14·764 | 3·398 | 7·711 | 39229 | 21631 | 49559 |
| Guangzhou Province | Renhua xian | 5·990 | 3·934 | 14·841 | 3·270 | 7·613 | 9400 | 5239 | 11888 |
| Guangzhou Province | Rucheng xian | 5·628 | 3·806 | 14·209 | 3·015 | 7·163 | 20209 | 10805 | 25692 |
| Guangzhou Province | Ruyuanyao zuzizhi xian | 6·148 | 4·023 | 15·169 | 3·363 | 7·792 | 9714 | 5325 | 12457 |
| Guangzhou Province | Shanshuishi | 5·684 | 3·692 | 13·982 | 3·112 | 7·193 | 49090 | 26745 | 62194 |
| Guangzhou Province | Shantoushi | 4·890 | 3·386 | 12·557 | 2·629 | 6·188 | 44751 | 23643 | 56596 |
| Guangzhou Province | Shaoguanshi | 6·128 | 4·010 | 14·964 | 3·348 | 7·691 | 26300 | 14333 | 33184 |
| Guangzhou Province | Shenzhen xian | 3·837 | 2·566 | 9·511 | 2·101 | 4·839 | 515852 | 283653 | 649846 |
| Guangzhou Province | Shenzhenshi | 3·632 | 2·526 | 9·293 | 1·958 | 4·542 | 101056 | 54300 | 126654 |
| Guangzhou Province | Shixing xian | 6·688 | 4·418 | 16·672 | 3·629 | 8·473 | 13060 | 7165 | 16490 |
| Guangzhou Province | Shundeshi | 4·807 | 3·033 | 11·561 | 2·721 | 6·049 | 153975 | 87018 | 193067 |
| Guangzhou Province | Sihuishi | 5·973 | 4·138 | 15·440 | 3·140 | 7·567 | 24233 | 12724 | 30926 |
| Guangzhou Province | Suixi xian | 3·638 | 2·555 | 9·545 | 1·914 | 4·608 | 31228 | 16638 | 39533 |
| Guangzhou Province | Taishanshi | 3·920 | 2·910 | 10·818 | 1·967 | 4·987 | 39959 | 20380 | 50792 |
| Guangzhou Province | Wongyuan xian | 6·870 | 4·597 | 17·226 | 3·682 | 8·735 | 22146 | 11803 | 28199 |
| Guangzhou Province | Wuchuanshi | 3·595 | 2·463 | 9·267 | 1·921 | 4·533 | 30498 | 16423 | 38335 |
| Guangzhou Province | Wuhua xian | 7·461 | 4·531 | 17·184 | 4·265 | 9·458 | 76633 | 44062 | 97137 |

| | | β-thalassaemia carrier prevalence | | | | | Estimated carrier counts | | |
|---|---|---|---|---|---|---|---|---|---|
| **ADM1** | **ADM2** | **Mean** | **SD** | **CI95%** | **Q25** | **Q75** | **Mean** | **Q25** | **Q75** |
| Guangzhou Province | Xinfeng xian | 6·762 | 4·561 | 17·171 | 3·621 | 8·623 | 13306 | 7201 | 17140 |
| Guangzhou Province | Xingningshi | 9·421 | 5·526 | 21·062 | 5·485 | 11·950 | 67163 | 39024 | 85287 |
| Guangzhou Province | Xinhuishi | 4·242 | 2·946 | 10·939 | 2·230 | 5·397 | 47350 | 25211 | 60187 |
| Guangzhou Province | Xinxing xian | 4·934 | 3·510 | 13·244 | 2·533 | 6·272 | 22579 | 11633 | 28482 |
| Guangzhou Province | Xinyishi | 4·916 | 3·289 | 12·278 | 2·649 | 6·271 | 49537 | 27261 | 62939 |
| Guangzhou Province | Yangchushi | 4·428 | 3·185 | 11·849 | 2·277 | 5·638 | 39350 | 20361 | 50186 |
| Guangzhou Province | Yangshan xian | 6·437 | 4·443 | 16·621 | 3·358 | 8·228 | 25466 | 13318 | 32375 |
| Guangzhou Province | Yangxi xian | 3·882 | 2·870 | 10·647 | 1·954 | 4·939 | 63007 | 32063 | 80104 |
| Guangzhou Province | Yingdeshi | 6·466 | 4·271 | 16·028 | 3·499 | 8·229 | 61429 | 33201 | 78329 |
| Guangzhou Province | Yizhang xian | 5·898 | 4·000 | 15·044 | 3·135 | 7·529 | 35561 | 18940 | 45471 |
| Guangzhou Province | Yunan xian | 5·782 | 4·186 | 15·676 | 2·913 | 7·374 | 14605 | 7320 | 18567 |
| Guangzhou Province | Yunfushi | 5·405 | 3·861 | 14·308 | 2·760 | 6·887 | 33765 | 17324 | 43144 |
| Guangzhou Province | Zengchengshi | 5·351 | 3·357 | 12·635 | 3·010 | 6·790 | 104134 | 59121 | 132252 |
| Guangzhou Province | Zhanjianghsi | 3·447 | 2·435 | 9·148 | 1·826 | 4·316 | 27432 | 14360 | 34140 |
| Guangzhou Province | Zhaoan xian | 5·635 | 3·788 | 14·267 | 3·038 | 7·121 | 27081 | 14493 | 34081 |
| Guangzhou Province | Zhaoqingshi | 5·565 | 3·846 | 14·466 | 2·927 | 7·083 | 34896 | 18204 | 44608 |
| Guangzhou Province | Zhijin xian | 6·042 | 3·966 | 14·858 | 3·305 | 7·672 | 31669 | 17322 | 40187 |
| Guangzhou Province | Zhongshanshi | 4·124 | 2·733 | 10·282 | 2·265 | 5·201 | 209190 | 115109 | 263954 |
| Guangzhou Province | Zhuhaishi | 3·628 | 2·486 | 9·252 | 1·960 | 4·576 | 64514 | 35300 | 81326 |
| Guizhou Province | Anlong xian | 4·492 | 3·502 | 12·977 | 2·144 | 5·732 | 17920 | 8712 | 22732 |
| Guizhou Province | Anshun xian | 3·542 | 3·297 | 11·989 | 1·419 | 4·519 | 32107 | 12835 | 40921 |
| Guizhou Province | Bijieshi | 2·139 | 2·133 | 7·663 | 0·823 | 2·670 | 30014 | 11511 | 37512 |
| Guizhou Province | Ceheng xian | 5·057 | 3·852 | 14·375 | 2·459 | 6·432 | 9399 | 4507 | 11946 |
| Guizhou Province | Changshun xian | 3·932 | 3·704 | 13·415 | 1·580 | 4·983 | 7352 | 2929 | 9314 |
| Guizhou Province | Chengong xian | 2·600 | 2·416 | 8·709 | 1·078 | 3·276 | 2033 | 852 | 2560 |
| Guizhou Province | Congjiang xian | 4·933 | 4·224 | 15·599 | 2·079 | 6·406 | 14268 | 6072 | 18491 |
| Guizhou Province | Dafang xian | 2·388 | 2·399 | 8·623 | 0·905 | 2·989 | 20052 | 7549 | 25258 |
| Guizhou Province | Danzhai xian | 3·936 | 3·752 | 13·699 | 1·539 | 5·014 | 5523 | 2196 | 7079 |
| Guizhou Province | Daozhenkelaozhumiaozhuzizhi xian | 1·675 | 1·713 | 5·998 | 0·639 | 2·082 | 4118 | 1596 | 5047 |
| Guizhou Province | Dejiang xian | 1·976 | 2·017 | 7·088 | 0·762 | 2·457 | 8234 | 3248 | 10335 |
| Guizhou Province | Dushan xian | 4·832 | 4·271 | 15·698 | 2·014 | 6·199 | 12496 | 5194 | 16161 |
| Guizhou Province | Duyunshi | 4·032 | 3·864 | 14·078 | 1·567 | 5·127 | 20721 | 8109 | 26628 |
| Guizhou Province | Fenggang xian | 2·183 | 2·279 | 7·999 | 0·816 | 2·716 | 6217 | 2346 | 7777 |
| Guizhou Province | Fuquan xian | 3·259 | 3·372 | 12·038 | 1·204 | 4·068 | 9758 | 3608 | 12322 |
| Guizhou Province | Guanlingbuyizumiao zuzizhi xian | 3·742 | 3·300 | 12·112 | 1·614 | 4·735 | 11277 | 4937 | 14270 |
| Guizhou Province | Guiding xian | 3·602 | 3·574 | 12·790 | 1·356 | 4·575 | 9169 | 3471 | 11833 |

| | | β-thalassaemia carrier prevalence | | | | | Estimated carrier counts | | |
|---|---|---|---|---|---|---|---|---|---|
| ADM1 | ADM2 | Mean | SD | CI95% | Q25 | Q75 | Mean | Q25 | Q75 |
| Guizhou Province | Guiyangshi | 3·333 | 3·295 | 11·841 | 1·261 | 4·218 | 156580 | 59954 | 198471 |
| Guizhou Province | Hezhang xian | 2·304 | 2·256 | 8·181 | 0·903 | 2·873 | 14393 | 5634 | 17881 |
| Guizhou Province | Huangping xian | 3·068 | 3·144 | 11·196 | 1·151 | 3·813 | 7393 | 2786 | 9327 |
| Guizhou Province | Huishui xian | 4·087 | 3·836 | 13·861 | 1·652 | 5·179 | 16109 | 6344 | 20428 |
| Guizhou Province | Jiangkou xian | 2·253 | 2·013 | 7·281 | 0·962 | 2·857 | 3940 | 1697 | 4979 |
| Guizhou Province | Jianhe xian | 3·469 | 3·272 | 11·716 | 1·404 | 4·406 | 5777 | 2364 | 7500 |
| Guizhou Province | Jingzhoumiaozhudong zuzizhi xian | 3·633 | 3·013 | 11·061 | 1·641 | 4·642 | 9032 | 4118 | 11606 |
| Guizhou Province | Jinping xian | 3·593 | 3·139 | 11·567 | 1·541 | 4·592 | 5086 | 2224 | 6448 |
| Guizhou Province | Jinsha xian | 2·356 | 2·458 | 8·762 | 0·859 | 2·943 | 12343 | 4569 | 15647 |
| Guizhou Province | Kailishi | 3·474 | 3·464 | 12·619 | 1·313 | 4·363 | 21711 | 8338 | 27280 |
| Guizhou Province | Kaiyang xian | 2·846 | 2·954 | 10·589 | 1·029 | 3·569 | 9975 | 3629 | 12545 |
| Guizhou Province | Leishan xian | 3·824 | 3·674 | 13·413 | 1·477 | 4·858 | 4542 | 1720 | 5759 |
| Guizhou Province | Libo xian | 5·308 | 4·512 | 16·705 | 2·276 | 6·874 | 8480 | 3632 | 10899 |
| Guizhou Province | Liping xian | 4·310 | 3·748 | 13·801 | 1·820 | 5·564 | 19978 | 8539 | 25939 |
| Guizhou Province | Liupanshuishi | 2·891 | 2·947 | 10·274 | 1·117 | 3·516 | 8605 | 3289 | 10628 |
| Guizhou Province | Liuzhitequ | 3·306 | 3·133 | 11·416 | 1·335 | 4·150 | 17757 | 7250 | 22407 |
| Guizhou Province | Longli xian | 3·445 | 3·445 | 12·362 | 1·286 | 4·347 | 8281 | 3088 | 10450 |
| Guizhou Province | Luodian xian | 4·858 | 4·251 | 15·671 | 2·112 | 6·147 | 12756 | 5532 | 16133 |
| Guizhou Province | Majiang xian | 3·684 | 3·704 | 13·495 | 1·378 | 4·640 | 8563 | 3160 | 10872 |
| Guizhou Province | Meitan xian | 2·308 | 2·447 | 8·553 | 0·843 | 2·872 | 8603 | 3178 | 10705 |
| Guizhou Province | Nayong xian | 2·723 | 2·710 | 9·707 | 1·058 | 3·397 | 19836 | 7718 | 24765 |
| Guizhou Province | Panxiantequ | 3·634 | 3·218 | 11·824 | 1·568 | 4·584 | 37529 | 15842 | 47285 |
| Guizhou Province | Pingba xian | 3·399 | 3·279 | 11·799 | 1·319 | 4·333 | 12439 | 4866 | 15821 |
| Guizhou Province | Pingtang xian | 4·461 | 4·110 | 15·101 | 1·821 | 5·665 | 10466 | 4253 | 13248 |
| Guizhou Province | Puan xian | 3·702 | 3·163 | 11·626 | 1·651 | 4·697 | 9431 | 4275 | 11989 |
| Guizhou Province | Puding xian | 3·263 | 3·092 | 11·280 | 1·305 | 4·126 | 11091 | 4394 | 14103 |
| Guizhou Province | Qianxi xian | 2·686 | 2·740 | 9·836 | 0·993 | 3·369 | 20488 | 7625 | 25563 |
| Guizhou Province | Qinglong xian | 3·679 | 3·294 | 11·946 | 1·575 | 4·651 | 7671 | 3303 | 9633 |
| Guizhou Province | Qingzhenshi | 3·094 | 3·023 | 10·931 | 1·184 | 3·911 | 16820 | 6338 | 21332 |
| Guizhou Province | Renhuai xian | 2·044 | 2·122 | 7·540 | 0·750 | 2·551 | 13772 | 5049 | 17431 |
| Guizhou Province | Rongjiang xian | 4·328 | 3·935 | 14·379 | 1·734 | 5·586 | 13307 | 5442 | 16924 |
| Guizhou Province | Sandushui zuzizhi xian | 4·488 | 4·050 | 14·843 | 1·834 | 5·770 | 12718 | 5255 | 16454 |
| Guizhou Province | Sansui xian | 3·013 | 2·736 | 9·898 | 1·264 | 3·800 | 5050 | 2101 | 6282 |
| Guizhou Province | Shibing xian | 2·952 | 2·964 | 10·666 | 1·125 | 3·686 | 5789 | 2228 | 7271 |
| Guizhou Province | Shiqian xian | 2·520 | 2·520 | 9·017 | 0·977 | 3·160 | 7462 | 2947 | 9336 |
| Guizhou Province | Shongtaomiao zuzizhi xian | 1·924 | 1·643 | 5·945 | 0·865 | 2·434 | 10195 | 4691 | 12899 |

| | | β-thalassaemia carrier prevalence | | | | | Estimated carrier counts | | |
|---|---|---|---|---|---|---|---|---|---|
| ADM1 | ADM2 | Mean | SD | CI95% | Q25 | Q75 | Mean | Q25 | Q75 |
| Guizhou Province | Shuicheng xian | 3·028 | 2·920 | 10·557 | 1·205 | 3·797 | 33249 | 13050 | 41459 |
| Guizhou Province | Sinan xian | 2·240 | 2·269 | 7·917 | 0·867 | 2·795 | 10094 | 3898 | 12682 |
| Guizhou Province | Suiyang xian | 2·025 | 2·147 | 7·575 | 0·738 | 2·515 | 8651 | 3145 | 10727 |
| Guizhou Province | Taijiang xian | 3·423 | 3·326 | 11·970 | 1·342 | 4·273 | 4440 | 1791 | 5608 |
| Guizhou Province | Tianzhu xian | 3·049 | 2·669 | 9·608 | 1·329 | 3·871 | 8282 | 3670 | 10673 |
| Guizhou Province | Tongrenshi | 2·223 | 1·891 | 6·895 | 1·000 | 2·823 | 11719 | 5209 | 14964 |
| Guizhou Province | Tongzhi xian | 1·812 | 1·910 | 6·661 | 0·672 | 2·244 | 9861 | 3780 | 12106 |
| Guizhou Province | Wangmo xian | 4·827 | 3·981 | 14·666 | 2·213 | 6·145 | 11924 | 5518 | 15098 |
| Guizhou Province | Wanshantequ | 2·362 | 1·997 | 7·246 | 1·078 | 2·978 | 1520 | 683 | 1916 |
| Guizhou Province | Weininglizuhuizumiao zuzizhi xian | 2·467 | 2·433 | 8·771 | 0·956 | 3·091 | 33007 | 12768 | 41241 |
| Guizhou Province | Wengan xian | 2·795 | 2·872 | 10·232 | 1·027 | 3·512 | 11359 | 4158 | 14063 |
| Guizhou Province | Wuchuankelaozhumiaozhuzizhi xian | 1·806 | 1·863 | 6·599 | 0·683 | 2·245 | 5596 | 2077 | 6937 |
| Guizhou Province | Xifeng xian | 2·711 | 2·833 | 9·942 | 0·975 | 3·402 | 5825 | 2104 | 7314 |
| Guizhou Province | Xingren xian | 4·030 | 3·296 | 12·159 | 1·858 | 5·122 | 16999 | 7807 | 21882 |
| Guizhou Province | Xingyishi | 4·351 | 3·359 | 12·362 | 2·106 | 5·535 | 45236 | 22095 | 57182 |
| Guizhou Province | Xinhuangdong zuzizhi xian | 2·651 | 2·308 | 8·312 | 1·162 | 3·353 | 6192 | 2701 | 7896 |
| Guizhou Province | Xishui xian | 1·741 | 1·856 | 6·498 | 0·647 | 2·143 | 11112 | 4184 | 13901 |
| Guizhou Province | Xiushantujiazumiao zuzizhi xian | 1·747 | 1·486 | 5·433 | 0·790 | 2·208 | 8583 | 3975 | 10724 |
| Guizhou Province | Xiuwen xian | 2·901 | 2·913 | 10·614 | 1·071 | 3·664 | 8050 | 3015 | 10069 |
| Guizhou Province | Yanhetujia zuzizhi xian | 1·768 | 1·689 | 6·063 | 0·720 | 2·210 | 4625 | 1887 | 5811 |
| Guizhou Province | Yinjiangtujiazumiao zuzizhi xian | 2·062 | 1·946 | 6·929 | 0·852 | 2·586 | 5548 | 2274 | 6895 |
| Guizhou Province | Yupingdong zuzizhi xian | 2·586 | 2·278 | 8·232 | 1·114 | 3·282 | 4408 | 1899 | 5591 |
| Guizhou Province | Yuqing xian | 2·625 | 2·770 | 9·767 | 0·954 | 3·279 | 4315 | 1552 | 5421 |
| Guizhou Province | Zhenfeng xian | 4·334 | 3·600 | 13·261 | 1·961 | 5·542 | 13125 | 5896 | 16776 |
| Guizhou Province | Zhengan xian | 1·837 | 1·912 | 6·775 | 0·679 | 2·291 | 6956 | 2651 | 8737 |
| Guizhou Province | Zhenningbuyizumiao zuzizhi xian | 3·882 | 3·460 | 12·491 | 1·668 | 4·932 | 12080 | 5070 | 15313 |
| Guizhou Province | Zhenyuan xian | 2·836 | 2·700 | 9·777 | 1·146 | 3·565 | 6157 | 2595 | 7879 |
| Guizhou Province | Zhijin xian | 2·971 | 2·907 | 10·528 | 1·153 | 3·741 | 23459 | 8954 | 29511 |
| Guizhou Province | Zhiyunmiaozubuyi zuzizhi xian | 4·195 | 3·819 | 13·719 | 1·784 | 5·269 | 12279 | 5285 | 15640 |
| Guizhou Province | Zhunyi xian | 2·295 | 2·418 | 8·548 | 0·826 | 2·867 | 28714 | 10293 | 35949 |
| Guizhou Province | Zhunyishi | 2·291 | 2·475 | 8·746 | 0·816 | 2·876 | 26173 | 9291 | 33118 |
| Hainan Province | Baishali zuzizhi xian | 3·014 | 2·175 | 8·071 | 1·560 | 3·803 | 4153 | 2127 | 5228 |
| Hainan Province | Baotinglizumiao zuzizhi xian | 2·924 | 2·199 | 8·116 | 1·467 | 3·686 | 9537 | 4798 | 12020 |
| Hainan Province | Changjiangli zuzizhi xian | 3·150 | 2·385 | 8·763 | 1·554 | 4·026 | 6874 | 3461 | 8746 |
| Hainan Province | Chanzhoushi | 2·998 | 2·142 | 7·965 | 1·553 | 3·798 | 26493 | 13766 | 33510 |
| Hainan Province | Chengmai xian | 2·708 | 1·891 | 6·970 | 1·452 | 3·412 | 10883 | 5820 | 13692 |

| | | β-thalassaemia carrier prevalence | | | | | Estimated carrier counts | | |
|---|---|---|---|---|---|---|---|---|---|
| ADM1 | ADM2 | Mean | SD | CI95% | Q25 | Q75 | Mean | Q25 | Q75 |
| Hainan Province | Dingan xian | 2·678 | 1·920 | 7·072 | 1·402 | 3·383 | 3732 | 1959 | 4724 |
| Hainan Province | Dongfangli zuzizhi xian | 3·153 | 2·520 | 9·147 | 1·485 | 4·022 | 13792 | 6300 | 17755 |
| Hainan Province | Haikoushi | 2·557 | 1·810 | 6·772 | 1·348 | 3·200 | 41585 | 21777 | 51549 |
| Hainan Province | Ledongli zuzizhi xian | 3·033 | 2·423 | 8·876 | 1·437 | 3·841 | 16601 | 7615 | 20966 |
| Hainan Province | Lingao xian | 2·820 | 1·970 | 7·309 | 1·496 | 3·563 | 10878 | 5690 | 13721 |
| Hainan Province | Lingshuilizumiao zuzizhi xian | 2·887 | 2·214 | 8·138 | 1·431 | 3·657 | 8009 | 3892 | 10225 |
| Hainan Province | Qionghaishi | 2·727 | 2·025 | 7·471 | 1·386 | 3·429 | 13796 | 7028 | 17239 |
| Hainan Province | Qiongshanshi | 2·577 | 1·848 | 6·833 | 1·344 | 3·251 | 26124 | 13758 | 32825 |
| Hainan Province | Qiongzhonglizumiao zuzizhi xian | 2·867 | 2·059 | 7·578 | 1·493 | 3·620 | 6316 | 3294 | 7976 |
| Hainan Province | Shanyashi | 2·937 | 2·330 | 8·717 | 1·413 | 3·708 | 11934 | 5773 | 14911 |
| Hainan Province | Tunchang xian | 2·739 | 1·913 | 7·125 | 1·448 | 3·455 | 6811 | 3581 | 8538 |
| Hainan Province | Wanningshi | 2·823 | 2·151 | 7·933 | 1·389 | 3·582 | 13067 | 6380 | 16571 |
| Hainan Province | Wenchang xian | 2·660 | 2·057 | 7·595 | 1·306 | 3·365 | 15237 | 7439 | 19186 |
| Hebei Province | Anguoshi | 3·210 | 5·134 | 17·907 | 0·573 | 3·553 | 10385 | 1837 | 11462 |
| Hebei Province | Anping xian | 3·142 | 5·024 | 17·213 | 0·566 | 3·461 | 9397 | 1658 | 10310 |
| Hebei Province | Anxin xian | 3·250 | 5·245 | 17·329 | 0·562 | 3·611 | 11478 | 1965 | 12713 |
| Hebei Province | Ba xian | 3·198 | 5·023 | 16·823 | 0·583 | 3·609 | 24708 | 4495 | 27914 |
| Hebei Province | Baodingshi | 3·290 | 5·381 | 17·872 | 0·585 | 3·672 | 31618 | 5616 | 34798 |
| Hebei Province | Botoushi | 2·900 | 4·623 | 15·994 | 0·548 | 3·189 | 15079 | 2894 | 16641 |
| Hebei Province | Boxiang xian | 3·210 | 5·111 | 18·363 | 0·596 | 3·491 | 3581 | 671 | 3884 |
| Hebei Province | Boye xian | 3·164 | 5·032 | 17·642 | 0·558 | 3·493 | 8151 | 1434 | 9033 |
| Hebei Province | Chang xian | 2·909 | 4·573 | 15·908 | 0·540 | 3·231 | 26159 | 4831 | 28947 |
| Hebei Province | Changli xian | 3·048 | 4·734 | 16·312 | 0·553 | 3·482 | 12760 | 2308 | 14624 |
| Hebei Province | Changzhoushi | 2·882 | 4·394 | 15·699 | 0·538 | 3·255 | 16448 | 2997 | 18574 |
| Hebei Province | Chengan xian | 3·009 | 4·782 | 16·898 | 0·571 | 3·289 | 11976 | 2243 | 13072 |
| Hebei Province | Chengde xian | 3·673 | 5·844 | 19·767 | 0·654 | 4·121 | 15978 | 2878 | 17965 |
| Hebei Province | Chengdeshi | 3·746 | 5·986 | 20·369 | 0·666 | 4·253 | 18062 | 3248 | 20491 |
| Hebei Province | Chongli xian | 4·131 | 6·822 | 24·068 | 0·684 | 4·388 | 4164 | 687 | 4486 |
| Hebei Province | Cicheng xian | 3·972 | 6·386 | 22·487 | 0·676 | 4·343 | 7756 | 1317 | 8461 |
| Hebei Province | Dacheng xian | 3·049 | 4·715 | 16·362 | 0·569 | 3·448 | 14085 | 2641 | 16157 |
| Hebei Province | Daming xian | 2·831 | 4·632 | 15·668 | 0·538 | 3·033 | 19676 | 3734 | 21217 |
| Hebei Province | Daxing xian | 3·455 | 5·496 | 18·620 | 0·611 | 3·883 | 51377 | 9185 | 57714 |
| Hebei Province | Dingxing xian | 3·349 | 5·310 | 17·707 | 0·591 | 3·798 | 15424 | 2725 | 17574 |
| Hebei Province | Dingzhoushi | 3·310 | 5·414 | 18·055 | 0·585 | 3·654 | 38385 | 6694 | 42030 |
| Hebei Province | Dongguang xian | 2·821 | 4·532 | 15·728 | 0·534 | 3·010 | 10662 | 2032 | 11421 |
| Hebei Province | Feixiang xian | 2·982 | 4·781 | 16·289 | 0·565 | 3·221 | 11297 | 2078 | 12069 |

| ADM1 | ADM2 | β-thalassaemia carrier prevalence Mean | SD | CI95% | Q25 | Q75 | Estimated carrier counts Mean | Q25 | Q75 |
|---|---|---|---|---|---|---|---|---|---|
| Hebei Province | Fengnan xian | 3·023 | 4·683 | 16·075 | 0·554 | 3·468 | 20832 | 3823 | 23879 |
| Hebei Province | Fengningman zuzizhi xian | 4·036 | 6·570 | 22·574 | 0·679 | 4·415 | 13098 | 2216 | 14292 |
| Hebei Province | Fengruan xian | 3·187 | 4·943 | 16·857 | 0·578 | 3·631 | 34265 | 6232 | 38893 |
| Hebei Province | Fucheng xian | 2·855 | 4·563 | 15·564 | 0·545 | 3·123 | 7539 | 1439 | 8280 |
| Hebei Province | Funing xian | 3·143 | 4·894 | 16·980 | 0·565 | 3·572 | 23745 | 4279 | 26934 |
| Hebei Province | Fuping xian | 3·640 | 6·018 | 20·193 | 0·631 | 3·986 | 6418 | 1092 | 6909 |
| Hebei Province | Gaocheng xian | 3·256 | 5·097 | 18·048 | 0·591 | 3·594 | 5307 | 967 | 5949 |
| Hebei Province | Gaoyang xian | 3·171 | 5·109 | 17·355 | 0·564 | 3·486 | 10859 | 1904 | 12019 |
| Hebei Province | Guan xian | 3·337 | 5·296 | 17·480 | 0·599 | 3·837 | 15725 | 2875 | 17970 |
| Hebei Province | Guan xian | 2·791 | 4·493 | 15·731 | 0·544 | 3·019 | 22226 | 4323 | 23929 |
| Hebei Province | Guangping xian | 2·954 | 4·803 | 16·778 | 0·557 | 3·130 | 10212 | 1916 | 10863 |
| Hebei Province | Guangzhong xian | 2·994 | 4·744 | 16·566 | 0·563 | 3·277 | 8517 | 1607 | 9308 |
| Hebei Province | Guantao xian | 2·889 | 4·708 | 16·152 | 0·556 | 3·119 | 7077 | 1387 | 7719 |
| Hebei Province | Gucheng xian | 2·856 | 4·690 | 15·611 | 0·551 | 3·075 | 11468 | 2208 | 12439 |
| Hebei Province | Guyuan xian | 4·157 | 6·902 | 23·600 | 0·701 | 4·431 | 7013 | 1180 | 7535 |
| Hebei Province | Handan xian | 3·036 | 4·715 | 16·413 | 0·590 | 3·381 | 33674 | 6561 | 37968 |
| Hebei Province | Handanshi | 3·024 | 4·621 | 15·252 | 0·569 | 3·376 | 16357 | 3066 | 18175 |
| Hebei Province | Handanshi | 3·121 | 4·725 | 16·843 | 0·594 | 3·530 | 10649 | 1998 | 11876 |
| Hebei Province | Haochengshi | 3·324 | 5·279 | 18·406 | 0·591 | 3·672 | 28051 | 4948 | 30834 |
| Hebei Province | Hejianshi | 3·041 | 4·807 | 16·119 | 0·555 | 3·398 | 22977 | 4218 | 25511 |
| Hebei Province | Hengshuishi | 3·056 | 4·908 | 16·937 | 0·570 | 3·345 | 20301 | 3644 | 21783 |
| Hebei Province | Huaian xian | 4·048 | 6·756 | 23·742 | 0·691 | 4·273 | 7065 | 1245 | 7556 |
| Hebei Province | Huailai xian | 3·867 | 6·224 | 22·006 | 0·660 | 4·238 | 12978 | 2208 | 13952 |
| Hebei Province | Huanghuashi | 2·839 | 4·371 | 15·411 | 0·528 | 3·206 | 15656 | 2893 | 17794 |
| Hebei Province | Ji xian | 3·053 | 4·919 | 16·942 | 0·571 | 3·282 | 10247 | 1916 | 11016 |
| Hebei Province | Jing xian | 2·872 | 4·678 | 15·931 | 0·544 | 3·086 | 12929 | 2436 | 13843 |
| Hebei Province | Jinghai xian | 3·053 | 4·774 | 15·983 | 0·557 | 3·473 | 23397 | 4257 | 26778 |
| Hebei Province | Jinzhoushi | 3·191 | 5·032 | 17·640 | 0·587 | 3·529 | 17710 | 3217 | 19760 |
| Hebei Province | Jizhe xian | 3·049 | 4·896 | 17·136 | 0·570 | 3·227 | 8921 | 1676 | 9429 |
| Hebei Province | Julu xian | 3·083 | 4·879 | 17·131 | 0·577 | 3·345 | 10112 | 1893 | 11013 |
| Hebei Province | Kangbao xian | 4·340 | 7·463 | 25·249 | 0·707 | 4·589 | 6021 | 975 | 6351 |
| Hebei Province | Kuanchengman zuzizhi xian | 3·407 | 5·358 | 18·280 | 0·618 | 3·824 | 8682 | 1547 | 9797 |
| Hebei Province | Laishui xian | 3·568 | 5·728 | 19·324 | 0·627 | 3·965 | 10942 | 1926 | 12297 |
| Hebei Province | Laiyuan xian | 3·610 | 5·985 | 19·755 | 0·636 | 3·963 | 8940 | 1594 | 9831 |
| Hebei Province | Langfangshi | 3·273 | 5·117 | 17·260 | 0·580 | 3·728 | 33244 | 5973 | 37722 |
| Hebei Province | Leting xian | 2·945 | 4·557 | 15·919 | 0·535 | 3·352 | 11603 | 2106 | 13348 |

| ADM1 | ADM2 | β-thalassaemia carrier prevalence Mean | SD | CI95% | Q25 | Q75 | Estimated carrier counts Mean | Q25 | Q75 |
|---|---|---|---|---|---|---|---|---|---|
| Hebei Province | Li xian | 3·164 | 5·086 | 17·013 | 0·565 | 3·522 | 13907 | 2483 | 15367 |
| Hebei Province | Lincheng xian | 3·268 | 5·146 | 18·346 | 0·595 | 3·611 | 6624 | 1207 | 7240 |
| Hebei Province | Lingshou xian | 3·546 | 5·738 | 19·840 | 0·618 | 3·894 | 8877 | 1582 | 9707 |
| Hebei Province | Lingxi xian | 2·898 | 4·638 | 16·172 | 0·544 | 3·203 | 9934 | 1842 | 11044 |
| Hebei Province | Longhua xian | 3·956 | 6·364 | 21·791 | 0·683 | 4·419 | 14543 | 2537 | 16267 |
| Hebei Province | Longyao xian | 3·140 | 4·900 | 17·651 | 0·579 | 3·432 | 18207 | 3384 | 19927 |
| Hebei Province | Luan xian | 3·117 | 4·846 | 16·810 | 0·566 | 3·543 | 15349 | 2812 | 17569 |
| Hebei Province | Luancheng xian | 3·307 | 5·196 | 17·979 | 0·593 | 3·706 | 21831 | 3953 | 24518 |
| Hebei Province | Luannan xian | 2·917 | 4·500 | 15·274 | 0·540 | 3·369 | 5072 | 934 | 5729 |
| Hebei Province | Luannan xian | 2·979 | 4·587 | 15·621 | 0·546 | 3·419 | 14559 | 2690 | 16608 |
| Hebei Province | Luanping xian | 3·799 | 6·151 | 21·079 | 0·651 | 4·220 | 12867 | 2208 | 14484 |
| Hebei Province | Lulong xian | 3·100 | 4·824 | 16·455 | 0·572 | 3·553 | 11492 | 2137 | 13285 |
| Hebei Province | Luquanshi | 3·472 | 5·603 | 19·272 | 0·617 | 3·846 | 29221 | 5223 | 32413 |
| Hebei Province | Mancheng xian | 3·342 | 5·402 | 17·516 | 0·600 | 3·743 | 18445 | 3290 | 20804 |
| Hebei Province | Mencunhui zuzizhi xian | 2·760 | 4·288 | 15·287 | 0·522 | 3·094 | 6214 | 1177 | 6983 |
| Hebei Province | Nangongshi | 3·020 | 4·808 | 17·078 | 0·559 | 3·274 | 12471 | 2304 | 13513 |
| Hebei Province | Nanhe xian | 3·076 | 4·903 | 17·642 | 0·580 | 3·310 | 9568 | 1823 | 10352 |
| Hebei Province | Nanpi xian | 2·805 | 4·390 | 15·559 | 0·526 | 3·084 | 11014 | 2075 | 12057 |
| Hebei Province | Neiqiu xian | 3·211 | 5·008 | 17·943 | 0·593 | 3·508 | 7924 | 1453 | 8623 |
| Hebei Province | Ningjin xian | 3·155 | 5·052 | 17·821 | 0·583 | 3·440 | 24074 | 4464 | 26091 |
| Hebei Province | Pingquan xian | 3·695 | 5·825 | 20·132 | 0·657 | 4·138 | 13630 | 2437 | 15304 |
| Hebei Province | Pingshan xian | 3·661 | 6·021 | 20·432 | 0·635 | 3·997 | 13029 | 2255 | 14118 |
| Hebei Province | Pingxiang xian | 3·059 | 4·903 | 17·290 | 0·566 | 3·305 | 9417 | 1740 | 10121 |
| Hebei Province | Qiananshi | 3·180 | 4·976 | 16·809 | 0·582 | 3·631 | 23967 | 4427 | 27372 |
| Hebei Province | Qianxi xian | 3·259 | 5·083 | 17·187 | 0·599 | 3·711 | 11672 | 2123 | 13674 |
| Hebei Province | Qing xian | 3·015 | 4·709 | 16·143 | 0·551 | 3·399 | 14018 | 2578 | 15972 |
| Hebei Province | Qinghe xian | 2·937 | 4·687 | 16·356 | 0·546 | 3·205 | 12329 | 2247 | 13352 |
| Hebei Province | Qinglongman zuzizhi xian | 3·311 | 5·228 | 17·883 | 0·593 | 3·714 | 15704 | 2810 | 17551 |
| Hebei Province | Qingyuan xian | 3·225 | 5·165 | 17·207 | 0·571 | 3·612 | 33461 | 5972 | 37552 |
| Hebei Province | Qinhuangdaoshi | 3·147 | 4·936 | 17·204 | 0·566 | 3·547 | 29302 | 5253 | 32751 |
| Hebei Province | Qiu xian | 2·960 | 4·808 | 16·418 | 0·564 | 3·170 | 5354 | 1013 | 5694 |
| Hebei Province | Quyang xian | 3·420 | 5·586 | 18·755 | 0·599 | 3·741 | 20526 | 3626 | 22316 |
| Hebei Province | Quzhou xian | 3·015 | 4·943 | 17·247 | 0·560 | 3·194 | 15124 | 2797 | 16012 |
| Hebei Province | Raoyang xian | 3·059 | 4·903 | 16·912 | 0·555 | 3·375 | 6694 | 1201 | 7400 |
| Hebei Province | Ren xian | 3·118 | 4·880 | 17·489 | 0·577 | 3·423 | 9921 | 1824 | 10838 |
| Hebei Province | Renqiushi | 3·137 | 4·973 | 16·722 | 0·566 | 3·524 | 27881 | 5091 | 31421 |

| | | β-thalassaemia carrier prevalence | | | | | Estimated carrier counts | | |
|---|---|---|---|---|---|---|---|---|---|
| ADM1 | ADM2 | Mean | SD | CI95% | Q25 | Q75 | Mean | Q25 | Q75 |
| Hebei Province | Rongcheng xian | 3·325 | 5·331 | 18·045 | 0·595 | 3·725 | 8644 | 1535 | 9659 |
| Hebei Province | Shaheshi | 3·142 | 4·856 | 16·973 | 0·593 | 3·507 | 15063 | 2843 | 16713 |
| Hebei Province | Shenzhoushi | 3·047 | 4·846 | 16·875 | 0·562 | 3·330 | 16262 | 2984 | 17901 |
| Hebei Province | Sheze xian | 3·188 | 5·187 | 17·589 | 0·573 | 3·492 | 7760 | 1408 | 8462 |
| Hebei Province | Shijiazhuanshi | 3·342 | 5·399 | 18·133 | 0·610 | 3·729 | 80632 | 14785 | 90893 |
| Hebei Province | Shuning xian | 3·079 | 4·916 | 16·744 | 0·569 | 3·411 | 9512 | 1766 | 10633 |
| Hebei Province | Tang xian | 3·448 | 5·683 | 18·838 | 0·601 | 3·785 | 16879 | 2905 | 18384 |
| Hebei Province | Tangshanshi | 3·152 | 4·886 | 17·114 | 0·564 | 3·590 | 50784 | 9171 | 57714 |
| Hebei Province | Wan xian | 3·397 | 5·624 | 18·075 | 0·593 | 3·777 | 8417 | 1471 | 9386 |
| Hebei Province | Wangdu xian | 3·260 | 5·366 | 17·443 | 0·588 | 3·626 | 9740 | 1754 | 10811 |
| Hebei Province | Wanquan xian | 4·083 | 6·715 | 23·503 | 0·695 | 4·361 | 6823 | 1175 | 7245 |
| Hebei Province | Wei xian | 3·798 | 6·275 | 21·588 | 0·659 | 4·082 | 15614 | 2675 | 16776 |
| Hebei Province | Wei xian | 2·964 | 4·733 | 16·483 | 0·554 | 3·221 | 15277 | 2866 | 16654 |
| Hebei Province | Weichang xian | 4·085 | 6·501 | 22·046 | 0·704 | 4·620 | 17499 | 2967 | 19713 |
| Hebei Province | Wenan xian | 3·114 | 4·793 | 16·317 | 0·569 | 3·594 | 13544 | 2473 | 15667 |
| Hebei Province | Wuanshi | 3·217 | 4·929 | 17·377 | 0·597 | 3·625 | 28567 | 5293 | 31743 |
| Hebei Province | Wuji xian | 3·312 | 5·239 | 18·050 | 0·588 | 3·676 | 11720 | 2077 | 12943 |
| Hebei Province | Wuqiang xian | 2·966 | 4·774 | 16·539 | 0·555 | 3·190 | 4656 | 865 | 4993 |
| Hebei Province | Wuqiao xian | 2·799 | 4·512 | 15·545 | 0·537 | 3·029 | 6390 | 1217 | 6853 |
| Hebei Province | Wuyi xian | 2·975 | 4·878 | 16·696 | 0·557 | 3·193 | 11553 | 2138 | 12406 |
| Hebei Province | Xian xian | 2·992 | 4·737 | 16·254 | 0·550 | 3·312 | 17055 | 3137 | 19047 |
| Hebei Province | Xincheng xian | 3·345 | 5·340 | 17·703 | 0·594 | 3·820 | 25880 | 4609 | 29174 |
| Hebei Province | Xinglong xian | 3·466 | 5·430 | 18·339 | 0·612 | 3·983 | 12364 | 2177 | 14250 |
| Hebei Province | Xingtai xian | 3·225 | 5·002 | 18·058 | 0·596 | 3·590 | 29829 | 5428 | 32499 |
| Hebei Province | Xingtaishi | 3·167 | 4·895 | 18·633 | 0·593 | 3·531 | 10709 | 2034 | 11856 |
| Hebei Province | Xingtang xian | 3·477 | 5·679 | 19·188 | 0·613 | 3·827 | 13050 | 2309 | 14493 |
| Hebei Province | Xinhe xian | 3·084 | 4·949 | 16·887 | 0·570 | 3·331 | 4063 | 743 | 4311 |
| Hebei Province | Xinjishi | 3·111 | 5·007 | 17·183 | 0·572 | 3·421 | 18271 | 3418 | 20329 |
| Hebei Province | Xinleshi | 3·357 | 5·438 | 18·450 | 0·590 | 3·711 | 14647 | 2574 | 16268 |
| Hebei Province | Xiong xian | 3·249 | 5·188 | 17·150 | 0·586 | 3·686 | 12902 | 2329 | 14665 |
| Hebei Province | Xuanhua xian | 3·988 | 6·507 | 22·927 | 0·673 | 4·267 | 24059 | 4086 | 25828 |
| Hebei Province | Xushui xian | 3·335 | 5·397 | 17·725 | 0·585 | 3·738 | 22774 | 3968 | 25591 |
| Hebei Province | Yi xian | 3·499 | 5·711 | 18·901 | 0·607 | 3·906 | 17662 | 3048 | 19848 |
| Hebei Province | Yongnian xian | 3·039 | 4·751 | 16·501 | 0·577 | 3·358 | 33249 | 6278 | 37118 |
| Hebei Province | Yongqing xian | 3·240 | 5·057 | 17·132 | 0·591 | 3·712 | 13822 | 2544 | 15803 |
| Hebei Province | Yuanshi xian | 3·353 | 5·346 | 18·778 | 0·610 | 3·670 | 12411 | 2243 | 13655 |

| | | β-thalassaemia carrier prevalence | | | | | Estimated carrier counts | | |
|---|---|---|---|---|---|---|---|---|---|
| ADM1 | ADM2 | Mean | SD | CI95% | Q25 | Q75 | Mean | Q25 | Q75 |
| Hebei Province | Zhangbei xian | 4·193 | 7·026 | 24·445 | 0·705 | 4·445 | 13234 | 2232 | 13985 |
| Hebei Province | Zhangjiakoushi | 4·083 | 6·725 | 23·875 | 0·699 | 4·316 | 35805 | 6215 | 37612 |
| Hebei Province | Zhanhuang xian | 3·354 | 5·332 | 18·757 | 0·612 | 3·668 | 6854 | 1263 | 7486 |
| Hebei Province | Zhao xian | 3·250 | 5·153 | 18·189 | 0·597 | 3·544 | 15622 | 2855 | 17013 |
| Hebei Province | Zhaoqian xian | 2·930 | 4·715 | 16·220 | 0·555 | 3·222 | 10145 | 1926 | 11171 |
| Hebei Province | Zhengding xian | 3·416 | 5·524 | 19·012 | 0·599 | 3·757 | 58660 | 10345 | 65100 |
| Hebei Province | Zhunhuashi | 3·281 | 5·045 | 17·313 | 0·587 | 3·801 | 22996 | 4116 | 26681 |
| Hebei Province | Zhuolu xian | 3·817 | 6·184 | 21·481 | 0·658 | 4·144 | 10862 | 1839 | 11837 |
| Hebei Province | Zhuozhoushi | 3·441 | 5·494 | 18·175 | 0·608 | 3·925 | 23530 | 4178 | 26801 |
| Heilongjiang Province | Achengshi | 4·460 | 6·695 | 23·364 | 0·765 | 5·204 | 27114 | 4704 | 31816 |
| Heilongjiang Province | Andashi | 4·506 | 6·851 | 24·288 | 0·803 | 5·113 | 31225 | 5573 | 35188 |
| Heilongjiang Province | Baiquan xian | 4·743 | 7·057 | 25·155 | 0·870 | 5·438 | 12881 | 2355 | 14821 |
| Heilongjiang Province | Baoqing xian | 4·860 | 7·399 | 26·125 | 0·867 | 5·569 | 17899 | 3196 | 20486 |
| Heilongjiang Province | Bayan xian | 4·653 | 6·864 | 24·386 | 0·795 | 5·499 | 21450 | 3682 | 25565 |
| Heilongjiang Province | Beianshi | 4·994 | 7·529 | 27·250 | 0·873 | 5·671 | 15522 | 2730 | 17667 |
| Heilongjiang Province | Bin xian | 4·542 | 6·793 | 24·034 | 0·771 | 5·281 | 22325 | 3801 | 26024 |
| Heilongjiang Province | Boli xian | 4·758 | 7·134 | 25·159 | 0·816 | 5·591 | 16412 | 2793 | 19223 |
| Heilongjiang Province | Daqingshi | 4·490 | 6·837 | 24·176 | 0·795 | 5·100 | 62905 | 11051 | 71416 |
| Heilongjiang Province | Dedu xian | 5·052 | 7·619 | 28·249 | 0·889 | 5·632 | 11987 | 2105 | 13522 |
| Heilongjiang Province | Dongning xian | 4·432 | 7·199 | 24·378 | 0·795 | 4·871 | 9114 | 1605 | 10030 |
| Heilongjiang Province | Duerbotemenggu zuzizhi xian | 4·489 | 6·786 | 24·035 | 0·807 | 5·105 | 8404 | 1539 | 9550 |
| Heilongjiang Province | Fangzheng xian | 4·657 | 6·950 | 24·512 | 0·786 | 5·498 | 8808 | 1503 | 10525 |
| Heilongjiang Province | Fujinshi | 5·048 | 7·700 | 27·392 | 0·888 | 5·749 | 21104 | 3709 | 23889 |
| Heilongjiang Province | Fuyu xian | 4·664 | 7·001 | 25·290 | 0·863 | 5·244 | 10888 | 1975 | 12224 |
| Heilongjiang Province | Fuyuan xian | 5·169 | 8·020 | 28·404 | 0·909 | 5·881 | 4793 | 840 | 5450 |
| Heilongjiang Province | Gannan xian | 4·722 | 7·142 | 25·436 | 0·863 | 5·377 | 15911 | 2955 | 18298 |
| Heilongjiang Province | Haerbinshi | 4·465 | 6·696 | 23·547 | 0·772 | 5·146 | 219944 | 37272 | 251700 |
| Heilongjiang Province | Hailinshi | 4·594 | 7·211 | 25·048 | 0·775 | 5·215 | 12118 | 1964 | 13530 |
| Heilongjiang Province | Hailunshi | 4·826 | 7·144 | 25·240 | 0·868 | 5·613 | 23750 | 4318 | 27557 |
| Heilongjiang Province | Hegangshi | 5·002 | 7·381 | 26·876 | 0·890 | 5·817 | 27360 | 4693 | 30996 |
| Heilongjiang Province | Heiheshi | 5·079 | 7·712 | 28·263 | 0·899 | 5·675 | 8933 | 1554 | 10028 |
| Heilongjiang Province | Huachuan xian | 4·982 | 7·321 | 26·729 | 0·888 | 5·789 | 8985 | 1627 | 10424 |
| Heilongjiang Province | Huanan xian | 4·837 | 7·109 | 25·383 | 0·835 | 5·727 | 14103 | 2416 | 16623 |
| Heilongjiang Province | Hulan xian | 4·545 | 6·786 | 24·188 | 0·781 | 5·268 | 36855 | 6341 | 42481 |
| Heilongjiang Province | Hulin xian | 4·799 | 7·354 | 25·557 | 0·874 | 5·516 | 12762 | 2303 | 14730 |
| Heilongjiang Province | Huma xian | 5·194 | 7·959 | 29·193 | 0·920 | 5·818 | 2808 | 487 | 3139 |

| ADM1 | ADM2 | β-thalassaemia carrier prevalence Mean | SD | CI95% | Q25 | Q75 | Estimated carrier counts Mean | Q25 | Q75 |
|---|---|---|---|---|---|---|---|---|---|
| Heilongjiang Province | Jiamusishi | 4·854 | 7·013 | 25·152 | 0·855 | 5·826 | 37545 | 6610 | 46494 |
| Heilongjiang Province | Jiayin xian | 5·105 | 7·590 | 27·886 | 0·935 | 5·833 | 2541 | 449 | 2884 |
| Heilongjiang Province | Jidong xian | 4·671 | 7·247 | 25·280 | 0·810 | 5·326 | 10437 | 1854 | 11847 |
| Heilongjiang Province | Jixian xian | 4·966 | 7·302 | 26·805 | 0·885 | 5·762 | 12141 | 2204 | 14073 |
| Heilongjiang Province | Jixishi | 4·672 | 7·213 | 25·459 | 0·795 | 5·323 | 30690 | 5298 | 35169 |
| Heilongjiang Province | Kedong xian | 4·954 | 7·416 | 26·686 | 0·876 | 5·633 | 8037 | 1400 | 9020 |
| Heilongjiang Province | Keshan xian | 4·945 | 7·458 | 27·390 | 0·876 | 5·530 | 12972 | 2301 | 14543 |
| Heilongjiang Province | Lanxi xian | 4·523 | 6·744 | 24·020 | 0·811 | 5·228 | 13956 | 2469 | 16238 |
| Heilongjiang Province | Lindian xian | 4·562 | 6·904 | 24·792 | 0·841 | 5·145 | 8485 | 1579 | 9533 |
| Heilongjiang Province | Linkou xian | 4·666 | 7·114 | 25·022 | 0·788 | 5·401 | 11705 | 1945 | 13411 |
| Heilongjiang Province | Longjiang xian | 4·652 | 7·109 | 25·047 | 0·843 | 5·265 | 18736 | 3468 | 21091 |
| Heilongjiang Province | Luobei xian | 5·074 | 7·541 | 27·336 | 0·917 | 5·848 | 10193 | 1888 | 11827 |
| Heilongjiang Province | Mingshui xian | 4·670 | 7·056 | 25·125 | 0·837 | 5·341 | 8887 | 1600 | 10142 |
| Heilongjiang Province | Mishanshi | 4·703 | 7·258 | 25·033 | 0·842 | 5·363 | 15634 | 2793 | 17820 |
| Heilongjiang Province | Mohe xian | 5·358 | 8·193 | 28·879 | 0·899 | 6·199 | 2748 | 459 | 3161 |
| Heilongjiang Province | Mudanjiangshi | 4·553 | 7·243 | 24·611 | 0·775 | 5·183 | 40807 | 6953 | 46086 |
| Heilongjiang Province | Mulan xian | 4·680 | 6·920 | 24·471 | 0·784 | 5·552 | 6331 | 1045 | 7601 |
| Heilongjiang Province | Muling xian | 4·575 | 7·272 | 25·171 | 0·780 | 5·124 | 9865 | 1667 | 11084 |
| Heilongjiang Province | Nehe xian | 4·907 | 7·425 | 27·394 | 0·874 | 5·447 | 23938 | 4225 | 26649 |
| Heilongjiang Province | Nenjiang xian | 5·064 | 7·701 | 28·523 | 0·884 | 5·625 | 17748 | 3097 | 19501 |
| Heilongjiang Province | Ningan xian | 4·507 | 7·270 | 24·791 | 0·783 | 5·020 | 15030 | 2636 | 16599 |
| Heilongjiang Province | Qingan xian | 4·827 | 7·097 | 25·042 | 0·842 | 5·683 | 12915 | 2225 | 15326 |
| Heilongjiang Province | Qinggang xian | 4·579 | 6·855 | 24·477 | 0·827 | 5·249 | 12124 | 2199 | 13833 |
| Heilongjiang Province | Qiqihaershi | 4·576 | 6·891 | 24·538 | 0·851 | 5·205 | 60806 | 11329 | 69742 |
| Heilongjiang Province | Qitaiheshi | 4·797 | 7·275 | 25·532 | 0·796 | 5·591 | 15124 | 2569 | 17695 |
| Heilongjiang Province | Raohe xian | 5·028 | 7·655 | 27·262 | 0·883 | 5·791 | 5644 | 972 | 6502 |
| Heilongjiang Province | Shangzhishi | 4·500 | 6·971 | 24·188 | 0·767 | 5·135 | 20813 | 3560 | 23626 |
| Heilongjiang Province | Shuangchengshi | 4·443 | 6·752 | 23·578 | 0·769 | 5·116 | 29047 | 4986 | 33445 |
| Heilongjiang Province | Shuileng xian | 4·883 | 7·201 | 25·632 | 0·864 | 5·702 | 11137 | 1939 | 13093 |
| Heilongjiang Province | Suangyashanshi | 4·959 | 7·430 | 26·761 | 0·867 | 5·700 | 18211 | 3202 | 21093 |
| Heilongjiang Province | Suibin xian | 5·095 | 7·715 | 27·971 | 0·914 | 5·780 | 6861 | 1223 | 7811 |
| Heilongjiang Province | Suifenheshi | 4·533 | 7·185 | 24·861 | 0·790 | 4·936 | 4822 | 857 | 5202 |
| Heilongjiang Province | Suihuashi | 4·669 | 6·831 | 24·447 | 0·832 | 5·477 | 32578 | 5764 | 37793 |
| Heilongjiang Province | Sunwu xian | 5·079 | 7·646 | 27·950 | 0·902 | 5·669 | 3321 | 592 | 3797 |
| Heilongjiang Province | Tahe xian | 5·233 | 8·025 | 29·201 | 0·928 | 5·896 | 2509 | 454 | 2907 |
| Heilongjiang Province | Tailai xian | 4·573 | 6·992 | 24·740 | 0·817 | 5·164 | 11130 | 1962 | 12594 |

| ADM1 | ADM2 | β-thalassaemia carrier prevalence Mean | SD | CI95% | Q25 | Q75 | Estimated carrier counts Mean | Q25 | Q75 |
|---|---|---|---|---|---|---|---|---|---|
| Heilongjiang Province | Tangyuan xian | 4·919 | 7·192 | 25·860 | 0·874 | 5·816 | 11450 | 1989 | 13727 |
| Heilongjiang Province | Tielishi | 4·839 | 7·141 | 25·155 | 0·839 | 5·719 | 12294 | 2144 | 14487 |
| Heilongjiang Province | Tonghe xian | 4·728 | 6·966 | 24·570 | 0·804 | 5·632 | 7127 | 1179 | 8652 |
| Heilongjiang Province | Tongjiangshi | 5·130 | 8·016 | 28·262 | 0·899 | 5·778 | 8494 | 1502 | 9581 |
| Heilongjiang Province | Wangkui xian | 4·657 | 6·827 | 24·448 | 0·843 | 5·433 | 12632 | 2320 | 14755 |
| Heilongjiang Province | Wuchangshi | 4·448 | 6·939 | 23·833 | 0·756 | 5·072 | 32496 | 5636 | 37343 |
| Heilongjiang Province | Wudalianchishi | 5·129 | 7·729 | 28·560 | 0·879 | 5·664 | 1350 | 228 | 1497 |
| Heilongjiang Province | Xunke xian | 5·051 | 7·510 | 27·483 | 0·920 | 5·761 | 4083 | 749 | 4709 |
| Heilongjiang Province | Yanshou xian | 4·523 | 6·791 | 24·195 | 0·768 | 5·277 | 8001 | 1348 | 9344 |
| Heilongjiang Province | Yian xian | 4·756 | 7·183 | 25·642 | 0·859 | 5·357 | 15792 | 2811 | 17645 |
| Heilongjiang Province | Yichunshi | 4·970 | 7·310 | 26·318 | 0·888 | 5·810 | 27002 | 4730 | 31618 |
| Heilongjiang Province | Yilan xian | 4·748 | 6·910 | 24·324 | 0·821 | 5·732 | 13767 | 2370 | 16643 |
| Heilongjiang Province | Youyi xian | 4·940 | 7·390 | 26·607 | 0·879 | 5·682 | 5372 | 960 | 6243 |
| Heilongjiang Province | Zhaodongshi | 4·464 | 6·769 | 23·866 | 0·790 | 5·043 | 29638 | 5227 | 33838 |
| Heilongjiang Province | Zhaoyuan xian | 4·493 | 6·821 | 24·181 | 0·782 | 5·125 | 14724 | 2561 | 16922 |
| Heilongjiang Province | Zhaozhou xian | 4·485 | 6·859 | 24·292 | 0·775 | 5·068 | 13348 | 2311 | 15140 |
| Henan Province | Anyang xian | 2·976 | 4·694 | 16·250 | 0·566 | 3·246 | 15441 | 2945 | 16745 |
| Henan Province | Anyang xian | 3·033 | 4·680 | 16·041 | 0·591 | 3·422 | 48193 | 9338 | 54780 |
| Henan Province | Anyangshi | 3·017 | 4·630 | 16·514 | 0·588 | 3·382 | 18634 | 3643 | 21190 |
| Henan Province | Baofeng xian | 2·983 | 4·046 | 14·221 | 0·762 | 3·454 | 24471 | 6285 | 28483 |
| Henan Province | Boai xian | 3·241 | 4·809 | 16·823 | 0·683 | 3·703 | 17946 | 3777 | 20353 |
| Henan Province | Cao xian | 2·302 | 3·485 | 11·896 | 0·478 | 2·599 | 35773 | 7385 | 40188 |
| Henan Province | Changgeshi | 2·823 | 3·998 | 14·700 | 0·661 | 3·219 | 18595 | 4400 | 21104 |
| Henan Province | Changyuan xian | 2·765 | 4·331 | 15·201 | 0·555 | 3·010 | 22896 | 4616 | 25310 |
| Henan Province | Ci xian | 3·113 | 4·821 | 16·767 | 0·586 | 3·470 | 23517 | 4482 | 26165 |
| Henan Province | Dan xian | 1·999 | 2·983 | 9·922 | 0·423 | 2·335 | 21711 | 4548 | 25167 |
| Henan Province | Dengzhoushi | 2·761 | 3·017 | 10·601 | 0·956 | 3·441 | 36397 | 12537 | 45181 |
| Henan Province | Dongfengshi | 3·117 | 4·324 | 15·551 | 0·734 | 3·593 | 20965 | 4875 | 23773 |
| Henan Province | Dongming xian | 2·640 | 4·124 | 14·499 | 0·531 | 2·889 | 18929 | 3796 | 20762 |
| Henan Province | Fangcheng xian | 2·840 | 3·629 | 12·744 | 0·809 | 3·373 | 24102 | 6988 | 28543 |
| Henan Province | Fengqiu xian | 2·807 | 4·334 | 15·029 | 0·569 | 3·100 | 22024 | 4483 | 24246 |
| Henan Province | Fugou xian | 2·557 | 3·626 | 12·676 | 0·602 | 2·946 | 14418 | 3380 | 16705 |
| Henan Province | Gong xian | 3·171 | 4·523 | 16·349 | 0·720 | 3·634 | 27632 | 6253 | 31593 |
| Henan Province | Guangshan xian | 2·010 | 2·236 | 7·790 | 0·702 | 2·470 | 12280 | 4248 | 15139 |
| Henan Province | Gushi xian | 1·630 | 1·923 | 6·650 | 0·518 | 1·983 | 15007 | 4752 | 18227 |
| Henan Province | Hebishi | 3·029 | 4·733 | 16·199 | 0·597 | 3·387 | 9133 | 1791 | 10267 |

| ADM1 | ADM2 | β-thalassaemia carrier prevalence Mean | SD | CI95% | Q25 | Q75 | Estimated carrier counts Mean | Q25 | Q75 |
|---|---|---|---|---|---|---|---|---|---|
| Henan Province | Hua xian | 2·805 | 4·372 | 14·989 | 0·559 | 3·107 | 31931 | 6436 | 35051 |
| Henan Province | Huaibin xian | 1·803 | 2·127 | 7·388 | 0·570 | 2·207 | 9484 | 2983 | 11552 |
| Henan Province | Huaiyang xian | 2·257 | 3·143 | 10·623 | 0·556 | 2·651 | 30644 | 7660 | 36069 |
| Henan Province | Huangchuan xian | 1·882 | 2·193 | 7·571 | 0·614 | 2·296 | 11981 | 3898 | 14738 |
| Henan Province | Huixianshi | 3·113 | 4·723 | 16·318 | 0·627 | 3·505 | 25526 | 5148 | 28517 |
| Henan Province | Huojia xian | 3·061 | 4·583 | 15·826 | 0·646 | 3·405 | 11747 | 2512 | 13019 |
| Henan Province | Jia xian | 2·973 | 4·078 | 14·670 | 0·742 | 3·426 | 15727 | 3899 | 18073 |
| Henan Province | Jiaozuoshi | 3·263 | 4·841 | 17·010 | 0·653 | 3·657 | 10824 | 2138 | 11987 |
| Henan Province | Jieshoushi | 1·860 | 2·442 | 8·125 | 0·505 | 2·258 | 15138 | 4096 | 18437 |
| Henan Province | Jinchengshi | 3·287 | 4·832 | 17·039 | 0·685 | 3·774 | 34711 | 7301 | 39960 |
| Henan Province | Jiyuanshi | 3·309 | 4·735 | 16·686 | 0·722 | 3·828 | 24062 | 5191 | 28119 |
| Henan Province | Jun xian | 2·932 | 4·648 | 15·905 | 0·570 | 3·229 | 27241 | 5295 | 30485 |
| Henan Province | Kaifeng xian | 2·732 | 4·127 | 14·647 | 0·582 | 3·015 | 26475 | 5663 | 29138 |
| Henan Province | Kaifengshi | 2·801 | 4·252 | 15·113 | 0·591 | 3·060 | 21230 | 4429 | 23488 |
| Henan Province | Lankao xian | 2·577 | 3·946 | 13·952 | 0·532 | 2·851 | 20678 | 4260 | 22889 |
| Henan Province | Lin xian | 3·129 | 4·775 | 16·609 | 0·610 | 3·545 | 28620 | 5534 | 32360 |
| Henan Province | Lingbaoshi | 3·198 | 4·320 | 14·378 | 0·787 | 3·880 | 21485 | 5261 | 25931 |
| Henan Province | Linquan xian | 1·891 | 2·369 | 8·142 | 0·546 | 2·302 | 29456 | 8437 | 35819 |
| Henan Province | Linying xian | 2·658 | 3·614 | 12·604 | 0·661 | 3·135 | 15483 | 3813 | 18116 |
| Henan Province | Luanchuan xian | 3·113 | 4·073 | 14·328 | 0·822 | 3·710 | 9736 | 2592 | 11558 |
| Henan Province | Luoheshi | 2·514 | 3·244 | 11·301 | 0·675 | 3·057 | 12177 | 3269 | 14815 |
| Henan Province | Luoning xian | 3·205 | 4·285 | 14·740 | 0·794 | 3·824 | 11909 | 2891 | 14286 |
| Henan Province | Luoshan xian | 2·150 | 2·322 | 8·192 | 0·765 | 2·672 | 9364 | 3238 | 11528 |
| Henan Province | Luoyangshi | 3·198 | 4·376 | 15·303 | 0·754 | 3·772 | 71489 | 16701 | 83833 |
| Henan Province | Lushan xian | 3·034 | 4·063 | 14·252 | 0·795 | 3·558 | 19823 | 5127 | 23257 |
| Henan Province | Lushi xian | 3·115 | 4·149 | 14·253 | 0·818 | 3·720 | 9722 | 2448 | 11798 |
| Henan Province | Luyi xian | 2·101 | 2·973 | 10·004 | 0·501 | 2·468 | 19508 | 4712 | 22770 |
| Henan Province | Meng xian | 3·217 | 4·545 | 16·091 | 0·720 | 3·687 | 12485 | 2798 | 14326 |
| Henan Province | Mengjin xian | 3·237 | 4·484 | 16·270 | 0·737 | 3·743 | 15814 | 3629 | 18289 |
| Henan Province | Mi xian | 2·987 | 4·218 | 14·997 | 0·699 | 3·395 | 24980 | 5917 | 28553 |
| Henan Province | Mianchi xian | 3·306 | 4·527 | 15·678 | 0·768 | 3·952 | 9927 | 2379 | 12118 |
| Henan Province | Minquan xian | 2·438 | 3·643 | 12·702 | 0·518 | 2·760 | 16697 | 3596 | 18937 |
| Henan Province | Miyang xian | 2·628 | 3·193 | 11·077 | 0·813 | 3·175 | 21227 | 6551 | 25783 |
| Henan Province | Nanle xian | 2·804 | 4·558 | 16·084 | 0·531 | 2·976 | 11770 | 2198 | 12422 |
| Henan Province | Nanyangshi | 2·819 | 3·360 | 11·678 | 0·889 | 3·424 | 35867 | 11361 | 43007 |
| Henan Province | Nanyangshi | 2·816 | 3·297 | 11·903 | 0·858 | 3·510 | 19765 | 5982 | 24513 |

| ADM1 | ADM2 | β-thalassaemia carrier prevalence Mean | SD | CI95% | Q25 | Q75 | Estimated carrier counts Mean | Q25 | Q75 |
|---|---|---|---|---|---|---|---|---|---|
| Henan Province | Nanzhao xian | 2·965 | 3·766 | 13·050 | 0·832 | 3·571 | 15049 | 4242 | 18003 |
| Henan Province | Neihuang xian | 2·893 | 4·632 | 16·261 | 0·553 | 3·132 | 20953 | 4018 | 22619 |
| Henan Province | Neixiang xian | 2·909 | 3·503 | 12·268 | 0·888 | 3·534 | 16154 | 5076 | 19503 |
| Henan Province | Ningling xian | 2·336 | 3·462 | 11·989 | 0·502 | 2·675 | 12335 | 2698 | 14211 |
| Henan Province | Pingdingshanshi | 2·895 | 4·062 | 13·975 | 0·722 | 3·299 | 16938 | 4207 | 19209 |
| Henan Province | Pingyu xian | 2·167 | 2·751 | 9·274 | 0·627 | 2·621 | 18155 | 5213 | 21782 |
| Henan Province | Puyangshi | 2·772 | 4·371 | 15·521 | 0·532 | 2·990 | 25671 | 4923 | 27555 |
| Henan Province | Qi xian | 2·987 | 4·656 | 16·033 | 0·585 | 3·331 | 11216 | 2202 | 12500 |
| Henan Province | Qi xian | 2·540 | 3·765 | 13·151 | 0·562 | 2·833 | 21195 | 4646 | 23589 |
| Henan Province | Qingfeng xian | 2·760 | 4·408 | 15·372 | 0·540 | 3·000 | 26122 | 5112 | 28453 |
| Henan Province | Qinyang xian | 3·259 | 4·732 | 16·693 | 0·701 | 3·685 | 10690 | 2342 | 12024 |
| Henan Province | Queshan xian | 2·410 | 2·874 | 10·078 | 0·759 | 2·924 | 26099 | 8087 | 32093 |
| Henan Province | Runan xian | 2·282 | 2·815 | 9·718 | 0·684 | 2·770 | 13802 | 4094 | 16834 |
| Henan Province | Ruyangshi | 3·151 | 4·300 | 15·137 | 0·789 | 3·716 | 9300 | 2285 | 11070 |
| Henan Province | Ruzhoushi | 3·102 | 4·282 | 15·160 | 0·755 | 3·603 | 31526 | 7727 | 36518 |
| Henan Province | Sanmenxiashi | 3·288 | 4·474 | 14·624 | 0·787 | 4·042 | 4480 | 1031 | 5400 |
| Henan Province | Shan xian | 3·298 | 4·509 | 15·460 | 0·766 | 3·952 | 11992 | 2783 | 14564 |
| Henan Province | Shancheng xian | 2·047 | 2·829 | 9·658 | 0·507 | 2·422 | 20361 | 4995 | 24340 |
| Henan Province | Shangcai xian | 2·328 | 3·068 | 10·317 | 0·633 | 2·799 | 23261 | 6387 | 28036 |
| Henan Province | Shangcheng xian | 1·761 | 1·948 | 6·825 | 0·601 | 2·184 | 8302 | 2759 | 10255 |
| Henan Province | Shangqiushi | 2·108 | 3·181 | 10·523 | 0·465 | 2·393 | 10412 | 2328 | 11744 |
| Henan Province | Shangqiushi | 2·113 | 3·082 | 10·332 | 0·470 | 2·475 | 29932 | 6714 | 35315 |
| Henan Province | Shangshui xian | 2·332 | 3·156 | 10·638 | 0·612 | 2·766 | 24762 | 6515 | 29340 |
| Henan Province | Shenqiu xian | 2·020 | 2·698 | 8·939 | 0·530 | 2·433 | 20185 | 5332 | 24217 |
| Henan Province | Sheqi xian | 2·738 | 3·329 | 11·488 | 0·849 | 3·314 | 14473 | 4479 | 17456 |
| Henan Province | Shui xian | 2·409 | 3·517 | 12·361 | 0·545 | 2·752 | 16029 | 3613 | 18180 |
| Henan Province | Shuiping xian | 2·498 | 3·165 | 10·932 | 0·721 | 2·996 | 13277 | 3843 | 15875 |
| Henan Province | Song xian | 3·141 | 4·173 | 14·715 | 0·810 | 3·713 | 17340 | 4325 | 20302 |
| Henan Province | Taikang xian | 2·399 | 3·408 | 11·680 | 0·557 | 2·796 | 25233 | 5882 | 29541 |
| Henan Province | Tanghe xian | 2·707 | 3·088 | 10·754 | 0·909 | 3·315 | 32309 | 10776 | 39413 |
| Henan Province | Tangyin xian | 2·938 | 4·633 | 15·908 | 0·572 | 3·258 | 13458 | 2638 | 14862 |
| Henan Province | Tongbo xian | 2·517 | 2·818 | 9·896 | 0·870 | 3·067 | 10550 | 3671 | 12676 |
| Henan Province | Tongxushi | 2·645 | 3·938 | 13·783 | 0·588 | 2·968 | 15662 | 3492 | 17476 |
| Henan Province | Tuocheng xian | 2·181 | 3·088 | 10·551 | 0·505 | 2·573 | 18766 | 4314 | 22296 |
| Henan Province | Wei xian | 2·912 | 4·660 | 16·118 | 0·564 | 3·172 | 23503 | 4559 | 25635 |
| Henan Province | Weihuishi | 2·988 | 4·560 | 15·713 | 0·605 | 3·343 | 18678 | 3771 | 20866 |

| | | β-thalassaemia carrier prevalence | | | | | Estimated carrier counts | | |
|---|---|---|---|---|---|---|---|---|---|
| **ADM1** | **ADM2** | **Mean** | **SD** | **CI95%** | **Q25** | **Q75** | **Mean** | **Q25** | **Q75** |
| Henan Province | Weishi xian | 2·765 | 4·031 | 14·298 | 0·625 | 3·111 | 23488 | 5282 | 26323 |
| Henan Province | Wen xian | 3·206 | 4·686 | 16·299 | 0·709 | 3·614 | 14365 | 3117 | 15955 |
| Henan Province | Wugangshi | 2·664 | 3·498 | 12·118 | 0·729 | 3·176 | 19918 | 5488 | 23771 |
| Henan Province | Wushi xian | 3·101 | 4·488 | 16·008 | 0·676 | 3·546 | 17127 | 3753 | 19546 |
| Henan Province | Xi xian | 1·994 | 2·342 | 8·117 | 0·642 | 2·442 | 12675 | 4148 | 15752 |
| Henan Province | Xiangcheng xian | 2·822 | 3·859 | 13·707 | 0·707 | 3·289 | 21602 | 5447 | 25088 |
| Henan Province | Xiangchengshi | 2·103 | 2·728 | 9·352 | 0·572 | 2·544 | 16422 | 4413 | 19839 |
| Henan Province | Xihua xian | 2·468 | 3·436 | 11·686 | 0·607 | 2·893 | 15729 | 3811 | 18138 |
| Henan Province | Xin xian | 1·959 | 2·038 | 7·142 | 0·722 | 2·455 | 5980 | 2159 | 7547 |
| Henan Province | Xinan xian | 3·296 | 4·566 | 16·274 | 0·757 | 3·873 | 13292 | 3140 | 15608 |
| Henan Province | Xincai xian | 2·022 | 2·483 | 8·573 | 0·600 | 2·468 | 15069 | 4534 | 18388 |
| Henan Province | Xingyang xian | 3·028 | 4·311 | 15·338 | 0·677 | 3·457 | 22536 | 5031 | 25804 |
| Henan Province | Xinxiang xian | 2·995 | 4·479 | 15·441 | 0·618 | 3·342 | 38352 | 7924 | 42705 |
| Henan Province | Xinxiangshi | 2·965 | 4·276 | 15·232 | 0·614 | 3·419 | 3454 | 718 | 3986 |
| Henan Province | Xinyangshi | 2·306 | 2·515 | 8·842 | 0·821 | 2·846 | 35634 | 12900 | 44371 |
| Henan Province | Xinye xian | 2·709 | 2·902 | 10·269 | 0·962 | 3·358 | 16741 | 5956 | 20745 |
| Henan Province | Xinzheng xian | 2·873 | 4·085 | 14·611 | 0·667 | 3·266 | 23176 | 5437 | 26383 |
| Henan Province | Xiping xian | 2·525 | 3·275 | 11·376 | 0·695 | 3·011 | 16446 | 4563 | 19939 |
| Henan Province | Xiuwu xian | 3·158 | 4·739 | 16·285 | 0·656 | 3·593 | 28373 | 5786 | 32377 |
| Henan Province | Xixia xian | 2·996 | 3·792 | 13·059 | 0·856 | 3·620 | 13492 | 4009 | 16264 |
| Henan Province | Xuchang xian | 2·805 | 3·926 | 13·921 | 0·670 | 3·248 | 31170 | 7465 | 36351 |
| Henan Province | Xuchangshi | 2·751 | 3·900 | 13·779 | 0·708 | 3·084 | 6482 | 1669 | 7267 |
| Henan Province | Yancheng xian | 2·558 | 3·473 | 11·792 | 0·667 | 3·011 | 20629 | 5455 | 24287 |
| Henan Province | Yanling xian | 2·652 | 3·770 | 13·135 | 0·631 | 3·043 | 16996 | 4087 | 19629 |
| Henan Province | Yanshishi | 3·204 | 4·472 | 15·972 | 0·741 | 3·714 | 20062 | 4589 | 23104 |
| Henan Province | Ye xian | 2·816 | 3·728 | 13·230 | 0·755 | 3·314 | 26456 | 7013 | 30720 |
| Henan Province | Yichuan xian | 3·179 | 4·323 | 15·534 | 0·771 | 3·738 | 28827 | 7040 | 33906 |
| Henan Province | Yimashi | 3·315 | 4·358 | 16·530 | 0·824 | 3·838 | 1103 | 274 | 1277 |
| Henan Province | Yiyang xian | 3·275 | 4·461 | 15·785 | 0·780 | 3·859 | 21973 | 5143 | 25945 |
| Henan Province | Yuanjin xian | 2·905 | 4·416 | 15·308 | 0·591 | 3·257 | 18574 | 3813 | 20735 |
| Henan Province | Yuanyangshi | 2·941 | 4·393 | 15·394 | 0·622 | 3·303 | 26582 | 5611 | 29850 |
| Henan Province | Yucheng xian | 1·960 | 2·817 | 9·536 | 0·436 | 2·303 | 17274 | 3876 | 20400 |
| Henan Province | Yuncheng xian | 3·253 | 4·463 | 14·634 | 0·770 | 3·970 | 11818 | 2817 | 14373 |
| Henan Province | Yuzhoushi | 2·993 | 4·205 | 14·771 | 0·702 | 3·458 | 34185 | 8036 | 39586 |
| Henan Province | Zhengyang xian | 2·154 | 2·499 | 8·728 | 0·694 | 2·643 | 13280 | 4282 | 16266 |
| Henan Province | Zhengzhoushi | 2·942 | 4·202 | 15·164 | 0·644 | 3·378 | 209997 | 45910 | 239213 |

| ADM1 | ADM2 | β-thalassaemia carrier prevalence Mean | SD | CI95% | Q25 | Q75 | Estimated carrier counts Mean | Q25 | Q75 |
|---|---|---|---|---|---|---|---|---|---|
| Henan Province | Zhenping xian | 2·849 | 3·394 | 11·822 | 0·888 | 3·467 | 22612 | 7111 | 27617 |
| Henan Province | Zhongmou xian | 2·833 | 4·131 | 14·832 | 0·628 | 3·224 | 47194 | 10413 | 53672 |
| Henan Province | Zhoukoushi | 2·224 | 2·764 | 9·457 | 0·608 | 2·868 | 9464 | 2563 | 12162 |
| Henan Province | Zhumadianshi | 2·361 | 2·846 | 10·175 | 0·714 | 2·841 | 5124 | 1565 | 6186 |
| Hong Kong Special Administrative Region | Xianggang | 3·401 | 2·448 | 9·037 | 1·768 | 4·287 | 144427 | 74272 | 183282 |
| Hubei Province | Anlushi | 2·264 | 1·923 | 7·017 | 1·004 | 2·885 | 11798 | 5336 | 15128 |
| Hubei Province | Badong xian | 2·030 | 1·596 | 5·853 | 0·984 | 2·567 | 8429 | 4077 | 10709 |
| Hubei Province | Baihe xian | 2·534 | 2·914 | 9·901 | 0·852 | 3·129 | 2328 | 786 | 2861 |
| Hubei Province | Baokang xian | 2·486 | 2·245 | 8·106 | 1·054 | 3·150 | 5587 | 2348 | 7064 |
| Hubei Province | Changyangtujia zuzizhi xian | 2·005 | 1·587 | 5·778 | 0·966 | 2·531 | 5824 | 2834 | 7321 |
| Hubei Province | Chongyang xian | 2·211 | 1·706 | 6·238 | 1·085 | 2·788 | 9220 | 4554 | 11644 |
| Hubei Province | Dangyangshi | 2·242 | 1·784 | 6·585 | 1·072 | 2·835 | 8796 | 4176 | 11194 |
| Hubei Province | Danjiangkoushi | 2·699 | 2·849 | 10·058 | 0·980 | 3·356 | 13764 | 4979 | 17194 |
| Hubei Province | Dawu xian | 2·134 | 2·033 | 7·363 | 0·851 | 2·689 | 10554 | 4211 | 13242 |
| Hubei Province | Dazhishi | 1·979 | 1·641 | 5·889 | 0·923 | 2·498 | 17808 | 8238 | 22268 |
| Hubei Province | Enshishi | 1·760 | 1·474 | 5·353 | 0·805 | 2·230 | 14987 | 6832 | 18756 |
| Hubei Province | Ezhoushi | 1·971 | 1·602 | 5·805 | 0·934 | 2·488 | 14009 | 6513 | 17639 |
| Hubei Province | Fang xian | 2·483 | 2·423 | 8·584 | 0·989 | 3·127 | 9001 | 3602 | 11498 |
| Hubei Province | Guangshuishi | 2·299 | 2·155 | 7·840 | 0·925 | 2·916 | 15597 | 6328 | 19840 |
| Hubei Province | Gucheng xian | 2·643 | 2·562 | 9·195 | 1·046 | 3·331 | 11440 | 4436 | 14607 |
| Hubei Province | Hanchuan xian | 2·171 | 1·669 | 6·155 | 1·074 | 2·740 | 18459 | 9113 | 23391 |
| Hubei Province | Hefeng xian | 1·742 | 1·381 | 5·039 | 0·842 | 2·198 | 2856 | 1376 | 3620 |
| Hubei Province | Hongan xian | 2·003 | 1·876 | 6·730 | 0·822 | 2·522 | 9854 | 4089 | 12433 |
| Hubei Province | Honghushi | 2·132 | 1·585 | 5·846 | 1·085 | 2·680 | 15599 | 7947 | 19582 |
| Hubei Province | Huangmei xian | 1·694 | 1·587 | 5·756 | 0·696 | 2·127 | 16212 | 6763 | 20278 |
| Hubei Province | Huangpi xian | 2·081 | 1·800 | 6·542 | 0·930 | 2·622 | 28575 | 13131 | 36131 |
| Hubei Province | Huangshishi | 1·918 | 1·604 | 5·741 | 0·871 | 2·451 | 9327 | 4221 | 11836 |
| Hubei Province | Huangzhoushi | 1·898 | 1·690 | 6·105 | 0·823 | 2·404 | 19028 | 8429 | 24237 |
| Hubei Province | Jiangling xian | 2·121 | 1·641 | 6·026 | 1·035 | 2·683 | 22005 | 10646 | 27839 |
| Hubei Province | Jianli xian | 2·076 | 1·535 | 5·671 | 1·052 | 2·623 | 23620 | 11932 | 29752 |
| Hubei Province | Jianshi xian | 1·897 | 1·528 | 5·512 | 0·906 | 2·394 | 7809 | 3688 | 9839 |
| Hubei Province | Jiayu xian | 2·108 | 1·573 | 5·739 | 1·077 | 2·645 | 4095 | 2076 | 5172 |
| Hubei Province | Jingmenshi | 2·279 | 1·811 | 6·665 | 1·100 | 2·874 | 25811 | 12581 | 32604 |
| Hubei Province | Jingshan xian | 2·281 | 1·800 | 6·616 | 1·096 | 2·888 | 11793 | 5777 | 14892 |
| Hubei Province | Laohekoushi | 2·682 | 2·690 | 9·598 | 0·996 | 3·380 | 9878 | 3826 | 12228 |

| | | β-thalassaemia carrier prevalence | | | | | Estimated carrier counts | | |
|---|---|---|---|---|---|---|---|---|---|
| ADM1 | ADM2 | Mean | SD | CI95% | Q25 | Q75 | Mean | Q25 | Q75 |
| Hubei Province | Lichuanshi | 1·642 | 1·494 | 5·378 | 0·703 | 2·062 | 12407 | 5365 | 15447 |
| Hubei Province | Luotian xian | 1·708 | 1·677 | 5·894 | 0·665 | 2·161 | 8058 | 3196 | 10180 |
| Hubei Province | Machengshi | 1·852 | 1·828 | 6·503 | 0·722 | 2·320 | 16347 | 6416 | 20467 |
| Hubei Province | Nanzhang xian | 2·511 | 2·225 | 8·069 | 1·082 | 3·173 | 10980 | 4788 | 13763 |
| Hubei Province | Puxishi | 2·150 | 1·591 | 5·907 | 1·100 | 2·704 | 10092 | 5095 | 12734 |
| Hubei Province | Qianjiangshi | 2·144 | 1·615 | 6·016 | 1·070 | 2·711 | 18519 | 9224 | 23328 |
| Hubei Province | Qichun xian | 1·749 | 1·632 | 5·863 | 0·720 | 2·202 | 15276 | 6396 | 19260 |
| Hubei Province | Shashi | 2·107 | 1·627 | 5·998 | 1·028 | 2·694 | 11206 | 5483 | 14341 |
| Hubei Province | Shennongjialinqu | 2·288 | 2·034 | 7·301 | 1·001 | 2·881 | 1532 | 650 | 1882 |
| Hubei Province | Shiyanshi | 2·678 | 2·938 | 10·265 | 0·935 | 3·309 | 25236 | 8453 | 31437 |
| Hubei Province | Shuizhoushi | 2·410 | 2·289 | 8·237 | 0·978 | 3·017 | 30594 | 12818 | 38416 |
| Hubei Province | Tianmenshi | 2·195 | 1·693 | 6·221 | 1·084 | 2·769 | 31108 | 15390 | 39266 |
| Hubei Province | Tongshan xian | 2·144 | 1·688 | 6·155 | 1·034 | 2·704 | 8503 | 4129 | 10725 |
| Hubei Province | Wufengtujia zuzizhi xian | 1·899 | 1·507 | 5·487 | 0·916 | 2·401 | 2760 | 1318 | 3491 |
| Hubei Province | Wuhanshi | 2·077 | 1·586 | 5·829 | 1·040 | 2·614 | 202328 | 99540 | 254466 |
| Hubei Province | Wuxueshi | 1·809 | 1·684 | 6·026 | 0·768 | 2·253 | 13136 | 5616 | 16295 |
| Hubei Province | Xiangfanshi | 2·586 | 2·290 | 8·413 | 1·058 | 3·363 | 15180 | 6369 | 19822 |
| Hubei Province | Xiangyang xian | 2·615 | 2·508 | 8·990 | 1·042 | 3·293 | 46296 | 18256 | 57951 |
| Hubei Province | Xianningshi | 2·096 | 1·600 | 5·921 | 1·038 | 2·656 | 14372 | 7168 | 18150 |
| Hubei Province | Xiantaoshi | 2·150 | 1·617 | 5·913 | 1·084 | 2·707 | 21816 | 10977 | 27450 |
| Hubei Province | Xiaoganshi | 2·179 | 1·843 | 6·690 | 0·985 | 2·755 | 32799 | 14995 | 41904 |
| Hubei Province | Xichuan xian | 2·836 | 3·285 | 11·604 | 0·919 | 3·465 | 14537 | 4574 | 17651 |
| Hubei Province | Xingshan xian | 2·290 | 1·942 | 7·050 | 1·039 | 2·888 | 2923 | 1300 | 3680 |
| Hubei Province | Xinzhou xian | 1·945 | 1·692 | 6·140 | 0·853 | 2·465 | 18449 | 8195 | 23394 |
| Hubei Province | Xishui xian | 1·824 | 1·655 | 5·941 | 0·768 | 2·303 | 16926 | 7295 | 21354 |
| Hubei Province | Xuanen xian | 1·684 | 1·402 | 5·113 | 0·780 | 2·133 | 4945 | 2319 | 6242 |
| Hubei Province | Yangxin xian | 1·992 | 1·706 | 6·133 | 0·916 | 2·490 | 15363 | 6992 | 19100 |
| Hubei Province | Yichang xian | 2·213 | 1·800 | 6·606 | 1·037 | 2·796 | 23121 | 11069 | 29139 |
| Hubei Province | Yichangshi | 2·136 | 1·684 | 6·278 | 1·017 | 2·707 | 10072 | 4792 | 12849 |
| Hubei Province | Yichengshi | 2·477 | 2·134 | 7·809 | 1·094 | 3·127 | 12125 | 5339 | 15325 |
| Hubei Province | Yingchengshi | 2·249 | 1·830 | 6·631 | 1·064 | 2·840 | 9739 | 4592 | 12194 |
| Hubei Province | Yingshan xian | 1·606 | 1·632 | 5·804 | 0·600 | 2·008 | 4867 | 1863 | 6097 |
| Hubei Province | Yuanan xian | 2·336 | 1·926 | 7·074 | 1·081 | 2·950 | 3664 | 1721 | 4621 |
| Hubei Province | Yun xian | 2·748 | 3·187 | 10·969 | 0·901 | 3·379 | 12121 | 3867 | 14895 |
| Hubei Province | Yunmeng xian | 2·233 | 1·813 | 6·562 | 1·051 | 2·825 | 10325 | 4828 | 13139 |
| Hubei Province | Yunxi xian | 2·693 | 3·239 | 10·926 | 0·861 | 3·289 | 10293 | 3335 | 12572 |

| | | β-thalassaemia carrier prevalence | | | | | Estimated carrier counts | | |
|---|---|---|---|---|---|---|---|---|---|
| ADM1 | ADM2 | Mean | SD | CI95% | Q25 | Q75 | Mean | Q25 | Q75 |
| Hubei Province | Zhaoyangshi | 2·530 | 2·412 | 8·803 | 1·010 | 3·171 | 22277 | 8751 | 27901 |
| Hubei Province | Zhichengshi | 2·014 | 1·597 | 5·856 | 0·963 | 2·544 | 7540 | 3562 | 9582 |
| Hubei Province | Zhigui xian | 2·101 | 1·661 | 6·007 | 1·020 | 2·654 | 5446 | 2670 | 7047 |
| Hubei Province | Zhijiang xian | 2·133 | 1·720 | 6·286 | 1·007 | 2·696 | 10227 | 4808 | 12938 |
| Hubei Province | Zhongxiangshi | 2·345 | 1·847 | 6·784 | 1·133 | 2·963 | 20058 | 9821 | 25366 |
| Hubei Province | Zhushan xian | 2·414 | 2·490 | 8·690 | 0·909 | 3·013 | 8681 | 3233 | 10925 |
| Hubei Province | Zhuxi xian | 2·281 | 2·325 | 8·164 | 0·878 | 2·839 | 6691 | 2420 | 8352 |
| Hunan Province | Anhua xian | 1·990 | 1·427 | 5·258 | 1·043 | 2·494 | 14623 | 7678 | 18388 |
| Hunan Province | Anren xian | 4·138 | 2·910 | 10·910 | 2·157 | 5·245 | 14765 | 7716 | 18742 |
| Hunan Province | Anxiang xian | 1·899 | 1·386 | 5·189 | 0·971 | 2·397 | 8115 | 4160 | 10251 |
| Hunan Province | Baojing xian | 1·635 | 1·281 | 4·689 | 0·796 | 2·062 | 4218 | 2057 | 5356 |
| Hunan Province | Chaling xian | 3·904 | 2·819 | 10·475 | 1·998 | 4·968 | 20502 | 10535 | 26089 |
| Hunan Province | Changdeshi | 1·803 | 1·307 | 4·819 | 0·936 | 2·268 | 25281 | 13060 | 31746 |
| Hunan Province | Changning xian | 4·520 | 3·158 | 11·762 | 2·363 | 5·759 | 35224 | 18405 | 44734 |
| Hunan Province | Changsha xian | 2·348 | 1·560 | 5·798 | 1·287 | 2·951 | 51670 | 28622 | 64678 |
| Hunan Province | Changshashi | 2·310 | 1·498 | 5·601 | 1·309 | 2·908 | 70733 | 40072 | 88534 |
| Hunan Province | Cheng xian | 5·345 | 3·573 | 13·371 | 2·881 | 6·805 | 30234 | 16298 | 38542 |
| Hunan Province | Chengbumiao zuzizhi xian | 4·279 | 3·333 | 12·362 | 2·051 | 5·444 | 9717 | 4687 | 12305 |
| Hunan Province | Chengzhoushi | 5·335 | 3·663 | 13·483 | 2·838 | 6·802 | 25560 | 13706 | 32549 |
| Hunan Province | Chenxi xian | 2·065 | 1·510 | 5·508 | 1·072 | 2·598 | 8008 | 4237 | 10084 |
| Hunan Province | Chili xian | 1·693 | 1·274 | 4·666 | 0·852 | 2·132 | 9860 | 4945 | 12427 |
| Hunan Province | Dao xian | 5·953 | 4·159 | 15·657 | 3·086 | 7·614 | 39088 | 20273 | 49972 |
| Hunan Province | Dayongshi | 1·595 | 1·175 | 4·320 | 0·817 | 2·007 | 8520 | 4398 | 10723 |
| Hunan Province | Dongan xian | 4·307 | 2·974 | 11·100 | 2·273 | 5·453 | 20375 | 10711 | 25763 |
| Hunan Province | Dongkou xian | 3·004 | 2·219 | 8·221 | 1·534 | 3·780 | 20628 | 10638 | 25944 |
| Hunan Province | Fenghuang xian | 1·931 | 1·517 | 5·542 | 0·937 | 2·440 | 6990 | 3407 | 8858 |
| Hunan Province | Gongan xian | 2·033 | 1·575 | 5·787 | 0·995 | 2·572 | 13465 | 6567 | 17097 |
| Hunan Province | Guidong xian | 5·063 | 3·551 | 13·354 | 2·639 | 6·454 | 7552 | 3964 | 9532 |
| Hunan Province | Guiyang xian | 5·216 | 3·543 | 13·227 | 2·791 | 6·606 | 37589 | 20274 | 47450 |
| Hunan Province | Guzhang xian | 1·662 | 1·230 | 4·488 | 0·847 | 2·102 | 1597 | 813 | 2026 |
| Hunan Province | Hanshou xian | 1·845 | 1·286 | 4·773 | 0·984 | 2·317 | 15012 | 8037 | 18791 |
| Hunan Province | Hengdong xian | 3·432 | 2·334 | 8·719 | 1·842 | 4·339 | 19869 | 10706 | 25105 |
| Hunan Province | Hengnan xian | 3·826 | 2·614 | 9·644 | 2·056 | 4·831 | 32817 | 17538 | 41418 |
| Hunan Province | Hengshan xian | 3·106 | 2·082 | 7·718 | 1·692 | 3·915 | 10673 | 5781 | 13427 |
| Hunan Province | Hengyang xian | 3·350 | 2·255 | 8·441 | 1·816 | 4·230 | 38714 | 20960 | 48977 |
| Hunan Province | Hengyangshi | 3·640 | 2·612 | 9·680 | 1·877 | 4·600 | 32923 | 17047 | 41591 |

| | | β-thalassaemia carrier prevalence | | | | | Estimated carrier counts | | |
|---|---|---|---|---|---|---|---|---|---|
| ADM1 | ADM2 | Mean | SD | CI95% | Q25 | Q75 | Mean | Q25 | Q75 |
| Hunan Province | Hongjiangshi | 2·703 | 2·056 | 7·586 | 1·340 | 3·417 | 10626 | 5299 | 13406 |
| Hunan Province | Huaihuashi | 2·340 | 1·751 | 6·430 | 1·184 | 2·953 | 22651 | 11150 | 28415 |
| Hunan Province | Huarong xian | 2·007 | 1·502 | 5·523 | 1·006 | 2·536 | 11563 | 5766 | 14665 |
| Hunan Province | Huayuan xian | 1·723 | 1·350 | 4·951 | 0·839 | 2·165 | 3882 | 1880 | 4902 |
| Hunan Province | Huitong xian | 3·117 | 2·534 | 9·275 | 1·451 | 3·951 | 8609 | 3983 | 10971 |
| Hunan Province | Jiahe xian | 5·663 | 3·889 | 14·546 | 2·980 | 7·202 | 20044 | 10592 | 25458 |
| Hunan Province | Jinshishi | 1·978 | 1·595 | 5·846 | 0·934 | 2·558 | 371 | 175 | 480 |
| Hunan Province | Jishoushi | 1·753 | 1·332 | 4·886 | 0·876 | 2·200 | 7369 | 3671 | 9353 |
| Hunan Province | Lanshan xian | 6·077 | 4·305 | 16·095 | 3·114 | 7·780 | 21630 | 11015 | 27905 |
| Hunan Province | Leiyangshi | 4·330 | 2·944 | 10·962 | 2·316 | 5·490 | 52303 | 27939 | 66596 |
| Hunan Province | Lengshuijiangshi | 2·552 | 1·807 | 6·554 | 1·355 | 3·204 | 8800 | 4643 | 11066 |
| Hunan Province | Lengshuitanshi | 4·263 | 2·924 | 10·894 | 2·271 | 5·418 | 25925 | 13909 | 33052 |
| Hunan Province | Li xian | 1·903 | 1·458 | 5·358 | 0·938 | 2·407 | 16542 | 8056 | 20962 |
| Hunan Province | Lianyuanshi | 2·511 | 1·720 | 6·371 | 1·361 | 3·145 | 24151 | 13075 | 30271 |
| Hunan Province | Lilingshi | 2·918 | 2·018 | 7·507 | 1·547 | 3·690 | 25270 | 13312 | 32114 |
| Hunan Province | Ling xian | 4·517 | 3·239 | 12·117 | 2·305 | 5·757 | 7153 | 3605 | 9118 |
| Hunan Province | Linli xian | 1·830 | 1·370 | 5·093 | 0·914 | 2·311 | 7623 | 3775 | 9669 |
| Hunan Province | Linwu xian | 5·960 | 4·120 | 15·509 | 3·135 | 7·584 | 20746 | 10924 | 26640 |
| Hunan Province | Linxiangshi | 2·105 | 1·507 | 5·592 | 1·098 | 2·651 | 8395 | 4400 | 10556 |
| Hunan Province | Liuxiang xian | 2·228 | 1·488 | 5·529 | 1·227 | 2·798 | 26995 | 14851 | 33989 |
| Hunan Province | Liuyang xian | 2·571 | 1·821 | 6·772 | 1·348 | 3·244 | 37425 | 19772 | 47210 |
| Hunan Province | Longhui xian | 2·804 | 1·993 | 7·392 | 1·468 | 3·532 | 29625 | 15575 | 37045 |
| Hunan Province | Longshan xian | 1·607 | 1·291 | 4·734 | 0·763 | 2·036 | 7344 | 3461 | 9317 |
| Hunan Province | Loudishi | 2·546 | 1·736 | 6·479 | 1·376 | 3·204 | 17007 | 9183 | 21338 |
| Hunan Province | Luxi xian | 1·849 | 1·360 | 4·974 | 0·951 | 2·322 | 4473 | 2303 | 5550 |
| Hunan Province | Mayangmiao zuzizhi xian | 2·088 | 1·586 | 5·833 | 1·036 | 2·643 | 6357 | 3241 | 7986 |
| Hunan Province | Miluoshi | 2·167 | 1·496 | 5·560 | 1·162 | 2·726 | 13865 | 7461 | 17458 |
| Hunan Province | Nan xian | 1·965 | 1·436 | 5·266 | 1·016 | 2·484 | 11233 | 5731 | 14156 |
| Hunan Province | Ninggang xian | 4·133 | 3·100 | 11·696 | 2·054 | 5·231 | 2538 | 1270 | 3212 |
| Hunan Province | Ningyuan xian | 5·754 | 4·057 | 15·199 | 2·967 | 7·360 | 38313 | 19670 | 49038 |
| Hunan Province | Pingjiang xian | 2·344 | 1·702 | 6·266 | 1·212 | 2·959 | 21942 | 11320 | 27677 |
| Hunan Province | Qidong xian | 3·788 | 2·581 | 9·709 | 2·026 | 4·793 | 29435 | 15725 | 37222 |
| Hunan Province | Qiyang xian | 4·467 | 3·089 | 11·524 | 2·362 | 5·652 | 33794 | 17834 | 42897 |
| Hunan Province | Shangzhi xian | 1·658 | 1·293 | 4·736 | 0·811 | 2·095 | 6163 | 3084 | 7742 |
| Hunan Province | Shaoyangshi | 3·125 | 2·227 | 7·747 | 1·666 | 3·982 | 16518 | 8737 | 21172 |
| Hunan Province | Shimen xian | 1·822 | 1·419 | 5·182 | 0·892 | 2·299 | 9729 | 4729 | 12240 |

| | | β-thalassaemia carrier prevalence | | | | | Estimated carrier counts | | |
|---|---|---|---|---|---|---|---|---|---|
| ADM1 | ADM2 | Mean | SD | CI95% | Q25 | Q75 | Mean | Q25 | Q75 |
| Hunan Province | Shishoushi | 2·022 | 1·544 | 5·647 | 1·010 | 2·545 | 10179 | 5074 | 12801 |
| Hunan Province | Shuangfeng xian | 2·888 | 1·952 | 7·204 | 1·575 | 3·636 | 21511 | 11722 | 27054 |
| Hunan Province | Shuangpai xian | 5·403 | 3·656 | 13·674 | 2·893 | 6·880 | 5143 | 2758 | 6505 |
| Hunan Province | Songzi xian | 2·000 | 1·550 | 5·701 | 0·971 | 2·531 | 12714 | 6186 | 16123 |
| Hunan Province | Suining xian | 3·521 | 2·724 | 10·004 | 1·714 | 4·459 | 9316 | 4538 | 11813 |
| Hunan Province | Taojiang xian | 1·953 | 1·343 | 4·999 | 1·049 | 2·456 | 11876 | 6398 | 14876 |
| Hunan Province | Taoyuan xian | 1·749 | 1·276 | 4·712 | 0·903 | 2·201 | 13381 | 6999 | 16906 |
| Hunan Province | Tongcheng xian | 2·234 | 1·687 | 6·158 | 1·124 | 2·808 | 10473 | 5288 | 13070 |
| Hunan Province | Tongzhoudong zuzizhi xian | 4·305 | 3·540 | 13·101 | 1·954 | 5·504 | 8577 | 3889 | 10944 |
| Hunan Province | Wangcheng xian | 2·216 | 1·470 | 5·421 | 1·230 | 2·775 | 47190 | 26438 | 59151 |
| Hunan Province | Wugangshi | 3·549 | 2·589 | 9·562 | 1·822 | 4·499 | 22066 | 11416 | 28072 |
| Hunan Province | Xiangtan xian | 2·695 | 1·801 | 6·723 | 1·479 | 3·389 | 47235 | 26307 | 59224 |
| Hunan Province | Xiangtanshi | 2·459 | 1·618 | 5·925 | 1·364 | 3·085 | 2502 | 1380 | 3143 |
| Hunan Province | Xiangxiangshi | 2·531 | 1·695 | 6·279 | 1·386 | 3·187 | 17319 | 9484 | 21619 |
| Hunan Province | Xiangyin xian | 2·058 | 1·406 | 5·219 | 1·121 | 2·584 | 13992 | 7611 | 17528 |
| Hunan Province | Xinhua xian | 2·281 | 1·613 | 5·975 | 1·204 | 2·878 | 26445 | 13949 | 33389 |
| Hunan Province | Xinling xian | 4·106 | 2·985 | 11·100 | 2·087 | 5·218 | 20284 | 10313 | 25761 |
| Hunan Province | Xintian xian | 5·314 | 3·735 | 13·886 | 2·748 | 6·773 | 16241 | 8451 | 20820 |
| Hunan Province | Xinzhao xian | 2·818 | 1·927 | 7·175 | 1·520 | 3·551 | 16238 | 8717 | 20503 |
| Hunan Province | Xupu xian | 2·193 | 1·584 | 5·817 | 1·136 | 2·766 | 16584 | 8592 | 20812 |
| Hunan Province | Yiyangshi | 1·981 | 1·346 | 5·034 | 1·080 | 2·480 | 26344 | 14110 | 32786 |
| Hunan Province | Yiyangshi | 1·920 | 1·305 | 4·826 | 1·052 | 2·400 | 378 | 208 | 474 |
| Hunan Province | Yongshun xian | 1·588 | 1·205 | 4·405 | 0·796 | 2·005 | 6644 | 3310 | 8384 |
| Hunan Province | Yongxing xian | 4·641 | 3·163 | 11·825 | 2·467 | 5·908 | 24529 | 13108 | 31209 |
| Hunan Province | Yongzhoushi | 5·057 | 3·429 | 12·781 | 2·693 | 6·416 | 32416 | 17301 | 40880 |
| Hunan Province | You xian | 3·450 | 2·517 | 9·304 | 1·765 | 4·370 | 22552 | 11611 | 28564 |
| Hunan Province | Yuanjiangshi | 1·971 | 1·365 | 5·021 | 1·056 | 2·490 | 8898 | 4802 | 11230 |
| Hunan Province | Yuanling xian | 1·745 | 1·274 | 4·692 | 0·899 | 2·197 | 8799 | 4486 | 11135 |
| Hunan Province | Yueyang xian | 2·106 | 1·501 | 5·557 | 1·105 | 2·651 | 33745 | 18005 | 42663 |
| Hunan Province | Yueyangshi | 2·073 | 1·515 | 5·592 | 1·079 | 2·597 | 3503 | 1836 | 4389 |
| Hunan Province | Zhaodong xian | 3·226 | 2·192 | 8·135 | 1·737 | 4·076 | 33893 | 18244 | 42741 |
| Hunan Province | Zhaoyang xian | 3·396 | 2·327 | 8·696 | 1·811 | 4·285 | 32294 | 17233 | 40745 |
| Hunan Province | Zhijiangdongzuzizhixain | 2·444 | 1·943 | 7·216 | 1·155 | 3·109 | 7124 | 3381 | 8973 |
| Hunan Province | Zhuzhou xian | 2·934 | 1·971 | 7·378 | 1·590 | 3·700 | 11102 | 6063 | 13975 |
| Hunan Province | Zhuzhoushi | 2·626 | 1·765 | 6·494 | 1·437 | 3·309 | 37232 | 20181 | 46669 |
| Hunan Province | Zixingshi | 5·053 | 3·466 | 12·891 | 2·688 | 6·403 | 15948 | 8524 | 20215 |

| | | β-thalassaemia carrier prevalence | | | | | Estimated carrier counts | | |
|---|---|---|---|---|---|---|---|---|---|
| ADM1 | ADM2 | Mean | SD | CI95% | Q25 | Q75 | Mean | Q25 | Q75 |
| Inner Mongolia Autonomous Region | Abagaqi | 4·824 | 7·873 | 27·750 | 0·779 | 5·251 | 1724 | 260 | 1853 |
| Inner Mongolia Autonomous Region | Alashanyouqi | 4·712 | 7·686 | 26·607 | 0·807 | 5·154 | 1175 | 178 | 1278 |
| Inner Mongolia Autonomous Region | Alashanzuoqi | 4·340 | 6·907 | 23·795 | 0·783 | 4·836 | 7952 | 1469 | 8723 |
| Inner Mongolia Autonomous Region | Alukeerqinqi | 4·346 | 6·693 | 23·967 | 0·774 | 4·911 | 10606 | 1833 | 11985 |
| Inner Mongolia Autonomous Region | Aohanqi | 3·959 | 6·289 | 21·396 | 0·671 | 4·517 | 17554 | 2982 | 19983 |
| Inner Mongolia Autonomous Region | Arongqi | 4·811 | 7·361 | 26·008 | 0·874 | 5·419 | 12405 | 2224 | 13840 |
| Inner Mongolia Autonomous Region | Balinyouqi | 4·324 | 6·715 | 23·409 | 0·752 | 4·902 | 7142 | 1258 | 8146 |
| Inner Mongolia Autonomous Region | Balinzuoqi | 4·473 | 6·954 | 24·721 | 0·769 | 5·028 | 12141 | 2093 | 13845 |
| Inner Mongolia Autonomous Region | Baotoushi | 4·501 | 7·109 | 24·458 | 0·771 | 5·082 | 101558 | 17450 | 113924 |
| Inner Mongolia Autonomous Region | Chahaeryouyihouqi | 4·413 | 7·311 | 25·143 | 0·725 | 4·776 | 4536 | 742 | 4733 |
| Inner Mongolia Autonomous Region | Chahaeryouyiqianqi | 4·225 | 7·078 | 23·884 | 0·720 | 4·533 | 17476 | 3033 | 19237 |
| Inner Mongolia Autonomous Region | Chahaeryouyizhongqi | 4·441 | 7·257 | 25·500 | 0·736 | 4·753 | 3688 | 610 | 3881 |
| Inner Mongolia Autonomous Region | Chenbaerhuqi | 5·263 | 8·178 | 28·106 | 0·897 | 5·965 | 4969 | 833 | 5720 |
| Inner Mongolia Autonomous Region | Cifengshi | 4·080 | 6·433 | 21·945 | 0·694 | 4·654 | 53289 | 9105 | 60426 |
| Inner Mongolia Autonomous Region | Daerhanmaominganlianheqi | 4·608 | 7·051 | 25·135 | 0·760 | 5·286 | 4213 | 673 | 4912 |
| Inner Mongolia Autonomous Region | Dalateqi | 4·488 | 7·287 | 24·705 | 0·775 | 4·981 | 14852 | 2519 | 16486 |
| Inner Mongolia Autonomous Region | Dengkou xian | 4·476 | 6·996 | 23·638 | 0·804 | 5·170 | 3943 | 711 | 4429 |
| Inner Mongolia Autonomous Region | Dongshengshi | 4·456 | 7·261 | 24·320 | 0·781 | 4·900 | 25989 | 4580 | 28928 |

| | | β-thalassaemia carrier prevalence | | | | | Estimated carrier counts | | |
|---|---|---|---|---|---|---|---|---|---|
| ADM1 | ADM2 | Mean | SD | CI95% | Q25 | Q75 | Mean | Q25 | Q75 |
| Inner Mongolia Autonomous Region | Dongwuzhumuqinqi | 4·883 | 7·637 | 27·099 | 0·845 | 5·449 | 4590 | 813 | 5197 |
| Inner Mongolia Autonomous Region | Duolun xian | 4·193 | 6·861 | 23·101 | 0·714 | 4·626 | 4330 | 717 | 4812 |
| Inner Mongolia Autonomous Region | Ejinaqi | 4·951 | 7·758 | 27·288 | 0·862 | 5·542 | 1786 | 286 | 1991 |
| Inner Mongolia Autonomous Region | Elunchunzizhiqi | 5·220 | 8·069 | 29·360 | 0·890 | 5·787 | 17548 | 2940 | 19332 |
| Inner Mongolia Autonomous Region | Erlianhaoteshi | 4·760 | 7·217 | 25·879 | 0·786 | 5·636 | 3542 | 582 | 4340 |
| Inner Mongolia Autonomous Region | Ertuokeqi | 4·282 | 6·723 | 23·501 | 0·766 | 4·830 | 10367 | 1874 | 11895 |
| Inner Mongolia Autonomous Region | Ertuokeqianqi | 4·142 | 6·351 | 22·208 | 0·779 | 4·711 | 3743 | 683 | 4300 |
| Inner Mongolia Autonomous Region | Ewenkezuzizhiqi | 5·231 | 7·931 | 28·305 | 0·908 | 5·946 | 2393 | 397 | 2688 |
| Inner Mongolia Autonomous Region | Fengzhenshi | 4·131 | 6·932 | 24·032 | 0·704 | 4·394 | 7084 | 1214 | 7739 |
| Inner Mongolia Autonomous Region | Geergunyouqi | 5·304 | 8·211 | 28·965 | 0·895 | 6·020 | 3584 | 614 | 4081 |
| Inner Mongolia Autonomous Region | Geergunzuoqi | 5·303 | 8·105 | 28·208 | 0·921 | 6·087 | 3762 | 643 | 4406 |
| Inner Mongolia Autonomous Region | Guyang xian | 4·561 | 7·076 | 25·002 | 0·756 | 5·207 | 5420 | 869 | 6224 |
| Inner Mongolia Autonomous Region | Hailaershi | 5·209 | 7·994 | 27·851 | 0·916 | 5·885 | 20364 | 3519 | 23112 |
| Inner Mongolia Autonomous Region | Hangjinhouqi | 4·465 | 6·847 | 23·162 | 0·801 | 5·210 | 10522 | 1874 | 12373 |
| Inner Mongolia Autonomous Region | Hangjinqi | 4·481 | 7·152 | 24·155 | 0·766 | 5·062 | 4796 | 781 | 5307 |
| Inner Mongolia Autonomous Region | Helingeer xian | 4·310 | 7·037 | 25·145 | 0·740 | 4·658 | 7388 | 1243 | 7957 |
| Inner Mongolia Autonomous Region | Huade xian | 4·438 | 7·603 | 25·412 | 0·712 | 4·766 | 4241 | 687 | 4581 |
| Inner Mongolia Autonomous Region | Huhehaoteshi | 4·385 | 7·008 | 25·387 | 0·745 | 4·777 | 116250 | 19539 | 124690 |

| | | β-thalassaemia carrier prevalence | | | | | Estimated carrier counts | | |
|---|---|---|---|---|---|---|---|---|---|
| ADM1 | ADM2 | Mean | SD | CI95% | Q25 | Q75 | Mean | Q25 | Q75 |
| Inner Mongolia Autonomous Region | Huinong xian | 4·177 | 6·475 | 22·579 | 0·765 | 4·795 | 1999 | 364 | 2322 |
| Inner Mongolia Autonomous Region | Huinong xian | 4·222 | 6·477 | 22·460 | 0·771 | 4·872 | 4204 | 770 | 4861 |
| Inner Mongolia Autonomous Region | Huolinguoleshi | 4·734 | 7·128 | 26·606 | 0·843 | 5·374 | 2324 | 396 | 2659 |
| Inner Mongolia Autonomous Region | Jiningshi | 4·234 | 7·077 | 22·242 | 0·745 | 4·632 | 5594 | 1030 | 6081 |
| Inner Mongolia Autonomous Region | Kailu xian | 4·230 | 6·607 | 22·544 | 0·729 | 4·820 | 13622 | 2341 | 15600 |
| Inner Mongolia Autonomous Region | Keerqinyouyiqianqi | 4·839 | 7·299 | 26·299 | 0·857 | 5·508 | 26180 | 4548 | 30301 |
| Inner Mongolia Autonomous Region | Keerqinyouyizhongqi | 4·507 | 6·887 | 24·504 | 0·814 | 5·097 | 9134 | 1594 | 10214 |
| Inner Mongolia Autonomous Region | Keerqinzuoyihouqi | 4·170 | 6·597 | 22·718 | 0·702 | 4·731 | 12868 | 2172 | 14709 |
| Inner Mongolia Autonomous Region | Keerqinzuoyizhongqi | 4·339 | 6·950 | 23·560 | 0·742 | 4·864 | 17164 | 2907 | 19094 |
| Inner Mongolia Autonomous Region | Kelinqinqi | 3·968 | 6·236 | 21·172 | 0·690 | 4·536 | 14921 | 2601 | 17005 |
| Inner Mongolia Autonomous Region | Keshenketengqi | 4·287 | 6·757 | 23·386 | 0·739 | 4·814 | 7609 | 1271 | 8542 |
| Inner Mongolia Autonomous Region | Kulunqi | 4·051 | 6·298 | 22·074 | 0·674 | 4·649 | 5860 | 970 | 6677 |
| Inner Mongolia Autonomous Region | Liangcheng xian | 4·256 | 7·024 | 24·598 | 0·722 | 4·563 | 4710 | 805 | 5130 |
| Inner Mongolia Autonomous Region | Linheshi | 4·493 | 6·855 | 23·027 | 0·785 | 5·295 | 25659 | 4468 | 30209 |
| Inner Mongolia Autonomous Region | Linxi xian | 4·368 | 6·782 | 23·824 | 0·753 | 4·943 | 8020 | 1379 | 9012 |
| Inner Mongolia Autonomous Region | Manzhoulishi | 5·278 | 8·122 | 28·360 | 0·901 | 6·041 | 11143 | 1874 | 12463 |
| Inner Mongolia Autonomous Region | Molidawadahanerzuzizhiqi | 4·961 | 7·614 | 27·796 | 0·875 | 5·499 | 10028 | 1775 | 11215 |
| Inner Mongolia Autonomous Region | Naimanqi | 4·081 | 6·366 | 22·034 | 0·681 | 4·699 | 14880 | 2469 | 17190 |

| | | β-thalassaemia carrier prevalence | | | | | Estimated carrier counts | | |
|---|---|---|---|---|---|---|---|---|---|
| ADM1 | ADM2 | Mean | SD | CI95% | Q25 | Q75 | Mean | Q25 | Q75 |
| Inner Mongolia Autonomous Region | Ningcheng xian | 3·877 | 6·090 | 21·030 | 0·678 | 4·375 | 15222 | 2651 | 17165 |
| Inner Mongolia Autonomous Region | Qingshuihe xian | 4·237 | 6·888 | 24·444 | 0·748 | 4·613 | 3114 | 526 | 3380 |
| Inner Mongolia Autonomous Region | Shangdu xian | 4·460 | 7·549 | 25·689 | 0·708 | 4·788 | 7591 | 1173 | 8110 |
| Inner Mongolia Autonomous Region | Shangyi xian | 4·259 | 7·241 | 24·819 | 0·694 | 4·530 | 4120 | 678 | 4484 |
| Inner Mongolia Autonomous Region | Shizhuishanshi | 4·182 | 6·487 | 23·143 | 0·772 | 4·780 | 5111 | 922 | 5822 |
| Inner Mongolia Autonomous Region | Shuniteyouqi | 4·586 | 7·403 | 25·384 | 0·757 | 5·098 | 2917 | 472 | 3157 |
| Inner Mongolia Autonomous Region | Shunitezuoqi | 4·722 | 7·439 | 25·704 | 0·789 | 5·368 | 1553 | 248 | 1738 |
| Inner Mongolia Autonomous Region | Siziwangqi | 4·570 | 7·173 | 25·760 | 0·755 | 5·068 | 5702 | 893 | 6295 |
| Inner Mongolia Autonomous Region | Taipusiqi | 4·296 | 7·363 | 24·561 | 0·706 | 4·548 | 4705 | 767 | 5044 |
| Inner Mongolia Autonomous Region | Tongliaoshi | 4·270 | 6·831 | 23·369 | 0·718 | 4·794 | 39122 | 6497 | 44148 |
| Inner Mongolia Autonomous Region | Tumuteyouqi | 4·414 | 7·076 | 24·884 | 0·762 | 4·860 | 10868 | 1889 | 11999 |
| Inner Mongolia Autonomous Region | Tumutezuoqi | 4·417 | 7·055 | 25·212 | 0·754 | 4·864 | 14568 | 2486 | 16072 |
| Inner Mongolia Autonomous Region | Tuoketuoqi | 4·341 | 7·017 | 24·600 | 0·741 | 4·746 | 7225 | 1257 | 7955 |
| Inner Mongolia Autonomous Region | Tuquan xian | 4·556 | 6·869 | 24·579 | 0·813 | 5·237 | 9810 | 1744 | 11108 |
| Inner Mongolia Autonomous Region | Wengniuteqi | 4·151 | 6·514 | 22·289 | 0·711 | 4·755 | 13975 | 2377 | 15975 |
| Inner Mongolia Autonomous Region | Wuchuang xian | 4·483 | 7·032 | 25·453 | 0·756 | 4·986 | 4407 | 742 | 4855 |
| Inner Mongolia Autonomous Region | Wuhaishi | 4·224 | 6·462 | 22·158 | 0·777 | 4·855 | 23064 | 4236 | 26498 |
| Inner Mongolia Autonomous Region | Wulanhaoteshi | 4·652 | 6·970 | 26·073 | 0·805 | 5·312 | 5938 | 1042 | 7010 |

| ADM1 | ADM2 | β-thalassaemia carrier prevalence Mean | SD | CI95% | Q25 | Q75 | Estimated carrier counts Mean | Q25 | Q75 |
|---|---|---|---|---|---|---|---|---|---|
| Inner Mongolia Autonomous Region | Wulatehouqi | 4·503 | 6·935 | 23·797 | 0·808 | 5·218 | 1836 | 315 | 2185 |
| Inner Mongolia Autonomous Region | Wulateqianqi | 4·617 | 7·182 | 24·776 | 0·758 | 5·331 | 11305 | 1845 | 13151 |
| Inner Mongolia Autonomous Region | Wulatezhongqi | 4·669 | 7·013 | 24·752 | 0·774 | 5·477 | 5778 | 916 | 6822 |
| Inner Mongolia Autonomous Region | Wushenqi | 4·330 | 6·902 | 23·882 | 0·781 | 4·797 | 6648 | 1175 | 7477 |
| Inner Mongolia Autonomous Region | Wuyuan xian | 4·596 | 7·062 | 24·311 | 0·764 | 5·374 | 10263 | 1721 | 12016 |
| Inner Mongolia Autonomous Region | Xianghuangqi | 4·451 | 7·582 | 24·953 | 0·725 | 4·823 | 1211 | 175 | 1285 |
| Inner Mongolia Autonomous Region | Xilinhaoteshi | 4·687 | 7·641 | 26·721 | 0·782 | 5·060 | 16559 | 2684 | 17524 |
| Inner Mongolia Autonomous Region | Xinbaerhuyouqi | 5·288 | 8·009 | 28·982 | 0·920 | 6·080 | 1992 | 344 | 2389 |
| Inner Mongolia Autonomous Region | Xinbaerhuzuoqi | 5·301 | 8·009 | 29·211 | 0·920 | 6·058 | 1955 | 336 | 2147 |
| Inner Mongolia Autonomous Region | Xinghe xian | 4·208 | 7·124 | 24·136 | 0·702 | 4·485 | 7098 | 1221 | 7618 |
| Inner Mongolia Autonomous Region | Xiwuzhumuqinqi | 4·647 | 7·351 | 25·758 | 0·787 | 5·193 | 4601 | 742 | 5260 |
| Inner Mongolia Autonomous Region | Yakesheishi | 5·144 | 7·804 | 27·538 | 0·912 | 5·829 | 15131 | 2629 | 17129 |
| Inner Mongolia Autonomous Region | Yijinhuoluoqi | 4·440 | 7·237 | 24·608 | 0·773 | 4·893 | 15695 | 2774 | 17460 |
| Inner Mongolia Autonomous Region | Zalantunshi | 4·806 | 7·261 | 25·637 | 0·872 | 5·472 | 14964 | 2674 | 16984 |
| Inner Mongolia Autonomous Region | Zaluteqi | 4·451 | 6·819 | 24·492 | 0·802 | 5·031 | 15473 | 2792 | 17558 |
| Inner Mongolia Autonomous Region | Zhafente | 4·701 | 7·177 | 25·315 | 0·839 | 5·340 | 14534 | 2618 | 16582 |
| Inner Mongolia Autonomous Region | Zhenglanqi | 4·345 | 7·230 | 24·418 | 0·728 | 4·709 | 2947 | 481 | 3211 |
| Inner Mongolia Autonomous Region | Zhengxiangbaiqi | 4·418 | 7·496 | 24·694 | 0·728 | 4·799 | 1918 | 305 | 2078 |

| ADM1 | ADM2 | β-thalassaemia carrier prevalence Mean | SD | CI95% | Q25 | Q75 | Estimated carrier counts Mean | Q25 | Q75 |
|---|---|---|---|---|---|---|---|---|---|
| Inner Mongolia Autonomous Region | Zungeerqi | 4·323 | 7·004 | 23·883 | 0·766 | 4·785 | 14888 | 2619 | 16612 |
| Inner Mongolia Autonomous Region | Zuozi xian | 4·349 | 7·063 | 24·974 | 0·734 | 4·695 | 3698 | 630 | 4011 |
| Jiangsu Province | Baoying xian | 0·612 | 0·889 | 2·837 | 0·163 | 0·695 | 4137 | 1102 | 4683 |
| Jiangsu Province | Binhai xian | 0·806 | 1·284 | 4·030 | 0·184 | 0·902 | 6551 | 1499 | 7333 |
| Jiangsu Province | Changshushi | 0·339 | 0·396 | 1·294 | 0·120 | 0·410 | 5215 | 1847 | 6349 |
| Jiangsu Province | Changzhoushi | 0·341 | 0·377 | 1·266 | 0·126 | 0·421 | 3551 | 1321 | 4378 |
| Jiangsu Province | Chongming xian | 0·416 | 0·525 | 1·748 | 0·131 | 0·491 | 1623 | 509 | 1914 |
| Jiangsu Province | Dafeng xian | 0·528 | 0·756 | 2·429 | 0·145 | 0·609 | 3497 | 957 | 4078 |
| Jiangsu Province | Dantu xian | 0·416 | 0·511 | 1·676 | 0·136 | 0·500 | 3186 | 1044 | 3796 |
| Jiangsu Province | Danyangshi | 0·385 | 0·444 | 1·489 | 0·135 | 0·467 | 3367 | 1196 | 4154 |
| Jiangsu Province | Donghai xian | 1·105 | 1·781 | 5·486 | 0·245 | 1·248 | 12172 | 2691 | 13864 |
| Jiangsu Province | Dongtaishi | 0·443 | 0·580 | 1·908 | 0·134 | 0·522 | 4361 | 1331 | 5148 |
| Jiangsu Province | Funing xian | 0·714 | 1·089 | 3·437 | 0·173 | 0·808 | 5657 | 1369 | 6351 |
| Jiangsu Province | Gaoyoushi | 0·515 | 0·692 | 2·245 | 0·152 | 0·602 | 3496 | 1046 | 4172 |
| Jiangsu Province | Guannan xian | 0·885 | 1·410 | 4·311 | 0·201 | 1·003 | 5654 | 1295 | 6452 |
| Jiangsu Province | Guanyun xian | 0·972 | 1·532 | 4·790 | 0·216 | 1·096 | 7509 | 1705 | 8461 |
| Jiangsu Province | Haian xian | 0·400 | 0·505 | 1·673 | 0·126 | 0·480 | 3230 | 1018 | 3881 |
| Jiangsu Province | Haimenshi | 0·387 | 0·466 | 1·571 | 0·126 | 0·467 | 3025 | 1007 | 3658 |
| Jiangsu Province | Hongze xian | 0·704 | 0·977 | 3·297 | 0·190 | 0·813 | 2622 | 706 | 3015 |
| Jiangsu Province | Huaianshi | 0·696 | 1·038 | 3·284 | 0·178 | 0·797 | 6241 | 1600 | 7244 |
| Jiangsu Province | Huaiyin xian | 0·793 | 1·201 | 3·657 | 0·201 | 0·906 | 7509 | 1941 | 8724 |
| Jiangsu Province | Huaiyinshi | 0·726 | 1·092 | 3·310 | 0·188 | 0·831 | 4759 | 1247 | 5489 |
| Jiangsu Province | Jiangdushi | 0·435 | 0·547 | 1·831 | 0·138 | 0·517 | 4488 | 1436 | 5346 |
| Jiangsu Province | Jiangning xian | 0·527 | 0·630 | 2·149 | 0·175 | 0·634 | 10843 | 3627 | 13048 |
| Jiangsu Province | Jiangyanshi | 0·405 | 0·519 | 1·673 | 0·129 | 0·481 | 4460 | 1423 | 5266 |
| Jiangsu Province | Jiangyinshi | 0·324 | 0·372 | 1·216 | 0·120 | 0·389 | 5166 | 1905 | 6214 |
| Jiangsu Province | Jianhu xian | 0·640 | 0·942 | 3·025 | 0·164 | 0·726 | 3634 | 948 | 4135 |
| Jiangsu Province | Jingjiangshi | 0·337 | 0·396 | 1·308 | 0·119 | 0·406 | 2221 | 794 | 2699 |
| Jiangsu Province | Jintanshi | 0·395 | 0·440 | 1·536 | 0·142 | 0·481 | 2172 | 793 | 2671 |
| Jiangsu Province | Jurong xian | 0·447 | 0·516 | 1·828 | 0·153 | 0·534 | 2624 | 897 | 3153 |
| Jiangsu Province | Kunshanshi | 0·372 | 0·450 | 1·481 | 0·128 | 0·442 | 7233 | 2486 | 8615 |
| Jiangsu Province | Lianshui xian | 0·800 | 1·260 | 3·846 | 0·193 | 0·904 | 6417 | 1563 | 7290 |
| Jiangsu Province | Lianyungangshi | 1·070 | 1·686 | 5·601 | 0·230 | 1·202 | 9274 | 2021 | 10603 |
| Jiangsu Province | Lishui xian | 0·479 | 0·547 | 1·897 | 0·166 | 0·581 | 2411 | 834 | 2912 |

| | | β-thalassaemia carrier prevalence | | | | | Estimated carrier counts | | |
|---|---|---|---|---|---|---|---|---|---|
| **ADM1** | **ADM2** | **Mean** | **SD** | **CI95%** | **Q25** | **Q75** | **Mean** | **Q25** | **Q75** |
| Jiangsu Province | Liuhe xian | 0·546 | 0·680 | 2·375 | 0·168 | 0·649 | 4086 | 1272 | 4854 |
| Jiangsu Province | Liyangshi | 0·423 | 0·472 | 1·653 | 0·149 | 0·517 | 3335 | 1156 | 4034 |
| Jiangsu Province | Nanjingshi | 0·530 | 0·633 | 2·201 | 0·172 | 0·639 | 27897 | 9059 | 33434 |
| Jiangsu Province | Nantongshi | 0·344 | 0·360 | 1·398 | 0·121 | 0·431 | 628 | 221 | 788 |
| Jiangsu Province | Pei xian | 1·638 | 2·454 | 8·195 | 0·361 | 1·875 | 14337 | 3126 | 16301 |
| Jiangsu Province | Pizhoushi | 1·254 | 1·917 | 6·261 | 0·282 | 1·418 | 16028 | 3643 | 18081 |
| Jiangsu Province | Qidongshi | 0·427 | 0·533 | 1·831 | 0·130 | 0·510 | 3242 | 998 | 3855 |
| Jiangsu Province | Rudong xian | 0·396 | 0·468 | 1·606 | 0·127 | 0·482 | 3277 | 1049 | 4014 |
| Jiangsu Province | Rugaoshi | 0·363 | 0·438 | 1·455 | 0·123 | 0·437 | 4180 | 1411 | 5017 |
| Jiangsu Province | Sheyang xian | 0·694 | 1·077 | 3·432 | 0·168 | 0·773 | 5852 | 1409 | 6487 |
| Jiangsu Province | Shuqianshi | 1·004 | 1·515 | 4·862 | 0·244 | 1·144 | 15331 | 3768 | 17514 |
| Jiangsu Province | Shuyang xian | 0·961 | 1·554 | 4·567 | 0·224 | 1·086 | 16427 | 3855 | 18357 |
| Jiangsu Province | Shuzhoushi | 0·355 | 0·403 | 1·362 | 0·129 | 0·416 | 4193 | 1518 | 4916 |
| Jiangsu Province | Sihong xian | 0·865 | 1·212 | 4·103 | 0·222 | 1·000 | 8892 | 2263 | 10267 |
| Jiangsu Province | Siyang xian | 0·857 | 1·271 | 4·016 | 0·215 | 0·994 | 7673 | 1923 | 8896 |
| Jiangsu Province | Suqian xian | 0·604 | 0·843 | 2·740 | 0·168 | 0·704 | 1746 | 500 | 2082 |
| Jiangsu Province | Taichang xian | 0·370 | 0·443 | 1·468 | 0·128 | 0·443 | 2899 | 999 | 3454 |
| Jiangsu Province | Taixing xian | 0·365 | 0·433 | 1·466 | 0·125 | 0·437 | 4212 | 1458 | 5081 |
| Jiangsu Province | Taizhoushi | 0·410 | 0·507 | 1·651 | 0·133 | 0·490 | 1717 | 561 | 2040 |
| Jiangsu Province | Tianchangshi | 0·576 | 0·750 | 2·545 | 0·170 | 0·680 | 3312 | 991 | 3858 |
| Jiangsu Province | Tongshan xian | 1·389 | 2·079 | 6·896 | 0·318 | 1·591 | 35541 | 8123 | 41118 |
| Jiangsu Province | Tongzhoushi | 0·365 | 0·428 | 1·441 | 0·124 | 0·445 | 8921 | 3077 | 10928 |
| Jiangsu Province | Wujin xian | 0·348 | 0·391 | 1·295 | 0·129 | 0·423 | 10339 | 3868 | 12517 |
| Jiangsu Province | Wuxi xian | 0·331 | 0·371 | 1·225 | 0·124 | 0·402 | 13997 | 5261 | 16999 |
| Jiangsu Province | Wuxianshi | 0·347 | 0·398 | 1·322 | 0·126 | 0·420 | 12421 | 4529 | 15013 |
| Jiangsu Province | Wuxishi | 0·327 | 0·338 | 1·255 | 0·123 | 0·410 | 150 | 54 | 189 |
| Jiangsu Province | Xiangshui xian | 0·870 | 1·375 | 4·392 | 0·192 | 0·978 | 4301 | 932 | 4836 |
| Jiangsu Province | Xinghuashi | 0·489 | 0·659 | 2·166 | 0·141 | 0·573 | 4675 | 1345 | 5518 |
| Jiangsu Province | Xinyi xian | 1·123 | 1·743 | 5·574 | 0·255 | 1·259 | 12969 | 2910 | 14446 |
| Jiangsu Province | Xuyu xian | 0·677 | 0·889 | 3·062 | 0·192 | 0·794 | 3094 | 863 | 3628 |
| Jiangsu Province | Xuzhoushi | 1·375 | 1·975 | 6·628 | 0·317 | 1·619 | 16584 | 3827 | 19422 |
| Jiangsu Province | Yanchengshi | 0·572 | 0·821 | 2·682 | 0·154 | 0·654 | 9674 | 2598 | 11059 |
| Jiangsu Province | Yangzhong | 0·373 | 0·432 | 1·514 | 0·128 | 0·454 | 1184 | 405 | 1442 |
| Jiangsu Province | Yangzhoushi | 0·445 | 0·550 | 1·870 | 0·140 | 0·528 | 7218 | 2265 | 8439 |
| Jiangsu Province | Yixingshi | 0·380 | 0·416 | 1·451 | 0·138 | 0·467 | 4776 | 1749 | 5890 |
| Jiangsu Province | Yizhengshi | 0·483 | 0·614 | 2·028 | 0·152 | 0·573 | 2708 | 862 | 3207 |

| ADM1 | ADM2 | β-thalassaemia carrier prevalence Mean | SD | CI95% | Q25 | Q75 | Estimated carrier counts Mean | Q25 | Q75 |
|---|---|---|---|---|---|---|---|---|---|
| Jiangsu Province | Zhangjiagangshi | 0·333 | 0·383 | 1·274 | 0·118 | 0·405 | 4502 | 1604 | 5497 |
| Jiangsu Province | Zhenjiangshi | 0·427 | 0·554 | 1·815 | 0·135 | 0·498 | 1781 | 565 | 2069 |
| Jiangxi Province | Anfu xian | 3·352 | 2·624 | 9·715 | 1·614 | 4·259 | 11578 | 5520 | 14636 |
| Jiangxi Province | Anyi xian | 2·096 | 1·653 | 5·989 | 1·023 | 2·629 | 5197 | 2520 | 6508 |
| Jiangxi Province | Anyuan xian | 7·231 | 4·831 | 18·185 | 3·882 | 9·201 | 26175 | 14110 | 33308 |
| Jiangxi Province | Boyang xian | 1·637 | 1·608 | 5·752 | 0·651 | 2·038 | 20959 | 8406 | 26054 |
| Jiangxi Province | Chongren xian | 2·621 | 2·197 | 7·996 | 1·185 | 3·330 | 7904 | 3561 | 10153 |
| Jiangxi Province | Chongyi xian | 5·606 | 3·887 | 14·624 | 2·947 | 7·116 | 10161 | 5270 | 13039 |
| Jiangxi Province | Dayu xian | 6·051 | 4·088 | 15·425 | 3·222 | 7·672 | 16275 | 8738 | 20622 |
| Jiangxi Province | Dean xian | 1·963 | 1·695 | 6·205 | 0·879 | 2·466 | 5107 | 2277 | 6397 |
| Jiangxi Province | Dexingshi | 1·516 | 1·631 | 5·644 | 0·553 | 1·876 | 4605 | 1647 | 5614 |
| Jiangxi Province | Dingnan xian | 8·200 | 5·249 | 19·860 | 4·489 | 10·493 | 17091 | 9519 | 21742 |
| Jiangxi Province | Dongxiang xian | 2·095 | 1·818 | 6·614 | 0·930 | 2·644 | 9467 | 4236 | 12005 |
| Jiangxi Province | Duchang xian | 1·718 | 1·604 | 5·750 | 0·722 | 2·143 | 9470 | 3979 | 11815 |
| Jiangxi Province | Fengchengshi | 2·421 | 1·944 | 7·153 | 1·145 | 3·064 | 26091 | 12402 | 32840 |
| Jiangxi Province | Fengxin xian | 2·308 | 1·808 | 6·618 | 1·119 | 2·916 | 6649 | 3183 | 8402 |
| Jiangxi Province | Fenyi xian | 2·944 | 2·367 | 8·650 | 1·382 | 3·754 | 8341 | 3871 | 10577 |
| Jiangxi Province | Gan xian | 5·243 | 3·633 | 13·569 | 2·768 | 6·665 | 32852 | 17219 | 42100 |
| Jiangxi Province | Ganzhoushi | 5·274 | 3·705 | 13·790 | 2·750 | 6·676 | 51072 | 26788 | 64741 |
| Jiangxi Province | Gaoanshi | 2·465 | 1·930 | 7·082 | 1·185 | 3·135 | 17947 | 8629 | 22840 |
| Jiangxi Province | Guangchang xian | 3·308 | 2·698 | 9·905 | 1·525 | 4·205 | 6388 | 2887 | 7995 |
| Jiangxi Province | Guangfeng xian | 1·626 | 1·723 | 6·144 | 0·581 | 2·016 | 12258 | 4363 | 15144 |
| Jiangxi Province | Guixi xian | 1·992 | 1·907 | 6·826 | 0·810 | 2·493 | 10256 | 4095 | 12862 |
| Jiangxi Province | Hengfeng xian | 1·746 | 1·805 | 6·276 | 0·645 | 2·182 | 3485 | 1292 | 4365 |
| Jiangxi Province | Huichang xian | 6·192 | 4·214 | 15·772 | 3·288 | 7·887 | 27807 | 14668 | 35445 |
| Jiangxi Province | Hukou xian | 1·632 | 1·538 | 5·542 | 0·674 | 2·034 | 3652 | 1507 | 4488 |
| Jiangxi Province | Jianning xian | 2·973 | 2·503 | 9·172 | 1·331 | 3·772 | 3330 | 1509 | 4217 |
| Jiangxi Province | Jianshi | 3·590 | 2·833 | 10·417 | 1·720 | 4·539 | 27163 | 13073 | 34411 |
| Jiangxi Province | Jianshi | 3·552 | 2·786 | 10·127 | 1·651 | 4·559 | 11062 | 5154 | 14197 |
| Jiangxi Province | Jingan xian | 2·192 | 1·750 | 6·395 | 1·049 | 2·756 | 2728 | 1326 | 3397 |
| Jiangxi Province | Jingdezheshi | 1·375 | 1·464 | 5·082 | 0·511 | 1·697 | 12503 | 4792 | 15405 |
| Jiangxi Province | Jinggangshanshi | 4·219 | 3·084 | 11·608 | 2·137 | 5·366 | 3291 | 1664 | 4184 |
| Jiangxi Province | Jinxi xian | 2·218 | 1·944 | 6·967 | 0·974 | 2·808 | 5716 | 2539 | 7288 |
| Jiangxi Province | Jinxian xian | 2·053 | 1·723 | 6·325 | 0·946 | 2·577 | 12228 | 5725 | 15362 |
| Jiangxi Province | Jishui xian | 3·407 | 2·766 | 10·112 | 1·583 | 4·336 | 14773 | 6875 | 18811 |
| Jiangxi Province | Jiujian xian | 1·790 | 1·616 | 5·704 | 0·782 | 2·241 | 3490 | 1476 | 4398 |

| | | β-thalassaemia carrier prevalence | | | | | Estimated carrier counts | | |
|---|---|---|---|---|---|---|---|---|---|
| ADM1 | ADM2 | Mean | SD | CI95% | Q25 | Q75 | Mean | Q25 | Q75 |
| Jiangxi Province | Jiujiang xian | 1·848 | 1·700 | 6·069 | 0·795 | 2·281 | 6418 | 2790 | 7943 |
| Jiangxi Province | Jiujiangshi | 1·759 | 1·603 | 5·801 | 0·755 | 2·194 | 9989 | 4308 | 12419 |
| Jiangxi Province | Kaihua xian | 1·210 | 1·381 | 4·732 | 0·423 | 1·474 | 3098 | 1075 | 3768 |
| Jiangxi Province | Lean xian | 3·007 | 2·547 | 9·324 | 1·357 | 3·804 | 8890 | 3985 | 11213 |
| Jiangxi Province | Lepingshi | 1·619 | 1·650 | 5·844 | 0·628 | 2·001 | 12531 | 4904 | 15488 |
| Jiangxi Province | Lianhua xian | 3·508 | 2·711 | 9·925 | 1·701 | 4·453 | 7417 | 3595 | 9359 |
| Jiangxi Province | Lichuan xian | 2·622 | 2·343 | 8·544 | 1·113 | 3·338 | 5549 | 2346 | 6953 |
| Jiangxi Province | Linchanshi | 2·374 | 2·015 | 7·294 | 1·065 | 3·013 | 26024 | 11836 | 32999 |
| Jiangxi Province | Longnan xian | 7·766 | 5·162 | 19·457 | 4·169 | 9·924 | 25531 | 13353 | 32903 |
| Jiangxi Province | Nanchang xian | 2·029 | 1·625 | 5·936 | 0·976 | 2·546 | 45762 | 22439 | 56968 |
| Jiangxi Province | Nanchangshi | 2·052 | 1·634 | 5·918 | 0·992 | 2·597 | 39792 | 19134 | 50412 |
| Jiangxi Province | Nancheng xian | 2·547 | 2·238 | 8·192 | 1·092 | 3·234 | 7199 | 3073 | 9209 |
| Jiangxi Province | Nanfeng xian | 2·925 | 2·489 | 9·092 | 1·294 | 3·728 | 7788 | 3456 | 9933 |
| Jiangxi Province | Nankangshi | 5·439 | 3·729 | 14·016 | 2·897 | 6·907 | 56870 | 30434 | 72681 |
| Jiangxi Province | Nanxiong xian | 6·578 | 4·391 | 16·408 | 3·521 | 8·375 | 23369 | 12507 | 29738 |
| Jiangxi Province | Ningdu xian | 3·772 | 2·976 | 10·901 | 1·789 | 4·810 | 28147 | 13503 | 36123 |
| Jiangxi Province | Pengze xian | 1·526 | 1·545 | 5·467 | 0·590 | 1·887 | 4074 | 1577 | 4996 |
| Jiangxi Province | Pingxiangshi | 3·070 | 2·277 | 8·451 | 1·532 | 3·907 | 48145 | 24236 | 61269 |
| Jiangxi Province | Qianshan xian | 1·896 | 1·907 | 6·868 | 0·724 | 2·362 | 6567 | 2498 | 8214 |
| Jiangxi Province | Quannan xian | 7·292 | 4·824 | 18·107 | 3·937 | 9·316 | 12387 | 6672 | 15765 |
| Jiangxi Province | Ruichangshi | 1·935 | 1·695 | 6·102 | 0·868 | 2·417 | 6729 | 2906 | 8390 |
| Jiangxi Province | Ruijinshi | 4·879 | 3·590 | 13·262 | 2·442 | 6·240 | 29793 | 14786 | 38279 |
| Jiangxi Province | Shanggao xian | 2·538 | 1·985 | 7·302 | 1·220 | 3·217 | 7389 | 3516 | 9350 |
| Jiangxi Province | Shanggao xian | 2·688 | 2·148 | 7·880 | 1·276 | 3·408 | 7832 | 3722 | 10038 |
| Jiangxi Province | Shangrao xian | 1·683 | 1·759 | 6·244 | 0·617 | 2·094 | 11995 | 4361 | 14985 |
| Jiangxi Province | Shangraoshi | 1·654 | 1·698 | 5·932 | 0·598 | 2·052 | 10257 | 3717 | 12504 |
| Jiangxi Province | Shangyou xian | 5·184 | 3·644 | 13·696 | 2·690 | 6·593 | 13472 | 6996 | 17193 |
| Jiangxi Province | Shicheng xian | 3·738 | 2·860 | 10·657 | 1·812 | 4·783 | 10509 | 5034 | 13484 |
| Jiangxi Province | Suichuan xian | 4·585 | 3·372 | 12·409 | 2·328 | 5·814 | 23789 | 11925 | 30328 |
| Jiangxi Province | Taihe xian | 3·947 | 3·050 | 11·210 | 1·925 | 5·003 | 17619 | 8609 | 22397 |
| Jiangxi Province | Tonggu xian | 2·489 | 1·937 | 7·066 | 1·220 | 3·141 | 2888 | 1400 | 3627 |
| Jiangxi Province | Wanan xian | 4·379 | 3·282 | 12·267 | 2·195 | 5·541 | 11634 | 5745 | 14773 |
| Jiangxi Province | Wannian xian | 1·767 | 1·685 | 6·059 | 0·723 | 2·213 | 6406 | 2633 | 8034 |
| Jiangxi Province | Wanzai xian | 2·696 | 2·048 | 7·618 | 1·324 | 3·431 | 14226 | 7005 | 18159 |
| Jiangxi Province | Wuning xian | 2·145 | 1·721 | 6·251 | 1·024 | 2·709 | 6888 | 3254 | 8703 |
| Jiangxi Province | Wuyuan xian | 1·312 | 1·420 | 5·006 | 0·480 | 1·603 | 4134 | 1508 | 4994 |

| ADM1 | ADM2 | β-thalassaemia carrier prevalence Mean | SD | CI95% | Q25 | Q75 | Estimated carrier counts Mean | Q25 | Q75 |
|---|---|---|---|---|---|---|---|---|---|
| Jiangxi Province | Xiajiang xian | 3·022 | 2·483 | 9·135 | 1·411 | 3·807 | 5026 | 2346 | 6329 |
| Jiangxi Province | Xinfeng xian | 6·824 | 4·527 | 17·043 | 3·677 | 8·709 | 47667 | 25537 | 61011 |
| Jiangxi Province | Xingan xian | 2·842 | 2·381 | 8·731 | 1·297 | 3·606 | 7090 | 3234 | 8926 |
| Jiangxi Province | Xingguo xian | 4·247 | 3·214 | 11·942 | 2·083 | 5·414 | 31897 | 15668 | 40814 |
| Jiangxi Province | Xingzi xian | 1·867 | 1·678 | 6·055 | 0·809 | 2·318 | 2351 | 1023 | 2923 |
| Jiangxi Province | Xinjian xian | 1·913 | 1·584 | 5·693 | 0·904 | 2·384 | 2898 | 1402 | 3630 |
| Jiangxi Province | Xinjian xian | 2·197 | 1·709 | 6·245 | 1·074 | 2·781 | 24358 | 12164 | 30575 |
| Jiangxi Province | Xinyushi | 2·850 | 2·317 | 8·393 | 1·344 | 3·589 | 26430 | 12329 | 33058 |
| Jiangxi Province | Xiushui xian | 2·325 | 1·798 | 6·610 | 1·135 | 2·940 | 16973 | 8260 | 21457 |
| Jiangxi Province | Xunwu xian | 8·584 | 5·255 | 19·880 | 4·871 | 10·940 | 24266 | 13821 | 30993 |
| Jiangxi Province | Yichunshi | 2·963 | 2·305 | 8·506 | 1·430 | 3·765 | 33743 | 16212 | 42503 |
| Jiangxi Province | Yihuang xian | 2·803 | 2·359 | 8·636 | 1·261 | 3·561 | 5177 | 2250 | 6540 |
| Jiangxi Province | Yingtanshi | 1·989 | 1·901 | 6·821 | 0·812 | 2·455 | 5895 | 2400 | 7271 |
| Jiangxi Province | Yiyang xian | 1·831 | 1·856 | 6·520 | 0·702 | 2·279 | 6234 | 2356 | 7754 |
| Jiangxi Province | Yongfeng xian | 3·363 | 2·754 | 10·155 | 1·560 | 4·253 | 12852 | 5990 | 16350 |
| Jiangxi Province | Yongxin xian | 3·788 | 2·832 | 10·441 | 1·878 | 4·824 | 14269 | 7138 | 18182 |
| Jiangxi Province | Yongxiu xian | 1·979 | 1·656 | 5·987 | 0·931 | 2·473 | 7698 | 3635 | 9592 |
| Jiangxi Province | Yudu xian | 5·089 | 3·577 | 13·397 | 2·639 | 6·475 | 45065 | 23250 | 57075 |
| Jiangxi Province | Yugan xian | 1·885 | 1·698 | 6·209 | 0·817 | 2·359 | 14847 | 6395 | 18486 |
| Jiangxi Province | Yujiang xian | 1·972 | 1·814 | 6·610 | 0·822 | 2·466 | 6316 | 2625 | 7868 |
| Jiangxi Province | Yushan xian | 1·473 | 1·607 | 5·751 | 0·513 | 1·810 | 7694 | 2657 | 9482 |
| Jiangxi Province | Zhangshushi | 2·645 | 2·170 | 7·975 | 1·233 | 3·334 | 13785 | 6413 | 17335 |
| Jiangxi Province | Zixi xian | 2·276 | 2·059 | 7·457 | 0·968 | 2·872 | 2012 | 841 | 2510 |
| Jilin Province | Antu xian | 4·095 | 6·568 | 22·467 | 0·756 | 4·505 | 6921 | 1257 | 7550 |
| Jilin Province | Baichengshi | 4·545 | 6·804 | 24·739 | 0·815 | 5·216 | 14723 | 2678 | 16809 |
| Jilin Province | Changbaichaoxianzu xian | 3·737 | 6·069 | 20·813 | 0·703 | 4·034 | 4350 | 819 | 4557 |
| Jilin Province | Changchunshi | 4·242 | 6·859 | 22·691 | 0·741 | 4·784 | 206356 | 36414 | 232700 |
| Jilin Province | Changling xian | 4·379 | 6·872 | 23·702 | 0·754 | 5·011 | 19531 | 3370 | 22274 |
| Jilin Province | Daanshi | 4·557 | 6·914 | 24·247 | 0·792 | 5·260 | 11845 | 2028 | 13643 |
| Jilin Province | Dehuishi | 4·367 | 6·833 | 23·108 | 0·759 | 5·013 | 28407 | 4956 | 32779 |
| Jilin Province | Dongfeng xian | 3·994 | 6·450 | 21·746 | 0·691 | 4·468 | 10596 | 1821 | 11908 |
| Jilin Province | Dongliao xian | 4·086 | 6·527 | 22·056 | 0·703 | 4·631 | 12952 | 2218 | 14646 |
| Jilin Province | Dunhuashi | 4·314 | 6·886 | 23·361 | 0·762 | 4·845 | 16491 | 2968 | 18399 |
| Jilin Province | Fuyu xian | 4·468 | 6·887 | 23·978 | 0·762 | 5·125 | 34406 | 5914 | 39427 |
| Jilin Province | Gongzhulingshi | 4·298 | 6·919 | 23·056 | 0·735 | 4·866 | 38351 | 6573 | 42739 |
| Jilin Province | Helongshi | 4·084 | 6·678 | 22·354 | 0·768 | 4·434 | 5360 | 1009 | 5728 |
| Jilin Province | Huadianshi | 4·077 | 6·463 | 21·820 | 0·729 | 4·616 | 13823 | 2424 | 15504 |
| Jilin Province | Huichunshi | 4·241 | 6·951 | 23·095 | 0·815 | 4·653 | 12605 | 2498 | 13766 |
| Jilin Province | Huinan xian | 3·945 | 6·296 | 21·082 | 0·715 | 4·419 | 9462 | 1695 | 10505 |
| Jilin Province | Jianshi | 3·444 | 5·413 | 18·761 | 0·655 | 3·830 | 5684 | 1075 | 6381 |
| Jilin Province | Jiaohe xian | 4·270 | 6·763 | 22·564 | 0·745 | 4·870 | 14168 | 2460 | 16303 |
| Jilin Province | Jilinshi | 4·288 | 6·865 | 22·822 | 0·732 | 4·829 | 73089 | 12401 | 81884 |
| Jilin Province | Jingyu xian | 3·916 | 6·226 | 20·943 | 0·719 | 4·380 | 3933 | 717 | 4450 |
| Jilin Province | Jiutaishi | 4·337 | 6·835 | 23·074 | 0·750 | 4·956 | 28480 | 4959 | 32442 |

| | | β-thalassaemia carrier prevalence | | | | | Estimated carrier counts | | |
|---|---|---|---|---|---|---|---|---|---|
| ADM1 | ADM2 | Mean | SD | CI95% | Q25 | Q75 | Mean | Q25 | Q75 |
| Jilin Province | Liaoyuanshi | 4·042 | 6·428 | 22·416 | 0·710 | 4·624 | 15310 | 2699 | 17615 |
| Jilin Province | Linjiangshi | 3·709 | 5·830 | 20·220 | 0·696 | 4·110 | 21937 | 4117 | 24396 |
| Jilin Province | Lishu xian | 4·233 | 6·788 | 22·658 | 0·710 | 4·854 | 29373 | 4947 | 33703 |
| Jilin Province | Liuhe xian | 3·835 | 6·113 | 20·848 | 0·684 | 4·275 | 10266 | 1789 | 11356 |
| Jilin Province | Longjingshi | 4·187 | 6·748 | 22·358 | 0·793 | 4·655 | 25150 | 4802 | 28506 |
| Jilin Province | Meihekoushi | 3·920 | 6·291 | 21·161 | 0·689 | 4·403 | 20977 | 3686 | 23782 |
| Jilin Province | Nongan xian | 4·178 | 6·793 | 22·530 | 0·731 | 4·661 | 14895 | 2585 | 16615 |
| Jilin Province | Nongan xian | 4·409 | 6·892 | 23·714 | 0·758 | 5·050 | 34220 | 5879 | 39224 |
| Jilin Province | Panshishi | 4·067 | 6·585 | 21·775 | 0·712 | 4·577 | 15564 | 2749 | 17682 |
| Jilin Province | Qianan xian | 4·454 | 6·729 | 23·482 | 0·771 | 5·218 | 9123 | 1564 | 10762 |
| Jilin Province | Qianguoerluomenggu zuzizhi xian | 4·499 | 6·895 | 24·074 | 0·763 | 5·204 | 35451 | 5940 | 41311 |
| Jilin Province | Shulanshi | 4·395 | 6·893 | 23·363 | 0·746 | 5·015 | 17432 | 2960 | 19819 |
| Jilin Province | Suanliao xian | 4·342 | 7·007 | 23·611 | 0·729 | 4·906 | 14476 | 2486 | 16262 |
| Jilin Province | Taonanshi | 4·542 | 6·870 | 24·347 | 0·804 | 5·216 | 18116 | 3199 | 20908 |
| Jilin Province | Tonghua xian | 3·671 | 5·833 | 20·207 | 0·665 | 4·080 | 6026 | 1096 | 6762 |
| Jilin Province | Tonghuashi | 3·639 | 5·774 | 19·888 | 0·665 | 4·042 | 16544 | 2974 | 18210 |
| Jilin Province | Tongyu xian | 4·405 | 6·824 | 23·661 | 0·779 | 5·023 | 12327 | 2204 | 13910 |
| Jilin Province | Tumenshi | 4·233 | 6·863 | 22·075 | 0·798 | 4·709 | 805 | 150 | 892 |
| Jilin Province | Wangqing xian | 4·323 | 6·984 | 23·358 | 0·799 | 4·799 | 6825 | 1261 | 7542 |
| Jilin Province | Wushong xian | 3·900 | 6·192 | 21·374 | 0·722 | 4·330 | 9543 | 1737 | 10576 |
| Jilin Province | Yanjishi | 4·225 | 6·853 | 22·644 | 0·788 | 4·607 | 12718 | 2458 | 14319 |
| Jilin Province | Yitongmazuzizi xian | 4·127 | 6·643 | 21·943 | 0·722 | 4·667 | 14705 | 2556 | 16736 |
| Jilin Province | Yongji xian | 4·264 | 6·845 | 22·914 | 0·734 | 4·832 | 22600 | 3828 | 25761 |
| Jilin Province | Yushushi | 4·408 | 6·815 | 23·260 | 0·748 | 5·093 | 37264 | 6287 | 43210 |
| Jilin Province | Zhenlai xian | 4·582 | 6·911 | 24·574 | 0·812 | 5·246 | 11933 | 2123 | 13679 |
| Liaoning Province | Anshanshi | 3·593 | 6·070 | 20·253 | 0·636 | 3·822 | 41166 | 7182 | 43237 |
| Liaoning Province | Beipiaoshi | 3·829 | 6·174 | 21·054 | 0·655 | 4·267 | 18665 | 3211 | 20614 |
| Liaoning Province | Beizhenman zuzizhi xian | 3·727 | 6·148 | 20·337 | 0·641 | 4·082 | 14410 | 2476 | 15847 |
| Liaoning Province | Benximan zuzizhi xian | 3·551 | 5·743 | 19·675 | 0·650 | 3·891 | 8574 | 1557 | 9434 |
| Liaoning Province | Benxishi | 3·660 | 6·043 | 20·246 | 0·641 | 3·991 | 29691 | 5247 | 32531 |
| Liaoning Province | Changtu xian | 4·094 | 6·517 | 22·044 | 0·694 | 4·693 | 28735 | 4884 | 33022 |
| Liaoning Province | Chaoyang xian | 3·683 | 5·833 | 20·443 | 0·639 | 4·094 | 16103 | 2786 | 17784 |
| Liaoning Province | Chaoyangshi | 3·733 | 5·869 | 20·509 | 0·653 | 4·218 | 24671 | 4198 | 27938 |
| Liaoning Province | Dalianshi | 2·762 | 4·492 | 15·624 | 0·495 | 2·967 | 58765 | 10493 | 63608 |
| Liaoning Province | Dandongshi | 3·105 | 5·032 | 17·142 | 0·612 | 3·339 | 14987 | 2908 | 16010 |
| Liaoning Province | Dawa xian | 3·545 | 5·907 | 19·781 | 0·628 | 3·813 | 8710 | 1542 | 9318 |
| Liaoning Province | Dengta xian | 3·697 | 6·120 | 20·661 | 0·652 | 4·009 | 16977 | 3005 | 18187 |
| Liaoning Province | Donggangshi | 3·090 | 5·094 | 17·389 | 0·596 | 3·297 | 12833 | 2436 | 13743 |
| Liaoning Province | Faku xian | 3·942 | 6·173 | 21·396 | 0·677 | 4·488 | 18247 | 3106 | 20781 |
| Liaoning Province | Fenchengman zuzizhi xian | 3·319 | 5·391 | 18·343 | 0·632 | 3·600 | 17021 | 3229 | 18135 |
| Liaoning Province | Fushun xian | 3·728 | 5·885 | 20·533 | 0·658 | 4·188 | 40862 | 7245 | 45363 |
| Liaoning Province | Fushunshi | 3·767 | 5·968 | 20·720 | 0·654 | 4·220 | 11728 | 1963 | 13100 |
| Liaoning Province | Fuxinmenggu zuzizhi xian | 3·895 | 6·138 | 21·233 | 0·665 | 4·386 | 24815 | 4267 | 27928 |
| Liaoning Province | Fuxinshi | 3·831 | 6·077 | 20·916 | 0·672 | 4·293 | 25597 | 4382 | 28768 |
| Liaoning Province | Gaizhoushi | 3·273 | 5·513 | 18·460 | 0·585 | 3·471 | 34928 | 6096 | 37215 |

| | | β-thalassaemia carrier prevalence | | | | | Estimated carrier counts | | |
|---|---|---|---|---|---|---|---|---|---|
| ADM1 | ADM2 | Mean | SD | CI95% | Q25 | Q75 | Mean | Q25 | Q75 |
| Liaoning Province | Haichengshi | 3·521 | 5·968 | 19·681 | 0·625 | 3·732 | 38665 | 6911 | 41545 |
| Liaoning Province | Heishan xian | 3·813 | 6·230 | 21·061 | 0·654 | 4·182 | 17749 | 3075 | 19325 |
| Liaoning Province | Huanrenman zuzizhi xian | 3·492 | 5·497 | 18·896 | 0·647 | 3·897 | 7646 | 1399 | 8439 |
| Liaoning Province | Huludaoishi | 3·515 | 5·653 | 19·910 | 0·620 | 3·829 | 27288 | 4930 | 29569 |
| Liaoning Province | Jianchangshi | 3·426 | 5·370 | 18·837 | 0·606 | 3·835 | 14713 | 2596 | 16574 |
| Liaoning Province | Jianping xian | 3·864 | 6·112 | 20·913 | 0·663 | 4·401 | 21607 | 3678 | 24347 |
| Liaoning Province | Jinzhoushi | 3·608 | 5·991 | 20·442 | 0·623 | 3·904 | 33941 | 5821 | 36418 |
| Liaoning Province | Kaiyuanshi | 3·941 | 6·248 | 21·426 | 0·672 | 4·476 | 21362 | 3618 | 24175 |
| Liaoning Province | Kalaqinzuoqimenggu zuzizhi xian | 3·609 | 5·665 | 19·855 | 0·635 | 4·063 | 12086 | 2136 | 13544 |
| Liaoning Province | Kangping xian | 4·104 | 6·530 | 22·323 | 0·690 | 4·650 | 10484 | 1755 | 11794 |
| Liaoning Province | Kuandianman zuzizhi xian | 3·328 | 5·272 | 18·131 | 0·642 | 3·654 | 11955 | 2349 | 12910 |
| Liaoning Province | Liaoyang xian | 3·594 | 6·059 | 20·246 | 0·636 | 3·853 | 6631 | 1146 | 7198 |
| Liaoning Province | Liaoyang xian | 3·708 | 6·261 | 20·846 | 0·638 | 3·961 | 28303 | 4973 | 30341 |
| Liaoning Province | Liaoyangshi | 3·669 | 6·281 | 21·266 | 0·645 | 3·924 | 22553 | 3981 | 23999 |
| Liaoning Province | Liaozhong xian | 3·750 | 6·151 | 20·616 | 0·653 | 4·095 | 15292 | 2656 | 16396 |
| Liaoning Province | Lingyuanshi | 3·582 | 5·593 | 19·596 | 0·633 | 4·025 | 20363 | 3594 | 23307 |
| Liaoning Province | Linhaishi | 3·597 | 5·931 | 20·335 | 0·635 | 3·876 | 19355 | 3496 | 20644 |
| Liaoning Province | Panjinshi | 3·594 | 5·939 | 19·882 | 0·634 | 3·889 | 35104 | 6330 | 37957 |
| Liaoning Province | Qinghemenqu | 3·775 | 5·902 | 20·500 | 0·656 | 4·129 | 1063 | 177 | 1156 |
| Liaoning Province | Qingyuanman zuzizhi xian | 3·831 | 6·114 | 20·937 | 0·659 | 4·323 | 9783 | 1706 | 10927 |
| Liaoning Province | Shenyangshi | 3·785 | 6·046 | 20·825 | 0·659 | 4·218 | 286245 | 49167 | 316285 |
| Liaoning Province | Sipingshi | 4·158 | 6·687 | 22·554 | 0·692 | 4·845 | 19997 | 3449 | 22625 |
| Liaoning Province | Suizhong xian | 3·255 | 5·114 | 18·035 | 0·579 | 3·635 | 12435 | 2228 | 13939 |
| Liaoning Province | Taian xian | 3·712 | 6·248 | 21·013 | 0·639 | 3·960 | 11954 | 2069 | 12700 |
| Liaoning Province | Tiefashi | 3·915 | 6·144 | 20·942 | 0·667 | 4·477 | 3330 | 576 | 3859 |
| Liaoning Province | Tieling xian | 3·838 | 6·026 | 20·757 | 0·665 | 4·365 | 25136 | 4258 | 28140 |
| Liaoning Province | Tielingshi | 3·796 | 6·094 | 21·247 | 0·649 | 4·165 | 2700 | 447 | 2929 |
| Liaoning Province | Wafangdianshi | 3·065 | 5·037 | 17·728 | 0·548 | 3·243 | 22823 | 4121 | 23862 |
| Liaoning Province | Xifeng xian | 3·997 | 6·293 | 21·499 | 0·693 | 4·557 | 8881 | 1503 | 10293 |
| Liaoning Province | Xinbinman zuzizhi xian | 3·646 | 5·793 | 19·628 | 0·652 | 4·113 | 7041 | 1235 | 7944 |
| Liaoning Province | Xingchengshi | 3·351 | 5·280 | 18·759 | 0·600 | 3·697 | 13049 | 2377 | 14306 |
| Liaoning Province | Xinjin xian | 2·985 | 4·956 | 17·056 | 0·534 | 3·172 | 17147 | 3030 | 18356 |
| Liaoning Province | Xinmin xian | 3·834 | 6·038 | 20·899 | 0·660 | 4·312 | 22641 | 3945 | 25566 |
| Liaoning Province | Yi xian | 3·714 | 6·078 | 20·537 | 0·644 | 4·055 | 11358 | 1998 | 12437 |
| Liaoning Province | Yingkoushi | 3·432 | 5·741 | 18·992 | 0·615 | 3·683 | 21639 | 3889 | 23392 |
| Liaoning Province | Yingkoushi | 3·396 | 5·650 | 18·705 | 0·615 | 3·677 | 19455 | 3530 | 21108 |

| | | β-thalassaemia carrier prevalence | | | | | Estimated carrier counts | | |
|---|---|---|---|---|---|---|---|---|---|
| ADM1 | ADM2 | Mean | SD | CI95% | Q25 | Q75 | Mean | Q25 | Q75 |
| Liaoning Province | Youyanman zuzizhi xian | 3·300 | 5·533 | 18·249 | 0·615 | 3·550 | 13424 | 2466 | 14554 |
| Liaoning Province | Zhangwu xian | 3·993 | 6·198 | 21·633 | 0·677 | 4·583 | 13512 | 2276 | 15583 |
| Liaoning Province | Zhuangheshi | 3·078 | 5·151 | 17·360 | 0·567 | 3·288 | 20452 | 3765 | 21788 |
| Ningxia Ningxia Hui Autonomous Region | Guyuan xian | 3·428 | 5·169 | 17·446 | 0·735 | 3·928 | 19246 | 4137 | 22123 |
| Ningxia Ningxia Hui Autonomous Region | Haiyuan xian | 3·642 | 5·694 | 18·506 | 0·728 | 4·161 | 9002 | 1800 | 10271 |
| Ningxia Ningxia Hui Autonomous Region | Helan xian | 4·123 | 6·409 | 22·332 | 0·757 | 4·648 | 22107 | 4221 | 24721 |
| Ningxia Ningxia Hui Autonomous Region | Lingwu xian | 4·022 | 6·065 | 21·589 | 0·757 | 4·614 | 13067 | 2447 | 14895 |
| Ningxia Ningxia Hui Autonomous Region | Pingluo xian | 4·176 | 6·505 | 22·618 | 0·764 | 4·746 | 17847 | 3259 | 20644 |
| Ningxia Ningxia Hui Autonomous Region | Qingtongxiashi | 4·039 | 6·188 | 21·723 | 0·754 | 4·573 | 9614 | 1799 | 10799 |
| Ningxia Ningxia Hui Autonomous Region | Taole xian | 4·145 | 6·435 | 22·710 | 0·755 | 4·713 | 1429 | 262 | 1631 |
| Ningxia Ningxia Hui Autonomous Region | Tongxin xian | 3·764 | 5·611 | 19·430 | 0·742 | 4·336 | 20413 | 3970 | 23344 |
| Ningxia Ningxia Hui Autonomous Region | Wuzhongshi | 4·025 | 6·159 | 21·856 | 0·760 | 4·563 | 19222 | 3547 | 21358 |
| Ningxia Ningxia Hui Autonomous Region | Xiji xian | 3·423 | 5·315 | 17·728 | 0·715 | 3·840 | 10991 | 2322 | 12379 |
| Ningxia Ningxia Hui Autonomous Region | Yanchi xian | 3·923 | 5·784 | 20·526 | 0·774 | 4·518 | 7796 | 1531 | 8922 |
| Ningxia Ningxia Hui Autonomous Region | Yinchuanshi | 4·090 | 6·353 | 22·266 | 0·751 | 4·578 | 67641 | 12489 | 75828 |
| Ningxia Ningxia Hui Autonomous Region | Yongning xian | 4·089 | 6·401 | 22·013 | 0·757 | 4·594 | 13586 | 2476 | 15238 |
| Ningxia Ningxia Hui Autonomous Region | Zhongning xian | 3·967 | 6·076 | 21·367 | 0·738 | 4·499 | 12379 | 2270 | 14092 |
| Ningxia Ningxia Hui Autonomous Region | Zhongwei xian | 3·922 | 6·143 | 20·676 | 0·734 | 4·411 | 16315 | 3005 | 18679 |
| Qinghai Province | Banma xian | 3·524 | 5·085 | 18·304 | 0·753 | 4·012 | 1360 | 289 | 1557 |
| Qinghai Province | Chengduo xian | 5·146 | 6·977 | 25·476 | 1·108 | 6·188 | 2809 | 634 | 3425 |
| Qinghai Province | Dari xian | 4·005 | 5·470 | 19·023 | 0·927 | 4·796 | 1533 | 353 | 1791 |

| ADM1 | ADM2 | β-thalassaemia carrier prevalence Mean | SD | CI95% | Q25 | Q75 | Estimated carrier counts Mean | Q25 | Q75 |
|---|---|---|---|---|---|---|---|---|---|
| Qinghai Province | Datonghuizutu zuzizhi xian | 4·238 | 6·650 | 22·963 | 0·797 | 4·704 | 16463 | 3028 | 18337 |
| Qinghai Province | Delinghashi | 5·703 | 8·470 | 30·830 | 1·002 | 6·554 | 2014 | 337 | 2307 |
| Qinghai Province | Dulan xian | 4·991 | 7·601 | 27·346 | 0·942 | 5·631 | 3348 | 608 | 3805 |
| Qinghai Province | Gande xian | 3·755 | 5·348 | 18·353 | 0·836 | 4·395 | 1577 | 354 | 1827 |
| Qinghai Province | Gangcha xian | 4·462 | 7·099 | 25·132 | 0·824 | 4·898 | 1888 | 346 | 2051 |
| Qinghai Province | Geermushi | 6·490 | 9·165 | 33·458 | 1·291 | 7·643 | 103 | 18 | 122 |
| Qinghai Province | Geermushi | 5·617 | 8·273 | 30·429 | 1·035 | 6·432 | 12260 | 2253 | 14043 |
| Qinghai Province | Gonghe xian | 4·190 | 6·339 | 22·608 | 0·809 | 4·740 | 5757 | 1098 | 6504 |
| Qinghai Province | Guide xian | 3·918 | 5·809 | 20·579 | 0·775 | 4·493 | 4156 | 838 | 4606 |
| Qinghai Province | Guinan xian | 3·933 | 5·827 | 20·602 | 0·789 | 4·498 | 2763 | 545 | 3246 |
| Qinghai Province | Haiyan xian | 4·292 | 6·809 | 23·897 | 0·800 | 4·745 | 1603 | 302 | 1818 |
| Qinghai Province | Hualonghui zuzizhi xian | 3·828 | 5·665 | 20·592 | 0·738 | 4·348 | 8144 | 1566 | 9236 |
| Qinghai Province | Huangyuan xian | 4·195 | 6·551 | 23·188 | 0·799 | 4·670 | 4904 | 900 | 5196 |
| Qinghai Province | Huangzhong xian | 4·080 | 6·137 | 21·957 | 0·782 | 4·646 | 24035 | 4597 | 27252 |
| Qinghai Province | Huzhutu zuzizhi xian | 4·095 | 6·314 | 21·880 | 0·771 | 4·594 | 13851 | 2590 | 15676 |
| Qinghai Province | Jianzha xian | 3·834 | 5·692 | 20·638 | 0·749 | 4·381 | 3743 | 716 | 4274 |
| Qinghai Province | Jiuzhi xian | 3·307 | 4·571 | 16·075 | 0·774 | 3·891 | 945 | 222 | 1110 |
| Qinghai Province | Ledu xian | 3·907 | 5·947 | 21·243 | 0·745 | 4·366 | 9144 | 1749 | 10493 |
| Qinghai Province | Maduo xian | 4·520 | 6·484 | 23·184 | 0·932 | 5·268 | 649 | 132 | 763 |
| Qinghai Province | Maqin xian | 3·944 | 5·633 | 19·685 | 0·848 | 4·613 | 2239 | 458 | 2560 |
| Qinghai Province | Maqu xian | 3·043 | 4·247 | 15·064 | 0·709 | 3·560 | 1768 | 403 | 1991 |
| Qinghai Province | Menyuanhui zuzizhi xian | 4·311 | 6·997 | 23·819 | 0·794 | 4·732 | 5940 | 1080 | 6444 |
| Qinghai Province | Minhehuizutu zuzizhi xian | 3·816 | 5·866 | 21·024 | 0·719 | 4·203 | 11337 | 2152 | 12428 |
| Qinghai Province | Nangqian xian | 5·954 | 7·710 | 28·691 | 1·373 | 7·152 | 5134 | 1259 | 6062 |
| Qinghai Province | Pingan xian | 3·945 | 5·877 | 20·848 | 0·759 | 4·464 | 4473 | 824 | 4914 |
| Qinghai Province | Qilian xian | 4·584 | 7·241 | 25·473 | 0·842 | 5·091 | 2159 | 395 | 2447 |
| Qinghai Province | Qumalai xian | 5·548 | 7·865 | 28·338 | 1·117 | 6·497 | 1737 | 359 | 2112 |
| Qinghai Province | Tianjun xian | 4·626 | 7·060 | 25·302 | 0·848 | 5·196 | 952 | 173 | 1095 |
| Qinghai Province | Tongde xian | 3·820 | 5·641 | 19·595 | 0·787 | 4·369 | 2223 | 474 | 2584 |
| Qinghai Province | Tongren xian | 3·701 | 5·401 | 19·803 | 0·745 | 4·190 | 3756 | 775 | 4263 |
| Qinghai Province | Wulan xian | 4·825 | 7·314 | 26·236 | 0·893 | 5·475 | 5721 | 1067 | 6404 |
| Qinghai Province | Xinghai xian | 4·057 | 5·859 | 20·970 | 0·810 | 4·703 | 3121 | 604 | 3636 |
| Qinghai Province | Xiningshi | 4·103 | 6·243 | 22·473 | 0·786 | 4·615 | 57179 | 11110 | 64726 |
| Qinghai Province | Xunhuasala zuzizhi xian | 3·686 | 5·559 | 19·867 | 0·715 | 4·135 | 3244 | 622 | 3637 |
| Qinghai Province | Yushu xian | 5·372 | 7·142 | 25·964 | 1·202 | 6·452 | 7709 | 1694 | 9530 |
| Qinghai Province | Zaduo xian | 6·067 | 7·980 | 29·091 | 1·293 | 7·436 | 4107 | 919 | 5035 |

| ADM1 | ADM2 | β-thalassaemia carrier prevalence Mean | SD | CI95% | Q25 | Q75 | Estimated carrier counts Mean | Q25 | Q75 |
|---|---|---|---|---|---|---|---|---|---|
| Qinghai Province | Zeku xian | 3·717 | 5·456 | 19·606 | 0·747 | 4·232 | 2789 | 542 | 3232 |
| Qinghai Province | Zhiduo xian | 5·897 | 8·321 | 29·913 | 1·174 | 6·953 | 1957 | 401 | 2325 |
| Shaanxi Province | Ankangshi | 2·253 | 2·604 | 9·137 | 0·744 | 2·753 | 22117 | 7520 | 26836 |
| Shaanxi Province | Ansai xian | 3·834 | 5·515 | 18·967 | 0·773 | 4·571 | 7125 | 1419 | 8587 |
| Shaanxi Province | Baishui xian | 3·310 | 4·471 | 15·396 | 0·756 | 4·020 | 7244 | 1652 | 8827 |
| Shaanxi Province | Baode xian | 4·099 | 6·510 | 22·526 | 0·745 | 4·557 | 4367 | 800 | 4975 |
| Shaanxi Province | Baoji xian | 2·635 | 3·528 | 12·232 | 0·659 | 3·136 | 31867 | 7985 | 37347 |
| Shaanxi Province | Baojishi | 2·570 | 3·495 | 11·785 | 0·641 | 3·103 | 6440 | 1588 | 7863 |
| Shaanxi Province | Bin xian | 3·081 | 4·126 | 14·675 | 0·716 | 3·708 | 10201 | 2376 | 12314 |
| Shaanxi Province | Changan xian | 2·776 | 3·659 | 12·518 | 0·734 | 3·341 | 53154 | 13669 | 63786 |
| Shaanxi Province | Changwu xian | 3·121 | 4·194 | 15·432 | 0·712 | 3·730 | 3865 | 856 | 4557 |
| Shaanxi Province | Chengcheng xian | 3·317 | 4·469 | 15·150 | 0·764 | 4·084 | 10275 | 2357 | 12828 |
| Shaanxi Province | Chenggu xian | 2·046 | 2·512 | 8·692 | 0·613 | 2·484 | 9274 | 2762 | 11415 |
| Shaanxi Province | Chunhua xian | 3·064 | 4·078 | 14·467 | 0·724 | 3·696 | 4047 | 950 | 4860 |
| Shaanxi Province | Dali xian | 3·301 | 4·577 | 14·811 | 0·762 | 4·031 | 19849 | 4552 | 24325 |
| Shaanxi Province | Danfeng xian | 2·958 | 3·879 | 13·149 | 0·825 | 3·523 | 7309 | 2000 | 8779 |
| Shaanxi Province | Dingbai xian | 3·906 | 5·741 | 20·164 | 0·772 | 4·524 | 13890 | 2616 | 16021 |
| Shaanxi Province | Feng xian | 2·362 | 3·171 | 10·765 | 0·621 | 2·815 | 1818 | 468 | 2144 |
| Shaanxi Province | Fengxiang xian | 2·792 | 3·746 | 13·181 | 0·676 | 3·342 | 11914 | 2916 | 14293 |
| Shaanxi Province | Foping xian | 2·358 | 3·014 | 10·217 | 0·679 | 2·832 | 562 | 170 | 674 |
| Shaanxi Province | Fu xian | 3·463 | 4·702 | 16·515 | 0·760 | 4·199 | 5207 | 1153 | 6318 |
| Shaanxi Province | Fufeng xian | 2·773 | 3·617 | 12·977 | 0·709 | 3·317 | 11415 | 2953 | 13716 |
| Shaanxi Province | Fugu xian | 4·234 | 6·781 | 23·179 | 0·765 | 4·678 | 15402 | 2842 | 16723 |
| Shaanxi Province | Fuping xian | 3·150 | 4·210 | 14·450 | 0·761 | 3·837 | 23599 | 5747 | 28428 |
| Shaanxi Province | Ganquan xian | 3·627 | 5·061 | 17·794 | 0·750 | 4·344 | 2751 | 559 | 3236 |
| Shaanxi Province | Gaolingshi | 3·013 | 4·076 | 13·812 | 0·748 | 3·595 | 12694 | 3147 | 15102 |
| Shaanxi Province | Hanchengshi | 3·442 | 4·691 | 16·073 | 0·760 | 4·213 | 11228 | 2547 | 13703 |
| Shaanxi Province | Hanyin xian | 2·182 | 2·577 | 9·048 | 0·710 | 2·651 | 4978 | 1604 | 5987 |
| Shaanxi Province | Hanzhongshi | 2·009 | 2·520 | 8·427 | 0·588 | 2·424 | 10581 | 3114 | 12851 |
| Shaanxi Province | Hengshan xian | 4·150 | 6·432 | 21·459 | 0·783 | 4·738 | 10245 | 1891 | 11519 |
| Shaanxi Province | Heyang xian | 3·367 | 4·634 | 15·205 | 0·765 | 4·112 | 11852 | 2662 | 14423 |
| Shaanxi Province | Hu xian | 2·724 | 3·530 | 12·343 | 0·714 | 3·279 | 15167 | 3980 | 18431 |
| Shaanxi Province | Hua xian | 3·134 | 4·221 | 13·957 | 0·787 | 3·815 | 8210 | 2042 | 10063 |
| Shaanxi Province | Huangling xian | 3·328 | 4·467 | 15·609 | 0·763 | 4·050 | 4613 | 1045 | 5568 |
| Shaanxi Province | Huanglong xian | 3·435 | 4·618 | 16·137 | 0·756 | 4·221 | 1852 | 409 | 2289 |
| Shaanxi Province | Huayinshi | 3·196 | 4·420 | 14·227 | 0·781 | 3·852 | 6276 | 1532 | 7663 |

| ADM1 | ADM2 | β-thalassaemia carrier prevalence Mean | SD | CI95% | Q25 | Q75 | Estimated carrier counts Mean | Q25 | Q75 |
|---|---|---|---|---|---|---|---|---|---|
| Shaanxi Province | Jia xian | 4·119 | 6·365 | 22·112 | 0·762 | 4·683 | 4063 | 756 | 4569 |
| Shaanxi Province | Jingbian xian | 4·032 | 6·113 | 20·545 | 0·790 | 4·646 | 15698 | 3021 | 18393 |
| Shaanxi Province | Jingyang xian | 2·976 | 3·883 | 13·455 | 0·740 | 3·615 | 15786 | 3958 | 19134 |
| Shaanxi Province | Langao xian | 2·097 | 2·319 | 8·131 | 0·744 | 2·566 | 2842 | 1002 | 3462 |
| Shaanxi Province | Lantian xian | 2·920 | 3·854 | 12·928 | 0·769 | 3·554 | 15742 | 4148 | 19151 |
| Shaanxi Province | Lin xian | 4·069 | 6·386 | 22·391 | 0·757 | 4·563 | 16703 | 3067 | 19171 |
| Shaanxi Province | Lintong xian | 3·051 | 4·037 | 13·528 | 0·766 | 3·723 | 25910 | 6582 | 31818 |
| Shaanxi Province | Linyou xian | 2·933 | 3·894 | 13·767 | 0·708 | 3·517 | 2082 | 506 | 2553 |
| Shaanxi Province | Liquan xian | 2·941 | 3·894 | 13·677 | 0·726 | 3·554 | 11631 | 2899 | 13990 |
| Shaanxi Province | Liuba xian | 2·204 | 2·905 | 9·744 | 0·609 | 2·649 | 763 | 207 | 920 |
| Shaanxi Province | Luochuan xian | 3·414 | 4·596 | 16·174 | 0·761 | 4·181 | 6280 | 1429 | 7784 |
| Shaanxi Province | Luonan xian | 3·100 | 4·204 | 13·832 | 0·795 | 3·731 | 11165 | 2906 | 13343 |
| Shaanxi Province | Mei xian | 2·628 | 3·385 | 11·903 | 0·690 | 3·166 | 5219 | 1357 | 6280 |
| Shaanxi Province | Mian xian | 1·948 | 2·373 | 8·201 | 0·589 | 2·376 | 6624 | 1996 | 8092 |
| Shaanxi Province | Mizhi xian | 4·061 | 6·185 | 21·046 | 0·757 | 4·625 | 6831 | 1219 | 7645 |
| Shaanxi Province | Nanzheng xian | 1·802 | 2·096 | 7·362 | 0·582 | 2·206 | 10389 | 3079 | 12455 |
| Shaanxi Province | Ningqiang xian | 1·763 | 2·030 | 7·112 | 0·564 | 2·177 | 4416 | 1431 | 5539 |
| Shaanxi Province | Ningshan xian | 2·467 | 3·140 | 10·790 | 0·700 | 2·975 | 1400 | 411 | 1697 |
| Shaanxi Province | Pingli xian | 2·222 | 2·414 | 8·425 | 0·792 | 2·743 | 4002 | 1348 | 4959 |
| Shaanxi Province | Pucheng xian | 3·247 | 4·387 | 14·798 | 0·757 | 3·958 | 21597 | 5108 | 26310 |
| Shaanxi Province | Qian xian | 2·910 | 3·833 | 13·482 | 0·722 | 3·482 | 14087 | 3520 | 16808 |
| Shaanxi Province | Qianyang xian | 2·811 | 3·725 | 13·099 | 0·686 | 3·388 | 2713 | 670 | 3267 |
| Shaanxi Province | Qingjian xian | 3·931 | 5·867 | 20·228 | 0·741 | 4·582 | 4253 | 806 | 5107 |
| Shaanxi Province | Qishan xian | 2·750 | 3·629 | 12·925 | 0·686 | 3·310 | 10344 | 2615 | 12348 |
| Shaanxi Province | Shangnan xian | 2·918 | 3·684 | 12·706 | 0·855 | 3·511 | 6006 | 1721 | 7186 |
| Shaanxi Province | Shangzhoushi | 2·936 | 3·926 | 12·883 | 0·794 | 3·534 | 13993 | 3698 | 16880 |
| Shaanxi Province | Shanyang xian | 2·780 | 3·534 | 11·762 | 0·820 | 3·364 | 10147 | 3001 | 12098 |
| Shaanxi Province | Shanyuan xian | 3·068 | 4·060 | 14·071 | 0·750 | 3·697 | 9923 | 2403 | 11820 |
| Shaanxi Province | Shenmu xian | 4·251 | 6·771 | 23·148 | 0·764 | 4·716 | 26033 | 4640 | 28896 |
| Shaanxi Province | Shiquan xian | 2·197 | 2·654 | 9·219 | 0·688 | 2·670 | 3416 | 1045 | 4141 |
| Shaanxi Province | Suide xian | 4·013 | 6·111 | 21·183 | 0·750 | 4·581 | 10220 | 1867 | 11609 |
| Shaanxi Province | Taibai xian | 2·459 | 3·247 | 11·056 | 0·654 | 2·966 | 1012 | 255 | 1179 |
| Shaanxi Province | Tongchuanshi | 3·266 | 4·469 | 15·367 | 0·750 | 3·936 | 7386 | 1758 | 8964 |
| Shaanxi Province | Tongguan xian | 3·239 | 4·452 | 14·484 | 0·775 | 3·873 | 3777 | 885 | 4521 |
| Shaanxi Province | Weinanshi | 3·140 | 4·234 | 14·126 | 0·780 | 3·842 | 28731 | 7072 | 35374 |
| Shaanxi Province | Wubao xian | 3·971 | 6·057 | 21·160 | 0·750 | 4·580 | 444 | 86 | 518 |

| ADM1 | ADM2 | β-thalassaemia carrier prevalence Mean | SD | CI95% | Q25 | Q75 | Estimated carrier counts Mean | Q25 | Q75 |
|---|---|---|---|---|---|---|---|---|---|
| Shaanxi Province | Wugong xian | 2·791 | 3·633 | 12·738 | 0·711 | 3·377 | 13869 | 3481 | 16832 |
| Shaanxi Province | Wuqi xian | 3·749 | 5·329 | 18·776 | 0·772 | 4·416 | 5399 | 1102 | 6411 |
| Shaanxi Province | Xianshi | 2·901 | 3·842 | 13·033 | 0·746 | 3·494 | 206996 | 53754 | 248319 |
| Shaanxi Province | Xianyangshi | 2·911 | 3·895 | 13·741 | 0·720 | 3·488 | 41703 | 10374 | 50191 |
| Shaanxi Province | Xingpingshi | 2·837 | 3·646 | 12·948 | 0·712 | 3·414 | 12029 | 3011 | 14458 |
| Shaanxi Province | Xixiang xian | 2·019 | 2·384 | 8·371 | 0·641 | 2·460 | 6570 | 2049 | 7871 |
| Shaanxi Province | Xunyang xian | 2·450 | 2·869 | 9·959 | 0·794 | 3·003 | 8631 | 2780 | 10508 |
| Shaanxi Province | Xunyi xian | 3·163 | 4·234 | 14·905 | 0·735 | 3·803 | 6895 | 1643 | 8302 |
| Shaanxi Province | Yananshi | 3·763 | 5·383 | 18·913 | 0·750 | 4·449 | 22980 | 4540 | 27098 |
| Shaanxi Province | Yanchang xian | 3·741 | 5·264 | 18·899 | 0·746 | 4·457 | 4528 | 903 | 5393 |
| Shaanxi Province | Yanchuan xian | 3·865 | 5·581 | 19·587 | 0·753 | 4·554 | 4428 | 860 | 5160 |
| Shaanxi Province | Yang xian | 2·213 | 2·794 | 9·563 | 0·642 | 2·668 | 7526 | 2176 | 9027 |
| Shaanxi Province | Yao xian | 3·166 | 4·267 | 14·714 | 0·749 | 3·837 | 12668 | 3068 | 15698 |
| Shaanxi Province | Yichuan xian | 3·604 | 5·020 | 17·684 | 0·742 | 4·369 | 3954 | 800 | 4734 |
| Shaanxi Province | Yijun xian | 3·317 | 4·473 | 15·560 | 0·755 | 4·016 | 2793 | 637 | 3393 |
| Shaanxi Province | Yongjishi | 3·350 | 4·642 | 15·220 | 0·766 | 4·072 | 13251 | 3046 | 16012 |
| Shaanxi Province | Yongshou xian | 3·016 | 3·999 | 14·196 | 0·723 | 3·644 | 4374 | 1085 | 5302 |
| Shaanxi Province | Yulinshi | 4·238 | 6·684 | 22·886 | 0·772 | 4·716 | 41760 | 7731 | 46017 |
| Shaanxi Province | Zhenan xian | 2·532 | 3·147 | 10·821 | 0·758 | 3·069 | 6595 | 1918 | 7812 |
| Shaanxi Province | Zhenba xian | 1·856 | 2·099 | 7·318 | 0·638 | 2·267 | 3937 | 1355 | 4760 |
| Shaanxi Province | Zhengning xian | 3·199 | 4·330 | 15·312 | 0·738 | 3·859 | 3412 | 789 | 4095 |
| Shaanxi Province | Zhidan xian | 3·716 | 5·221 | 18·426 | 0·777 | 4·444 | 5802 | 1205 | 6870 |
| Shaanxi Province | Zhiyang xian | 2·016 | 2·285 | 8·078 | 0·690 | 2·456 | 3600 | 1224 | 4400 |
| Shaanxi Province | Zhouzhi xian | 2·666 | 3·472 | 12·159 | 0·695 | 3·215 | 17329 | 4375 | 20889 |
| Shaanxi Province | Zichang xian | 3·992 | 5·938 | 20·071 | 0·770 | 4·654 | 9637 | 1873 | 11061 |
| Shaanxi Province | Zizhou xian | 4·033 | 6·055 | 20·736 | 0·771 | 4·662 | 5495 | 1057 | 6446 |
| Shaanxi Province | Zuoshui xian | 2·716 | 3·545 | 11·834 | 0·761 | 3·268 | 3559 | 1006 | 4280 |
| Shandong Province | Anqiushi | 1·843 | 3·204 | 10·132 | 0·329 | 1·984 | 17999 | 3184 | 19561 |
| Shandong Province | Binzhoushi | 2·477 | 4·164 | 14·131 | 0·437 | 2·646 | 19858 | 3494 | 21329 |
| Shandong Province | Boxing xian | 2·295 | 3·879 | 12·735 | 0·416 | 2·508 | 13323 | 2413 | 14482 |
| Shandong Province | Changle xian | 1·963 | 3·416 | 10·581 | 0·353 | 2·114 | 8906 | 1607 | 9587 |
| Shandong Province | Changqing xian | 2·345 | 3·799 | 12·775 | 0·454 | 2·554 | 13252 | 2543 | 14687 |
| Shandong Province | Changshang xian | 1·386 | 2·121 | 7·124 | 0·300 | 1·566 | 14914 | 3245 | 16566 |
| Shandong Province | Changyishi | 1·980 | 3·338 | 10·909 | 0·354 | 2·192 | 12869 | 2295 | 14188 |
| Shandong Province | Chengwu xian | 2·179 | 3·258 | 11·412 | 0·450 | 2·517 | 10963 | 2273 | 12769 |
| Shandong Province | Cheping xian | 2·612 | 4·214 | 14·233 | 0·504 | 2·834 | 17161 | 3290 | 18597 |

| | | β-thalassaemia carrier prevalence | | | | | Estimated carrier counts | | |
|---|---|---|---|---|---|---|---|---|---|
| ADM1 | ADM2 | Mean | SD | CI95% | Q25 | Q75 | Mean | Q25 | Q75 |
| Shandong Province | Dezhoushi | 2·770 | 4·427 | 15·161 | 0·533 | 2·999 | 20156 | 3897 | 21784 |
| Shandong Province | Dingtao xian | 2·400 | 3·742 | 12·622 | 0·478 | 2·695 | 12415 | 2470 | 14005 |
| Shandong Province | Donger xian | 2·508 | 4·068 | 13·974 | 0·482 | 2·699 | 10469 | 2000 | 11380 |
| Shandong Province | Dongping xian | 2·345 | 3·785 | 12·695 | 0·459 | 2·537 | 13206 | 2573 | 14326 |
| Shandong Province | Dongyingshi | 2·275 | 3·792 | 12·481 | 0·417 | 2·484 | 3891 | 709 | 4189 |
| Shandong Province | Dongyingshi | 2·518 | 4·137 | 13·942 | 0·458 | 2·768 | 3900 | 703 | 4249 |
| Shandong Province | Fan xian | 2·594 | 4·097 | 13·982 | 0·520 | 2·815 | 8436 | 1670 | 9184 |
| Shandong Province | Fei xian | 1·522 | 2·351 | 7·866 | 0·325 | 1·700 | 13271 | 2844 | 14768 |
| Shandong Province | Feicheng xian | 2·309 | 3·779 | 12·749 | 0·442 | 2·495 | 20772 | 3992 | 22388 |
| Shandong Province | Ganyu xian | 1·192 | 1·880 | 6·099 | 0·251 | 1·342 | 10185 | 2148 | 11516 |
| Shandong Province | Gaomishi | 1·766 | 2·973 | 9·562 | 0·318 | 1·935 | 16348 | 2944 | 17999 |
| Shandong Province | Gaoqing xian | 2·395 | 4·033 | 13·170 | 0·434 | 2·620 | 7703 | 1366 | 8389 |
| Shandong Province | Gaotangixan | 2·661 | 4·348 | 14·732 | 0·513 | 2·881 | 12831 | 2456 | 13789 |
| Shandong Province | Guangrao xian | 2·228 | 3·897 | 12·040 | 0·401 | 2·428 | 12456 | 2216 | 13559 |
| Shandong Province | Haixing xian | 2·740 | 4·326 | 15·138 | 0·511 | 3·026 | 5515 | 1001 | 6193 |
| Shandong Province | Haiyang xian | 1·818 | 2·983 | 9·602 | 0·336 | 2·026 | 8432 | 1525 | 9559 |
| Shandong Province | Hezheshi | 2·520 | 3·989 | 13·613 | 0·501 | 2·770 | 43851 | 8676 | 48091 |
| Shandong Province | Huantan xian | 2·282 | 3·921 | 12·516 | 0·417 | 2·455 | 17535 | 3218 | 18832 |
| Shandong Province | Huimin xian | 2·493 | 4·104 | 13·814 | 0·454 | 2·698 | 14386 | 2609 | 15558 |
| Shandong Province | Jiaonanshi | 1·500 | 2·411 | 8·011 | 0·280 | 1·665 | 11771 | 2200 | 13122 |
| Shandong Province | Jiaozhoushi | 1·659 | 2·743 | 9·031 | 0·305 | 1·813 | 16724 | 2978 | 18405 |
| Shandong Province | Jiaxiang xian | 2·234 | 3·490 | 12·015 | 0·446 | 2·472 | 18118 | 3617 | 20224 |
| Shandong Province | Jimoshi | 1·719 | 2·775 | 9·301 | 0·315 | 1·895 | 26167 | 4879 | 29132 |
| Shandong Province | Jinanshi | 2·328 | 3·921 | 12·644 | 0·443 | 2·533 | 111775 | 21236 | 121072 |
| Shandong Province | Jiningshi | 2·095 | 3·251 | 11·105 | 0·415 | 2·325 | 29126 | 5756 | 31731 |
| Shandong Province | Jinxiang xian | 2·074 | 3·209 | 10·757 | 0·416 | 2·361 | 13962 | 2782 | 15802 |
| Shandong Province | Jiyang xian | 2·434 | 4·015 | 13·423 | 0·465 | 2·650 | 13579 | 2594 | 15029 |
| Shandong Province | Ju xian | 1·559 | 2·596 | 8·645 | 0·296 | 1·681 | 14877 | 2805 | 16129 |
| Shandong Province | Junan xian | 1·361 | 2·215 | 7·156 | 0·270 | 1·507 | 11845 | 2354 | 13055 |
| Shandong Province | Juye xian | 2·311 | 3·687 | 12·375 | 0·451 | 2·564 | 20787 | 4091 | 23224 |
| Shandong Province | Kenli xian | 2·363 | 3·924 | 12·991 | 0·432 | 2·604 | 24640 | 4484 | 27196 |
| Shandong Province | Laiwushi | 2·059 | 3·436 | 11·033 | 0·391 | 2·245 | 24527 | 4647 | 26639 |
| Shandong Province | Laixishi | 1·902 | 3·180 | 10·401 | 0·337 | 2·073 | 13226 | 2375 | 14476 |
| Shandong Province | Laiyangshi | 1·874 | 3·090 | 10·075 | 0·338 | 2·085 | 15102 | 2726 | 16593 |
| Shandong Province | Laizhoushi | 2·059 | 3·465 | 11·301 | 0·366 | 2·273 | 16531 | 2931 | 18130 |
| Shandong Province | Leningshi | 2·685 | 4·373 | 15·019 | 0·497 | 2·905 | 14554 | 2670 | 15759 |

| | | β-thalassaemia carrier prevalence | | | | | Estimated carrier counts | | |
|---|---|---|---|---|---|---|---|---|---|
| ADM1 | ADM2 | Mean | SD | CI95% | Q25 | Q75 | Mean | Q25 | Q75 |
| Shandong Province | Liangshan xian | 2·435 | 3·929 | 13·165 | 0·478 | 2·652 | 18380 | 3634 | 19996 |
| Shandong Province | Liaochengshi | 2·644 | 4·299 | 14·898 | 0·516 | 2·843 | 34591 | 6810 | 37073 |
| Shandong Province | Lijin xian | 2·421 | 3·952 | 13·422 | 0·448 | 2·644 | 6541 | 1208 | 7044 |
| Shandong Province | Ling xian | 2·707 | 4·480 | 14·917 | 0·518 | 2·904 | 21189 | 4054 | 22799 |
| Shandong Province | Linju xian | 1·925 | 3·396 | 10·468 | 0·352 | 2·051 | 15797 | 2915 | 16930 |
| Shandong Province | Linqingshi | 2·778 | 4·433 | 15·473 | 0·537 | 3·028 | 21618 | 4237 | 23675 |
| Shandong Province | Linshu xian | 1·268 | 2·041 | 6·431 | 0·270 | 1·414 | 7358 | 1560 | 8247 |
| Shandong Province | Linyi xian | 2·558 | 4·189 | 13·996 | 0·493 | 2·762 | 12461 | 2394 | 13576 |
| Shandong Province | Linyishi | 1·394 | 2·230 | 7·220 | 0·292 | 1·552 | 47493 | 10009 | 52958 |
| Shandong Province | Longkoushi | 2·210 | 3·737 | 11·761 | 0·387 | 2·432 | 12795 | 2212 | 13985 |
| Shandong Province | Mengyin xian | 1·722 | 2·777 | 9·199 | 0·344 | 1·892 | 7720 | 1526 | 8466 |
| Shandong Province | Muping xian | 1·963 | 3·297 | 10·517 | 0·361 | 2·169 | 11392 | 2066 | 12706 |
| Shandong Province | Ningjin xian | 2·729 | 4·404 | 15·542 | 0·515 | 2·942 | 13066 | 2429 | 14070 |
| Shandong Province | Ningyang xian | 2·108 | 3·391 | 11·386 | 0·410 | 2·319 | 14051 | 2763 | 15434 |
| Shandong Province | Penglai xian | 2·200 | 3·693 | 11·833 | 0·392 | 2·426 | 10930 | 1930 | 11954 |
| Shandong Province | Pingdushi | 1·919 | 3·209 | 10·777 | 0·338 | 2·085 | 23011 | 4040 | 25087 |
| Shandong Province | Pingyi xian | 1·672 | 2·579 | 8·798 | 0·344 | 1·881 | 15283 | 3107 | 17133 |
| Shandong Province | Pingyin xian | 2·446 | 3·988 | 13·631 | 0·470 | 2·640 | 8493 | 1631 | 9032 |
| Shandong Province | Pingyuan xian | 2·705 | 4·476 | 14·941 | 0·513 | 2·934 | 11438 | 2217 | 12376 |
| Shandong Province | Puyang xian | 2·692 | 4·282 | 14·897 | 0·525 | 2·945 | 18764 | 3649 | 20564 |
| Shandong Province | Qihe xian | 2·481 | 4·064 | 13·486 | 0·480 | 2·708 | 18481 | 3595 | 20126 |
| Shandong Province | Qingdaoshi | 1·604 | 2·525 | 8·588 | 0·298 | 1·789 | 44711 | 8300 | 50426 |
| Shandong Province | Qingdaoshi | 1·584 | 2·598 | 8·474 | 0·290 | 1·727 | 1183 | 213 | 1277 |
| Shandong Province | Qingyun xian | 2·648 | 4·193 | 15·158 | 0·491 | 2·927 | 8274 | 1539 | 9005 |
| Shandong Province | Qingzhoushi | 2·058 | 3·563 | 11·214 | 0·374 | 2·228 | 19003 | 3421 | 20231 |
| Shandong Province | Qixiashi | 2·041 | 3·453 | 11·080 | 0·364 | 2·250 | 9050 | 1620 | 10013 |
| Shandong Province | Qufushi | 1·988 | 3·137 | 10·448 | 0·400 | 2·195 | 12986 | 2611 | 14415 |
| Shandong Province | Rizhaoshi | 1·383 | 2·271 | 7·455 | 0·264 | 1·529 | 16553 | 3145 | 18271 |
| Shandong Province | Rushanshi | 1·821 | 3·007 | 9·617 | 0·340 | 2·032 | 7580 | 1404 | 8453 |
| Shandong Province | Shanghe xian | 2·561 | 4·275 | 14·340 | 0·475 | 2·744 | 11929 | 2192 | 12824 |
| Shandong Province | Shouguangshi | 2·124 | 3·676 | 11·440 | 0·383 | 2·319 | 27800 | 5015 | 30105 |
| Shandong Province | Sishui xian | 1·872 | 2·915 | 9·937 | 0·373 | 2·071 | 9880 | 1970 | 11061 |
| Shandong Province | Taianshi | 2·165 | 3·581 | 11·699 | 0·415 | 2·353 | 42146 | 7905 | 45492 |
| Shandong Province | Taiqian xian | 2·545 | 4·103 | 14·099 | 0·496 | 2·755 | 8420 | 1643 | 9180 |
| Shandong Province | Tancheng xian | 1·241 | 1·956 | 6·224 | 0·272 | 1·391 | 12242 | 2690 | 13858 |
| Shandong Province | Tengzhoushi | 1·693 | 2·563 | 8·697 | 0·358 | 1·918 | 33298 | 7010 | 37910 |

| | | β-thalassaemia carrier prevalence | | | | | Estimated carrier counts | | |
|---|---|---|---|---|---|---|---|---|---|
| **ADM1** | **ADM2** | **Mean** | **SD** | **CI95%** | **Q25** | **Q75** | **Mean** | **Q25** | **Q75** |
| Shandong Province | Weifangshi | 1·977 | 3·366 | 10·768 | 0·361 | 2·181 | 42445 | 7874 | 47176 |
| Shandong Province | Weishan xian | 1·716 | 2·613 | 8·798 | 0·364 | 1·945 | 13789 | 2933 | 15847 |
| Shandong Province | Wenshang xian | 2·209 | 3·446 | 11·870 | 0·443 | 2·430 | 14915 | 3006 | 16270 |
| Shandong Province | Wucheng xian | 2·800 | 4·563 | 15·360 | 0·539 | 3·015 | 9857 | 1893 | 10645 |
| Shandong Province | Wuli xian | 2·618 | 4·137 | 14·306 | 0·487 | 2·924 | 13539 | 2506 | 15193 |
| Shandong Province | Wulian xian | 1·531 | 2·511 | 8·290 | 0·289 | 1·679 | 6315 | 1190 | 7115 |
| Shandong Province | Xiajin xian | 2·800 | 4·537 | 15·380 | 0·542 | 3·032 | 11836 | 2303 | 12874 |
| Shandong Province | Xin xian | 2·702 | 4·434 | 15·107 | 0·521 | 2·887 | 24453 | 4703 | 26149 |
| Shandong Province | Xintaishi | 1·884 | 2·983 | 10·022 | 0·373 | 2·083 | 25881 | 5206 | 28539 |
| Shandong Province | Yanggu xian | 2·558 | 4·082 | 14·149 | 0·509 | 2·755 | 20121 | 3971 | 21814 |
| Shandong Province | Yangxin xian | 2·585 | 4·173 | 14·264 | 0·468 | 2·835 | 10497 | 1909 | 11485 |
| Shandong Province | Yanshan xian | 2·753 | 4·315 | 15·381 | 0·512 | 3·039 | 10461 | 1964 | 11710 |
| Shandong Province | Yantaishi | 2·098 | 3·595 | 11·389 | 0·379 | 2·293 | 33526 | 5999 | 36410 |
| Shandong Province | Yanzhoushi | 2·093 | 3·307 | 11·171 | 0·416 | 2·306 | 14485 | 2823 | 16038 |
| Shandong Province | Yinan xian | 1·575 | 2·601 | 8·612 | 0·313 | 1·706 | 13535 | 2659 | 14583 |
| Shandong Province | Yishui xian | 1·741 | 2·994 | 9·690 | 0·324 | 1·851 | 16894 | 3061 | 17651 |
| Shandong Province | Yiyuan xian | 1·851 | 3·091 | 10·003 | 0·356 | 2·003 | 10325 | 1973 | 11195 |
| Shandong Province | Yucheng xian | 2·581 | 4·301 | 14·208 | 0·496 | 2·776 | 12027 | 2317 | 12992 |
| Shandong Province | Yuncheng xian | 2·462 | 3·927 | 13·463 | 0·480 | 2·708 | 29939 | 5836 | 32830 |
| Shandong Province | Yutai xian | 1·942 | 3·016 | 9·948 | 0·401 | 2·211 | 7316 | 1523 | 8370 |
| Shandong Province | Zhangqiushi | 2·282 | 3·809 | 12·256 | 0·428 | 2·477 | 25500 | 4770 | 27615 |
| Shandong Province | Zhanhua xian | 2·523 | 4·063 | 14·048 | 0·461 | 2·771 | 8447 | 1524 | 9246 |
| Shandong Province | Zhaoyuanshi | 2·107 | 3·558 | 11·564 | 0·376 | 2·313 | 11685 | 2110 | 12907 |
| Shandong Province | Zhaozhuangshi | 1·481 | 2·266 | 7·575 | 0·318 | 1·686 | 28589 | 6142 | 32557 |
| Shandong Province | Zhencheng xian | 2·561 | 4·081 | 13·789 | 0·499 | 2·794 | 15759 | 3154 | 17367 |
| Shandong Province | Zhouchengshi | 1·853 | 2·846 | 9·469 | 0·384 | 2·088 | 23055 | 4873 | 25864 |
| Shandong Province | Zhouping xian | 2·337 | 3·969 | 12·527 | 0·431 | 2·532 | 20548 | 3739 | 22099 |
| Shandong Province | Zhuchengshi | 1·617 | 2·667 | 8·783 | 0·303 | 1·770 | 17947 | 3380 | 19808 |
| Shandong Province | Ziboshi | 2·114 | 3·573 | 11·270 | 0·392 | 2·315 | 64468 | 11828 | 71254 |
| Shanghai Municipality | Chongming xian | 0·405 | 0·498 | 1·614 | 0·137 | 0·486 | 5130 | 1721 | 6125 |
| Shanghai Municipality | Jinshan xian | 0·431 | 0·524 | 1·755 | 0·141 | 0·516 | 6988 | 2267 | 8307 |
| Shanghai Municipality | Shanghaishi | 0·418 | 0·513 | 1·748 | 0·137 | 0·497 | 58219 | 18739 | 69258 |
| Shanghai Municipality | Songjiang xian | 0·495 | 0·658 | 2·209 | 0·148 | 0·577 | 5577 | 1683 | 6485 |
| Shanxi Province | Anzhe xian | 3·579 | 5·384 | 19·110 | 0·682 | 4·082 | 2566 | 482 | 2953 |
| Shanxi Province | Changzhi xian | 3·339 | 4·992 | 17·594 | 0·648 | 3·816 | 15049 | 2945 | 17628 |
| Shanxi Province | Changzhishi | 3·403 | 5·116 | 18·130 | 0·649 | 3·870 | 26832 | 5188 | 30387 |

| ADM1 | ADM2 | β-thalassaemia carrier prevalence Mean | SD | CI95% | Q25 | Q75 | Estimated carrier counts Mean | Q25 | Q75 |
|---|---|---|---|---|---|---|---|---|---|
| Shanxi Province | Changzi xian | 3·491 | 5·221 | 18·589 | 0·663 | 3·999 | 9372 | 1834 | 10793 |
| Shanxi Province | Dagu xian | 3·672 | 5·964 | 19·793 | 0·661 | 4·080 | 12145 | 2215 | 13629 |
| Shanxi Province | Dai xian | 3·930 | 6·524 | 22·529 | 0·695 | 4·211 | 7021 | 1228 | 7514 |
| Shanxi Province | Daling xian | 3·692 | 5·208 | 18·603 | 0·735 | 4·430 | 1889 | 366 | 2268 |
| Shanxi Province | Datong xian | 3·970 | 6·709 | 23·026 | 0·693 | 4·193 | 14824 | 2550 | 15761 |
| Shanxi Province | Datongshi | 4·072 | 6·913 | 23·721 | 0·706 | 4·268 | 68601 | 12159 | 73455 |
| Shanxi Province | Dingxiang xian | 3·853 | 6·299 | 21·999 | 0·680 | 4·160 | 7433 | 1327 | 8113 |
| Shanxi Province | Fangshan xian | 4·035 | 6·489 | 22·315 | 0·739 | 4·468 | 4350 | 794 | 4898 |
| Shanxi Province | Fanshi xian | 3·887 | 6·507 | 22·265 | 0·673 | 4·165 | 9955 | 1698 | 10760 |
| Shanxi Province | Fenxi xian | 3·773 | 5·767 | 19·746 | 0·701 | 4·376 | 3761 | 679 | 4397 |
| Shanxi Province | Fenyang xian | 3·837 | 6·146 | 20·973 | 0·696 | 4·288 | 27807 | 5046 | 31084 |
| Shanxi Province | Fushan xian | 3·591 | 5·341 | 18·625 | 0·706 | 4·144 | 3601 | 697 | 4327 |
| Shanxi Province | Gaopingshi | 3·314 | 4·887 | 17·433 | 0·654 | 3·843 | 14036 | 2796 | 16540 |
| Shanxi Province | Gu xian | 3·684 | 5·707 | 19·726 | 0·683 | 4·175 | 3031 | 565 | 3468 |
| Shanxi Province | Guangling xian | 3·868 | 6·502 | 22·338 | 0·674 | 4·117 | 5820 | 1022 | 6216 |
| Shanxi Province | Gujiaoshi | 3·903 | 6·390 | 21·720 | 0·713 | 4·275 | 8268 | 1502 | 9083 |
| Shanxi Province | Hejin xian | 3·479 | 4·849 | 16·373 | 0·748 | 4·187 | 15629 | 3344 | 18992 |
| Shanxi Province | Hequ xian | 4·151 | 6·688 | 22·388 | 0·753 | 4·642 | 2381 | 424 | 2688 |
| Shanxi Province | Heshun xian | 3·455 | 5·418 | 18·968 | 0·627 | 3·864 | 3802 | 703 | 4226 |
| Shanxi Province | Hongtong xian | 3·701 | 5·651 | 19·724 | 0·700 | 4·238 | 23755 | 4502 | 27029 |
| Shanxi Province | Houmashi | 3·594 | 5·316 | 18·148 | 0·727 | 4·246 | 7501 | 1487 | 8947 |
| Shanxi Province | Huairen xian | 4·065 | 6·934 | 24·227 | 0·714 | 4·194 | 13844 | 2442 | 14144 |
| Shanxi Province | Huguan xian | 3·274 | 4·909 | 17·273 | 0·642 | 3·727 | 8408 | 1584 | 9435 |
| Shanxi Province | Hunyuan xian | 3·959 | 6·689 | 22·999 | 0·687 | 4·154 | 9362 | 1573 | 9706 |
| Shanxi Province | Huozhoushi | 3·792 | 6·045 | 20·837 | 0·679 | 4·265 | 10376 | 1909 | 11743 |
| Shanxi Province | Ji xian | 3·616 | 5·048 | 17·774 | 0·735 | 4·384 | 3189 | 634 | 3912 |
| Shanxi Province | Jiang xian | 3·490 | 5·020 | 17·842 | 0·731 | 4·072 | 7764 | 1610 | 8960 |
| Shanxi Province | Jiaocheng xian | 3·858 | 6·206 | 20·871 | 0·709 | 4·297 | 12005 | 2237 | 13469 |
| Shanxi Province | Jiaokou xian | 3·792 | 5·853 | 19·964 | 0·714 | 4·362 | 2681 | 503 | 3037 |
| Shanxi Province | Jiexiushi | 3·747 | 5·940 | 20·331 | 0·676 | 4·211 | 15048 | 2720 | 16932 |
| Shanxi Province | Jingjing xian | 3·535 | 5·737 | 19·548 | 0·623 | 3·897 | 13275 | 2323 | 14717 |
| Shanxi Province | Jingle xian | 3·943 | 6·460 | 22·130 | 0·717 | 4·370 | 4773 | 871 | 5273 |
| Shanxi Province | Jishan xian | 3·490 | 4·940 | 17·014 | 0·747 | 4·163 | 10812 | 2302 | 12749 |
| Shanxi Province | Kelan xian | 4·007 | 6·417 | 21·418 | 0·738 | 4·489 | 2800 | 514 | 3184 |
| Shanxi Province | Lan xian | 3·972 | 6·445 | 21·294 | 0·734 | 4·454 | 5760 | 1061 | 6251 |
| Shanxi Province | Licheng xian | 3·400 | 5·204 | 18·422 | 0·625 | 3·846 | 4489 | 813 | 5044 |

| | | β-thalassaemia carrier prevalence | | | | | Estimated carrier counts | | |
|---|---|---|---|---|---|---|---|---|---|
| ADM1 | ADM2 | Mean | SD | CI95% | Q25 | Q75 | Mean | Q25 | Q75 |
| Shanxi Province | Linfenshi | 3·688 | 5·482 | 19·319 | 0·703 | 4·316 | 37683 | 7113 | 44453 |
| Shanxi Province | Lingchuan xian | 3·212 | 4·795 | 16·804 | 0·648 | 3·659 | 6971 | 1404 | 7931 |
| Shanxi Province | Lingqiu xian | 3·772 | 6·317 | 21·142 | 0·658 | 4·066 | 8066 | 1383 | 8703 |
| Shanxi Province | Lingshi xian | 3·787 | 5·979 | 20·603 | 0·695 | 4·278 | 9332 | 1697 | 10581 |
| Shanxi Province | Linqi xian | 3·404 | 4·723 | 16·024 | 0·765 | 4·107 | 16077 | 3571 | 19349 |
| Shanxi Province | Lishi xian | 3·909 | 6·132 | 21·277 | 0·729 | 4·376 | 18932 | 3544 | 20971 |
| Shanxi Province | Liulin xian | 3·919 | 6·050 | 21·202 | 0·734 | 4·431 | 12038 | 2248 | 13616 |
| Shanxi Province | Loufan xian | 3·931 | 6·470 | 21·837 | 0·721 | 4·298 | 3635 | 662 | 3889 |
| Shanxi Province | Luchengshi | 3·376 | 5·142 | 18·106 | 0·643 | 3·852 | 9130 | 1709 | 10369 |
| Shanxi Province | Meng xian | 3·777 | 6·202 | 21·835 | 0·658 | 4·064 | 10927 | 1930 | 11831 |
| Shanxi Province | Ningwu xian | 3·998 | 6·483 | 22·548 | 0·710 | 4·382 | 5450 | 971 | 6041 |
| Shanxi Province | Pianguan xian | 4·167 | 6·806 | 23·073 | 0·743 | 4·588 | 3136 | 561 | 3445 |
| Shanxi Province | Pingding xian | 3·546 | 5·652 | 19·516 | 0·633 | 3·928 | 11966 | 2184 | 13343 |
| Shanxi Province | Pinglu xian | 3·368 | 4·725 | 16·227 | 0·763 | 4·007 | 13518 | 3179 | 16133 |
| Shanxi Province | Pingluqu | 4·116 | 6·819 | 23·595 | 0·722 | 4·437 | 5746 | 996 | 6130 |
| Shanxi Province | Pingshun xian | 3·335 | 5·084 | 18·047 | 0·626 | 3·788 | 4345 | 819 | 4962 |
| Shanxi Province | Pingyao xian | 3·729 | 6·010 | 20·228 | 0·678 | 4·150 | 17406 | 3187 | 19579 |
| Shanxi Province | Pu xian | 3·745 | 5·481 | 19·088 | 0·719 | 4·370 | 3374 | 632 | 3854 |
| Shanxi Province | Qi xian | 3·755 | 6·158 | 20·472 | 0·663 | 4·143 | 9256 | 1621 | 9934 |
| Shanxi Province | Qin xian | 3·624 | 5·668 | 19·599 | 0·654 | 4·059 | 4647 | 808 | 5147 |
| Shanxi Province | Qingxu xian | 3·837 | 6·290 | 20·991 | 0·678 | 4·188 | 13079 | 2319 | 14383 |
| Shanxi Province | Qinshui xian | 3·458 | 5·112 | 18·387 | 0·696 | 3·956 | 7165 | 1442 | 8177 |
| Shanxi Province | Qinyuan xian | 3·716 | 5·931 | 20·163 | 0·672 | 4·138 | 5697 | 997 | 6328 |
| Shanxi Province | Qizhoushi | 3·933 | 6·441 | 22·602 | 0·692 | 4·279 | 22220 | 3895 | 24219 |
| Shanxi Province | Quwo xian | 3·540 | 5·158 | 17·860 | 0·722 | 4·145 | 8744 | 1795 | 10293 |
| Shanxi Province | Shanyin xian | 4·020 | 6·718 | 23·424 | 0·712 | 4·238 | 7884 | 1382 | 8293 |
| Shanxi Province | She xian | 3·305 | 5·064 | 17·861 | 0·609 | 3·731 | 12700 | 2296 | 14288 |
| Shanxi Province | Shenci xian | 4·091 | 6·637 | 23·152 | 0·729 | 4·462 | 3118 | 554 | 3372 |
| Shanxi Province | Shilou xian | 3·856 | 5·814 | 20·158 | 0·721 | 4·441 | 3961 | 716 | 4532 |
| Shanxi Province | Shouyang xian | 3·713 | 6·021 | 20·729 | 0·659 | 4·076 | 7664 | 1314 | 8304 |
| Shanxi Province | Shouzhoushi | 4·024 | 6·646 | 22·740 | 0·710 | 4·328 | 23139 | 4136 | 25161 |
| Shanxi Province | Taiyuanshi | 3·846 | 6·266 | 21·663 | 0·698 | 4·167 | 174879 | 31948 | 189220 |
| Shanxi Province | Tianzhen xian | 4·009 | 6·757 | 23·431 | 0·691 | 4·214 | 6670 | 1115 | 7018 |
| Shanxi Province | Tunliu xian | 3·512 | 5·342 | 18·960 | 0·662 | 3·995 | 5799 | 1118 | 6649 |
| Shanxi Province | Wanrong xian | 3·439 | 4·775 | 16·414 | 0·764 | 4·136 | 12826 | 2825 | 15294 |
| Shanxi Province | Wenshui xian | 3·869 | 6·229 | 21·329 | 0·709 | 4·262 | 11321 | 2075 | 12818 |

| ADM1 | ADM2 | β-thalassaemia carrier prevalence Mean | SD | CI95% | Q25 | Q75 | Estimated carrier counts Mean | Q25 | Q75 |
|---|---|---|---|---|---|---|---|---|---|
| Shanxi Province | Wenxi xian | 3·550 | 5·186 | 18·247 | 0·742 | 4·096 | 13264 | 2734 | 15109 |
| Shanxi Province | Wutai xian | 3·814 | 6·278 | 21·523 | 0·667 | 4·124 | 9191 | 1607 | 9820 |
| Shanxi Province | Wuxiang xian | 3·615 | 5·708 | 19·891 | 0·641 | 4·031 | 5828 | 1014 | 6420 |
| Shanxi Province | Wuzai xian | 4·104 | 6·637 | 22·524 | 0·722 | 4·549 | 4059 | 752 | 4487 |
| Shanxi Province | Xia xian | 3·438 | 4·921 | 16·897 | 0·760 | 4·039 | 11257 | 2458 | 13242 |
| Shanxi Province | Xiangfen xian | 3·635 | 5·336 | 18·575 | 0·719 | 4·308 | 14422 | 2864 | 17136 |
| Shanxi Province | Xiangning xian | 3·575 | 5·058 | 17·834 | 0·731 | 4·278 | 7849 | 1567 | 9388 |
| Shanxi Province | Xiangyuan xian | 3·497 | 5·391 | 18·944 | 0·642 | 3·922 | 9661 | 1787 | 10922 |
| Shanxi Province | Xiaoyishi | 3·836 | 6·107 | 20·774 | 0·699 | 4·309 | 8092 | 1494 | 8838 |
| Shanxi Province | Xing xian | 4·083 | 6·467 | 22·189 | 0·759 | 4·542 | 6587 | 1230 | 7473 |
| Shanxi Province | Xinjiang xian | 3·539 | 5·207 | 17·566 | 0·738 | 4·146 | 10191 | 2092 | 11975 |
| Shanxi Province | Xiyang xian | 3·467 | 5·513 | 19·491 | 0·620 | 3·828 | 6552 | 1184 | 7394 |
| Shanxi Province | Yangcheng xian | 3·354 | 4·784 | 16·920 | 0·712 | 3·912 | 10147 | 2104 | 11839 |
| Shanxi Province | Yanggao xian | 3·996 | 6·750 | 23·140 | 0·694 | 4·220 | 7315 | 1252 | 7722 |
| Shanxi Province | Yangqu xian | 3·889 | 6·352 | 22·036 | 0·694 | 4·224 | 5814 | 1030 | 6161 |
| Shanxi Province | Yangquanshi | 3·682 | 6·020 | 20·655 | 0·639 | 4·028 | 25196 | 4425 | 27219 |
| Shanxi Province | Yangyuan xian | 3·931 | 6·592 | 22·704 | 0·679 | 4·167 | 8133 | 1434 | 8465 |
| Shanxi Province | Yicheng xian | 3·554 | 5·206 | 18·180 | 0·713 | 4·108 | 10244 | 2073 | 11827 |
| Shanxi Province | Yin xian | 3·797 | 5·650 | 19·589 | 0·716 | 4·400 | 3723 | 696 | 4378 |
| Shanxi Province | Ying xian | 3·996 | 6·759 | 23·232 | 0·696 | 4·190 | 9971 | 1752 | 10293 |
| Shanxi Province | Yonghe xian | 3·798 | 5·456 | 19·515 | 0·730 | 4·515 | 2024 | 400 | 2447 |
| Shanxi Province | Youyu xian | 4·168 | 6·931 | 24·143 | 0·718 | 4·421 | 3659 | 619 | 3825 |
| Shanxi Province | Yuanpingshi | 3·929 | 6·417 | 22·365 | 0·694 | 4·267 | 15939 | 2770 | 17108 |
| Shanxi Province | Yuanqu xian | 3·413 | 4·855 | 17·288 | 0·746 | 3·989 | 6831 | 1459 | 7941 |
| Shanxi Province | Yucishi | 3·693 | 5·989 | 20·349 | 0·670 | 4·053 | 34531 | 6307 | 37957 |
| Shanxi Province | Yunchengshi | 3·381 | 4·719 | 16·246 | 0·753 | 4·039 | 30668 | 6875 | 36514 |
| Shanxi Province | Yushe xian | 3·569 | 5·643 | 19·252 | 0·645 | 3·978 | 3859 | 718 | 4398 |
| Shanxi Province | Zhongyan xian | 3·869 | 6·025 | 20·929 | 0·709 | 4·353 | 5549 | 1025 | 6341 |
| Shanxi Province | Zuoquan xian | 3·399 | 5·252 | 18·654 | 0·621 | 3·835 | 5141 | 930 | 5889 |
| Shanxi Province | Zuoyun xian | 4·149 | 7·050 | 24·747 | 0·721 | 4·307 | 4680 | 818 | 4905 |
| Sichuan Province | Aba xian | 2·764 | 3·836 | 12·764 | 0·697 | 3·267 | 2323 | 583 | 2718 |
| Sichuan Province | An xian | 1·357 | 1·312 | 4·540 | 0·564 | 1·680 | 6333 | 2635 | 7866 |
| Sichuan Province | Anyue xian | 1·118 | 0·992 | 3·570 | 0·500 | 1·390 | 10079 | 4515 | 12618 |
| Sichuan Province | Baiyu xian | 4·131 | 5·351 | 19·054 | 1·048 | 4·972 | 1898 | 471 | 2258 |
| Sichuan Province | Banma xian | 3·520 | 4·937 | 16·608 | 0·834 | 4·137 | 1099 | 253 | 1283 |
| Sichuan Province | Baoxing xian | 1·592 | 1·515 | 5·318 | 0·651 | 1·991 | 743 | 310 | 921 |

| ADM1 | ADM2 | β-thalassaemia carrier prevalence Mean | SD | CI95% | Q25 | Q75 | Estimated carrier counts Mean | Q25 | Q75 |
|---|---|---|---|---|---|---|---|---|---|
| Sichuan Province | Batang xian | 4·241 | 5·372 | 19·556 | 1·115 | 5·110 | 3011 | 796 | 3534 |
| Sichuan Province | Bazhongshi | 1·276 | 1·187 | 4·207 | 0·540 | 1·600 | 13097 | 5539 | 16495 |
| Sichuan Province | Beichuan xian | 1·475 | 1·537 | 5·247 | 0·564 | 1·836 | 590 | 232 | 733 |
| Sichuan Province | Bi xian | 1·256 | 1·004 | 3·593 | 0·619 | 1·579 | 15489 | 7719 | 19512 |
| Sichuan Province | Butuo xian | 2·196 | 2·318 | 8·116 | 0·808 | 2·735 | 4315 | 1606 | 5415 |
| Sichuan Province | Changning xian | 1·338 | 1·238 | 4·355 | 0·581 | 1·662 | 4727 | 2050 | 5880 |
| Sichuan Province | Changxi xian | 1·284 | 1·206 | 4·269 | 0·533 | 1·615 | 6339 | 2690 | 8011 |
| Sichuan Province | Chengdushi | 1·151 | 0·863 | 3·193 | 0·581 | 1·447 | 106929 | 53650 | 134022 |
| Sichuan Province | Chengkou xian | 1·911 | 2·000 | 6·964 | 0·721 | 2·372 | 3689 | 1360 | 4529 |
| Sichuan Province | Chishuishi | 1·577 | 1·627 | 5·639 | 0·604 | 1·959 | 3672 | 1476 | 4628 |
| Sichuan Province | Chongzhoushi | 1·315 | 1·059 | 3·892 | 0·623 | 1·661 | 14443 | 7031 | 18242 |
| Sichuan Province | Da xian | 1·284 | 1·142 | 4·073 | 0·571 | 1·606 | 12155 | 5373 | 15236 |
| Sichuan Province | Dachuanshi | 1·289 | 1·099 | 4·127 | 0·598 | 1·585 | 11161 | 5133 | 13820 |
| Sichuan Province | Daguan xian | 1·616 | 1·493 | 5·358 | 0·684 | 2·022 | 3405 | 1441 | 4185 |
| Sichuan Province | Danba xian | 2·258 | 2·664 | 9·275 | 0·704 | 2·774 | 1120 | 353 | 1386 |
| Sichuan Province | Danling xian | 1·299 | 1·104 | 3·962 | 0·595 | 1·630 | 1910 | 875 | 2395 |
| Sichuan Province | Daocheng xian | 3·565 | 4·409 | 15·694 | 0·989 | 4·324 | 1123 | 329 | 1401 |
| Sichuan Province | Daofu xian | 2·642 | 3·317 | 11·366 | 0·751 | 3·241 | 1476 | 414 | 1799 |
| Sichuan Province | Dayi xian | 1·364 | 1·138 | 4·135 | 0·632 | 1·722 | 5845 | 2828 | 7437 |
| Sichuan Province | Dazhu xian | 1·238 | 1·087 | 3·915 | 0·551 | 1·557 | 10614 | 4697 | 13376 |
| Sichuan Province | Dechang xian | 2·755 | 3·078 | 11·070 | 0·900 | 3·406 | 5832 | 1906 | 7129 |
| Sichuan Province | Dege xian | 4·214 | 5·531 | 20·044 | 1·031 | 5·047 | 2881 | 686 | 3429 |
| Sichuan Province | Deyangshi | 1·241 | 1·087 | 3·874 | 0·558 | 1·555 | 11409 | 5236 | 14255 |
| Sichuan Province | Dujiangyanshi | 1·361 | 1·151 | 4·216 | 0·621 | 1·714 | 9375 | 4285 | 11774 |
| Sichuan Province | Erbianyi zuzizhi xian | 1·384 | 1·266 | 4·546 | 0·596 | 1·729 | 1588 | 696 | 2023 |
| Sichuan Province | Ermeishanshi | 1·312 | 1·169 | 4·187 | 0·580 | 1·637 | 5128 | 2296 | 6460 |
| Sichuan Province | Fushun xian | 1·195 | 1·091 | 3·889 | 0·517 | 1·494 | 8340 | 3619 | 10400 |
| Sichuan Province | Ganluo xian | 1·658 | 1·719 | 5·973 | 0·637 | 2·039 | 3250 | 1247 | 3896 |
| Sichuan Province | Ganzhi xian | 3·627 | 4·838 | 16·610 | 0·891 | 4·374 | 2600 | 672 | 3231 |
| Sichuan Province | Gao xian | 1·326 | 1·189 | 4·285 | 0·578 | 1·667 | 8675 | 3678 | 10913 |
| Sichuan Province | Gong xian | 1·477 | 1·361 | 4·837 | 0·628 | 1·855 | 2440 | 1048 | 3053 |
| Sichuan Province | Gongyan xian | 1·142 | 0·947 | 3·379 | 0·536 | 1·442 | 3291 | 1538 | 4154 |
| Sichuan Province | Guangan xian | 1·158 | 1·003 | 3·595 | 0·521 | 1·458 | 11576 | 5282 | 14544 |
| Sichuan Province | Guanghanshi | 1·205 | 0·993 | 3·562 | 0·577 | 1·504 | 9113 | 4405 | 11413 |
| Sichuan Province | Guanyuanshi | 1·532 | 1·651 | 5·733 | 0·546 | 1·911 | 15518 | 5456 | 19275 |
| Sichuan Province | Gulin xian | 1·873 | 1·941 | 6·836 | 0·702 | 2·332 | 12119 | 4571 | 15102 |

| ADM1 | ADM2 | β-thalassaemia carrier prevalence Mean | SD | CI95% | Q25 | Q75 | Estimated carrier counts Mean | Q25 | Q75 |
|---|---|---|---|---|---|---|---|---|---|
| Sichuan Province | Hanyuan xian | 1·657 | 1·711 | 5·964 | 0·642 | 2·042 | 4939 | 1943 | 6149 |
| Sichuan Province | Heishui xian | 1·897 | 2·176 | 7·641 | 0·610 | 2·326 | 800 | 263 | 976 |
| Sichuan Province | Hejiang xian | 1·460 | 1·479 | 5·110 | 0·582 | 1·800 | 8981 | 3649 | 11215 |
| Sichuan Province | Hongya xian | 1·389 | 1·262 | 4·530 | 0·599 | 1·739 | 4021 | 1780 | 5026 |
| Sichuan Province | Hongyuan xian | 2·393 | 3·109 | 10·506 | 0·653 | 2·903 | 1114 | 300 | 1376 |
| Sichuan Province | Huayingshi | 1·181 | 1·070 | 3·876 | 0·513 | 1·470 | 3051 | 1345 | 3779 |
| Sichuan Province | Huidong xian | 3·009 | 3·307 | 11·581 | 1·020 | 3·737 | 9344 | 3138 | 11478 |
| Sichuan Province | Huili xian | 3·234 | 3·734 | 13·060 | 1·032 | 3·996 | 14148 | 4524 | 17672 |
| Sichuan Province | Jiajiang xian | 1·265 | 1·081 | 3·852 | 0·577 | 1·607 | 3671 | 1672 | 4658 |
| Sichuan Province | Jiangan xian | 1·328 | 1·235 | 4·455 | 0·559 | 1·656 | 5663 | 2386 | 7116 |
| Sichuan Province | Jiange xian | 1·309 | 1·259 | 4·429 | 0·528 | 1·650 | 4635 | 1886 | 5859 |
| Sichuan Province | Jiangyoushi | 1·385 | 1·402 | 4·805 | 0·542 | 1·731 | 10137 | 4079 | 12616 |
| Sichuan Province | Jianwei xian | 1·163 | 0·958 | 3·520 | 0·543 | 1·467 | 4721 | 2228 | 6004 |
| Sichuan Province | Jianyangshi | 1·095 | 0·859 | 3·109 | 0·536 | 1·377 | 12149 | 5928 | 15210 |
| Sichuan Province | Jinchuan xian | 2·415 | 2·962 | 10·135 | 0·698 | 2·980 | 1233 | 367 | 1547 |
| Sichuan Province | Jinkouhequ | 1·463 | 1·396 | 4·955 | 0·611 | 1·806 | 536 | 227 | 673 |
| Sichuan Province | Jintang xian | 1·116 | 0·871 | 3·184 | 0·547 | 1·413 | 7492 | 3678 | 9486 |
| Sichuan Province | Jinyang xian | 1·984 | 1·974 | 7·074 | 0·774 | 2·477 | 3082 | 1182 | 3819 |
| Sichuan Province | Jiulong xian | 2·422 | 2·874 | 10·094 | 0·769 | 2·925 | 1306 | 408 | 1536 |
| Sichuan Province | Kaijiang xian | 1·422 | 1·307 | 4·600 | 0·612 | 1·779 | 6474 | 2809 | 8156 |
| Sichuan Province | Kangding xian | 2·210 | 2·611 | 8·809 | 0·725 | 2·693 | 2715 | 913 | 3388 |
| Sichuan Province | Langzhongshi | 1·189 | 1·047 | 3·725 | 0·519 | 1·501 | 7171 | 3173 | 9007 |
| Sichuan Province | Leibo xian | 1·520 | 1·387 | 4·949 | 0·653 | 1·909 | 4265 | 1875 | 5361 |
| Sichuan Province | Leshanshi | 1·200 | 1·016 | 3·628 | 0·556 | 1·505 | 14838 | 6942 | 18592 |
| Sichuan Province | Lezhi xian | 1·074 | 0·871 | 3·198 | 0·514 | 1·347 | 5284 | 2561 | 6625 |
| Sichuan Province | Li xian | 1·679 | 1·737 | 6·136 | 0·628 | 2·091 | 610 | 230 | 766 |
| Sichuan Province | Linshui xian | 1·239 | 1·159 | 4·117 | 0·529 | 1·536 | 8592 | 3663 | 10645 |
| Sichuan Province | Litang xian | 3·526 | 4·502 | 16·003 | 0·943 | 4·244 | 2412 | 642 | 2960 |
| Sichuan Province | Longchang xian | 1·185 | 1·075 | 3·814 | 0·511 | 1·484 | 7512 | 3262 | 9492 |
| Sichuan Province | Lu xian | 1·294 | 1·263 | 4·369 | 0·534 | 1·608 | 19371 | 8001 | 24169 |
| Sichuan Province | Luding xian | 1·982 | 2·214 | 7·625 | 0·702 | 2·412 | 1547 | 561 | 1967 |
| Sichuan Province | Luohuo xian | 3·198 | 4·297 | 14·342 | 0·804 | 3·916 | 1478 | 364 | 1795 |
| Sichuan Province | Lushan xian | 1·480 | 1·334 | 4·703 | 0·640 | 1·865 | 1423 | 613 | 1792 |
| Sichuan Province | Luzhoushi | 1·323 | 1·249 | 4·362 | 0·554 | 1·642 | 5933 | 2428 | 7436 |
| Sichuan Province | Mabianyi zuzizhi xian | 1·314 | 1·134 | 4·089 | 0·595 | 1·650 | 2414 | 1095 | 3082 |
| Sichuan Province | Maerkang xian | 2·450 | 3·079 | 10·609 | 0·684 | 2·995 | 1333 | 396 | 1631 |

| | | β-thalassaemia carrier prevalence | | | | | Estimated carrier counts | | |
|---|---|---|---|---|---|---|---|---|---|
| ADM1 | ADM2 | Mean | SD | CI95% | Q25 | Q75 | Mean | Q25 | Q75 |
| Sichuan Province | Mao xian | 1·583 | 1·629 | 5·652 | 0·596 | 1·970 | 1444 | 572 | 1792 |
| Sichuan Province | Meigu xian | 1·658 | 1·610 | 5·715 | 0·675 | 2·065 | 3961 | 1614 | 4954 |
| Sichuan Province | Meishan xian | 1·217 | 1·008 | 3·620 | 0·572 | 1·529 | 11345 | 5336 | 14113 |
| Sichuan Province | Mianning xian | 2·324 | 2·696 | 9·306 | 0·774 | 2·818 | 8863 | 2992 | 10726 |
| Sichuan Province | Mianyangshi | 1·250 | 1·181 | 4·105 | 0·532 | 1·550 | 23963 | 9972 | 29414 |
| Sichuan Province | Mianzhu xian | 1·358 | 1·241 | 4·436 | 0·579 | 1·696 | 6011 | 2614 | 7460 |
| Sichuan Province | Mingshan xian | 1·388 | 1·224 | 4·392 | 0·623 | 1·734 | 3070 | 1358 | 3857 |
| Sichuan Province | Miyi xian | 3·084 | 3·560 | 12·760 | 0·973 | 3·795 | 7131 | 2248 | 8788 |
| Sichuan Province | Muchuan xian | 1·198 | 0·996 | 3·634 | 0·561 | 1·501 | 2271 | 1083 | 2845 |
| Sichuan Province | Mulicang zuzizhi xian | 3·137 | 3·922 | 13·557 | 0·907 | 3·786 | 3754 | 1055 | 4495 |
| Sichuan Province | Nanbu xian | 1·129 | 0·970 | 3·492 | 0·514 | 1·425 | 11643 | 5340 | 14694 |
| Sichuan Province | Nanchongshi | 1·093 | 0·917 | 3·302 | 0·507 | 1·374 | 21552 | 10003 | 27154 |
| Sichuan Province | Nanjiang xian | 1·523 | 1·630 | 5·732 | 0·554 | 1·884 | 6948 | 2582 | 8566 |
| Sichuan Province | Nanping xian | 2·146 | 2·793 | 9·527 | 0·603 | 2·561 | 1376 | 395 | 1727 |
| Sichuan Province | Nanxi xian | 1·243 | 1·116 | 3·947 | 0·535 | 1·570 | 5484 | 2363 | 6913 |
| Sichuan Province | Naxi xian | 1·382 | 1·331 | 4·701 | 0·571 | 1·716 | 5795 | 2441 | 7136 |
| Sichuan Province | Neijiangshi | 1·131 | 1·006 | 3·664 | 0·494 | 1·420 | 13160 | 5738 | 16609 |
| Sichuan Province | Ningnan xian | 2·638 | 2·836 | 10·284 | 0·910 | 3·278 | 7997 | 2866 | 10046 |
| Sichuan Province | Panzhihuashi | 3·478 | 4·211 | 14·740 | 1·046 | 4·240 | 30736 | 9400 | 37680 |
| Sichuan Province | Pengan xian | 1·083 | 0·884 | 3·175 | 0·520 | 1·360 | 5005 | 2436 | 6274 |
| Sichuan Province | Pengshan xian | 1·191 | 0·930 | 3·362 | 0·591 | 1·493 | 3841 | 1900 | 4825 |
| Sichuan Province | Pengxi xian | 1·076 | 0·896 | 3·225 | 0·501 | 1·349 | 8616 | 4006 | 10820 |
| Sichuan Province | Pengzhoushi | 1·351 | 1·185 | 4·269 | 0·604 | 1·689 | 8802 | 4142 | 11103 |
| Sichuan Province | Pingchang xian | 1·279 | 1·172 | 4·156 | 0·558 | 1·594 | 7815 | 3371 | 9673 |
| Sichuan Province | Pingshan xian | 1·221 | 1·008 | 3·677 | 0·573 | 1·535 | 1388 | 656 | 1756 |
| Sichuan Province | Pingwu xian | 1·634 | 1·813 | 6·226 | 0·562 | 2·025 | 1894 | 658 | 2338 |
| Sichuan Province | Puge xian | 2·438 | 2·654 | 9·433 | 0·853 | 3·009 | 4587 | 1572 | 5691 |
| Sichuan Province | Pujiang xian | 1·318 | 1·119 | 4·026 | 0·612 | 1·644 | 3230 | 1492 | 4036 |
| Sichuan Province | Qingchuan xian | 1·623 | 1·822 | 6·195 | 0·554 | 2·011 | 1737 | 599 | 2156 |
| Sichuan Province | Qingshen xian | 1·185 | 0·993 | 3·530 | 0·552 | 1·501 | 1760 | 816 | 2204 |
| Sichuan Province | Qionglai xian | 1·345 | 1·129 | 4·105 | 0·622 | 1·694 | 8405 | 3968 | 10646 |
| Sichuan Province | Qu xian | 1·154 | 0·979 | 3·480 | 0·538 | 1·445 | 9913 | 4631 | 12378 |
| Sichuan Province | Rangtang xian | 2·969 | 3·983 | 13·438 | 0·742 | 3·587 | 1258 | 325 | 1482 |
| Sichuan Province | Renshou xian | 1·123 | 0·907 | 3·305 | 0·539 | 1·412 | 12934 | 6231 | 16235 |
| Sichuan Province | Rong xian | 1·135 | 0·956 | 3·451 | 0·519 | 1·435 | 6597 | 3000 | 8297 |
| Sichuan Province | Rongchang xian | 1·191 | 1·120 | 3·973 | 0·508 | 1·472 | 7683 | 3262 | 9530 |

| ADM1 | ADM2 | β-thalassaemia carrier prevalence Mean | SD | CI95% | Q25 | Q75 | Estimated carrier counts Mean | Q25 | Q75 |
|---|---|---|---|---|---|---|---|---|---|
| Sichuan Province | Ruoergai xian | 2·809 | 4·153 | 14·169 | 0·658 | 3·176 | 2170 | 530 | 2463 |
| Sichuan Province | Shantai xian | 1·140 | 0·967 | 3·457 | 0·527 | 1·432 | 10697 | 4938 | 13365 |
| Sichuan Province | Sheda xian | 3·404 | 4·670 | 15·806 | 0·821 | 4·088 | 2164 | 498 | 2684 |
| Sichuan Province | Shefang xian | 1·352 | 1·226 | 4·342 | 0·592 | 1·685 | 5074 | 2289 | 6289 |
| Sichuan Province | Shehong xian | 1·088 | 0·917 | 3·308 | 0·502 | 1·370 | 8022 | 3749 | 10116 |
| Sichuan Province | Shimian xian | 1·918 | 2·116 | 7·351 | 0·679 | 2·350 | 2194 | 763 | 2594 |
| Sichuan Province | Shiqu xian | 4·766 | 6·340 | 22·949 | 1·092 | 5·709 | 4996 | 1188 | 5978 |
| Sichuan Province | Shongpan xian | 1·947 | 2·329 | 8·110 | 0·605 | 2·378 | 1271 | 397 | 1564 |
| Sichuan Province | Shuifu xian | 1·244 | 1·049 | 3·731 | 0·581 | 1·555 | 335 | 156 | 416 |
| Sichuan Province | Suangliu xian | 1·154 | 0·866 | 3·198 | 0·586 | 1·447 | 29310 | 14889 | 36671 |
| Sichuan Province | Suijiang xian | 1·235 | 1·017 | 3·689 | 0·577 | 1·549 | 1841 | 863 | 2318 |
| Sichuan Province | Suiningshi | 1·073 | 0·893 | 3·211 | 0·498 | 1·352 | 13801 | 6402 | 17434 |
| Sichuan Province | Tianquan xian | 1·619 | 1·579 | 5·587 | 0·649 | 2·027 | 2156 | 875 | 2691 |
| Sichuan Province | Tongjiang xian | 1·541 | 1·612 | 5·645 | 0·585 | 1·898 | 7847 | 3125 | 9775 |
| Sichuan Province | Wangchang xian | 1·496 | 1·600 | 5·583 | 0·541 | 1·854 | 4666 | 1704 | 5756 |
| Sichuan Province | Wanyuan xian | 1·672 | 1·734 | 6·131 | 0·635 | 2·061 | 7158 | 2672 | 8569 |
| Sichuan Province | Weiyuan xian | 1·114 | 0·934 | 3·391 | 0·515 | 1·397 | 5903 | 2739 | 7432 |
| Sichuan Province | Wenchuan xian | 1·505 | 1·399 | 5·065 | 0·626 | 1·878 | 1232 | 521 | 1551 |
| Sichuan Province | Wenjaing xian | 1·246 | 0·962 | 3·604 | 0·617 | 1·572 | 10305 | 5162 | 13108 |
| Sichuan Province | Xiangcheng xian | 3·896 | 4·807 | 17·581 | 1·053 | 4·743 | 1230 | 336 | 1433 |
| Sichuan Province | Xiaojin xian | 1·875 | 1·989 | 6·994 | 0·663 | 2·350 | 1204 | 417 | 1443 |
| Sichuan Province | Xichangshi | 2·545 | 2·939 | 10·304 | 0·831 | 3·097 | 23886 | 7705 | 29360 |
| Sichuan Province | Xichong xian | 1·091 | 0·923 | 3·332 | 0·506 | 1·371 | 4463 | 2059 | 5686 |
| Sichuan Province | Xide xian | 2·191 | 2·490 | 8·622 | 0·756 | 2·662 | 3099 | 1104 | 3815 |
| Sichuan Province | Xindu xian | 1·191 | 0·897 | 3·266 | 0·605 | 1·504 | 16916 | 8585 | 21432 |
| Sichuan Province | Xingwen xian | 1·519 | 1·465 | 5·135 | 0·622 | 1·899 | 5556 | 2268 | 6954 |
| Sichuan Province | Xinjin xian | 1·215 | 0·940 | 3·427 | 0·604 | 1·535 | 4459 | 2202 | 5609 |
| Sichuan Province | Xinlong xian | 3·422 | 4·532 | 15·134 | 0·876 | 4·171 | 1108 | 283 | 1347 |
| Sichuan Province | Xuanhan xian | 1·521 | 1·478 | 5·221 | 0·622 | 1·890 | 14569 | 6069 | 18121 |
| Sichuan Province | Xuyong xian | 1·650 | 1·647 | 5·782 | 0·646 | 2·058 | 8243 | 3257 | 10478 |
| Sichuan Province | Yaanshi | 1·455 | 1·339 | 4·805 | 0·627 | 1·816 | 5690 | 2485 | 7109 |
| Sichuan Province | Yajiang xian | 2·831 | 3·556 | 12·354 | 0·825 | 3·414 | 933 | 274 | 1118 |
| Sichuan Province | Yanbian xian | 3·287 | 4·001 | 14·232 | 0·983 | 3·972 | 1407 | 414 | 1702 |
| Sichuan Province | Yanjin xian | 1·462 | 1·313 | 4·681 | 0·636 | 1·836 | 4511 | 1961 | 5688 |
| Sichuan Province | Yanting xian | 1·124 | 0·959 | 3·429 | 0·516 | 1·416 | 3822 | 1773 | 4828 |
| Sichuan Province | Yanyuan xian | 3·051 | 3·703 | 13·100 | 0·929 | 3·668 | 10587 | 3260 | 12623 |

| ADM1 | ADM2 | β-thalassaemia carrier prevalence Mean | SD | CI95% | Q25 | Q75 | Estimated carrier counts Mean | Q25 | Q75 |
|---|---|---|---|---|---|---|---|---|---|
| Sichuan Province | Yibin xian | 1·184 | 0·997 | 3·606 | 0·543 | 1·494 | 10258 | 4722 | 12953 |
| Sichuan Province | Yibinshi | 1·208 | 1·048 | 3·698 | 0·546 | 1·513 | 12753 | 5745 | 16173 |
| Sichuan Province | Yilong xian | 1·145 | 0·967 | 3·467 | 0·531 | 1·436 | 6055 | 2803 | 7585 |
| Sichuan Province | Yingshan xian | 1·106 | 0·910 | 3·264 | 0·525 | 1·390 | 6457 | 3107 | 8130 |
| Sichuan Province | Yongjing xian | 1·570 | 1·550 | 5·328 | 0·630 | 1·957 | 1999 | 803 | 2452 |
| Sichuan Province | Yongshan xian | 1·640 | 1·548 | 5·516 | 0·686 | 2·050 | 4625 | 2041 | 5919 |
| Sichuan Province | Yuechi xian | 1·135 | 0·992 | 3·604 | 0·509 | 1·419 | 8933 | 4004 | 11175 |
| Sichuan Province | Yuexi xian | 1·906 | 2·101 | 7·185 | 0·692 | 2·325 | 5857 | 2046 | 7109 |
| Sichuan Province | Zhaojue xian | 2·010 | 2·128 | 7·422 | 0·748 | 2·492 | 5142 | 1922 | 6437 |
| Sichuan Province | Zhitong xian | 1·246 | 1·152 | 4·050 | 0·531 | 1·562 | 3090 | 1332 | 3917 |
| Sichuan Province | Zhongjiang xian | 1·126 | 0·937 | 3·315 | 0·534 | 1·412 | 10279 | 4862 | 12884 |
| Sichuan Province | Zigongshi | 1·155 | 1·004 | 3·574 | 0·515 | 1·459 | 12840 | 5829 | 16264 |
| Sichuan Province | Ziyangshi | 1·077 | 0·864 | 3·141 | 0·517 | 1·358 | 9090 | 4408 | 11494 |
| Sichuan Province | Zizhong xian | 1·100 | 0·930 | 3·355 | 0·508 | 1·378 | 8991 | 4129 | 11293 |
| Tianjin Municipality | Baodi xian | 3·163 | 4·884 | 16·720 | 0·577 | 3·623 | 23680 | 4333 | 26913 |
| Tianjin Municipality | Ji xian | 3·322 | 5·163 | 17·853 | 0·592 | 3·804 | 26553 | 4724 | 30229 |
| Tianjin Municipality | Ninghe xian | 3·060 | 4·740 | 16·200 | 0·570 | 3·510 | 18762 | 3513 | 21278 |
| Tianjin Municipality | Tianjinshi | 3·025 | 4·753 | 16·002 | 0·555 | 3·432 | 289358 | 53377 | 326775 |
| Tianjin Municipality | Wuqing xian | 3·188 | 4·995 | 16·864 | 0·579 | 3·658 | 38176 | 6986 | 43779 |
| Tianjin Municipality | Xianhae xian | 3·256 | 5·044 | 17·501 | 0·582 | 3·780 | 14205 | 2537 | 16353 |
| Tianjin Municipality | Yutian xian | 3·197 | 4·934 | 17·134 | 0·579 | 3·665 | 20032 | 3597 | 23065 |
| Tibet Autonomous Region | Anduo xian | 6·976 | 9·353 | 35·063 | 1·505 | 8·353 | 2437 | 551 | 2940 |
| Tibet Autonomous Region | Angren xian | 6·776 | 8·477 | 31·357 | 1·699 | 8·294 | 3395 | 862 | 4166 |
| Tibet Autonomous Region | Bailang xian | 7·448 | 8·281 | 30·449 | 2·179 | 9·464 | 3212 | 924 | 4117 |
| Tibet Autonomous Region | Bange xian | 6·714 | 8·886 | 32·030 | 1·452 | 8·257 | 2864 | 657 | 3361 |
| Tibet Autonomous Region | Baqing xian | 6·785 | 8·461 | 31·335 | 1·609 | 8·381 | 2704 | 644 | 3452 |
| Tibet Autonomous Region | Bashu xian | 6·329 | 7·145 | 26·209 | 1·964 | 7·849 | 2603 | 802 | 3225 |
| Tibet Autonomous Region | Bianba xian | 7·176 | 8·016 | 29·592 | 2·045 | 9·129 | 2991 | 861 | 3801 |
| Tibet Autonomous Region | Biru xian | 7·114 | 8·671 | 32·031 | 1·780 | 8·885 | 3279 | 836 | 4130 |
| Tibet Autonomous Region | Bitu xian | 5·170 | 6·224 | 22·716 | 1·499 | 6·372 | 55 | 15 | 68 |
| Tibet Autonomous Region | Bomi xian | 6·945 | 7·533 | 27·972 | 2·216 | 8·731 | 2436 | 788 | 3050 |

| | | β-thalassaemia carrier prevalence | | | | | Estimated carrier counts | | |
|---|---|---|---|---|---|---|---|---|---|
| ADM1 | ADM2 | Mean | SD | CI95% | Q25 | Q75 | Mean | Q25 | Q75 |
| Tibet Autonomous Region | Changdu xian | 5·715 | 7·370 | 27·057 | 1·415 | 6·833 | 8530 | 2154 | 10177 |
| Tibet Autonomous Region | Chaya xian | 5·532 | 6·815 | 24·679 | 1·460 | 6·759 | 2892 | 755 | 3576 |
| Tibet Autonomous Region | Chayu xian | 6·155 | 6·533 | 23·975 | 2·065 | 7·733 | 1725 | 579 | 2200 |
| Tibet Autonomous Region | Cuomei xian | 8·206 | 7·987 | 30·168 | 2·934 | 10·561 | 1108 | 391 | 1441 |
| Tibet Autonomous Region | Cuona xian | 8·323 | 7·585 | 28·539 | 3·232 | 10·809 | 1268 | 484 | 1642 |
| Tibet Autonomous Region | Cuoqin xian | 5·961 | 7·996 | 29·249 | 1·340 | 7·095 | 1018 | 224 | 1198 |
| Tibet Autonomous Region | Dangxiong xian | 7·770 | 9·149 | 33·876 | 2·011 | 9·812 | 3623 | 946 | 4612 |
| Tibet Autonomous Region | Dazhi xian | 8·123 | 8·868 | 32·779 | 2·367 | 10·560 | 3085 | 914 | 3998 |
| Tibet Autonomous Region | Dingjie xian | 6·936 | 7·368 | 26·851 | 2·246 | 8·883 | 1120 | 348 | 1459 |
| Tibet Autonomous Region | Dingqing xian | 6·548 | 8·002 | 29·742 | 1·713 | 8·084 | 6026 | 1512 | 7323 |
| Tibet Autonomous Region | Dingri xian | 7·008 | 7·803 | 28·896 | 2·088 | 8·870 | 2778 | 811 | 3463 |
| Tibet Autonomous Region | Duinongdeqing xian | 7·932 | 9·044 | 33·563 | 2·193 | 10·079 | 9958 | 2884 | 12612 |
| Tibet Autonomous Region | Gaize xian | 5·483 | 7·933 | 29·066 | 1·090 | 6·296 | 1190 | 244 | 1332 |
| Tibet Autonomous Region | Gangba xian | 7·100 | 7·370 | 27·189 | 2·377 | 9·061 | 432 | 140 | 552 |
| Tibet Autonomous Region | Geer xian | 3·509 | 4·207 | 14·991 | 1·035 | 4·295 | 1166 | 315 | 1401 |
| Tibet Autonomous Region | Geji xian | 4·040 | 5·118 | 18·384 | 1·073 | 4·864 | 569 | 154 | 698 |
| Tibet Autonomous Region | Gongbujiangda xian | 8·153 | 8·584 | 32·106 | 2·551 | 10·523 | 2661 | 831 | 3406 |
| Tibet Autonomous Region | Gongga xian | 8·033 | 8·580 | 32·013 | 2·489 | 10·307 | 4163 | 1288 | 5344 |
| Tibet Autonomous Region | Gongjue xian | 4·933 | 6·270 | 22·365 | 1·273 | 6·009 | 1857 | 504 | 2305 |
| Tibet Autonomous Region | Jiahe xian | 7·730 | 8·876 | 33·282 | 2·106 | 9·779 | 3024 | 811 | 3848 |
| Tibet Autonomous Region | Jiangda xian | 4·679 | 6·098 | 22·623 | 1·168 | 5·512 | 2018 | 501 | 2374 |
| Tibet Autonomous Region | Jiangzi xian | 7·649 | 8·492 | 31·351 | 2·274 | 9·721 | 5434 | 1632 | 6911 |
| Tibet Autonomous Region | Jiazha xian | 8·564 | 8·621 | 32·201 | 2·883 | 11·078 | 2213 | 751 | 2877 |
| Tibet Autonomous Region | Jilong xian | 6·358 | 7·626 | 28·129 | 1·720 | 7·845 | 1070 | 276 | 1310 |

| | | β-thalassaemia carrier prevalence | | | | | Estimated carrier counts | | |
|---|---|---|---|---|---|---|---|---|---|
| ADM1 | ADM2 | Mean | SD | CI95% | Q25 | Q75 | Mean | Q25 | Q75 |
| Tibet Autonomous Region | Kangma xian | 7·631 | 8·102 | 29·932 | 2·432 | 9·754 | 1493 | 474 | 1958 |
| Tibet Autonomous Region | Lang xian | 8·738 | 8·320 | 31·252 | 3·199 | 11·284 | 1482 | 534 | 1891 |
| Tibet Autonomous Region | Langkazhi xian | 7·940 | 8·422 | 31·203 | 2·512 | 10·181 | 2496 | 773 | 3186 |
| Tibet Autonomous Region | Lashashi | 7·999 | 8·778 | 31·974 | 2·315 | 10·329 | 41338 | 11949 | 53467 |
| Tibet Autonomous Region | Lazhi xian | 7·084 | 8·197 | 30·136 | 2·012 | 8·844 | 3851 | 1085 | 4788 |
| Tibet Autonomous Region | Leiwuqi xian | 6·104 | 7·747 | 28·585 | 1·546 | 7·406 | 3595 | 942 | 4446 |
| Tibet Autonomous Region | Linzhi xian | 7·858 | 7·824 | 29·092 | 2·674 | 10·220 | 6661 | 2258 | 8716 |
| Tibet Autonomous Region | Linzhou xian | 7·989 | 9·088 | 33·618 | 2·183 | 10·210 | 3476 | 976 | 4474 |
| Tibet Autonomous Region | Longgeer xian | 5·411 | 7·329 | 26·455 | 1·213 | 6·484 | 116 | 24 | 137 |
| Tibet Autonomous Region | Longzhi xian | 8·644 | 7·998 | 30·029 | 3·296 | 11·173 | 2778 | 1043 | 3577 |
| Tibet Autonomous Region | Luolong xian | 6·590 | 7·733 | 28·309 | 1·867 | 8·142 | 3429 | 1016 | 4310 |
| Tibet Autonomous Region | Luozha xian | 7·961 | 7·891 | 29·425 | 2·748 | 10·288 | 1400 | 473 | 1823 |
| Tibet Autonomous Region | Manggang xian | 4·975 | 6·194 | 22·615 | 1·329 | 6·042 | 3081 | 868 | 3713 |
| Tibet Autonomous Region | Milin xian | 8·240 | 7·798 | 29·163 | 3·022 | 10·697 | 2228 | 828 | 2914 |
| Tibet Autonomous Region | Motuo xian | 7·446 | 7·186 | 26·529 | 2·779 | 9·524 | 1128 | 415 | 1420 |
| Tibet Autonomous Region | Mozhugongka xian | 8·182 | 8·877 | 32·843 | 2·424 | 10·521 | 4127 | 1197 | 5264 |
| Tibet Autonomous Region | Naidong xian | 8·265 | 8·364 | 31·262 | 2·727 | 10·737 | 6903 | 2163 | 8912 |
| Tibet Autonomous Region | Nanmulin xian | 7·395 | 8·498 | 31·495 | 2·002 | 9·424 | 6081 | 1638 | 7658 |
| Tibet Autonomous Region | Naqu xian | 7·557 | 9·309 | 34·609 | 1·807 | 9·385 | 9249 | 2206 | 11662 |
| Tibet Autonomous Region | Nielamu xian | 6·865 | 8·026 | 29·713 | 1·875 | 8·596 | 870 | 234 | 1095 |
| Tibet Autonomous Region | Nierong xian | 7·070 | 9·125 | 33·764 | 1·565 | 8·684 | 2566 | 556 | 3113 |
| Tibet Autonomous Region | Nima xian | 6·270 | 8·852 | 32·490 | 1·206 | 7·363 | 2057 | 376 | 2486 |
| Tibet Autonomous Region | Nimu xian | 7·732 | 8·893 | 33·206 | 2·169 | 9·742 | 2303 | 653 | 2873 |
| Tibet Autonomous Region | Pulan xian | 3·734 | 4·364 | 15·515 | 1·163 | 4·565 | 442 | 131 | 550 |

| | | β-thalassaemia carrier prevalence | | | | | Estimated carrier counts | | |
|---|---|---|---|---|---|---|---|---|---|
| **ADM1** | **ADM2** | **Mean** | **SD** | **CI95%** | **Q25** | **Q75** | **Mean** | **Q25** | **Q75** |
| Tibet Autonomous Region | Qiongjie xian | 8·181 | 8·211 | 30·947 | 2·773 | 10·480 | 1196 | 414 | 1535 |
| Tibet Autonomous Region | Qushong xian | 8·472 | 8·319 | 31·113 | 2·957 | 10·986 | 1061 | 361 | 1383 |
| Tibet Autonomous Region | Qushui xian | 7·933 | 8·684 | 31·927 | 2·359 | 10·200 | 2920 | 879 | 3765 |
| Tibet Autonomous Region | Renbu xian | 7·735 | 8·670 | 32·449 | 2·257 | 9·767 | 2623 | 792 | 3400 |
| Tibet Autonomous Region | Rikezeshi | 7·343 | 8·271 | 30·492 | 2·065 | 9·405 | 12780 | 3546 | 16078 |
| Tibet Autonomous Region | Ritu xian | 4·024 | 5·305 | 18·739 | 0·957 | 4·877 | 320 | 71 | 394 |
| Tibet Autonomous Region | Saxun xian | 7·196 | 8·087 | 30·040 | 2·083 | 9·090 | 3388 | 987 | 4333 |
| Tibet Autonomous Region | Shaga xian | 6·102 | 7·715 | 27·926 | 1·549 | 7·426 | 777 | 193 | 940 |
| Tibet Autonomous Region | Shangri xian | 8·327 | 8·356 | 31·125 | 2·786 | 10·785 | 1460 | 492 | 1927 |
| Tibet Autonomous Region | Shengda xian | 5·188 | 6·839 | 25·022 | 1·250 | 6·147 | 387 | 90 | 458 |
| Tibet Autonomous Region | Shenzha xian | 6·973 | 8·915 | 32·954 | 1·589 | 8·667 | 1478 | 327 | 1835 |
| Tibet Autonomous Region | Suo xian | 6·801 | 8·235 | 30·594 | 1·721 | 8·478 | 1332 | 334 | 1673 |
| Tibet Autonomous Region | Tuoba xian | 5·468 | 7·093 | 25·906 | 1·354 | 6·497 | 1066 | 266 | 1227 |
| Tibet Autonomous Region | Xietongmen xian | 7·169 | 8·525 | 31·438 | 1·857 | 8·975 | 2500 | 658 | 3076 |
| Tibet Autonomous Region | Yadong xian | 7·071 | 6·772 | 25·164 | 2·647 | 8·989 | 1096 | 417 | 1375 |
| Tibet Autonomous Region | Yanjing xian | 4·819 | 5·933 | 21·835 | 1·319 | 5·869 | 786 | 220 | 958 |
| Tibet Autonomous Region | Zhada xian | 3·210 | 3·493 | 12·344 | 1·112 | 3·978 | 255 | 77 | 315 |
| Tibet Autonomous Region | Zhaxiang xian | 8·178 | 8·568 | 31·980 | 2·561 | 10·549 | 3037 | 957 | 3939 |
| Tibet Autonomous Region | Zhongba xian | 5·124 | 6·550 | 23·237 | 1·364 | 6·150 | 1231 | 310 | 1437 |
| Tibet Autonomous Region | Zuogong xian | 5·907 | 6·887 | 25·098 | 1·722 | 7·351 | 2602 | 746 | 3229 |
| Xinjiang Uyghur Autonomous Region | Aheqi xian | 5·407 | 8·331 | 29·573 | 0·998 | 6·007 | 2416 | 436 | 2665 |
| Xinjiang Uyghur Autonomous Region | Akeshushi | 5·339 | 8·122 | 27·597 | 0·989 | 6·158 | 47790 | 9103 | 55491 |
| Xinjiang Uyghur Autonomous Region | Aletaishi | 5·003 | 7·619 | 26·411 | 0·887 | 5·779 | 12085 | 2138 | 13909 |

| ADM1 | ADM2 | β-thalassaemia carrier prevalence Mean | SD | CI95% | Q25 | Q75 | Estimated carrier counts Mean | Q25 | Q75 |
|---|---|---|---|---|---|---|---|---|---|
| Xinjiang Uyghur Autonomous Region | Anketao xian | 5·506 | 8·190 | 29·841 | 1·016 | 6·272 | 10403 | 1947 | 11912 |
| Xinjiang Uyghur Autonomous Region | Atushenshi | 5·490 | 8·504 | 30·615 | 1·013 | 6·046 | 18556 | 3393 | 20903 |
| Xinjiang Uyghur Autonomous Region | Awati xian | 5·342 | 8·094 | 27·937 | 1·004 | 6·154 | 14197 | 2664 | 16612 |
| Xinjiang Uyghur Autonomous Region | Bachu xian | 5·520 | 8·550 | 30·471 | 1·008 | 6·088 | 33613 | 6094 | 37373 |
| Xinjiang Uyghur Autonomous Region | Baicheng xian | 5·273 | 8·241 | 28·346 | 0·989 | 5·903 | 12335 | 2322 | 13550 |
| Xinjiang Uyghur Autonomous Region | Balikunhashakezizhi xian | 5·240 | 8·345 | 28·842 | 0·901 | 5·833 | 3499 | 574 | 3891 |
| Xinjiang Uyghur Autonomous Region | Bohu xian | 5·347 | 8·021 | 28·743 | 0·956 | 6·119 | 2314 | 426 | 2584 |
| Xinjiang Uyghur Autonomous Region | Boleshi | 5·358 | 8·224 | 29·163 | 0·959 | 6·075 | 17009 | 2981 | 19113 |
| Xinjiang Uyghur Autonomous Region | Buerjin xian | 5·144 | 7·772 | 27·666 | 0·895 | 5·873 | 3745 | 642 | 4293 |
| Xinjiang Uyghur Autonomous Region | Cele xian | 5·234 | 7·918 | 28·261 | 0·959 | 5·971 | 8501 | 1548 | 9723 |
| Xinjiang Uyghur Autonomous Region | Chabuchaerxibo zuzizhi xian | 5·188 | 7·898 | 27·121 | 0·981 | 5·900 | 25386 | 4902 | 29005 |
| Xinjiang Uyghur Autonomous Region | Changjishi | 5·278 | 8·112 | 29·866 | 0·914 | 5·949 | 36360 | 6197 | 41224 |
| Xinjiang Uyghur Autonomous Region | Ermin xian | 5·288 | 8·454 | 29·856 | 0·886 | 5·829 | 9904 | 1726 | 11094 |
| Xinjiang Uyghur Autonomous Region | Fuhai xian | 5·070 | 7·684 | 27·183 | 0·885 | 5·845 | 3889 | 671 | 4512 |
| Xinjiang Uyghur Autonomous Region | Fukangshi | 5·283 | 8·254 | 29·867 | 0·933 | 5·835 | 9405 | 1637 | 10359 |
| Xinjiang Uyghur Autonomous Region | Fuyun xian | 4·963 | 7·450 | 26·332 | 0·910 | 5·706 | 4942 | 886 | 5701 |
| Xinjiang Uyghur Autonomous Region | Gongliu xian | 5·272 | 8·163 | 28·257 | 0·961 | 5·945 | 10072 | 1849 | 11372 |
| Xinjiang Uyghur Autonomous Region | Habahe xian | 5·235 | 7·982 | 28·810 | 0·899 | 5·883 | 4290 | 719 | 5048 |

| | | β-thalassaemia carrier prevalence | | | | | Estimated carrier counts | | |
|---|---|---|---|---|---|---|---|---|---|
| ADM1 | ADM2 | Mean | SD | CI95% | Q25 | Q75 | Mean | Q25 | Q75 |
| Xinjiang Uyghur Autonomous Region | Hamishi | 5·075 | 8·253 | 27·670 | 0·888 | 5·637 | 26139 | 4483 | 29176 |
| Xinjiang Uyghur Autonomous Region | Hebukesaiermengguzizhi xian | 5·268 | 8·362 | 30·208 | 0·893 | 5·761 | 3352 | 545 | 3632 |
| Xinjiang Uyghur Autonomous Region | Hejing xian | 5·241 | 8·167 | 28·357 | 0·935 | 5·950 | 10204 | 1811 | 11895 |
| Xinjiang Uyghur Autonomous Region | Heshuo xian | 5·313 | 8·054 | 28·811 | 0·935 | 6·075 | 3568 | 625 | 4117 |
| Xinjiang Uyghur Autonomous Region | Hetian xian | 5·336 | 8·195 | 28·885 | 0·962 | 6·073 | 20078 | 3581 | 22577 |
| Xinjiang Uyghur Autonomous Region | Hetianshi | 5·501 | 8·422 | 28·986 | 1·019 | 6·305 | 20110 | 3638 | 22773 |
| Xinjiang Uyghur Autonomous Region | Huocheng xian | 5·198 | 7·862 | 27·386 | 1·002 | 5·896 | 18513 | 3572 | 21054 |
| Xinjiang Uyghur Autonomous Region | Hutubi xian | 5·275 | 8·161 | 29·728 | 0·908 | 5·942 | 10303 | 1735 | 11619 |
| Xinjiang Uyghur Autonomous Region | Jiashi xian | 5·520 | 8·625 | 31·099 | 1·024 | 6·016 | 23392 | 4313 | 26141 |
| Xinjiang Uyghur Autonomous Region | Jimunai xian | 5·204 | 7·969 | 28·475 | 0·878 | 5·834 | 1694 | 266 | 1885 |
| Xinjiang Uyghur Autonomous Region | Jimushaer xian | 5·224 | 8·162 | 29·127 | 0·929 | 5·800 | 7977 | 1374 | 8599 |
| Xinjiang Uyghur Autonomous Region | Jinghe xian | 5·414 | 8·525 | 29·787 | 0·936 | 6·071 | 6669 | 1130 | 7516 |
| Xinjiang Uyghur Autonomous Region | Kelamayishi | 5·386 | 8·784 | 31·286 | 0·909 | 5·860 | 16059 | 2620 | 17812 |
| Xinjiang Uyghur Autonomous Region | Keping xian | 5·371 | 8·320 | 28·706 | 0·996 | 6·027 | 2666 | 483 | 2989 |
| Xinjiang Uyghur Autonomous Region | Keshishi | 5·550 | 8·587 | 30·918 | 1·019 | 6·087 | 43758 | 8140 | 48620 |
| Xinjiang Uyghur Autonomous Region | Kuche xian | 5·396 | 8·586 | 29·358 | 0·971 | 6·025 | 30516 | 5498 | 34610 |
| Xinjiang Uyghur Autonomous Region | Kuerleshi | 5·307 | 8·147 | 28·074 | 0·942 | 6·092 | 42458 | 7541 | 48103 |
| Xinjiang Uyghur Autonomous Region | Kuitunshi | 5·302 | 8·475 | 29·829 | 0·888 | 5·900 | 16161 | 2696 | 17636 |

| | | β-thalassaemia carrier prevalence | | | | | Estimated carrier counts | | |
|---|---|---|---|---|---|---|---|---|---|
| **ADM1** | **ADM2** | **Mean** | **SD** | **CI95%** | **Q25** | **Q75** | **Mean** | **Q25** | **Q75** |
| Xinjiang Uyghur Autonomous Region | Luntai xian | 5·336 | 8·460 | 28·953 | 0·957 | 5·967 | 6820 | 1249 | 7528 |
| Xinjiang Uyghur Autonomous Region | Luopu xian | 5·387 | 8·273 | 29·077 | 0·964 | 6·155 | 17748 | 3177 | 20285 |
| Xinjiang Uyghur Autonomous Region | Maigaiti xian | 5·504 | 8·665 | 30·718 | 0·991 | 6·146 | 12738 | 2298 | 14416 |
| Xinjiang Uyghur Autonomous Region | Manashi xian | 5·277 | 8·277 | 29·982 | 0·905 | 5·880 | 9247 | 1551 | 10249 |
| Xinjiang Uyghur Autonomous Region | Minfeng xian | 5·408 | 8·012 | 27·784 | 0·981 | 6·380 | 2308 | 415 | 2702 |
| Xinjiang Uyghur Autonomous Region | Miquanshi | 5·284 | 8·143 | 30·091 | 0·924 | 5·911 | 24599 | 4298 | 27347 |
| Xinjiang Uyghur Autonomous Region | Moyu xian | 5·387 | 8·099 | 28·854 | 0·992 | 6·158 | 35159 | 6268 | 39052 |
| Xinjiang Uyghur Autonomous Region | Muleihashakezizhi xian | 5·210 | 8·175 | 28·930 | 0·893 | 5·762 | 3536 | 600 | 3803 |
| Xinjiang Uyghur Autonomous Region | Nileke xian | 5·340 | 8·471 | 29·184 | 0·944 | 5·968 | 8116 | 1420 | 9262 |
| Xinjiang Uyghur Autonomous Region | Pishan xian | 5·348 | 8·165 | 29·418 | 0·980 | 6·020 | 18374 | 3470 | 20653 |
| Xinjiang Uyghur Autonomous Region | Qiemo xian | 5·447 | 8·255 | 28·842 | 0·977 | 6·296 | 3789 | 619 | 4325 |
| Xinjiang Uyghur Autonomous Region | Qinghe xian | 5·030 | 7·538 | 27·349 | 0·931 | 5·712 | 3058 | 574 | 3651 |
| Xinjiang Uyghur Autonomous Region | Qishan xian | 5·212 | 8·107 | 28·800 | 0·917 | 5·816 | 12629 | 2250 | 13989 |
| Xinjiang Uyghur Autonomous Region | Qitai xian | 5·165 | 8·045 | 28·844 | 0·907 | 5·738 | 11416 | 1974 | 12510 |
| Xinjiang Uyghur Autonomous Region | Ruoqiang xian | 5·818 | 8·800 | 31·262 | 1·005 | 6·730 | 2933 | 481 | 3446 |
| Xinjiang Uyghur Autonomous Region | Shache xian | 5·452 | 8·341 | 29·815 | 0·977 | 6·249 | 48818 | 8380 | 56382 |
| Xinjiang Uyghur Autonomous Region | Shawan xian | 5·274 | 8·308 | 29·627 | 0·893 | 5·916 | 19342 | 3237 | 21656 |
| Xinjiang Uyghur Autonomous Region | Shaya xian | 5·363 | 8·273 | 28·379 | 0·949 | 6·152 | 14401 | 2535 | 16510 |

| | | β-thalassaemia carrier prevalence | | | | | Estimated carrier counts | | |
|---|---|---|---|---|---|---|---|---|---|
| ADM1 | ADM2 | Mean | SD | CI95% | Q25 | Q75 | Mean | Q25 | Q75 |
| Xinjiang Uyghur Autonomous Region | Shihezhishi | 5·223 | 8·064 | 28·780 | 0·890 | 5·893 | 23726 | 3927 | 27436 |
| Xinjiang Uyghur Autonomous Region | Shufu xian | 5·487 | 8·250 | 30·138 | 1·027 | 6·151 | 15458 | 2891 | 17236 |
| Xinjiang Uyghur Autonomous Region | Shule xian | 5·538 | 8·480 | 31·070 | 1·003 | 6·193 | 21702 | 3987 | 24410 |
| Xinjiang Uyghur Autonomous Region | Tachengshi | 5·236 | 8·444 | 29·311 | 0·892 | 5·764 | 8523 | 1435 | 9479 |
| Xinjiang Uyghur Autonomous Region | Tashenkuergantajike | 5·509 | 7·970 | 28·152 | 1·040 | 6·518 | 2158 | 393 | 2600 |
| Xinjiang Uyghur Autonomous Region | Tekeshi xian | 5·137 | 7·830 | 27·151 | 0·964 | 5·816 | 7718 | 1445 | 8756 |
| Xinjiang Uyghur Autonomous Region | Tulufanshi | 5·254 | 8·040 | 28·797 | 0·937 | 5·941 | 17021 | 3016 | 19387 |
| Xinjiang Uyghur Autonomous Region | Tuokexun xian | 5·315 | 8·121 | 29·309 | 0·945 | 5·996 | 6997 | 1278 | 7982 |
| Xinjiang Uyghur Autonomous Region | Tuoli xian | 5·415 | 8·809 | 31·028 | 0·892 | 5·924 | 11559 | 1921 | 12503 |
| Xinjiang Uyghur Autonomous Region | Weili xian | 5·421 | 8·193 | 28·802 | 0·981 | 6·255 | 6130 | 1090 | 7024 |
| Xinjiang Uyghur Autonomous Region | Wenquan xian | 5·248 | 7·939 | 28·429 | 0·980 | 5·988 | 2774 | 512 | 3146 |
| Xinjiang Uyghur Autonomous Region | Wenshu xian | 5·155 | 7·838 | 26·562 | 1·003 | 5·907 | 19304 | 3772 | 22418 |
| Xinjiang Uyghur Autonomous Region | Wulumuqi xian | 5·264 | 8·088 | 29·188 | 0·927 | 5·959 | 193031 | 33109 | 221129 |
| Xinjiang Uyghur Autonomous Region | Wulumuqishi | 5·357 | 8·194 | 28·992 | 0·922 | 6·165 | 3046 | 499 | 3492 |
| Xinjiang Uyghur Autonomous Region | Wuqia xian | 5·508 | 8·475 | 30·056 | 1·019 | 6·161 | 3364 | 625 | 3827 |
| Xinjiang Uyghur Autonomous Region | Wushen xian | 5·215 | 7·955 | 27·322 | 0·997 | 5·937 | 11548 | 2210 | 12967 |
| Xinjiang Uyghur Autonomous Region | Wushu xian | 5·379 | 8·676 | 30·515 | 0·894 | 5·934 | 18219 | 3030 | 20088 |
| Xinjiang Uyghur Autonomous Region | Xinhe xian | 5·407 | 8·487 | 28·859 | 0·971 | 6·098 | 10383 | 1837 | 11593 |

| ADM1 | ADM2 | β-thalassaemia carrier prevalence Mean | SD | CI95% | Q25 | Q75 | Estimated carrier counts Mean | Q25 | Q75 |
|---|---|---|---|---|---|---|---|---|---|
| Xinjiang Uyghur Autonomous Region | Xinyuan xian | 5·325 | 8·499 | 29·637 | 0·940 | 5·875 | 16686 | 2959 | 18619 |
| Xinjiang Uyghur Autonomous Region | Yanqihui zuzizhi xian | 5·279 | 8·050 | 28·353 | 0·934 | 6·030 | 7979 | 1440 | 9259 |
| Xinjiang Uyghur Autonomous Region | Yecheng xian | 5·370 | 8·162 | 29·338 | 0·973 | 6·111 | 28957 | 5051 | 32254 |
| Xinjiang Uyghur Autonomous Region | Yingjisha xian | 5·462 | 8·121 | 29·291 | 1·011 | 6·303 | 15339 | 2800 | 17590 |
| Xinjiang Uyghur Autonomous Region | Yining xian | 5·277 | 8·149 | 28·039 | 0·977 | 5·960 | 27357 | 5080 | 31041 |
| Xinjiang Uyghur Autonomous Region | Yiningshi | 5·254 | 8·017 | 28·568 | 0·978 | 5·774 | 15511 | 2985 | 16943 |
| Xinjiang Uyghur Autonomous Region | Yiwu xian | 5·090 | 8·342 | 27·624 | 0·890 | 5·649 | 1953 | 331 | 2169 |
| Xinjiang Uyghur Autonomous Region | Yuepuhu xian | 5·508 | 8·530 | 30·404 | 1·006 | 6·194 | 8041 | 1431 | 8838 |
| Xinjiang Uyghur Autonomous Region | Yumin xian | 5·270 | 8·436 | 29·425 | 0·894 | 5·823 | 2629 | 439 | 2962 |
| Xinjiang Uyghur Autonomous Region | Yutian xian | 5·317 | 8·001 | 28·282 | 0·971 | 6·134 | 13892 | 2539 | 15962 |
| Xinjiang Uyghur Autonomous Region | Zepu xian | 5·463 | 8·363 | 30·210 | 0·960 | 6·186 | 11436 | 2019 | 12801 |
| Xinjiang Uyghur Autonomous Region | Zhaoshu xian | 5·152 | 7·857 | 27·566 | 0·979 | 5·809 | 7522 | 1406 | 8443 |
| Yunnan Province | Anning xian | 4·379 | 5·175 | 18·477 | 1·289 | 5·432 | 20560 | 6153 | 25336 |
| Yunnan Province | Baoshanshi | 4·105 | 5·472 | 18·959 | 1·028 | 4·939 | 37815 | 9598 | 46002 |
| Yunnan Province | Binchuan xian | 4·017 | 5·235 | 17·799 | 1·075 | 4·846 | 14058 | 3803 | 16900 |
| Yunnan Province | Changning xian | 4·181 | 5·721 | 19·636 | 1·016 | 4·946 | 14035 | 3336 | 16658 |
| Yunnan Province | Changyuanwa zuzizhi xian | 4·146 | 5·525 | 19·628 | 0·932 | 5·058 | 6869 | 1505 | 8038 |
| Yunnan Province | Chenggong xian | 4·351 | 4·905 | 17·196 | 1·381 | 5·428 | 39623 | 12828 | 48765 |
| Yunnan Province | Chengjiang xian | 4·393 | 4·824 | 17·377 | 1·449 | 5·509 | 7600 | 2474 | 9481 |
| Yunnan Province | Chuxiongshi | 4·292 | 5·459 | 19·158 | 1·141 | 5·231 | 27679 | 7592 | 33805 |
| Yunnan Province | Dalishi | 4·064 | 5·314 | 18·123 | 1·074 | 4·938 | 26399 | 7035 | 32248 |
| Yunnan Province | Dayao xian | 3·927 | 4·965 | 17·165 | 1·075 | 4·785 | 8952 | 2386 | 10676 |
| Yunnan Province | Deqin xian | 4·401 | 5·419 | 19·323 | 1·229 | 5·366 | 2557 | 715 | 3107 |
| Yunnan Province | Dongchuanshi | 3·240 | 3·496 | 12·469 | 1·138 | 4·024 | 8193 | 2869 | 10214 |

| | | β-thalassaemia carrier prevalence | | | | | Estimated carrier counts | | |
|---|---|---|---|---|---|---|---|---|---|
| ADM1 | ADM2 | Mean | SD | CI95% | Q25 | Q75 | Mean | Q25 | Q75 |
| Yunnan Province | Ershanyi zuzizhi xian | 4·765 | 5·870 | 20·601 | 1·311 | 5·873 | 7309 | 1979 | 9098 |
| Yunnan Province | Eryuan xian | 4·115 | 5·437 | 18·425 | 1·069 | 4·969 | 11547 | 3008 | 13897 |
| Yunnan Province | Fengqing xian | 4·254 | 5·862 | 20·395 | 1·017 | 5·020 | 15744 | 3901 | 18596 |
| Yunnan Province | Fugong xian | 4·390 | 5·398 | 18·808 | 1·235 | 5·398 | 5074 | 1447 | 6281 |
| Yunnan Province | Fumin xian | 4·012 | 4·581 | 16·061 | 1·276 | 4·990 | 6305 | 2018 | 7908 |
| Yunnan Province | Funing xian | 6·217 | 4·703 | 17·546 | 3·011 | 7·973 | 25436 | 12148 | 32414 |
| Yunnan Province | Fuyuan xian | 3·871 | 3·417 | 12·564 | 1·680 | 4·862 | 25908 | 11271 | 32748 |
| Yunnan Province | Gejiushi | 5·331 | 6·153 | 21·849 | 1·595 | 6·659 | 25279 | 7652 | 31751 |
| Yunnan Province | Gengmadaizhuwa zuzizhi xian | 4·184 | 5·633 | 20·281 | 0·955 | 5·004 | 11510 | 2684 | 13874 |
| Yunnan Province | Gongshandulongzunu zuzizhi xian | 4·965 | 5·739 | 20·752 | 1·511 | 6·109 | 1909 | 564 | 2330 |
| Yunnan Province | Guangnan xian | 5·538 | 4·552 | 16·902 | 2·491 | 7·089 | 43535 | 19366 | 56486 |
| Yunnan Province | Hekouyao zuzizhi xian | 5·636 | 6·315 | 22·587 | 1·731 | 7·060 | 4134 | 1257 | 5173 |
| Yunnan Province | Heqing xian | 3·955 | 5·156 | 17·148 | 1·088 | 4·816 | 9618 | 2637 | 11750 |
| Yunnan Province | Honghe xian | 4·975 | 6·209 | 22·175 | 1·309 | 6·123 | 14232 | 3871 | 17851 |
| Yunnan Province | Huaning xian | 4·745 | 5·319 | 18·848 | 1·517 | 5·940 | 8551 | 2635 | 10613 |
| Yunnan Province | Huaping xian | 3·618 | 4·550 | 15·878 | 1·024 | 4·391 | 6392 | 1770 | 7647 |
| Yunnan Province | Huizhe xian | 2·932 | 3·052 | 10·928 | 1·075 | 3·647 | 23705 | 8513 | 29467 |
| Yunnan Province | Jianchuan xian | 4·128 | 5·413 | 18·308 | 1·086 | 5·002 | 6582 | 1770 | 8032 |
| Yunnan Province | Jiangchenghanizhuyi zuzizhi xian | 4·784 | 6·150 | 21·787 | 1·213 | 5·802 | 5068 | 1313 | 6211 |
| Yunnan Province | Jiangchuan xian | 4·726 | 5·539 | 19·612 | 1·414 | 5·813 | 10853 | 3232 | 13383 |
| Yunnan Province | Jianshan xian | 5·465 | 5·278 | 19·269 | 2·060 | 6·981 | 26418 | 9953 | 33601 |
| Yunnan Province | Jianshui xian | 5·096 | 5·944 | 21·123 | 1·503 | 6·351 | 28420 | 8228 | 35122 |
| Yunnan Province | Jingdongyi zuzizhi xian | 4·429 | 5·936 | 20·608 | 1·087 | 5·323 | 13803 | 3471 | 16702 |
| Yunnan Province | Jinggudaizhuyi zuzizhi xian | 4·396 | 5·752 | 20·299 | 1·021 | 5·420 | 12059 | 2835 | 14894 |
| Yunnan Province | Jinghongshi | 4·566 | 5·950 | 21·302 | 1·081 | 5·590 | 30152 | 7200 | 36579 |
| Yunnan Province | Jinning xian | 4·502 | 5·244 | 18·813 | 1·364 | 5·593 | 15990 | 4943 | 19658 |
| Yunnan Province | Jinpingmiaozhuyaozhudai zuzizhi xian | 5·409 | 6·458 | 23·306 | 1·506 | 6·720 | 18428 | 5223 | 23084 |
| Yunnan Province | Junlian xian | 1·517 | 1·386 | 5·013 | 0·642 | 1·915 | 3543 | 1483 | 4498 |
| Yunnan Province | Kaiyuanshi | 5·177 | 5·681 | 20·449 | 1·690 | 6·500 | 15702 | 5051 | 19977 |
| Yunnan Province | Kunmingshi | 4·149 | 4·656 | 16·816 | 1·323 | 5·151 | 187987 | 59053 | 234953 |
| Yunnan Province | Lanchanglagu zuzizhi xian | 4·272 | 5·549 | 19·742 | 0·984 | 5·265 | 19093 | 4397 | 23318 |
| Yunnan Province | Lanpingbazupumi zuzizhi xian | 4·140 | 5·205 | 17·869 | 1·135 | 5·084 | 8074 | 2205 | 9900 |
| Yunnan Province | Lianghe xian | 3·849 | 5·072 | 17·780 | 0·941 | 4·658 | 5382 | 1326 | 6381 |
| Yunnan Province | Lijiangnaxi zuzizhi xian | 3·920 | 4·971 | 17·082 | 1·102 | 4·750 | 20102 | 5563 | 24568 |
| Yunnan Province | Linchang xian | 4·314 | 5·746 | 20·477 | 1·005 | 5·200 | 15607 | 3649 | 18985 |
| Yunnan Province | Longchuan xian | 3·600 | 4·664 | 16·349 | 0·894 | 4·369 | 4487 | 1133 | 5409 |

| ADM1 | ADM2 | β-thalassaemia carrier prevalence Mean | SD | CI95% | Q25 | Q75 | Estimated carrier counts Mean | Q25 | Q75 |
|---|---|---|---|---|---|---|---|---|---|
| Yunnan Province | Longling xian | 4·004 | 5·470 | 18·795 | 0·951 | 4·804 | 7979 | 1909 | 9524 |
| Yunnan Province | Luchun xian | 4·969 | 6·213 | 22·145 | 1·291 | 6·089 | 10488 | 2770 | 12879 |
| Yunnan Province | Ludingshi | 2·316 | 2·315 | 8·471 | 0·872 | 2·910 | 8244 | 3144 | 10400 |
| Yunnan Province | Lufeng xian | 4·118 | 4·839 | 17·148 | 1·221 | 5·138 | 14660 | 4340 | 18170 |
| Yunnan Province | Luliang xian | 4·160 | 4·039 | 14·848 | 1·600 | 5·254 | 23614 | 9114 | 29772 |
| Yunnan Province | Lunanyi zuzizhi xian | 4·373 | 4·405 | 16·031 | 1·594 | 5·519 | 6026 | 2147 | 7640 |
| Yunnan Province | Luoping xian | 4·287 | 3·677 | 13·436 | 1·893 | 5·430 | 23671 | 10581 | 30282 |
| Yunnan Province | Luquanyizumiao zuzizhi xian | 3·553 | 4·020 | 14·052 | 1·161 | 4·412 | 9672 | 3135 | 12001 |
| Yunnan Province | Lushui xian | 4·159 | 5·310 | 18·351 | 1·096 | 5·096 | 7631 | 2000 | 9262 |
| Yunnan Province | Luxi xian | 4·604 | 4·478 | 16·297 | 1·765 | 5·787 | 19593 | 7473 | 24668 |
| Yunnan Province | Luxi xian | 3·820 | 5·110 | 17·836 | 0·924 | 4·577 | 19052 | 4733 | 23134 |
| Yunnan Province | Maguan xian | 5·817 | 6·080 | 22·270 | 1·947 | 7·397 | 18349 | 6218 | 23790 |
| Yunnan Province | Malipo xian | 6·054 | 5·662 | 21·039 | 2·351 | 7·739 | 16306 | 6275 | 20809 |
| Yunnan Province | Malong xian | 3·892 | 3·976 | 14·197 | 1·460 | 4·854 | 7544 | 2842 | 9324 |
| Yunnan Province | Menghai xian | 4·447 | 5·770 | 20·603 | 1·031 | 5·463 | 15158 | 3514 | 18537 |
| Yunnan Province | Mengla xian | 4·656 | 6·128 | 21·887 | 1·129 | 5·627 | 13399 | 3249 | 16188 |
| Yunnan Province | Mengliandaizhulaguzhuva zuzizhi xian | 4·181 | 5·460 | 19·388 | 0·978 | 5·120 | 9231 | 2078 | 11330 |
| Yunnan Province | Mengzi xian | 5·417 | 5·988 | 21·528 | 1·701 | 6·843 | 28454 | 8703 | 35471 |
| Yunnan Province | Midu xian | 4·150 | 5·391 | 18·766 | 1·079 | 5·009 | 10892 | 2813 | 13040 |
| Yunnan Province | Mile xian | 4·825 | 5·122 | 18·678 | 1·664 | 6·046 | 26713 | 9214 | 33112 |
| Yunnan Province | Mojianghani zuzizhi xian | 4·744 | 6·161 | 21·854 | 1·169 | 5·776 | 13789 | 3312 | 16355 |
| Yunnan Province | Mouding xian | 4·028 | 4·862 | 17·148 | 1·153 | 4·988 | 5949 | 1643 | 7333 |
| Yunnan Province | Nanhua xian | 4·200 | 5·389 | 18·985 | 1·101 | 5·086 | 8502 | 2272 | 10227 |
| Yunnan Province | Nanjianyi zuzizhi xian | 4·221 | 5·653 | 19·513 | 1·067 | 5·054 | 7754 | 1968 | 9258 |
| Yunnan Province | Naxishulishu zuzizhi xian | 4·338 | 5·236 | 18·321 | 1·259 | 5·359 | 5959 | 1759 | 7491 |
| Yunnan Province | Ninglangyi zuzizhi xian | 3·579 | 4·567 | 15·813 | 1·019 | 4·289 | 9084 | 2541 | 10776 |
| Yunnan Province | Pingbianmiao zuzizhi xian | 5·552 | 6·067 | 21·779 | 1·768 | 7·049 | 7060 | 2284 | 9018 |
| Yunnan Province | Puerhanizhuyi zuzizhi xian | 4·575 | 5·964 | 21·133 | 1·099 | 5·592 | 7220 | 1708 | 9124 |
| Yunnan Province | Qiaojia xian | 2·559 | 2·706 | 9·523 | 0·918 | 3·191 | 9576 | 3364 | 11956 |
| Yunnan Province | Qiubei xian | 5·045 | 4·804 | 17·601 | 1·973 | 6·373 | 23554 | 9255 | 29771 |
| Yunnan Province | Qujingshi | 3·675 | 3·601 | 12·994 | 1·447 | 4·591 | 53396 | 20561 | 66690 |
| Yunnan Province | Ruilishi | 3·392 | 4·530 | 15·337 | 0·837 | 4·115 | 7013 | 1728 | 8525 |
| Yunnan Province | Shangjianglahuzhuvazhublang zuzizhi xian | 4·244 | 5·559 | 19·971 | 0·984 | 5·202 | 7336 | 1691 | 8810 |
| Yunnan Province | Shidian xian | 4·095 | 5·631 | 19·213 | 0·981 | 4·861 | 12204 | 2942 | 14421 |
| Yunnan Province | Shiping xian | 4·959 | 6·071 | 21·743 | 1·361 | 6·120 | 13477 | 3662 | 16632 |
| Yunnan Province | Shizong xian | 4·549 | 4·082 | 14·985 | 1·902 | 5·774 | 15581 | 6391 | 19662 |

| | | β-thalassaemia carrier prevalence | | | | | Estimated carrier counts | | |
|---|---|---|---|---|---|---|---|---|---|
| ADM1 | ADM2 | Mean | SD | CI95% | Q25 | Q75 | Mean | Q25 | Q75 |
| Yunnan Province | Shuangbo xian | 4·482 | 5·673 | 19·948 | 1·198 | 5·455 | 6476 | 1757 | 7854 |
| Yunnan Province | Simao xian | 4·501 | 5·894 | 20·899 | 1·058 | 5·523 | 19160 | 4418 | 23950 |
| Yunnan Province | Songming xian | 3·983 | 4·305 | 15·184 | 1·363 | 4·991 | 18128 | 6272 | 22710 |
| Yunnan Province | Tengchong xian | 4·009 | 5·215 | 18·386 | 1·027 | 4·848 | 25804 | 6629 | 31376 |
| Yunnan Province | Tonghai xian | 4·886 | 5·786 | 20·315 | 1·422 | 6·072 | 13842 | 4029 | 17193 |
| Yunnan Province | Wandingshi | 3·633 | 4·853 | 18·131 | 0·860 | 4·311 | 909 | 218 | 1055 |
| Yunnan Province | Weishanyizuhui zuzizhi xian | 4·123 | 5·519 | 19·180 | 1·054 | 4·914 | 10883 | 2822 | 13028 |
| Yunnan Province | Weixin xian | 1·714 | 1·664 | 5·910 | 0·678 | 2·152 | 6060 | 2375 | 7457 |
| Yunnan Province | Wenshan xian | 5·548 | 5·649 | 20·627 | 1·958 | 7·045 | 36376 | 13258 | 46266 |
| Yunnan Province | Wuding xian | 3·781 | 4·429 | 15·533 | 1·168 | 4·686 | 13430 | 4203 | 16622 |
| Yunnan Province | Xangyun xian | 4·070 | 5·241 | 18·177 | 1·091 | 4·905 | 16445 | 4357 | 19698 |
| Yunnan Province | Xichou xian | 5·837 | 5·517 | 20·357 | 2·248 | 7·431 | 10001 | 3883 | 12723 |
| Yunnan Province | Ximengwa zuzizhi xian | 4·154 | 5·390 | 19·212 | 0·959 | 5·112 | 3354 | 772 | 4148 |
| Yunnan Province | Xinpingyizhudai zuzizhi xian | 4·719 | 6·037 | 21·445 | 1·220 | 5·732 | 12618 | 3278 | 15396 |
| Yunnan Province | Xuanweishi | 3·109 | 3·074 | 11·050 | 1·205 | 3·894 | 38149 | 15018 | 48054 |
| Yunnan Province | Xundianhuizuyi zuzizhi xian | 3·668 | 3·956 | 14·078 | 1·285 | 4·558 | 17327 | 6155 | 21394 |
| Yunnan Province | Yangbiyi zuzizhi xian | 4·160 | 5·509 | 18·745 | 1·061 | 4·999 | 4209 | 1068 | 5052 |
| Yunnan Province | Yaoan xian | 4·011 | 5·001 | 17·594 | 1·108 | 4·902 | 6848 | 1894 | 8182 |
| Yunnan Province | Yiliang xian | 4·270 | 4·451 | 16·015 | 1·480 | 5·401 | 22339 | 7721 | 27986 |
| Yunnan Province | Yiliang xian | 1·816 | 1·709 | 6·149 | 0·743 | 2·279 | 9451 | 3761 | 11926 |
| Yunnan Province | Yimen xian | 4·433 | 5·403 | 19·221 | 1·271 | 5·436 | 6547 | 1937 | 7999 |
| Yunnan Province | Yingjiang xian | 3·723 | 4·814 | 16·994 | 0·961 | 4·489 | 10917 | 2785 | 13243 |
| Yunnan Province | Yongde xian | 4·180 | 5·722 | 20·031 | 0·968 | 5·004 | 13462 | 3130 | 15898 |
| Yunnan Province | Yongping xian | 4·161 | 5·519 | 19·103 | 1·060 | 4·990 | 6966 | 1684 | 8394 |
| Yunnan Province | Yongren xian | 3·726 | 4·600 | 15·955 | 1·076 | 4·566 | 3915 | 1116 | 4926 |
| Yunnan Province | Yongsheng xian | 3·838 | 4·963 | 16·502 | 1·066 | 4·654 | 12211 | 3428 | 15016 |
| Yunnan Province | Yuanjianghanizhudai zuzizhi xian | 4·879 | 6·177 | 21·946 | 1·266 | 5·946 | 9239 | 2430 | 11041 |
| Yunnan Province | Yuanmou xian | 3·800 | 4·545 | 15·826 | 1·124 | 4·710 | 7381 | 2161 | 9142 |
| Yunnan Province | Yuanyang xian | 5·192 | 6·189 | 22·340 | 1·476 | 6·453 | 18910 | 5423 | 23507 |
| Yunnan Province | Yun xian | 4·318 | 5·901 | 20·561 | 1·025 | 5·128 | 17316 | 4070 | 20480 |
| Yunnan Province | Yunlong xian | 4·190 | 5·503 | 18·961 | 1·076 | 5·061 | 8124 | 2065 | 9842 |
| Yunnan Province | Yuxishi | 4·734 | 5·700 | 20·015 | 1·344 | 5·860 | 27924 | 8333 | 34848 |
| Yunnan Province | Zhaotongshi | 2·042 | 1·990 | 7·194 | 0·797 | 2·571 | 20466 | 8037 | 25733 |
| Yunnan Province | Zhenkang xian | 4·052 | 5·560 | 19·610 | 0·930 | 4·825 | 6055 | 1377 | 7230 |
| Yunnan Province | Zhenxiong xian | 1·899 | 1·815 | 6·540 | 0·767 | 2·383 | 26492 | 10752 | 33190 |
| Yunnan Province | Zhenyuanyizhuhanizhulahuzuz izhixain | 4·520 | 5·987 | 21·010 | 1·084 | 5·485 | 7848 | 1875 | 9478 |

| ADM1 | ADM2 | β-thalassaemia carrier prevalence Mean | SD | CI95% | Q25 | Q75 | Estimated carrier counts Mean | Q25 | Q75 |
|---|---|---|---|---|---|---|---|---|---|
| Yunnan Province | Zhongdian xian | 3·939 | 4·949 | 17·433 | 1·100 | 4·753 | 7088 | 2035 | 8558 |
| Zhejiang Province | Anji xian | 0·506 | 0·586 | 2·011 | 0·175 | 0·615 | 2906 | 995 | 3549 |
| Zhejiang Province | Changnan xian | 1·659 | 1·811 | 6·420 | 0·573 | 2·034 | 19243 | 6475 | 23597 |
| Zhejiang Province | Changshan xian | 1·273 | 1·462 | 4·986 | 0·439 | 1·543 | 3319 | 1159 | 3999 |
| Zhejiang Province | Changxing xian | 0·415 | 0·466 | 1·583 | 0·150 | 0·505 | 2518 | 911 | 3054 |
| Zhejiang Province | Cixishi | 0·618 | 0·947 | 2·641 | 0·180 | 0·723 | 7107 | 2060 | 8335 |
| Zhejiang Province | Cunan xian | 0·922 | 1·118 | 3·694 | 0·303 | 1·118 | 2951 | 967 | 3568 |
| Zhejiang Province | Deqing xian | 0·476 | 0·555 | 1·845 | 0·164 | 0·584 | 2888 | 1007 | 3602 |
| Zhejiang Province | Dongyangshi | 0·908 | 1·140 | 3·772 | 0·282 | 1·092 | 9967 | 3108 | 12000 |
| Zhejiang Province | Fenghuashi | 0·805 | 1·141 | 3·541 | 0·227 | 0·952 | 4299 | 1226 | 5121 |
| Zhejiang Province | Fuyangshi | 0·648 | 0·762 | 2·586 | 0·214 | 0·789 | 6051 | 2002 | 7317 |
| Zhejiang Province | Hainingshi | 0·495 | 0·596 | 1·954 | 0·163 | 0·603 | 4040 | 1334 | 4936 |
| Zhejiang Province | Haiyan xian | 0·490 | 0·595 | 1·993 | 0·158 | 0·593 | 1660 | 540 | 2015 |
| Zhejiang Province | Hangzhoushi | 0·571 | 0·677 | 2·239 | 0·187 | 0·700 | 9233 | 2998 | 11133 |
| Zhejiang Province | Huzhoushi | 0·423 | 0·476 | 1·637 | 0·150 | 0·515 | 5264 | 1879 | 6451 |
| Zhejiang Province | Jiandeshi | 0·887 | 1·065 | 3·542 | 0·291 | 1·080 | 4166 | 1362 | 5073 |
| Zhejiang Province | Jiangshanshi | 1·440 | 1·621 | 5·601 | 0·500 | 1·759 | 7084 | 2472 | 8669 |
| Zhejiang Province | Jiashan xian | 0·416 | 0·498 | 1·628 | 0·142 | 0·504 | 2962 | 1000 | 3573 |
| Zhejiang Province | Jiaxingshi | 0·442 | 0·536 | 1·762 | 0·146 | 0·533 | 6968 | 2321 | 8430 |
| Zhejiang Province | Jingningshe zuzizhi xian | 1·547 | 1·663 | 5·745 | 0·566 | 1·903 | 1539 | 540 | 1895 |
| Zhejiang Province | Jinhuashi | 1·042 | 1·212 | 4·167 | 0·344 | 1·270 | 14604 | 4842 | 17776 |
| Zhejiang Province | Jinyun xian | 1·142 | 1·364 | 4·693 | 0·368 | 1·389 | 4475 | 1461 | 5469 |
| Zhejiang Province | Lanxishi | 0·941 | 1·101 | 3·682 | 0·315 | 1·150 | 5601 | 1869 | 6843 |
| Zhejiang Province | Leqing xian | 1·314 | 1·656 | 5·680 | 0·383 | 1·581 | 15418 | 4563 | 18439 |
| Zhejiang Province | Linan xian | 0·646 | 0·757 | 2·587 | 0·214 | 0·789 | 3562 | 1178 | 4356 |
| Zhejiang Province | Linhaishi | 1·072 | 1·397 | 4·686 | 0·308 | 1·276 | 11736 | 3329 | 14289 |
| Zhejiang Province | Lishuishi | 1·276 | 1·490 | 5·113 | 0·422 | 1·565 | 7602 | 2481 | 9227 |
| Zhejiang Province | Longquanshi | 1·567 | 1·715 | 5·909 | 0·562 | 1·928 | 3886 | 1391 | 4790 |
| Zhejiang Province | Longyou xian | 1·109 | 1·264 | 4·326 | 0·375 | 1·364 | 3191 | 1069 | 3953 |
| Zhejiang Province | Ningboshi | 0·751 | 1·217 | 3·347 | 0·203 | 0·862 | 11924 | 3240 | 13639 |
| Zhejiang Province | Ninghai xian | 0·925 | 1·306 | 4·133 | 0·256 | 1·078 | 5804 | 1581 | 6738 |
| Zhejiang Province | Ouhai xian | 1·410 | 1·675 | 5·733 | 0·453 | 1·736 | 11285 | 3551 | 13920 |
| Zhejiang Province | Panan xian | 0·993 | 1·244 | 4·285 | 0·302 | 1·191 | 1455 | 433 | 1740 |
| Zhejiang Province | Pinghushi | 0·470 | 0·593 | 1·899 | 0·150 | 0·568 | 3174 | 1027 | 3843 |
| Zhejiang Province | Pingyang xian | 1·547 | 1·706 | 6·095 | 0·527 | 1·903 | 11209 | 3771 | 13719 |
| Zhejiang Province | Pujiang xian | 0·813 | 0·982 | 3·280 | 0·263 | 0·983 | 3202 | 1024 | 3785 |

| ADM1 | ADM2 | β-thalassaemia carrier prevalence Mean | SD | CI95% | Q25 | Q75 | Estimated carrier counts Mean | Q25 | Q75 |
|---|---|---|---|---|---|---|---|---|---|
| Zhejiang Province | Qingpu xian | 0·458 | 0·591 | 1·892 | 0·145 | 0·543 | 3926 | 1236 | 4667 |
| Zhejiang Province | Qingtian xian | 1·345 | 1·536 | 5·257 | 0·456 | 1·657 | 4642 | 1573 | 5841 |
| Zhejiang Province | Qu xian | 1·229 | 1·575 | 4·781 | 0·404 | 1·467 | 3776 | 1227 | 4476 |
| Zhejiang Province | Qu xian | 1·189 | 1·384 | 4·695 | 0·401 | 1·447 | 7241 | 2410 | 8862 |
| Zhejiang Province | Ruianshi | 1·478 | 1·699 | 5·883 | 0·490 | 1·810 | 23701 | 7642 | 29294 |
| Zhejiang Province | Sanmen xian | 1·047 | 1·474 | 4·649 | 0·292 | 1·231 | 3105 | 875 | 3644 |
| Zhejiang Province | Shangyushi | 0·663 | 0·859 | 2·806 | 0·200 | 0·793 | 4699 | 1426 | 5623 |
| Zhejiang Province | Shaoxing xian | 0·636 | 0·773 | 2·639 | 0·200 | 0·775 | 8418 | 2673 | 10299 |
| Zhejiang Province | Shaoxingshi | 0·654 | 0·891 | 2·690 | 0·200 | 0·775 | 2158 | 661 | 2558 |
| Zhejiang Province | Sheng xian | 0·781 | 0·995 | 3·359 | 0·234 | 0·936 | 5674 | 1692 | 6830 |
| Zhejiang Province | Shuzhoushi | 0·367 | 0·424 | 1·416 | 0·133 | 0·443 | 2284 | 828 | 2762 |
| Zhejiang Province | Songyang xian | 1·344 | 1·521 | 5·250 | 0·458 | 1·654 | 2425 | 818 | 3063 |
| Zhejiang Province | Suichang xian | 1·339 | 1·512 | 5·202 | 0·460 | 1·637 | 2637 | 888 | 3214 |
| Zhejiang Province | Taizhoushi | 1·177 | 1·527 | 5·157 | 0·340 | 1·402 | 25935 | 7312 | 30862 |
| Zhejiang Province | Tiantai xian | 0·957 | 1·262 | 4·161 | 0·277 | 1·141 | 4611 | 1333 | 5451 |
| Zhejiang Province | Tonglu xian | 0·731 | 0·875 | 2·936 | 0·241 | 0·890 | 2887 | 935 | 3496 |
| Zhejiang Province | Tongxiangshi | 0·459 | 0·523 | 1·776 | 0·157 | 0·565 | 5174 | 1776 | 6332 |
| Zhejiang Province | Wencheng xian | 1·530 | 1·724 | 5·982 | 0·533 | 1·865 | 4289 | 1513 | 5176 |
| Zhejiang Province | Wenlingshi | 1·266 | 1·620 | 5·667 | 0·362 | 1·501 | 13825 | 3939 | 16338 |
| Zhejiang Province | Wenzhoushi | 1·421 | 1·751 | 5·921 | 0·456 | 1·690 | 13116 | 4164 | 15474 |
| Zhejiang Province | Wujiangshi | 0·389 | 0·457 | 1·529 | 0·137 | 0·468 | 5891 | 2062 | 7081 |
| Zhejiang Province | Wuyi xian | 1·132 | 1·299 | 4·469 | 0·380 | 1·391 | 4327 | 1436 | 5292 |
| Zhejiang Province | Xiangshan xian | 0·938 | 1·494 | 4·245 | 0·243 | 1·088 | 3958 | 1058 | 4592 |
| Zhejiang Province | Xianju xian | 1·100 | 1·346 | 4·634 | 0·339 | 1·324 | 4520 | 1353 | 5461 |
| Zhejiang Province | Xiaoshanshi | 0·588 | 0·728 | 2·360 | 0·191 | 0·713 | 15926 | 5172 | 19315 |
| Zhejiang Province | Xinchang xian | 0·845 | 1·106 | 3·639 | 0·247 | 1·009 | 2803 | 814 | 3390 |
| Zhejiang Province | Ying xian | 0·779 | 1·204 | 3·428 | 0·216 | 0·900 | 18800 | 5217 | 21527 |
| Zhejiang Province | Yiwushi | 0·901 | 1·141 | 3·656 | 0·285 | 1·083 | 18413 | 5809 | 22137 |
| Zhejiang Province | Yongjia xian | 1·266 | 1·529 | 5·278 | 0·394 | 1·534 | 10247 | 3181 | 12449 |
| Zhejiang Province | Yongkangshi | 1·042 | 1·272 | 4·308 | 0·334 | 1·258 | 10415 | 3261 | 12467 |
| Zhejiang Province | Yuhang xian | 0·526 | 0·619 | 2·063 | 0·178 | 0·644 | 24840 | 8295 | 30420 |
| Zhejiang Province | Yuhuan xian | 1·351 | 1·763 | 6·114 | 0·391 | 1·585 | 6567 | 1913 | 7793 |
| Zhejiang Province | Yunhe xian | 1·446 | 1·612 | 5·442 | 0·511 | 1·776 | 1848 | 628 | 2225 |
| Zhejiang Province | Yuyaoshi | 0·662 | 0·932 | 2·793 | 0·196 | 0·782 | 11080 | 3332 | 13049 |
| Zhejiang Province | Zhujishi | 0·746 | 0·935 | 3·129 | 0·232 | 0·897 | 8999 | 2782 | 10745 |

ADM1: administrative level 1. ADM2: administrative level 2. SD: standard deviation. CI95%: 95% credible interval. Q25: 25th percentile. Q75: 75th percentile. SD and CI95% are measures of prediction uncertainty.

**Supplementary Table S20. Subnational areal predictions of β-thalassaemia carrier prevalence and estimated carrier count at administrative level 2 of Cyprus.**

| | | β-thalassaemia carrier prevalence | | | | | Estimated carrier counts | | |
|---|---|---|---|---|---|---|---|---|---|
| **ADM1** | **ADM2** | **Mean** | **SD** | **CI95%** | **Q25** | **Q75** | **Mean** | **Q25** | **Q75** |
| Famagusta | Acheritou | 12·529 | 7·356 | 27·776 | 7·130 | 15·658 | 188 | 107 | 234 |
| Famagusta | Achna | 12·795 | 7·282 | 27·458 | 7·469 | 16·419 | 247 | 144 | 318 |
| Famagusta | Afanteia-Orniti | 12·660 | 6·903 | 25·720 | 7·408 | 16·133 | 205 | 120 | 261 |
| Famagusta | Agia Napa | 11·896 | 7·068 | 27·372 | 6·830 | 14·903 | 110 | 63 | 137 |
| Famagusta | Agia Trias | 9·546 | 6·282 | 22·724 | 5·139 | 12·254 | 304 | 163 | 388 |
| Famagusta | Agios Andronikos Karpasias | 9·832 | 6·287 | 22·753 | 5·355 | 12·738 | 279 | 154 | 355 |
| Famagusta | Agios Andronikos Trikomou | 11·391 | 6·810 | 25·621 | 6·514 | 14·506 | 33 | 19 | 42 |
| Famagusta | Agios Chariton | 12·265 | 6·973 | 26·844 | 7·238 | 15·636 | 849 | 502 | 1085 |
| Famagusta | Agios Efstathios | 10·845 | 7·188 | 26·297 | 5·683 | 14·091 | 129 | 68 | 168 |
| Famagusta | Agios Georgios Ammochostou | 11·689 | 6·898 | 26·760 | 6·737 | 14·870 | 1303 | 751 | 1672 |
| Famagusta | Agios Iakovos | 11·563 | 6·960 | 26·402 | 6·473 | 14·965 | 72 | 41 | 94 |
| Famagusta | Agios Ilias | 11·185 | 6·941 | 26·147 | 6·344 | 14·355 | 298 | 169 | 383 |
| Famagusta | Agios Nikolaos Ammochostou | 12·239 | 7·084 | 27·442 | 7·268 | 15·522 | 85 | 50 | 107 |
| Famagusta | Agios Sergios | 11·441 | 6·627 | 25·319 | 6·821 | 14·372 | 758 | 452 | 953 |
| Famagusta | Agios Symeon | 9·693 | 6·311 | 22·533 | 5·218 | 12·563 | 194 | 105 | 247 |
| Famagusta | Agios Theodoros Ammochostou | 11·490 | 7·062 | 27·931 | 6·385 | 14·723 | 28 | 16 | 36 |
| Famagusta | Agkomi Ammochostou | 12·224 | 7·447 | 29·842 | 6·986 | 15·622 | 1215 | 695 | 1553 |
| Famagusta | Aigialousa | 9·415 | 6·288 | 22·899 | 5·137 | 11·902 | 27 | 15 | 34 |
| Famagusta | Akanthou | 11·228 | 6·746 | 25·305 | 6·379 | 14·443 | 144 | 82 | 179 |
| Famagusta | Aloda | 11·823 | 6·876 | 26·794 | 6·830 | 15·190 | 534 | 310 | 682 |
| Famagusta | Ammochostos | 11·780 | 6·776 | 25·200 | 6·858 | 15·113 | 2915 | 1697 | 3740 |
| Famagusta | Angastina | 11·994 | 6·491 | 25·721 | 7·460 | 15·059 | 236 | 147 | 297 |
| Famagusta | Ardana | 11·964 | 7·936 | 30·110 | 6·507 | 14·850 | 98 | 53 | 122 |
| Famagusta | Arnadi | 11·838 | 6·971 | 27·621 | 6·841 | 14·919 | 579 | 336 | 734 |
| Famagusta | Artemi | 11·970 | 6·844 | 25·926 | 6·972 | 15·528 | 256 | 150 | 335 |
| Famagusta | Askeia | 12·092 | 6·475 | 24·828 | 7·337 | 15·437 | 364 | 221 | 465 |
| Famagusta | Avgolida | 11·139 | 6·938 | 27·201 | 6·181 | 14·305 | 790 | 441 | 1019 |
| Famagusta | Avgorou | 12·749 | 7·551 | 29·964 | 7·264 | 16·332 | 595 | 339 | 762 |
| Famagusta | Davios | 11·080 | 7·059 | 26·602 | 5·927 | 14·200 | 42 | 22 | 53 |
| Famagusta | Deryneia | 11·714 | 6·825 | 25·903 | 6·840 | 14·986 | 5593 | 3254 | 7160 |
| Famagusta | Eptakomi | 10·728 | 6·977 | 27·088 | 5·857 | 13·752 | 280 | 155 | 362 |
| Famagusta | Flamoudi | 11·278 | 6·770 | 27·306 | 6·651 | 14·243 | 19 | 11 | 24 |
| Famagusta | Frenaros | 12·086 | 7·000 | 26·948 | 6·999 | 15·582 | 506 | 293 | 653 |
| Famagusta | Gaidouras | 12·059 | 6·933 | 26·201 | 7·067 | 15·470 | 118 | 69 | 151 |
| Famagusta | Galateia | 10·684 | 6·885 | 25·717 | 5·746 | 13·781 | 899 | 483 | 1162 |
| Famagusta | Galinoporni | 9·512 | 6·210 | 22·756 | 4·986 | 12·339 | 83 | 43 | 107 |
| Famagusta | Gastria | 11·330 | 7·020 | 28·323 | 6·339 | 14·562 | 224 | 125 | 288 |
| Famagusta | Genagra | 12·148 | 6·934 | 27·368 | 7·144 | 15·473 | 80 | 47 | 102 |
| Famagusta | Gerani | 11·241 | 7·162 | 27·175 | 6·160 | 14·259 | 491 | 273 | 626 |
| Famagusta | Goufes | 12·066 | 6·808 | 25·268 | 7·114 | 15·584 | 455 | 269 | 588 |
| Famagusta | Gypsou | 11·896 | 6·801 | 26·168 | 6·886 | 15·389 | 185 | 107 | 239 |
| Famagusta | Kalopsida | 12·802 | 7·446 | 28·178 | 7·279 | 16·363 | 123 | 70 | 158 |
| Famagusta | Kiados | 12·455 | 7·084 | 26·821 | 7·297 | 15·991 | 192 | 113 | 247 |
| Famagusta | Knodara | 12·012 | 6·836 | 25·798 | 7·130 | 15·265 | 212 | 126 | 269 |
| Famagusta | Koilanemos | 10·052 | 6·473 | 23·926 | 5·574 | 12·711 | 256 | 142 | 324 |
| Famagusta | Koma tou Gialou | 10·517 | 6·848 | 25·216 | 5·638 | 13·395 | 592 | 316 | 761 |
| Famagusta | Komi Kepir | 11·307 | 7·079 | 28·495 | 6·177 | 14·673 | 215 | 118 | 280 |
| Famagusta | Kontea | 12·207 | 6·695 | 25·550 | 7·500 | 15·771 | 210 | 129 | 271 |
| Famagusta | Kornokipos | 12·307 | 6·972 | 26·611 | 7·242 | 15·290 | 181 | 107 | 225 |
| Famagusta | Koroveia | 9·562 | 6·155 | 22·168 | 5·061 | 12·552 | 106 | 55 | 138 |
| Famagusta | Kouklia Ammochostou | 12·787 | 7·118 | 26·738 | 7·659 | 16·475 | 124 | 74 | 160 |
| Famagusta | Krideia | 11·020 | 6·896 | 27·167 | 6·021 | 14·152 | 268 | 147 | 348 |
| Famagusta | Lapathos | 11·664 | 6·918 | 26·879 | 6·661 | 14·891 | 423 | 243 | 541 |
| Famagusta | Lefkonoiko | 11·921 | 6·881 | 25·744 | 6·881 | 15·312 | 345 | 201 | 447 |

| | | β-thalassaemia carrier prevalence | | | | | Estimated carrier counts | | |
|---|---|---|---|---|---|---|---|---|---|
| ADM1 | ADM2 | Mean | SD | CI95% | Q25 | Q75 | Mean | Q25 | Q75 |
| Famagusta | Leonarisso | 10·526 | 6·949 | 24·901 | 5·452 | 13·270 | 205 | 106 | 258 |
| Famagusta | Limnia | 11·733 | 6·804 | 25·729 | 6·823 | 14·987 | 695 | 404 | 886 |
| Famagusta | Liopetri | 11·932 | 7·040 | 26·847 | 6·790 | 15·533 | 439 | 250 | 572 |
| Famagusta | Livadia Ammochostou | 10·845 | 7·188 | 26·297 | 5·683 | 14·091 | 129 | 68 | 168 |
| Famagusta | Lysi | 12·544 | 6·922 | 27·014 | 7·697 | 15·766 | 527 | 324 | 663 |
| Famagusta | Lythragkomi | 10·162 | 6·635 | 24·933 | 5·615 | 12·759 | 354 | 196 | 447 |
| Famagusta | Makrasyka | 12·497 | 6·907 | 26·144 | 7·579 | 16·123 | 334 | 203 | 431 |
| Famagusta | Mandres | 11·410 | 6·806 | 25·920 | 6·522 | 14·682 | 160 | 91 | 206 |
| Famagusta | Maratha | 11·998 | 7·101 | 28·511 | 6·967 | 15·403 | 404 | 236 | 516 |
| Famagusta | Marathovounos | 12·517 | 7·127 | 27·400 | 7·301 | 15·997 | 253 | 149 | 322 |
| Famagusta | Melanarga | 9·775 | 6·523 | 23·487 | 5·302 | 12·361 | 171 | 93 | 216 |
| Famagusta | Melounta | 11·821 | 6·908 | 27·113 | 6·847 | 15·153 | 13 | 7 | 16 |
| Famagusta | Milia Ammochostou | 11·882 | 6·988 | 27·530 | 6·861 | 15·126 | 1270 | 737 | 1617 |
| Famagusta | Mousoulita | 12·390 | 6·976 | 26·528 | 7·288 | 15·932 | 977 | 579 | 1259 |
| Famagusta | Neta | 9·775 | 6·523 | 23·487 | 5·302 | 12·361 | 171 | 93 | 216 |
| Famagusta | Ovgoros | 10·734 | 6·713 | 25·840 | 5·864 | 13·631 | 53 | 29 | 68 |
| Famagusta | Paralimni | 11·863 | 7·031 | 26·812 | 6·887 | 15·174 | 2300 | 1322 | 2940 |
| Famagusta | Patanissos | 10·645 | 6·938 | 25·909 | 5·662 | 13·564 | 373 | 198 | 475 |
| Famagusta | Patriki | 11·231 | 7·058 | 27·439 | 6·186 | 14·481 | 894 | 496 | 1155 |
| Famagusta | Peristerona Ammochostou | 12·219 | 7·291 | 29·349 | 7·188 | 15·559 | 316 | 186 | 402 |
| Famagusta | Perivolia Trikomou | 11·390 | 6·753 | 25·039 | 6·529 | 14·772 | 724 | 415 | 938 |
| Famagusta | Pigi | 11·967 | 7·030 | 27·784 | 6·936 | 15·250 | 586 | 341 | 746 |
| Famagusta | Platani | 11·561 | 6·754 | 25·948 | 6·666 | 14·782 | 32 | 18 | 41 |
| Famagusta | Prastio Ammochostou | 12·369 | 7·020 | 26·050 | 7·249 | 15·806 | 225 | 132 | 288 |
| Famagusta | Psyllatos | 12·185 | 6·939 | 26·290 | 7·140 | 15·659 | 558 | 329 | 720 |
| Famagusta | Pyrga Ammochostou | 12·230 | 7·040 | 27·538 | 7·167 | 15·630 | 854 | 501 | 1092 |
| Famagusta | Rizokarpaso | 8·821 | 5·971 | 22·391 | 4·594 | 11·286 | 334 | 175 | 425 |
| Famagusta | Santalaris | 11·777 | 6·911 | 27·673 | 6·745 | 15·248 | 88 | 50 | 114 |
| Famagusta | Sinta | 12·185 | 6·912 | 27·387 | 7·088 | 15·681 | 233 | 136 | 300 |
| Famagusta | Sotira Ammochostou | 11·659 | 6·602 | 26·168 | 7·059 | 14·887 | 1122 | 679 | 1432 |
| Famagusta | Spathariko | 11·567 | 6·740 | 25·425 | 6·755 | 14·759 | 1928 | 1127 | 2459 |
| Famagusta | Strongylos | 12·262 | 6·825 | 25·656 | 7·276 | 15·868 | 724 | 430 | 937 |
| Famagusta | Stylloi | 11·825 | 6·863 | 26·333 | 6·864 | 15·173 | 901 | 523 | 1151 |
| Famagusta | Sygkrasi | 11·807 | 7·102 | 29·326 | 6·768 | 14·706 | 133 | 76 | 166 |
| Famagusta | Tavrou | 10·753 | 6·799 | 24·806 | 5·804 | 14·109 | 289 | 156 | 380 |
| Famagusta | Trikomo | 11·390 | 6·753 | 25·039 | 6·529 | 14·772 | 724 | 415 | 938 |
| Famagusta | Trypimeni | 12·206 | 6·985 | 26·819 | 7·219 | 15·576 | 624 | 369 | 796 |
| Famagusta | Vasili | 10·273 | 6·796 | 25·941 | 5·656 | 12·806 | 98 | 54 | 123 |
| Famagusta | Vatili | 12·433 | 7·175 | 26·484 | 7·215 | 16·300 | 360 | 209 | 472 |
| Famagusta | Vitsada | 12·375 | 7·044 | 26·855 | 7·257 | 15·812 | 657 | 388 | 838 |
| Famagusta | Vokolida | 10·799 | 6·993 | 25·552 | 5·743 | 14·100 | 418 | 224 | 548 |
| Famagusta | Vothylakas | 10·033 | 6·598 | 24·451 | 5·511 | 12·626 | 525 | 289 | 663 |
| Famagusta | Xylofagou | 12·432 | 6·870 | 26·022 | 7·358 | 15·894 | 790 | 468 | 1010 |
| Kyrenia | Agia Eirini Keryneias | 12·881 | 7·141 | 27·226 | 7·544 | 16·475 | 49 | 29 | 63 |
| Kyrenia | Agios Amvrosios Keryneias | 12·181 | 6·898 | 26·727 | 7·230 | 15·737 | 341 | 202 | 440 |
| Kyrenia | Agios Epiktitos | 12·512 | 6·977 | 26·461 | 7·670 | 15·693 | 1415 | 856 | 1795 |
| Kyrenia | Agios Ermolaos | 12·799 | 7·129 | 27·310 | 7·694 | 16·032 | 137 | 82 | 171 |
| Kyrenia | Agios Georgios Keryneias | 12·236 | 6·851 | 25·961 | 7·392 | 15·209 | 1402 | 847 | 1743 |
| Kyrenia | Agirda | 12·616 | 6·920 | 26·118 | 7·615 | 16·053 | 2710 | 1636 | 3425 |
| Kyrenia | Agridaki | 12·748 | 6·985 | 27·063 | 7·717 | 16·123 | 2404 | 1432 | 3049 |
| Kyrenia | Asomatos Keryneias | 12·650 | 6·757 | 25·829 | 7·851 | 16·062 | 68 | 42 | 86 |
| Kyrenia | Belapais | 12·441 | 6·862 | 25·951 | 7·527 | 15·765 | 8023 | 4823 | 10180 |
| Kyrenia | Charkeia | 12·268 | 7·347 | 27·651 | 6·972 | 15·758 | 252 | 143 | 324 |
| Kyrenia | Diorios | 13·053 | 7·052 | 26·523 | 7·882 | 16·676 | 416 | 249 | 529 |
| Kyrenia | Ella Keryneias | 12·205 | 6·515 | 24·849 | 7·391 | 15·400 | 1318 | 798 | 1664 |
| Kyrenia | Fota | 12·933 | 7·003 | 27·642 | 7·909 | 16·376 | 163 | 100 | 206 |
| Kyrenia | Ftericha | 12·205 | 6·515 | 24·849 | 7·391 | 15·400 | 1318 | 798 | 1664 |

| | | β-thalassaemia carrier prevalence | | | | | Estimated carrier counts | | |
|---|---|---|---|---|---|---|---|---|---|
| **ADM1** | **ADM2** | **Mean** | **SD** | **CI95%** | **Q25** | **Q75** | **Mean** | **Q25** | **Q75** |
| Kyrenia | Kalograia | 12·119 | 6·937 | 27·216 | 7·181 | 15·526 | 135 | 80 | 173 |
| Kyrenia | Kampyli | 12·777 | 7·002 | 26·875 | 7·780 | 16·255 | 1351 | 816 | 1711 |
| Kyrenia | Karavas | 12·274 | 6·746 | 25·829 | 7·337 | 15·474 | 3990 | 2386 | 5029 |
| Kyrenia | Karmi | 12·347 | 6·842 | 25·826 | 7·470 | 15·492 | 5236 | 3168 | 6576 |
| Kyrenia | Karpaseia | 12·967 | 6·981 | 27·022 | 7·860 | 16·544 | 301 | 180 | 385 |
| Kyrenia | Kato Dikomo | 12·414 | 6·623 | 24·773 | 7·583 | 15·714 | 650 | 397 | 823 |
| Kyrenia | Kazafani | 12·428 | 6·791 | 25·154 | 7·320 | 16·103 | 2045 | 1205 | 2650 |
| Kyrenia | Keryneia | 12·599 | 7·161 | 26·667 | 7·628 | 15·869 | 2516 | 1523 | 3169 |
| Kyrenia | Kiomourtzou | 12·717 | 7·068 | 26·791 | 7·631 | 16·222 | 2060 | 1239 | 2602 |
| Kyrenia | Klepini | 12·435 | 6·930 | 26·881 | 7·562 | 15·631 | 1971 | 1189 | 2496 |
| Kyrenia | Kontemenos | 12·944 | 6·904 | 26·918 | 8·095 | 16·446 | 66 | 41 | 84 |
| Kyrenia | Kormakitis | 12·409 | 6·872 | 26·716 | 7·408 | 15·741 | 74 | 44 | 92 |
| Kyrenia | Koutsoventis | 12·381 | 7·021 | 28·311 | 7·510 | 15·262 | 70 | 42 | 86 |
| Kyrenia | Krini | 13·197 | 7·286 | 27·622 | 7·871 | 17·236 | 658 | 392 | 859 |
| Kyrenia | Lapithos | 12·381 | 6·873 | 26·678 | 7·228 | 15·813 | 1270 | 741 | 1622 |
| Kyrenia | Larnakas Lapithou | 12·731 | 6·937 | 26·980 | 7·725 | 16·153 | 2267 | 1350 | 2878 |
| Kyrenia | Livera | 12·464 | 6·867 | 25·769 | 7·583 | 15·653 | 42 | 26 | 53 |
| Kyrenia | Motides | 12·205 | 6·515 | 24·849 | 7·391 | 15·400 | 1318 | 798 | 1664 |
| Kyrenia | Myrtou | 12·643 | 7·314 | 27·409 | 7·320 | 16·314 | 286 | 165 | 369 |
| Kyrenia | Orga | 12·840 | 6·957 | 25·610 | 7·864 | 16·103 | 161 | 98 | 202 |
| Kyrenia | Palaiosofos | 12·205 | 6·515 | 24·849 | 7·391 | 15·400 | 1318 | 798 | 1664 |
| Kyrenia | Panagra | 12·742 | 7·136 | 26·509 | 7·592 | 16·208 | 447 | 263 | 571 |
| Kyrenia | Pano Dikomo | 12·190 | 6·659 | 26·359 | 7·489 | 15·425 | 84 | 52 | 106 |
| Kyrenia | Pileri | 12·646 | 6·882 | 26·600 | 7·665 | 15·936 | 1618 | 980 | 2041 |
| Kyrenia | Sichari | 12·498 | 6·758 | 26·556 | 7·695 | 15·784 | 7233 | 4449 | 9092 |
| Kyrenia | Sysklipos | 12·579 | 6·880 | 26·620 | 7·555 | 15·905 | 2888 | 1721 | 3663 |
| Kyrenia | Templos | 12·418 | 7·006 | 26·314 | 7·510 | 15·539 | 3918 | 2370 | 4912 |
| Kyrenia | Thermeia | 12·514 | 6·976 | 25·911 | 7·474 | 15·986 | 4561 | 2728 | 5819 |
| Kyrenia | Trapeza | 12·230 | 6·723 | 26·982 | 7·544 | 15·518 | 48 | 30 | 61 |
| Kyrenia | Trimithi | 12·236 | 6·851 | 25·961 | 7·392 | 15·209 | 1402 | 847 | 1743 |
| Kyrenia | Vasileia | 12·869 | 7·033 | 27·346 | 7·853 | 16·199 | 931 | 568 | 1172 |
| Kyrenia | Vouno | 12·706 | 6·963 | 26·591 | 7·742 | 16·068 | 131 | 80 | 165 |
| Larnaca | Agia Anna | 13·125 | 7·076 | 27·153 | 8·011 | 16·710 | 686 | 417 | 872 |
| Larnaca | Agios Theodoros Larnakas | 13·470 | 6·857 | 25·454 | 8·523 | 17·203 | 176 | 111 | 224 |
| Larnaca | Agloi Vavatsinias | 13·691 | 7·216 | 27·996 | 8·434 | 17·343 | 122 | 74 | 154 |
| Larnaca | Alaminos | 13·829 | 7·532 | 28·267 | 8·231 | 17·542 | 129 | 77 | 164 |
| Larnaca | Alethriko | 13·394 | 7·025 | 27·346 | 8·137 | 17·017 | 275 | 167 | 350 |
| Larnaca | Anafotida | 13·509 | 7·339 | 27·613 | 8·080 | 17·364 | 486 | 291 | 626 |
| Larnaca | Anglisides | 13·556 | 7·609 | 29·128 | 7·913 | 17·489 | 175 | 102 | 225 |
| Larnaca | Aplanta | 13·364 | 7·170 | 27·306 | 8·048 | 17·189 | 632 | 381 | 812 |
| Larnaca | Aradippou | 12·813 | 6·797 | 26·366 | 7·759 | 16·235 | 3602 | 2208 | 4530 |
| Larnaca | Arsos Larnakas | 13·024 | 7·485 | 30·093 | 7·845 | 16·011 | 197 | 119 | 242 |
| Larnaca | Athienou | 12·852 | 6·967 | 26·482 | 7·748 | 16·617 | 733 | 442 | 950 |
| Larnaca | Avdellero | 12·830 | 6·902 | 26·785 | 7·722 | 16·467 | 1443 | 868 | 1850 |
| Larnaca | Choirokoitia | 13·795 | 7·363 | 27·885 | 8·308 | 17·573 | 226 | 136 | 289 |
| Larnaca | Delikipos | 13·211 | 6·824 | 25·219 | 8·284 | 16·813 | 57 | 35 | 72 |
| Larnaca | Dromolaxia | 13·139 | 7·194 | 27·071 | 7·950 | 16·773 | 12082 | 7242 | 15298 |
| Larnaca | Kalavasos | 13·534 | 7·209 | 28·346 | 8·234 | 17·381 | 213 | 129 | 272 |
| Larnaca | Kalo Chorio Larnakas | 13·262 | 7·013 | 27·094 | 8·187 | 16·993 | 292 | 178 | 374 |
| Larnaca | Kato Drys | 13·630 | 7·253 | 27·479 | 8·233 | 17·468 | 280 | 168 | 359 |
| Larnaca | Kato Lefkara | 13·464 | 7·220 | 27·536 | 8·157 | 17·259 | 250 | 151 | 321 |
| Larnaca | Kellia | 12·994 | 7·437 | 26·912 | 7·470 | 16·613 | 886 | 509 | 1132 |
| Larnaca | Kiti | 12·887 | 7·307 | 27·003 | 7·739 | 16·543 | 1637 | 983 | 2101 |
| Larnaca | Kivisili | 13·279 | 7·031 | 26·740 | 8·139 | 16·980 | 685 | 419 | 876 |
| Larnaca | Klavdia | 13·305 | 7·061 | 27·241 | 8·174 | 16·966 | 795 | 485 | 1013 |
| Larnaca | Kofinou | 13·363 | 7·272 | 27·830 | 8·021 | 17·324 | 58 | 35 | 75 |
| Larnaca | Kosi | 12·888 | 7·081 | 27·592 | 7·782 | 16·358 | 152 | 92 | 192 |

| | | β-thalassaemia carrier prevalence | | | | | Estimated carrier counts | | |
|---|---|---|---|---|---|---|---|---|---|
| ADM1 | ADM2 | Mean | SD | CI95% | Q25 | Q75 | Mean | Q25 | Q75 |
| Larnaca | Lageia | 13·801 | 7·369 | 27·932 | 8·466 | 17·609 | 111 | 67 | 142 |
| Larnaca | Larnaka | 13·294 | 7·372 | 27·411 | 7·954 | 17·061 | 7137 | 4224 | 9036 |
| Larnaca | Livadia Larnakas | 12·992 | 7·365 | 27·665 | 7·613 | 16·485 | 8975 | 5293 | 11359 |
| Larnaca | Mari | 13·743 | 7·373 | 28·519 | 8·437 | 17·417 | 218 | 133 | 277 |
| Larnaca | Maroni | 13·524 | 7·132 | 28·280 | 8·187 | 17·232 | 210 | 127 | 268 |
| Larnaca | Mazotos | 13·143 | 6·875 | 25·442 | 8·095 | 17·059 | 182 | 112 | 237 |
| Larnaca | Melini | 13·883 | 7·641 | 29·568 | 8·505 | 17·556 | 156 | 95 | 197 |
| Larnaca | Melouseia | 12·798 | 7·055 | 26·732 | 7·720 | 16·385 | 408 | 246 | 522 |
| Larnaca | Meneou | 13·037 | 7·225 | 26·578 | 7·861 | 16·859 | 1957 | 1176 | 2517 |
| Larnaca | Menogeia | 13·583 | 7·471 | 28·409 | 8·055 | 17·452 | 362 | 214 | 464 |
| Larnaca | Mosfiloti | 13·383 | 7·350 | 27·979 | 8·164 | 16·930 | 1484 | 903 | 1874 |
| Larnaca | Odou | 13·764 | 7·407 | 28·580 | 8·482 | 17·467 | 258 | 158 | 327 |
| Larnaca | Ora | 13·824 | 7·396 | 28·623 | 8·538 | 17·421 | 104 | 63 | 131 |
| Larnaca | Pano Lefkara | 13·671 | 7·244 | 27·263 | 8·240 | 17·541 | 317 | 192 | 410 |
| Larnaca | Pergamos | 12·562 | 6·903 | 26·223 | 7·538 | 16·164 | 43 | 26 | 56 |
| Larnaca | Perivolia Larnakas | 13·037 | 7·225 | 26·578 | 7·861 | 16·859 | 1957 | 1176 | 2517 |
| Larnaca | Petrofani | 13·074 | 7·196 | 27·071 | 7·844 | 16·759 | 750 | 452 | 971 |
| Larnaca | Psematismenos | 13·622 | 7·166 | 27·547 | 8·335 | 17·406 | 687 | 420 | 877 |
| Larnaca | Psevdas | 13·299 | 7·279 | 27·785 | 8·111 | 16·768 | 942 | 566 | 1179 |
| Larnaca | Pyla | 12·572 | 6·980 | 26·818 | 7·492 | 16·030 | 1315 | 779 | 1663 |
| Larnaca | Pyrga Larnakas | 13·328 | 7·194 | 26·395 | 8·121 | 16·850 | 242 | 147 | 306 |
| Larnaca | Sia | 13·483 | 7·283 | 27·790 | 8·301 | 17·312 | 331 | 204 | 425 |
| Larnaca | Skarinou | 13·615 | 7·275 | 27·769 | 8·258 | 17·401 | 467 | 286 | 597 |
| Larnaca | Softades | 13·095 | 7·250 | 27·217 | 7·961 | 16·703 | 1865 | 1123 | 2390 |
| Larnaca | Tersefanou | 13·302 | 7·193 | 27·431 | 8·184 | 16·862 | 228 | 140 | 289 |
| Larnaca | Tochni | 13·641 | 7·253 | 28·078 | 8·252 | 17·380 | 584 | 353 | 745 |
| Larnaca | Tremetousia | 12·880 | 7·221 | 27·988 | 7·755 | 16·223 | 566 | 341 | 714 |
| Larnaca | Troulloi | 13·118 | 7·246 | 27·611 | 7·945 | 16·599 | 150 | 91 | 190 |
| Larnaca | Vavatsinia | 13·601 | 7·137 | 27·282 | 8·338 | 17·291 | 76 | 46 | 97 |
| Larnaca | Vavla | 13·630 | 7·253 | 27·479 | 8·233 | 17·468 | 280 | 168 | 359 |
| Larnaca | Voroklini | 12·581 | 7·058 | 27·413 | 7·446 | 15·895 | 1272 | 753 | 1607 |
| Larnaca | Zygi | 13·487 | 7·144 | 28·270 | 8·196 | 17·186 | 358 | 217 | 456 |
| Limassol | Agios Amvrosios Lemesou | 14·300 | 7·618 | 29·224 | 8·863 | 17·610 | 24 | 15 | 30 |
| Limassol | Agios Athanasios | 14·153 | 7·562 | 28·498 | 8·699 | 18·022 | 15644 | 9497 | 19745 |
| Limassol | Agios Dimitrios | 14·000 | 7·150 | 27·226 | 8·961 | 17·825 | 47 | 30 | 60 |
| Limassol | Agios Georgios Lemesou | 14·507 | 7·808 | 29·632 | 8·837 | 18·492 | 297 | 181 | 379 |
| Limassol | Agios Ioannis Lemesou | 13·900 | 7·471 | 28·680 | 8·419 | 17·558 | 524 | 317 | 663 |
| Limassol | Agios Konstantinos | 14·286 | 7·370 | 28·985 | 8·821 | 17·972 | 40 | 25 | 50 |
| Limassol | Agios Mamas | 14·112 | 7·415 | 27·895 | 8·633 | 18·299 | 36 | 22 | 47 |
| Limassol | Agios Pavlos | 13·379 | 7·495 | 28·978 | 8·104 | 16·469 | 117 | 71 | 144 |
| Limassol | Agios Theodoros Lemesou | 13·800 | 7·284 | 28·506 | 8·496 | 17·331 | 438 | 271 | 551 |
| Limassol | Agios Therapon | 14·042 | 7·372 | 28·624 | 8·690 | 17·872 | 40 | 25 | 51 |
| Limassol | Agios Thomas | 13·987 | 7·456 | 28·243 | 8·564 | 17·827 | 153 | 94 | 194 |
| Limassol | Agios Tychon | 13·837 | 7·519 | 27·807 | 8·419 | 17·844 | 1283 | 778 | 1660 |
| Limassol | Agridia | 14·124 | 7·623 | 29·700 | 8·479 | 17·920 | 228 | 137 | 289 |
| Limassol | Agros | 13·943 | 7·330 | 28·490 | 8·436 | 17·898 | 140 | 85 | 180 |
| Limassol | Akapnou | 13·862 | 7·480 | 28·459 | 8·537 | 17·540 | 118 | 72 | 149 |
| Limassol | Akrounta | 13·900 | 7·459 | 29·217 | 8·555 | 17·511 | 1128 | 699 | 1426 |
| Limassol | Alassa | 14·542 | 7·934 | 30·272 | 8·715 | 18·591 | 503 | 301 | 644 |
| Limassol | Alektora | 13·813 | 7·693 | 29·361 | 8·025 | 17·979 | 156 | 91 | 203 |
| Limassol | Amiantos | 14·067 | 7·600 | 29·486 | 8·690 | 18·056 | 237 | 148 | 306 |
| Limassol | Anogyra | 14·339 | 7·609 | 29·162 | 8·712 | 18·386 | 49 | 30 | 63 |
| Limassol | Apesia | 14·309 | 7·719 | 29·445 | 8·663 | 18·519 | 1412 | 864 | 1808 |
| Limassol | Apsiou | 14·207 | 7·270 | 27·033 | 8·651 | 18·349 | 61 | 37 | 78 |
| Limassol | Arakapas | 14·162 | 7·570 | 29·717 | 8·769 | 17·847 | 110 | 68 | 138 |
| Limassol | Armenochori | 13·755 | 7·309 | 27·528 | 8·537 | 17·391 | 527 | 327 | 666 |
| Limassol | Arsos Lemesou | 13·687 | 7·361 | 27·269 | 8·355 | 17·213 | 38 | 23 | 47 |

| | | β-thalassaemia carrier prevalence | | | | | Estimated carrier counts | | |
|---|---|---|---|---|---|---|---|---|---|
| **ADM1** | **ADM2** | **Mean** | **SD** | **CI95%** | **Q25** | **Q75** | **Mean** | **Q25** | **Q75** |
| Limassol | Asgata | 13·875 | 7·565 | 28·641 | 8·440 | 17·714 | 399 | 240 | 511 |
| Limassol | Avdimou | 14·115 | 7·524 | 28·650 | 8·589 | 17·891 | 92 | 56 | 117 |
| Limassol | Dierona | 13·827 | 7·056 | 26·934 | 8·647 | 17·511 | 9 | 6 | 12 |
| Limassol | Dora | 14·142 | 7·663 | 30·313 | 8·356 | 17·996 | 20 | 12 | 25 |
| Limassol | Doros | 14·414 | 7·899 | 30·184 | 8·615 | 18·581 | 453 | 270 | 583 |
| Limassol | Dymes | 14·039 | 7·524 | 29·318 | 8·638 | 17·980 | 454 | 279 | 581 |
| Limassol | Episkopi Lemesou | 14·655 | 7·568 | 29·290 | 9·036 | 19·017 | 1082 | 667 | 1404 |
| Limassol | Eptagoneia | 14·039 | 7·770 | 30·449 | 8·718 | 17·721 | 70 | 43 | 88 |
| Limassol | Erimi | 14·648 | 7·581 | 29·802 | 9·115 | 18·851 | 4146 | 2591 | 5315 |
| Limassol | Fasoula Lemesou | 13·881 | 7·371 | 27·489 | 8·604 | 18·006 | 1463 | 907 | 1897 |
| Limassol | Foini | 13·817 | 7·136 | 26·408 | 8·502 | 18·073 | 32 | 20 | 42 |
| Limassol | Foinikaria | 13·864 | 7·422 | 28·795 | 8·551 | 17·481 | 1655 | 1026 | 2092 |
| Limassol | Gerasa | 13·891 | 7·444 | 27·873 | 8·509 | 18·302 | 85 | 52 | 112 |
| Limassol | Germasogeia | 14·086 | 7·402 | 29·086 | 8·731 | 17·826 | 1081 | 670 | 1367 |
| Limassol | Gerovasa | 14·174 | 7·778 | 30·587 | 8·404 | 18·041 | 71 | 43 | 90 |
| Limassol | Kalo Chorio Lemesou | 13·957 | 7·345 | 27·893 | 8·565 | 17·758 | 351 | 215 | 446 |
| Limassol | Kaminaria | 13·974 | 7·286 | 28·361 | 8·605 | 17·821 | 54 | 34 | 69 |
| Limassol | Kantou | 14·655 | 7·568 | 29·290 | 9·036 | 19·017 | 1082 | 667 | 1404 |
| Limassol | Kapileio | 14·110 | 7·495 | 28·126 | 8·637 | 18·280 | 276 | 169 | 359 |
| Limassol | Kato Kivides | 14·345 | 7·587 | 29·210 | 8·784 | 18·284 | 373 | 227 | 476 |
| Limassol | Kato Mylos | 14·139 | 7·529 | 28·626 | 8·568 | 17·932 | 267 | 161 | 339 |
| Limassol | Kato Platres | 14·103 | 7·351 | 27·614 | 8·745 | 17·964 | 198 | 124 | 251 |
| Limassol | Kato Polemidia | 14·657 | 7·958 | 29·665 | 8·788 | 18·886 | 8223 | 4931 | 10596 |
| Limassol | Kellaki | 14·208 | 7·527 | 29·103 | 9·014 | 17·762 | 67 | 43 | 84 |
| Limassol | Kissousa | 14·153 | 7·686 | 29·993 | 8·557 | 17·993 | 195 | 120 | 248 |
| Limassol | Klonari | 13·968 | 7·519 | 28·895 | 8·653 | 17·576 | 153 | 95 | 193 |
| Limassol | Koilani | 14·178 | 7·430 | 28·353 | 8·808 | 17·897 | 206 | 129 | 260 |
| Limassol | Kolossi | 14·648 | 7·581 | 29·802 | 9·115 | 18·851 | 4146 | 2591 | 5315 |
| Limassol | Korfi | 14·263 | 7·773 | 29·536 | 8·576 | 18·556 | 407 | 243 | 527 |
| Limassol | Kouka | 14·257 | 7·690 | 29·493 | 8·600 | 18·082 | 122 | 73 | 154 |
| Limassol | Kyperounta | 13·878 | 7·353 | 28·263 | 8·594 | 17·653 | 534 | 329 | 681 |
| Limassol | Laneia | 14·250 | 7·654 | 29·098 | 8·652 | 18·333 | 405 | 245 | 519 |
| Limassol | Lemesos | 14·492 | 7·914 | 28·920 | 8·762 | 18·233 | 13100 | 7920 | 16481 |
| Limassol | Lemithou | 13·976 | 7·167 | 27·681 | 8·750 | 17·737 | 133 | 84 | 169 |
| Limassol | Limnatis | 14·285 | 7·688 | 29·183 | 8·748 | 18·577 | 116 | 71 | 150 |
| Limassol | Lofou | 14·290 | 7·557 | 28·782 | 8·809 | 18·165 | 427 | 263 | 544 |
| Limassol | Louvaras | 13·826 | 7·316 | 27·704 | 8·478 | 17·658 | 272 | 166 | 346 |
| Limassol | Malia | 14·206 | 7·893 | 30·860 | 8·453 | 18·086 | 51 | 31 | 65 |
| Limassol | Mandria Lemesou | 14·246 | 7·459 | 28·218 | 8·866 | 17·909 | 166 | 104 | 209 |
| Limassol | Mathikoloni | 14·000 | 7·275 | 27·635 | 8·658 | 17·923 | 2614 | 1620 | 3354 |
| Limassol | Mesa Geitonia | 14·187 | 7·643 | 28·204 | 8·683 | 18·119 | 14563 | 8827 | 18378 |
| Limassol | Monagri | 14·346 | 7·823 | 29·819 | 8·625 | 18·376 | 131 | 79 | 168 |
| Limassol | Monagroulli | 13·909 | 7·759 | 29·226 | 8·294 | 17·760 | 250 | 149 | 320 |
| Limassol | Moni | 13·913 | 7·744 | 28·657 | 8·297 | 18·029 | 1006 | 600 | 1314 |
| Limassol | Moniatis | 14·257 | 7·690 | 29·493 | 8·600 | 18·082 | 122 | 73 | 154 |
| Limassol | Mouttagiaka | 13·921 | 7·355 | 28·307 | 8·634 | 17·609 | 1608 | 997 | 2033 |
| Limassol | Omodos | 14·388 | 7·679 | 29·193 | 8·863 | 18·015 | 76 | 47 | 95 |
| Limassol | Pachna | 14·218 | 7·912 | 31·513 | 8·594 | 18·303 | 64 | 39 | 83 |
| Limassol | Palaiomylos | 13·908 | 7·143 | 26·817 | 8·732 | 17·949 | 79 | 50 | 102 |
| Limassol | Palodeia | 14·447 | 7·558 | 29·171 | 8·924 | 18·409 | 1005 | 621 | 1281 |
| Limassol | Pano Kivides | 14·222 | 7·528 | 29·136 | 8·722 | 17·951 | 231 | 140 | 295 |
| Limassol | Pano Platres | 14·179 | 7·775 | 30·036 | 8·584 | 18·073 | 11 | 6 | 14 |
| Limassol | Pano Polemidia | 14·552 | 7·758 | 29·418 | 8·856 | 18·647 | 9228 | 5552 | 11877 |
| Limassol | Paramali | 14·207 | 7·571 | 28·937 | 8·726 | 17·750 | 116 | 71 | 147 |
| Limassol | Paramytha | 14·316 | 7·729 | 29·532 | 8·634 | 18·500 | 1296 | 793 | 1658 |
| Limassol | Parekklisia | 13·918 | 7·728 | 28·087 | 8·301 | 18·297 | 756 | 451 | 994 |
| Limassol | Pelendri | 13·955 | 7·425 | 28·936 | 8·796 | 18·040 | 226 | 142 | 292 |

| | | β-thalassaemia carrier prevalence | | | | | Estimated carrier counts | | |
|---|---|---|---|---|---|---|---|---|---|
| ADM1 | ADM2 | Mean | SD | CI95% | Q25 | Q75 | Mean | Q25 | Q75 |
| Limassol | Pentakomo | 14·034 | 7·589 | 28·778 | 8·668 | 17·695 | 70 | 43 | 89 |
| Limassol | Pera Pedi | 14·104 | 7·239 | 27·242 | 8·870 | 17·802 | 90 | 57 | 114 |
| Limassol | Pissouri | 13·635 | 7·302 | 27·324 | 8·416 | 17·268 | 104 | 64 | 131 |
| Limassol | Platanisteia | 13·878 | 7·484 | 28·459 | 8·413 | 17·800 | 313 | 188 | 403 |
| Limassol | Potamiou | 14·244 | 7·579 | 29·160 | 8·762 | 17·958 | 220 | 136 | 278 |
| Limassol | Potamitissa | 14·139 | 7·529 | 28·626 | 8·568 | 17·932 | 267 | 161 | 339 |
| Limassol | Prastio Avdimou | 14·211 | 7·685 | 29·795 | 8·682 | 17·935 | 180 | 110 | 230 |
| Limassol | Prastio Kellakiou | 13·786 | 7·920 | 31·633 | 8·289 | 17·196 | 38 | 23 | 47 |
| Limassol | Prodromos | 13·887 | 7·229 | 27·315 | 8·658 | 17·758 | 170 | 107 | 217 |
| Limassol | Pyrgos Lemesou | 13·994 | 7·666 | 28·637 | 8·536 | 17·899 | 1087 | 652 | 1416 |
| Limassol | Sanida | 14·074 | 7·587 | 28·617 | 8·774 | 17·769 | 81 | 52 | 102 |
| Limassol | Silikou | 14·187 | 7·531 | 28·794 | 8·696 | 18·033 | 383 | 234 | 487 |
| Limassol | Sotira Lemesou | 14·426 | 7·593 | 29·358 | 8·837 | 18·333 | 1273 | 782 | 1648 |
| Limassol | Souni Zanakia | 14·496 | 7·694 | 29·503 | 8·831 | 18·490 | 333 | 202 | 425 |
| Limassol | Spitali | 14·169 | 7·501 | 28·522 | 8·716 | 18·355 | 1090 | 673 | 1393 |
| Limassol | Sykopetra | 13·927 | 7·391 | 28·904 | 8·669 | 17·597 | 343 | 215 | 433 |
| Limassol | Trachoni Lemesou | 14·641 | 7·594 | 30·314 | 9·195 | 18·685 | 3064 | 1924 | 3911 |
| Limassol | Treis Elies | 13·974 | 7·286 | 28·361 | 8·605 | 17·821 | 54 | 34 | 69 |
| Limassol | Trimiklini | 14·184 | 7·553 | 28·694 | 8·617 | 18·190 | 158 | 95 | 201 |
| Limassol | Tserkezoi | 14·657 | 7·958 | 29·665 | 8·788 | 18·886 | 8223 | 4931 | 10596 |
| Limassol | Vasa Kellakiou | 13·939 | 7·648 | 28·131 | 8·535 | 17·777 | 14 | 9 | 18 |
| Limassol | Vasa Koilaniou | 14·095 | 7·603 | 29·112 | 8·583 | 17·804 | 309 | 190 | 390 |
| Limassol | Vikla | 13·949 | 7·492 | 28·620 | 8·656 | 17·595 | 185 | 115 | 233 |
| Limassol | Vouni | 14·101 | 7·478 | 29·127 | 8·661 | 17·900 | 144 | 89 | 183 |
| Limassol | Ypsonas | 14·626 | 7·890 | 30·933 | 8·833 | 18·737 | 3270 | 2044 | 4176 |
| Limassol | Zoopigi | 14·154 | 7·435 | 27·553 | 8·657 | 17·943 | 39 | 24 | 50 |
| Nicosia | Agia Eirini Lefkosias | 13·716 | 7·182 | 27·209 | 8·550 | 17·326 | 80 | 50 | 100 |
| Nicosia | Agia Kepir | 12·499 | 6·933 | 26·259 | 7·476 | 16·058 | 219 | 131 | 282 |
| Nicosia | Agia Marina Skyllouras | 13·084 | 7·322 | 27·246 | 7·919 | 16·380 | 28 | 17 | 35 |
| Nicosia | Agia Marina Xyliatou | 13·474 | 7·050 | 27·296 | 8·416 | 17·039 | 223 | 141 | 284 |
| Nicosia | Agia Varvara | 13·048 | 7·036 | 26·924 | 8·040 | 16·756 | 202 | 125 | 260 |
| Nicosia | Agia Varvara Lefkosias | 12·895 | 6·819 | 27·170 | 8·016 | 16·249 | 346 | 215 | 436 |
| Nicosia | Agioi Iliofotoi | 13·335 | 7·012 | 26·946 | 8·291 | 16·647 | 347 | 216 | 433 |
| Nicosia | Agioi Trimithias | 13·270 | 6·786 | 25·844 | 8·213 | 16·849 | 356 | 220 | 451 |
| Nicosia | Agios Dometios | 12·904 | 7·021 | 26·767 | 7·935 | 16·272 | 22075 | 13532 | 27797 |
| Nicosia | Agios Epifanios Oreinis | 13·549 | 7·093 | 27·327 | 8·446 | 16·985 | 296 | 187 | 371 |
| Nicosia | Agios Epifanios Soleas | 13·859 | 7·277 | 27·266 | 8·497 | 17·764 | 1189 | 732 | 1519 |
| Nicosia | Agios Georgios Kafkaliou | 14·043 | 7·026 | 27·378 | 8·917 | 17·676 | 10 | 6 | 13 |
| Nicosia | Agios Georgios Soleas | 13·805 | 7·509 | 27·892 | 8·339 | 17·620 | 1364 | 829 | 1743 |
| Nicosia | Agios Ioannis Malountas | 13·310 | 7·052 | 27·089 | 8·105 | 16·822 | 1232 | 751 | 1562 |
| Nicosia | Agios Ioannis Selemani | 13·468 | 7·275 | 27·031 | 8·177 | 17·433 | 180 | 110 | 233 |
| Nicosia | Agios Nikolaos Soleas | 13·884 | 7·411 | 27·441 | 8·443 | 17·709 | 855 | 526 | 1093 |
| Nicosia | Agios Sozomenos | 13·120 | 6·820 | 27·003 | 8·027 | 16·802 | 846 | 515 | 1086 |
| Nicosia | Agios Theodoros Soleas | 13·639 | 7·054 | 26·789 | 8·439 | 17·371 | 620 | 383 | 789 |
| Nicosia | Agios Theodoros Tillrias | 13·212 | 6·994 | 26·035 | 8·058 | 16·893 | 11 | 7 | 14 |
| Nicosia | Agios Vasileios | 12·862 | 7·048 | 26·393 | 7·843 | 16·335 | 316 | 191 | 401 |
| Nicosia | Aglantzia | 12·999 | 6·971 | 25·783 | 7·998 | 16·399 | 5253 | 3198 | 6596 |
| Nicosia | Agrokipia | 13·359 | 7·101 | 27·295 | 8·192 | 16·768 | 1018 | 622 | 1287 |
| Nicosia | Akaki | 13·316 | 7·254 | 27·422 | 8·315 | 16·834 | 478 | 299 | 605 |
| Nicosia | Alampra | 13·146 | 7·158 | 27·540 | 8·221 | 16·708 | 413 | 258 | 525 |
| Nicosia | Alaykoy | 13·039 | 6·913 | 26·187 | 7·901 | 16·643 | 643 | 389 | 820 |
| Nicosia | Alevga | 13·226 | 7·113 | 26·600 | 7·935 | 17·038 | 23 | 14 | 29 |
| Nicosia | Alithinou | 13·822 | 7·233 | 27·944 | 8·461 | 17·569 | 142 | 87 | 183 |
| Nicosia | Alona | 13·768 | 7·135 | 28·030 | 8·530 | 17·441 | 281 | 175 | 357 |
| Nicosia | Ammadies | 13·511 | 7·255 | 26·866 | 8·252 | 17·496 | 180 | 110 | 233 |
| Nicosia | Ampelikou | 13·642 | 7·225 | 28·478 | 8·262 | 17·375 | 51 | 31 | 65 |
| Nicosia | Anageia | 13·162 | 6·867 | 26·112 | 8·171 | 16·634 | 2150 | 1334 | 2714 |

| | | β-thalassaemia carrier prevalence | | | | | Estimated carrier counts | | |
|---|---|---|---|---|---|---|---|---|---|
| ADM1 | ADM2 | Mean | SD | CI95% | Q25 | Q75 | Mean | Q25 | Q75 |
| Nicosia | Analiontas | 13·372 | 7·115 | 26·728 | 8·228 | 16·938 | 1229 | 758 | 1555 |
| Nicosia | Angolemi | 13·799 | 7·265 | 27·468 | 8·472 | 17·698 | 48 | 30 | 62 |
| Nicosia | Apliki | 13·691 | 7·212 | 28·090 | 8·570 | 17·347 | 233 | 147 | 295 |
| Nicosia | Arediou | 13·417 | 7·069 | 27·050 | 8·201 | 16·978 | 1143 | 697 | 1449 |
| Nicosia | Argaki | 13·320 | 7·099 | 26·259 | 8·192 | 16·871 | 2311 | 1419 | 2939 |
| Nicosia | Askas | 13·795 | 7·259 | 28·299 | 8·511 | 17·392 | 284 | 177 | 361 |
| Nicosia | Astromeritis | 13·481 | 7·074 | 26·941 | 8·419 | 17·133 | 2330 | 1435 | 2961 |
| Nicosia | Avlona | 13·268 | 7·070 | 27·068 | 8·337 | 16·934 | 968 | 609 | 1236 |
| Nicosia | Beikioi | 12·314 | 6·804 | 25·534 | 7·347 | 15·724 | 1334 | 801 | 1716 |
| Nicosia | Dali | 13·215 | 7·294 | 28·842 | 7·950 | 17·010 | 1542 | 928 | 1985 |
| Nicosia | Deneia | 13·049 | 7·186 | 26·919 | 7·967 | 16·551 | 1190 | 733 | 1517 |
| Nicosia | Dyo Potamoi | 12·842 | 6·857 | 25·973 | 7·961 | 16·310 | 228 | 141 | 289 |
| Nicosia | Egkomi Lefkosias | 12·935 | 6·959 | 27·076 | 8·127 | 16·101 | 6166 | 3874 | 7675 |
| Nicosia | Elia Lefkosias | 13·648 | 7·706 | 28·794 | 8·131 | 17·441 | 509 | 303 | 650 |
| Nicosia | Epicho | 12·417 | 6·857 | 25·644 | 7·438 | 15·818 | 1786 | 1075 | 2288 |
| Nicosia | Episkopeio | 13·529 | 7·237 | 27·935 | 8·223 | 16·891 | 271 | 164 | 338 |
| Nicosia | Ergates | 13·377 | 7·043 | 26·877 | 8·253 | 16·848 | 1218 | 753 | 1534 |
| Nicosia | Evrychou | 14·031 | 7·385 | 28·435 | 8·646 | 17·889 | 320 | 197 | 408 |
| Nicosia | Exo Metochi | 12·728 | 7·014 | 25·974 | 7·711 | 16·099 | 452 | 274 | 572 |
| Nicosia | Farmakas | 13·716 | 7·288 | 28·208 | 8·500 | 17·431 | 273 | 168 | 346 |
| Nicosia | Fikardou | 13·534 | 6·970 | 26·444 | 8·371 | 17·288 | 83 | 51 | 106 |
| Nicosia | Flasou | 13·859 | 7·277 | 27·266 | 8·497 | 17·764 | 1189 | 732 | 1519 |
| Nicosia | Frodisia | 13·555 | 7·404 | 27·883 | 8·124 | 17·366 | 2 | 1 | 3 |
| Nicosia | Fterikoudi | 13·848 | 7·505 | 28·837 | 8·472 | 17·296 | 3 | 2 | 4 |
| Nicosia | Fyllia | 13·163 | 7·105 | 26·875 | 8·166 | 16·701 | 1156 | 725 | 1474 |
| Nicosia | Galata | 13·710 | 7·374 | 28·079 | 8·365 | 17·363 | 257 | 159 | 327 |
| Nicosia | Galini | 13·548 | 7·261 | 27·392 | 8·227 | 17·289 | 112 | 67 | 142 |
| Nicosia | Gerakies | 14·080 | 7·402 | 29·730 | 8·479 | 17·749 | 4 | 2 | 5 |
| Nicosia | Geri | 12·989 | 6·801 | 26·372 | 8·067 | 16·415 | 3491 | 2155 | 4404 |
| Nicosia | Gourri | 13·499 | 6·863 | 26·574 | 8·403 | 17·048 | 10 | 6 | 12 |
| Nicosia | Kakopetria | 13·533 | 7·068 | 26·620 | 8·368 | 17·230 | 248 | 154 | 316 |
| Nicosia | Kalapanagiotis | 13·776 | 7·011 | 26·358 | 8·696 | 17·611 | 8 | 5 | 11 |
| Nicosia | Kaliana | 13·772 | 7·259 | 27·585 | 8·457 | 17·591 | 591 | 365 | 753 |
| Nicosia | Kalo Chorio Kapouti | 13·060 | 6·955 | 26·181 | 8·034 | 16·648 | 1698 | 1060 | 2164 |
| Nicosia | Kalo Chorio Oreinis | 13·498 | 7·043 | 26·919 | 8·366 | 17·090 | 842 | 517 | 1070 |
| Nicosia | Kalo Chorio Soleas | 13·884 | 7·411 | 27·441 | 8·443 | 17·709 | 855 | 526 | 1093 |
| Nicosia | Kalyvakia | 12·134 | 6·753 | 24·816 | 7·230 | 15·398 | 49 | 29 | 62 |
| Nicosia | Kampi | 13·789 | 7·484 | 28·611 | 8·516 | 17·710 | 92 | 57 | 118 |
| Nicosia | Kampia | 13·207 | 6·590 | 25·436 | 8·507 | 16·643 | 17 | 11 | 21 |
| Nicosia | Kampos | 13·702 | 7·183 | 27·294 | 8·397 | 17·381 | 33 | 20 | 41 |
| Nicosia | Kanli | 12·811 | 7·031 | 26·223 | 7·694 | 16·403 | 508 | 305 | 651 |
| Nicosia | Kannavia | 13·790 | 7·287 | 27·258 | 8·655 | 17·479 | 35 | 22 | 44 |
| Nicosia | Kapedes | 13·511 | 7·096 | 25·833 | 8·476 | 17·013 | 158 | 99 | 199 |
| Nicosia | Kataliontas | 13·227 | 7·176 | 27·768 | 8·065 | 16·694 | 283 | 172 | 357 |
| Nicosia | Kato Deftera | 12·878 | 6·841 | 25·766 | 7·931 | 16·150 | 6579 | 4039 | 8222 |
| Nicosia | Kato Koutrafas | 13·732 | 7·225 | 26·265 | 8·443 | 17·560 | 108 | 66 | 138 |
| Nicosia | Kato Moni | 13·343 | 6·850 | 26·610 | 8·387 | 16·766 | 50 | 32 | 63 |
| Nicosia | Kato Pyrgos | 13·511 | 7·255 | 26·866 | 8·252 | 17·496 | 180 | 110 | 233 |
| Nicosia | Kato Zodeia | 13·433 | 6·997 | 26·970 | 8·266 | 17·136 | 807 | 503 | 1024 |
| Nicosia | Katokopia | 13·207 | 7·053 | 26·531 | 8·130 | 16·779 | 1938 | 1179 | 2466 |
| Nicosia | Katydata | 13·884 | 7·411 | 27·441 | 8·443 | 17·709 | 855 | 526 | 1093 |
| Nicosia | Katydata | 13·946 | 7·610 | 29·209 | 8·332 | 17·703 | 143 | 85 | 181 |
| Nicosia | Kazivera | 13·543 | 7·498 | 28·159 | 8·018 | 17·414 | 777 | 460 | 996 |
| Nicosia | Kioneli | 13·075 | 7·237 | 27·461 | 7·979 | 16·692 | 3427 | 2091 | 4374 |
| Nicosia | Klirou | 13·452 | 7·184 | 27·370 | 8·169 | 17·193 | 516 | 313 | 660 |
| Nicosia | Kokkina | 13·226 | 7·113 | 26·600 | 7·935 | 17·038 | 23 | 14 | 29 |
| Nicosia | Kokkinotrimithia | 13·092 | 6·929 | 26·394 | 8·029 | 16·921 | 1294 | 793 | 1673 |

| | | β-thalassaemia carrier prevalence | | | | | Estimated carrier counts | | |
|---|---|---|---|---|---|---|---|---|---|
| **ADM1** | **ADM2** | **Mean** | **SD** | **CI95%** | **Q25** | **Q75** | **Mean** | **Q25** | **Q75** |
| Nicosia | Korakou | 13·638 | 6·903 | 25·745 | 8·454 | 17·750 | 14 | 9 | 18 |
| Nicosia | Kotsiatis | 13·148 | 7·029 | 27·081 | 8·042 | 16·735 | 1028 | 630 | 1307 |
| Nicosia | Kourou Monastiri | 12·361 | 6·753 | 25·848 | 7·585 | 15·579 | 688 | 421 | 869 |
| Nicosia | Kyra | 13·004 | 6·922 | 25·907 | 8·043 | 16·406 | 352 | 218 | 444 |
| Nicosia | Kythrea | 12·269 | 6·923 | 26·481 | 7·233 | 15·552 | 307 | 181 | 389 |
| Nicosia | Lagoudera | 13·701 | 7·136 | 27·398 | 8·487 | 17·241 | 2 | 2 | 3 |
| Nicosia | Lakatameia | 12·871 | 6·972 | 25·892 | 7·886 | 16·050 | 5376 | 3294 | 6704 |
| Nicosia | Latsia | 12·973 | 6·821 | 26·280 | 8·084 | 16·568 | 2291 | 1428 | 2926 |
| Nicosia | Lazanias | 13·616 | 7·209 | 28·164 | 8·258 | 17·482 | 8 | 5 | 11 |
| Nicosia | Lefka | 13·822 | 7·212 | 25·673 | 8·554 | 17·715 | 712 | 441 | 912 |
| Nicosia | Lefkosia | 12·700 | 6·866 | 25·764 | 7·699 | 16·024 | 12482 | 7567 | 15748 |
| Nicosia | Linou | 13·988 | 7·497 | 28·822 | 8·489 | 17·796 | 463 | 282 | 589 |
| Nicosia | Livadia Lefosias | 13·890 | 7·344 | 28·211 | 8·514 | 17·634 | 405 | 246 | 516 |
| Nicosia | Louroukina | 13·120 | 7·316 | 26·872 | 7·847 | 16·628 | 81 | 49 | 103 |
| Nicosia | Loutros | 13·411 | 7·509 | 27·798 | 8·019 | 17·148 | 57 | 34 | 72 |
| Nicosia | Lympia | 13·239 | 7·486 | 29·089 | 7·928 | 16·562 | 621 | 369 | 773 |
| Nicosia | Lythrodontas | 13·183 | 6·974 | 26·783 | 8·047 | 16·671 | 319 | 194 | 403 |
| Nicosia | Malounta | 13·491 | 7·211 | 27·653 | 8·196 | 17·042 | 787 | 477 | 998 |
| Nicosia | Mammari | 12·977 | 6·988 | 26·393 | 7·936 | 16·628 | 1610 | 984 | 2074 |
| Nicosia | Mandres Lefkosias | 12·685 | 6·602 | 25·814 | 7·980 | 16·107 | 2015 | 1267 | 2558 |
| Nicosia | Mansoura | 13·226 | 7·113 | 26·600 | 7·935 | 17·038 | 23 | 14 | 29 |
| Nicosia | Margi | 13·232 | 7·099 | 27·051 | 8·051 | 16·897 | 682 | 415 | 871 |
| Nicosia | Margo | 13·130 | 7·146 | 27·322 | 8·037 | 16·588 | 23 | 14 | 29 |
| Nicosia | Masari | 13·076 | 6·910 | 26·176 | 8·148 | 16·615 | 842 | 528 | 1075 |
| Nicosia | Mathiatis | 13·251 | 6·932 | 26·378 | 8·179 | 16·859 | 854 | 525 | 1083 |
| Nicosia | Meniko | 13·194 | 7·203 | 27·560 | 7·926 | 16·539 | 155 | 93 | 194 |
| Nicosia | Mia Milia | 12·591 | 6·933 | 26·510 | 7·652 | 15·877 | 2768 | 1710 | 3518 |
| Nicosia | Mitsero | 13·467 | 6·982 | 26·667 | 8·559 | 16·635 | 142 | 91 | 176 |
| Nicosia | Mora | 12·709 | 6·931 | 26·167 | 7·478 | 16·556 | 163 | 96 | 213 |
| Nicosia | Morfou | 13·368 | 7·158 | 26·837 | 8·324 | 16·946 | 1150 | 726 | 1466 |
| Nicosia | Moutoulias | 13·866 | 7·315 | 27·813 | 8·585 | 17·567 | 91 | 57 | 115 |
| Nicosia | Mylikouri | 13·698 | 7·223 | 27·681 | 8·418 | 17·357 | 8 | 5 | 10 |
| Nicosia | Neo Chorio Lefkosias | 12·707 | 7·175 | 25·406 | 7·465 | 16·261 | 683 | 401 | 874 |
| Nicosia | Nikitari | 13·351 | 6·683 | 24·941 | 8·297 | 17·191 | 7 | 4 | 9 |
| Nicosia | Nikitas | 13·396 | 7·338 | 27·857 | 8·033 | 17·095 | 395 | 234 | 505 |
| Nicosia | Nisou | 13·095 | 7·077 | 27·397 | 8·074 | 16·761 | 4448 | 2739 | 5696 |
| Nicosia | Oikos | 13·836 | 7·213 | 27·328 | 8·622 | 17·582 | 99 | 62 | 126 |
| Nicosia | Orounta | 13·549 | 7·300 | 27·514 | 8·392 | 17·104 | 136 | 84 | 172 |
| Nicosia | Ortakioi | 12·729 | 6·970 | 26·896 | 7·748 | 16·084 | 20912 | 12708 | 26385 |
| Nicosia | Pachyammos | 13·240 | 7·233 | 27·166 | 7·812 | 17·183 | 12 | 7 | 15 |
| Nicosia | Palaichori Morfou | 13·592 | 6·940 | 27·570 | 8·624 | 16·983 | 141 | 90 | 177 |
| Nicosia | Palaichori Oreinis | 13·538 | 7·035 | 27·196 | 8·364 | 17·244 | 101 | 62 | 128 |
| Nicosia | Palaikythro | 12·537 | 6·737 | 25·304 | 7·578 | 16·221 | 978 | 591 | 1265 |
| Nicosia | Palaiometocho | 13·324 | 7·034 | 27·584 | 8·113 | 16·706 | 205 | 125 | 257 |
| Nicosia | Pano Deftera | 12·884 | 6·711 | 25·641 | 7·976 | 16·249 | 1203 | 745 | 1518 |
| Nicosia | Pano Koutrafas | 13·580 | 7·116 | 26·549 | 8·280 | 17·459 | 431 | 257 | 555 |
| Nicosia | Pano Pyrgos | 13·380 | 7·181 | 26·910 | 8·319 | 17·256 | 103 | 64 | 133 |
| Nicosia | Pano Zodeia | 13·601 | 7·480 | 27·597 | 8·074 | 17·268 | 1096 | 651 | 1391 |
| Nicosia | Pedoulas | 13·850 | 7·255 | 27·344 | 8·558 | 17·736 | 123 | 77 | 157 |
| Nicosia | Pentageia | 13·735 | 7·459 | 27·234 | 8·343 | 17·578 | 1221 | 744 | 1562 |
| Nicosia | Pera | 13·375 | 7·094 | 26·656 | 8·186 | 17·022 | 788 | 487 | 999 |
| Nicosia | Pera Chorio | 13·136 | 7·163 | 27·769 | 8·070 | 16·825 | 2157 | 1311 | 2770 |
| Nicosia | Peristerona Lefkosias | 13·220 | 6·886 | 26·714 | 8·359 | 17·034 | 490 | 310 | 631 |
| Nicosia | Peristeronari | 13·822 | 7·212 | 25·673 | 8·554 | 17·715 | 712 | 441 | 912 |
| Nicosia | Petra | 13·798 | 7·527 | 28·491 | 8·312 | 17·614 | 700 | 418 | 893 |
| Nicosia | Petra tou Digeni | 12·291 | 6·807 | 25·781 | 7·411 | 15·462 | 918 | 557 | 1156 |
| Nicosia | Pigenia | 13·296 | 7·088 | 26·473 | 8·188 | 17·075 | 114 | 71 | 147 |

| | | β-thalassaemia carrier prevalence | | | | | Estimated carrier counts | | |
|---|---|---|---|---|---|---|---|---|---|
| **ADM1** | **ADM2** | **Mean** | **SD** | **CI95%** | **Q25** | **Q75** | **Mean** | **Q25** | **Q75** |
| Nicosia | Platanistasa | 13·709 | 7·205 | 27·833 | 8·445 | 17·300 | 195 | 121 | 250 |
| Nicosia | Politiko | 13·400 | 6·975 | 26·667 | 8·357 | 16·906 | 962 | 595 | 1210 |
| Nicosia | Polystypos | 14·034 | 7·477 | 29·095 | 8·457 | 17·909 | 368 | 222 | 469 |
| Nicosia | Potami | 13·422 | 7·073 | 27·710 | 8·593 | 16·919 | 97 | 62 | 123 |
| Nicosia | Potamia | 13·206 | 6·994 | 27·163 | 7·899 | 17·095 | 513 | 307 | 664 |
| Nicosia | Prastio Lefkosias | 13·480 | 7·460 | 28·169 | 8·065 | 17·210 | 904 | 537 | 1155 |
| Nicosia | Psimolofou | 13·127 | 6·968 | 26·211 | 8·059 | 16·586 | 7367 | 4526 | 9221 |
| Nicosia | Pyrogi | 13·095 | 7·036 | 27·012 | 8·013 | 16·575 | 437 | 271 | 554 |
| Nicosia | Saranti | 13·746 | 7·212 | 27·328 | 8·571 | 17·360 | 37 | 24 | 47 |
| Nicosia | Selladi tou Appi | 13·212 | 6·994 | 26·035 | 8·058 | 16·893 | 11 | 7 | 14 |
| Nicosia | Sinaoros | 13·782 | 7·227 | 27·527 | 8·507 | 17·559 | 568 | 351 | 724 |
| Nicosia | Skouriotissa | 13·884 | 7·411 | 27·441 | 8·443 | 17·709 | 855 | 526 | 1093 |
| Nicosia | Skylloura | 12·940 | 7·148 | 27·049 | 7·857 | 16·313 | 447 | 269 | 564 |
| Nicosia | Spilia | 13·642 | 7·077 | 27·160 | 8·445 | 17·173 | 45 | 28 | 56 |
| Nicosia | Strovolos | 12·885 | 6·915 | 26·183 | 7·955 | 16·213 | 29473 | 18110 | 37035 |
| Nicosia | Syrianochori | 13·354 | 7·386 | 28·190 | 8·160 | 16·803 | 127 | 77 | 159 |
| Nicosia | Temvria | 13·835 | 7·144 | 27·090 | 8·550 | 17·819 | 334 | 206 | 426 |
| Nicosia | Trachonas | 12·412 | 6·807 | 27·462 | 7·567 | 15·537 | 5003 | 3050 | 6263 |
| Nicosia | Trachoni Lefkosias | 12·537 | 6·737 | 25·304 | 7·578 | 16·221 | 978 | 591 | 1265 |
| Nicosia | Tsakistra | 13·794 | 7·274 | 28·037 | 8·433 | 17·386 | 38 | 23 | 47 |
| Nicosia | Tseri | 13·078 | 6·978 | 26·389 | 8·014 | 16·654 | 8066 | 4965 | 10144 |
| Nicosia | Tymvou | 12·731 | 7·387 | 29·060 | 7·401 | 16·145 | 134 | 78 | 170 |
| Nicosia | Variseia | 13·444 | 7·241 | 27·055 | 8·188 | 17·281 | 75 | 44 | 95 |
| Nicosia | Voni | 12·633 | 6·876 | 25·639 | 7·644 | 16·160 | 1430 | 865 | 1837 |
| Nicosia | Vyzakia | 13·657 | 7·116 | 27·205 | 8·537 | 17·346 | 289 | 179 | 369 |
| Nicosia | Xerovounos | 13·485 | 7·247 | 27·144 | 8·166 | 17·544 | 91 | 54 | 118 |
| Nicosia | Xyliatos | 13·429 | 7·139 | 27·467 | 8·196 | 17·230 | 74 | 45 | 95 |
| Paphos | Acheleia | 13·702 | 7·794 | 30·227 | 8·161 | 17·362 | 1987 | 1180 | 2521 |
| Paphos | Agia Marina Chrysochous | 13·421 | 7·581 | 28·798 | 7·930 | 16·937 | 236 | 139 | 303 |
| Paphos | Agia Marinouda | 13·702 | 7·794 | 30·227 | 8·161 | 17·362 | 1987 | 1180 | 2521 |
| Paphos | Agia Varvara Pafou | 13·704 | 7·770 | 30·225 | 8·145 | 17·447 | 597 | 354 | 761 |
| Paphos | Agios Dimitrianos | 13·401 | 7·298 | 26·796 | 8·019 | 16·971 | 91 | 55 | 115 |
| Paphos | Agios Georgios | 13·818 | 7·442 | 28·472 | 8·411 | 17·580 | 49 | 30 | 63 |
| Paphos | Agios Ioannis Pafou | 13·761 | 7·503 | 28·481 | 8·261 | 17·612 | 33 | 20 | 42 |
| Paphos | Agios Isidoros | 13·747 | 7·566 | 28·845 | 8·273 | 17·508 | 215 | 129 | 273 |
| Paphos | Agios Nikolaos Pafou | 13·950 | 7·529 | 28·624 | 8·442 | 17·768 | 135 | 83 | 169 |
| Paphos | Akoursos | 13·827 | 7·785 | 30·004 | 8·188 | 17·609 | 854 | 504 | 1092 |
| Paphos | Amargeti | 13·484 | 7·439 | 28·898 | 7·961 | 17·484 | 46 | 27 | 60 |
| Paphos | Anadiou | 13·725 | 7·578 | 28·253 | 8·275 | 17·549 | 87 | 52 | 111 |
| Paphos | Anarita | 13·664 | 7·897 | 30·279 | 7·993 | 17·402 | 243 | 142 | 310 |
| Paphos | Androlikou | 13·697 | 7·940 | 29·982 | 7·882 | 17·726 | 0 | 0 | 1 |
| Paphos | Argaka | 13·648 | 7·615 | 28·961 | 8·138 | 17·362 | 240 | 141 | 309 |
| Paphos | Arminou | 14·291 | 8·103 | 31·182 | 8·525 | 18·159 | 18 | 11 | 23 |
| Paphos | Armou | 13·833 | 7·614 | 29·368 | 8·306 | 17·711 | 169 | 102 | 217 |
| Paphos | Asprogia | 13·873 | 7·581 | 28·941 | 8·437 | 17·623 | 93 | 57 | 119 |
| Paphos | Axylou | 13·601 | 7·752 | 29·545 | 7·974 | 17·258 | 80 | 47 | 102 |
| Paphos | Ayia Marina Kelokedharon | 13·712 | 7·573 | 29·412 | 8·101 | 17·476 | 116 | 68 | 149 |
| Paphos | Chloraka | 13·140 | 7·198 | 27·327 | 7·875 | 16·395 | 3272 | 1961 | 4083 |
| Paphos | Choletria | 13·484 | 7·221 | 27·612 | 8·142 | 17·110 | 108 | 65 | 137 |
| Paphos | Choli | 13·976 | 8·073 | 32·399 | 8·438 | 17·810 | 104 | 63 | 132 |
| Paphos | Choulou | 13·366 | 7·513 | 27·693 | 7·883 | 17·031 | 32 | 19 | 41 |
| Paphos | Chrysochou | 13·976 | 8·073 | 32·399 | 8·438 | 17·810 | 104 | 63 | 132 |
| Paphos | Drouseia | 13·203 | 7·492 | 28·770 | 7·746 | 16·665 | 118 | 70 | 149 |
| Paphos | Drymou | 13·858 | 7·815 | 31·325 | 8·237 | 17·455 | 127 | 76 | 160 |
| Paphos | Eledio | 13·601 | 7·752 | 29·545 | 7·974 | 17·258 | 80 | 47 | 102 |
| Paphos | Empa | 13·322 | 7·313 | 27·784 | 7·983 | 16·862 | 5920 | 3548 | 7482 |
| Paphos | Episkopi Pafou | 13·719 | 8·065 | 30·192 | 7·986 | 17·032 | 34 | 20 | 42 |

| | | β-thalassaemia carrier prevalence | | | | | Estimated carrier counts | | |
|---|---|---|---|---|---|---|---|---|---|
| **ADM1** | **ADM2** | **Mean** | **SD** | **CI95%** | **Q25** | **Q75** | **Mean** | **Q25** | **Q75** |
| Paphos | Evretou | 13·711 | 7·679 | 30·473 | 8·187 | 17·545 | 169 | 101 | 215 |
| Paphos | Faleia | 13·962 | 7·522 | 28·141 | 8·179 | 17·660 | 10 | 6 | 12 |
| Paphos | Fasli | 13·417 | 7·753 | 29·767 | 7·706 | 17·178 | 13 | 7 | 17 |
| Paphos | Fasoula Pafou | 13·437 | 7·188 | 27·664 | 8·091 | 17·284 | 10 | 6 | 12 |
| Paphos | Filousa Chrysochous | 13·445 | 7·285 | 28·546 | 7·937 | 17·280 | 65 | 38 | 83 |
| Paphos | Filousa Kelokedaron | 13·989 | 7·732 | 29·225 | 8·440 | 17·686 | 56 | 34 | 70 |
| Paphos | Foinikas | 13·574 | 7·450 | 28·303 | 8·158 | 17·069 | 305 | 180 | 387 |
| Paphos | Fyti | 13·435 | 7·083 | 25·899 | 8·155 | 16·911 | 59 | 36 | 74 |
| Paphos | Galataria | 13·766 | 7·619 | 29·369 | 8·140 | 17·471 | 68 | 40 | 86 |
| Paphos | Geroskipou | 13·789 | 7·787 | 29·427 | 8·186 | 17·496 | 1802 | 1070 | 2287 |
| Paphos | Gialia | 13·563 | 7·709 | 29·485 | 8·037 | 17·201 | 166 | 97 | 215 |
| Paphos | Giolou | 13·839 | 7·788 | 30·541 | 8·224 | 17·589 | 416 | 248 | 527 |
| Paphos | Goudi | 13·623 | 7·745 | 30·193 | 8·200 | 17·256 | 209 | 126 | 265 |
| Paphos | Ineia | 13·171 | 8·013 | 29·303 | 7·445 | 16·785 | 2 | 1 | 3 |
| Paphos | Kallepeia | 13·833 | 7·576 | 27·169 | 8·284 | 17·777 | 258 | 155 | 332 |
| Paphos | Kannaviou | 14·026 | 7·599 | 28·305 | 8·254 | 18·349 | 26 | 15 | 34 |
| Paphos | Karamoullides | 13·677 | 7·686 | 29·643 | 8·233 | 17·349 | 620 | 373 | 786 |
| Paphos | Kathikas | 13·712 | 7·527 | 29·186 | 8·282 | 17·276 | 77 | 47 | 97 |
| Paphos | Kato Akourdaleia | 14·026 | 8·025 | 31·146 | 8·361 | 17·811 | 224 | 134 | 284 |
| Paphos | Kato Arodes | 13·426 | 7·593 | 28·387 | 7·931 | 17·025 | 218 | 131 | 276 |
| Paphos | Kedares | 14·217 | 7·883 | 30·747 | 8·440 | 18·078 | 38 | 23 | 48 |
| Paphos | Kelokedara | 13·880 | 7·543 | 29·080 | 8·321 | 17·613 | 35 | 21 | 45 |
| Paphos | Kidasi | 13·949 | 7·502 | 29·089 | 8·428 | 17·771 | 34 | 21 | 43 |
| Paphos | Kios | 13·640 | 7·632 | 29·197 | 8·105 | 17·537 | 67 | 39 | 86 |
| Paphos | Kissonerga | 13·383 | 7·504 | 28·702 | 7·919 | 17·127 | 4285 | 2554 | 5398 |
| Paphos | Koili | 13·899 | 7·850 | 31·080 | 8·096 | 17·806 | 364 | 212 | 466 |
| Paphos | Koilineia | 13·848 | 7·484 | 29·044 | 8·388 | 17·611 | 135 | 82 | 171 |
| Paphos | Konia | 13·636 | 7·672 | 29·565 | 8·137 | 17·351 | 4635 | 2767 | 5920 |
| Paphos | Kouklia Pafou | 13·684 | 7·811 | 30·126 | 8·132 | 17·185 | 159 | 94 | 201 |
| Paphos | Kourdaka | 13·366 | 7·513 | 27·693 | 7·883 | 17·031 | 32 | 19 | 41 |
| Paphos | Kritou Marottou | 13·688 | 7·444 | 27·721 | 8·275 | 17·467 | 85 | 51 | 108 |
| Paphos | Kritou Tera | 13·759 | 7·749 | 29·866 | 8·241 | 17·400 | 406 | 244 | 514 |
| Paphos | Kynousa | 13·615 | 7·513 | 28·393 | 8·148 | 17·315 | 74 | 44 | 94 |
| Paphos | Lapithiou | 14·026 | 7·599 | 28·305 | 8·254 | 18·349 | 26 | 15 | 34 |
| Paphos | Lasa | 13·579 | 7·394 | 28·590 | 8·110 | 17·215 | 251 | 150 | 317 |
| Paphos | Lemona | 13·664 | 7·517 | 27·917 | 8·031 | 17·346 | 42 | 25 | 53 |
| Paphos | Lempa | 13·140 | 7·198 | 27·327 | 7·875 | 16·395 | 3272 | 1961 | 4083 |
| Paphos | Letymvou | 13·688 | 7·692 | 28·605 | 8·115 | 17·387 | 493 | 296 | 632 |
| Paphos | Livadi | 13·431 | 7·539 | 28·049 | 7·765 | 16·795 | 0 | 0 | 0 |
| Paphos | Loukrounou | 13·832 | 7·778 | 30·279 | 8·219 | 17·634 | 289 | 172 | 367 |
| Paphos | Lysos | 13·547 | 7·481 | 28·025 | 8·080 | 17·226 | 0 | 0 | 0 |
| Paphos | Makounta | 13·747 | 7·566 | 28·845 | 8·273 | 17·508 | 215 | 129 | 273 |
| Paphos | Mamonia | 13·571 | 7·293 | 27·992 | 8·174 | 17·279 | 153 | 92 | 194 |
| Paphos | Mamountali | 14·018 | 7·405 | 28·349 | 8·569 | 18·120 | 93 | 57 | 119 |
| Paphos | Mandria Pafou | 13·639 | 7·849 | 30·653 | 8·064 | 17·315 | 428 | 252 | 544 |
| Paphos | Marathounta | 13·724 | 7·707 | 30·198 | 8·221 | 17·469 | 354 | 212 | 451 |
| Paphos | Maronas | 13·699 | 7·350 | 28·150 | 8·353 | 17·503 | 63 | 39 | 81 |
| Paphos | Meladeia | 13·445 | 7·285 | 28·546 | 7·937 | 17·280 | 65 | 38 | 83 |
| Paphos | Melandra | 13·640 | 7·632 | 29·197 | 8·105 | 17·537 | 67 | 39 | 86 |
| Paphos | Mesa Chorio | 13·504 | 7·428 | 28·240 | 8·091 | 17·329 | 2648 | 1587 | 3399 |
| Paphos | Mesana | 14·065 | 7·756 | 29·992 | 8·380 | 17·922 | 71 | 43 | 90 |
| Paphos | Mesogi | 13·668 | 7·521 | 28·804 | 8·198 | 17·520 | 2817 | 1689 | 3616 |
| Paphos | Milia Pafou | 13·646 | 7·449 | 28·612 | 8·196 | 17·183 | 186 | 112 | 234 |
| Paphos | Miliou | 14·026 | 8·025 | 31·146 | 8·361 | 17·811 | 224 | 134 | 284 |
| Paphos | Mousere | 14·068 | 7·534 | 29·155 | 8·522 | 17·984 | 73 | 45 | 94 |
| Paphos | Nata | 13·587 | 7·601 | 28·924 | 8·066 | 17·163 | 385 | 227 | 489 |
| Paphos | Nea Dimmata | 13·127 | 7·368 | 28·173 | 7·880 | 16·550 | 70 | 42 | 88 |

| ADM1 | ADM2 | β-thalassaemia carrier prevalence Mean | SD | CI95% | Q25 | Q75 | Estimated carrier counts Mean | Q25 | Q75 |
|---|---|---|---|---|---|---|---|---|---|
| Paphos | Neo Chorio Pafou | 13·546 | 7·763 | 28·972 | 8·007 | 16·954 | 63 | 37 | 79 |
| Paphos | Nikokleia | 13·574 | 7·450 | 28·303 | 8·158 | 17·069 | 305 | 180 | 387 |
| Paphos | Pano Akourdaleia | 14·026 | 8·025 | 31·146 | 8·361 | 17·811 | 224 | 134 | 284 |
| Paphos | Pano Archimandrita | 13·724 | 7·329 | 27·991 | 8·497 | 17·569 | 4 | 3 | 6 |
| Paphos | Pano Arodes | 13·553 | 7·413 | 27·072 | 8·034 | 17·338 | 34 | 20 | 43 |
| Paphos | Pano Panagia | 13·822 | 7·574 | 29·153 | 8·498 | 17·382 | 67 | 42 | 85 |
| Paphos | Paphos | 13·478 | 7·471 | 28·331 | 8·051 | 17·074 | 7722 | 4618 | 9769 |
| Paphos | Pegeia | 13·505 | 7·657 | 29·390 | 7·940 | 17·493 | 1013 | 593 | 1315 |
| Paphos | Pelathousa | 13·747 | 7·566 | 28·845 | 8·273 | 17·508 | 215 | 129 | 273 |
| Paphos | Pentalia | 13·771 | 7·736 | 30·258 | 8·020 | 17·330 | 35 | 20 | 44 |
| Paphos | Peristerona Pafou | 13·679 | 7·623 | 29·780 | 8·174 | 17·469 | 243 | 145 | 309 |
| Paphos | Pitargou | 13·542 | 7·789 | 28·942 | 7·934 | 17·031 | 66 | 39 | 83 |
| Paphos | Polemi | 13·623 | 7·497 | 28·021 | 8·140 | 17·293 | 476 | 286 | 607 |
| Paphos | Polis | 13·732 | 7·627 | 29·092 | 8·266 | 17·443 | 411 | 247 | 521 |
| Paphos | Pomos | 13·127 | 7·368 | 28·173 | 7·880 | 16·550 | 70 | 42 | 88 |
| Paphos | Praitori | 14·082 | 7·755 | 29·906 | 8·422 | 17·864 | 127 | 77 | 160 |
| Paphos | Prastio Pafou | 13·874 | 7·444 | 28·723 | 8·451 | 17·704 | 38 | 24 | 49 |
| Paphos | Psathi | 13·401 | 7·298 | 26·796 | 8·019 | 16·971 | 91 | 55 | 115 |
| Paphos | Salamiou | 13·944 | 7·669 | 29·620 | 8·332 | 17·729 | 120 | 72 | 152 |
| Paphos | Sarama | 13·572 | 7·449 | 28·098 | 8·122 | 17·328 | 126 | 75 | 160 |
| Paphos | Simou | 13·651 | 7·550 | 29·936 | 8·087 | 17·367 | 192 | 114 | 243 |
| Paphos | Skoulli | 13·976 | 8·073 | 32·399 | 8·438 | 17·810 | 104 | 63 | 132 |
| Paphos | Souskiou | 13·483 | 7·002 | 26·327 | 8·322 | 16·736 | 62 | 38 | 77 |
| Paphos | Statos Agios Fotios | 13·942 | 7·517 | 28·774 | 8·334 | 17·808 | 138 | 83 | 175 |
| Paphos | Stavrokonnou | 13·601 | 7·390 | 28·547 | 8·124 | 17·460 | 91 | 54 | 117 |
| Paphos | Steni | 13·729 | 7·622 | 29·659 | 8·230 | 17·526 | 384 | 230 | 488 |
| Paphos | Stroumpi | 13·917 | 7·805 | 29·866 | 8·225 | 17·712 | 869 | 514 | 1110 |
| Paphos | Tala | 13·541 | 7·565 | 29·126 | 8·038 | 17·268 | 6577 | 3934 | 8325 |
| Paphos | Tera | 13·623 | 7·745 | 30·193 | 8·200 | 17·256 | 209 | 126 | 265 |
| Paphos | Theletra | 14·076 | 7·978 | 29·892 | 8·284 | 17·811 | 120 | 71 | 152 |
| Paphos | Thrinia | 13·646 | 7·449 | 28·612 | 8·196 | 17·183 | 186 | 112 | 234 |
| Paphos | Timi | 13·614 | 7·800 | 31·027 | 8·135 | 17·228 | 185 | 110 | 234 |
| Paphos | Trachypedoula | 13·755 | 7·341 | 27·864 | 8·501 | 17·547 | 14 | 9 | 18 |
| Paphos | Trimithousa | 13·711 | 7·679 | 30·473 | 8·187 | 17·545 | 169 | 101 | 215 |
| Paphos | Trimithousa | 13·514 | 7·492 | 28·882 | 8·021 | 17·177 | 6284 | 3760 | 7948 |
| Paphos | Tsada | 13·757 | 7·706 | 29·210 | 8·153 | 17·531 | 3473 | 2076 | 4456 |
| Paphos | Vretsia | 13·795 | 7·376 | 28·711 | 8·463 | 17·443 | 135 | 82 | 171 |
| Paphos | Zacharia | 13·835 | 7·979 | 29·848 | 8·273 | 17·794 | 2 | 1 | 3 |

ADM1: administrative level 1. ADM2: administrative level 2. SD: standard deviation. CI95%: 95% credible interval. Q25: 25th percentile. Q75: 75th percentile. SD and CI95% are measures of prediction uncertainty.

**Supplementary Table S21. Subnational areal predictions of β-thalassaemia carrier prevalence and estimated carrier count at administrative level 2 of Greece.**

| ADM1 | ADM2 | β-thalassaemia carrier prevalence Mean | SD | CI95% | Q25 | Q75 | Estimated carrier counts Mean | Q25 | Q75 |
|---|---|---|---|---|---|---|---|---|---|
| Agion Oros | Agion Oros | 7·799 | 5·556 | 20·786 | 3·945 | 9·994 | 93 | 47 | 118 |
| Attica | Attikis | 7·187 | 7·213 | 26·864 | 2·471 | 9·271 | 247886 | 86159 | 323721 |
| Crete | Kritis | 6·085 | 6·983 | 25·389 | 1·741 | 7·684 | 33158 | 9513 | 41495 |
| Egean | Notioy Aigaioy | 5·301 | 4·711 | 17·349 | 2·208 | 6·796 | 10495 | 4554 | 13499 |
| Egean | Voreioy Aigaioy | 4·546 | 3·286 | 12·259 | 2·325 | 5·755 | 5970 | 3096 | 7565 |
| Epirus-Western Macedonia | Dytikis Makedonias | 10·870 | 8·187 | 30·759 | 5·084 | 14·240 | 24720 | 11593 | 32358 |
| Epirus-Western Macedonia | Ipeiroy | 9·959 | 8·598 | 32·155 | 4·067 | 12·987 | 27524 | 10966 | 36053 |
| Macedonia-Thrace | Anatolikis Makedonias kai Thrakis | 6·523 | 4·792 | 17·979 | 3·227 | 8·377 | 30310 | 15036 | 38808 |
| Macedonia-Thrace | Kentrikis Makedonias | 10·517 | 7·265 | 27·437 | 5·347 | 13·671 | 156595 | 80299 | 204262 |
| Peloponisos-W· Greece & Ionian | Dytikis Elladas | 8·531 | 8·479 | 31·619 | 2·954 | 10·993 | 44776 | 15434 | 57330 |

| ADM1 | ADM2 | β-thalassaemia carrier prevalence Mean | SD | CI95% | Q25 | Q75 | Estimated carrier counts Mean | Q25 | Q75 |
|---|---|---|---|---|---|---|---|---|---|
| Peloponisos-W· Greece & Ionian | Ionion Nison | 8·788 | 8·717 | 32·806 | 3·068 | 11·297 | 12612 | 4557 | 16338 |
| Peloponisos-W· Greece & Ionian | Peloponnisoy | 7·512 | 8·191 | 30·490 | 2·305 | 9·521 | 33285 | 10374 | 42284 |
| Thessalia-Central Greece | Stereas Elladas | 8·247 | 7·545 | 28·098 | 3·192 | 10·679 | 30606 | 11699 | 39669 |
| Thessalia-Central Greece | Thessalias | 9·770 | 7·853 | 29·507 | 4·324 | 12·741 | 59373 | 26445 | 77344 |

ADM1: administrative level 1. ADM2: administrative level 2. SD: standard deviation. CI95%: 95% credible interval. Q25: 25th percentile. Q75: 75th percentile. SD and CI95% are measures of prediction uncertainty.

**Supplementary Table S22. Subnational areal predictions of β-thalassaemia carrier prevalence and estimated carrier count at administrative level 2 of India.**

| ADM1 | ADM2 | β-thalassaemia carrier prevalence Mean | SD | CI95% | Q25 | Q75 | Estimated carrier counts Mean | Q25 | Q75 |
|---|---|---|---|---|---|---|---|---|---|
| Andaman and Nicobar Islands | Nicobars | 6·177 | 8·779 | 31·713 | 1·300 | 7·234 | 1753 | 403 | 2025 |
| Andaman and Nicobar Islands | North & Middle Andaman | 6·396 | 7·934 | 28·887 | 1·734 | 7·755 | 5535 | 1502 | 6691 |
| Andaman and Nicobar Islands | South Andaman | 6·203 | 7·957 | 28·481 | 1·625 | 7·405 | 10234 | 2651 | 12058 |
| Andhra Pradesh | Anantapur | 2·875 | 2·972 | 10·587 | 1·046 | 3·608 | 124893 | 45649 | 157398 |
| Andhra Pradesh | Chittoor | 3·528 | 3·828 | 13·715 | 1·213 | 4·379 | 194127 | 66228 | 240474 |
| Andhra Pradesh | East Godavari | 3·978 | 5·163 | 18·498 | 0·975 | 4·827 | 201414 | 48261 | 244995 |
| Andhra Pradesh | Guntur | 3·392 | 4·301 | 15·384 | 0·948 | 4·044 | 188268 | 51435 | 223895 |
| Andhra Pradesh | Kadapa (YSR) | 3·277 | 3·713 | 13·317 | 1·044 | 4·058 | 99745 | 31975 | 123360 |
| Andhra Pradesh | Krishna | 3·498 | 4·395 | 15·813 | 0·930 | 4·230 | 168969 | 44823 | 204758 |
| Andhra Pradesh | Kurnool | 2·973 | 3·421 | 12·108 | 0·949 | 3·658 | 96881 | 30937 | 119388 |
| Andhra Pradesh | Prakasam | 3·441 | 4·381 | 15·611 | 0·945 | 4·117 | 125926 | 33881 | 149385 |
| Andhra Pradesh | Sri Potti Sriramulu Nellore | 3·714 | 4·522 | 16·273 | 1·028 | 4·561 | 132221 | 36033 | 161907 |
| Andhra Pradesh | Srikakulam | 5·900 | 7·534 | 27·096 | 1·478 | 7·173 | 147195 | 36553 | 178676 |
| Andhra Pradesh | Visakhapatnam | 4·484 | 5·894 | 21·179 | 1·103 | 5·367 | 209322 | 48923 | 248869 |
| Andhra Pradesh | Vizianagaram | 5·211 | 6·781 | 24·328 | 1·306 | 6·265 | 104954 | 25801 | 126050 |
| Andhra Pradesh | West Godavari | 3·651 | 4·605 | 16·427 | 0·940 | 4·472 | 165783 | 42190 | 203320 |
| Andhra Pradesh | Yanam | 4·378 | 5·828 | 20·789 | 1·028 | 5·184 | 1168 | 279 | 1395 |
| Arunachal Pradesh | Anjaw | 6·241 | 5·727 | 21·342 | 2·506 | 7·957 | 960 | 394 | 1239 |
| Arunachal Pradesh | Changlang | 6·023 | 4·714 | 17·553 | 2·833 | 7·745 | 11449 | 5762 | 14730 |
| Arunachal Pradesh | Dibang Valley | 7·019 | 6·412 | 23·847 | 2·830 | 8·938 | 691 | 289 | 900 |
| Arunachal Pradesh | East Kameng | 8·362 | 6·787 | 25·317 | 3·723 | 10·838 | 2496 | 1162 | 3233 |
| Arunachal Pradesh | East Siang | 7·209 | 4·937 | 18·599 | 3·781 | 9·226 | 14367 | 7585 | 18411 |
| Arunachal Pradesh | Kamle | 8·193 | 5·652 | 21·244 | 4·243 | 10·515 | 1263 | 673 | 1609 |
| Arunachal Pradesh | Kra Daadi | 8·648 | 6·650 | 24·861 | 4·034 | 11·223 | 1228 | 582 | 1595 |
| Arunachal Pradesh | Kurung Kumey | 8·720 | 7·109 | 26·587 | 3·827 | 11·360 | 2179 | 986 | 2860 |
| Arunachal Pradesh | Leparada | 7·465 | 4·988 | 18·736 | 3·974 | 9·541 | 2467 | 1326 | 3181 |
| Arunachal Pradesh | Lohit | 6·511 | 5·040 | 18·756 | 3·081 | 8·372 | 5240 | 2550 | 6670 |
| Arunachal Pradesh | Longding | 6·073 | 4·136 | 15·275 | 3·222 | 7·731 | 2556 | 1370 | 3263 |
| Arunachal Pradesh | Lower Dibang Valley | 6·956 | 5·326 | 19·997 | 3·334 | 8·934 | 7104 | 3533 | 9186 |
| Arunachal Pradesh | Lower Siang | 7·531 | 4·819 | 18·177 | 4·139 | 9·616 | 1932 | 1070 | 2471 |
| Arunachal Pradesh | Lower Subansiri | 8·431 | 5·968 | 22·159 | 4·267 | 10·892 | 5908 | 3041 | 7589 |
| Arunachal Pradesh | Namsai | 6·526 | 4·731 | 17·750 | 3·269 | 8·387 | 10535 | 5396 | 13460 |
| Arunachal Pradesh | Pakke Kessang | 7·804 | 5·945 | 22·321 | 3·697 | 10·071 | 1807 | 857 | 2320 |
| Arunachal Pradesh | Papum Pare | 8·257 | 5·828 | 21·878 | 4·207 | 10·616 | 26731 | 13966 | 34565 |
| Arunachal Pradesh | Shi Yomi | 8·174 | 6·821 | 25·624 | 3·526 | 10·596 | 1496 | 640 | 1948 |
| Arunachal Pradesh | Siang | 7·543 | 5·598 | 21·115 | 3·652 | 9·756 | 1983 | 949 | 2554 |
| Arunachal Pradesh | Tawang | 7·928 | 7·171 | 26·971 | 3·104 | 10·224 | 2801 | 1104 | 3620 |
| Arunachal Pradesh | Tirap | 6·096 | 4·120 | 15·422 | 3·245 | 7·813 | 2986 | 1616 | 3813 |
| Arunachal Pradesh | Upper Siang | 7·727 | 6·607 | 24·712 | 3·263 | 9·999 | 2923 | 1286 | 3764 |
| Arunachal Pradesh | Upper Subansiri | 8·402 | 6·642 | 25·025 | 3·843 | 10·882 | 4597 | 2232 | 5954 |
| Arunachal Pradesh | West Kameng | 7·569 | 6·386 | 23·858 | 3·236 | 9·813 | 5776 | 2510 | 7525 |
| Arunachal Pradesh | West Siang | 7·810 | 5·796 | 21·875 | 3·814 | 10·012 | 7489 | 3698 | 9578 |
| Assam | Baksa | 6·041 | 5·376 | 19·858 | 2·428 | 7·818 | 82550 | 33381 | 107071 |
| Assam | Barpeta | 5·539 | 4·927 | 18·285 | 2·248 | 7·161 | 97732 | 39610 | 126622 |

| | | β-thalassaemia carrier prevalence | | | | | Estimated carrier counts | | |
|---|---|---|---|---|---|---|---|---|---|
| ADM1 | ADM2 | Mean | SD | CI95% | Q25 | Q75 | Mean | Q25 | Q75 |
| Assam | Biswanath | 7·251 | 5·079 | 19·062 | 3·738 | 9·303 | 60492 | 31073 | 77624 |
| Assam | Bongaigaon | 5·710 | 5·118 | 19·047 | 2·309 | 7·316 | 45708 | 18332 | 58324 |
| Assam | Cachar | 1·779 | 1·499 | 5·399 | 0·812 | 2·247 | 25993 | 11775 | 32735 |
| Assam | Charaideo | 6·620 | 4·092 | 15·553 | 3·727 | 8·422 | 39308 | 22201 | 49976 |
| Assam | Chirang | 6·139 | 5·626 | 20·838 | 2·422 | 7·896 | 37157 | 14780 | 47864 |
| Assam | Darrang | 5·579 | 4·772 | 17·701 | 2·380 | 7·182 | 70137 | 29790 | 90522 |
| Assam | Dhemaji | 7·414 | 4·621 | 17·442 | 4·173 | 9·419 | 75249 | 42393 | 95525 |
| Assam | Dhubri | 5·827 | 5·193 | 19·142 | 2·433 | 7·426 | 67636 | 28188 | 86176 |
| Assam | Dibrugarh | 6·960 | 4·253 | 16·037 | 3·973 | 8·828 | 129078 | 73706 | 163535 |
| Assam | Dima Hasao | 2·503 | 2·009 | 7·409 | 1·180 | 3·173 | 8030 | 3806 | 10164 |
| Assam | Goalpara | 5·295 | 4·752 | 17·530 | 2·182 | 6·769 | 63643 | 26261 | 81143 |
| Assam | Golaghat | 6·026 | 3·948 | 14·852 | 3·275 | 7·691 | 115902 | 63514 | 147807 |
| Assam | Hailakandi | 1·724 | 1·532 | 5·527 | 0·763 | 2·167 | 9847 | 4422 | 12434 |
| Assam | Hojai | 4·021 | 3·103 | 11·426 | 1·929 | 5·151 | 38532 | 18453 | 49318 |
| Assam | Jorhat | 7·020 | 4·340 | 16·321 | 3·969 | 8·909 | 86765 | 48777 | 110383 |
| Assam | Kamrup | 4·928 | 4·354 | 15·953 | 2·055 | 6·319 | 111020 | 46182 | 142709 |
| Assam | Kamrup Metropolitan | 4·717 | 4·011 | 14·629 | 2·025 | 6·089 | 62605 | 26476 | 81390 |
| Assam | Karbi Anglong East | 4·848 | 3·466 | 12·903 | 2·484 | 6·177 | 55734 | 28629 | 71210 |
| Assam | Karbi Anglong West | 3·788 | 3·037 | 11·244 | 1·758 | 4·852 | 13347 | 6283 | 16999 |
| Assam | Karimganj | 1·873 | 1·632 | 5·856 | 0·847 | 2·344 | 19930 | 9132 | 25058 |
| Assam | Kokrajhar | 6·104 | 5·467 | 20·266 | 2·481 | 7·844 | 66138 | 27055 | 84572 |
| Assam | Lakhimpur | 7·865 | 4·950 | 18·637 | 4·345 | 10·081 | 108312 | 59470 | 139192 |
| Assam | Majuli | 7·535 | 4·650 | 17·591 | 4·237 | 9·615 | 16865 | 9474 | 21492 |
| Assam | Morigaon | 5·115 | 4·282 | 15·885 | 2·232 | 6·625 | 50945 | 22348 | 65870 |
| Assam | Nagaon | 5·347 | 4·142 | 15·508 | 2·538 | 6·871 | 106994 | 50185 | 137615 |
| Assam | Nalbari | 5·450 | 4·884 | 17·958 | 2·178 | 7·040 | 50786 | 20370 | 65846 |
| Assam | Sivasagar | 7·064 | 4·293 | 16·327 | 4·049 | 8·951 | 75763 | 43382 | 95775 |
| Assam | Sonitpur | 6·512 | 5·142 | 19·252 | 3·001 | 8·394 | 117938 | 54513 | 151688 |
| Assam | South Salmara-Mankachar | 5·461 | 4·728 | 17·375 | 2·362 | 6·933 | 17842 | 7745 | 22631 |
| Assam | Tinsukia | 6·781 | 4·447 | 16·748 | 3·670 | 8·648 | 103812 | 56430 | 132427 |
| Assam | Udalguri | 6·176 | 5·294 | 19·745 | 2·608 | 7·975 | 77961 | 33028 | 100742 |
| Bihar | Araria | 7·019 | 5·806 | 21·684 | 3·069 | 9·069 | 278225 | 121691 | 359951 |
| Bihar | Arwal | 8·212 | 8·186 | 31·268 | 2·938 | 10·474 | 53702 | 19188 | 68531 |
| Bihar | Aurangabad | 8·352 | 8·089 | 30·272 | 3·039 | 10·730 | 298339 | 108487 | 382507 |
| Bihar | Banka | 7·728 | 6·199 | 22·951 | 3·563 | 9·912 | 195602 | 89954 | 250866 |
| Bihar | Begusarai | 8·011 | 7·404 | 27·544 | 3·097 | 10·348 | 317000 | 122279 | 409929 |
| Bihar | Bhagalpur | 7·429 | 6·157 | 22·925 | 3·329 | 9·492 | 247932 | 110999 | 316070 |
| Bihar | Bhojpur | 7·924 | 8·094 | 29·929 | 2·720 | 10·103 | 228320 | 78032 | 291595 |
| Bihar | Buxar | 7·573 | 7·819 | 29·184 | 2·590 | 9·562 | 130196 | 44541 | 164766 |
| Bihar | Darbhanga | 7·622 | 7·167 | 26·719 | 2·841 | 9·917 | 361088 | 133353 | 469731 |
| Bihar | Gaya | 8·706 | 8·023 | 29·801 | 3·312 | 11·334 | 566214 | 214783 | 737010 |
| Bihar | Gopalganj | 6·877 | 7·104 | 26·424 | 2·276 | 8·814 | 265770 | 88039 | 340656 |
| Bihar | Jamui | 8·191 | 6·842 | 25·374 | 3·573 | 10·584 | 209530 | 91234 | 269486 |
| Bihar | Jehanabad | 8·373 | 8·136 | 30·558 | 3·019 | 10·804 | 116585 | 42133 | 150586 |
| Bihar | Kaimur (Bhabua) | 7·866 | 7·926 | 29·862 | 2·723 | 10·031 | 158742 | 55027 | 201908 |
| Bihar | Katihar | 6·978 | 5·480 | 20·606 | 3·245 | 8·975 | 310887 | 145013 | 398936 |
| Bihar | Khagaria | 7·722 | 6·841 | 25·436 | 3·168 | 9·912 | 123602 | 50792 | 158357 |
| Bihar | Kishanganj | 6·620 | 5·074 | 18·823 | 3·161 | 8·537 | 198292 | 94925 | 254918 |
| Bihar | Lakhisarai | 8·149 | 7·264 | 26·842 | 3·304 | 10·492 | 86475 | 34976 | 111565 |
| Bihar | Madhepura | 7·449 | 6·447 | 24·098 | 3·097 | 9·663 | 224054 | 92510 | 291263 |
| Bihar | Madhubani | 7·398 | 7·017 | 26·220 | 2·714 | 9·626 | 432264 | 158226 | 562778 |
| Bihar | Munger | 7·896 | 6·882 | 25·719 | 3·326 | 10·139 | 115806 | 48021 | 148376 |
| Bihar | Muzaffarpur | 7·622 | 7·564 | 28·179 | 2·622 | 9·890 | 518550 | 177944 | 674576 |
| Bihar | Nalanda | 8·328 | 7·865 | 29·059 | 3·132 | 10·790 | 251251 | 95106 | 326644 |
| Bihar | Nawada | 8·671 | 7·814 | 29·119 | 3·436 | 11·250 | 222633 | 87667 | 288642 |
| Bihar | Pashchim Champaran | 6·357 | 6·806 | 25·054 | 2·029 | 8·092 | 306260 | 98668 | 390825 |
| Bihar | Patna | 8·168 | 8·030 | 29·703 | 2·891 | 10·562 | 593039 | 204783 | 761654 |

| | | β-thalassaemia carrier prevalence | | | | | Estimated carrier counts | | |
|---|---|---|---|---|---|---|---|---|---|
| ADM1 | ADM2 | Mean | SD | CI95% | Q25 | Q75 | Mean | Q25 | Q75 |
| Bihar | Purba Champaran | 6·987 | 7·200 | 26·492 | 2·322 | 9·015 | 450822 | 149675 | 581590 |
| Bihar | Purnia | 7·091 | 5·767 | 21·672 | 3·189 | 9·128 | 371686 | 167294 | 477073 |
| Bihar | Rohtas | 8·099 | 8·177 | 30·626 | 2·838 | 10·315 | 249528 | 87784 | 317651 |
| Bihar | Saharsa | 7·629 | 6·799 | 25·262 | 3·056 | 9·873 | 190055 | 76493 | 247004 |
| Bihar | Samastipur | 7·938 | 7·536 | 28·119 | 2·899 | 10·332 | 424766 | 155027 | 552912 |
| Bihar | Saran | 7·620 | 7·715 | 28·668 | 2·602 | 9·779 | 427660 | 146310 | 548621 |
| Bihar | Sheikhpura | 8·348 | 7·645 | 28·357 | 3·267 | 10·772 | 48115 | 18692 | 62264 |
| Bihar | Sheohar | 7·173 | 7·162 | 26·379 | 2·455 | 9·287 | 53453 | 18373 | 69159 |
| Bihar | Sitamarhi | 7·262 | 7·310 | 26·778 | 2·468 | 9·425 | 244637 | 83384 | 317371 |
| Bihar | Siwan | 7·083 | 7·285 | 26·668 | 2·386 | 9·092 | 267311 | 89956 | 343056 |
| Bihar | Supaul | 7·327 | 6·558 | 24·522 | 2·933 | 9·482 | 272701 | 109455 | 352480 |
| Bihar | Vaishali | 7·983 | 7·868 | 29·022 | 2·797 | 10·337 | 419977 | 146863 | 545880 |
| Chandigarh | Chandigarh | 3·594 | 2·813 | 10·241 | 1·768 | 4·502 | 49609 | 24415 | 62241 |
| Chhattisgarh | Balod | 4·193 | 3·905 | 14·183 | 1·687 | 5·324 | 40540 | 16226 | 51399 |
| Chhattisgarh | Baloda Bazar | 6·538 | 6·729 | 24·517 | 2·241 | 8·356 | 70112 | 24220 | 89402 |
| Chhattisgarh | Balrampur | 8·645 | 8·528 | 31·780 | 3·088 | 11·048 | 66440 | 23681 | 84896 |
| Chhattisgarh | Bastar | 3·522 | 4·012 | 14·404 | 1·095 | 4·382 | 34857 | 10739 | 43398 |
| Chhattisgarh | Bemetra | 6·361 | 6·221 | 22·754 | 2·362 | 8·125 | 46893 | 17384 | 59910 |
| Chhattisgarh | Bijapur | 2·740 | 2·866 | 10·079 | 1·004 | 3·422 | 8994 | 3222 | 11195 |
| Chhattisgarh | Bilaspur | 7·304 | 7·240 | 26·475 | 2·627 | 9·385 | 161717 | 57752 | 208155 |
| Chhattisgarh | Dakshin Bastar Dantewada | 3·099 | 3·455 | 12·236 | 1·014 | 3·858 | 13489 | 4452 | 16745 |
| Chhattisgarh | Dhamtari | 4·497 | 4·617 | 16·797 | 1·585 | 5·688 | 40368 | 14494 | 51028 |
| Chhattisgarh | Durg | 5·302 | 5·131 | 18·714 | 2·005 | 6·762 | 108324 | 41546 | 138189 |
| Chhattisgarh | Gariaband | 4·853 | 5·251 | 19·024 | 1·593 | 6·114 | 31700 | 10473 | 39942 |
| Chhattisgarh | Gaurella Pendra Marwahi | 8·044 | 7·799 | 28·836 | 2·925 | 10·395 | 45467 | 16641 | 58627 |
| Chhattisgarh | Janjgir-Champa | 7·267 | 7·437 | 27·061 | 2·493 | 9·336 | 152846 | 52578 | 196366 |
| Chhattisgarh | Jashpur | 8·804 | 8·402 | 31·128 | 3·264 | 11·384 | 85038 | 31432 | 110035 |
| Chhattisgarh | Kabeerdham | 7·040 | 6·590 | 24·280 | 2·741 | 9·036 | 61620 | 23813 | 78567 |
| Chhattisgarh | Kondagaon | 3·532 | 3·758 | 13·426 | 1·227 | 4·414 | 26907 | 9299 | 33620 |
| Chhattisgarh | Korba | 7·900 | 7·863 | 28·767 | 2·804 | 10·164 | 129729 | 45861 | 167804 |
| Chhattisgarh | Koriya | 8·553 | 8·464 | 31·720 | 3·052 | 10·958 | 94918 | 34016 | 121571 |
| Chhattisgarh | Mahasamund | 6·239 | 6·610 | 24·082 | 2·053 | 7·980 | 78007 | 25668 | 99953 |
| Chhattisgarh | Mungeli | 7·288 | 7·180 | 26·416 | 2·662 | 9·309 | 55158 | 20047 | 70397 |
| Chhattisgarh | Narayanpur | 2·937 | 2·882 | 10·423 | 1·136 | 3·694 | 5195 | 1978 | 6543 |
| Chhattisgarh | Raigarh | 7·911 | 8·001 | 29·208 | 2·750 | 10·183 | 142825 | 49435 | 183792 |
| Chhattisgarh | Raipur | 5·734 | 5·776 | 21·186 | 2·034 | 7·341 | 159915 | 58051 | 205817 |
| Chhattisgarh | Rajnandgaon | 4·723 | 4·148 | 15·207 | 2·023 | 6·009 | 96710 | 40677 | 122994 |
| Chhattisgarh | Sukma | 3·220 | 3·749 | 13·324 | 0·976 | 3·997 | 12502 | 3743 | 15587 |
| Chhattisgarh | Surajpur | 8·535 | 8·572 | 32·077 | 3·029 | 10·816 | 79421 | 28173 | 100654 |
| Chhattisgarh | Surguja | 8·430 | 8·353 | 31·112 | 3·021 | 10·736 | 98092 | 34878 | 124851 |
| Chhattisgarh | Uttar Bastar Kanker | 3·363 | 3·184 | 11·573 | 1·344 | 4·250 | 46053 | 17984 | 58235 |
| Dadra and Nagar Haveli and Daman and Diu | Dadra & Nagar Haveli | 3·951 | 2·669 | 9·954 | 2·128 | 5·006 | 25346 | 13663 | 31898 |
| Dadra and Nagar Haveli and Daman and Diu | Daman | 3·958 | 2·832 | 11·077 | 2·076 | 4·955 | 15322 | 8029 | 19131 |
| Dadra and Nagar Haveli and Daman and Diu | Diu | 6·815 | 4·792 | 17·879 | 3·468 | 8·772 | 2936 | 1494 | 3779 |
| Delhi | Central | 4·466 | 4·492 | 16·012 | 1·660 | 5·607 | 61209 | 22799 | 77134 |
| Delhi | East | 4·439 | 4·477 | 16·169 | 1·679 | 5·590 | 60309 | 22697 | 75597 |
| Delhi | New Delhi | 4·546 | 4·539 | 16·666 | 1·716 | 5·705 | 171062 | 64531 | 214431 |
| Delhi | North | 4·352 | 4·364 | 15·664 | 1·683 | 5·422 | 116908 | 44672 | 145678 |
| Delhi | North East | 4·483 | 4·445 | 15·975 | 1·682 | 5·625 | 112735 | 42437 | 140330 |
| Delhi | North West | 4·533 | 4·641 | 16·190 | 1·733 | 5·594 | 153081 | 58402 | 187485 |
| Delhi | Shahdara | 4·478 | 4·654 | 15·681 | 1·665 | 5·570 | 84009 | 31226 | 104468 |
| Delhi | South | 4·640 | 4·605 | 16·483 | 1·737 | 5·837 | 68030 | 25471 | 84844 |
| Delhi | South East | 4·585 | 4·743 | 17·889 | 1·714 | 5·670 | 109861 | 40762 | 135823 |
| Delhi | South West | 4·558 | 4·626 | 16·442 | 1·696 | 5·706 | 78137 | 29456 | 98220 |
| Delhi | West | 4·563 | 4·569 | 16·786 | 1·680 | 5·735 | 125005 | 45820 | 157538 |

| | | β-thalassaemia carrier prevalence | | | | | Estimated carrier counts | | |
|---|---|---|---|---|---|---|---|---|---|
| **ADM1** | **ADM2** | **Mean** | **SD** | **CI95%** | **Q25** | **Q75** | **Mean** | **Q25** | **Q75** |
| Goa | North Goa | 2·081 | 1·804 | 6·511 | 0·944 | 2·608 | 15969 | 7211 | 19938 |
| Goa | South Goa | 2·198 | 2·179 | 7·519 | 0·900 | 2·711 | 14276 | 5938 | 17731 |
| Gujarat | Ahmadabad | 3·267 | 2·123 | 7·883 | 1·812 | 4·120 | 162212 | 91207 | 204072 |
| Gujarat | Amreli | 5·886 | 3·820 | 14·333 | 3·228 | 7·473 | 128996 | 70808 | 163688 |
| Gujarat | Anand | 3·293 | 2·124 | 7·974 | 1·841 | 4·134 | 78538 | 44175 | 98307 |
| Gujarat | Aravali | 2·720 | 1·938 | 7·186 | 1·422 | 3·431 | 35558 | 18685 | 44828 |
| Gujarat | Banas Kantha | 3·372 | 2·488 | 9·181 | 1·706 | 4·268 | 159907 | 81541 | 202214 |
| Gujarat | Batod | 4·566 | 3·010 | 11·278 | 2·492 | 5·789 | 42618 | 23122 | 54120 |
| Gujarat | Bharuch | 3·486 | 2·271 | 8·464 | 1·933 | 4·384 | 73572 | 40760 | 92611 |
| Gujarat | Bhavnagar | 4·716 | 3·118 | 11·682 | 2·567 | 5·978 | 141665 | 77067 | 179309 |
| Gujarat | Chhota Udaipur | 3·496 | 2·352 | 8·732 | 1·901 | 4·415 | 39524 | 21449 | 49913 |
| Gujarat | Devbhumi Dwarka | 7·582 | 5·331 | 19·998 | 3·892 | 9·702 | 120576 | 62279 | 154203 |
| Gujarat | Dohad | 3·390 | 2·395 | 8·833 | 1·788 | 4·286 | 62736 | 33017 | 79347 |
| Gujarat | Gandhinagar | 2·635 | 1·720 | 6·352 | 1·475 | 3·309 | 58729 | 32957 | 73849 |
| Gujarat | Gir Somnath | 7·036 | 4·837 | 18·177 | 3·669 | 9·009 | 137758 | 70628 | 176977 |
| Gujarat | Jamnagar | 6·289 | 4·107 | 15·507 | 3·439 | 7·992 | 176192 | 96176 | 223913 |
| Gujarat | Junagadh | 7·357 | 4·756 | 17·943 | 4·035 | 9·359 | 184317 | 101158 | 234580 |
| Gujarat | Kachchh | 5·669 | 4·194 | 15·567 | 2·827 | 7·235 | 283963 | 144632 | 361065 |
| Gujarat | Kheda | 2·998 | 1·941 | 7·258 | 1·676 | 3·774 | 69640 | 38891 | 87878 |
| Gujarat | Mahesana | 2·589 | 1·750 | 6·501 | 1·408 | 3·252 | 66081 | 35847 | 83002 |
| Gujarat | Mahisagar | 3·022 | 2·131 | 7·849 | 1·587 | 3·817 | 30979 | 16328 | 39140 |
| Gujarat | Morbi | 4·972 | 3·399 | 12·714 | 2·650 | 6·309 | 100727 | 53150 | 127595 |
| Gujarat | Narmada | 3·517 | 2·303 | 8·539 | 1·945 | 4·436 | 17164 | 9547 | 21602 |
| Gujarat | Navsari | 3·737 | 2·481 | 9·279 | 2·050 | 4·709 | 62576 | 34437 | 78867 |
| Gujarat | Panch Mahals | 3·287 | 2·176 | 8·059 | 1·803 | 4·148 | 53285 | 29031 | 67202 |
| Gujarat | Patan | 3·414 | 2·326 | 8·668 | 1·842 | 4·303 | 48501 | 26250 | 60995 |
| Gujarat | Porbandar | 7·869 | 5·193 | 19·501 | 4·233 | 10·074 | 88708 | 47304 | 113666 |
| Gujarat | Rajkot | 5·917 | 3·757 | 14·171 | 3·307 | 7·499 | 267381 | 148646 | 339941 |
| Gujarat | Sabar Kantha | 2·574 | 1·875 | 6·929 | 1·325 | 3·247 | 47408 | 24796 | 59680 |
| Gujarat | Surat | 3·582 | 2·357 | 8·771 | 1·969 | 4·522 | 150892 | 83325 | 190859 |
| Gujarat | Surendranagar | 4·096 | 2·689 | 10·074 | 2·247 | 5·183 | 92762 | 50686 | 117348 |
| Gujarat | Tapi | 3·565 | 2·352 | 8·868 | 1·960 | 4·502 | 35149 | 19276 | 44455 |
| Gujarat | The Dangs | 3·729 | 2·500 | 9·358 | 2·027 | 4·711 | 6514 | 3545 | 8199 |
| Gujarat | Vadodara | 3·336 | 2·116 | 7·903 | 1·883 | 4·203 | 110721 | 62703 | 139459 |
| Gujarat | Valsad | 3·903 | 2·655 | 9·901 | 2·097 | 4·950 | 91389 | 49166 | 115770 |
| Haryana | Ambala | 3·612 | 2·900 | 10·571 | 1·719 | 4·581 | 49059 | 23075 | 62057 |
| Haryana | Bhiwani | 4·959 | 5·402 | 19·374 | 1·691 | 6·145 | 112606 | 38547 | 139959 |
| Haryana | Faridabad | 4·682 | 4·760 | 17·038 | 1·766 | 5·840 | 90051 | 34597 | 112100 |
| Haryana | Fatehabad | 4·741 | 4·835 | 17·529 | 1·755 | 5·923 | 63304 | 23459 | 79382 |
| Haryana | Gurgaon | 4·877 | 5·101 | 18·474 | 1·725 | 6·092 | 156337 | 56074 | 195298 |
| Haryana | Hisar | 4·791 | 4·985 | 18·160 | 1·728 | 5·978 | 123879 | 44714 | 153641 |
| Haryana | Jhajjar | 4·733 | 4·955 | 18·051 | 1·692 | 5·911 | 67655 | 24357 | 84645 |
| Haryana | Jind | 4·385 | 4·196 | 15·370 | 1·716 | 5·535 | 65655 | 25620 | 82889 |
| Haryana | Kaithal | 4·095 | 3·631 | 13·307 | 1·764 | 5·182 | 54909 | 23591 | 69424 |
| Haryana | Karnal | 3·869 | 3·369 | 12·379 | 1·683 | 4·897 | 63618 | 27619 | 80347 |
| Haryana | Kurukshetra | 3·765 | 3·111 | 11·436 | 1·729 | 4·785 | 45532 | 20922 | 57649 |
| Haryana | Mahendragarh | 5·105 | 5·615 | 20·032 | 1·706 | 6·341 | 62960 | 21073 | 78264 |
| Haryana | Mewat | 5·106 | 5·319 | 19·471 | 1·798 | 6·433 | 45190 | 15963 | 56840 |
| Haryana | Palwal | 4·972 | 5·156 | 18·456 | 1·790 | 6·239 | 47537 | 17108 | 59672 |
| Haryana | Panchkula | 3·508 | 2·700 | 9·930 | 1·703 | 4·469 | 27184 | 13385 | 34425 |
| Haryana | Panipat | 4·075 | 3·713 | 13·763 | 1·690 | 5·159 | 52461 | 21787 | 66598 |
| Haryana | Rewari | 5·058 | 5·425 | 19·682 | 1·741 | 6·271 | 71107 | 24456 | 88487 |
| Haryana | Rohtak | 4·573 | 4·638 | 16·748 | 1·699 | 5·734 | 56181 | 20763 | 70383 |
| Haryana | Sirsa | 4·965 | 5·290 | 19·274 | 1·702 | 6·197 | 100213 | 34402 | 125073 |
| Haryana | Sonipat | 4·280 | 4·107 | 14·917 | 1·685 | 5·393 | 71936 | 28028 | 90803 |
| Haryana | Yamunanagar | 3·415 | 2·657 | 9·750 | 1·642 | 4·344 | 41738 | 19952 | 53118 |
| Himachal Pradesh | Bilaspur | 3·668 | 3·039 | 11·135 | 1·669 | 4·671 | 20221 | 9182 | 25782 |

| ADM1 | ADM2 | β-thalassaemia carrier prevalence Mean | SD | CI95% | Q25 | Q75 | Estimated carrier counts Mean | Q25 | Q75 |
|---|---|---|---|---|---|---|---|---|---|
| Himachal Pradesh | Chamba | 4·135 | 4·541 | 16·279 | 1·342 | 5·174 | 14697 | 4862 | 18497 |
| Himachal Pradesh | Hamirpur | 3·757 | 3·332 | 12·196 | 1·600 | 4·776 | 19196 | 8237 | 24414 |
| Himachal Pradesh | Kangra | 3·962 | 3·933 | 14·066 | 1·485 | 5·009 | 75968 | 28664 | 96031 |
| Himachal Pradesh | Kinnaur | 3·106 | 2·940 | 10·565 | 1·254 | 3·919 | 2435 | 987 | 3086 |
| Himachal Pradesh | Kullu | 3·530 | 3·456 | 12·445 | 1·365 | 4·442 | 15458 | 6083 | 19510 |
| Himachal Pradesh | Lahul & Spiti | 3·751 | 4·144 | 14·747 | 1·238 | 4·681 | 968 | 309 | 1215 |
| Himachal Pradesh | Mandi | 3·603 | 3·267 | 11·881 | 1·497 | 4·575 | 39106 | 16221 | 49648 |
| Himachal Pradesh | Shimla | 3·245 | 2·730 | 9·978 | 1·472 | 4·114 | 23492 | 10754 | 29848 |
| Himachal Pradesh | Sirmaur | 3·274 | 2·546 | 9·384 | 1·591 | 4·141 | 17817 | 8782 | 22611 |
| Himachal Pradesh | Solan | 3·554 | 2·866 | 10·426 | 1·677 | 4·525 | 26107 | 12382 | 33270 |
| Himachal Pradesh | Una | 3·853 | 3·318 | 12·128 | 1·688 | 4·912 | 33500 | 14687 | 42702 |
| Jammu and Kashmir | Anantnag | 4·722 | 5·982 | 21·382 | 1·213 | 5·784 | 35561 | 9197 | 43750 |
| Jammu and Kashmir | Badgam | 4·855 | 6·140 | 22·249 | 1·220 | 5·982 | 31935 | 7986 | 39121 |
| Jammu and Kashmir | Bandipore | 5·037 | 6·723 | 24·319 | 1·163 | 6·052 | 22278 | 5209 | 26431 |
| Jammu and Kashmir | Baramula | 4·991 | 6·492 | 23·156 | 1·186 | 6·088 | 53197 | 12519 | 64992 |
| Jammu and Kashmir | Doda | 4·410 | 5·139 | 18·443 | 1·295 | 5·496 | 12756 | 3735 | 15813 |
| Jammu and Kashmir | Ganderbal | 4·914 | 6·468 | 23·537 | 1·172 | 5·923 | 23310 | 5550 | 27895 |
| Jammu and Kashmir | Jammu | 4·643 | 5·484 | 19·982 | 1·383 | 5·676 | 110022 | 33149 | 134922 |
| Jammu and Kashmir | Kathua | 4·297 | 4·687 | 16·874 | 1·416 | 5·417 | 46899 | 15732 | 59035 |
| Jammu and Kashmir | Kishtwar | 4·517 | 5·542 | 19·835 | 1·228 | 5·571 | 7992 | 2225 | 10066 |
| Jammu and Kashmir | Kulgam | 4·763 | 5·910 | 21·393 | 1·254 | 5·874 | 19235 | 5073 | 23971 |
| Jammu and Kashmir | Kupwara | 5·197 | 6·984 | 25·267 | 1·170 | 6·229 | 36730 | 8388 | 43963 |
| Jammu and Kashmir | Pulwama | 4·779 | 6·082 | 21·980 | 1·206 | 5·890 | 25342 | 6422 | 31202 |
| Jammu and Kashmir | Punch | 4·938 | 6·218 | 22·570 | 1·234 | 6·065 | 29536 | 7342 | 36625 |
| Jammu and Kashmir | Rajouri | 4·849 | 6·017 | 21·674 | 1·311 | 5·939 | 42977 | 11569 | 52761 |
| Jammu and Kashmir | Ramban | 4·638 | 5·648 | 20·249 | 1·276 | 5·767 | 10800 | 2984 | 13450 |
| Jammu and Kashmir | Reasi | 4·707 | 5·752 | 20·712 | 1·285 | 5·778 | 23143 | 6436 | 28365 |
| Jammu and Kashmir | Samba | 4·508 | 5·089 | 18·648 | 1·430 | 5·554 | 26291 | 8290 | 32227 |
| Jammu and Kashmir | Shupiyan | 4·767 | 5·951 | 21·624 | 1·226 | 5·916 | 12476 | 3211 | 15352 |
| Jammu and Kashmir | Srinagar | 4·851 | 6·309 | 22·203 | 1·186 | 5·942 | 63566 | 15220 | 78024 |
| Jammu and Kashmir | Udhampur | 4·484 | 5·191 | 18·738 | 1·344 | 5·555 | 42294 | 12653 | 51866 |
| Jharkhand | Bokaro | 9·148 | 6·597 | 24·923 | 4·563 | 11·789 | 247350 | 124519 | 318798 |
| Jharkhand | Chatra | 9·137 | 8·140 | 30·632 | 3·619 | 11·903 | 120565 | 47859 | 156680 |
| Jharkhand | Deoghar | 8·020 | 5·912 | 22·052 | 3·961 | 10·322 | 153418 | 75176 | 197440 |
| Jharkhand | Dhanbad | 8·647 | 5·933 | 22·460 | 4·490 | 11·125 | 293748 | 152711 | 376589 |
| Jharkhand | Dumka | 7·508 | 5·224 | 19·678 | 3·878 | 9·625 | 120823 | 62161 | 155271 |
| Jharkhand | Garhwa | 8·580 | 8·408 | 31·510 | 3·037 | 11·019 | 142894 | 50504 | 183149 |
| Jharkhand | Giridih | 8·687 | 6·737 | 25·270 | 4·026 | 11·278 | 259026 | 121016 | 336126 |
| Jharkhand | Godda | 7·333 | 5·589 | 20·810 | 3·531 | 9·418 | 88509 | 42133 | 114051 |
| Jharkhand | Gumla | 9·242 | 8·266 | 31·004 | 3·694 | 11·950 | 112735 | 45634 | 145853 |
| Jharkhand | Hazaribagh | 9·267 | 7·700 | 28·881 | 3·997 | 12·041 | 234083 | 100507 | 303298 |
| Jharkhand | Jamtara | 8·095 | 5·551 | 21·018 | 4·227 | 10·416 | 80747 | 41851 | 103811 |
| Jharkhand | Khunti | 9·545 | 7·684 | 29·041 | 4·270 | 12·318 | 57341 | 25570 | 73929 |
| Jharkhand | Kodarma | 8·876 | 7·545 | 28·196 | 3·715 | 11·588 | 100053 | 42115 | 130754 |
| Jharkhand | Latehar | 9·115 | 8·392 | 31·232 | 3·524 | 11·780 | 98841 | 38550 | 127353 |
| Jharkhand | Lohardaga | 9·331 | 8·333 | 31·019 | 3·732 | 12·094 | 66786 | 27206 | 86576 |
| Jharkhand | Pakur | 7·154 | 5·143 | 19·257 | 3·591 | 9·183 | 53797 | 26751 | 69125 |
| Jharkhand | Palamu | 8·700 | 8·261 | 30·991 | 3·200 | 11·258 | 195088 | 71817 | 251783 |
| Jharkhand | Pashchimi Singhbhum | 9·778 | 7·846 | 29·402 | 4·374 | 12·736 | 194285 | 88518 | 253192 |
| Jharkhand | Purbi Singhbhum | 9·795 | 6·562 | 24·703 | 5·177 | 12·633 | 290118 | 151276 | 375679 |
| Jharkhand | Ramgarh | 9·405 | 7·340 | 27·521 | 4·381 | 12·148 | 130115 | 60307 | 167745 |
| Jharkhand | Ranchi | 9·500 | 7·490 | 28·026 | 4·381 | 12·263 | 422563 | 189729 | 547218 |
| Jharkhand | Sahibganj | 7·080 | 5·421 | 20·294 | 3·385 | 9·122 | 80442 | 38619 | 103690 |
| Jharkhand | Saraikela-Kharsawan | 9·666 | 6·892 | 25·938 | 4·871 | 12·461 | 134714 | 68134 | 174323 |
| Jharkhand | Simdega | 9·212 | 8·266 | 30·964 | 3·656 | 11·952 | 75456 | 29895 | 97825 |
| Karnataka | Bagalkot | 2·461 | 2·268 | 8·099 | 1·051 | 3·074 | 53769 | 23369 | 67126 |
| Karnataka | Bangalore | 2·839 | 2·343 | 8·487 | 1·328 | 3·587 | 237536 | 111644 | 299934 |

| ADM1 | ADM2 | β-thalassaemia carrier prevalence Mean | SD | CI95% | Q25 | Q75 | Estimated carrier counts Mean | Q25 | Q75 |
|---|---|---|---|---|---|---|---|---|---|
| Karnataka | Bangalore Rural | 2·804 | 2·353 | 8·455 | 1·283 | 3·556 | 57017 | 26260 | 72164 |
| Karnataka | Belgaum | 2·328 | 1·931 | 6·990 | 1·088 | 2·929 | 123929 | 58966 | 155802 |
| Karnataka | Bellary | 2·672 | 2·812 | 9·989 | 0·967 | 3·326 | 55574 | 20167 | 69253 |
| Karnataka | Bidar | 2·587 | 2·176 | 7·890 | 1·181 | 3·273 | 33423 | 15153 | 42392 |
| Karnataka | Bijapur | 2·521 | 2·195 | 7·970 | 1·112 | 3·179 | 71769 | 31978 | 90339 |
| Karnataka | Chamarajanagar | 3·455 | 3·489 | 12·649 | 1·264 | 4·357 | 38175 | 13947 | 48135 |
| Karnataka | Chikkaballapura | 2·894 | 2·642 | 9·464 | 1·211 | 3·669 | 44410 | 18774 | 56293 |
| Karnataka | Chikmagalur | 2·841 | 3·070 | 10·796 | 1·007 | 3·517 | 47768 | 17035 | 59061 |
| Karnataka | Chitradurga | 2·737 | 2·757 | 9·780 | 1·031 | 3·441 | 52065 | 19672 | 65392 |
| Karnataka | Dakshina Kannada | 3·085 | 3·505 | 12·150 | 1·018 | 3·810 | 134101 | 43764 | 165509 |
| Karnataka | Davanagere | 2·652 | 2·787 | 9·823 | 0·976 | 3·292 | 46447 | 17228 | 58120 |
| Karnataka | Dharwad | 2·362 | 2·341 | 8·242 | 0·951 | 2·918 | 34326 | 13962 | 42318 |
| Karnataka | Gadag | 2·463 | 2·520 | 8·889 | 0·960 | 3·037 | 23666 | 9238 | 29195 |
| Karnataka | Gulbarga | 2·561 | 2·304 | 8·324 | 1·084 | 3·252 | 58452 | 24626 | 74176 |
| Karnataka | Hassan | 2·967 | 3·049 | 10·949 | 1·099 | 3·701 | 82401 | 30745 | 103172 |
| Karnataka | Haveri | 2·489 | 2·598 | 9·099 | 0·927 | 3·085 | 37854 | 14075 | 46981 |
| Karnataka | Kodagu | 3·352 | 3·782 | 13·125 | 1·107 | 4·169 | 34623 | 11427 | 43149 |
| Karnataka | Kolar | 3·069 | 2·788 | 9·976 | 1·302 | 3·870 | 60009 | 25777 | 75583 |
| Karnataka | Koppal | 2·587 | 2·730 | 9·636 | 0·963 | 3·191 | 37401 | 13816 | 46132 |
| Karnataka | Mandya | 3·005 | 2·827 | 10·314 | 1·201 | 3·805 | 70906 | 28423 | 89880 |
| Karnataka | Mysore | 3·323 | 3·453 | 12·397 | 1·182 | 4·168 | 104947 | 38279 | 132100 |
| Karnataka | Raichur | 2·672 | 2·788 | 9·856 | 0·971 | 3·345 | 51557 | 18654 | 64485 |
| Karnataka | Ramanagara | 2·925 | 2·512 | 9·166 | 1·302 | 3·693 | 50943 | 22877 | 64370 |
| Karnataka | Shimoga | 2·619 | 2·873 | 10·089 | 0·921 | 3·224 | 66815 | 23560 | 82478 |
| Karnataka | Tumkur | 2·771 | 2·547 | 9·141 | 1·149 | 3·509 | 109545 | 45798 | 138818 |
| Karnataka | Udupi | 2·757 | 3·154 | 11·139 | 0·918 | 3·364 | 75783 | 24765 | 92120 |
| Karnataka | Uttara Kannada | 2·356 | 2·479 | 8·652 | 0·896 | 2·891 | 37733 | 14026 | 46199 |
| Karnataka | Yadgir | 2·606 | 2·570 | 9·141 | 1·008 | 3·295 | 26807 | 10403 | 33905 |
| Kerala | Alappuzha | 5·401 | 6·860 | 24·578 | 1·387 | 6·542 | 132217 | 34075 | 160243 |
| Kerala | Ernakulam | 4·972 | 6·050 | 21·811 | 1·342 | 6·162 | 249302 | 67030 | 308474 |
| Kerala | Idukki | 5·230 | 6·284 | 22·842 | 1·417 | 6·514 | 47739 | 13020 | 59812 |
| Kerala | Kannur | 3·554 | 4·148 | 14·377 | 1·123 | 4·393 | 90755 | 28462 | 112473 |
| Kerala | Kasaragod | 3·275 | 3·777 | 12·997 | 1·047 | 4·039 | 54428 | 17361 | 66856 |
| Kerala | Kollam | 5·760 | 7·189 | 26·462 | 1·500 | 7·054 | 155245 | 39526 | 188547 |
| Kerala | Kottayam | 5·289 | 6·513 | 23·502 | 1·377 | 6·570 | 119055 | 31003 | 148054 |
| Kerala | Kozhikode | 3·926 | 4·653 | 16·008 | 1·182 | 4·870 | 78068 | 23085 | 97138 |
| Kerala | Malappuram | 4·133 | 4·837 | 17·061 | 1·231 | 5·151 | 134049 | 39283 | 166373 |
| Kerala | Palakkad | 4·405 | 5·142 | 18·280 | 1·282 | 5·543 | 128960 | 37187 | 162004 |
| Kerala | Pathanamthitta | 5·645 | 7·036 | 25·748 | 1·445 | 6·932 | 96977 | 24644 | 119011 |
| Kerala | Thiruvananthapuram | 5·940 | 7·399 | 27·070 | 1·553 | 7·246 | 203262 | 52875 | 246181 |
| Kerala | Thrissur | 4·632 | 5·576 | 19·715 | 1·289 | 5·776 | 206357 | 57384 | 257000 |
| Kerala | Wayanad | 3·731 | 4·244 | 15·016 | 1·170 | 4·679 | 29310 | 9161 | 36704 |
| Ladakh | IND Region 1 | 4·961 | 6·829 | 24·453 | 1·110 | 5·831 | 1148 | 259 | 1384 |
| Ladakh | Kargil | 4·658 | 6·127 | 21·474 | 1·172 | 5·619 | 5810 | 1404 | 6964 |
| Ladakh | Leh(Ladakh) | 4·538 | 6·219 | 22·035 | 1·063 | 5·347 | 12483 | 2949 | 14852 |
| Madhya Pradesh | Agar | 5·042 | 4·358 | 16·047 | 2·162 | 6·460 | 31890 | 13716 | 40769 |
| Madhya Pradesh | Alirajpur | 3·558 | 2·516 | 9·311 | 1·856 | 4·498 | 41156 | 21463 | 52015 |
| Madhya Pradesh | Anuppur | 8·352 | 7·927 | 29·640 | 3·096 | 10·773 | 108259 | 39655 | 139412 |
| Madhya Pradesh | Ashoknagar | 9·184 | 6·862 | 25·824 | 4·386 | 11·925 | 90255 | 43098 | 117080 |
| Madhya Pradesh | Balaghat | 6·830 | 5·633 | 20·795 | 3·065 | 8·750 | 161681 | 73877 | 207102 |
| Madhya Pradesh | Barwani | 3·496 | 2·537 | 9·400 | 1·786 | 4·435 | 58246 | 29648 | 73797 |
| Madhya Pradesh | Betul | 5·675 | 4·259 | 15·874 | 2·789 | 7·243 | 124614 | 60993 | 159353 |
| Madhya Pradesh | Bhind | 6·802 | 6·030 | 22·313 | 2·833 | 8·708 | 90033 | 37574 | 115038 |
| Madhya Pradesh | Bhopal | 7·220 | 5·881 | 21·934 | 3·239 | 9·290 | 213895 | 95821 | 273555 |
| Madhya Pradesh | Burhanpur | 3·368 | 2·501 | 9·271 | 1·690 | 4·269 | 21169 | 10755 | 26739 |
| Madhya Pradesh | Chhatarpur | 9·510 | 7·300 | 27·344 | 4·410 | 12·420 | 188766 | 87412 | 245913 |
| Madhya Pradesh | Chhindwara | 7·870 | 5·825 | 21·877 | 3·832 | 10·135 | 209461 | 102508 | 269396 |

| | | β-thalassaemia carrier prevalence | | | | | Estimated carrier counts | | |
|---|---|---|---|---|---|---|---|---|---|
| **ADM1** | **ADM2** | **Mean** | **SD** | **CI95%** | **Q25** | **Q75** | **Mean** | **Q25** | **Q75** |
| Madhya Pradesh | Damoh | 10·370 | 8·003 | 30·109 | 4·748 | 13·576 | 141955 | 65228 | 186155 |
| Madhya Pradesh | Datia | 7·912 | 6·516 | 24·499 | 3·493 | 10·208 | 54975 | 24242 | 71019 |
| Madhya Pradesh | Dewas | 4·570 | 3·584 | 13·307 | 2·186 | 5·804 | 98104 | 47195 | 124569 |
| Madhya Pradesh | Dhar | 3·601 | 2·681 | 9·902 | 1·809 | 4·566 | 115853 | 58197 | 146637 |
| Madhya Pradesh | Dindori | 8·420 | 7·540 | 28·072 | 3·347 | 10·955 | 60511 | 23996 | 78782 |
| Madhya Pradesh | Guna | 7·661 | 6·386 | 23·943 | 3·322 | 9·919 | 108631 | 47536 | 140665 |
| Madhya Pradesh | Gwalior | 7·516 | 6·494 | 24·333 | 3·150 | 9·700 | 170346 | 70928 | 218942 |
| Madhya Pradesh | Harda | 4·879 | 3·805 | 14·158 | 2·335 | 6·215 | 32040 | 15313 | 40898 |
| Madhya Pradesh | Hoshangabad | 7·267 | 5·753 | 21·518 | 3·343 | 9·374 | 106814 | 48668 | 138542 |
| Madhya Pradesh | Indore | 3·823 | 2·822 | 10·474 | 1·931 | 4·854 | 161270 | 81120 | 204780 |
| Madhya Pradesh | Jabalpur | 9·689 | 7·912 | 29·723 | 4·228 | 12·638 | 275129 | 120068 | 359848 |
| Madhya Pradesh | Jhabua | 3·519 | 2·598 | 9·577 | 1·781 | 4·462 | 58427 | 29765 | 74100 |
| Madhya Pradesh | Katni | 9·654 | 8·086 | 30·253 | 4·073 | 12·632 | 144350 | 61150 | 189418 |
| Madhya Pradesh | Khandwa (East Nimar) | 3·879 | 2·939 | 10·883 | 1·907 | 4·929 | 57607 | 28332 | 73391 |
| Madhya Pradesh | Khargone (West Nimar) | 3·510 | 2·626 | 9·706 | 1·752 | 4·455 | 79233 | 39534 | 100538 |
| Madhya Pradesh | Mandla | 8·345 | 7·114 | 26·569 | 3·525 | 10·820 | 108683 | 45983 | 140476 |
| Madhya Pradesh | Mandsaur | 4·477 | 4·097 | 15·035 | 1·827 | 5·695 | 75883 | 31125 | 96614 |
| Madhya Pradesh | Morena | 6·723 | 6·251 | 22·990 | 2·609 | 8·639 | 113480 | 44290 | 145364 |
| Madhya Pradesh | Narsimhapur | 9·445 | 7·377 | 27·725 | 4·321 | 12·314 | 135191 | 61678 | 176100 |
| Madhya Pradesh | Neemuch | 4·374 | 4·161 | 15·281 | 1·713 | 5·568 | 46306 | 18441 | 58957 |
| Madhya Pradesh | Niwari | 9·274 | 6·957 | 26·163 | 4·427 | 12·048 | 51629 | 24618 | 67022 |
| Madhya Pradesh | Panna | 9·541 | 7·767 | 29·100 | 4·127 | 12·526 | 107121 | 46106 | 140677 |
| Madhya Pradesh | Raisen | 8·441 | 6·511 | 24·400 | 3·911 | 11·007 | 123278 | 57164 | 160212 |
| Madhya Pradesh | Rajgarh | 6·284 | 5·379 | 20·004 | 2·692 | 8·070 | 106341 | 45725 | 136655 |
| Madhya Pradesh | Ratlam | 3·815 | 3·147 | 11·507 | 1·748 | 4·852 | 76085 | 34996 | 96806 |
| Madhya Pradesh | Rewa | 8·048 | 7·492 | 27·998 | 3·056 | 10·421 | 278628 | 106622 | 361409 |
| Madhya Pradesh | Sagar | 10·216 | 7·407 | 27·838 | 4·958 | 13·352 | 228999 | 111305 | 299348 |
| Madhya Pradesh | Satna | 8·766 | 7·589 | 28·451 | 3·615 | 11·398 | 263327 | 108461 | 341906 |
| Madhya Pradesh | Sehore | 6·015 | 4·883 | 18·155 | 2·751 | 7·700 | 120612 | 55212 | 154120 |
| Madhya Pradesh | Seoni | 8·324 | 6·489 | 24·306 | 3·865 | 10·762 | 172443 | 79851 | 222429 |
| Madhya Pradesh | Shahdol | 8·780 | 8·169 | 30·588 | 3·327 | 11·364 | 172879 | 65568 | 223917 |
| Madhya Pradesh | Shajapur | 5·417 | 4·429 | 16·559 | 2·455 | 6·913 | 62232 | 28195 | 79310 |
| Madhya Pradesh | Sheopur | 6·985 | 6·605 | 24·573 | 2·642 | 8·984 | 44497 | 16661 | 57339 |
| Madhya Pradesh | Shivpuri | 8·394 | 6·848 | 25·669 | 3·690 | 10·895 | 165156 | 73375 | 214298 |
| Madhya Pradesh | Sidhi | 8·500 | 8·085 | 30·374 | 3·135 | 10·995 | 172022 | 63415 | 223091 |
| Madhya Pradesh | Singrauli | 8·401 | 8·298 | 31·108 | 2·954 | 10·776 | 254311 | 89904 | 325950 |
| Madhya Pradesh | Tikamgarh | 9·905 | 7·056 | 26·649 | 4·912 | 12·884 | 114866 | 56997 | 149376 |
| Madhya Pradesh | Ujjain | 4·203 | 3·411 | 12·498 | 1·959 | 5·344 | 119474 | 56046 | 151883 |
| Madhya Pradesh | Umaria | 9·133 | 8·135 | 30·571 | 3·627 | 11·890 | 81173 | 32277 | 105381 |
| Madhya Pradesh | Vidisha | 8·709 | 6·639 | 24·949 | 4·069 | 11·342 | 141669 | 65876 | 184438 |
| Maharashtra | Ahmadnagar | 4·020 | 2·699 | 10·061 | 2·170 | 5·089 | 429408 | 232363 | 543765 |
| Maharashtra | Akola | 3·178 | 2·208 | 8·205 | 1·683 | 4·023 | 51255 | 27006 | 64994 |
| Maharashtra | Amravati | 4·008 | 2·837 | 10·553 | 2·090 | 5·079 | 116967 | 61700 | 147870 |
| Maharashtra | Aurangabad | 3·616 | 2·463 | 9·192 | 1·939 | 4·576 | 182129 | 97911 | 231067 |
| Maharashtra | Bhandara | 5·223 | 3·706 | 13·862 | 2·701 | 6·651 | 85398 | 44316 | 108781 |
| Maharashtra | Bid | 3·467 | 2·321 | 8·639 | 1·882 | 4·392 | 121837 | 66021 | 154341 |
| Maharashtra | Buldana | 3·119 | 2·163 | 8·047 | 1·654 | 3·939 | 84078 | 44727 | 106116 |
| Maharashtra | Chandrapur | 3·387 | 2·429 | 9·029 | 1·752 | 4·287 | 91586 | 47346 | 115882 |
| Maharashtra | Dhule | 3·641 | 2·518 | 9·392 | 1·937 | 4·603 | 92601 | 49454 | 117372 |
| Maharashtra | Gadchiroli | 2·872 | 2·347 | 8·577 | 1·341 | 3·631 | 40903 | 19816 | 51734 |
| Maharashtra | Gondiya | 5·042 | 3·863 | 14·424 | 2·434 | 6·459 | 91539 | 43779 | 117560 |
| Maharashtra | Hingoli | 2·980 | 2·083 | 7·739 | 1·585 | 3·756 | 32308 | 17223 | 40720 |
| Maharashtra | Jalgaon | 3·316 | 2·342 | 8·711 | 1·736 | 4·199 | 129904 | 68312 | 164853 |
| Maharashtra | Jalna | 3·243 | 2·207 | 8·187 | 1·751 | 4·099 | 81106 | 43605 | 102619 |
| Maharashtra | Kolhapur | 2·334 | 1·668 | 6·164 | 1·214 | 2·953 | 93087 | 49034 | 117647 |
| Maharashtra | Latur | 2·705 | 1·970 | 7·267 | 1·382 | 3·432 | 73590 | 37733 | 93292 |

| ADM1 | ADM2 | β-thalassaemia carrier prevalence Mean | SD | CI95% | Q25 | Q75 | Estimated carrier counts Mean | Q25 | Q75 |
|---|---|---|---|---|---|---|---|---|---|
| Maharashtra | Mumbai | 3·533 | 2·517 | 8·681 | 1·951 | 4·410 | 13645 | 7536 | 17032 |
| Maharashtra | Mumbai Suburban | 3·594 | 2·319 | 8·688 | 2·012 | 4·524 | 94480 | 53001 | 118556 |
| Maharashtra | Nagpur | 6·213 | 4·160 | 15·657 | 3·331 | 7·925 | 389275 | 209337 | 495843 |
| Maharashtra | Nanded | 2·785 | 2·046 | 7·578 | 1·418 | 3·526 | 72546 | 37159 | 91768 |
| Maharashtra | Nandurbar | 3·609 | 2·488 | 9·309 | 1·921 | 4·562 | 64365 | 34391 | 81538 |
| Maharashtra | Nashik | 3·980 | 2·687 | 10·013 | 2·150 | 5·037 | 460840 | 249532 | 582763 |
| Maharashtra | Osmanabad | 2·870 | 2·054 | 7·591 | 1·484 | 3·644 | 68019 | 35166 | 86348 |
| Maharashtra | Palghar | 3·967 | 2·656 | 9·937 | 2·154 | 5·017 | 128137 | 70040 | 161800 |
| Maharashtra | Parbhani | 3·022 | 2·050 | 7·631 | 1·630 | 3·818 | 51537 | 27801 | 65104 |
| Maharashtra | Pune | 3·672 | 2·476 | 9·226 | 1·984 | 4·648 | 553751 | 300415 | 699870 |
| Maharashtra | Raigarh | 3·614 | 2·405 | 8·977 | 1·974 | 4·567 | 117247 | 65143 | 147612 |
| Maharashtra | Ratnagiri | 2·909 | 2·050 | 7·599 | 1·526 | 3·677 | 66776 | 34941 | 84325 |
| Maharashtra | Sangli | 2·702 | 1·916 | 7·108 | 1·416 | 3·416 | 123940 | 65481 | 156678 |
| Maharashtra | Satara | 3·235 | 2·198 | 8·219 | 1·740 | 4·096 | 160962 | 86608 | 203944 |
| Maharashtra | Sindhudurg | 2·141 | 1·662 | 6·041 | 1·061 | 2·693 | 29477 | 14572 | 37054 |
| Maharashtra | Solapur | 2·853 | 2·057 | 7·619 | 1·474 | 3·608 | 218679 | 113281 | 276910 |
| Maharashtra | Thane | 3·928 | 2·553 | 9·520 | 2·176 | 4·956 | 189133 | 105664 | 238272 |
| Maharashtra | Wardha | 4·695 | 3·243 | 12·133 | 2·477 | 5·971 | 71273 | 37454 | 90294 |
| Maharashtra | Washim | 3·110 | 2·189 | 8·099 | 1·642 | 3·917 | 31838 | 16843 | 40072 |
| Maharashtra | Yavatmal | 3·202 | 2·256 | 8·392 | 1·681 | 4·050 | 81021 | 42624 | 102570 |
| Manipur | Bishnupur | 1·182 | 1·038 | 3·742 | 0·511 | 1·486 | 3459 | 1500 | 4349 |
| Manipur | Chandel | 1·139 | 1·136 | 4·045 | 0·447 | 1·422 | 611 | 248 | 771 |
| Manipur | Churachandpur | 1·192 | 1·122 | 4·010 | 0·495 | 1·489 | 2726 | 1132 | 3436 |
| Manipur | Imphal East | 1·242 | 1·054 | 3·807 | 0·572 | 1·557 | 10166 | 4726 | 12786 |
| Manipur | Imphal West | 1·266 | 1·095 | 3·941 | 0·578 | 1·587 | 8302 | 3782 | 10462 |
| Manipur | Jiribam | 1·506 | 1·267 | 4·798 | 0·671 | 1·903 | 864 | 388 | 1093 |
| Manipur | Kakching | 1·132 | 1·039 | 3·648 | 0·495 | 1·401 | 1928 | 853 | 2391 |
| Manipur | Kamjong | 1·424 | 1·284 | 4·603 | 0·617 | 1·791 | 455 | 200 | 576 |
| Manipur | Kangpokpi | 1·539 | 1·269 | 4·571 | 0·719 | 1·941 | 2730 | 1265 | 3444 |
| Manipur | Noney | 1·348 | 1·178 | 4·242 | 0·605 | 1·685 | 526 | 236 | 666 |
| Manipur | Pherzawl | 1·310 | 1·210 | 4·272 | 0·562 | 1·639 | 303 | 128 | 373 |
| Manipur | Senapati | 2·219 | 1·810 | 6·581 | 1·051 | 2·793 | 2529 | 1179 | 3183 |
| Manipur | Tamenglong | 1·685 | 1·404 | 5·057 | 0·782 | 2·127 | 1563 | 718 | 1970 |
| Manipur | Tengnoupal | 1·174 | 1·121 | 3·951 | 0·484 | 1·463 | 976 | 412 | 1239 |
| Manipur | Thoubal | 1·176 | 1·041 | 3·855 | 0·515 | 1·481 | 4466 | 1955 | 5600 |
| Manipur | Ukhrul | 1·956 | 1·674 | 5·990 | 0·895 | 2·451 | 2189 | 1009 | 2761 |
| Meghalaya | East Garo Hills | 4·707 | 4·229 | 15·338 | 1·976 | 5·972 | 11422 | 4766 | 14509 |
| Meghalaya | East Jaintia Hills | 2·536 | 2·038 | 7·479 | 1·194 | 3·222 | 6436 | 3014 | 8170 |
| Meghalaya | East Khasi Hills | 3·194 | 2·623 | 9·627 | 1·470 | 4·061 | 28744 | 13049 | 36863 |
| Meghalaya | North Garo Hills | 4·921 | 4·430 | 16·348 | 2·051 | 6·243 | 11763 | 4908 | 14948 |
| Meghalaya | Ribhoi | 4·153 | 3·536 | 13·069 | 1·814 | 5·313 | 17625 | 7635 | 22601 |
| Meghalaya | South Garo Hills | 4·370 | 3·905 | 14·271 | 1·865 | 5·524 | 8200 | 3524 | 10414 |
| Meghalaya | South West Garo Hills | 5·162 | 4·551 | 16·903 | 2·212 | 6·564 | 8265 | 3569 | 10494 |
| Meghalaya | South West Khasi Hills | 3·630 | 3·176 | 11·555 | 1·583 | 4·598 | 6248 | 2737 | 7923 |
| Meghalaya | West Garo Hills | 5·068 | 4·457 | 16·298 | 2·160 | 6·455 | 27422 | 11719 | 35025 |
| Meghalaya | West Jaintia Hills | 3·008 | 2·424 | 8·907 | 1·406 | 3·829 | 9783 | 4566 | 12446 |
| Meghalaya | West Khasi Hills | 4·047 | 3·524 | 12·894 | 1·748 | 5·163 | 13356 | 5768 | 17048 |
| Mizoram | Aizawl | 1·504 | 1·569 | 5·404 | 0·588 | 1·849 | 5679 | 2197 | 7055 |
| Mizoram | Champhai | 1·438 | 1·641 | 5·457 | 0·511 | 1·765 | 1136 | 396 | 1374 |
| Mizoram | Hnahthial | 1·759 | 2·128 | 7·113 | 0·587 | 2·113 | 519 | 173 | 622 |
| Mizoram | Khawzawl | 1·438 | 1·634 | 5·309 | 0·514 | 1·770 | 580 | 209 | 721 |
| Mizoram | Kolasib | 1·548 | 1·522 | 5·249 | 0·636 | 1·916 | 2769 | 1130 | 3393 |
| Mizoram | Lawngtlai | 2·213 | 2·703 | 9·275 | 0·687 | 2·682 | 3375 | 1056 | 4069 |
| Mizoram | Lunglei | 1·897 | 2·235 | 7·522 | 0·643 | 2·302 | 2766 | 919 | 3329 |
| Mizoram | Mamit | 1·722 | 1·808 | 6·295 | 0·667 | 2·109 | 2006 | 786 | 2466 |
| Mizoram | Saiha | 2·242 | 2·829 | 9·487 | 0·679 | 2·703 | 1262 | 387 | 1528 |

| ADM1 | ADM2 | β-thalassaemia carrier prevalence Mean | SD | CI95% | Q25 | Q75 | Estimated carrier counts Mean | Q25 | Q75 |
|---|---|---|---|---|---|---|---|---|---|
| Mizoram | Saitual | 1·327 | 1·337 | 4·715 | 0·521 | 1·652 | 401 | 156 | 501 |
| Mizoram | Serchhip | 1·603 | 1·837 | 6·053 | 0·564 | 1·964 | 1277 | 452 | 1560 |
| Nagaland | Dimapur | 3·382 | 2·577 | 9·515 | 1·671 | 4·264 | 27043 | 13712 | 34231 |
| Nagaland | Kiphire | 3·612 | 2·857 | 10·568 | 1·726 | 4·592 | 1998 | 960 | 2541 |
| Nagaland | Kohima | 3·284 | 2·469 | 9·047 | 1·645 | 4·145 | 8783 | 4329 | 11025 |
| Nagaland | Longleng | 6·042 | 3·980 | 14·983 | 3·268 | 7·666 | 2347 | 1263 | 2984 |
| Nagaland | Mokokchung | 6·048 | 3·914 | 14·735 | 3·324 | 7·708 | 12051 | 6566 | 15406 |
| Nagaland | Mon | 6·035 | 3·987 | 14·911 | 3·252 | 7·692 | 12253 | 6629 | 15668 |
| Nagaland | Peren | 2·542 | 1·969 | 7·208 | 1·250 | 3·207 | 3725 | 1841 | 4729 |
| Nagaland | Phek | 2·988 | 2·441 | 8·841 | 1·412 | 3·762 | 3916 | 1883 | 4908 |
| Nagaland | Tuensang | 4·778 | 3·517 | 13·024 | 2·395 | 6·089 | 5701 | 2885 | 7303 |
| Nagaland | Wokha | 4·934 | 3·292 | 12·316 | 2·669 | 6·238 | 5497 | 2968 | 6909 |
| Nagaland | Zunheboto | 4·151 | 3·019 | 11·274 | 2·126 | 5·254 | 5023 | 2587 | 6347 |
| Odisha | Anugul | 8·573 | 8·438 | 31·428 | 2·998 | 11·079 | 212916 | 74474 | 276007 |
| Odisha | Balangir | 6·090 | 6·686 | 24·327 | 1·886 | 7·747 | 122437 | 37990 | 155697 |
| Odisha | Baleshwar | 9·632 | 7·494 | 28·345 | 4·453 | 12·452 | 211640 | 98628 | 273764 |
| Odisha | Bargarh | 6·972 | 7·357 | 26·505 | 2·304 | 8·921 | 115487 | 38338 | 147641 |
| Odisha | Baudh | 7·360 | 7·672 | 27·936 | 2·353 | 9·544 | 40649 | 12989 | 52767 |
| Odisha | Bhadrak | 9·486 | 8·342 | 31·306 | 3·828 | 12·307 | 130027 | 52600 | 169092 |
| Odisha | Cuttack | 8·772 | 8·936 | 32·971 | 2·940 | 11·313 | 228014 | 76599 | 293709 |
| Odisha | Debagarh | 8·849 | 8·411 | 31·136 | 3·245 | 11·523 | 47492 | 17365 | 61927 |
| Odisha | Dhenkanal | 9·059 | 8·943 | 32·981 | 3·182 | 11·714 | 115656 | 40437 | 149724 |
| Odisha | Gajapati | 6·281 | 7·562 | 27·555 | 1·685 | 7·800 | 28659 | 7648 | 35503 |
| Odisha | Ganjam | 7·138 | 8·118 | 29·733 | 2·059 | 9·042 | 213776 | 60937 | 268956 |
| Odisha | Jagatsinghapur | 8·867 | 8·941 | 33·161 | 3·008 | 11·414 | 89366 | 30578 | 115049 |
| Odisha | Jajapur | 9·400 | 8·896 | 33·138 | 3·507 | 12·140 | 157001 | 58443 | 201917 |
| Odisha | Jharsuguda | 8·286 | 8·160 | 29·429 | 2·956 | 10·776 | 96371 | 34534 | 125286 |
| Odisha | Kalahandi | 5·356 | 6·209 | 22·598 | 1·568 | 6·669 | 92268 | 27201 | 114831 |
| Odisha | Kandhamal | 6·763 | 7·456 | 27·244 | 2·041 | 8·641 | 51314 | 15628 | 65802 |
| Odisha | Kendrapara | 9·129 | 8·682 | 32·401 | 3·365 | 11·843 | 110189 | 40550 | 142760 |
| Odisha | Kendujhar | 9·646 | 8·491 | 31·770 | 3·907 | 12·513 | 250174 | 102953 | 324859 |
| Odisha | Khordha | 8·274 | 8·780 | 32·112 | 2·610 | 10·687 | 276795 | 89724 | 357416 |
| Odisha | Koraput | 4·395 | 5·361 | 19·432 | 1·208 | 5·414 | 53775 | 14907 | 66017 |
| Odisha | Malkangiri | 3·643 | 4·392 | 15·990 | 1·017 | 4·503 | 30706 | 8652 | 38038 |
| Odisha | Mayurbhanj | 10·038 | 7·751 | 29·114 | 4·658 | 13·023 | 295788 | 139127 | 383379 |
| Odisha | Nabarangapur | 4·185 | 4·804 | 17·262 | 1·271 | 5·209 | 51692 | 15707 | 64390 |
| Odisha | Nayagarh | 7·929 | 8·390 | 30·668 | 2·492 | 10·198 | 68071 | 21513 | 87471 |
| Odisha | Nuapada | 5·248 | 5·772 | 21·036 | 1·663 | 6·608 | 45677 | 14429 | 57643 |
| Odisha | Puri | 8·275 | 8·755 | 32·140 | 2·599 | 10·730 | 146858 | 46353 | 190395 |
| Odisha | Rayagada | 5·576 | 6·714 | 24·261 | 1·521 | 6·917 | 51179 | 13814 | 63264 |
| Odisha | Sambalpur | 8·306 | 8·167 | 29·903 | 2·937 | 10·809 | 161125 | 56884 | 210617 |
| Odisha | Subarnapur | 7·206 | 7·587 | 27·418 | 2·351 | 9·241 | 47047 | 15281 | 60468 |
| Odisha | Sundargarh | 8·925 | 8·214 | 30·336 | 3·433 | 11·597 | 332195 | 129026 | 431592 |
| Puducherry | Karaikal | 5·402 | 6·683 | 22·543 | 1·573 | 6·480 | 13205 | 3815 | 15927 |
| Puducherry | Mahe | 3·885 | 4·573 | 15·843 | 1·141 | 4·888 | 8682 | 2543 | 10978 |
| Puducherry | Puducherry | 4·790 | 5·821 | 20·583 | 1·430 | 5·776 | 45931 | 13678 | 54669 |
| Punjab | Amritsar | 4·376 | 4·398 | 15·978 | 1·630 | 5·512 | 96590 | 36278 | 121806 |
| Punjab | Barnala | 4·277 | 3·795 | 13·773 | 1·875 | 5·375 | 34862 | 15293 | 43748 |
| Punjab | Bathinda | 4·595 | 4·512 | 16·352 | 1·762 | 5·803 | 92308 | 35428 | 116641 |
| Punjab | Faridkot | 4·500 | 4·283 | 15·527 | 1·761 | 5·724 | 33332 | 13109 | 42372 |
| Punjab | Fatehgarh Sahib | 3·801 | 2·920 | 10·739 | 1·856 | 4·814 | 27016 | 13249 | 34173 |
| Punjab | Fazilka | 4·856 | 5·196 | 18·818 | 1·630 | 6·105 | 72148 | 24260 | 90706 |
| Punjab | Firozpur | 4·402 | 4·136 | 15·049 | 1·752 | 5·599 | 40614 | 16156 | 51487 |
| Punjab | Gurdaspur | 4·258 | 4·254 | 15·334 | 1·606 | 5·368 | 68251 | 25842 | 86077 |
| Punjab | Hoshiarpur | 3·979 | 3·498 | 12·752 | 1·712 | 5·062 | 70390 | 30423 | 89394 |
| Punjab | Jalandhar | 4·099 | 3·401 | 12·560 | 1·872 | 5·205 | 100963 | 45961 | 127711 |
| Punjab | Kapurthala | 4·158 | 3·636 | 13·298 | 1·801 | 5·294 | 43031 | 18936 | 55095 |

| ADM1 | ADM2 | β-thalassaemia carrier prevalence Mean | SD | CI95% | Q25 | Q75 | Estimated carrier counts Mean | Q25 | Q75 |
|---|---|---|---|---|---|---|---|---|---|
| Punjab | Ludhiana | 4·017 | 3·145 | 11·608 | 1·943 | 5·086 | 161609 | 78296 | 204247 |
| Punjab | Mansa | 4·585 | 4·523 | 16·134 | 1·769 | 5·763 | 44352 | 17142 | 55977 |
| Punjab | Moga | 4·318 | 3·854 | 13·909 | 1·847 | 5·484 | 51645 | 22086 | 65487 |
| Punjab | Muktsar | 4·733 | 4·836 | 17·514 | 1·681 | 6·004 | 61002 | 21550 | 77152 |
| Punjab | Pathankot | 4·213 | 4·372 | 15·840 | 1·497 | 5·298 | 30533 | 10918 | 38449 |
| Punjab | Patiala | 3·961 | 3·287 | 12·050 | 1·826 | 4·998 | 88215 | 40798 | 111457 |
| Punjab | Rupnagar | 3·722 | 2·910 | 10·717 | 1·787 | 4·724 | 33877 | 16237 | 42981 |
| Punjab | Sahibzada Ajit Singh Nagar | 3·631 | 2·821 | 10·326 | 1·747 | 4·611 | 52804 | 25490 | 66825 |
| Punjab | Sangrur | 4·268 | 3·768 | 13·751 | 1·866 | 5·382 | 81922 | 36052 | 103436 |
| Punjab | Shahid Bhagat Singh Nagar | 3·841 | 2·963 | 10·952 | 1·876 | 4·870 | 27435 | 13396 | 34719 |
| Punjab | Tarn Taran | 4·373 | 4·141 | 15·085 | 1·725 | 5·554 | 57916 | 22794 | 73479 |
| Rajasthan | Ajmer | 4·881 | 5·232 | 18·944 | 1·586 | 6·210 | 150616 | 48897 | 192117 |
| Rajasthan | Alwar | 5·399 | 5·773 | 20·968 | 1·823 | 6·788 | 213683 | 72162 | 268964 |
| Rajasthan | Banswara | 3·386 | 2·696 | 9·923 | 1·601 | 4·301 | 65460 | 31006 | 83138 |
| Rajasthan | Baran | 6·816 | 6·130 | 22·740 | 2·709 | 8·812 | 78452 | 31039 | 101354 |
| Rajasthan | Barmer | 4·717 | 4·882 | 17·586 | 1·663 | 5·949 | 163766 | 58674 | 205958 |
| Rajasthan | Bharatpur | 5·713 | 5·847 | 21·304 | 2·013 | 7·254 | 103935 | 36750 | 132409 |
| Rajasthan | Bhilwara | 4·569 | 4·710 | 17·245 | 1·587 | 5·797 | 116646 | 40591 | 148072 |
| Rajasthan | Bikaner | 5·342 | 6·390 | 23·201 | 1·451 | 6·662 | 203955 | 55963 | 254778 |
| Rajasthan | Bundi | 5·538 | 5·545 | 20·412 | 1·955 | 7·105 | 78703 | 27743 | 101070 |
| Rajasthan | Chittaurgarh | 4·170 | 4·097 | 14·839 | 1·597 | 5·282 | 73353 | 28267 | 92632 |
| Rajasthan | Churu | 5·230 | 5·834 | 21·218 | 1·598 | 6·593 | 139981 | 42868 | 175954 |
| Rajasthan | Dausa | 5·817 | 6·198 | 22·657 | 1·976 | 7·308 | 104829 | 35619 | 131444 |
| Rajasthan | Dhaulpur | 6·363 | 6·126 | 22·453 | 2·364 | 8·190 | 51814 | 19244 | 66426 |
| Rajasthan | Dungarpur | 3·058 | 2·429 | 8·961 | 1·460 | 3·875 | 51763 | 24666 | 65486 |
| Rajasthan | Ganganagar | 5·298 | 6·298 | 23·129 | 1·528 | 6·488 | 154530 | 44911 | 188795 |
| Rajasthan | Hanumangarh | 5·155 | 5·711 | 20·941 | 1·639 | 6·424 | 125917 | 40093 | 156573 |
| Rajasthan | Jaipur | 5·423 | 5·944 | 21·333 | 1·738 | 6·858 | 491097 | 156242 | 620857 |
| Rajasthan | Jaisalmer | 5·515 | 6·313 | 22·864 | 1·557 | 7·028 | 52638 | 14968 | 66855 |
| Rajasthan | Jalor | 3·834 | 3·578 | 12·902 | 1·567 | 4·836 | 90733 | 37648 | 114420 |
| Rajasthan | Jhalawar | 5·611 | 5·041 | 18·741 | 2·279 | 7·197 | 72650 | 29336 | 93123 |
| Rajasthan | Jhunjhunun | 5·248 | 5·776 | 20·968 | 1·673 | 6·576 | 173772 | 55296 | 217482 |
| Rajasthan | Jodhpur | 4·813 | 5·589 | 20·135 | 1·415 | 5·999 | 269907 | 82216 | 336469 |
| Rajasthan | Karauli | 6·287 | 6·371 | 23·289 | 2·241 | 7·988 | 67827 | 24001 | 85841 |
| Rajasthan | Kota | 5·815 | 5·570 | 20·549 | 2·174 | 7·467 | 115955 | 42678 | 148177 |
| Rajasthan | Nagaur | 4·975 | 5·582 | 20·226 | 1·500 | 6·293 | 252604 | 76463 | 318907 |
| Rajasthan | Pali | 4·072 | 4·184 | 15·183 | 1·415 | 5·178 | 118035 | 40858 | 150234 |
| Rajasthan | Pratapgarh | 3·725 | 3·297 | 12·002 | 1·600 | 4·732 | 36170 | 15547 | 45901 |
| Rajasthan | Rajsamand | 3·868 | 3·892 | 14·086 | 1·410 | 4·915 | 61688 | 22501 | 78549 |
| Rajasthan | Sawai Madhopur | 6·114 | 6·250 | 22·519 | 2·146 | 7·795 | 81155 | 28334 | 102990 |
| Rajasthan | Sikar | 5·315 | 5·915 | 21·423 | 1·633 | 6·702 | 224386 | 68471 | 282295 |
| Rajasthan | Sirohi | 3·231 | 2·820 | 10·253 | 1·394 | 4·111 | 40589 | 17738 | 51575 |
| Rajasthan | Tonk | 5·469 | 5·763 | 20·894 | 1·859 | 6·927 | 83955 | 28594 | 106200 |
| Rajasthan | Udaipur | 3·179 | 2·825 | 10·269 | 1·366 | 4·025 | 115609 | 49572 | 146416 |
| Sikkim | East District | 6·733 | 5·862 | 21·890 | 2·815 | 8·696 | 19409 | 8249 | 24862 |
| Sikkim | North District | 6·945 | 6·672 | 24·785 | 2·590 | 8·888 | 3840 | 1495 | 4938 |
| Sikkim | South District | 6·726 | 5·935 | 21·892 | 2·811 | 8·653 | 14116 | 6027 | 18222 |
| Sikkim | West District | 6·806 | 6·188 | 22·587 | 2·710 | 8·799 | 9904 | 4071 | 12821 |
| Tamil Nadu | Ariyalur | 4·929 | 5·715 | 20·533 | 1·503 | 6·065 | 45770 | 14025 | 56237 |
| Tamil Nadu | Chengalputtu | 4·218 | 4·977 | 17·865 | 1·281 | 5·158 | 71751 | 21521 | 88307 |
| Tamil Nadu | Chennai | 4·366 | 5·254 | 18·908 | 1·223 | 5·379 | 73109 | 20469 | 89998 |
| Tamil Nadu | Coimbatore | 4·346 | 4·937 | 17·861 | 1·316 | 5·458 | 225810 | 69510 | 283396 |
| Tamil Nadu | Cuddalore | 4·771 | 5·588 | 20·230 | 1·469 | 5·821 | 115145 | 35272 | 139859 |
| Tamil Nadu | Dharmapuri | 3·613 | 3·562 | 12·851 | 1·359 | 4·586 | 70441 | 26909 | 89591 |
| Tamil Nadu | Dindigul | 4·919 | 5·433 | 19·904 | 1·468 | 6·281 | 127062 | 38200 | 162247 |
| Tamil Nadu | Erode | 3·853 | 4·095 | 14·911 | 1·321 | 4·828 | 153086 | 51611 | 191667 |

| | | β-thalassaemia carrier prevalence | | | | | Estimated carrier counts | | |
|---|---|---|---|---|---|---|---|---|---|
| ADM1 | ADM2 | Mean | SD | CI95% | Q25 | Q75 | Mean | Q25 | Q75 |
| Tamil Nadu | Kallakurichi | 4·367 | 4·796 | 17·309 | 1·426 | 5·451 | 51404 | 16689 | 63843 |
| Tamil Nadu | Kancheepuram | 4·419 | 5·389 | 19·381 | 1·286 | 5·332 | 150138 | 42798 | 181273 |
| Tamil Nadu | Kanniyakumari | 6·032 | 7·328 | 26·544 | 1·678 | 7·434 | 92498 | 25553 | 113822 |
| Tamil Nadu | Karur | 4·702 | 5·134 | 18·859 | 1·470 | 5·941 | 78367 | 24608 | 98926 |
| Tamil Nadu | Krishnagiri | 3·258 | 2·972 | 10·716 | 1·368 | 4·131 | 77411 | 32766 | 98067 |
| Tamil Nadu | Madurai | 5·374 | 6·080 | 22·094 | 1·568 | 6·806 | 130529 | 38679 | 164375 |
| Tamil Nadu | Mayiladuthurai | 5·043 | 5·921 | 21·267 | 1·529 | 6·238 | 38190 | 11642 | 47374 |
| Tamil Nadu | Nagapattinam | 5·391 | 6·292 | 22·512 | 1·638 | 6·676 | 50053 | 15203 | 61877 |
| Tamil Nadu | Namakkal | 4·245 | 4·501 | 16·427 | 1·424 | 5·370 | 127981 | 42837 | 161481 |
| Tamil Nadu | Perambalur | 4·685 | 5·223 | 18·953 | 1·476 | 5·833 | 31081 | 9792 | 38682 |
| Tamil Nadu | Pudukkottai | 5·394 | 6·145 | 22·334 | 1·642 | 6·716 | 123412 | 37707 | 153808 |
| Tamil Nadu | Ramanathapuram | 5·847 | 6·495 | 23·526 | 1·784 | 7·395 | 66246 | 20385 | 83849 |
| Tamil Nadu | Ranipet | 3·953 | 4·480 | 16·007 | 1·267 | 4·902 | 47784 | 15437 | 59243 |
| Tamil Nadu | Salem | 3·989 | 4·144 | 15·017 | 1·396 | 5·030 | 165221 | 58181 | 208532 |
| Tamil Nadu | Sivaganga | 5·582 | 6·264 | 22·839 | 1·669 | 7·054 | 89237 | 26682 | 112735 |
| Tamil Nadu | Tenkasi | 5·846 | 7·071 | 25·645 | 1·578 | 7·214 | 83544 | 22409 | 103186 |
| Tamil Nadu | Thanjavur | 5·185 | 5·938 | 21·236 | 1·603 | 6·431 | 126481 | 39060 | 156877 |
| Tamil Nadu | The Nilgiris | 3·860 | 4·297 | 15·362 | 1·233 | 4·828 | 22795 | 7269 | 28636 |
| Tamil Nadu | Theni | 5·264 | 6·142 | 22·412 | 1·455 | 6·695 | 47564 | 13108 | 60490 |
| Tamil Nadu | Thiruvallur | 4·147 | 4·952 | 17·695 | 1·208 | 5·108 | 166356 | 47733 | 203510 |
| Tamil Nadu | Thiruvarur | 5·240 | 6·062 | 21·523 | 1·615 | 6·486 | 66342 | 20477 | 82072 |
| Tamil Nadu | Thoothukkudi | 5·987 | 6·910 | 24·675 | 1·755 | 7·544 | 105937 | 31220 | 132856 |
| Tamil Nadu | Tiruchirappalli | 4·859 | 5·362 | 19·652 | 1·525 | 6·090 | 135558 | 42386 | 169652 |
| Tamil Nadu | Tirunelveli | 5·988 | 7·081 | 25·319 | 1·695 | 7·499 | 112178 | 32117 | 141201 |
| Tamil Nadu | Tirupathur | 3·596 | 3·603 | 12·968 | 1·364 | 4·495 | 41045 | 15895 | 51535 |
| Tamil Nadu | Tiruppur | 4·455 | 4·978 | 18·142 | 1·369 | 5·606 | 218292 | 68372 | 274128 |
| Tamil Nadu | Tiruvannamalai | 4·049 | 4·434 | 15·976 | 1·359 | 5·034 | 99323 | 32941 | 123455 |
| Tamil Nadu | Vellore | 3·608 | 3·796 | 13·673 | 1·320 | 4·471 | 45623 | 16372 | 56525 |
| Tamil Nadu | Viluppuram | 4·419 | 5·133 | 18·473 | 1·390 | 5·380 | 103407 | 32469 | 125891 |
| Tamil Nadu | Virudhunagar | 5·720 | 6·695 | 24·325 | 1·623 | 7·167 | 121002 | 33980 | 151459 |
| Telangana | Adilabad | 2·847 | 2·170 | 8·029 | 1·413 | 3·593 | 18451 | 9233 | 23311 |
| Telangana | Bhadradri | 3·157 | 3·755 | 13·356 | 0·935 | 3·885 | 53132 | 15695 | 65426 |
| Telangana | Hydrabad | 2·743 | 2·870 | 10·118 | 0·999 | 3·430 | 115820 | 42087 | 144703 |
| Telangana | Jagtial | 2·616 | 2·283 | 8·160 | 1·160 | 3·306 | 36397 | 16103 | 45982 |
| Telangana | Jangaon | 2·746 | 2·898 | 10·321 | 0·984 | 3·416 | 20633 | 7434 | 25755 |
| Telangana | Jayashankar | 2·608 | 2·643 | 9·456 | 0·985 | 3·258 | 11921 | 4517 | 14846 |
| Telangana | Jogulamba | 2·873 | 3·249 | 11·378 | 0·951 | 3·567 | 15270 | 5057 | 18980 |
| Telangana | Kamareddy | 2·627 | 2·262 | 8·170 | 1·160 | 3·339 | 23429 | 10292 | 29733 |
| Telangana | Karimnagar | 2·597 | 2·513 | 8·832 | 1·044 | 3·273 | 40477 | 16409 | 51303 |
| Telangana | Khammam | 3·161 | 3·759 | 13·413 | 0·924 | 3·898 | 54568 | 16251 | 67753 |
| Telangana | Komaram Bheem | 2·633 | 2·124 | 7·760 | 1·241 | 3·337 | 17767 | 8395 | 22568 |
| Telangana | Mahabubabad | 2·924 | 3·303 | 11·856 | 0·936 | 3·627 | 26656 | 8464 | 33096 |
| Telangana | Mahabubnagar | 2·795 | 3·030 | 10·419 | 0·984 | 3·492 | 30017 | 10478 | 37499 |
| Telangana | Mancherial | 2·548 | 2·223 | 8·107 | 1·120 | 3·223 | 27350 | 11919 | 34593 |
| Telangana | Medak | 2·649 | 2·465 | 8·906 | 1·084 | 3·351 | 23024 | 9318 | 29027 |
| Telangana | Medchal | 2·736 | 2·800 | 9·968 | 1·011 | 3·439 | 88852 | 32603 | 111574 |
| Telangana | Mulugu | 2·755 | 2·998 | 10·625 | 0·963 | 3·399 | 8035 | 2816 | 9925 |
| Telangana | Nagarkurnool | 3·028 | 3·551 | 12·463 | 0·953 | 3·714 | 26510 | 8607 | 32694 |
| Telangana | Nalgonda | 2·979 | 3·421 | 12·176 | 0·957 | 3·665 | 65238 | 20951 | 79928 |
| Telangana | Narayanpet | 2·706 | 2·852 | 9·851 | 0·987 | 3·391 | 14153 | 5141 | 17771 |
| Telangana | Nirmal | 2·730 | 2·140 | 7·929 | 1·317 | 3·458 | 20917 | 10156 | 26541 |
| Telangana | Nizamabad | 2·654 | 2·196 | 7·989 | 1·223 | 3·368 | 43066 | 20030 | 54580 |
| Telangana | Peddapalli | 2·575 | 2·420 | 8·635 | 1·056 | 3·255 | 31449 | 12901 | 39480 |
| Telangana | Rajanna Sircilla | 2·605 | 2·406 | 8·523 | 1·095 | 3·279 | 17967 | 7510 | 22648 |
| Telangana | Rangareddy | 2·800 | 2·990 | 10·498 | 0·993 | 3·508 | 111441 | 40039 | 140220 |
| Telangana | Sangareddy | 2·639 | 2·446 | 8·733 | 1·084 | 3·352 | 53466 | 20925 | 67734 |
| Telangana | Siddipet | 2·659 | 2·622 | 9·212 | 1·031 | 3·365 | 37442 | 14518 | 47517 |

| ADM1 | ADM2 | β-thalassaemia carrier prevalence Mean | SD | CI95% | Q25 | Q75 | Estimated carrier counts Mean | Q25 | Q75 |
|---|---|---|---|---|---|---|---|---|---|
| Telangana | Suryapet | 3·029 | 3·483 | 12·467 | 0·945 | 3·751 | 40836 | 12682 | 50281 |
| Telangana | Vikarabad | 2·663 | 2·628 | 9·203 | 1·027 | 3·389 | 28558 | 11047 | 36533 |
| Telangana | Wanaparthy | 2·872 | 3·222 | 11·153 | 0·963 | 3·580 | 17419 | 5869 | 21784 |
| Telangana | Warangal (R) | 2·782 | 3·057 | 10·890 | 0·956 | 3·443 | 21468 | 7374 | 26546 |
| Telangana | Warangal (U) | 2·683 | 2·806 | 10·049 | 0·973 | 3·352 | 38315 | 13715 | 47185 |
| Telangana | Yadadri Bhongiri | 2·801 | 2·980 | 10·535 | 0·991 | 3·505 | 35528 | 12539 | 44543 |
| Tripura | Dhalai | 2·104 | 2·139 | 7·440 | 0·826 | 2·607 | 9621 | 3815 | 11949 |
| Tripura | Gomati | 2·391 | 2·534 | 8·791 | 0·893 | 2·950 | 13621 | 5091 | 17014 |
| Tripura | Khowai | 2·334 | 2·321 | 8·201 | 0·925 | 2·888 | 8888 | 3505 | 10940 |
| Tripura | North Tripura | 1·877 | 1·819 | 6·387 | 0·777 | 2·327 | 9495 | 4102 | 11765 |
| Tripura | Sipahijula | 2·643 | 2·707 | 9·510 | 1·023 | 3·296 | 18828 | 7319 | 23560 |
| Tripura | South Tripura | 2·507 | 2·742 | 9·381 | 0·912 | 3·100 | 14940 | 5474 | 18567 |
| Tripura | Unokoti | 2·005 | 1·901 | 6·705 | 0·833 | 2·510 | 6842 | 2853 | 8617 |
| Tripura | West Tripura | 2·549 | 2·497 | 8·919 | 1·019 | 3·179 | 25827 | 10397 | 32358 |
| Uttar Pradesh | Agra | 5·880 | 5·743 | 20·701 | 2·218 | 7·497 | 394942 | 147390 | 500510 |
| Uttar Pradesh | Aligarh | 4·899 | 4·862 | 17·689 | 1·879 | 6·116 | 198377 | 76644 | 245442 |
| Uttar Pradesh | Allahabad | 7·044 | 6·653 | 24·639 | 2·672 | 9·063 | 622571 | 236552 | 798059 |
| Uttar Pradesh | Ambedkar Nagar | 5·687 | 5·561 | 20·410 | 2·132 | 7·240 | 175043 | 65720 | 222972 |
| Uttar Pradesh | Amethi | 5·255 | 4·678 | 17·342 | 2·259 | 6·616 | 152352 | 65534 | 191855 |
| Uttar Pradesh | Auraiya | 5·885 | 5·067 | 18·645 | 2·559 | 7·484 | 104803 | 45239 | 133434 |
| Uttar Pradesh | Azamgarh | 6·383 | 6·496 | 24·191 | 2·236 | 8·104 | 424843 | 148005 | 539689 |
| Uttar Pradesh | Baghpat | 4·120 | 3·905 | 13·990 | 1·668 | 5·189 | 56146 | 22683 | 70640 |
| Uttar Pradesh | Bahraich | 4·269 | 3·846 | 14·090 | 1·794 | 5·411 | 145950 | 61226 | 185464 |
| Uttar Pradesh | Ballia | 7·239 | 7·499 | 27·644 | 2·439 | 9·215 | 278478 | 94056 | 355234 |
| Uttar Pradesh | Balrampur | 4·808 | 4·739 | 17·213 | 1·812 | 6·103 | 102097 | 38705 | 129261 |
| Uttar Pradesh | Banda | 7·343 | 6·136 | 22·918 | 3·183 | 9·511 | 191481 | 83061 | 249016 |
| Uttar Pradesh | Bara Banki | 4·540 | 3·809 | 14·049 | 2·047 | 5·751 | 170923 | 77821 | 216567 |
| Uttar Pradesh | Bareilly | 3·842 | 3·287 | 11·949 | 1·735 | 4·846 | 162117 | 73378 | 205224 |
| Uttar Pradesh | Basti | 5·330 | 5·273 | 19·286 | 1·989 | 6·726 | 181442 | 67480 | 229588 |
| Uttar Pradesh | Bijnor | 3·278 | 2·567 | 9·481 | 1·574 | 4·154 | 113837 | 54615 | 143982 |
| Uttar Pradesh | Budaun | 4·387 | 3·992 | 14·532 | 1·849 | 5·539 | 98799 | 41821 | 124763 |
| Uttar Pradesh | Bulandshahr | 4·480 | 4·343 | 15·737 | 1·766 | 5·615 | 152057 | 60082 | 190497 |
| Uttar Pradesh | Chandauli | 7·579 | 7·665 | 28·444 | 2·605 | 9·675 | 199840 | 69154 | 253600 |
| Uttar Pradesh | Chitrakoot | 7·660 | 6·855 | 25·427 | 3·087 | 9·898 | 128305 | 52275 | 165690 |
| Uttar Pradesh | Deoria | 6·488 | 6·658 | 24·939 | 2·187 | 8·280 | 248387 | 83667 | 317036 |
| Uttar Pradesh | Etah | 5·213 | 5·099 | 18·289 | 2·037 | 6·538 | 95599 | 37002 | 119828 |
| Uttar Pradesh | Etawah | 6·055 | 5·438 | 20·032 | 2·527 | 7·729 | 150194 | 62528 | 191527 |
| Uttar Pradesh | Faizabad | 5·004 | 4·565 | 16·714 | 2·071 | 6·352 | 164395 | 67762 | 208409 |
| Uttar Pradesh | Farrukhabad | 4·877 | 4·409 | 15·974 | 2·065 | 6·190 | 78368 | 33168 | 99015 |
| Uttar Pradesh | Fatehpur | 6·166 | 5·127 | 19·040 | 2·747 | 7·892 | 153465 | 68500 | 196492 |
| Uttar Pradesh | Firozabad | 5·689 | 5·499 | 19·743 | 2·214 | 7·201 | 167718 | 64760 | 213362 |
| Uttar Pradesh | Gautam Buddha Nagar | 4·609 | 4·643 | 16·978 | 1·731 | 5·780 | 308400 | 116745 | 385698 |
| Uttar Pradesh | Ghaziabad | 4·327 | 4·249 | 15·298 | 1·688 | 5·420 | 224381 | 86174 | 280945 |
| Uttar Pradesh | Ghazipur | 7·111 | 7·312 | 27·103 | 2·448 | 8·988 | 361769 | 125184 | 457371 |
| Uttar Pradesh | Gonda | 4·742 | 4·366 | 15·949 | 1·961 | 5·979 | 197654 | 81538 | 248962 |
| Uttar Pradesh | Gorakhpur | 6·015 | 6·234 | 22·868 | 2·063 | 7·614 | 388672 | 133233 | 492153 |
| Uttar Pradesh | Hamirpur | 7·283 | 5·913 | 21·958 | 3·296 | 9·343 | 108949 | 49450 | 139619 |
| Uttar Pradesh | Hapur | 4·146 | 3·863 | 14·173 | 1·696 | 5·228 | 61057 | 24659 | 76902 |
| Uttar Pradesh | Hardoi | 4·584 | 3·898 | 14·250 | 2·049 | 5·816 | 141619 | 63302 | 179935 |
| Uttar Pradesh | Jalaun | 6·939 | 5·687 | 21·174 | 3·105 | 8·925 | 160382 | 71944 | 206327 |
| Uttar Pradesh | Jaunpur | 6·529 | 6·383 | 23·672 | 2·373 | 8·347 | 424506 | 154283 | 542622 |
| Uttar Pradesh | Jhansi | 8·491 | 6·557 | 24·644 | 3·951 | 10·995 | 324051 | 150835 | 422976 |
| Uttar Pradesh | Jyotiba Phule Nagar | 3·858 | 3·409 | 12·488 | 1·679 | 4·847 | 71740 | 31456 | 90313 |
| Uttar Pradesh | Kannauj | 5·230 | 4·561 | 16·607 | 2·257 | 6·644 | 73776 | 31714 | 93574 |
| Uttar Pradesh | Kanpur Dehat | 5·857 | 4·843 | 17·968 | 2·626 | 7·468 | 98701 | 44160 | 126100 |
| Uttar Pradesh | Kanpur Nagar | 5·614 | 4·609 | 17·003 | 2·561 | 7·166 | 281298 | 127679 | 361531 |
| Uttar Pradesh | Kanshiram Nagar | 4·823 | 4·498 | 16·309 | 1·966 | 6·101 | 63313 | 25725 | 79495 |

| | | β-thalassaemia carrier prevalence | | | | | Estimated carrier counts | | |
|---|---|---|---|---|---|---|---|---|---|
| ADM1 | ADM2 | Mean | SD | CI95% | Q25 | Q75 | Mean | Q25 | Q75 |
| Uttar Pradesh | Kaushambi | 6·896 | 6·256 | 23·321 | 2·769 | 8·806 | 147964 | 59136 | 188879 |
| Uttar Pradesh | Kheri | 4·004 | 3·523 | 12·900 | 1·729 | 5·060 | 165117 | 71642 | 208585 |
| Uttar Pradesh | Kushinagar | 6·265 | 6·575 | 24·201 | 2·052 | 8·021 | 244029 | 79998 | 312237 |
| Uttar Pradesh | Lalitpur | 10·241 | 7·049 | 26·571 | 5·224 | 13·309 | 214827 | 109740 | 278866 |
| Uttar Pradesh | Lucknow | 4·536 | 3·555 | 13·200 | 2·175 | 5·766 | 426760 | 204998 | 541271 |
| Uttar Pradesh | Mahamaya Nagar | 5·275 | 5·272 | 19·003 | 2·010 | 6·619 | 89448 | 33913 | 112700 |
| Uttar Pradesh | Mahoba | 8·344 | 6·644 | 24·804 | 3·809 | 10·764 | 111200 | 50835 | 143854 |
| Uttar Pradesh | Mahrajganj | 5·852 | 6·292 | 23·035 | 1·925 | 7·360 | 169592 | 56045 | 213068 |
| Uttar Pradesh | Mainpuri | 5·416 | 5·061 | 18·368 | 2·206 | 6·837 | 109865 | 44744 | 138652 |
| Uttar Pradesh | Mathura | 5·323 | 5·387 | 19·489 | 1·937 | 6·734 | 244131 | 88867 | 307712 |
| Uttar Pradesh | Mau | 6·671 | 6·878 | 25·679 | 2·287 | 8·408 | 212166 | 72973 | 267069 |
| Uttar Pradesh | Meerut | 3·901 | 3·571 | 12·859 | 1·635 | 4·924 | 176535 | 73012 | 222662 |
| Uttar Pradesh | Mirzapur | 7·545 | 7·430 | 27·701 | 2·658 | 9·730 | 278389 | 98361 | 359178 |
| Uttar Pradesh | Moradabad | 3·600 | 3·002 | 10·992 | 1·658 | 4·528 | 108929 | 49938 | 137678 |
| Uttar Pradesh | Muzaffarnagar | 3·583 | 3·012 | 11·137 | 1·615 | 4·536 | 105095 | 47342 | 132111 |
| Uttar Pradesh | Pilibhit | 3·721 | 3·138 | 11·500 | 1·698 | 4·694 | 71165 | 32627 | 89414 |
| Uttar Pradesh | Pratapgarh | 6·101 | 5·659 | 20·889 | 2·446 | 7·739 | 304645 | 121853 | 386549 |
| Uttar Pradesh | Rae Bareli | 5·379 | 4·460 | 16·638 | 2·417 | 6·864 | 198524 | 88481 | 253806 |
| Uttar Pradesh | Rampur | 3·581 | 2·881 | 10·574 | 1·689 | 4·538 | 94614 | 44685 | 120321 |
| Uttar Pradesh | Saharanpur | 3·364 | 2·645 | 9·753 | 1·612 | 4·262 | 132018 | 63409 | 167613 |
| Uttar Pradesh | Sambhal | 4·179 | 3·787 | 13·768 | 1·772 | 5·251 | 75311 | 32343 | 94715 |
| Uttar Pradesh | Samli | 3·765 | 3·222 | 11·916 | 1·663 | 4·801 | 47302 | 20855 | 60456 |
| Uttar Pradesh | Sant Kabir Nagar | 5·666 | 5·789 | 21·456 | 2·004 | 7·131 | 126989 | 44893 | 159661 |
| Uttar Pradesh | Sant Ravidas Nagar (Bhadohi) | 6·943 | 6·740 | 24·924 | 2·550 | 8·911 | 164490 | 60450 | 211194 |
| Uttar Pradesh | Shahjahanpur | 4·234 | 3·736 | 13·524 | 1·852 | 5·353 | 124992 | 54759 | 157674 |
| Uttar Pradesh | Shrawasti | 4·387 | 4·086 | 15·050 | 1·756 | 5·599 | 54415 | 21888 | 69364 |
| Uttar Pradesh | Siddharthnagar | 5·284 | 5·481 | 20·145 | 1·879 | 6·618 | 127005 | 44989 | 159164 |
| Uttar Pradesh | Sitapur | 4·255 | 3·574 | 13·062 | 1·912 | 5·404 | 148598 | 66850 | 188744 |
| Uttar Pradesh | Sonbhadra | 8·225 | 8·231 | 30·935 | 2·814 | 10·555 | 311402 | 106803 | 400160 |
| Uttar Pradesh | Sultanpur | 5·659 | 5·353 | 19·704 | 2·232 | 7·145 | 181219 | 71726 | 228275 |
| Uttar Pradesh | Unnao | 4·990 | 4·000 | 14·743 | 2·344 | 6·331 | 155381 | 72695 | 197079 |
| Uttar Pradesh | Varanasi | 7·117 | 7·138 | 26·454 | 2·502 | 9·107 | 459392 | 160272 | 588085 |
| Uttarakhand | Almora | 2·913 | 2·161 | 7·972 | 1·473 | 3·676 | 23299 | 11829 | 29383 |
| Uttarakhand | Bageshwar | 2·912 | 2·369 | 8·644 | 1·364 | 3·681 | 10988 | 5282 | 13891 |
| Uttarakhand | Chamoli | 2·825 | 2·387 | 8·666 | 1·276 | 3·580 | 13493 | 6540 | 17108 |
| Uttarakhand | Champawat | 3·213 | 2·589 | 9·586 | 1·513 | 4·076 | 10467 | 4915 | 13316 |
| Uttarakhand | Dehradun | 2·975 | 2·194 | 8·096 | 1·506 | 3·764 | 65718 | 33758 | 82995 |
| Uttarakhand | Garhwal | 2·816 | 2·045 | 7·560 | 1·446 | 3·558 | 25643 | 13132 | 32565 |
| Uttarakhand | Hardwar | 3·130 | 2·377 | 8·814 | 1·542 | 3·970 | 52722 | 25786 | 66711 |
| Uttarakhand | Nainital | 3·108 | 2·301 | 8·538 | 1·575 | 3·926 | 43177 | 21798 | 54749 |
| Uttarakhand | Pithoragarh | 3·173 | 2·979 | 10·751 | 1·299 | 3·983 | 17561 | 7790 | 22127 |
| Uttarakhand | Rudraprayag | 2·677 | 2·066 | 7·613 | 1·304 | 3·393 | 6082 | 3037 | 7699 |
| Uttarakhand | Tehri Garhwal | 2·718 | 1·966 | 7·249 | 1·403 | 3·436 | 16618 | 8666 | 21026 |
| Uttarakhand | Udham Singh Nagar | 3·310 | 2·533 | 9·288 | 1·632 | 4·204 | 69408 | 34383 | 88136 |
| Uttarakhand | Uttarkashi | 2·861 | 2·323 | 8·517 | 1·339 | 3·627 | 8293 | 4012 | 10509 |
| West Bengal | Alipurduar | 6·270 | 5·280 | 19·710 | 2·763 | 8·015 | 108856 | 48497 | 138532 |
| West Bengal | Bankura | 8·619 | 5·152 | 19·449 | 4·951 | 10·983 | 427064 | 245463 | 544218 |
| West Bengal | Barddhaman | 7·554 | 4·557 | 17·278 | 4·346 | 9·591 | 449603 | 258980 | 570629 |
| West Bengal | Birbhum | 7·330 | 4·689 | 17·696 | 4·044 | 9·328 | 341099 | 188197 | 433528 |
| West Bengal | Dakshin Dinajpur | 6·351 | 5·001 | 18·675 | 2·992 | 8·091 | 161891 | 76156 | 206319 |
| West Bengal | Darjiling | 6·619 | 5·386 | 19·880 | 2·961 | 8·556 | 140908 | 65501 | 182925 |
| West Bengal | Haora | 7·224 | 4·328 | 16·245 | 4·183 | 9·145 | 357304 | 207723 | 452746 |
| West Bengal | Hugli | 7·196 | 4·281 | 16·217 | 4·187 | 9·110 | 513774 | 297238 | 650286 |
| West Bengal | Jalpaiguri | 6·399 | 4·981 | 18·654 | 3·025 | 8·192 | 246251 | 117457 | 315644 |
| West Bengal | Jhargram | 9·597 | 6·078 | 22·966 | 5·284 | 12·281 | 144551 | 80251 | 184809 |
| West Bengal | Kalimpong | 6·546 | 5·353 | 20·077 | 2·946 | 8·400 | 23994 | 10671 | 31010 |
| West Bengal | Koch Bihar | 6·044 | 4·908 | 18·256 | 2·748 | 7·757 | 252510 | 114969 | 324388 |

| ADM1 | ADM2 | β-thalassaemia carrier prevalence Mean | SD | CI95% | Q25 | Q75 | Estimated carrier counts Mean | Q25 | Q75 |
|---|---|---|---|---|---|---|---|---|---|
| West Bengal | Kolkata | 6·318 | 3·821 | 14·376 | 3·654 | 8·083 | 116985 | 67612 | 149498 |
| West Bengal | Maldah | 6·727 | 5·107 | 19·107 | 3·236 | 8·635 | 268330 | 129429 | 343689 |
| West Bengal | Murshidabad | 6·701 | 4·710 | 17·667 | 3·449 | 8·585 | 409395 | 209648 | 524558 |
| West Bengal | Nadia | 6·347 | 4·213 | 15·827 | 3·425 | 8·087 | 355308 | 193167 | 451784 |
| West Bengal | North Twenty Four Parganas | 5·765 | 3·799 | 14·234 | 3·142 | 7·317 | 527863 | 294754 | 670817 |
| West Bengal | Paschim Barddhaman | 7·966 | 5·040 | 19·011 | 4·418 | 10·176 | 353853 | 196655 | 451399 |
| West Bengal | Paschim Medinipur | 9·016 | 5·388 | 20·379 | 5·194 | 11·463 | 563206 | 325621 | 716024 |
| West Bengal | Purba Medinipur | 8·384 | 5·348 | 20·225 | 4·639 | 10·674 | 435876 | 243809 | 553717 |
| West Bengal | Puruliya | 9·139 | 5·893 | 22·256 | 4·989 | 11·712 | 287116 | 157302 | 367050 |
| West Bengal | South Twenty Four Parganas | 6·509 | 4·511 | 16·918 | 3·394 | 8·316 | 640209 | 354775 | 812230 |
| West Bengal | Uttar Dinajpur | 6·581 | 4·962 | 18·603 | 3·196 | 8·443 | 245242 | 118511 | 314555 |

ADM1: administrative level 1. ADM2: administrative level 2. SD: standard deviation. CI95%: 95% credible interval. Q25: 25th percentile. Q75: 75th percentile. SD and CI95% are measures of prediction uncertainty.

**Supplementary Table S23. Subnational areal predictions of β-thalassaemia carrier prevalence and estimated carrier count at administrative level 2 of Iran.**

| ADM1 | ADM2 | β-thalassaemia carrier prevalence Mean | SD | CI95% | Q25 | Q75 | Estimated carrier counts Mean | Q25 | Q75 |
|---|---|---|---|---|---|---|---|---|---|
| Alborz | Eshtehard | 6·054 | 4·954 | 18·395 | 2·744 | 7·735 | 3055 | 1368 | 3880 |
| Alborz | Karaj | 7·763 | 5·774 | 21·638 | 3·766 | 10·016 | 145485 | 69811 | 187168 |
| Alborz | Nazarabad | 6·807 | 5·436 | 20·274 | 3·111 | 8·835 | 13372 | 6200 | 17251 |
| Alborz | Savojbolagh | 7·384 | 5·746 | 21·196 | 3·451 | 9·558 | 20628 | 9671 | 26658 |
| Alborz | Shahariar | 6·755 | 5·156 | 19·224 | 3·320 | 8·509 | 34338 | 16858 | 43281 |
| Alborz | Taleghan | 7·846 | 6·185 | 22·958 | 3·618 | 10·172 | 1541 | 722 | 1984 |
| Ardabil | Ardebil | 2·910 | 2·109 | 7·819 | 1·495 | 3·679 | 20755 | 10690 | 26159 |
| Ardabil | Bilehsavar | 2·676 | 2·361 | 8·667 | 1·152 | 3·398 | 1556 | 643 | 2011 |
| Ardabil | Garmi | 2·620 | 2·181 | 7·992 | 1·210 | 3·292 | 2351 | 1086 | 2955 |
| Ardabil | Kahlkhal | 3·845 | 3·081 | 11·340 | 1·806 | 4·907 | 3734 | 1841 | 4707 |
| Ardabil | Kosar | 3·336 | 2·542 | 9·377 | 1·632 | 4·244 | 850 | 415 | 1084 |
| Ardabil | Meshkinshahr | 2·614 | 1·999 | 7·332 | 1·296 | 3·306 | 4618 | 2262 | 5915 |
| Ardabil | Namin | 2·944 | 2·181 | 8·014 | 1·479 | 3·747 | 1909 | 950 | 2444 |
| Ardabil | Nir | 2·988 | 2·181 | 8·019 | 1·533 | 3·779 | 735 | 374 | 925 |
| Ardabil | Parsabad | 2·619 | 2·394 | 8·782 | 1·079 | 3·338 | 5146 | 2076 | 6635 |
| Ardabil | Sareyn | 2·803 | 2·031 | 7·598 | 1·454 | 3·524 | 540 | 278 | 686 |
| Bushehr | Asaluyeh | 1·007 | 0·804 | 2·912 | 0·482 | 1·270 | 804 | 384 | 1021 |
| Bushehr | Bushehr | 1·206 | 0·998 | 3·594 | 0·565 | 1·524 | 4040 | 1890 | 5093 |
| Bushehr | Dashtestan | 0·989 | 0·807 | 2·928 | 0·469 | 1·244 | 2972 | 1391 | 3730 |
| Bushehr | Dashti | 1·003 | 0·771 | 2·802 | 0·499 | 1·265 | 1017 | 499 | 1294 |
| Bushehr | Deylam | 1·379 | 1·136 | 4·119 | 0·646 | 1·732 | 247 | 113 | 311 |
| Bushehr | Deyr | 1·094 | 0·855 | 3·113 | 0·540 | 1·371 | 744 | 361 | 928 |
| Bushehr | Genaveh | 1·295 | 1·067 | 3·878 | 0·607 | 1·636 | 1543 | 736 | 1934 |
| Bushehr | Jam | 0·928 | 0·721 | 2·579 | 0·463 | 1·165 | 762 | 380 | 960 |
| Bushehr | Kangan | 1·000 | 0·794 | 2·900 | 0·489 | 1·260 | 963 | 463 | 1210 |
| Bushehr | Tangestan | 1·099 | 0·880 | 3·202 | 0·529 | 1·388 | 989 | 468 | 1246 |
| Chaharmahal and Bakhtiari | Ardal | 0·684 | 0·587 | 2·104 | 0·312 | 0·858 | 348 | 157 | 434 |
| Chaharmahal and Bakhtiari | Ben | 0·570 | 0·490 | 1·756 | 0·263 | 0·712 | 170 | 77 | 212 |
| Chaharmahal and Bakhtiari | Borujen | 0·444 | 0·359 | 1·293 | 0·215 | 0·558 | 587 | 289 | 734 |
| Chaharmahal and Bakhtiari | Farsan | 0·617 | 0·518 | 1·838 | 0·288 | 0·779 | 680 | 321 | 848 |
| Chaharmahal and Bakhtiari | Kiar | 0·547 | 0·452 | 1·660 | 0·258 | 0·688 | 299 | 142 | 374 |
| Chaharmahal and Bakhtiari | Kuhrang | 0·861 | 0·760 | 2·750 | 0·380 | 1·081 | 417 | 190 | 529 |
| Chaharmahal and Bakhtiari | Lordegan | 0·572 | 0·463 | 1·690 | 0·274 | 0·716 | 1387 | 664 | 1765 |
| Chaharmahal and Bakhtiari | Saman | 0·487 | 0·412 | 1·511 | 0·228 | 0·606 | 195 | 91 | 237 |

| | | β-thalassaemia carrier prevalence | | | | | Estimated carrier counts | | |
|---|---|---|---|---|---|---|---|---|---|
| ADM1 | ADM2 | Mean | SD | CI95% | Q25 | Q75 | Mean | Q25 | Q75 |
| Chaharmahal and Bakhtiari | Sharekurd | 0·526 | 0·442 | 1·587 | 0·248 | 0·653 | 1909 | 911 | 2337 |
| East Azerbaijan | Ahar | 2·551 | 2·090 | 7·645 | 1·189 | 3·225 | 4621 | 2165 | 5916 |
| East Azerbaijan | Ajabshir | 3·491 | 2·934 | 10·647 | 1·569 | 4·440 | 2900 | 1334 | 3718 |
| East Azerbaijan | Azarshahr | 3·341 | 2·895 | 10·509 | 1·469 | 4·207 | 4218 | 1866 | 5285 |
| East Azerbaijan | Bonab | 3·614 | 2·948 | 10·926 | 1·674 | 4·593 | 6054 | 2885 | 7755 |
| East Azerbaijan | Bostanabad | 3·070 | 2·522 | 9·160 | 1·428 | 3·903 | 3323 | 1536 | 4247 |
| East Azerbaijan | Charuymaq | 3·728 | 3·082 | 11·223 | 1·709 | 4·737 | 1300 | 594 | 1639 |
| East Azerbaijan | Hashtrud | 3·383 | 2·794 | 10·107 | 1·569 | 4·270 | 2250 | 1050 | 2845 |
| East Azerbaijan | Heris | 2·759 | 2·274 | 8·286 | 1·268 | 3·512 | 2224 | 1042 | 2838 |
| East Azerbaijan | Jolfa | 2·521 | 2·397 | 8·740 | 1·020 | 3·145 | 2283 | 906 | 2826 |
| East Azerbaijan | Kaleybar | 2·475 | 2·144 | 7·789 | 1·105 | 3·117 | 1321 | 589 | 1675 |
| East Azerbaijan | Khoda Afarin | 2·478 | 2·232 | 8·135 | 1·047 | 3·128 | 802 | 337 | 1016 |
| East Azerbaijan | Malekan | 3·745 | 3·018 | 11·118 | 1·771 | 4·752 | 5313 | 2511 | 6779 |
| East Azerbaijan | Maragheh | 3·540 | 2·894 | 10·687 | 1·630 | 4·480 | 10418 | 4766 | 13223 |
| East Azerbaijan | Marand | 2·664 | 2·476 | 8·871 | 1·120 | 3·334 | 7915 | 3302 | 9939 |
| East Azerbaijan | Mianeh | 3·390 | 2·675 | 9·782 | 1·612 | 4·322 | 7336 | 3497 | 9312 |
| East Azerbaijan | Osku | 3·193 | 2·783 | 10·065 | 1·398 | 4·062 | 5126 | 2319 | 6487 |
| East Azerbaijan | Sarab | 2·891 | 2·228 | 8·158 | 1·420 | 3·670 | 4304 | 2096 | 5436 |
| East Azerbaijan | Shabestar | 2·883 | 2·569 | 9·408 | 1·242 | 3·630 | 4512 | 1934 | 5719 |
| East Azerbaijan | Tabriz | 2·954 | 2·549 | 9·303 | 1·301 | 3·732 | 61969 | 27264 | 77626 |
| East Azerbaijan | Varzaqan | 2·615 | 2·295 | 8·420 | 1·143 | 3·294 | 1554 | 680 | 1971 |
| Fars | Abadeh | 0·367 | 0·333 | 1·175 | 0·160 | 0·460 | 431 | 187 | 540 |
| Fars | Arsanjan | 0·521 | 0·460 | 1·652 | 0·232 | 0·652 | 255 | 110 | 319 |
| Fars | Bavanat | 0·460 | 0·440 | 1·569 | 0·188 | 0·576 | 271 | 109 | 332 |
| Fars | Darab | 0·591 | 0·458 | 1·656 | 0·293 | 0·745 | 1369 | 658 | 1733 |
| Fars | Eqlid | 0·448 | 0·374 | 1·342 | 0·209 | 0·563 | 461 | 203 | 579 |
| Fars | Estahban | 0·564 | 0·487 | 1·758 | 0·257 | 0·706 | 449 | 200 | 564 |
| Fars | Farashband | 0·757 | 0·581 | 2·124 | 0·379 | 0·949 | 380 | 182 | 472 |
| Fars | Fasa | 0·585 | 0·485 | 1·760 | 0·275 | 0·732 | 1380 | 642 | 1735 |
| Fars | Firuzabad | 0·667 | 0·509 | 1·845 | 0·337 | 0·835 | 884 | 458 | 1105 |
| Fars | Gerash | 0·767 | 0·623 | 2·233 | 0·371 | 0·961 | 464 | 232 | 583 |
| Fars | Jahrom | 0·618 | 0·489 | 1·773 | 0·302 | 0·776 | 1628 | 789 | 2046 |
| Fars | Kavar | 0·616 | 0·472 | 1·715 | 0·311 | 0·771 | 611 | 305 | 755 |
| Fars | Kazerun | 0·754 | 0·596 | 2·149 | 0·371 | 0·945 | 2344 | 1154 | 2952 |
| Fars | Kharameh | 0·552 | 0·462 | 1·682 | 0·257 | 0·691 | 345 | 162 | 438 |
| Fars | Khonj | 0·764 | 0·585 | 2·124 | 0·384 | 0·957 | 336 | 167 | 429 |
| Fars | Khorambid | 0·432 | 0·398 | 1·428 | 0·184 | 0·540 | 249 | 103 | 318 |
| Fars | Lamerd | 0·872 | 0·694 | 2·522 | 0·423 | 1·093 | 954 | 462 | 1188 |
| Fars | Larestan | 0·677 | 0·518 | 1·881 | 0·342 | 0·850 | 1681 | 839 | 2108 |
| Fars | Mamasani | 0·750 | 0·584 | 2·128 | 0·370 | 0·941 | 873 | 445 | 1091 |
| Fars | Marvdasht | 0·517 | 0·413 | 1·484 | 0·252 | 0·649 | 1957 | 960 | 2451 |
| Fars | Mehr | 0·906 | 0·702 | 2·572 | 0·447 | 1·135 | 679 | 338 | 859 |
| Fars | Neyriz | 0·550 | 0·493 | 1·761 | 0·242 | 0·689 | 708 | 309 | 894 |
| Fars | Pasargad | 0·479 | 0·412 | 1·487 | 0·217 | 0·603 | 170 | 78 | 216 |
| Fars | Qir and Karzin | 0·682 | 0·525 | 1·916 | 0·342 | 0·853 | 489 | 245 | 611 |
| Fars | Rostam | 0·667 | 0·509 | 1·853 | 0·335 | 0·839 | 355 | 176 | 447 |
| Fars | Sarvestan | 0·588 | 0·464 | 1·688 | 0·287 | 0·739 | 256 | 124 | 321 |
| Fars | Sepidan | 0·559 | 0·426 | 1·546 | 0·284 | 0·702 | 654 | 329 | 818 |
| Fars | Shiraz | 0·614 | 0·466 | 1·695 | 0·312 | 0·769 | 12883 | 6533 | 16130 |
| Fars | Zarindasht | 0·610 | 0·456 | 1·666 | 0·311 | 0·767 | 505 | 255 | 632 |
| Gilan | Amlash | 6·806 | 5·863 | 22·034 | 2·925 | 8·719 | 4282 | 1810 | 5570 |
| Gilan | Astan-e-Ashrafieh | 6·192 | 5·525 | 20·362 | 2·607 | 7·884 | 7732 | 3292 | 9764 |
| Gilan | Astara | 3·157 | 2·375 | 8·735 | 1·557 | 3·984 | 3634 | 1807 | 4568 |
| Gilan | Bandar-e-Anzali | 5·172 | 4·551 | 17·045 | 2·195 | 6·649 | 6525 | 2761 | 8363 |
| Gilan | Fuman | 4·818 | 4·146 | 15·120 | 2·090 | 6·172 | 5970 | 2532 | 7589 |
| Gilan | Lahijan | 6·420 | 5·705 | 21·213 | 2·662 | 8·282 | 12251 | 5139 | 15763 |
| Gilan | Langerud | 6·633 | 5·824 | 21·417 | 2·809 | 8·459 | 9816 | 4132 | 12549 |

| ADM1 | ADM2 | β-thalassaemia carrier prevalence Mean | SD | CI95% | Q25 | Q75 | Estimated carrier counts Mean | Q25 | Q75 |
|---|---|---|---|---|---|---|---|---|---|
| Gilan | Masal | 4·414 | 3·613 | 13·198 | 2·008 | 5·671 | 2921 | 1341 | 3771 |
| Gilan | Rasht | 5·701 | 5·008 | 18·362 | 2·409 | 7·316 | 60538 | 25679 | 78084 |
| Gilan | Rezvanshahr | 4·158 | 3·353 | 12·481 | 1·926 | 5·335 | 3275 | 1492 | 4148 |
| Gilan | Rudbar | 6·013 | 5·239 | 19·116 | 2·550 | 7·720 | 6393 | 2708 | 8192 |
| Gilan | Rudsar | 7·168 | 6·027 | 22·477 | 3·129 | 9·242 | 11627 | 4998 | 15026 |
| Gilan | Shaft | 5·361 | 4·798 | 17·209 | 2·255 | 6·845 | 3995 | 1678 | 5075 |
| Gilan | Siahkal | 6·346 | 5·466 | 20·366 | 2·710 | 8·132 | 3277 | 1370 | 4168 |
| Gilan | Sowme'eh Sara | 4·883 | 4·231 | 15·460 | 2·121 | 6·222 | 6559 | 2859 | 8351 |
| Gilan | Talesh | 3·625 | 2·790 | 10·389 | 1·761 | 4·614 | 8036 | 3829 | 10293 |
| Golestan | Aliabad | 8·251 | 6·261 | 23·404 | 3·934 | 10·598 | 14813 | 6997 | 19175 |
| Golestan | Aqqala | 9·101 | 6·894 | 26·085 | 4·332 | 11·744 | 13919 | 6744 | 17962 |
| Golestan | Azadshahr | 7·990 | 6·656 | 24·961 | 3·451 | 10·295 | 9244 | 4094 | 11993 |
| Golestan | Galikash | 8·230 | 7·435 | 27·545 | 3·207 | 10·666 | 6199 | 2543 | 8049 |
| Golestan | Gomishan | 9·394 | 6·982 | 26·228 | 4·544 | 12·138 | 7668 | 3749 | 9951 |
| Golestan | Gonbad-e-Kavus | 9·058 | 7·811 | 29·250 | 3·744 | 11·753 | 34806 | 15049 | 45214 |
| Golestan | Gorgan | 8·440 | 6·085 | 22·980 | 4·218 | 10·856 | 48317 | 24182 | 62619 |
| Golestan | Kalaleh | 8·717 | 8·025 | 29·743 | 3·369 | 11·270 | 12187 | 4823 | 15660 |
| Golestan | Kordkuy | 8·440 | 6·163 | 23·109 | 4·167 | 10·858 | 8003 | 3969 | 10378 |
| Golestan | Maraveh Tapeh | 8·710 | 8·551 | 32·076 | 3·076 | 11·225 | 6135 | 2195 | 7838 |
| Golestan | Minudasht | 8·128 | 6·976 | 25·968 | 3·419 | 10·530 | 7000 | 2945 | 8999 |
| Golestan | Ramian | 8·440 | 6·761 | 25·104 | 3·812 | 10·830 | 7840 | 3513 | 10061 |
| Golestan | Turkman | 9·019 | 6·527 | 24·948 | 4·477 | 11·532 | 7170 | 3516 | 9189 |
| Hamadan | Asadabad | 4·456 | 4·024 | 14·689 | 1·824 | 5·707 | 5130 | 2075 | 6499 |
| Hamadan | Bahar | 4·442 | 4·140 | 15·303 | 1·761 | 5·658 | 6113 | 2400 | 7727 |
| Hamadan | Famenin | 4·280 | 3·887 | 14·380 | 1·724 | 5·497 | 1991 | 794 | 2561 |
| Hamadan | Hamedan | 4·055 | 3·821 | 14·026 | 1·579 | 5·170 | 33385 | 12817 | 42834 |
| Hamadan | Kabutarahang | 4·654 | 4·300 | 15·678 | 1·863 | 5·931 | 6743 | 2678 | 8586 |
| Hamadan | Malayer | 3·528 | 3·411 | 12·332 | 1·356 | 4·492 | 12088 | 4659 | 15508 |
| Hamadan | Nahavand | 4·019 | 3·777 | 13·924 | 1·560 | 5·116 | 8082 | 3130 | 10319 |
| Hamadan | Razan | 4·749 | 4·307 | 15·888 | 1·919 | 6·100 | 5924 | 2391 | 7661 |
| Hamadan | Tuyserkan | 4·130 | 3·792 | 13·835 | 1·656 | 5·275 | 4700 | 1894 | 6053 |
| Hormozgan | Bandar-e-Abbas | 0·672 | 0·521 | 1·894 | 0·334 | 0·843 | 4502 | 2253 | 5666 |
| Hormozgan | Bandar-e-Lengeh | 0·878 | 0·693 | 2·528 | 0·426 | 1·103 | 1382 | 660 | 1752 |
| Hormozgan | Bashagard | 1·076 | 0·968 | 3·478 | 0·471 | 1·346 | 423 | 173 | 530 |
| Hormozgan | Bastak | 0·769 | 0·593 | 2·170 | 0·383 | 0·966 | 692 | 348 | 882 |
| Hormozgan | Hajiabad | 0·628 | 0·519 | 1·860 | 0·297 | 0·788 | 483 | 222 | 598 |
| Hormozgan | Jask | 1·463 | 1·431 | 5·077 | 0·587 | 1·821 | 452 | 168 | 569 |
| Hormozgan | Khamir | 0·702 | 0·533 | 1·954 | 0·353 | 0·882 | 411 | 200 | 519 |
| Hormozgan | Minab | 0·819 | 0·655 | 2·377 | 0·400 | 1·025 | 2249 | 1097 | 2808 |
| Hormozgan | Parsian | 0·987 | 0·807 | 2·948 | 0·464 | 1·238 | 581 | 274 | 730 |
| Hormozgan | Qeshm | 0·819 | 0·657 | 2·373 | 0·394 | 1·029 | 863 | 413 | 1081 |
| Hormozgan | Rudan | 0·721 | 0·611 | 2·192 | 0·335 | 0·901 | 1094 | 507 | 1365 |
| Hormozgan | Sirik | 0·959 | 0·791 | 2·847 | 0·456 | 1·201 | 474 | 230 | 600 |
| Ilam | Abdanan County | 4·676 | 4·336 | 16·007 | 1·863 | 5·976 | 2653 | 1028 | 3463 |
| Ilam | Badreh | 5·433 | 4·962 | 18·391 | 2·147 | 6·958 | 884 | 346 | 1144 |
| Ilam | Chardavol | 5·558 | 4·953 | 18·373 | 2·261 | 7·151 | 3931 | 1608 | 5063 |
| Ilam | Darreh Shahr County | 4·776 | 4·477 | 16·372 | 1·854 | 6·125 | 2426 | 955 | 3092 |
| Ilam | Dehloran County | 5·087 | 4·514 | 16·634 | 2·112 | 6·530 | 3849 | 1593 | 4885 |
| Ilam | Eyvan County | 6·165 | 5·126 | 18·872 | 2·744 | 7·900 | 3512 | 1545 | 4555 |
| Ilam | Ilam | 6·250 | 5·432 | 20·068 | 2·631 | 8·045 | 16639 | 6598 | 21768 |
| Ilam | Malekshahi | 5·881 | 5·280 | 19·189 | 2·372 | 7·610 | 1414 | 564 | 1845 |
| Ilam | Mehran County | 6·147 | 5·491 | 20·211 | 2·491 | 7·929 | 2114 | 843 | 2758 |
| Ilam | Sirvan | 5·814 | 5·187 | 19·369 | 2·367 | 7·498 | 1002 | 415 | 1319 |
| Isfahan | Aran and Bidgol | 1·193 | 1·022 | 3·669 | 0·543 | 1·498 | 1504 | 695 | 1893 |
| Isfahan | Ardestan | 0·515 | 0·446 | 1·580 | 0·237 | 0·643 | 217 | 93 | 269 |
| Isfahan | Barkhar | 0·374 | 0·301 | 1·080 | 0·182 | 0·467 | 429 | 209 | 533 |
| Isfahan | Buin and Miandasht | 1·061 | 0·988 | 3·500 | 0·459 | 1·319 | 306 | 129 | 384 |

| ADM1 | ADM2 | β-thalassaemia carrier prevalence Mean | SD | CI95% | Q25 | Q75 | Estimated carrier counts Mean | Q25 | Q75 |
|---|---|---|---|---|---|---|---|---|---|
| Isfahan | Chadegan | 0·653 | 0·562 | 2·011 | 0·302 | 0·813 | 243 | 113 | 302 |
| Isfahan | Dehaqan | 0·338 | 0·269 | 0·971 | 0·165 | 0·423 | 133 | 63 | 169 |
| Isfahan | Falaverjan | 0·329 | 0·248 | 0·885 | 0·166 | 0·419 | 838 | 420 | 1068 |
| Isfahan | Fereydan | 0·775 | 0·693 | 2·442 | 0·348 | 0·958 | 425 | 192 | 521 |
| Isfahan | Fereydunshahr | 1·069 | 1·004 | 3·490 | 0·457 | 1·322 | 365 | 159 | 451 |
| Isfahan | Golpayegan | 1·002 | 0·901 | 3·254 | 0·440 | 1·248 | 1079 | 474 | 1332 |
| Isfahan | Isfahan | 0·334 | 0·258 | 0·941 | 0·168 | 0·419 | 4566 | 2286 | 5721 |
| Isfahan | Isfahan County | 0·306 | 0·261 | 0·931 | 0·142 | 0·383 | 3929 | 1947 | 4918 |
| Isfahan | Kashan | 1·083 | 0·938 | 3·367 | 0·493 | 1·353 | 4239 | 1929 | 5308 |
| Isfahan | Khansar | 0·877 | 0·805 | 2·853 | 0·387 | 1·078 | 350 | 156 | 431 |
| Isfahan | Khomeini Shahr | 0·349 | 0·268 | 0·995 | 0·177 | 0·434 | 1578 | 805 | 1963 |
| Isfahan | Khur and Biabanak | 0·796 | 0·795 | 2·795 | 0·314 | 0·993 | 172 | 66 | 220 |
| Isfahan | Lenjan | 0·388 | 0·304 | 1·114 | 0·193 | 0·485 | 1036 | 517 | 1308 |
| Isfahan | Mobarakeh | 0·331 | 0·256 | 0·940 | 0·165 | 0·415 | 573 | 288 | 723 |
| Isfahan | Nain | 0·434 | 0·384 | 1·368 | 0·196 | 0·540 | 138 | 64 | 178 |
| Isfahan | Najafabad | 0·475 | 0·377 | 1·349 | 0·234 | 0·594 | 1579 | 800 | 1987 |
| Isfahan | Natanz | 0·578 | 0·484 | 1·746 | 0·271 | 0·724 | 305 | 141 | 386 |
| Isfahan | Sahreza | 0·328 | 0·270 | 0·964 | 0·156 | 0·412 | 582 | 274 | 732 |
| Isfahan | Semirom | 0·444 | 0·358 | 1·307 | 0·212 | 0·559 | 370 | 176 | 471 |
| Isfahan | Shahin Shahr and Meymeh | 0·554 | 0·448 | 1·629 | 0·267 | 0·692 | 991 | 490 | 1249 |
| Isfahan | Tiran and Karvan | 0·479 | 0·385 | 1·377 | 0·236 | 0·596 | 365 | 177 | 459 |
| Kerman | Anar | 0·540 | 0·647 | 2·187 | 0·179 | 0·652 | 213 | 72 | 263 |
| Kerman | Anbarabad | 0·960 | 1·053 | 3·687 | 0·342 | 1·182 | 901 | 317 | 1118 |
| Kerman | Arzuiyeh | 0·659 | 0·598 | 2·132 | 0·285 | 0·826 | 284 | 117 | 365 |
| Kerman | Baft | 0·685 | 0·700 | 2·451 | 0·263 | 0·851 | 663 | 244 | 832 |
| Kerman | Bam | 1·034 | 1·239 | 4·276 | 0·330 | 1·256 | 812 | 254 | 968 |
| Kerman | Bardsir | 0·741 | 0·883 | 2·987 | 0·246 | 0·899 | 706 | 224 | 863 |
| Kerman | Fahraj | 1·336 | 1·729 | 5·883 | 0·390 | 1·588 | 950 | 281 | 1122 |
| Kerman | Faryab | 0·747 | 0·721 | 2·553 | 0·311 | 0·926 | 287 | 119 | 355 |
| Kerman | Ghalehganj | 1·022 | 1·015 | 3·577 | 0·411 | 1·266 | 795 | 325 | 977 |
| Kerman | Jiroft | 0·844 | 0·924 | 3·213 | 0·300 | 1·036 | 2909 | 1041 | 3542 |
| Kerman | Kahnuj | 0·795 | 0·767 | 2·684 | 0·333 | 0·984 | 906 | 362 | 1122 |
| Kerman | Kerman | 0·908 | 1·214 | 3·910 | 0·272 | 1·071 | 7201 | 2160 | 8641 |
| Kerman | Kuhbanan | 0·755 | 1·069 | 3·322 | 0·216 | 0·882 | 182 | 51 | 208 |
| Kerman | Manujan | 0·807 | 0·698 | 2·483 | 0·366 | 1·015 | 574 | 262 | 728 |
| Kerman | Narmashir | 1·170 | 1·453 | 4·940 | 0·357 | 1·415 | 144 | 44 | 175 |
| Kerman | Rabor | 0·768 | 0·855 | 2·972 | 0·266 | 0·946 | 300 | 104 | 375 |
| Kerman | Rafsanjan | 0·639 | 0·806 | 2·602 | 0·208 | 0·765 | 2307 | 766 | 2793 |
| Kerman | Ravar | 0·913 | 1·340 | 4·044 | 0·252 | 1·057 | 427 | 115 | 502 |
| Kerman | Rigan | 1·222 | 1·493 | 5·135 | 0·385 | 1·470 | 1251 | 358 | 1487 |
| Kerman | Rudbar-e-Jonub | 1·004 | 1·095 | 3·824 | 0·368 | 1·231 | 1140 | 425 | 1401 |
| Kerman | Shahr-e-Babak | 0·543 | 0·616 | 2·063 | 0·194 | 0·662 | 639 | 221 | 786 |
| Kerman | Sirjan | 0·624 | 0·629 | 2·192 | 0·244 | 0·777 | 2304 | 893 | 2940 |
| Kerman | Zarand | 0·726 | 0·985 | 3·068 | 0·220 | 0·855 | 1256 | 368 | 1446 |
| Kermanshah | Dalahu | 6·002 | 4·750 | 17·746 | 2·794 | 7·701 | 2306 | 1078 | 2936 |
| Kermanshah | Eslamabad-e Gharb | 5·796 | 4·977 | 18·402 | 2·479 | 7·460 | 9176 | 4003 | 11823 |
| Kermanshah | Gilan-e-Gharb | 6·341 | 5·143 | 19·303 | 2·870 | 8·134 | 4092 | 1870 | 5264 |
| Kermanshah | Harsin | 4·919 | 4·373 | 15·919 | 2·047 | 6·341 | 4079 | 1680 | 5315 |
| Kermanshah | Javanrud | 5·568 | 4·136 | 15·451 | 2·748 | 7·082 | 4714 | 2273 | 6048 |
| Kermanshah | kangavar | 4·446 | 4·056 | 14·964 | 1·800 | 5·666 | 3874 | 1552 | 4965 |
| Kermanshah | Kermanshah | 5·383 | 4·664 | 17·132 | 2·289 | 6·902 | 67184 | 29113 | 85793 |
| Kermanshah | Paveh | 5·513 | 4·007 | 15·013 | 2·771 | 7·052 | 3819 | 1892 | 4831 |
| Kermanshah | Qasr-e-Shirin | 6·444 | 4·868 | 18·275 | 3·117 | 8·303 | 1749 | 882 | 2240 |
| Kermanshah | Ravansar | 5·462 | 4·331 | 15·829 | 2·556 | 6·989 | 3210 | 1513 | 4127 |
| Kermanshah | Sahneh | 4·872 | 4·349 | 16·113 | 2·022 | 6·217 | 3966 | 1631 | 5027 |
| Kermanshah | Salas-e Babajani | 5·829 | 4·156 | 15·382 | 2·975 | 7·447 | 2374 | 1183 | 3075 |
| Kermanshah | Sarpol-e-Zahab | 6·153 | 4·492 | 16·777 | 3·098 | 7·862 | 6380 | 3122 | 8125 |

| | | β-thalassaemia carrier prevalence | | | | | Estimated carrier counts | | |
|---|---|---|---|---|---|---|---|---|---|
| ADM1 | ADM2 | Mean | SD | CI95% | Q25 | Q75 | Mean | Q25 | Q75 |
| Kermanshah | Sonqor | 4·864 | 4·282 | 15·813 | 2·052 | 6·210 | 4738 | 1959 | 6049 |
| Khuzestan | Abadan | 3·932 | 2·910 | 10·832 | 1·971 | 5·006 | 9041 | 4669 | 11591 |
| Khuzestan | Aghajari | 1·490 | 1·203 | 4·348 | 0·706 | 1·877 | 324 | 152 | 411 |
| Khuzestan | Ahvaz | 3·187 | 2·515 | 9·258 | 1·526 | 4·046 | 48903 | 22790 | 62154 |
| Khuzestan | Andika | 1·536 | 1·440 | 5·138 | 0·646 | 1·910 | 814 | 345 | 1014 |
| Khuzestan | Andimeshk | 3·515 | 3·351 | 12·160 | 1·369 | 4·469 | 7538 | 3021 | 9795 |
| Khuzestan | Baghmalek | 1·296 | 1·145 | 4·099 | 0·585 | 1·613 | 1504 | 679 | 1874 |
| Khuzestan | Bandar-e-Mahshahr | 2·585 | 2·092 | 7·558 | 1·219 | 3·271 | 8650 | 4097 | 10797 |
| Khuzestan | Bavi | 2·528 | 2·186 | 7·911 | 1·130 | 3·183 | 2882 | 1300 | 3679 |
| Khuzestan | Behbahan | 1·283 | 1·066 | 3·873 | 0·601 | 1·613 | 2726 | 1266 | 3412 |
| Khuzestan | Dasht-e-Azadegan | 4·089 | 3·282 | 12·061 | 1·909 | 5·233 | 5543 | 2624 | 7108 |
| Khuzestan | Dezful | 2·670 | 2·507 | 9·074 | 1·080 | 3·371 | 16136 | 6636 | 20353 |
| Khuzestan | Gotvand | 2·554 | 2·377 | 8·548 | 1·056 | 3·210 | 2008 | 823 | 2526 |
| Khuzestan | Haftgol | 1·834 | 1·624 | 5·849 | 0·812 | 2·303 | 456 | 201 | 574 |
| Khuzestan | Hamidieh | 3·510 | 2·856 | 10·406 | 1·642 | 4·451 | 1837 | 874 | 2284 |
| Khuzestan | Hendijan | 1·864 | 1·583 | 5·653 | 0·861 | 2·350 | 858 | 400 | 1081 |
| Khuzestan | Hoveyzeh | 4·573 | 3·327 | 12·412 | 2·324 | 5·817 | 1871 | 933 | 2413 |
| Khuzestan | Izeh | 1·216 | 1·124 | 3·972 | 0·524 | 1·511 | 2736 | 1173 | 3438 |
| Khuzestan | Karun | 3·391 | 2·593 | 9·693 | 1·657 | 4·303 | 2868 | 1377 | 3635 |
| Khuzestan | Khoramshahr | 4·298 | 2·960 | 11·109 | 2·257 | 5·475 | 10674 | 5559 | 13719 |
| Khuzestan | Lali | 2·055 | 1·985 | 7·113 | 0·821 | 2·585 | 926 | 372 | 1135 |
| Khuzestan | Masjedsoleyman | 1·893 | 1·773 | 6·342 | 0·795 | 2·362 | 2500 | 1069 | 3078 |
| Khuzestan | Omidieh | 1·767 | 1·496 | 5·433 | 0·816 | 2·225 | 1897 | 885 | 2441 |
| Khuzestan | Ramhormoz | 1·634 | 1·408 | 5·060 | 0·744 | 2·052 | 2142 | 960 | 2674 |
| Khuzestan | Ramshir | 2·089 | 1·740 | 6·310 | 0·968 | 2·629 | 1227 | 573 | 1549 |
| Khuzestan | Shadegan | 3·486 | 2·615 | 9·703 | 1·729 | 4·415 | 5502 | 2673 | 7043 |
| Khuzestan | Shush | 3·746 | 3·256 | 11·823 | 1·618 | 4·794 | 8386 | 3518 | 10686 |
| Khuzestan | Shushtar | 2·552 | 2·260 | 8·104 | 1·102 | 3·237 | 5781 | 2530 | 7273 |
| Kohgiluyeh and Boyer-Ahmad | Bahmai | 1·201 | 1·010 | 3·634 | 0·558 | 1·507 | 427 | 200 | 543 |
| Kohgiluyeh and Boyer-Ahmad | Basht | 0·745 | 0·568 | 2·084 | 0·373 | 0·934 | 198 | 98 | 251 |
| Kohgiluyeh and Boyer-Ahmad | Buyerahmad | 0·591 | 0·456 | 1·668 | 0·294 | 0·741 | 1960 | 967 | 2472 |
| Kohgiluyeh and Boyer-Ahmad | Charam | 0·744 | 0·578 | 2·096 | 0·371 | 0·927 | 335 | 164 | 421 |
| Kohgiluyeh and Boyer-Ahmad | Dena | 0·552 | 0·429 | 1·575 | 0·274 | 0·691 | 284 | 138 | 355 |
| Kohgiluyeh and Boyer-Ahmad | Gachsaran | 0·953 | 0·760 | 2·766 | 0·462 | 1·194 | 1335 | 647 | 1676 |
| Kohgiluyeh and Boyer-Ahmad | Kohkiluyeh | 0·894 | 0·718 | 2·606 | 0·432 | 1·117 | 1305 | 638 | 1655 |
| Kohgiluyeh and Boyer-Ahmad | Landeh | 0·986 | 0·827 | 2·955 | 0·466 | 1·236 | 233 | 110 | 282 |
| Kurdistan | Baneh | 4·566 | 3·128 | 11·813 | 2·428 | 5·797 | 8650 | 4478 | 11072 |
| Kurdistan | Bijar | 4·685 | 4·136 | 15·163 | 1·996 | 5·970 | 4771 | 2010 | 6135 |
| Kurdistan | Dehgolan | 4·951 | 4·314 | 15·608 | 2·118 | 6·330 | 3714 | 1573 | 4719 |
| Kurdistan | Divandareh | 4·614 | 3·909 | 14·429 | 2·058 | 5·854 | 4222 | 1882 | 5366 |
| Kurdistan | Ghorveh | 4·745 | 4·249 | 15·493 | 1·970 | 6·073 | 7868 | 3245 | 10058 |
| Kurdistan | Kamyaran | 5·183 | 4·288 | 15·853 | 2·345 | 6·624 | 6182 | 2728 | 7819 |
| Kurdistan | Marivan | 4·950 | 3·624 | 13·630 | 2·486 | 6·289 | 11274 | 5895 | 14227 |
| Kurdistan | Sanandaj | 5·029 | 4·139 | 15·261 | 2·291 | 6·402 | 29277 | 13296 | 38045 |
| Kurdistan | Saqqez | 4·423 | 3·412 | 12·619 | 2·127 | 5·645 | 11905 | 5790 | 14981 |
| Kurdistan | Sarvabad | 5·257 | 3·881 | 14·353 | 2·630 | 6·674 | 2785 | 1384 | 3510 |
| Lorestan | Aligudarz | 1·730 | 1·717 | 6·168 | 0·684 | 2·144 | 2783 | 1108 | 3530 |
| Lorestan | Azna | 1·960 | 1·977 | 7·011 | 0·760 | 2·436 | 1695 | 645 | 2133 |
| Lorestan | Borujerd | 3·142 | 3·046 | 11·096 | 1·202 | 3·996 | 12067 | 4567 | 15111 |
| Lorestan | Delfan | 4·395 | 4·018 | 14·698 | 1·743 | 5·660 | 6898 | 2771 | 8927 |
| Lorestan | Doreh | 4·111 | 3·954 | 14·366 | 1·555 | 5·244 | 2093 | 794 | 2685 |
| Lorestan | Dorud | 2·435 | 2·403 | 8·615 | 0·938 | 3·055 | 4838 | 1901 | 6022 |
| Lorestan | Khoramabad | 3·248 | 3·139 | 11·448 | 1·221 | 4·139 | 20013 | 7491 | 25710 |

| | | β-thalassaemia carrier prevalence | | | | | Estimated carrier counts | | |
|---|---|---|---|---|---|---|---|---|---|
| **ADM1** | **ADM2** | **Mean** | **SD** | **CI95%** | **Q25** | **Q75** | **Mean** | **Q25** | **Q75** |
| Lorestan | Kuhdasht | 4·989 | 4·658 | 17·032 | 1·936 | 6·394 | 9292 | 3522 | 11900 |
| Lorestan | Pol-e-Dokhtar | 4·160 | 3·975 | 14·517 | 1·593 | 5·324 | 3837 | 1470 | 4932 |
| Lorestan | Rumeshkhan | 4·908 | 4·533 | 16·589 | 1·954 | 6·269 | 2094 | 820 | 2687 |
| Lorestan | Selseleh | 3·910 | 3·727 | 13·630 | 1·472 | 5·033 | 3511 | 1321 | 4574 |
| Markazi | Arak | 2·280 | 2·217 | 7·902 | 0·915 | 2·850 | 15297 | 6184 | 19204 |
| Markazi | Ashtian | 2·572 | 2·400 | 8·610 | 1·049 | 3·250 | 527 | 212 | 665 |
| Markazi | Delijan | 1·390 | 1·224 | 4·464 | 0·612 | 1·745 | 780 | 337 | 998 |
| Markazi | Farahan | 3·167 | 2·983 | 10·857 | 1·261 | 4·005 | 1017 | 408 | 1288 |
| Markazi | Khomein | 1·485 | 1·459 | 5·185 | 0·607 | 1·827 | 1709 | 707 | 2107 |
| Markazi | Khondab | 3·061 | 2·943 | 10·816 | 1·196 | 3·847 | 1888 | 731 | 2373 |
| Markazi | Komijan | 3·604 | 3·393 | 12·414 | 1·419 | 4·573 | 1494 | 580 | 1912 |
| Markazi | Mahallat | 1·358 | 1·242 | 4·415 | 0·581 | 1·703 | 860 | 363 | 1079 |
| Markazi | Saveh | 4·124 | 3·669 | 13·453 | 1·731 | 5·263 | 12921 | 5625 | 16256 |
| Markazi | Shazand | 2·463 | 2·429 | 8·736 | 0·964 | 3·080 | 3597 | 1403 | 4518 |
| Markazi | Tafresh | 3·505 | 3·224 | 11·670 | 1·439 | 4·457 | 941 | 392 | 1201 |
| Markazi | Zarandieh | 5·123 | 4·353 | 16·046 | 2·242 | 6·550 | 4455 | 1989 | 5713 |
| Mazandaran | Abbasabad | 8·658 | 6·803 | 25·777 | 3·948 | 11·195 | 4250 | 1907 | 5500 |
| Mazandaran | Amol | 8·364 | 6·289 | 23·666 | 4·023 | 10·793 | 43446 | 20831 | 56381 |
| Mazandaran | Babol | 8·542 | 6·512 | 24·419 | 4·034 | 11·066 | 53070 | 24847 | 68730 |
| Mazandaran | Babolsar | 9·462 | 7·200 | 26·968 | 4·490 | 12·359 | 16744 | 7887 | 21947 |
| Mazandaran | Bandar-e-Gaz | 8·585 | 6·428 | 23·789 | 4·088 | 11·207 | 3801 | 1815 | 4993 |
| Mazandaran | Behshahr | 8·637 | 6·686 | 24·788 | 4·114 | 11·089 | 16067 | 7694 | 20819 |
| Mazandaran | Chalus | 8·632 | 6·505 | 24·346 | 4·102 | 11·154 | 11337 | 5251 | 14619 |
| Mazandaran | Fereydunkenar | 9·595 | 7·248 | 27·692 | 4·532 | 12·411 | 7713 | 3597 | 9990 |
| Mazandaran | Galugah | 8·601 | 6·536 | 24·601 | 4·121 | 10·997 | 4542 | 2173 | 5793 |
| Mazandaran | Ghaemshahr | 8·686 | 6·770 | 25·151 | 4·110 | 11·202 | 31918 | 15161 | 41081 |
| Mazandaran | Juybar | 9·331 | 7·152 | 26·878 | 4·389 | 12·181 | 7409 | 3431 | 9621 |
| Mazandaran | Kelardasht | 8·465 | 6·467 | 24·433 | 3·979 | 10·939 | 2178 | 1040 | 2809 |
| Mazandaran | Mahmudabad | 9·766 | 7·477 | 28·651 | 4·554 | 12·832 | 8427 | 3925 | 11119 |
| Mazandaran | Miandorud | 8·836 | 6·847 | 25·336 | 4·183 | 11·345 | 5127 | 2425 | 6585 |
| Mazandaran | Neka | 8·315 | 6·492 | 24·149 | 3·905 | 10·696 | 12316 | 5695 | 15913 |
| Mazandaran | Noshahr | 9·101 | 6·826 | 25·746 | 4·364 | 11·777 | 13610 | 6368 | 17472 |
| Mazandaran | Nur | 8·996 | 6·694 | 25·098 | 4·351 | 11·663 | 12243 | 5798 | 15830 |
| Mazandaran | Ramsar | 7·638 | 6·388 | 24·184 | 3·321 | 9·871 | 5541 | 2487 | 7056 |
| Mazandaran | Sari | 7·937 | 6·390 | 23·675 | 3·657 | 10·175 | 51255 | 24055 | 65885 |
| Mazandaran | Savadkuh-e-Shomali | 8·254 | 6·441 | 23·905 | 3·864 | 10·689 | 2264 | 1056 | 2932 |
| Mazandaran | Savdkuh | 7·424 | 5·987 | 22·170 | 3·412 | 9·541 | 3876 | 1792 | 4948 |
| Mazandaran | Simorgh | 9·116 | 7·086 | 26·836 | 4·304 | 11·793 | 2306 | 1091 | 2970 |
| Mazandaran | Tonekabon | 8·131 | 6·516 | 24·448 | 3·662 | 10·517 | 12919 | 5749 | 16798 |
| North Khorasan | Bojnurd | 7·149 | 7·983 | 29·692 | 2·122 | 9·037 | 26998 | 7967 | 34397 |
| North Khorasan | Esfarayen County | 6·298 | 7·092 | 26·200 | 1·812 | 7·992 | 8978 | 2597 | 11389 |
| North Khorasan | Faruj | 6·274 | 7·398 | 27·595 | 1·663 | 7·885 | 3534 | 957 | 4434 |
| North Khorasan | Garmeh | 7·412 | 7·286 | 27·031 | 2·646 | 9·526 | 2168 | 774 | 2789 |
| North Khorasan | Jajrom | 6·876 | 7·239 | 26·617 | 2·247 | 8·799 | 2954 | 986 | 3785 |
| North Khorasan | Maneh and Samalqan | 7·938 | 8·275 | 30·512 | 2·592 | 10·142 | 8928 | 2779 | 11474 |
| North Khorasan | Raz and Jargalan | 8·201 | 8·958 | 33·328 | 2·468 | 10·462 | 5463 | 1647 | 7010 |
| North Khorasan | Shirwan | 6·764 | 7·866 | 28·850 | 1·870 | 8·543 | 11210 | 3087 | 14069 |
| Qazvin | Abyek | 6·967 | 5·744 | 21·429 | 3·098 | 8·954 | 7436 | 3373 | 9553 |
| Qazvin | Alborz | 6·782 | 5·667 | 21·093 | 2·949 | 8·780 | 19428 | 8602 | 25141 |
| Qazvin | Avaj | 5·069 | 4·501 | 16·602 | 2·090 | 6·516 | 2582 | 1053 | 3320 |
| Qazvin | Buin Zahra | 5·871 | 4·987 | 18·462 | 2·545 | 7·540 | 8746 | 3839 | 11200 |
| Qazvin | Qazvin | 6·458 | 5·527 | 20·333 | 2·769 | 8·320 | 43247 | 18525 | 56197 |
| Qazvin | Takestan | 5·706 | 4·958 | 18·171 | 2·414 | 7·352 | 12901 | 5518 | 16619 |
| Qom | Qom | 2·820 | 2·454 | 8·917 | 1·237 | 3·570 | 39740 | 17697 | 50452 |
| Razavi Khorasan | Bajestan | 2·958 | 4·117 | 14·205 | 0·733 | 3·466 | 1071 | 266 | 1269 |
| Razavi Khorasan | Bakharz | 4·253 | 6·119 | 21·613 | 0·893 | 4·904 | 2658 | 549 | 3105 |
| Razavi Khorasan | Bardaskan | 3·664 | 4·810 | 16·996 | 0·985 | 4·340 | 3226 | 853 | 3768 |

| | | β-thalassaemia carrier prevalence | | | | | Estimated carrier counts | | |
|---|---|---|---|---|---|---|---|---|---|
| **ADM1** | **ADM2** | **Mean** | **SD** | **CI95%** | **Q25** | **Q75** | **Mean** | **Q25** | **Q75** |
| Razavi Khorasan | Binalud | 5·126 | 6·608 | 23·860 | 1·215 | 6·321 | 6942 | 1610 | 8590 |
| Razavi Khorasan | Chenaran | 5·561 | 7·052 | 25·797 | 1·342 | 6·824 | 10010 | 2404 | 12195 |
| Razavi Khorasan | Dargaz | 6·117 | 7·687 | 28·351 | 1·503 | 7·516 | 5246 | 1290 | 6398 |
| Razavi Khorasan | Davarzan | 5·284 | 5·794 | 21·141 | 1·631 | 6·734 | 1335 | 418 | 1715 |
| Razavi Khorasan | Fariman | 4·577 | 6·143 | 21·983 | 1·033 | 5·538 | 5285 | 1196 | 6448 |
| Razavi Khorasan | Firuzeh | 5·130 | 6·281 | 22·981 | 1·334 | 6·318 | 2352 | 607 | 2897 |
| Razavi Khorasan | Gonabad | 3·023 | 4·485 | 15·047 | 0·695 | 3·455 | 3065 | 712 | 3493 |
| Razavi Khorasan | Joghatai | 6·047 | 6·553 | 23·967 | 1·888 | 7·680 | 3449 | 1077 | 4368 |
| Razavi Khorasan | Jowayin | 5·736 | 6·398 | 23·843 | 1·664 | 7·305 | 3687 | 1076 | 4672 |
| Razavi Khorasan | Kalat | 5·570 | 7·373 | 26·580 | 1·274 | 6·756 | 2389 | 536 | 2904 |
| Razavi Khorasan | Kashmar | 4·060 | 5·569 | 19·414 | 1·008 | 4·765 | 7860 | 1910 | 9056 |
| Razavi Khorasan | Khaf | 3·751 | 5·567 | 19·503 | 0·769 | 4·274 | 6105 | 1256 | 7083 |
| Razavi Khorasan | Khalilabad | 3·704 | 5·067 | 17·942 | 0·943 | 4·293 | 2215 | 558 | 2559 |
| Razavi Khorasan | Khoshab | 5·396 | 6·368 | 23·591 | 1·466 | 6·709 | 2313 | 626 | 2846 |
| Razavi Khorasan | Mahvelat | 3·769 | 5·403 | 18·571 | 0·881 | 4·353 | 2318 | 531 | 2664 |
| Razavi Khorasan | Mashhad | 5·054 | 6·629 | 23·678 | 1·166 | 6·181 | 200489 | 46909 | 245573 |
| Razavi Khorasan | Neyshabur | 5·023 | 6·409 | 23·102 | 1·253 | 6·132 | 26110 | 6560 | 31238 |
| Razavi Khorasan | Quchan | 6·063 | 7·406 | 27·240 | 1·548 | 7·532 | 12254 | 3203 | 15232 |
| Razavi Khorasan | Roshtkhar | 3·883 | 5·717 | 19·565 | 0·844 | 4·463 | 2772 | 597 | 3181 |
| Razavi Khorasan | Sabzevar | 4·581 | 5·474 | 19·955 | 1·287 | 5·671 | 17777 | 4982 | 22075 |
| Razavi Khorasan | Sarakhs | 5·087 | 6·916 | 24·582 | 1·127 | 6·129 | 5547 | 1220 | 6798 |
| Razavi Khorasan | Taybad | 4·203 | 6·166 | 21·945 | 0·847 | 4·843 | 5913 | 1221 | 6978 |
| Razavi Khorasan | Torbat-e-Heydarieh | 4·292 | 5·909 | 20·525 | 1·003 | 5·110 | 10868 | 2482 | 12778 |
| Razavi Khorasan | Torbat-e-jam | 4·610 | 6·413 | 22·840 | 0·982 | 5·454 | 13622 | 2858 | 16232 |
| Razavi Khorasan | Zaveh | 4·267 | 6·052 | 20·745 | 0·939 | 5·050 | 3306 | 725 | 3890 |
| Semnan | Aradan | 3·705 | 3·080 | 11·277 | 1·691 | 4·717 | 680 | 313 | 860 |
| Semnan | Damghan | 5·539 | 4·654 | 17·066 | 2·500 | 7·036 | 6982 | 3177 | 8852 |
| Semnan | Garmsar | 4·159 | 3·349 | 12·303 | 1·954 | 5·292 | 4241 | 2001 | 5399 |
| Semnan | Mehdishahr | 5·713 | 4·824 | 17·934 | 2·495 | 7·334 | 2256 | 974 | 2938 |
| Semnan | Miami | 6·404 | 6·199 | 22·881 | 2·377 | 8·187 | 3245 | 1237 | 4154 |
| Semnan | Semnan | 4·034 | 3·550 | 12·957 | 1·740 | 5·108 | 11167 | 4887 | 14240 |
| Semnan | Shahmirzad | 6·413 | 5·284 | 19·515 | 2·881 | 8·224 | 670 | 302 | 866 |
| Semnan | Shahrud | 5·281 | 4·848 | 17·697 | 2·153 | 6·713 | 16945 | 7769 | 21681 |
| Semnan | Sorkheh | 3·841 | 3·319 | 12·114 | 1·683 | 4·877 | 857 | 370 | 1089 |
| Sistan and Baluchestan | Chabahar | 3·121 | 3·628 | 13·062 | 0·911 | 3·924 | 5185 | 1500 | 6475 |
| Sistan and Baluchestan | Dalagan | 1·417 | 1·712 | 5·706 | 0·459 | 1·725 | 876 | 284 | 1051 |
| Sistan and Baluchestan | Fanuj | 1·680 | 1·990 | 6·817 | 0·541 | 2·052 | 977 | 313 | 1212 |
| Sistan and Baluchestan | Hamun | 2·726 | 4·091 | 13·591 | 0·608 | 3·106 | 1438 | 320 | 1619 |
| Sistan and Baluchestan | Hirmand | 3·149 | 5·109 | 16·068 | 0·667 | 3·468 | 2789 | 586 | 3065 |
| Sistan and Baluchestan | Iranshahr | 1·945 | 2·466 | 8·522 | 0·552 | 2·346 | 6422 | 1777 | 7636 |
| Sistan and Baluchestan | Khash | 2·336 | 3·184 | 10·923 | 0·604 | 2·757 | 4469 | 1137 | 5286 |
| Sistan and Baluchestan | Konarak | 2·328 | 2·575 | 9·179 | 0·753 | 2·923 | 2692 | 839 | 3415 |
| Sistan and Baluchestan | Mehrestan | 2·992 | 3·937 | 13·665 | 0·781 | 3·600 | 2384 | 611 | 2893 |
| Sistan and Baluchestan | Mirjaveh | 2·460 | 3·366 | 11·717 | 0·606 | 2·894 | 1268 | 304 | 1468 |
| Sistan and Baluchestan | Nikshahr | 2·022 | 2·349 | 8·262 | 0·626 | 2·500 | 3376 | 1017 | 4128 |
| Sistan and Baluchestan | Nimruz | 2·608 | 4·141 | 13·292 | 0·572 | 2·895 | 1084 | 230 | 1238 |
| Sistan and Baluchestan | Qasr-e-Ghand | 2·662 | 3·192 | 11·299 | 0·777 | 3·273 | 1831 | 527 | 2291 |
| Sistan and Baluchestan | Saravan | 3·609 | 4·796 | 16·249 | 0·932 | 4·347 | 7640 | 1894 | 9105 |
| Sistan and Baluchestan | Sarbaz | 2·966 | 3·718 | 13·145 | 0·816 | 3·613 | 6240 | 1712 | 7566 |
| Sistan and Baluchestan | Sib and Suran | 3·060 | 4·122 | 14·019 | 0·783 | 3·657 | 3035 | 756 | 3632 |
| Sistan and Baluchestan | Zabol | 3·052 | 4·738 | 15·134 | 0·662 | 3·406 | 6065 | 1351 | 6830 |
| Sistan and Baluchestan | Zahak | 3·053 | 4·672 | 15·565 | 0·655 | 3·455 | 2692 | 573 | 3093 |
| Sistan and Baluchestan | Zahedan | 2·102 | 2·980 | 10·268 | 0·511 | 2·432 | 17712 | 4234 | 20828 |
| South Khorasan | Birjand | 2·453 | 3·729 | 12·455 | 0·521 | 2·781 | 7042 | 1506 | 7973 |
| South Khorasan | Boshruyeh | 1·904 | 2·674 | 8·726 | 0·498 | 2·236 | 550 | 141 | 664 |
| South Khorasan | Darmian | 2·860 | 4·355 | 15·023 | 0·586 | 3·255 | 1756 | 357 | 2003 |
| South Khorasan | Ferdows | 2·377 | 3·476 | 11·369 | 0·576 | 2·774 | 1344 | 307 | 1571 |

| | | β-thalassaemia carrier prevalence | | | | | Estimated carrier counts | | |
|---|---|---|---|---|---|---|---|---|---|
| ADM1 | ADM2 | Mean | SD | CI95% | Q25 | Q75 | Mean | Q25 | Q75 |
| South Khorasan | Ghaenat | 2·734 | 4·168 | 13·876 | 0·591 | 3·109 | 3490 | 763 | 4013 |
| South Khorasan | Khusf | 2·007 | 3·199 | 10·155 | 0·433 | 2·253 | 699 | 144 | 793 |
| South Khorasan | Nehbandan | 2·362 | 3·852 | 12·148 | 0·502 | 2·610 | 1339 | 283 | 1485 |
| South Khorasan | Sarayan | 2·160 | 3·335 | 10·475 | 0·494 | 2·477 | 965 | 212 | 1106 |
| South Khorasan | Sarbisheh | 2·621 | 4·109 | 13·655 | 0·542 | 2·937 | 1236 | 239 | 1378 |
| South Khorasan | Tabas | 1·495 | 2·074 | 6·716 | 0·407 | 1·757 | 1313 | 342 | 1537 |
| South Khorasan | Zirkuh | 3·157 | 4·780 | 16·663 | 0·636 | 3·559 | 1666 | 329 | 1890 |
| Tehran | Baharestan | 6·438 | 4·916 | 18·285 | 3·234 | 8·258 | 29038 | 14588 | 37253 |
| Tehran | City of Tehran | 7·343 | 5·296 | 19·582 | 3·662 | 9·482 | 690333 | 345137 | 890215 |
| Tehran | Damavand | 6·229 | 4·728 | 17·485 | 3·026 | 7·970 | 9522 | 4792 | 12195 |
| Tehran | Firuzkuh | 6·131 | 4·928 | 18·198 | 2·849 | 7·850 | 2435 | 1116 | 3101 |
| Tehran | Gharchak | 5·753 | 4·281 | 15·991 | 2·917 | 7·304 | 21512 | 10982 | 27359 |
| Tehran | Islamshahr | 6·631 | 5·020 | 18·477 | 3·247 | 8·521 | 43318 | 21325 | 55456 |
| Tehran | Malard | 6·045 | 4·797 | 17·806 | 2·793 | 7·757 | 30396 | 14381 | 38954 |
| Tehran | Pakdasht | 5·875 | 4·438 | 16·509 | 2·885 | 7·505 | 25682 | 12598 | 32476 |
| Tehran | Paveh | 6·930 | 4·974 | 18·503 | 3·558 | 8·924 | 14648 | 7545 | 19082 |
| Tehran | Pishva | 5·313 | 4·084 | 15·248 | 2·585 | 6·778 | 4069 | 1970 | 5166 |
| Tehran | Qods | 6·725 | 5·016 | 18·614 | 3·292 | 8·543 | 13010 | 6369 | 16531 |
| Tehran | Rey | 5·490 | 4·231 | 15·773 | 2·646 | 7·013 | 36375 | 17984 | 46849 |
| Tehran | Robatkarim | 6·105 | 4·664 | 17·386 | 2·955 | 7·756 | 26362 | 12543 | 33628 |
| Tehran | Shahriar | 6·410 | 4·871 | 18·279 | 3·103 | 8·244 | 64001 | 31360 | 82851 |
| Tehran | Shemiranat | 8·017 | 5·727 | 21·149 | 4·036 | 10·372 | 22223 | 11027 | 28560 |
| Tehran | Tehran | 7·326 | 5·318 | 19·998 | 3·646 | 9·413 | 39964 | 19864 | 50981 |
| Tehran | Varamin | 4·338 | 3·419 | 12·541 | 2·049 | 5·535 | 15209 | 7300 | 19316 |
| West Azerbaijan | Ashnavieh | 4·289 | 3·041 | 11·262 | 2·236 | 5·445 | 3746 | 1956 | 4803 |
| West Azerbaijan | Bukan | 4·173 | 3·202 | 11·899 | 2·024 | 5·315 | 11786 | 5766 | 14819 |
| West Azerbaijan | Chaldoran County | 2·123 | 2·027 | 7·248 | 0·847 | 2·689 | 1116 | 455 | 1390 |
| West Azerbaijan | Chaypareh | 2·396 | 2·253 | 8·222 | 0·981 | 3·018 | 1378 | 554 | 1711 |
| West Azerbaijan | Khoy | 2·556 | 2·356 | 8·528 | 1·063 | 3·229 | 10724 | 4410 | 13398 |
| West Azerbaijan | Mahabad | 4·194 | 3·091 | 11·451 | 2·108 | 5·333 | 11398 | 5682 | 14364 |
| West Azerbaijan | Maku | 2·020 | 2·041 | 7·205 | 0·761 | 2·543 | 2271 | 874 | 2855 |
| West Azerbaijan | Miandoab | 3·858 | 3·075 | 11·290 | 1·839 | 4·885 | 11820 | 5655 | 14995 |
| West Azerbaijan | Naghadeh | 4·128 | 3·051 | 11·424 | 2·067 | 5·249 | 6192 | 3111 | 7870 |
| West Azerbaijan | Piranshahr | 4·448 | 2·976 | 11·200 | 2·405 | 5·627 | 7222 | 3845 | 9191 |
| West Azerbaijan | Poldasht | 2·237 | 2·196 | 7·867 | 0·875 | 2·817 | 1137 | 436 | 1438 |
| West Azerbaijan | Salmas | 2·968 | 2·549 | 9·270 | 1·317 | 3·756 | 6633 | 2910 | 8268 |
| West Azerbaijan | Sardasht | 4·534 | 2·946 | 11·079 | 2·510 | 5·718 | 6337 | 3492 | 7950 |
| West Azerbaijan | Shahindezh | 4·092 | 3·323 | 12·287 | 1·881 | 5·221 | 4224 | 1961 | 5260 |
| West Azerbaijan | Showt | 2·190 | 2·077 | 7·499 | 0·874 | 2·786 | 1411 | 559 | 1805 |
| West Azerbaijan | Takab | 4·216 | 3·586 | 13·212 | 1·855 | 5·374 | 3988 | 1758 | 5057 |
| West Azerbaijan | Urumia | 3·594 | 2·813 | 10·321 | 1·733 | 4·570 | 44337 | 21661 | 56363 |
| Yazd | Abarkuh | 0·374 | 0·375 | 1·303 | 0·149 | 0·463 | 219 | 85 | 271 |
| Yazd | Ardekan | 0·448 | 0·488 | 1·676 | 0·166 | 0·548 | 287 | 112 | 357 |
| Yazd | Ashkezar | 0·337 | 0·342 | 1·218 | 0·130 | 0·416 | 160 | 59 | 201 |
| Yazd | Bafq | 0·541 | 0·668 | 2·232 | 0·173 | 0·650 | 311 | 96 | 375 |
| Yazd | Bahabad | 0·700 | 0·935 | 3·038 | 0·205 | 0·829 | 126 | 39 | 154 |
| Yazd | Khatam | 0·474 | 0·482 | 1·702 | 0·185 | 0·587 | 195 | 75 | 246 |
| Yazd | Mehriz | 0·413 | 0·463 | 1·585 | 0·145 | 0·504 | 232 | 80 | 285 |
| Yazd | Meybod | 0·315 | 0·317 | 1·109 | 0·125 | 0·386 | 457 | 178 | 576 |
| Yazd | Taft | 0·354 | 0·367 | 1·291 | 0·134 | 0·438 | 181 | 60 | 223 |
| Yazd | Yazd | 0·379 | 0·410 | 1·402 | 0·137 | 0·468 | 2777 | 1011 | 3474 |
| Zanjan | Abhar | 5·402 | 4·819 | 17·575 | 2·244 | 6·964 | 8396 | 3522 | 10790 |
| Zanjan | Ijrud | 4·709 | 4·195 | 15·246 | 1·953 | 6·044 | 2004 | 834 | 2558 |
| Zanjan | Khodabandeh | 4·891 | 4·387 | 15·938 | 2·021 | 6·283 | 9412 | 3881 | 12115 |
| Zanjan | Khoramdareh | 5·547 | 4·918 | 17·991 | 2·293 | 7·160 | 5395 | 2234 | 7042 |
| Zanjan | Mahneshan | 4·218 | 3·616 | 13·280 | 1·826 | 5·414 | 1925 | 814 | 2504 |
| Zanjan | Soltanieh | 5·061 | 4·435 | 16·162 | 2·113 | 6·578 | 1671 | 686 | 2163 |

| ADM1 | ADM2 | β-thalassaemia carrier prevalence Mean | SD | CI95% | Q25 | Q75 | Estimated carrier counts Mean | Q25 | Q75 |
|---|---|---|---|---|---|---|---|---|---|
| Zanjan | Tarom | 4·763 | 4·085 | 15·046 | 2·065 | 6·114 | 2645 | 1137 | 3447 |
| Zanjan | Zanjan | 4·315 | 3·672 | 13·501 | 1·880 | 5·559 | 27666 | 11991 | 35779 |

ADM1: administrative level 1. ADM2: administrative level 2. SD: standard deviation. CI95%: 95% credible interval. Q25: 25th percentile. Q75: 75th percentile. SD and CI95% are measures of prediction uncertainty.

**Supplementary Table S24. Subnational areal predictions of β-thalassaemia carrier prevalence and estimated carrier count at administrative level 2 of Iraq.**

| ADM1 | ADM2 | β-thalassaemia carrier prevalence Mean | SD | CI95% | Q25 | Q75 | Estimated carrier counts Mean | Q25 | Q75 |
|---|---|---|---|---|---|---|---|---|---|
| Al-Anbar | Al-Falluja | 5·522 | 5·342 | 19·374 | 2·129 | 7·007 | 47038 | 17983 | 60382 |
| Al-Anbar | Al-Kadhmiyah | 5·977 | 5·629 | 20·507 | 2·377 | 7·614 | 117311 | 46422 | 149551 |
| Al-Anbar | Al-Kaim | 4·640 | 4·899 | 17·876 | 1·604 | 5·794 | 10800 | 3692 | 13346 |
| Al-Anbar | Al-Ramadi | 4·989 | 4·945 | 17·925 | 1·852 | 6·341 | 34641 | 13242 | 44200 |
| Al-Anbar | Al-Rutba | 2·719 | 3·130 | 10·812 | 0·913 | 3·326 | 1482 | 477 | 1829 |
| Al-Anbar | Ana | 5·136 | 5·085 | 18·651 | 1·884 | 6·524 | 3702 | 1384 | 4681 |
| Al-Anbar | Haditha | 5·329 | 5·164 | 18·944 | 1·994 | 6·806 | 6973 | 2559 | 8810 |
| Al-Anbar | Heet | 5·071 | 4·945 | 18·070 | 1·890 | 6·464 | 10738 | 4052 | 13487 |
| Al-Basrah | Abu Al-Khaseeb | 4·517 | 3·011 | 11·215 | 2·440 | 5·754 | 14663 | 7967 | 18700 |
| Al-Basrah | Al-Basrah | 5·065 | 3·245 | 12·169 | 2·833 | 6·397 | 79070 | 43645 | 99456 |
| Al-Basrah | Al-Faw | 4·193 | 3·001 | 11·294 | 2·145 | 5·348 | 1320 | 661 | 1639 |
| Al-Basrah | Al-Midaina | 5·448 | 3·524 | 13·155 | 3·003 | 6·922 | 13287 | 7350 | 16866 |
| Al-Basrah | Al-Qurna | 5·246 | 3·383 | 12·701 | 2·900 | 6·654 | 17128 | 9415 | 21731 |
| Al-Basrah | Al-Zubair | 4·506 | 2·951 | 11·073 | 2·473 | 5·711 | 28523 | 15803 | 36133 |
| Al-Basrah | Shat Al-Arab | 4·819 | 3·143 | 11·809 | 2·655 | 6·110 | 24924 | 13951 | 31483 |
| Al-Muthanna | Al-Khidhir | 4·151 | 3·792 | 13·706 | 1·745 | 5·218 | 4916 | 2075 | 6185 |
| Al-Muthanna | Al-Rumaitha | 4·265 | 4·133 | 14·868 | 1·673 | 5·344 | 16868 | 6593 | 20941 |
| Al-Muthanna | Al-Salman | 2·314 | 2·070 | 7·478 | 1·007 | 2·905 | 495 | 198 | 616 |
| Al-Muthanna | Al-Samawa | 3·798 | 3·562 | 13·064 | 1·544 | 4·779 | 17950 | 7326 | 22242 |
| Al-Qadisiyah | Afaq | 5·132 | 4·965 | 18·056 | 1·993 | 6·480 | 10723 | 4156 | 13551 |
| Al-Qadisiyah | Al-Diwaniya | 4·630 | 4·653 | 16·675 | 1·755 | 5·785 | 35049 | 13303 | 43337 |
| Al-Qadisiyah | Al-Hamza | 3·984 | 4·055 | 14·545 | 1·501 | 4·955 | 11440 | 4355 | 14307 |
| Al-Qadisiyah | Al-Shamiya | 4·172 | 4·278 | 15·402 | 1·536 | 5·179 | 14675 | 5316 | 18329 |
| Al-Sulaimaniyah | Al-Sulaymaniyah | 5·371 | 3·313 | 12·489 | 3·078 | 6·776 | 59193 | 34112 | 74271 |
| Al-Sulaimaniyah | Chamchamal | 5·930 | 3·670 | 13·822 | 3·371 | 7·485 | 12564 | 7218 | 15736 |
| Al-Sulaimaniyah | Derbendikhan | 5·532 | 3·596 | 13·672 | 3·047 | 6·964 | 6103 | 3325 | 7720 |
| Al-Sulaimaniyah | Dokan | 5·098 | 3·075 | 11·620 | 2·954 | 6·416 | 3898 | 2283 | 4919 |
| Al-Sulaimaniyah | Halabcha | 5·445 | 3·728 | 14·011 | 2·890 | 6·921 | 12741 | 6884 | 16063 |
| Al-Sulaimaniyah | Kalar | 6·252 | 4·067 | 15·285 | 3·434 | 7·950 | 13373 | 7050 | 17076 |
| Al-Sulaimaniyah | Panjwin | 4·837 | 3·347 | 12·399 | 2·561 | 6·134 | 2722 | 1463 | 3446 |
| Al-Sulaimaniyah | Pshdar | 4·744 | 3·005 | 11·284 | 2·664 | 5·991 | 7993 | 4547 | 10091 |
| Al-Sulaimaniyah | Rania | 4·985 | 3·062 | 11·520 | 2·852 | 6·316 | 14825 | 8472 | 18627 |
| Al-Sulaimaniyah | Sharbazher | 4·905 | 3·124 | 11·766 | 2·738 | 6·207 | 2884 | 1603 | 3662 |
| An-Najaf | Al-Kufa | 4·402 | 4·599 | 16·680 | 1·594 | 5·452 | 20007 | 7267 | 24842 |
| An-Najaf | Al-Manathera | 3·820 | 4·025 | 14·277 | 1·403 | 4·743 | 14321 | 5232 | 17808 |
| An-Najaf | Al-Najaf | 2·900 | 3·081 | 10·937 | 1·059 | 3·580 | 42674 | 15291 | 51759 |
| Babil | Al-Hashimiya | 5·169 | 5·110 | 18·632 | 1·959 | 6·507 | 31386 | 11718 | 39345 |
| Babil | Al-Hilla | 4·869 | 5·004 | 18·228 | 1·791 | 6·065 | 51420 | 18730 | 64006 |
| Babil | Al-Hindiya | 4·773 | 4·916 | 17·792 | 1·724 | 5·963 | 19621 | 7045 | 24505 |
| Babil | Al-Mahaweel | 5·494 | 5·478 | 19·807 | 2·067 | 6·909 | 22231 | 8307 | 27947 |
| Babil | Al-Mahmoudiya | 5·635 | 5·461 | 19·835 | 2·172 | 7·129 | 46030 | 17994 | 58767 |
| Babil | Al-Mussyab | 5·357 | 5·400 | 19·474 | 1·984 | 6·759 | 25479 | 9404 | 32152 |
| Baghdad | Al-Adhamiya | 6·225 | 5·720 | 21·037 | 2·471 | 8·030 | 84231 | 33393 | 109990 |
| Baghdad | Al-Karkh | 5·831 | 5·508 | 19·636 | 2·277 | 7·428 | 88672 | 34595 | 112485 |
| Baghdad | Al-Risafa | 6·022 | 5·604 | 20·260 | 2·404 | 7·723 | 126798 | 50642 | 162888 |
| Baghdad | Al-Thawra | 5·971 | 5·306 | 19·408 | 2·490 | 7·802 | 106497 | 44373 | 138700 |
| Dhi Qar | Al-Chibayish | 5·151 | 3·458 | 12·958 | 2·760 | 6·541 | 6157 | 3270 | 7866 |
| Dhi Qar | Al-Nasiriya | 4·804 | 3·807 | 14·064 | 2·265 | 6·130 | 45908 | 21442 | 58358 |

| ADM1 | ADM2 | β-thalassaemia carrier prevalence Mean | SD | CI95% | Q25 | Q75 | Estimated carrier counts Mean | Q25 | Q75 |
|---|---|---|---|---|---|---|---|---|---|
| Dhi Qar | Al-Rifai | 5·377 | 4·729 | 17·491 | 2·289 | 6·871 | 29525 | 12418 | 37886 |
| Dhi Qar | Al-Shatra | 5·189 | 4·196 | 15·515 | 2·380 | 6·631 | 30017 | 13830 | 38114 |
| Dhi Qar | Suq Al-Shoyokh | 4·858 | 3·646 | 13·573 | 2·411 | 6·171 | 20078 | 9827 | 25386 |
| Diyala | Al-Khalis | 6·514 | 5·191 | 19·301 | 3·005 | 8·374 | 27967 | 12331 | 36053 |
| Diyala | Al-Mada'in | 6·015 | 5·528 | 20·210 | 2·434 | 7·640 | 34138 | 13777 | 43263 |
| Diyala | Al-Muqdadiya | 6·554 | 5·324 | 19·834 | 2·929 | 8·446 | 20865 | 9272 | 26977 |
| Diyala | Baladruz | 6·472 | 5·521 | 20·554 | 2·773 | 8·320 | 12005 | 5236 | 15553 |
| Diyala | Baquba | 6·356 | 5·579 | 20·722 | 2·677 | 8·133 | 49229 | 20762 | 63086 |
| Diyala | Khanaqin | 6·451 | 4·687 | 17·509 | 3·225 | 8·267 | 21251 | 10371 | 27228 |
| Diyala | Kifri | 6·585 | 4·638 | 17·440 | 3·374 | 8·444 | 10001 | 5190 | 12882 |
| Dohuk | Al-Amadiya | 4·136 | 2·694 | 10·097 | 2·269 | 5·228 | 6243 | 3421 | 7864 |
| Dohuk | Duhok | 4·084 | 2·627 | 9·916 | 2·277 | 5·156 | 19271 | 10686 | 24396 |
| Dohuk | Sumail | 4·001 | 2·667 | 10·023 | 2·173 | 5·070 | 10989 | 6052 | 13807 |
| Dohuk | Zakho | 3·571 | 2·382 | 8·844 | 1·952 | 4·493 | 11875 | 6546 | 15179 |
| Erbil | Al-Zibar | 4·436 | 2·871 | 10·864 | 2·451 | 5·626 | 1169 | 648 | 1481 |
| Erbil | Erbil | 6·088 | 3·871 | 14·501 | 3·403 | 7·722 | 93541 | 52754 | 118282 |
| Erbil | Koysinjaq | 5·557 | 3·420 | 12·847 | 3·178 | 7·022 | 7842 | 4502 | 9891 |
| Erbil | Makhmour | 6·340 | 4·338 | 16·301 | 3·344 | 8·090 | 13656 | 7278 | 17339 |
| Erbil | Rawanduz | 4·936 | 3·192 | 11·981 | 2·736 | 6·243 | 17730 | 9839 | 22506 |
| Erbil | Shaqlawa | 5·549 | 3·389 | 12·820 | 3·186 | 7·021 | 11387 | 6495 | 14412 |
| Karbala | Ain Al-Tamur | 4·146 | 4·328 | 15·333 | 1·497 | 5·193 | 1484 | 514 | 1829 |
| Karbala | Kerbela | 4·740 | 4·880 | 17·478 | 1·736 | 5·913 | 54179 | 19861 | 67732 |
| Kirkuk | Al-Hawiga | 6·603 | 4·783 | 17·931 | 3·315 | 8·450 | 23084 | 11615 | 29505 |
| Kirkuk | Daquq | 6·588 | 4·702 | 17·551 | 3·360 | 8·408 | 7608 | 3936 | 9733 |
| Kirkuk | Dibis | 6·380 | 4·170 | 15·768 | 3·480 | 8·107 | 8246 | 4495 | 10494 |
| Kirkuk | Kirkuk | 6·242 | 3·993 | 15·018 | 3·468 | 7·899 | 87156 | 47606 | 110364 |
| Maysan | Al-Amara | 5·411 | 4·524 | 16·701 | 2·390 | 6·951 | 4915 | 2194 | 6293 |
| Maysan | Ali Al-Gharbi | 5·643 | 4·852 | 17·842 | 2·419 | 7·245 | 3615 | 1565 | 4656 |
| Maysan | Al-Kahla | 5·184 | 4·070 | 15·079 | 2·431 | 6·651 | 41502 | 19560 | 53474 |
| Maysan | Al-Maimouna | 5·614 | 4·335 | 16·072 | 2·685 | 7·206 | 7609 | 3636 | 9724 |
| Maysan | Al-Mejar Al-Kabir | 5·306 | 3·748 | 14·043 | 2·738 | 6·772 | 12846 | 6529 | 16280 |
| Maysan | Qalat Saleh | 5·328 | 3·795 | 14·141 | 2·741 | 6·798 | 4130 | 2091 | 5261 |
| Ninawa | Al-Baaj | 4·893 | 4·036 | 14·801 | 2·196 | 6·290 | 11747 | 5426 | 15108 |
| Ninawa | Al-Hamdaniya | 5·725 | 3·815 | 14·297 | 3·083 | 7·266 | 29879 | 16080 | 38044 |
| Ninawa | Al-Hatra | 5·776 | 4·620 | 17·136 | 2·661 | 7·414 | 4542 | 2169 | 5793 |
| Ninawa | Al-Mosul | 5·603 | 3·972 | 14·839 | 2·887 | 7·145 | 120328 | 63423 | 153536 |
| Ninawa | Al-Shikhan | 4·806 | 3·080 | 11·523 | 2·684 | 6·078 | 8725 | 4872 | 11006 |
| Ninawa | Aqra | 5·156 | 3·278 | 12·333 | 2·886 | 6·524 | 19658 | 10929 | 24886 |
| Ninawa | Sinjar | 4·448 | 3·451 | 12·852 | 2·117 | 5·692 | 15453 | 7431 | 19797 |
| Ninawa | Telafar | 4·371 | 3·127 | 11·564 | 2·249 | 5·561 | 28115 | 14507 | 35434 |
| Ninawa | Tilkaef | 4·781 | 3·130 | 11·812 | 2·609 | 6·057 | 15239 | 8168 | 19401 |
| Salah al-Din | Al-Daur | 6·510 | 5·068 | 18·829 | 3·078 | 8·380 | 5448 | 2493 | 7123 |
| Salah al-Din | Al-Shirqat | 6·376 | 4·907 | 18·187 | 3·057 | 8·176 | 17996 | 8743 | 23128 |
| Salah al-Din | Balad | 6·212 | 5·437 | 19·951 | 2·619 | 7·971 | 33310 | 14278 | 42739 |
| Salah al-Din | Beygee | 6·298 | 5·221 | 19·357 | 2·800 | 8·065 | 19555 | 9074 | 25096 |
| Salah al-Din | Samarra | 6·221 | 5·254 | 19·433 | 2·704 | 8·038 | 19869 | 8905 | 26005 |
| Salah al-Din | Tikrit | 6·440 | 5·095 | 19·030 | 2·994 | 8·260 | 18477 | 8379 | 23714 |
| Salah al-Din | Tooz Khurmato | 6·562 | 4·531 | 16·830 | 3·446 | 8·373 | 16454 | 8705 | 21051 |
| Wasit | Al-Hai | 5·737 | 5·230 | 19·043 | 2·345 | 7·349 | 13098 | 5332 | 16635 |
| Wasit | Al-Kut | 5·903 | 5·264 | 19·454 | 2·425 | 7·595 | 37994 | 15210 | 47991 |
| Wasit | Al-Namaniya | 5·826 | 5·515 | 20·005 | 2·262 | 7·440 | 14595 | 5553 | 18611 |
| Wasit | Al-Suwaira | 5·884 | 5·507 | 20·117 | 2·323 | 7·504 | 31809 | 12619 | 40833 |
| Wasit | Badra | 6·227 | 5·584 | 20·580 | 2·509 | 8·036 | 2297 | 912 | 2986 |
| Al-Anbar | Al-Falluja | 5·522 | 5·342 | 19·374 | 2·129 | 7·007 | 47038 | 17983 | 60382 |
| Al-Anbar | Al-Kadhmiyah | 5·977 | 5·629 | 20·507 | 2·377 | 7·614 | 117311 | 46422 | 149551 |
| Al-Anbar | Al-Kaim | 4·640 | 4·899 | 17·876 | 1·604 | 5·794 | 10800 | 3692 | 13346 |
| Al-Anbar | Al-Ramadi | 4·989 | 4·945 | 17·925 | 1·852 | 6·341 | 34641 | 13242 | 44200 |

| | | β-thalassaemia carrier prevalence | | | | | Estimated carrier counts | | |
|---|---|---|---|---|---|---|---|---|---|
| **ADM1** | **ADM2** | **Mean** | **SD** | **CI95%** | **Q25** | **Q75** | **Mean** | **Q25** | **Q75** |
| Al-Anbar | Al-Rutba | 2·719 | 3·130 | 10·812 | 0·913 | 3·326 | 1482 | 477 | 1829 |
| Al-Anbar | Ana | 5·136 | 5·085 | 18·651 | 1·884 | 6·524 | 3702 | 1384 | 4681 |
| Al-Anbar | Haditha | 5·329 | 5·164 | 18·944 | 1·994 | 6·806 | 6973 | 2559 | 8810 |
| Al-Anbar | Heet | 5·071 | 4·945 | 18·070 | 1·890 | 6·464 | 10738 | 4052 | 13487 |
| Al-Basrah | Abu Al-Khaseeb | 4·517 | 3·011 | 11·215 | 2·440 | 5·754 | 14663 | 7967 | 18700 |
| Al-Basrah | Al-Basrah | 5·065 | 3·245 | 12·169 | 2·833 | 6·397 | 79070 | 43645 | 99456 |
| Al-Basrah | Al-Faw | 4·193 | 3·001 | 11·294 | 2·145 | 5·348 | 1320 | 661 | 1639 |
| Al-Basrah | Al-Midaina | 5·448 | 3·524 | 13·155 | 3·003 | 6·922 | 13287 | 7350 | 16866 |
| Al-Basrah | Al-Qurna | 5·246 | 3·383 | 12·701 | 2·900 | 6·654 | 17128 | 9415 | 21731 |
| Al-Basrah | Al-Zubair | 4·506 | 2·951 | 11·073 | 2·473 | 5·711 | 28523 | 15803 | 36133 |
| Al-Basrah | Shat Al-Arab | 4·819 | 3·143 | 11·809 | 2·655 | 6·110 | 24924 | 13951 | 31483 |
| Al-Muthanna | Al-Khidhir | 4·151 | 3·792 | 13·706 | 1·745 | 5·218 | 4916 | 2075 | 6185 |
| Al-Muthanna | Al-Rumaitha | 4·265 | 4·133 | 14·868 | 1·673 | 5·344 | 16868 | 6593 | 20941 |
| Al-Muthanna | Al-Salman | 2·314 | 2·070 | 7·478 | 1·007 | 2·905 | 495 | 198 | 616 |
| Al-Muthanna | Al-Samawa | 3·798 | 3·562 | 13·064 | 1·544 | 4·779 | 17950 | 7326 | 22242 |
| Al-Qadisiyah | Afaq | 5·132 | 4·965 | 18·056 | 1·993 | 6·480 | 10723 | 4156 | 13551 |
| Al-Qadisiyah | Al-Diwaniya | 4·630 | 4·653 | 16·675 | 1·755 | 5·785 | 35049 | 13303 | 43337 |
| Al-Qadisiyah | Al-Hamza | 3·984 | 4·055 | 14·545 | 1·501 | 4·955 | 11440 | 4355 | 14307 |
| Al-Qadisiyah | Al-Shamiya | 4·172 | 4·278 | 15·402 | 1·536 | 5·179 | 14675 | 5316 | 18329 |
| Al-Sulaimaniyah | Al-Sulaymaniyah | 5·371 | 3·313 | 12·489 | 3·078 | 6·776 | 59193 | 34112 | 74271 |
| Al-Sulaimaniyah | Chamchamal | 5·930 | 3·670 | 13·822 | 3·371 | 7·485 | 12564 | 7218 | 15736 |
| Al-Sulaimaniyah | Derbendikhan | 5·532 | 3·596 | 13·672 | 3·047 | 6·964 | 6103 | 3325 | 7720 |
| Al-Sulaimaniyah | Dokan | 5·098 | 3·075 | 11·620 | 2·954 | 6·416 | 3898 | 2283 | 4919 |
| Al-Sulaimaniyah | Halabcha | 5·445 | 3·728 | 14·011 | 2·890 | 6·921 | 12741 | 6884 | 16063 |
| Al-Sulaimaniyah | Kalar | 6·252 | 4·067 | 15·285 | 3·434 | 7·950 | 13373 | 7050 | 17076 |
| Al-Sulaimaniyah | Panjwin | 4·837 | 3·347 | 12·399 | 2·561 | 6·134 | 2722 | 1463 | 3446 |
| Al-Sulaimaniyah | Pshdar | 4·744 | 3·005 | 11·284 | 2·664 | 5·991 | 7993 | 4547 | 10091 |
| Al-Sulaimaniyah | Rania | 4·985 | 3·062 | 11·520 | 2·852 | 6·316 | 14825 | 8472 | 18627 |
| Al-Sulaimaniyah | Sharbazher | 4·905 | 3·124 | 11·766 | 2·738 | 6·207 | 2884 | 1603 | 3662 |
| An-Najaf | Al-Kufa | 4·402 | 4·599 | 16·680 | 1·594 | 5·452 | 20007 | 7267 | 24842 |
| An-Najaf | Al-Manathera | 3·820 | 4·025 | 14·277 | 1·403 | 4·743 | 14321 | 5232 | 17808 |
| An-Najaf | Al-Najaf | 2·900 | 3·081 | 10·937 | 1·059 | 3·580 | 42674 | 15291 | 51759 |
| Babil | Al-Hashimiya | 5·169 | 5·110 | 18·632 | 1·959 | 6·507 | 31386 | 11718 | 39345 |
| Babil | Al-Hilla | 4·869 | 5·004 | 18·228 | 1·791 | 6·065 | 51420 | 18730 | 64006 |
| Babil | Al-Hindiya | 4·773 | 4·916 | 17·792 | 1·724 | 5·963 | 19621 | 7045 | 24505 |
| Babil | Al-Mahaweel | 5·494 | 5·478 | 19·807 | 2·067 | 6·909 | 22231 | 8307 | 27947 |
| Babil | Al-Mahmoudiya | 5·635 | 5·461 | 19·835 | 2·172 | 7·129 | 46030 | 17994 | 58767 |
| Babil | Al-Mussyab | 5·357 | 5·400 | 19·474 | 1·984 | 6·759 | 25479 | 9404 | 32152 |
| Baghdad | Al-Adhamiya | 6·225 | 5·720 | 21·037 | 2·471 | 8·030 | 84231 | 33393 | 109990 |
| Baghdad | Al-Karkh | 5·831 | 5·508 | 19·636 | 2·277 | 7·428 | 88672 | 34595 | 112485 |
| Baghdad | Al-Risafa | 6·022 | 5·604 | 20·260 | 2·404 | 7·723 | 126798 | 50642 | 162888 |
| Baghdad | Al-Thawra | 5·971 | 5·306 | 19·408 | 2·490 | 7·802 | 106497 | 44373 | 138700 |
| Dhi Qar | Al-Chibayish | 5·151 | 3·458 | 12·958 | 2·760 | 6·541 | 6157 | 3270 | 7866 |
| Dhi Qar | Al-Nasiriya | 4·804 | 3·807 | 14·064 | 2·265 | 6·130 | 45908 | 21442 | 58358 |
| Dhi Qar | Al-Rifai | 5·377 | 4·729 | 17·491 | 2·289 | 6·871 | 29525 | 12418 | 37886 |
| Dhi Qar | Al-Shatra | 5·189 | 4·196 | 15·515 | 2·380 | 6·631 | 30017 | 13830 | 38114 |
| Dhi Qar | Suq Al-Shoyokh | 4·858 | 3·646 | 13·573 | 2·411 | 6·171 | 20078 | 9827 | 25386 |
| Diyala | Al-Khalis | 6·514 | 5·191 | 19·301 | 3·005 | 8·374 | 27967 | 12331 | 36053 |
| Diyala | Al-Mada'in | 6·015 | 5·528 | 20·210 | 2·434 | 7·640 | 34138 | 13777 | 43263 |
| Diyala | Al-Muqdadiya | 6·554 | 5·324 | 19·834 | 2·929 | 8·446 | 20865 | 9272 | 26977 |
| Diyala | Baladruz | 6·472 | 5·521 | 20·554 | 2·773 | 8·320 | 12005 | 5236 | 15553 |
| Diyala | Baquba | 6·356 | 5·579 | 20·722 | 2·677 | 8·133 | 49229 | 20762 | 63086 |
| Diyala | Khanaqin | 6·451 | 4·687 | 17·509 | 3·225 | 8·267 | 21251 | 10371 | 27228 |
| Diyala | Kifri | 6·585 | 4·638 | 17·440 | 3·374 | 8·444 | 10001 | 5190 | 12882 |
| Dohuk | Al-Amadiya | 4·136 | 2·694 | 10·097 | 2·269 | 5·228 | 6243 | 3421 | 7864 |
| Dohuk | Duhok | 4·084 | 2·627 | 9·916 | 2·277 | 5·156 | 19271 | 10686 | 24396 |
| Dohuk | Sumail | 4·001 | 2·667 | 10·023 | 2·173 | 5·070 | 10989 | 6052 | 13807 |

| ADM1 | ADM2 | β-thalassaemia carrier prevalence Mean | SD | CI95% | Q25 | Q75 | Estimated carrier counts Mean | Q25 | Q75 |
|---|---|---|---|---|---|---|---|---|---|
| Dohuk | Zakho | 3·571 | 2·382 | 8·844 | 1·952 | 4·493 | 11875 | 6546 | 15179 |
| Erbil | Al-Zibar | 4·436 | 2·871 | 10·864 | 2·451 | 5·626 | 1169 | 648 | 1481 |
| Erbil | Erbil | 6·088 | 3·871 | 14·501 | 3·403 | 7·722 | 93541 | 52754 | 118282 |
| Erbil | Koysinjaq | 5·557 | 3·420 | 12·847 | 3·178 | 7·022 | 7842 | 4502 | 9891 |
| Erbil | Makhmour | 6·340 | 4·338 | 16·301 | 3·344 | 8·090 | 13656 | 7278 | 17339 |
| Erbil | Rawanduz | 4·936 | 3·192 | 11·981 | 2·736 | 6·243 | 17730 | 9839 | 22506 |
| Erbil | Shaqlawa | 5·549 | 3·389 | 12·820 | 3·186 | 7·021 | 11387 | 6495 | 14412 |
| Karbala | Ain Al-Tamur | 4·146 | 4·328 | 15·333 | 1·497 | 5·193 | 1484 | 514 | 1829 |
| Karbala | Kerbela | 4·740 | 4·880 | 17·478 | 1·736 | 5·913 | 54179 | 19861 | 67732 |
| Kirkuk | Al-Hawiga | 6·603 | 4·783 | 17·931 | 3·315 | 8·450 | 23084 | 11615 | 29505 |
| Kirkuk | Daquq | 6·588 | 4·702 | 17·551 | 3·360 | 8·408 | 7608 | 3936 | 9733 |
| Kirkuk | Dibis | 6·380 | 4·170 | 15·768 | 3·480 | 8·107 | 8246 | 4495 | 10494 |
| Kirkuk | Kirkuk | 6·242 | 3·993 | 15·018 | 3·468 | 7·899 | 87156 | 47606 | 110364 |
| Maysan | Al-Amara | 5·411 | 4·524 | 16·701 | 2·390 | 6·951 | 4915 | 2194 | 6293 |
| Maysan | Ali Al-Gharbi | 5·643 | 4·852 | 17·842 | 2·419 | 7·245 | 3615 | 1565 | 4656 |
| Maysan | Al-Kahla | 5·184 | 4·070 | 15·079 | 2·431 | 6·651 | 41502 | 19560 | 53474 |
| Maysan | Al-Maimouna | 5·614 | 4·335 | 16·072 | 2·685 | 7·206 | 7609 | 3636 | 9724 |
| Maysan | Al-Mejar Al-Kabir | 5·306 | 3·748 | 14·043 | 2·738 | 6·772 | 12846 | 6529 | 16280 |
| Maysan | Qalat Saleh | 5·328 | 3·795 | 14·141 | 2·741 | 6·798 | 4130 | 2091 | 5261 |
| Ninawa | Al-Baaj | 4·893 | 4·036 | 14·801 | 2·196 | 6·290 | 11747 | 5426 | 15108 |
| Ninawa | Al-Hamdaniya | 5·725 | 3·815 | 14·297 | 3·083 | 7·266 | 29879 | 16080 | 38044 |
| Ninawa | Al-Hatra | 5·776 | 4·620 | 17·136 | 2·661 | 7·414 | 4542 | 2169 | 5793 |
| Ninawa | Al-Mosul | 5·603 | 3·972 | 14·839 | 2·887 | 7·145 | 120328 | 63423 | 153536 |
| Ninawa | Al-Shikhan | 4·806 | 3·080 | 11·523 | 2·684 | 6·078 | 8725 | 4872 | 11006 |
| Ninawa | Aqra | 5·156 | 3·278 | 12·333 | 2·886 | 6·524 | 19658 | 10929 | 24886 |
| Ninawa | Sinjar | 4·448 | 3·451 | 12·852 | 2·117 | 5·692 | 15453 | 7431 | 19797 |
| Ninawa | Telafar | 4·371 | 3·127 | 11·564 | 2·249 | 5·561 | 28115 | 14507 | 35434 |
| Ninawa | Tilkaef | 4·781 | 3·130 | 11·812 | 2·609 | 6·057 | 15239 | 8168 | 19401 |
| Salah al-Din | Al-Daur | 6·510 | 5·068 | 18·829 | 3·078 | 8·380 | 5448 | 2493 | 7123 |
| Salah al-Din | Al-Shirqat | 6·376 | 4·907 | 18·187 | 3·057 | 8·176 | 17996 | 8743 | 23128 |
| Salah al-Din | Balad | 6·212 | 5·437 | 19·951 | 2·619 | 7·971 | 33310 | 14278 | 42739 |
| Salah al-Din | Beygee | 6·298 | 5·221 | 19·357 | 2·800 | 8·065 | 19555 | 9074 | 25096 |
| Salah al-Din | Samarra | 6·221 | 5·254 | 19·433 | 2·704 | 8·038 | 19869 | 8905 | 26005 |
| Salah al-Din | Tikrit | 6·440 | 5·095 | 19·030 | 2·994 | 8·260 | 18477 | 8379 | 23714 |
| Salah al-Din | Tooz Khurmato | 6·562 | 4·531 | 16·830 | 3·446 | 8·373 | 16454 | 8705 | 21051 |
| Wasit | Al-Hai | 5·737 | 5·230 | 19·043 | 2·345 | 7·349 | 13098 | 5332 | 16635 |
| Wasit | Al-Kut | 5·903 | 5·264 | 19·454 | 2·425 | 7·595 | 37994 | 15210 | 47991 |
| Wasit | Al-Namaniya | 5·826 | 5·515 | 20·005 | 2·262 | 7·440 | 14595 | 5553 | 18611 |
| Wasit | Al-Suwaira | 5·884 | 5·507 | 20·117 | 2·323 | 7·504 | 31809 | 12619 | 40833 |
| Wasit | Badra | 6·227 | 5·584 | 20·580 | 2·509 | 8·036 | 2297 | 912 | 2986 |

ADM1: administrative level 1. ADM2: administrative level 2. SD: standard deviation. CI95%: 95% credible interval. Q25: 25th percentile. Q75: 75th percentile. SD and CI95% are measures of prediction uncertainty.

**Supplementary Table S25. Subnational areal predictions of β-thalassaemia carrier prevalence and estimated carrier count at administrative level 2 of Italy.**

| ADM1 | ADM2 | β-thalassaemia carrier prevalence Mean | SD | CI95% | Q25 | Q75 | Estimated carrier counts Mean | Q25 | Q75 |
|---|---|---|---|---|---|---|---|---|---|
| Abruzzo | Chieti | 7·112 | 7·675 | 27·990 | 2·232 | 9·070 | 23790 | 7338 | 30275 |
| Abruzzo | L'Aquila | 7·126 | 7·441 | 27·611 | 2·291 | 9·159 | 20508 | 6542 | 26418 |
| Abruzzo | Pescara | 6·889 | 7·473 | 27·087 | 2·154 | 8·837 | 21651 | 6590 | 27433 |
| Abruzzo | Teramo | 6·391 | 6·896 | 25·361 | 1·995 | 8·158 | 16324 | 5019 | 20835 |
| Basilicata | Matera | 8·385 | 8·874 | 32·901 | 2·786 | 10·577 | 16446 | 5585 | 20680 |
| Basilicata | Potenza | 8·359 | 8·988 | 33·197 | 2·661 | 10·583 | 27952 | 8822 | 35389 |
| Calabria | Catanzaro | 7·894 | 8·670 | 32·473 | 2·464 | 9·911 | 25617 | 8007 | 31904 |
| Calabria | Cosenza | 8·349 | 9·030 | 33·859 | 2·634 | 10·510 | 48441 | 15109 | 61230 |
| Calabria | Crotone | 8·156 | 8·753 | 32·757 | 2·587 | 10·326 | 11415 | 3640 | 14521 |
| Calabria | Reggio di Calabria | 7·068 | 7·637 | 28·377 | 2·276 | 8·908 | 29611 | 9562 | 36810 |

| | | β-thalassaemia carrier prevalence | | | | | Estimated carrier counts | | |
|---|---|---|---|---|---|---|---|---|---|
| ADM1 | ADM2 | Mean | SD | CI95% | Q25 | Q75 | Mean | Q25 | Q75 |
| Calabria | Vibo Valentia | 7·487 | 8·194 | 30·483 | 2·373 | 9·449 | 10146 | 3164 | 12870 |
| Campania | Avellino | 8·196 | 8·830 | 32·493 | 2·606 | 10·401 | 32573 | 10413 | 41039 |
| Campania | Benevento | 8·088 | 8·673 | 31·794 | 2·588 | 10·283 | 19657 | 6307 | 25053 |
| Campania | Caserta | 8·056 | 8·444 | 31·286 | 2·616 | 10·295 | 72623 | 23512 | 92789 |
| Campania | Napoli | 8·367 | 8·927 | 32·908 | 2·656 | 10·720 | 232668 | 73215 | 297560 |
| Campania | Salerno | 8·441 | 9·188 | 34·130 | 2·633 | 10·693 | 87002 | 27528 | 110267 |
| Emilia-Romagna | Bologna | 3·285 | 2·903 | 10·466 | 1·441 | 4·134 | 33018 | 14652 | 41229 |
| Emilia-Romagna | Ferrara | 3·232 | 3·069 | 10·961 | 1·316 | 4·066 | 10338 | 4223 | 13014 |
| Emilia-Romagna | Forli'-Cesena | 3·832 | 3·689 | 13·156 | 1·541 | 4·818 | 14388 | 5770 | 18222 |
| Emilia-Romagna | Modena | 3·185 | 2·732 | 9·938 | 1·437 | 4·009 | 22777 | 10321 | 28611 |
| Emilia-Romagna | Parma | 3·389 | 3·082 | 11·180 | 1·433 | 4·264 | 14657 | 6231 | 18480 |
| Emilia-Romagna | Piacenza | 3·500 | 3·392 | 12·183 | 1·376 | 4·395 | 9973 | 3877 | 12657 |
| Emilia-Romagna | Ravenna | 3·491 | 3·305 | 11·809 | 1·428 | 4·394 | 13003 | 5301 | 16411 |
| Emilia-Romagna | Reggio nell'Emilia | 3·211 | 2·784 | 10·095 | 1·429 | 4·048 | 15355 | 6961 | 19395 |
| Emilia-Romagna | Rimini | 4·124 | 4·205 | 15·129 | 1·554 | 5·156 | 11505 | 4230 | 14296 |
| Friuli Venezia Giulia | Gorizia | 4·161 | 5·500 | 17·963 | 1·149 | 4·949 | 5268 | 1435 | 6180 |
| Friuli Venezia Giulia | Pordenone | 3·741 | 4·613 | 15·637 | 1·107 | 4·561 | 11276 | 3408 | 13765 |
| Friuli Venezia Giulia | Trieste | 4·297 | 5·551 | 18·854 | 1·208 | 5·242 | 7326 | 2048 | 8721 |
| Friuli Venezia Giulia | Udine | 3·962 | 5·074 | 17·007 | 1·107 | 4·809 | 20369 | 5859 | 24622 |
| Lazio | Frosinone | 7·803 | 8·021 | 30·030 | 2·557 | 10·037 | 36540 | 12070 | 47208 |
| Lazio | Latina | 8·130 | 8·148 | 30·677 | 2·717 | 10·514 | 42453 | 14234 | 55097 |
| Lazio | Rieti | 6·773 | 7·019 | 26·128 | 2·240 | 8·672 | 9899 | 3309 | 12682 |
| Lazio | Roma | 7·587 | 7·601 | 28·445 | 2·605 | 9·761 | 316418 | 109604 | 407768 |
| Lazio | Viterbo | 6·714 | 6·752 | 25·060 | 2·367 | 8·551 | 21130 | 7356 | 26945 |
| Liguria | Genova | 4·016 | 3·973 | 14·389 | 1·519 | 5·061 | 20970 | 7726 | 26438 |
| Liguria | Imperia | 5·399 | 5·971 | 20·931 | 1·787 | 6·743 | 8357 | 2776 | 10489 |
| Liguria | La Spezia | 3·851 | 3·559 | 12·905 | 1·583 | 4·868 | 6754 | 2821 | 8582 |
| Liguria | Savona | 4·746 | 5·182 | 18·170 | 1·597 | 5·972 | 9041 | 3030 | 11506 |
| Lombardia | Bergamo | 3·484 | 3·888 | 13·661 | 1·173 | 4·295 | 37923 | 13092 | 47003 |
| Lombardia | Brescia | 3·328 | 3·516 | 12·415 | 1·189 | 4·158 | 41208 | 15062 | 51646 |
| Lombardia | Como | 3·736 | 4·410 | 15·420 | 1·165 | 4·571 | 22116 | 7090 | 27221 |
| Lombardia | Cremona | 3·302 | 3·241 | 11·704 | 1·288 | 4·148 | 12057 | 4608 | 15159 |
| Lombardia | Lecco | 3·642 | 4·196 | 14·943 | 1·178 | 4·470 | 12449 | 4105 | 15291 |
| Lombardia | Lodi | 3·460 | 3·567 | 12·929 | 1·280 | 4·310 | 8093 | 2950 | 10039 |
| Lombardia | Mantova | 3·107 | 2·915 | 10·360 | 1·280 | 3·916 | 12882 | 5303 | 16204 |
| Lombardia | Milano | 3·729 | 4·155 | 14·946 | 1·238 | 4·591 | 118747 | 39689 | 145446 |
| Lombardia | Monza e della Brianza | 3·676 | 4·186 | 14·686 | 1·222 | 4·508 | 33163 | 11066 | 40731 |
| Lombardia | Pavia | 3·790 | 4·043 | 14·542 | 1·325 | 4·688 | 20413 | 7094 | 25331 |
| Lombardia | Sondrio | 3·590 | 4·250 | 14·963 | 1·097 | 4·386 | 6381 | 1973 | 7773 |
| Lombardia | Varese | 3·887 | 4·655 | 16·190 | 1·196 | 4·754 | 32251 | 10095 | 39434 |
| Marche | Ancona | 5·004 | 5·505 | 19·726 | 1·648 | 6·281 | 19618 | 6395 | 24482 |
| Marche | Ascoli Piceno | 6·007 | 6·477 | 23·571 | 1·887 | 7·711 | 10783 | 3334 | 13879 |
| Marche | Fermo | 5·761 | 6·360 | 23·250 | 1·797 | 7·239 | 8934 | 2789 | 11233 |
| Marche | Macerata | 5·556 | 6·123 | 22·202 | 1·779 | 6·989 | 13972 | 4418 | 17609 |
| Marche | Pesaro e Urbino | 4·502 | 4·789 | 17·036 | 1·591 | 5·626 | 11329 | 3986 | 14141 |
| Molise | Campobasso | 7·570 | 8·093 | 29·257 | 2·438 | 9·668 | 15490 | 4953 | 19870 |
| Molise | Isernia | 7·598 | 8·089 | 29·778 | 2·449 | 9·637 | 6077 | 1955 | 7642 |
| Piemonte | Alessandria | 4·125 | 4·457 | 15·817 | 1·422 | 5·147 | 16925 | 5789 | 21229 |
| Piemonte | Asti | 4·436 | 5·064 | 17·654 | 1·430 | 5·505 | 8639 | 2782 | 10805 |
| Piemonte | Biella | 4·239 | 5·202 | 17·925 | 1·257 | 5·130 | 7312 | 2188 | 8862 |
| Piemonte | Cuneo | 5·024 | 5·824 | 20·141 | 1·572 | 6·240 | 28909 | 9141 | 35817 |
| Piemonte | Novara | 4·005 | 4·737 | 16·506 | 1·246 | 4·897 | 14505 | 4582 | 17778 |
| Piemonte | Torino | 4·736 | 5·783 | 20·361 | 1·371 | 5·769 | 103552 | 30576 | 126602 |
| Piemonte | Verbano-Cusio-Ossola | 4·140 | 5·375 | 18·654 | 1·153 | 4·909 | 5264 | 1485 | 6358 |
| Piemonte | Vercelli | 4·219 | 5·044 | 17·740 | 1·265 | 5·162 | 6975 | 2138 | 8619 |
| Puglia | Bari | 8·333 | 8·518 | 31·279 | 2·874 | 10·645 | 90213 | 30993 | 115385 |
| Puglia | Barletta-Andria-Trani | 8·162 | 8·560 | 30·865 | 2·722 | 10·488 | 23681 | 7813 | 29971 |

| ADM1 | ADM2 | β-thalassaemia carrier prevalence Mean | SD | CI95% | Q25 | Q75 | Estimated carrier counts Mean | Q25 | Q75 |
|---|---|---|---|---|---|---|---|---|---|
| Puglia | Brindisi | 8·806 | 8·661 | 32·211 | 3·153 | 11·265 | 25325 | 9037 | 32445 |
| Puglia | Foggia | 7·899 | 8·409 | 30·068 | 2·536 | 10·155 | 44227 | 14329 | 57055 |
| Puglia | Lecce | 8·949 | 8·506 | 31·886 | 3·326 | 11·522 | 65877 | 24406 | 84913 |
| Puglia | Taranto | 8·600 | 8·736 | 32·797 | 2·988 | 10·901 | 43534 | 15061 | 55126 |
| Sardegna | Cagliari | 16·226 | 9·288 | 35·359 | 9·395 | 21·001 | 83437 | 48495 | 107703 |
| Sardegna | Carbonia-Iglesias | 16·743 | 9·972 | 37·720 | 9·343 | 21·905 | 17791 | 9932 | 23239 |
| Sardegna | Medio Campidano | 16·534 | 9·205 | 35·140 | 9·727 | 21·344 | 14863 | 8731 | 19137 |
| Sardegna | Nuoro | 14·175 | 8·120 | 30·888 | 8·252 | 18·225 | 19926 | 11569 | 25685 |
| Sardegna | Ogliastra | 14·949 | 8·576 | 32·729 | 8·677 | 19·286 | 7232 | 4147 | 9356 |
| Sardegna | Olbia-Tempio | 12·334 | 8·011 | 30·336 | 6·589 | 15·956 | 14881 | 7785 | 19373 |
| Sardegna | Oristano | 15·891 | 8·734 | 33·222 | 9·486 | 20·384 | 24119 | 14449 | 30892 |
| Sardegna | Sassari | 13·404 | 7·973 | 30·279 | 7·639 | 17·220 | 35908 | 20240 | 46034 |
| Sicilia | Agrigento | 6·930 | 6·759 | 25·121 | 2·498 | 8·882 | 24205 | 8777 | 30960 |
| Sicilia | Caltanissetta | 6·304 | 6·069 | 22·259 | 2·361 | 8·076 | 14942 | 5583 | 19172 |
| Sicilia | Catania | 6·141 | 6·139 | 22·363 | 2·228 | 7·809 | 61533 | 22159 | 78006 |
| Sicilia | Enna | 6·403 | 6·385 | 23·598 | 2·318 | 8·154 | 10155 | 3682 | 12930 |
| Sicilia | Messina | 6·989 | 7·400 | 27·338 | 2·323 | 8·818 | 31962 | 10431 | 40665 |
| Sicilia | Palermo | 7·568 | 7·513 | 27·929 | 2·626 | 9·766 | 89710 | 30872 | 115618 |
| Sicilia | Ragusa | 5·336 | 5·049 | 18·380 | 2·097 | 6·801 | 16865 | 6642 | 21479 |
| Sicilia | Siracusa | 5·468 | 5·405 | 19·533 | 2·061 | 6·933 | 18943 | 7111 | 23751 |
| Sicilia | Trapani | 8·450 | 8·196 | 30·483 | 2·925 | 11·011 | 30192 | 10403 | 39021 |
| Toscana | Arezzo | 4·427 | 4·310 | 15·434 | 1·721 | 5·598 | 15017 | 5838 | 19053 |
| Toscana | Firenze | 3·925 | 3·613 | 13·057 | 1·642 | 4·953 | 39302 | 16508 | 49630 |
| Toscana | Grosseto | 5·834 | 5·584 | 20·337 | 2·254 | 7·430 | 11276 | 4411 | 14336 |
| Toscana | Livorno | 5·244 | 4·834 | 17·501 | 2·094 | 6·751 | 11641 | 4650 | 15124 |
| Toscana | Lucca | 3·786 | 3·344 | 12·143 | 1·623 | 4·805 | 12468 | 5340 | 15778 |
| Toscana | Massa Carrara | 3·658 | 3·282 | 12·056 | 1·554 | 4·593 | 5799 | 2439 | 7255 |
| Toscana | Pisa | 4·440 | 4·042 | 14·769 | 1·836 | 5·644 | 17503 | 7268 | 22243 |
| Toscana | Pistoia | 3·709 | 3·246 | 11·844 | 1·599 | 4·709 | 11604 | 4970 | 14843 |
| Toscana | Prato | 3·736 | 3·297 | 12·103 | 1·577 | 4·736 | 9374 | 3960 | 11878 |
| Toscana | Siena | 4·960 | 4·785 | 17·460 | 1·924 | 6·278 | 12429 | 4876 | 15730 |
| Trentino-Alto Adige | Bolzano | 3·624 | 4·467 | 15·661 | 1·017 | 4·451 | 19069 | 5513 | 23428 |
| Trentino-Alto Adige | Trento | 3·380 | 3·841 | 13·293 | 1·115 | 4·184 | 18268 | 6204 | 22520 |
| Umbria | Perugia | 5·455 | 5·692 | 20·623 | 1·875 | 6·898 | 34804 | 12048 | 44109 |
| Umbria | Terni | 6·164 | 6·297 | 23·089 | 2·134 | 7·868 | 13694 | 4676 | 17308 |
| Valle D'Aosta | Aosta | 4·578 | 5·855 | 20·465 | 1·236 | 5·495 | 5561 | 1514 | 6647 |
| Veneto | Belluno | 3·614 | 4·395 | 15·185 | 1·067 | 4·449 | 7077 | 2151 | 8700 |
| Veneto | Padova | 3·236 | 3·306 | 11·599 | 1·227 | 4·064 | 28683 | 10784 | 36058 |
| Veneto | Rovigo | 3·218 | 3·179 | 11·145 | 1·268 | 4·035 | 7103 | 2815 | 8891 |
| Veneto | Treviso | 3·462 | 3·953 | 13·480 | 1·154 | 4·292 | 31404 | 10632 | 39219 |
| Veneto | Venezia | 3·519 | 3·960 | 13·508 | 1·185 | 4·382 | 25716 | 8929 | 32074 |
| Veneto | Verona | 3·156 | 3·097 | 10·990 | 1·238 | 3·979 | 29344 | 11571 | 36892 |
| Veneto | Vicenza | 3·241 | 3·376 | 11·825 | 1·185 | 4·079 | 27432 | 10143 | 34391 |

ADM1: administrative level 1. ADM2: administrative level 2. SD: standard deviation. CI95%: 95% credible interval. Q25: 25th percentile. Q75: 75th percentile. SD and CI95% are measures of prediction uncertainty.

**Supplementary Table S26. Subnational areal predictions of β-thalassaemia carrier prevalence and estimated carrier count at administrative level 2 of Jordan.**

| ADM1 | ADM2 | β-thalassaemia carrier prevalence Mean | SD | CI95% | Q25 | Q75 | Estimated carrier counts Mean | Q25 | Q75 |
|---|---|---|---|---|---|---|---|---|---|
| Ajloun | Aljun | 3·681 | 2·501 | 9·305 | 1·990 | 4·645 | 5421 | 2937 | 6825 |
| Ajloun | Kofranjah | 3·596 | 2·469 | 8·915 | 1·939 | 4·565 | 453 | 245 | 567 |
| Amman | Al-Jizah | 2·056 | 1·565 | 5·743 | 1·025 | 2·597 | 3826 | 1947 | 4831 |
| Amman | Al-Muwaqqar | 2·360 | 1·713 | 6·353 | 1·214 | 2·985 | 6003 | 3151 | 7526 |
| Amman | Amman | 2·968 | 2·038 | 7·702 | 1·568 | 3·747 | 45017 | 23230 | 57148 |
| Amman | Na'ur | 2·802 | 2·021 | 7·241 | 1·447 | 3·576 | 18209 | 9369 | 23266 |

| | | β-thalassaemia carrier prevalence | | | | | Estimated carrier counts | | |
|---|---|---|---|---|---|---|---|---|---|
| **ADM1** | **ADM2** | **Mean** | **SD** | **CI95%** | **Q25** | **Q75** | **Mean** | **Q25** | **Q75** |
| Amman | Sahab | 2·557 | 1·810 | 6·710 | 1·348 | 3·242 | 25738 | 13548 | 32921 |
| Amman | Um al-Basatin | 2·953 | 2·085 | 7·648 | 1·560 | 3·729 | 5575 | 2972 | 7024 |
| Amman | Wadi al-Sayr | 3·069 | 2·121 | 7·871 | 1·622 | 3·897 | 41471 | 21775 | 52653 |
| Aqaba | Al-Aqaba | 1·607 | 1·620 | 5·731 | 0·623 | 2·003 | 2883 | 1089 | 3516 |
| Aqaba | Al-Quwayra | 1·412 | 1·407 | 4·948 | 0·563 | 1·755 | 480 | 194 | 605 |
| Aqaba | Wadi Araba | 2·024 | 1·904 | 6·749 | 0·846 | 2·539 | 223 | 89 | 284 |
| Balqa | Al-Balqa | 3·435 | 2·273 | 8·595 | 1·834 | 4·341 | 353 | 188 | 446 |
| Balqa | Ardhah | 3·311 | 2·245 | 8·417 | 1·786 | 4·184 | 3648 | 1946 | 4513 |
| Balqa | As-Salt | 3·232 | 2·198 | 8·084 | 1·761 | 4·078 | 4966 | 2702 | 6216 |
| Balqa | Dair Alla | 3·640 | 2·422 | 9·255 | 1·983 | 4·597 | 2233 | 1209 | 2808 |
| Balqa | Shuna al-Jabiniyya | 3·282 | 2·298 | 8·578 | 1·737 | 4·166 | 2711 | 1431 | 3451 |
| Irbid | Al-Aghwar Shamaliyyeh | 3·418 | 2·245 | 8·429 | 1·882 | 4·317 | 9478 | 5194 | 12095 |
| Irbid | Ar Ramtha | 3·379 | 2·350 | 8·777 | 1·793 | 4·232 | 8687 | 4617 | 10976 |
| Irbid | Bani Knana | 3·629 | 2·380 | 9·457 | 1·980 | 4·643 | 1917 | 1060 | 2452 |
| Irbid | Hariema | 3·809 | 2·575 | 9·678 | 2·047 | 4·810 | 10267 | 5495 | 12949 |
| Irbid | Irbid | 3·981 | 2·728 | 10·259 | 2·160 | 4·962 | 4463 | 2395 | 5536 |
| Irbid | Kora | 3·883 | 2·734 | 10·243 | 2·075 | 4·835 | 311 | 166 | 387 |
| Irbid | Mazar Shamaliyyeh | 3·819 | 2·629 | 10·107 | 2·062 | 4·774 | 8214 | 4409 | 10316 |
| Irbid | Tayybeh | 3·927 | 2·684 | 9·974 | 2·122 | 4·937 | 12997 | 7014 | 16319 |
| Irbid | Wastiyyeh | 3·698 | 2·553 | 9·414 | 1·981 | 4·629 | 30007 | 16056 | 37498 |
| Jerash | Jerash | 3·351 | 2·260 | 8·359 | 1·827 | 4·232 | 12281 | 6745 | 15446 |
| Karak | Al-Karak | 2·298 | 1·822 | 6·320 | 1·123 | 2·836 | 158 | 77 | 195 |
| Karak | Al-Mazar al-Janubiyya | 2·076 | 1·743 | 6·230 | 0·969 | 2·601 | 2785 | 1316 | 3472 |
| Karak | Al-Mazra'a | 2·638 | 2·135 | 7·702 | 1·262 | 3·309 | 1292 | 622 | 1609 |
| Karak | Al-Qasr | 2·048 | 1·644 | 6·012 | 0·978 | 2·595 | 3126 | 1482 | 3986 |
| Karak | Al-Safi | 2·421 | 1·717 | 6·455 | 1·272 | 3·038 | 29 | 16 | 37 |
| Karak | Ayy | 2·465 | 2·073 | 7·562 | 1·122 | 3·114 | 993 | 448 | 1258 |
| Karak | Faqqu | 2·472 | 1·851 | 6·976 | 1·225 | 3·164 | 367 | 181 | 467 |
| Ma'an | Al-Husanyniyya | 1·681 | 1·520 | 5·449 | 0·730 | 2·110 | 370 | 160 | 463 |
| Ma'an | Ash-Shibek | 1·879 | 1·712 | 6·174 | 0·818 | 2·358 | 421 | 182 | 527 |
| Ma'an | Ayi | 1·573 | 1·518 | 5·325 | 0·649 | 1·975 | 297 | 127 | 380 |
| Ma'an | Ma'an | 1·178 | 1·103 | 3·889 | 0·500 | 1·466 | 996 | 409 | 1247 |
| Ma'an | Wadi Musa | 1·743 | 1·592 | 5·657 | 0·740 | 2·219 | 753 | 323 | 953 |
| Madaba | Dhiban | 2·602 | 1·940 | 7·137 | 1·316 | 3·278 | 1004 | 508 | 1273 |
| Madaba | Madaba | 2·861 | 2·096 | 7·688 | 1·454 | 3·633 | 5248 | 2689 | 6731 |
| Mafraq | Al-Mafraq | 2·813 | 1·921 | 7·222 | 1·514 | 3·549 | 10979 | 5983 | 13884 |
| Mafraq | Ar-Ruwayshid | 1·989 | 1·870 | 6·577 | 0·862 | 2·464 | 725 | 348 | 905 |
| Mafraq | Bal'ama | 3·046 | 2·068 | 7·679 | 1·654 | 3·827 | 3656 | 1981 | 4575 |
| Mafraq | Sabha | 2·326 | 1·780 | 6·459 | 1·164 | 2·926 | 1262 | 644 | 1595 |
| Mafraq | Sama as-Sarhan | 3·108 | 2·158 | 7·995 | 1·655 | 3·913 | 2967 | 1578 | 3719 |
| Tafilah | Al-Hasa | 1·756 | 1·541 | 5·543 | 0·781 | 2·214 | 212 | 93 | 266 |
| Tafilah | Al-Tafila | 2·154 | 1·861 | 6·723 | 0·971 | 2·709 | 1854 | 843 | 2303 |
| Tafilah | Birsayra | 1·902 | 1·727 | 6·074 | 0·846 | 2·358 | 311 | 136 | 383 |
| Zarqa | Al-Azraq | 1·751 | 1·328 | 4·848 | 0·880 | 2·206 | 987 | 499 | 1236 |
| Zarqa | Az-Zarqa | 2·746 | 1·942 | 7·058 | 1·457 | 3·448 | 42166 | 22233 | 52983 |
| Zarqa | Birin | 3·140 | 2·178 | 8·243 | 1·686 | 3·957 | 3010 | 1604 | 3785 |

ADM1: administrative level 1. ADM2: administrative level 2. SD: standard deviation. CI95%: 95% credible interval. Q25: 25th percentile. Q75: 75th percentile. SD and CI95% are measures of prediction uncertainty.

**Supplementary Table S27. Subnational areal predictions of β-thalassaemia carrier prevalence and estimated carrier count at administrative level 2 of Malaysia.**

| | | β-thalassaemia carrier prevalence | | | | | Estimated carrier counts | | |
|---|---|---|---|---|---|---|---|---|---|
| **ADM1** | **ADM2** | **Mean** | **SD** | **CI95%** | **Q25** | **Q75** | **Mean** | **Q25** | **Q75** |
| Johor | Batu Pahat | 4·732 | 5·697 | 18·931 | 1·433 | 5·901 | 25540 | 7690 | 31894 |
| Johor | Johor Bahru | 5·144 | 6·652 | 22·893 | 1·342 | 6·296 | 92150 | 24144 | 113416 |
| Johor | Kluang | 4·888 | 6·127 | 20·276 | 1·415 | 6·033 | 16781 | 4928 | 20785 |

| | | β-thalassaemia carrier prevalence | | | | | Estimated carrier counts | | |
|---|---|---|---|---|---|---|---|---|---|
| ADM1 | ADM2 | Mean | SD | CI95% | Q25 | Q75 | Mean | Q25 | Q75 |
| Johor | Kota Tinggi | 5·207 | 6·742 | 23·468 | 1·343 | 6·379 | 12207 | 3112 | 15043 |
| Johor | Kulai Jaya | 5·001 | 6·485 | 21·668 | 1·354 | 6·087 | 15691 | 4257 | 19236 |
| Johor | Ledang | 4·521 | 4·837 | 16·731 | 1·561 | 5·737 | 10068 | 3409 | 12702 |
| Johor | Mersing | 4·942 | 6·214 | 20·835 | 1·402 | 6·090 | 3159 | 896 | 3958 |
| Johor | Muar | 4·570 | 5·150 | 17·636 | 1·504 | 5·706 | 13020 | 4372 | 16162 |
| Johor | Pontian | 4·950 | 6·267 | 21·017 | 1·354 | 6·106 | 10081 | 2723 | 12408 |
| Johor | Segamat | 4·535 | 5·059 | 17·175 | 1·530 | 5·685 | 9654 | 3304 | 12199 |
| Johor | Tioman Island | 4·952 | 6·342 | 21·785 | 1·346 | 5·993 | 85 | 23 | 102 |
| Kedah | Baling | 5·404 | 4·853 | 17·820 | 2·186 | 6·928 | 7181 | 2878 | 9261 |
| Kedah | Bandar Baharu | 5·375 | 5·090 | 18·189 | 2·094 | 6·836 | 2280 | 883 | 2892 |
| Kedah | Kota Setar | 5·656 | 4·837 | 17·667 | 2·415 | 7·249 | 20696 | 8754 | 26669 |
| Kedah | Kuala Muda | 5·528 | 5·108 | 18·729 | 2·217 | 7·061 | 34377 | 13768 | 43818 |
| Kedah | Kubang Pasu | 5·673 | 4·747 | 17·623 | 2·483 | 7·313 | 17874 | 7824 | 23029 |
| Kedah | Kulim | 5·426 | 5·039 | 18·830 | 2·162 | 6·936 | 16703 | 6652 | 21250 |
| Kedah | Langkawi | 5·932 | 5·488 | 20·446 | 2·343 | 7·575 | 4738 | 1877 | 6086 |
| Kedah | Padang Terap | 5·512 | 4·624 | 17·203 | 2·426 | 7·083 | 4098 | 1798 | 5246 |
| Kedah | Pendang | 5·553 | 4·860 | 17·692 | 2·361 | 7·135 | 6989 | 2994 | 8975 |
| Kedah | Pokok Sena | 5·505 | 4·720 | 17·619 | 2·400 | 7·054 | 2849 | 1233 | 3657 |
| Kedah | Sik | 5·430 | 4·775 | 17·685 | 2·285 | 6·969 | 4649 | 1945 | 5988 |
| Kedah | Yan | 5·619 | 5·054 | 18·860 | 2·296 | 7·178 | 4724 | 1941 | 5992 |
| Kelantan | Bachok | 5·024 | 5·331 | 19·661 | 1·715 | 6·296 | 7285 | 2476 | 9076 |
| Kelantan | Gua Musang | 4·968 | 4·955 | 18·063 | 1·816 | 6·310 | 6304 | 2242 | 7979 |
| Kelantan | Jeli | 5·138 | 5·038 | 18·478 | 1·904 | 6·538 | 3004 | 1111 | 3813 |
| Kelantan | Kota Bharu | 5·034 | 5·135 | 18·769 | 1·756 | 6·439 | 30747 | 10779 | 39566 |
| Kelantan | Kuala Krai | 5·103 | 5·256 | 19·138 | 1·796 | 6·462 | 5956 | 2060 | 7614 |
| Kelantan | Machang | 5·067 | 5·194 | 19·081 | 1·799 | 6·418 | 6114 | 2144 | 7767 |
| Kelantan | Pasir Mas | 5·067 | 5·082 | 18·912 | 1·824 | 6·431 | 13325 | 4801 | 16923 |
| Kelantan | Pasir Puteh | 5·008 | 5·154 | 18·696 | 1·717 | 6·413 | 7603 | 2596 | 9755 |
| Kelantan | Tanah Merah | 5·118 | 5·186 | 18·838 | 1·847 | 6·424 | 9078 | 3294 | 11466 |
| Kelantan | Tumpat | 5·056 | 5·071 | 19·172 | 1·774 | 6·441 | 8366 | 2924 | 10666 |
| Kuala Lumpur | Kuala Lumpur | 4·102 | 3·441 | 12·415 | 1·843 | 5·207 | 94664 | 42591 | 120182 |
| Labuan | Labuan | 11·722 | 9·159 | 33·989 | 5·279 | 15·418 | 5410 | 2437 | 7085 |
| Malacca | Alor Gajah | 4·226 | 4·164 | 14·767 | 1·637 | 5·323 | 12787 | 4951 | 16077 |
| Malacca | Central Melaka | 4·341 | 4·498 | 15·587 | 1·612 | 5·447 | 25415 | 9450 | 32026 |
| Malacca | Jasin | 4·388 | 4·514 | 15·980 | 1·610 | 5·531 | 5405 | 1950 | 6858 |
| Negeri Sembilan | Jelebu | 4·104 | 3·605 | 13·232 | 1·751 | 5·234 | 2409 | 1024 | 3079 |
| Negeri Sembilan | Jempol | 4·285 | 4·225 | 15·164 | 1·635 | 5·397 | 5288 | 2023 | 6693 |
| Negeri Sembilan | Kuala Pilah | 4·168 | 3·887 | 13·833 | 1·666 | 5·334 | 3482 | 1389 | 4443 |
| Negeri Sembilan | Port Dickson | 4·185 | 3·817 | 13·720 | 1·700 | 5·376 | 5873 | 2383 | 7444 |
| Negeri Sembilan | Rembau | 4·160 | 3·895 | 13·844 | 1·673 | 5·342 | 2326 | 935 | 2974 |
| Negeri Sembilan | Seremban | 4·083 | 3·581 | 13·011 | 1·746 | 5·217 | 33963 | 14477 | 43025 |
| Negeri Sembilan | Tampin | 4·312 | 4·313 | 15·045 | 1·627 | 5·463 | 3046 | 1154 | 3858 |
| Pahang | Bentong | 4·192 | 3·675 | 13·484 | 1·789 | 5·350 | 5201 | 2226 | 6718 |
| Pahang | Bera | 4·324 | 4·236 | 15·417 | 1·661 | 5·427 | 4538 | 1783 | 5726 |
| Pahang | Cameron Highlands | 4·748 | 4·446 | 16·474 | 1·863 | 6·036 | 1627 | 624 | 2096 |
| Pahang | Jerantut | 4·620 | 4·537 | 16·618 | 1·673 | 5·917 | 4876 | 1855 | 6227 |
| Pahang | Kuantan | 4·692 | 4·941 | 17·955 | 1·551 | 5·988 | 26186 | 8252 | 33350 |
| Pahang | Lipis | 4·643 | 4·397 | 16·138 | 1·806 | 5·916 | 4647 | 1779 | 5906 |
| Pahang | Maran | 4·483 | 4·362 | 15·951 | 1·668 | 5·732 | 5114 | 1930 | 6542 |
| Pahang | Pekan | 4·639 | 5·045 | 18·150 | 1·527 | 5·808 | 6172 | 1973 | 7655 |
| Pahang | Raub | 4·417 | 4·010 | 14·600 | 1·812 | 5·617 | 5036 | 2086 | 6387 |
| Pahang | Rompin | 4·548 | 4·998 | 17·486 | 1·537 | 5·674 | 4592 | 1536 | 5704 |
| Pahang | Temerloh | 4·359 | 4·006 | 14·813 | 1·743 | 5·570 | 9046 | 3566 | 11615 |
| Penang | Central Seberang Perai | 5·672 | 5·459 | 20·145 | 2·182 | 7·252 | 28159 | 10730 | 35852 |
| Penang | North Seberang Perai | 5·486 | 5·082 | 18·558 | 2·153 | 7·012 | 15952 | 6273 | 20432 |
| Penang | Northeast Penang Island | 5·569 | 5·280 | 19·006 | 2·172 | 7·100 | 21503 | 8369 | 27751 |
| Penang | South Seberang Perai | 5·487 | 5·274 | 19·088 | 2·121 | 6·929 | 13111 | 5044 | 16548 |

| | | β-thalassaemia carrier prevalence | | | | | Estimated carrier counts | | |
|---|---|---|---|---|---|---|---|---|---|
| ADM1 | ADM2 | Mean | SD | CI95% | Q25 | Q75 | Mean | Q25 | Q75 |
| Penang | Southwest Penang Island | 5·725 | 5·531 | 19·921 | 2·118 | 7·373 | 15748 | 5864 | 19891 |
| Perak | Bagan Datuk | 4·694 | 4·404 | 16·257 | 1·865 | 5·990 | 3508 | 1358 | 4457 |
| Perak | Batang Padang | 4·635 | 4·280 | 15·690 | 1·883 | 5·890 | 6558 | 2634 | 8276 |
| Perak | Hilir Perak | 4·673 | 4·374 | 16·089 | 1·865 | 5·918 | 6830 | 2690 | 8637 |
| Perak | Hulu Perak | 5·229 | 4·976 | 18·366 | 2·019 | 6·662 | 5214 | 2043 | 6654 |
| Perak | Kampar | 4·842 | 4·600 | 17·172 | 1·889 | 6·111 | 5395 | 2151 | 6810 |
| Perak | Kerian | 5·357 | 5·172 | 18·888 | 2·060 | 6·820 | 9057 | 3456 | 11588 |
| Perak | Kinta | 4·997 | 4·894 | 17·927 | 1·883 | 6·334 | 49374 | 18461 | 62935 |
| Perak | Kuala Kangsar | 5·092 | 4·922 | 18·058 | 1·943 | 6·462 | 9881 | 3821 | 12529 |
| Perak | Larut and Selama | 5·217 | 4·973 | 18·200 | 2·033 | 6·644 | 17165 | 6645 | 21819 |
| Perak | Manjung | 5·039 | 4·990 | 18·210 | 1·881 | 6·359 | 11272 | 4206 | 14449 |
| Perak | Muallim | 4·481 | 4·103 | 15·141 | 1·833 | 5·685 | 2447 | 1006 | 3123 |
| Perak | Perak Tengah | 4·909 | 4·782 | 17·500 | 1·883 | 6·184 | 5758 | 2199 | 7252 |
| Perlis | Perlis | 5·711 | 4·814 | 17·705 | 2·509 | 7·293 | 18210 | 7928 | 23406 |
| Putrajaya | Putrajaya | 3·942 | 3·162 | 11·539 | 1·805 | 5·094 | 3432 | 1572 | 4435 |
| Sabah | Beaufort | 12·412 | 8·848 | 33·498 | 6·090 | 16·259 | 10220 | 5057 | 13363 |
| Sabah | Beluran | 13·410 | 8·806 | 33·286 | 7·082 | 17·447 | 17534 | 9325 | 22813 |
| Sabah | Keningau | 12·967 | 8·340 | 31·786 | 7·001 | 16·792 | 21371 | 11610 | 27559 |
| Sabah | Kinabatangan | 12·772 | 8·928 | 33·579 | 6·398 | 16·630 | 21664 | 10999 | 28165 |
| Sabah | Kota Belud | 12·986 | 8·434 | 32·098 | 6·902 | 16·906 | 15352 | 8037 | 20139 |
| Sabah | Kota Kinabalu | 12·914 | 8·615 | 33·041 | 6·669 | 16·824 | 53004 | 27552 | 68939 |
| Sabah | Kota Marudu | 13·039 | 8·430 | 32·196 | 6·957 | 16·943 | 10152 | 5427 | 13236 |
| Sabah | Kuala Penyu | 12·365 | 9·318 | 35·423 | 5·737 | 16·177 | 2295 | 1063 | 3011 |
| Sabah | Kudat | 12·158 | 8·935 | 34·031 | 5·825 | 15·951 | 8843 | 4389 | 11583 |
| Sabah | Kunak | 12·605 | 8·713 | 32·539 | 6·343 | 16·561 | 7272 | 3604 | 9542 |
| Sabah | Lahad Datu | 12·555 | 8·933 | 33·717 | 6·189 | 16·370 | 29398 | 14046 | 38498 |
| Sabah | Nabawan | 12·673 | 8·947 | 33·854 | 6·327 | 16·480 | 3974 | 2089 | 5108 |
| Sabah | Papar | 12·858 | 8·569 | 32·548 | 6·678 | 16·736 | 18319 | 9405 | 23965 |
| Sabah | Penamnang | 13·006 | 8·357 | 31·507 | 6·945 | 17·045 | 40659 | 21453 | 53268 |
| Sabah | Pitas | 12·892 | 9·055 | 34·063 | 6·430 | 16·833 | 4965 | 2496 | 6493 |
| Sabah | Ranau | 13·491 | 8·454 | 32·130 | 7·366 | 17·478 | 12806 | 6956 | 16601 |
| Sabah | Sandakan | 13·056 | 8·894 | 33·687 | 6·662 | 16·983 | 55401 | 27764 | 72428 |
| Sabah | Semporna | 12·323 | 9·591 | 36·061 | 5·492 | 16·246 | 22202 | 9525 | 29207 |
| Sabah | Sipitang | 12·092 | 9·258 | 35·024 | 5·550 | 15·861 | 4861 | 2241 | 6378 |
| Sabah | Tambunan | 13·238 | 8·312 | 31·516 | 7·251 | 17·132 | 4834 | 2646 | 6246 |
| Sabah | Tawau | 12·727 | 9·021 | 34·143 | 6·233 | 16·702 | 51066 | 24055 | 66731 |
| Sabah | Tenom | 12·425 | 8·796 | 33·416 | 6·160 | 16·227 | 7241 | 3714 | 9369 |
| Sabah | Tongod | 13·420 | 8·607 | 32·651 | 7·207 | 17·437 | 6829 | 3735 | 8890 |
| Sabah | Tuaran | 13·046 | 8·450 | 32·072 | 6·942 | 16·926 | 19565 | 10255 | 25390 |
| Sarawak | Asajaya | 6·342 | 8·353 | 30·473 | 1·408 | 7·732 | 1544 | 345 | 1907 |
| Sarawak | Bau | 6·137 | 8·196 | 30·577 | 1·310 | 7·445 | 3599 | 766 | 4341 |
| Sarawak | Belaga | 9·258 | 10·596 | 39·211 | 2·533 | 11·804 | 2234 | 606 | 2854 |
| Sarawak | Betong | 6·649 | 8·948 | 32·460 | 1·484 | 7·978 | 2536 | 570 | 3008 |
| Sarawak | Bintulu | 8·703 | 10·125 | 37·657 | 2·397 | 10·888 | 18048 | 4788 | 22239 |
| Sarawak | Bukit Mabong | 7·929 | 9·993 | 36·885 | 1·890 | 9·768 | 3077 | 737 | 3801 |
| Sarawak | Dalat | 7·212 | 9·203 | 34·359 | 1·775 | 8·758 | 1834 | 463 | 2232 |
| Sarawak | Daro | 6·897 | 8·957 | 32·464 | 1·576 | 8·485 | 794 | 179 | 983 |
| Sarawak | Julau | 6·954 | 9·129 | 33·772 | 1·574 | 8·405 | 799 | 175 | 972 |
| Sarawak | Kabong | 6·566 | 8·519 | 31·073 | 1·475 | 8·046 | 1873 | 421 | 2296 |
| Sarawak | Kanowit | 7·160 | 9·309 | 34·341 | 1·658 | 8·665 | 1880 | 424 | 2273 |
| Sarawak | Kapit | 8·254 | 10·193 | 37·700 | 2·015 | 10·283 | 899 | 204 | 1133 |
| Sarawak | Kuching | 6·183 | 8·159 | 30·383 | 1·339 | 7·570 | 44029 | 9569 | 54382 |
| Sarawak | Lawas | 11·793 | 9·652 | 36·382 | 5·062 | 15·453 | 4854 | 2152 | 6378 |
| Sarawak | Limbang | 11·215 | 9·971 | 37·683 | 4·354 | 14·724 | 6088 | 2408 | 8034 |
| Sarawak | Lubok Antu | 6·829 | 9·093 | 33·477 | 1·502 | 8·269 | 1776 | 384 | 2123 |
| Sarawak | Lundu | 6·054 | 7·909 | 29·352 | 1·325 | 7·407 | 2588 | 570 | 3145 |
| Sarawak | Marudi | 10·294 | 10·400 | 38·900 | 3·374 | 13·391 | 8455 | 2750 | 10983 |

| ADM1 | ADM2 | β-thalassaemia carrier prevalence | | | | | Estimated carrier counts | | |
|---|---|---|---|---|---|---|---|---|---|
| | | **Mean** | **SD** | **CI95%** | **Q25** | **Q75** | **Mean** | **Q25** | **Q75** |
| Sarawak | Matu | 7·093 | 9·118 | 33·586 | 1·684 | 8·668 | 1196 | 283 | 1470 |
| Sarawak | Meradong | 6·876 | 8·931 | 33·111 | 1·562 | 8·350 | 1425 | 328 | 1733 |
| Sarawak | Miri | 9·300 | 9·915 | 36·656 | 2·852 | 12·020 | 28747 | 9326 | 37558 |
| Sarawak | Mukah | 7·607 | 9·409 | 35·241 | 1·931 | 9·328 | 2232 | 562 | 2743 |
| Sarawak | Pakan | 6·848 | 9·042 | 33·364 | 1·553 | 8·288 | 1144 | 260 | 1392 |
| Sarawak | Pusa | 6·462 | 8·484 | 30·986 | 1·423 | 7·878 | 543 | 122 | 664 |
| Sarawak | Samarahan | 6·198 | 8·144 | 30·028 | 1·361 | 7·568 | 5926 | 1317 | 7093 |
| Sarawak | Saratok | 6·672 | 8·740 | 32·289 | 1·501 | 8·156 | 1443 | 322 | 1717 |
| Sarawak | Sarikei | 6·699 | 8·630 | 31·392 | 1·555 | 8·175 | 3303 | 769 | 3990 |
| Sarawak | Selangau | 7·568 | 9·689 | 36·301 | 1·804 | 9·158 | 1611 | 383 | 1958 |
| Sarawak | Serian | 6·271 | 8·654 | 32·072 | 1·338 | 7·458 | 1453 | 309 | 1727 |
| Sarawak | Sibu | 7·116 | 9·239 | 34·121 | 1·650 | 8·674 | 19982 | 4620 | 24636 |
| Sarawak | Simunjan | 6·366 | 8·521 | 30·738 | 1·380 | 7·678 | 2709 | 585 | 3288 |
| Sarawak | Song | 7·279 | 9·384 | 34·901 | 1·680 | 8·841 | 712 | 164 | 853 |
| Sarawak | Sri Aman | 6·508 | 8·795 | 31·886 | 1·434 | 7·801 | 4235 | 969 | 5103 |
| Sarawak | Tanjung Manis | 6·758 | 8·694 | 32·097 | 1·546 | 8·272 | 535 | 122 | 657 |
| Sarawak | Tatau | 8·094 | 9·970 | 37·173 | 2·037 | 9·998 | 2539 | 638 | 3130 |
| Sarawak | Tebedu | 6·303 | 8·515 | 30·943 | 1·353 | 7·602 | 6345 | 1388 | 7738 |
| Selangor | Gombak | 4·124 | 3·442 | 12·751 | 1·860 | 5·251 | 40205 | 17975 | 50937 |
| Selangor | Hulu Langat | 4·068 | 3·441 | 12·647 | 1·805 | 5·161 | 53277 | 23695 | 67354 |
| Selangor | Hulu Selangor | 4·269 | 3·715 | 13·570 | 1·834 | 5·473 | 11654 | 5099 | 14965 |
| Selangor | Klang | 4·197 | 3·653 | 13·581 | 1·821 | 5·323 | 57628 | 25158 | 73435 |
| Selangor | Kuala Langat | 4·193 | 3·618 | 13·331 | 1·795 | 5·319 | 14239 | 6145 | 18068 |
| Selangor | Kuala Selangor | 4·295 | 3·782 | 13·832 | 1·834 | 5·450 | 15741 | 6808 | 19883 |
| Selangor | Petaling | 4·104 | 3·421 | 12·561 | 1·850 | 5·236 | 91404 | 41273 | 117070 |
| Selangor | Sabak Bernam | 4·583 | 4·307 | 15·554 | 1·846 | 5·826 | 5455 | 2196 | 6912 |
| Selangor | Sepang | 4·129 | 3·508 | 12·931 | 1·782 | 5·305 | 23180 | 10144 | 29443 |
| Terengganu | Besut | 4·952 | 5·194 | 18·522 | 1·685 | 6·295 | 8555 | 2876 | 10885 |
| Terengganu | Dungun | 4·781 | 5·189 | 18·747 | 1·503 | 6·084 | 5838 | 1773 | 7465 |
| Terengganu | Hulu Terengganu | 4·850 | 5·160 | 18·783 | 1·593 | 6·154 | 4231 | 1324 | 5382 |
| Terengganu | Kemanman | 4·772 | 5·252 | 19·005 | 1·496 | 6·033 | 10472 | 3155 | 13099 |
| Terengganu | Kuala Nerus | 4·849 | 5·401 | 19·109 | 1·513 | 6·190 | 11009 | 3357 | 14162 |
| Terengganu | Kuala Terengganu | 4·825 | 5·314 | 19·030 | 1·494 | 6·205 | 4585 | 1429 | 5910 |
| Terengganu | Marang | 4·803 | 5·331 | 19·772 | 1·467 | 6·071 | 4631 | 1419 | 5855 |
| Terengganu | Setiu | 4·863 | 5·320 | 19·042 | 1·573 | 6·120 | 3102 | 990 | 3920 |

ADM1: administrative level 1. ADM2: administrative level 2. SD: standard deviation. CI95%: 95% credible interval. Q25: 25$^{th}$ percentile. Q75: 75$^{th}$ percentile. SD and CI95% are measures of prediction uncertainty.

**Supplementary Table S28. Subnational areal predictions of β-thalassaemia carrier prevalence and estimated carrier count at administrative level 2 of Malta.**

| ADM1 | ADM2 | β-thalassaemia carrier prevalence | | | | | Estimated carrier counts | | |
|---|---|---|---|---|---|---|---|---|---|
| | | **Mean** | **SD** | **CI95%** | **Q25** | **Q75** | **Mean** | **Q25** | **Q75** |
| Attard | Attard | 4·055 | 3·380 | 12·262 | 1·852 | 5·160 | 10471 | 4737 | 13279 |
| Balzan | Balzan | 4·091 | 3·508 | 12·661 | 1·838 | 5·141 | 7810 | 3502 | 9858 |
| Birgu | Birgu | 3·961 | 3·325 | 12·405 | 1·874 | 4·915 | 3434 | 1625 | 4261 |
| Birkirkara | Birkirkara | 4·116 | 3·486 | 12·859 | 1·855 | 5·257 | 10664 | 4797 | 13620 |
| Birzebbuga | Birzebbuga | 3·961 | 3·325 | 12·405 | 1·874 | 4·915 | 3434 | 1625 | 4261 |
| Bormla | Bormla | 3·961 | 3·325 | 12·405 | 1·874 | 4·915 | 3434 | 1625 | 4261 |
| Dingli | Dingli | 4·010 | 3·305 | 12·309 | 1·787 | 5·107 | 590 | 263 | 751 |
| Fgura | Fgura | 3·961 | 3·325 | 12·405 | 1·874 | 4·915 | 3434 | 1625 | 4261 |
| Floriana | Floriana | 4·053 | 3·362 | 12·525 | 1·855 | 5·189 | 11104 | 5069 | 14189 |
| Fontana | Fontana | 4·276 | 3·514 | 13·671 | 1·959 | 5·391 | 1179 | 539 | 1477 |
| Ghajnsielem | Ghajnsielem | 4·228 | 3·536 | 13·213 | 1·936 | 5·401 | 1693 | 775 | 2145 |
| Gharb | Gharb | 4·261 | 3·360 | 13·131 | 1·961 | 5·431 | 260 | 120 | 331 |
| Gharghur | Gharghur | 4·091 | 3·508 | 12·661 | 1·838 | 5·141 | 7810 | 3502 | 9858 |
| Ghasri | Ghasri | 4·276 | 3·514 | 13·671 | 1·959 | 5·391 | 1179 | 539 | 1477 |

| ADM1 | ADM2 | β-thalassaemia carrier prevalence Mean | SD | CI95% | Q25 | Q75 | Estimated carrier counts Mean | Q25 | Q75 |
|---|---|---|---|---|---|---|---|---|---|
| Ghaxaq | Ghaxaq | 3·961 | 3·325 | 12·405 | 1·874 | 4·915 | 3434 | 1625 | 4261 |
| Gudja | Gudja | 4·062 | 3·383 | 12·831 | 1·882 | 5·202 | 6288 | 2920 | 8023 |
| Gzira | Gzira | 4·033 | 3·320 | 11·913 | 1·800 | 5·164 | 4816 | 2149 | 6166 |
| Hamrun | Hamrun | 4·099 | 3·380 | 12·585 | 1·845 | 5·326 | 7670 | 3444 | 9928 |
| Iklin | Iklin | 4·091 | 3·508 | 12·661 | 1·838 | 5·141 | 7810 | 3502 | 9858 |
| Isla | Isla | 3·961 | 3·325 | 12·405 | 1·874 | 4·915 | 3434 | 1625 | 4261 |
| Kalkara | Kalkara | 3·961 | 3·325 | 12·405 | 1·874 | 4·915 | 3434 | 1625 | 4261 |
| Kercem | Kercem | 4·261 | 3·360 | 13·131 | 1·961 | 5·431 | 260 | 120 | 331 |
| Kirkop | Kirkop | 4·164 | 3·441 | 13·256 | 1·890 | 5·489 | 2854 | 1295 | 3762 |
| Lija | Lija | 4·149 | 3·697 | 13·408 | 1·876 | 5·117 | 2994 | 1353 | 3692 |
| Luqa | Luqa | 4·164 | 3·441 | 13·256 | 1·890 | 5·489 | 2854 | 1295 | 3762 |
| Marsa | Marsa | 4·164 | 3·441 | 13·256 | 1·890 | 5·489 | 2854 | 1295 | 3762 |
| Marsaskala | Marsaskala | 4·231 | 3·645 | 12·525 | 1·891 | 5·417 | 915 | 409 | 1171 |
| Marsaxlokk | Marsaxlokk | 4·096 | 3·485 | 12·465 | 1·882 | 5·166 | 4349 | 2034 | 5432 |
| Mdina | Mdina | 4·010 | 3·305 | 12·309 | 1·787 | 5·107 | 590 | 263 | 751 |
| Mellieha | Mellieha | 4·158 | 3·324 | 11·974 | 1·876 | 5·427 | 334 | 151 | 436 |
| Mgarr | Mgarr | 4·211 | 3·549 | 13·443 | 1·952 | 5·365 | 235 | 109 | 299 |
| Mosta | Mosta | 4·111 | 3·488 | 12·746 | 1·908 | 5·169 | 3943 | 1805 | 4909 |
| Mqabba | Mqabba | 4·087 | 3·369 | 12·426 | 1·875 | 5·341 | 3976 | 1815 | 5215 |
| Msida | Msida | 4·099 | 3·380 | 12·585 | 1·845 | 5·326 | 7670 | 3444 | 9928 |
| Mtarfa | Mtarfa | 4·042 | 3·292 | 12·197 | 1·863 | 5·163 | 1539 | 715 | 1968 |
| Munxar | Munxar | 4·276 | 3·514 | 13·671 | 1·959 | 5·391 | 1179 | 539 | 1477 |
| Nadur | Nadur | 4·234 | 3·617 | 13·455 | 1·975 | 5·424 | 440 | 205 | 563 |
| Naxxar | Naxxar | 4·149 | 3·697 | 13·408 | 1·876 | 5·117 | 2994 | 1353 | 3692 |
| Paola | Paola | 4·062 | 3·383 | 12·831 | 1·882 | 5·202 | 6288 | 2920 | 8023 |
| Pembroke | Pembroke | 4·033 | 3·320 | 11·913 | 1·800 | 5·164 | 4816 | 2149 | 6166 |
| Pieta | Pieta | 4·099 | 3·380 | 12·585 | 1·845 | 5·326 | 7670 | 3444 | 9928 |
| Qala | Qala | 4·234 | 3·617 | 13·455 | 1·975 | 5·424 | 440 | 205 | 563 |
| Qormi | Qormi | 4·069 | 3·353 | 12·255 | 1·850 | 5·282 | 8792 | 3964 | 11381 |
| Qrendi | Qrendi | 4·087 | 3·369 | 12·426 | 1·875 | 5·341 | 3976 | 1815 | 5215 |
| Rabat Gozo | Rabat Gozo | 4·276 | 3·514 | 13·671 | 1·959 | 5·391 | 1179 | 539 | 1477 |
| Rabat Malta | Rabat Malta | 4·091 | 3·426 | 12·568 | 1·883 | 5·200 | 5890 | 2697 | 7412 |
| Safi | Safi | 4·062 | 3·383 | 12·831 | 1·882 | 5·202 | 6288 | 2920 | 8023 |
| Saint John | Saint John | 4·033 | 3·320 | 11·913 | 1·800 | 5·164 | 4816 | 2149 | 6166 |
| Saint Julian's | Saint Julian's | 4·033 | 3·320 | 11·913 | 1·800 | 5·164 | 4816 | 2149 | 6166 |
| Saint Lawerence | Saint Lawerence | 4·261 | 3·360 | 13·131 | 1·961 | 5·431 | 260 | 120 | 331 |
| Saint Lucia's | Saint Lucia's | 4·062 | 3·383 | 12·831 | 1·882 | 5·202 | 6288 | 2920 | 8023 |
| Saint Paul's Bay | Saint Paul's Bay | 4·073 | 3·280 | 12·085 | 1·940 | 5·220 | 949 | 452 | 1217 |
| Sannat | Sannat | 4·276 | 3·514 | 13·671 | 1·959 | 5·391 | 1179 | 539 | 1477 |
| Santa Venera | Santa Venera | 4·099 | 3·380 | 12·585 | 1·845 | 5·326 | 7670 | 3444 | 9928 |
| Siggiewi | Siggiewi | 4·061 | 3·348 | 12·387 | 1·845 | 5·263 | 4566 | 2078 | 5966 |
| Sliema | Sliema | 4·033 | 3·320 | 11·913 | 1·800 | 5·164 | 4816 | 2149 | 6166 |
| Swieqi | Swieqi | 4·033 | 3·320 | 11·913 | 1·800 | 5·164 | 4816 | 2149 | 6166 |
| Ta' Xbiex | Ta' Xbiex | 4·033 | 3·320 | 11·913 | 1·800 | 5·164 | 4816 | 2149 | 6166 |
| Tarxien | Tarxien | 4·062 | 3·383 | 12·831 | 1·882 | 5·202 | 6288 | 2920 | 8023 |
| Xaghra | Xaghra | 4·096 | 3·485 | 12·465 | 1·882 | 5·166 | 4349 | 2034 | 5432 |
| Xewkija | Xewkija | 4·291 | 3·667 | 14·210 | 1·957 | 5·351 | 919 | 419 | 1146 |
| Xghajra | Xghajra | 4·263 | 3·642 | 13·833 | 1·966 | 5·387 | 1359 | 624 | 1709 |
| Zabbar | Zabbar | 4·096 | 3·485 | 12·465 | 1·882 | 5·166 | 4349 | 2034 | 5432 |
| Zebbug Gozo | Zebbug Gozo | 4·276 | 3·514 | 13·671 | 1·959 | 5·391 | 1179 | 539 | 1477 |
| Zebbug Malta | Zebbug Malta | 4·009 | 3·298 | 11·596 | 1·860 | 5·192 | 1122 | 520 | 1453 |
| Zejtun | Zejtun | 3·961 | 3·325 | 12·405 | 1·874 | 4·915 | 3434 | 1625 | 4261 |
| Zurrieq | Zurrieq | 4·062 | 3·383 | 12·831 | 1·882 | 5·202 | 6288 | 2920 | 8023 |

ADM1: administrative level 1. ADM2: administrative level 2. SD: standard deviation. CI95%: 95% credible interval. Q25: 25$^{th}$ percentile. Q75: 75$^{th}$ percentile. SD and CI95% are measures of prediction uncertainty.

**Supplementary Table S29. Subnational areal predictions of β-thalassaemia carrier prevalence and estimated carrier count at administrative level 2 of Myanmar.**

| | | β-thalassaemia carrier prevalence | | | | | Estimated carrier counts | | |
|---|---|---|---|---|---|---|---|---|---|
| **ADM1** | **ADM2** | **Mean** | **SD** | **CI95%** | **Q25** | **Q75** | **Mean** | **Q25** | **Q75** |
| Ayeyarwady | Hinthada | 7·432 | 6·375 | 23·599 | 3·138 | 9·613 | 85009 | 36793 | 109622 |
| Ayeyarwady | Labutta | 8·127 | 7·253 | 27·234 | 3·252 | 10·514 | 48161 | 19814 | 62421 |
| Ayeyarwady | Maubin | 8·046 | 6·021 | 22·596 | 3·867 | 10·417 | 78458 | 37939 | 101487 |
| Ayeyarwady | Myaungmya | 8·230 | 6·987 | 26·161 | 3·485 | 10·674 | 64726 | 27463 | 83842 |
| Ayeyarwady | Pathein | 7·972 | 7·111 | 26·432 | 3·213 | 10·316 | 122451 | 51181 | 158887 |
| Ayeyarwady | Pyapon | 8·058 | 6·639 | 24·664 | 3·516 | 10·449 | 80566 | 36240 | 104428 |
| Bago | Bago | 6·809 | 5·421 | 20·234 | 3·145 | 8·729 | 122336 | 57854 | 156618 |
| Bago | Pyay | 6·375 | 6·254 | 23·149 | 2·324 | 8·150 | 61292 | 22738 | 78053 |
| Bago | Taungoo | 6·087 | 5·994 | 22·126 | 2·245 | 7·728 | 68347 | 25475 | 86994 |
| Bago | Thayarwady | 7·178 | 6·094 | 22·625 | 3·078 | 9·272 | 79226 | 34004 | 102045 |
| Chin | Falam | 1·397 | 1·616 | 5·452 | 0·480 | 1·704 | 2636 | 900 | 3243 |
| Chin | Hakha | 1·940 | 2·484 | 8·340 | 0·586 | 2·316 | 2073 | 626 | 2469 |
| Chin | Mindat | 2·882 | 3·640 | 12·650 | 0·830 | 3·468 | 6470 | 1861 | 7853 |
| Kachin | Bhamo | 3·095 | 4·074 | 14·011 | 0·798 | 3·733 | 13679 | 3555 | 16544 |
| Kachin | Mohnyin | 3·294 | 3·641 | 12·997 | 1·078 | 4·122 | 27154 | 9004 | 34043 |
| Kachin | Myitkyina | 4·098 | 4·747 | 16·930 | 1·261 | 5·084 | 25510 | 7348 | 31665 |
| Kachin | Puta-O | 5·072 | 5·471 | 19·661 | 1·705 | 6·357 | 5628 | 1927 | 7022 |
| Kayah | Bawlake | 5·549 | 5·857 | 21·410 | 1·916 | 6·948 | 2839 | 969 | 3548 |
| Kayah | Loikaw | 5·237 | 5·924 | 21·361 | 1·657 | 6·513 | 15003 | 4685 | 18390 |
| Kayin | Hpa-An | 5·400 | 5·118 | 18·740 | 2·144 | 6·818 | 43849 | 18561 | 55364 |
| Kayin | Hpapun | 5·750 | 5·311 | 19·587 | 2·302 | 7·337 | 2469 | 996 | 3131 |
| Kayin | Kawkareik | 4·212 | 3·904 | 14·298 | 1·707 | 5·346 | 21345 | 8771 | 27074 |
| Kayin | Myawaddy | 3·977 | 3·682 | 13·444 | 1·640 | 5·043 | 7281 | 3113 | 9350 |
| Magway | Gangaw | 2·622 | 3·492 | 11·867 | 0·738 | 3·095 | 6142 | 1732 | 7235 |
| Magway | Magway | 4·430 | 5·274 | 19·087 | 1·321 | 5·422 | 52926 | 15756 | 64331 |
| Magway | Minbu | 4·211 | 5·147 | 18·448 | 1·234 | 5·084 | 28765 | 8423 | 34686 |
| Magway | Pakokku | 2·977 | 3·884 | 13·378 | 0·828 | 3·549 | 31109 | 8576 | 37085 |
| Magway | Thayet | 5·454 | 5·926 | 21·910 | 1·785 | 6·811 | 39568 | 13059 | 49251 |
| Mandalay | Det Khi Na | 5·236 | 5·870 | 21·114 | 1·690 | 6·472 | 39345 | 12700 | 48882 |
| Mandalay | Kyaukse | 3·290 | 4·465 | 15·459 | 0·817 | 3·908 | 27787 | 6926 | 32931 |
| Mandalay | Mandalay | 2·967 | 4·037 | 14·348 | 0·731 | 3·517 | 55652 | 13801 | 66893 |
| Mandalay | Meiktila | 3·769 | 4·765 | 16·771 | 1·004 | 4·591 | 35696 | 9452 | 43420 |
| Mandalay | Myingyan | 3·153 | 4·160 | 14·395 | 0·823 | 3·779 | 27020 | 7038 | 32544 |
| Mandalay | Nyaung-U | 3·630 | 4·667 | 16·494 | 0·991 | 4·352 | 18048 | 4947 | 21651 |
| Mandalay | Oke Ta Ra | 4·743 | 5·500 | 19·816 | 1·442 | 5·860 | 27919 | 8690 | 34720 |
| Mandalay | Pyinoolwin | 2·641 | 3·615 | 12·403 | 0·667 | 3·122 | 28567 | 7104 | 33603 |
| Mandalay | Yamethin | 4·226 | 5·051 | 18·245 | 1·207 | 5·214 | 23211 | 6568 | 28790 |
| Mon | Mawlamyine | 4·738 | 4·435 | 16·304 | 1·884 | 6·023 | 51188 | 20794 | 65010 |
| Mon | Thaton | 6·036 | 5·024 | 18·474 | 2·706 | 7·702 | 41554 | 18745 | 53403 |
| Rakhine | Kyaukpyu | 5·525 | 6·336 | 23·345 | 1·646 | 6·877 | 31189 | 9273 | 38810 |
| Rakhine | Maungdaw | 3·759 | 4·582 | 16·211 | 1·055 | 4·624 | 6124 | 1692 | 7584 |
| Rakhine | Mrauk-U | 4·286 | 5·356 | 18·930 | 1·189 | 5·205 | 39294 | 10876 | 47678 |
| Rakhine | Sittwe | 4·502 | 5·701 | 20·410 | 1·207 | 5·476 | 33400 | 8700 | 40923 |
| Rakhine | Thandwe | 6·549 | 6·761 | 24·954 | 2·225 | 8·336 | 33199 | 11086 | 42256 |
| Saigang | Hkamti | 3·534 | 3·022 | 10·995 | 1·561 | 4·500 | 12462 | 5397 | 15797 |
| Saigang | Kale | 1·581 | 2·054 | 6·690 | 0·485 | 1·881 | 8145 | 2531 | 9668 |
| Saigang | Kanbalu | 1·890 | 2·440 | 8·233 | 0·546 | 2·264 | 8043 | 2302 | 9608 |
| Saigang | Katha | 2·060 | 2·578 | 8·896 | 0·610 | 2·479 | 18593 | 5420 | 22257 |
| Saigang | Mawlaik | 1·353 | 1·528 | 5·144 | 0·481 | 1·653 | 2283 | 805 | 2794 |
| Saigang | Monywa | 2·375 | 3·186 | 10·897 | 0·638 | 2·815 | 18071 | 4909 | 21660 |
| Saigang | Sagaing | 2·716 | 3·625 | 12·640 | 0·712 | 3·223 | 12759 | 3397 | 15156 |
| Saigang | Shwebo | 2·111 | 2·816 | 9·506 | 0·577 | 2·511 | 24478 | 6572 | 29105 |
| Saigang | Tamu | 1·220 | 1·256 | 4·441 | 0·469 | 1·506 | 1347 | 518 | 1643 |
| Saigang | Yinmarbin | 2·225 | 2·969 | 9·994 | 0·624 | 2·635 | 14292 | 3989 | 16937 |
| Shan | Hopang | 4·016 | 5·282 | 18·788 | 0·919 | 4·924 | 10698 | 2447 | 13042 |

| | | β-thalassaemia carrier prevalence | | | | | Estimated carrier counts | | |
|---|---|---|---|---|---|---|---|---|---|
| ADM1 | ADM2 | Mean | SD | CI95% | Q25 | Q75 | Mean | Q25 | Q75 |
| Shan | Kengtung | 4·310 | 5·594 | 20·003 | 1·032 | 5·265 | 22119 | 5321 | 26990 |
| Shan | Kyaukme | 3·230 | 4·349 | 15·225 | 0·771 | 3·881 | 24001 | 5712 | 28647 |
| Shan | Langkho | 4·617 | 5·607 | 20·018 | 1·275 | 5·730 | 7335 | 1966 | 9057 |
| Shan | Lashio | 3·733 | 4·901 | 17·334 | 0·871 | 4·576 | 26756 | 6236 | 32593 |
| Shan | Laukkaing | 3·884 | 5·296 | 18·781 | 0·899 | 4·633 | 8251 | 1918 | 9879 |
| Shan | Loilen | 4·039 | 5·195 | 18·616 | 0·987 | 4·920 | 25822 | 6351 | 31208 |
| Shan | Matman | 4·067 | 5·286 | 19·019 | 0·950 | 5·007 | 8310 | 1941 | 10310 |
| Shan | Monghsat | 4·444 | 5·439 | 19·178 | 1·215 | 5·520 | 8388 | 2276 | 10395 |
| Shan | Mongmit | 2·658 | 3·583 | 12·406 | 0·679 | 3·168 | 3239 | 824 | 3868 |
| Shan | Muse | 3·554 | 4·743 | 16·612 | 0·841 | 4·309 | 20822 | 5010 | 25369 |
| Shan | Tachileik | 4·438 | 5·686 | 20·386 | 1·113 | 5·412 | 14316 | 3744 | 17297 |
| Shan | Taunggyi | 4·282 | 5·331 | 18·846 | 1·167 | 5·246 | 81313 | 21989 | 99925 |
| Tanitharyi | Dawei | 3·343 | 3·620 | 12·708 | 1·148 | 4·191 | 16390 | 5447 | 20456 |
| Tanitharyi | Kawthoung | 3·765 | 4·623 | 16·661 | 1·019 | 4·613 | 5621 | 1503 | 6887 |
| Tanitharyi | Myeik | 3·054 | 3·612 | 12·845 | 0·912 | 3·749 | 17659 | 5383 | 21893 |
| Yangon | Yangon (East) | 7·512 | 5·084 | 19·076 | 4·002 | 9·547 | 162585 | 86957 | 205653 |
| Yangon | Yangon (North) | 7·661 | 5·298 | 19·847 | 3·957 | 9·858 | 236263 | 124557 | 302993 |
| Yangon | Yangon (South) | 7·489 | 5·331 | 19·999 | 3·787 | 9·631 | 135716 | 69663 | 174320 |
| Yangon | Yangon (West) | 7·771 | 5·095 | 19·063 | 4·252 | 9·937 | 121058 | 66154 | 154495 |

ADM1: administrative level 1. ADM2: administrative level 2. SD: standard deviation. CI95%: 95% credible interval. Q25: 25th percentile. Q75: 75th percentile. SD and CI95% are measures of prediction uncertainty.

**Supplementary Table S30. Subnational areal predictions of β-thalassaemia carrier prevalence and estimated carrier count at administrative level 2 of Oman.**

| | | β-thalassaemia carrier prevalence | | | | | Estimated carrier counts | | |
|---|---|---|---|---|---|---|---|---|---|
| ADM1 | ADM2 | Mean | SD | CI95% | Q25 | Q75 | Mean | Q25 | Q75 |
| Ad Dakhiliyah | Wilayat Adam | 2·865 | 2·218 | 8·168 | 1·398 | 3·632 | 950 | 461 | 1183 |
| Ad Dakhiliyah | Wilayat Al Hamra | 2·625 | 1·749 | 6·532 | 1·432 | 3·314 | 1091 | 601 | 1379 |
| Ad Dakhiliyah | Wilayat Bahla | 2·720 | 1·903 | 7·080 | 1·438 | 3·438 | 2682 | 1455 | 3392 |
| Ad Dakhiliyah | Wilayat Bidbid | 2·518 | 1·807 | 6·759 | 1·312 | 3·160 | 1215 | 642 | 1538 |
| Ad Dakhiliyah | Wilayat Izki | 2·622 | 1·793 | 6·674 | 1·413 | 3·298 | 1836 | 1004 | 2312 |
| Ad Dakhiliyah | Wilayat Manah | 2·675 | 1·840 | 6·785 | 1·428 | 3·387 | 866 | 468 | 1090 |
| Ad Dakhiliyah | Wilayat Nizwa | 2·624 | 1·777 | 6·559 | 1·426 | 3·302 | 4419 | 2436 | 5529 |
| Ad Dakhiliyah | Wilayat Samail | 2·550 | 1·790 | 6·616 | 1·350 | 3·220 | 2489 | 1309 | 3164 |
| Al Batinah | Wilayat Al Awabi | 2·567 | 1·750 | 6·463 | 1·412 | 3·186 | 630 | 351 | 784 |
| Al Batinah | Wilayat Al Khaburah | 2·558 | 1·817 | 6·747 | 1·336 | 3·229 | 2844 | 1464 | 3593 |
| Al Batinah | Wilayat Al Musanaah | 2·539 | 1·856 | 6·924 | 1·292 | 3·231 | 2619 | 1319 | 3345 |
| Al Batinah | Wilayat Ar Rustaq | 2·564 | 1·761 | 6·524 | 1·383 | 3·235 | 3638 | 1958 | 4596 |
| Al Batinah | Wilayat As Suwayq | 2·525 | 1·796 | 6·717 | 1·315 | 3·200 | 4198 | 2160 | 5281 |
| Al Batinah | Wilayat Barka | 2·495 | 1·834 | 6·783 | 1·271 | 3·158 | 6100 | 3101 | 7709 |
| Al Batinah | Wilayat Liwa | 2·280 | 1·610 | 5·975 | 1·186 | 2·879 | 1385 | 698 | 1737 |
| Al Batinah | Wilayat Nakhal | 2·499 | 1·728 | 6·448 | 1·345 | 3·144 | 845 | 449 | 1065 |
| Al Batinah | Wilayat Saham | 2·512 | 1·776 | 6·543 | 1·318 | 3·176 | 3522 | 1837 | 4471 |
| Al Batinah | Wilayat Shinas | 2·146 | 1·606 | 5·873 | 1·084 | 2·701 | 1680 | 839 | 2140 |
| Al Batinah | Wilayat Sohar | 2·425 | 1·707 | 6·315 | 1·273 | 3·078 | 5516 | 2852 | 7055 |
| Al Batinah | Wilayat Wadi Al Maawil | 2·494 | 1·714 | 6·455 | 1·334 | 3·155 | 719 | 382 | 904 |
| Al Wusta | Wilayat Ad Duqm | 3·288 | 3·189 | 11·500 | 1·303 | 4·130 | 710 | 281 | 900 |
| Al Wusta | Wilayat Al Jazir | 3·020 | 2·668 | 9·649 | 1·322 | 3·812 | 188 | 77 | 237 |
| Al Wusta | Wilayat Hayma | 2·971 | 2·414 | 8·826 | 1·392 | 3·762 | 357 | 160 | 456 |
| Al Wusta | Wilayat Mahawt | 3·096 | 2·721 | 10·018 | 1·336 | 3·913 | 629 | 263 | 788 |
| Ash Sharqiyah | Wilayat Al Kamil Wa Al Wafi | 2·824 | 2·180 | 8·048 | 1·365 | 3·598 | 1269 | 618 | 1614 |
| Ash Sharqiyah | Wilayat Al Mudaybi | 2·682 | 1·893 | 6·981 | 1·410 | 3·389 | 3767 | 1998 | 4734 |
| Ash Sharqiyah | Wilayat Al Qabil | 2·670 | 1·953 | 7·258 | 1·370 | 3·360 | 881 | 453 | 1120 |
| Ash Sharqiyah | Wilayat Bidiyah | 2·724 | 2·007 | 7·472 | 1·377 | 3·443 | 1388 | 715 | 1769 |
| Ash Sharqiyah | Wilayat Dama Wa At Taiyin | 2·628 | 1·933 | 7·063 | 1·345 | 3·322 | 874 | 449 | 1099 |
| Ash Sharqiyah | Wilayat Ibra | 2·658 | 1·955 | 7·219 | 1·370 | 3·335 | 1618 | 830 | 2032 |

| ADM1 | ADM2 | β-thalassaemia carrier prevalence Mean | SD | CI95% | Q25 | Q75 | Estimated carrier counts Mean | Q25 | Q75 |
|---|---|---|---|---|---|---|---|---|---|
| Ash Sharqiyah | Wilayat Jaalan Bani Bu Ali | 2·970 | 2·392 | 8·768 | 1·390 | 3·755 | 3806 | 1823 | 4817 |
| Ash Sharqiyah | Wilayat Jaalan Bani Bu Hasan | 2·904 | 2·292 | 8·411 | 1·392 | 3·681 | 1460 | 698 | 1850 |
| Ash Sharqiyah | Wilayat Sur | 2·909 | 2·411 | 8·821 | 1·324 | 3·690 | 3377 | 1473 | 4305 |
| Ash Sharqiyah | Wilayat Wadi Bani Khalid | 2·768 | 2·154 | 7·920 | 1·360 | 3·474 | 456 | 220 | 575 |
| Az Zahirah | Wilayat Al Buraymi | 2·611 | 1·880 | 6·915 | 1·349 | 3·306 | 3575 | 1855 | 4538 |
| Az Zahirah | Wilayat As Sunaynah | 2·866 | 2·135 | 7·939 | 1·444 | 3·631 | 590 | 288 | 742 |
| Az Zahirah | Wilayat Bukha | 1·196 | 0·958 | 3·541 | 0·565 | 1·526 | 35 | 18 | 45 |
| Az Zahirah | Wilayat Daba | 1·412 | 1·236 | 4·500 | 0·639 | 1·758 | 187 | 86 | 230 |
| Az Zahirah | Wilayat Dank | 2·739 | 1·969 | 7·302 | 1·419 | 3·451 | 817 | 417 | 1028 |
| Az Zahirah | Wilayat Ibri | 2·893 | 2·162 | 7·949 | 1·458 | 3·655 | 5528 | 2881 | 6987 |
| Az Zahirah | Wilayat Khasab | 1·200 | 1·018 | 3·615 | 0·549 | 1·515 | 284 | 130 | 367 |
| Az Zahirah | Wilayat Madha | 1·790 | 1·427 | 5·714 | 0·843 | 2·304 | 159 | 75 | 204 |
| Az Zahirah | Wilayat Mahadah | 2·320 | 1·703 | 6·250 | 1·194 | 2·927 | 714 | 361 | 904 |
| Az Zahirah | Wilayat Masirah | 3·343 | 3·242 | 11·556 | 1·328 | 4·207 | 408 | 167 | 509 |
| Az Zahirah | Wilayat Yanqul | 2·683 | 1·899 | 7·047 | 1·406 | 3·390 | 872 | 458 | 1104 |
| Dhofar | Wilayat Al Mazyunah | 2·832 | 2·622 | 9·434 | 1·174 | 3·568 | 256 | 99 | 321 |
| Dhofar | Wilayat Dalkut | 2·819 | 2·769 | 9·777 | 1·117 | 3·552 | 79 | 29 | 102 |
| Dhofar | Wilayat Mirbat | 2·785 | 2·328 | 8·462 | 1·280 | 3·529 | 593 | 275 | 754 |
| Dhofar | Wilayat Muqshin | 2·994 | 2·516 | 9·242 | 1·361 | 3·788 | 19 | 8 | 23 |
| Dhofar | Wilayat Rakhyut | 2·792 | 2·515 | 8·997 | 1·191 | 3·538 | 154 | 65 | 195 |
| Dhofar | Wilayat Sadah | 2·807 | 2·368 | 8·605 | 1·265 | 3·577 | 146 | 65 | 191 |
| Dhofar | Wilayat Salalah | 2·726 | 2·220 | 8·062 | 1·284 | 3·447 | 9887 | 4780 | 12619 |
| Dhofar | Wilayat Shalim Wa Juzor Al Hallaniyat | 2·794 | 2·193 | 8·027 | 1·349 | 3·536 | 174 | 81 | 230 |
| Dhofar | Wilayat Taqah | 2·739 | 2·180 | 7·942 | 1·315 | 3·471 | 714 | 340 | 898 |
| Dhofar | Wilayat Thumrayt | 2·735 | 2·124 | 7·819 | 1·337 | 3·454 | 580 | 280 | 748 |
| Muscat | Wilayat Al Amrat | 2·553 | 1·893 | 6·964 | 1·308 | 3·210 | 3636 | 1878 | 4570 |
| Muscat | Wilayat As Seeb | 2·503 | 1·834 | 6·733 | 1·273 | 3·180 | 14018 | 7096 | 17759 |
| Muscat | Wilayat Bawshar | 2·535 | 1·882 | 6·975 | 1·278 | 3·214 | 8745 | 4298 | 11088 |
| Muscat | Wilayat Muscat | 2·590 | 1·987 | 7·403 | 1·274 | 3·295 | 350 | 170 | 442 |
| Muscat | Wilayat Mutrah | 2·547 | 1·975 | 7·270 | 1·251 | 3·252 | 7914 | 3886 | 10170 |
| Muscat | Wilayat Qurayyat | 2·694 | 2·138 | 7·720 | 1·305 | 3·393 | 1642 | 786 | 2086 |

ADM1: administrative level 1. ADM2: administrative level 2. SD: standard deviation. CI95%: 95% credible interval. Q25: 25th percentile. Q75: 75th percentile. SD and CI95% are measures of prediction uncertainty.

**Supplementary Table S31. Subnational areal predictions of β-thalassaemia carrier prevalence and estimated carrier count at administrative level 2 of Pakistan.**

| ADM1 | ADM2 | β-thalassaemia carrier prevalence Mean | SD | CI95% | Q25 | Q75 | Estimated carrier counts Mean | Q25 | Q75 |
|---|---|---|---|---|---|---|---|---|---|
| Azad Kashmir | Azad Kashmir | 5·149 | 6·786 | 24·463 | 1·203 | 6·224 | 267568 | 63784 | 323668 |
| Balochistan | Awaran | 5·933 | 6·282 | 23·034 | 1·967 | 7·510 | 9311 | 2972 | 11682 |
| Balochistan | Barkhan | 6·045 | 8·072 | 29·532 | 1·302 | 7·302 | 9450 | 1996 | 11475 |
| Balochistan | Chagai | 4·000 | 5·551 | 19·280 | 0·927 | 4·724 | 12237 | 2782 | 14350 |
| Balochistan | Dera Bugti | 5·986 | 7·672 | 28·122 | 1·380 | 7·361 | 27312 | 6437 | 33414 |
| Balochistan | Gwadar | 4·577 | 5·249 | 18·936 | 1·378 | 5·719 | 6791 | 2007 | 8485 |
| Balochistan | Jafarabad | 5·996 | 7·432 | 26·601 | 1·514 | 7·426 | 59313 | 15030 | 73126 |
| Balochistan | Jhal Magsi | 5·887 | 7·461 | 27·100 | 1·479 | 7·183 | 13207 | 3364 | 16257 |
| Balochistan | Kachhi | 5·804 | 7·676 | 28·227 | 1·322 | 6·974 | 25991 | 5922 | 31432 |
| Balochistan | Kalat | 5·573 | 7·177 | 26·376 | 1·327 | 6·781 | 27525 | 6554 | 33375 |
| Balochistan | Kech | 4·303 | 5·214 | 18·770 | 1·212 | 5·284 | 46023 | 12882 | 56173 |
| Balochistan | Kharan | 4·985 | 6·454 | 23·157 | 1·245 | 6·025 | 20942 | 5068 | 25167 |
| Balochistan | Khuzdar | 6·012 | 6·842 | 25·215 | 1·766 | 7·544 | 55022 | 15263 | 69155 |
| Balochistan | Kohlu | 5·946 | 7·961 | 29·296 | 1·298 | 7·141 | 16621 | 3569 | 20074 |
| Balochistan | Lasbela | 6·426 | 5·772 | 21·305 | 2·663 | 8·196 | 45116 | 20479 | 57245 |
| Balochistan | Loralai | 5·915 | 8·072 | 29·792 | 1·224 | 7·011 | 35501 | 7177 | 41529 |
| Balochistan | Mastung | 5·541 | 7·522 | 27·390 | 1·208 | 6·628 | 16704 | 3638 | 19997 |
| Balochistan | Musakhel | 5·856 | 7·874 | 28·577 | 1·248 | 7·110 | 11577 | 2462 | 13773 |

| | | β-thalassaemia carrier prevalence | | | | | Estimated carrier counts | | |
|---|---|---|---|---|---|---|---|---|---|
| ADM1 | ADM2 | Mean | SD | CI95% | Q25 | Q75 | Mean | Q25 | Q75 |
| Balochistan | Nasirabad | 5·945 | 7·631 | 27·749 | 1·434 | 7·247 | 49490 | 12009 | 60619 |
| Balochistan | Nushki | 5·193 | 6·935 | 24·933 | 1·198 | 6·251 | 11358 | 2559 | 13505 |
| Balochistan | Panjgur | 4·932 | 5·911 | 21·205 | 1·369 | 6·122 | 18074 | 4864 | 22586 |
| Balochistan | Pishin | 5·581 | 7·890 | 28·977 | 1·130 | 6·510 | 37337 | 7693 | 43666 |
| Balochistan | Qilla Abdullah | 5·494 | 7·889 | 28·618 | 1·117 | 6·403 | 66682 | 13435 | 76830 |
| Balochistan | Qilla Saifullah | 5·802 | 8·169 | 29·924 | 1·166 | 6·778 | 21903 | 4374 | 25582 |
| Balochistan | Quetta | 5·478 | 7·586 | 27·391 | 1·168 | 6·555 | 150927 | 31670 | 181857 |
| Balochistan | Sibi | 5·763 | 7·816 | 28·730 | 1·225 | 6·848 | 13366 | 2815 | 15875 |
| Balochistan | Zhob | 5·811 | 8·180 | 29·580 | 1·177 | 6·830 | 32406 | 6652 | 38249 |
| Balochistan | Ziarat | 5·633 | 7·834 | 28·435 | 1·153 | 6·648 | 4627 | 943 | 5462 |
| Gilgit-Baltistan | Astore | 5·287 | 7·363 | 26·624 | 1·120 | 6·249 | 5983 | 1239 | 7071 |
| Gilgit-Baltistan | Diamer | 5·535 | 7·752 | 27·978 | 1·098 | 6·585 | 10332 | 2016 | 12340 |
| Gilgit-Baltistan | Ghanche | 5·046 | 7·285 | 24·958 | 1·105 | 5·888 | 4982 | 1105 | 5795 |
| Gilgit-Baltistan | Ghizer | 5·588 | 7·893 | 28·764 | 1·074 | 6·643 | 9146 | 1721 | 10872 |
| Gilgit-Baltistan | Gilgit | 5·575 | 7·921 | 28·774 | 1·085 | 6·547 | 7115 | 1359 | 8220 |
| Gilgit-Baltistan | Hunza | 5·594 | 7·986 | 28·755 | 1·058 | 6·649 | 4745 | 883 | 5674 |
| Gilgit-Baltistan | Kharmang | 4·984 | 7·021 | 24·063 | 1·125 | 5·895 | 3615 | 811 | 4207 |
| Gilgit-Baltistan | Nagar | 5·590 | 7·938 | 29·088 | 1·063 | 6·620 | 3453 | 639 | 4130 |
| Gilgit-Baltistan | Shigar | 5·341 | 7·667 | 27·136 | 1·086 | 6·306 | 3155 | 649 | 3699 |
| Gilgit-Baltistan | Skardu | 5·167 | 7·229 | 25·751 | 1·125 | 6·107 | 8497 | 1812 | 10050 |
| Islamabad Capital Territory | Islamabad Capital Territory | 5·312 | 7·152 | 26·183 | 1·199 | 6·337 | 218981 | 49339 | 263037 |
| Khyber Pakhtunkhwa | Abbottabad | 5·373 | 7·369 | 26·849 | 1·155 | 6·366 | 84280 | 18063 | 99752 |
| Khyber Pakhtunkhwa | Bajaur | 5·726 | 8·256 | 29·783 | 1·096 | 6·692 | 77010 | 14711 | 90313 |
| Khyber Pakhtunkhwa | Bannu | 5·650 | 7·734 | 28·389 | 1·178 | 6·719 | 49087 | 10203 | 58323 |
| Khyber Pakhtunkhwa | Battagram | 5·564 | 7·801 | 27·913 | 1·128 | 6·575 | 36728 | 7545 | 43120 |
| Khyber Pakhtunkhwa | Buner | 5·701 | 8·133 | 29·607 | 1·129 | 6·636 | 58502 | 11551 | 68297 |
| Khyber Pakhtunkhwa | Charsadda | 5·727 | 8·417 | 30·057 | 1·105 | 6·607 | 117497 | 22705 | 135331 |
| Khyber Pakhtunkhwa | Chitral | 5·536 | 7·641 | 27·533 | 1·091 | 6·670 | 30191 | 6004 | 36687 |
| Khyber Pakhtunkhwa | Dera Ismail Khan | 5·664 | 7·569 | 27·928 | 1·257 | 6·801 | 111370 | 24999 | 133142 |
| Khyber Pakhtunkhwa | Hangu | 5·711 | 8·047 | 29·642 | 1·138 | 6·728 | 16291 | 3230 | 19050 |
| Khyber Pakhtunkhwa | Haripur | 5·475 | 7·635 | 27·524 | 1·150 | 6·436 | 73934 | 15624 | 86894 |
| Khyber Pakhtunkhwa | Karak | 5·645 | 7·834 | 28·684 | 1·183 | 6·667 | 52257 | 10961 | 61951 |
| Khyber Pakhtunkhwa | Khyber | 5·667 | 8·122 | 29·236 | 1·118 | 6·662 | 60703 | 11842 | 71139 |
| Khyber Pakhtunkhwa | Kohat | 5·645 | 8·046 | 29·755 | 1·149 | 6·551 | 56079 | 11358 | 64588 |
| Khyber Pakhtunkhwa | Kohistan | 5·564 | 7·680 | 28·021 | 1·104 | 6·694 | 54349 | 10750 | 65048 |
| Khyber Pakhtunkhwa | Kurram | 5·698 | 7·891 | 28·834 | 1·131 | 6·855 | 46395 | 9305 | 56025 |
| Khyber Pakhtunkhwa | Lakki Marwat | 5·615 | 7·596 | 27·791 | 1·220 | 6·706 | 42978 | 9403 | 51309 |
| Khyber Pakhtunkhwa | Lower Dir | 5·639 | 8·076 | 28·601 | 1·100 | 6·617 | 104056 | 20243 | 121950 |
| Khyber Pakhtunkhwa | Malakand | 5·708 | 8·272 | 29·232 | 1·088 | 6·685 | 53044 | 9997 | 62358 |
| Khyber Pakhtunkhwa | Mansehra | 5·472 | 7·558 | 27·625 | 1·130 | 6·525 | 103276 | 21357 | 122793 |
| Khyber Pakhtunkhwa | Mardan | 5·707 | 8·286 | 29·748 | 1·109 | 6·582 | 165128 | 32181 | 189997 |
| Khyber Pakhtunkhwa | Mohmand | 5·747 | 8·364 | 30·043 | 1·094 | 6·711 | 53242 | 10325 | 62357 |
| Khyber Pakhtunkhwa | North Waziristan | 5·663 | 7·794 | 28·633 | 1·154 | 6·768 | 90504 | 18593 | 107388 |
| Khyber Pakhtunkhwa | Nowshera | 5·700 | 8·248 | 29·587 | 1·127 | 6·555 | 90162 | 17655 | 103384 |
| Khyber Pakhtunkhwa | Orakzai | 5·657 | 8·074 | 29·172 | 1·124 | 6·628 | 46129 | 9217 | 54255 |
| Khyber Pakhtunkhwa | Peshawar | 5·685 | 8·333 | 30·293 | 1·114 | 6·536 | 278755 | 55168 | 323055 |
| Khyber Pakhtunkhwa | Shangla | 5·593 | 7·788 | 28·592 | 1·113 | 6·639 | 43129 | 8595 | 50968 |
| Khyber Pakhtunkhwa | South Waziristan | 5·702 | 7·805 | 28·556 | 1·178 | 6·830 | 52093 | 10656 | 62463 |
| Khyber Pakhtunkhwa | Swabi | 5·651 | 8·061 | 29·462 | 1·136 | 6·550 | 109974 | 22064 | 127631 |
| Khyber Pakhtunkhwa | Swat | 5·573 | 7·779 | 28·246 | 1·097 | 6·651 | 153618 | 30244 | 180335 |
| Khyber Pakhtunkhwa | Tank | 5·674 | 7·667 | 27·961 | 1·212 | 6·775 | 22733 | 4805 | 26798 |
| Khyber Pakhtunkhwa | Upper Dir | 5·575 | 7·810 | 28·452 | 1·090 | 6·603 | 75587 | 14766 | 89212 |
| Punjab | Attock | 5·546 | 7·818 | 28·046 | 1·168 | 6·515 | 137350 | 28948 | 160608 |
| Punjab | Bahawalnagar | 5·352 | 6·572 | 24·082 | 1·461 | 6·506 | 201117 | 55635 | 243311 |
| Punjab | Bahawalpur | 5·681 | 7·154 | 26·144 | 1·410 | 6·980 | 280678 | 67527 | 342893 |
| Punjab | Bhakkar | 5·587 | 7·367 | 27·237 | 1·297 | 6·717 | 109589 | 25396 | 132032 |

| | | β-thalassaemia carrier prevalence | | | | | Estimated carrier counts | | |
|---|---|---|---|---|---|---|---|---|---|
| ADM1 | ADM2 | Mean | SD | CI95% | Q25 | Q75 | Mean | Q25 | Q75 |
| Punjab | Chakwal | 5·465 | 7·401 | 26·729 | 1·246 | 6·489 | 95083 | 21993 | 113141 |
| Punjab | Dera Ghazi Khan | 5·995 | 7·876 | 28·870 | 1·313 | 7·293 | 213377 | 46843 | 258079 |
| Punjab | Faisalabad | 5·205 | 6·362 | 23·157 | 1·437 | 6·354 | 492743 | 136415 | 600345 |
| Punjab | Gujranwala | 4·749 | 5·554 | 20·284 | 1·467 | 5·802 | 285735 | 88090 | 350741 |
| Punjab | Gujrat | 4·889 | 6·052 | 21·998 | 1·359 | 5·903 | 165560 | 46176 | 199893 |
| Punjab | Hafizabad | 5·000 | 6·123 | 22·188 | 1·405 | 6·112 | 78879 | 22420 | 96448 |
| Punjab | Jhang | 5·509 | 7·121 | 26·118 | 1·342 | 6·627 | 259988 | 63757 | 312281 |
| Punjab | Jhelum | 5·176 | 6·703 | 24·295 | 1·307 | 6·224 | 77298 | 19677 | 93147 |
| Punjab | Kasur | 4·695 | 4·908 | 17·869 | 1·631 | 5·901 | 184278 | 64075 | 231028 |
| Punjab | Khanewal | 5·710 | 7·395 | 27·345 | 1·351 | 6·898 | 211400 | 50141 | 254857 |
| Punjab | Khushab | 5·577 | 7·427 | 27·360 | 1·279 | 6·661 | 82123 | 19032 | 97771 |
| Punjab | Lahore | 4·577 | 4·731 | 17·265 | 1·636 | 5·748 | 615335 | 218904 | 767932 |
| Punjab | Layyah | 5·767 | 7·588 | 27·969 | 1·308 | 6·985 | 122990 | 27761 | 149145 |
| Punjab | Lodhran | 5·791 | 7·478 | 27·542 | 1·361 | 7·034 | 140238 | 33021 | 170016 |
| Punjab | Mandi Bahauddin | 5·072 | 6·398 | 23·114 | 1·364 | 6·136 | 95673 | 25676 | 115594 |
| Punjab | Mianwali | 5·633 | 7·705 | 28·427 | 1·230 | 6·640 | 95822 | 20902 | 112606 |
| Punjab | Multan | 5·910 | 7·755 | 28·538 | 1·352 | 7·145 | 333676 | 75592 | 406337 |
| Punjab | Muzaffargarh | 5·952 | 7·758 | 28·388 | 1·345 | 7·256 | 310251 | 70075 | 378253 |
| Punjab | Narowal | 4·465 | 4·882 | 17·654 | 1·502 | 5·544 | 99780 | 33671 | 123142 |
| Punjab | Okara | 4·987 | 5·683 | 20·630 | 1·536 | 6·173 | 174590 | 53121 | 216220 |
| Punjab | Pakpattan | 5·250 | 6·397 | 23·481 | 1·476 | 6·397 | 119701 | 33561 | 146035 |
| Punjab | Rahim Yar Khan | 5·772 | 6·954 | 25·416 | 1·446 | 7·305 | 321889 | 79701 | 406655 |
| Punjab | Rajanpur | 5·922 | 7·467 | 27·196 | 1·385 | 7·334 | 130523 | 30296 | 161093 |
| Punjab | Rawalpindi | 5·281 | 7·053 | 25·460 | 1·219 | 6·313 | 261497 | 59945 | 308854 |
| Punjab | Sahiwal | 5·359 | 6·690 | 24·516 | 1·425 | 6·499 | 152291 | 41049 | 183960 |
| Punjab | Sargodha | 5·370 | 6·910 | 25·389 | 1·338 | 6·460 | 235002 | 58852 | 283729 |
| Punjab | Sheikhpura | 4·788 | 5·383 | 19·352 | 1·531 | 5·952 | 307991 | 99220 | 384170 |
| Punjab | Sialkot | 4·651 | 5·402 | 19·590 | 1·442 | 5·696 | 215848 | 66244 | 265026 |
| Punjab | Toba Tek Singh | 5·496 | 7·017 | 25·857 | 1·366 | 6·643 | 140721 | 35065 | 170294 |
| Punjab | Vihari | 5·562 | 7·114 | 26·221 | 1·388 | 6·722 | 165674 | 41435 | 199651 |
| Sindh | Badin | 6·497 | 5·297 | 19·772 | 2·896 | 8·384 | 137359 | 60530 | 177502 |
| Sindh | Dadu | 6·362 | 6·739 | 24·554 | 2·091 | 8·093 | 122294 | 38841 | 154853 |
| Sindh | Ghotki | 5·961 | 7·069 | 25·442 | 1·533 | 7·613 | 105894 | 26432 | 134553 |
| Sindh | Hyderabad | 6·481 | 5·536 | 20·652 | 2·744 | 8·402 | 195987 | 83580 | 253266 |
| Sindh | Jacobabad | 6·009 | 7·299 | 26·462 | 1·530 | 7·546 | 63405 | 16093 | 79789 |
| Sindh | Jamshoro | 6·593 | 5·684 | 21·096 | 2·816 | 8·472 | 98842 | 40819 | 127219 |
| Sindh | Karachi | 6·721 | 4·835 | 17·941 | 3·428 | 8·582 | 1306608 | 686172 | 1673758 |
| Sindh | Kashmore | 5·976 | 7·265 | 26·480 | 1·472 | 7·495 | 82100 | 20258 | 103076 |
| Sindh | Khairpur | 6·172 | 6·754 | 24·406 | 1·883 | 7·910 | 167783 | 47735 | 213432 |
| Sindh | Matiari | 6·510 | 6·080 | 22·546 | 2·494 | 8·440 | 34930 | 13452 | 45353 |
| Sindh | Mirpurkhas | 6·183 | 5·603 | 20·780 | 2·466 | 7·960 | 100811 | 39973 | 129238 |
| Sindh | Naushehro Feroze | 6·369 | 6·793 | 24·784 | 2·039 | 8·168 | 132665 | 42013 | 169872 |
| Sindh | Nawabshah | 6·449 | 6·430 | 23·559 | 2·255 | 8·310 | 136414 | 47789 | 175911 |
| Sindh | Qambar Shahdadkot | 6·227 | 7·382 | 26·455 | 1·729 | 7·818 | 220465 | 60040 | 277028 |
| Sindh | Sanghar | 6·132 | 6·036 | 22·269 | 2·172 | 7·915 | 169948 | 61596 | 220396 |
| Sindh | Shikarpur | 6·096 | 7·256 | 26·383 | 1·575 | 7·714 | 86929 | 22361 | 110770 |
| Sindh | Sukkur | 6·077 | 7·045 | 25·359 | 1·649 | 7·755 | 102771 | 26850 | 128793 |
| Sindh | Tando Allahyar | 6·393 | 5·757 | 21·441 | 2·546 | 8·254 | 58520 | 23450 | 75706 |
| Sindh | Tando Muhammad Khan | 6·578 | 5·326 | 19·799 | 2·959 | 8·485 | 26490 | 11914 | 34206 |
| Sindh | Tharparkar | 5·350 | 4·638 | 16·965 | 2·289 | 6·851 | 99992 | 41904 | 128026 |
| Sindh | Thatta | 6·916 | 5·179 | 19·407 | 3·356 | 8·909 | 138260 | 67461 | 178083 |
| Sindh | Umerkot | 5·737 | 5·361 | 19·816 | 2·215 | 7·366 | 74279 | 28985 | 95196 |

ADM1: administrative level 1. ADM2: administrative level 2. SD: standard deviation. CI95%: 95% credible interval. Q25: 25$^{th}$ percentile. Q75: 75$^{th}$ percentile. SD and CI95% are measures of prediction uncertainty.

**Supplementary Table S32. Subnational areal predictions of β-thalassaemia carrier prevalence and estimated carrier count at administrative level 2 of Palestine.**

| | | β-thalassaemia carrier prevalence | | | | | Estimated carrier counts | | |
|---|---|---|---|---|---|---|---|---|---|
| ADM1 | ADM2 | Mean | SD | CI95% | Q25 | Q75 | Mean | Q25 | Q75 |
| Gaza | Deir Al Balah | 4·517 | 3·186 | 12·163 | 2·273 | 5·853 | 8593 | 4324 | 11134 |
| Gaza | Gaza | 4·716 | 3·505 | 13·249 | 2·345 | 6·019 | 40682 | 20314 | 52317 |
| Gaza | Khan Yunis | 4·638 | 3·465 | 12·523 | 2·314 | 5·941 | 23537 | 11714 | 30241 |
| Gaza | North Gaza | 4·573 | 3·297 | 12·184 | 2·345 | 5·752 | 15257 | 7844 | 19189 |
| Gaza | Rafah | 4·694 | 3·525 | 13·243 | 2·314 | 6·004 | 15613 | 7669 | 20145 |
| West Bank | Bethlehem | 3·439 | 2·570 | 9·490 | 1·730 | 4·351 | 7974 | 4038 | 10119 |
| West Bank | Hebron | 3·514 | 2·603 | 9·604 | 1·789 | 4·431 | 29679 | 15281 | 37585 |
| West Bank | Jenin | 4·582 | 3·197 | 11·937 | 2·411 | 5·794 | 16588 | 8700 | 20873 |
| West Bank | Jericho & Al Aghwar | 3·581 | 2·538 | 9·318 | 1·861 | 4·546 | 1897 | 982 | 2368 |
| West Bank | Jerusalem | 3·674 | 2·715 | 10·160 | 1·886 | 4·629 | 8378 | 4274 | 10544 |
| West Bank | Nablus | 4·139 | 2·943 | 10·855 | 2·163 | 5·221 | 18329 | 9662 | 23068 |
| West Bank | Qalqiliya | 4·642 | 3·287 | 12·212 | 2·422 | 5·866 | 4693 | 2450 | 5866 |
| West Bank | Ramallah & Al Bireh | 4·022 | 2·843 | 10·552 | 2·093 | 5·065 | 12878 | 6671 | 16099 |
| West Bank | Salfit | 4·393 | 3·084 | 11·637 | 2·332 | 5·501 | 4260 | 2257 | 5348 |
| West Bank | Tubas | 4·045 | 2·762 | 10·322 | 2·170 | 5·103 | 3145 | 1704 | 3976 |
| West Bank | Tulkarm | 4·874 | 3·529 | 12·970 | 2·521 | 6·149 | 11695 | 6051 | 14770 |

ADM1: administrative level 1. ADM2: administrative level 2. SD: standard deviation. CI95%: 95% credible interval. Q25: 25th percentile. Q75: 75th percentile. SD and CI95% are measures of prediction uncertainty.

**Supplementary Table S33. Subnational areal predictions of β-thalassaemia carrier prevalence and estimated carrier count at administrative level 2 of Saudi Arabia.**

| | | β-thalassaemia carrier prevalence | | | | | Estimated carrier counts | | |
|---|---|---|---|---|---|---|---|---|---|
| ADM1 | ADM2 | Mean | SD | CI95% | Q25 | Q75 | Mean | Q25 | Q75 |
| Al Bahah Region | Al Aqiq | 0·752 | 0·576 | 2·096 | 0·378 | 0·945 | 254 | 130 | 322 |
| Al Bahah Region | Al Bahah | 1·003 | 0·719 | 2·700 | 0·523 | 1·254 | 727 | 379 | 907 |
| Al Bahah Region | Al Hajrah | 1·568 | 1·148 | 4·181 | 0·814 | 1·970 | 140 | 71 | 177 |
| Al Bahah Region | Al Mandaq | 1·196 | 0·885 | 3·206 | 0·613 | 1·523 | 176 | 92 | 225 |
| Al Bahah Region | Al Mukhwah | 1·420 | 0·997 | 3·657 | 0·755 | 1·790 | 481 | 254 | 611 |
| Al Bahah Region | Al Qara | 1·045 | 0·791 | 2·889 | 0·527 | 1·319 | 187 | 93 | 240 |
| Al Bahah Region | Bani Hasan | 1·113 | 0·778 | 2·860 | 0·592 | 1·404 | 164 | 88 | 208 |
| Al Bahah Region | Biljurashi | 0·751 | 0·546 | 1·989 | 0·392 | 0·940 | 355 | 190 | 447 |
| Al Bahah Region | Qilwah | 1·499 | 1·069 | 3·936 | 0·790 | 1·883 | 367 | 194 | 466 |
| Al Jawf Region | Al Qurayyat | 0·772 | 0·622 | 2·246 | 0·375 | 0·966 | 2064 | 1011 | 2575 |
| Al Jawf Region | Dawamat Al Jandal | 0·201 | 0·176 | 0·616 | 0·092 | 0·250 | 114 | 52 | 143 |
| Al Jawf Region | Sakaka | 0·250 | 0·210 | 0·759 | 0·116 | 0·313 | 556 | 263 | 701 |
| Al Jawf Region | Tubarjal | 0·303 | 0·246 | 0·887 | 0·146 | 0·379 | 423 | 190 | 536 |
| Al Madinah Region | Al Hinakiyah | 0·190 | 0·169 | 0·599 | 0·085 | 0·236 | 62 | 27 | 87 |
| Al Madinah Region | Al Is | 0·314 | 0·254 | 0·909 | 0·153 | 0·393 | 63 | 27 | 80 |
| Al Madinah Region | Al Madinah Al Munawwarah | 0·325 | 0·263 | 0·948 | 0·157 | 0·406 | 5229 | 2574 | 6558 |
| Al Madinah Region | Al Mahd | 0·579 | 0·496 | 1·761 | 0·268 | 0·722 | 276 | 119 | 357 |
| Al Madinah Region | Al Ula | 0·339 | 0·276 | 0·992 | 0·163 | 0·425 | 203 | 96 | 260 |
| Al Madinah Region | Badr | 0·544 | 0·442 | 1·595 | 0·260 | 0·685 | 336 | 153 | 432 |
| Al Madinah Region | Khaybar | 0·141 | 0·118 | 0·426 | 0·066 | 0·177 | 64 | 30 | 80 |
| Al Madinah Region | Wadi Al Fara | 0·670 | 0·579 | 2·059 | 0·308 | 0·837 | 150 | 62 | 196 |
| Al Madinah Region | Yanbu | 0·475 | 0·377 | 1·374 | 0·231 | 0·598 | 1886 | 902 | 2360 |
| Al-Qassim Region | Al Asyah | 0·209 | 0·171 | 0·614 | 0·100 | 0·262 | 83 | 39 | 111 |
| Al-Qassim Region | Al Badai | 0·304 | 0·222 | 0·812 | 0·158 | 0·380 | 142 | 72 | 175 |
| Al-Qassim Region | Al Bukayriyah | 0·138 | 0·109 | 0·394 | 0·068 | 0·173 | 108 | 54 | 137 |
| Al-Qassim Region | Al Midhnab | 0·433 | 0·317 | 1·167 | 0·225 | 0·543 | 179 | 89 | 222 |
| Al-Qassim Region | An Nabhaniyah | 0·201 | 0·155 | 0·562 | 0·100 | 0·252 | 75 | 36 | 105 |
| Al-Qassim Region | Ar Rass | 0·333 | 0·253 | 0·924 | 0·168 | 0·416 | 321 | 162 | 400 |
| Al-Qassim Region | Ash Shimasiyah | 0·440 | 0·332 | 1·198 | 0·223 | 0·552 | 27 | 10 | 35 |
| Al-Qassim Region | Buraydah | 0·194 | 0·149 | 0·542 | 0·098 | 0·243 | 1598 | 807 | 2018 |

| | | β-thalassaemia carrier prevalence | | | | | Estimated carrier counts | | |
|---|---|---|---|---|---|---|---|---|---|
| ADM1 | ADM2 | Mean | SD | CI95% | Q25 | Q75 | Mean | Q25 | Q75 |
| Al-Qassim Region | Dariyah | 0·345 | 0·289 | 1·036 | 0·162 | 0·432 | 69 | 30 | 91 |
| Al-Qassim Region | Riyadh Al Khabra | 0·124 | 0·097 | 0·348 | 0·061 | 0·156 | 84 | 41 | 107 |
| Al-Qassim Region | Unayzah | 0·322 | 0·235 | 0·860 | 0·167 | 0·402 | 550 | 280 | 690 |
| Al-Qassim Region | Uqlat As Suqur | 0·111 | 0·093 | 0·332 | 0·052 | 0·138 | 15 | 5 | 18 |
| Al-Qassim Region | Uyun Al Jiwa | 0·131 | 0·102 | 0·368 | 0·065 | 0·165 | 45 | 23 | 59 |
| 'Asir Region | Abha | 1·612 | 1·124 | 4·150 | 0·857 | 2·038 | 6560 | 3523 | 8232 |
| 'Asir Region | Ahad Rufaydah | 1·212 | 0·841 | 3·107 | 0·653 | 1·520 | 1278 | 687 | 1607 |
| 'Asir Region | Al Harjah | 0·579 | 0·452 | 1·642 | 0·286 | 0·728 | 127 | 64 | 163 |
| 'Asir Region | Al Majardah | 1·005 | 0·700 | 2·560 | 0·538 | 1·258 | 429 | 227 | 537 |
| 'Asir Region | An Namas | 0·583 | 0·417 | 1·547 | 0·307 | 0·729 | 288 | 150 | 364 |
| 'Asir Region | Balqarn | 0·625 | 0·449 | 1·647 | 0·329 | 0·782 | 410 | 220 | 512 |
| 'Asir Region | Bariq | 1·177 | 0·819 | 3·028 | 0·634 | 1·480 | 567 | 301 | 709 |
| 'Asir Region | Bishah | 0·347 | 0·267 | 0·969 | 0·174 | 0·435 | 640 | 320 | 814 |
| 'Asir Region | Khamis Mushayt | 0·665 | 0·482 | 1·772 | 0·348 | 0·829 | 7045 | 3760 | 8805 |
| 'Asir Region | Muhayil | 2·094 | 1·448 | 5·351 | 1·120 | 2·638 | 4339 | 2356 | 5448 |
| 'Asir Region | Rijal Al Ma | 2·923 | 2·041 | 7·598 | 1·547 | 3·683 | 1261 | 671 | 1585 |
| 'Asir Region | Sarat Abidah | 0·970 | 0·695 | 2·564 | 0·510 | 1·219 | 570 | 293 | 713 |
| 'Asir Region | Tanumah | 0·648 | 0·464 | 1·737 | 0·343 | 0·812 | 126 | 66 | 157 |
| 'Asir Region | Tarib | 0·371 | 0·296 | 1·082 | 0·180 | 0·466 | 91 | 37 | 116 |
| 'Asir Region | Tathlith | 0·276 | 0·250 | 0·878 | 0·122 | 0·344 | 106 | 43 | 140 |
| 'Asir Region | Zahran Al Janub | 0·756 | 0·581 | 2·116 | 0·377 | 0·951 | 288 | 138 | 357 |
| Eastern Region | Abqaiq Governorate | 4·850 | 3·785 | 14·031 | 2·322 | 6·183 | 3211 | 1536 | 4098 |
| Eastern Region | Al Ahsa Governorate | 4·306 | 3·544 | 13·035 | 1·982 | 5·462 | 59527 | 27078 | 76232 |
| Eastern Region | Al Jubayl Governorate | 4·577 | 3·387 | 12·663 | 2·273 | 5·845 | 23069 | 11459 | 29568 |
| Eastern Region | Al Khafji Governorate | 3·822 | 3·207 | 11·695 | 1·724 | 4·865 | 2375 | 1032 | 2996 |
| Eastern Region | Al Khubar Governorate | 4·492 | 3·412 | 12·716 | 2·236 | 5·657 | 29072 | 14489 | 36822 |
| Eastern Region | Al Nuayriyah Governorate | 4·271 | 3·418 | 12·627 | 1·988 | 5·470 | 2984 | 1364 | 3818 |
| Eastern Region | Al Qatif Governorate | 4·354 | 3·273 | 12·160 | 2·175 | 5·475 | 26756 | 13303 | 33774 |
| Eastern Region | Al Udayd Governorate | 4·366 | 3·956 | 14·525 | 1·812 | 5·541 | 445 | 175 | 563 |
| Eastern Region | Hafar Al Batin Governorate | 1·240 | 0·979 | 3·571 | 0·600 | 1·566 | 4481 | 2215 | 5624 |
| Eastern Region | Qaryah Al Ulya Governorate | 2·729 | 2·309 | 8·479 | 1·225 | 3·468 | 645 | 280 | 839 |
| Eastern Region | Ras Tannurah Governorate | 4·322 | 3·300 | 12·226 | 2·146 | 5·406 | 1414 | 704 | 1783 |
| Eastern Region | Sharurah | 1·534 | 1·985 | 6·623 | 0·446 | 1·835 | 1030 | 287 | 1228 |
| Hayel Region | Al Ghazalah | 0·039 | 0·035 | 0·125 | 0·017 | 0·049 | 3 | 1 | 4 |
| Hayel Region | Al Hait | 0·058 | 0·049 | 0·176 | 0·027 | 0·072 | 27 | 6 | 37 |
| Hayel Region | As Sulaymi | 0·055 | 0·048 | 0·170 | 0·025 | 0·069 | 6 | 1 | 8 |
| Hayel Region | Ash Shamli | 0·048 | 0·042 | 0·148 | 0·022 | 0·060 | 8 | 3 | 9 |
| Hayel Region | Ash Shinan | 0·059 | 0·052 | 0·182 | 0·027 | 0·074 | 4 | 2 | 9 |
| Hayel Region | Baqa | 0·058 | 0·051 | 0·181 | 0·027 | 0·073 | 14 | 5 | 24 |
| Hayel Region | Hail | 0·039 | 0·035 | 0·123 | 0·017 | 0·048 | 188 | 75 | 236 |
| Hayel Region | Mawqaq | 0·036 | 0·032 | 0·112 | 0·016 | 0·044 | 2 | 0 | 2 |
| Hayel Region | Simira | 0·062 | 0·055 | 0·194 | 0·028 | 0·077 | 8 | 1 | 13 |
| Jazan Region | Abu Arish | 3·144 | 2·211 | 8·156 | 1·643 | 4·003 | 6139 | 3220 | 7806 |
| Jazan Region | Ad Dair | 1·501 | 1·098 | 4·043 | 0·774 | 1·881 | 1005 | 520 | 1253 |
| Jazan Region | Ad Darb | 3·771 | 2·632 | 9·797 | 1·971 | 4·801 | 2804 | 1476 | 3551 |
| Jazan Region | Ahad Al Masarihah | 3·445 | 2·449 | 9·192 | 1·779 | 4·356 | 4550 | 2339 | 5766 |
| Jazan Region | Al Aridah | 2·210 | 1·621 | 5·867 | 1·151 | 2·792 | 2285 | 1199 | 2885 |
| Jazan Region | Al Aydabi | 2·049 | 1·419 | 5·258 | 1·089 | 2·591 | 768 | 404 | 970 |
| Jazan Region | Al Harth | 2·428 | 1·795 | 6·497 | 1·230 | 3·082 | 451 | 230 | 574 |
| Jazan Region | Ar Rayth | 2·013 | 1·392 | 5·197 | 1·065 | 2·543 | 373 | 196 | 476 |
| Jazan Region | At Tuwal | 3·641 | 2·696 | 10·143 | 1·791 | 4·624 | 1384 | 684 | 1761 |
| Jazan Region | Baysh | 3·731 | 2·578 | 9·588 | 1·989 | 4·729 | 3023 | 1606 | 3788 |
| Jazan Region | Damad | 2·797 | 1·975 | 7·125 | 1·472 | 3·533 | 1921 | 993 | 2455 |
| Jazan Region | Farasan | 6·561 | 5·085 | 18·729 | 3·125 | 8·422 | 557 | 260 | 704 |
| Jazan Region | Fayfa | 1·847 | 1·329 | 4·926 | 0·962 | 2·302 | 353 | 182 | 439 |
| Jazan Region | Harub | 2·096 | 1·452 | 5·470 | 1·122 | 2·620 | 641 | 346 | 805 |
| Jazan Region | Jazan | 3·139 | 2·189 | 8·155 | 1·665 | 3·954 | 5850 | 3110 | 7384 |

| | | β-thalassaemia carrier prevalence | | | | | Estimated carrier counts | | |
|---|---|---|---|---|---|---|---|---|---|
| ADM1 | ADM2 | Mean | SD | CI95% | Q25 | Q75 | Mean | Q25 | Q75 |
| Jazan Region | Sabya | 3·351 | 2·293 | 8·570 | 1·780 | 4·259 | 8232 | 4378 | 10404 |
| Jazan Region | Samtah | 3·294 | 2·410 | 8·978 | 1·654 | 4·193 | 4703 | 2373 | 5999 |
| Makkah Region | Adam | 1·629 | 1·264 | 4·629 | 0·797 | 2·061 | 581 | 286 | 736 |
| Makkah Region | Al Ardiyat | 1·016 | 0·717 | 2·608 | 0·540 | 1·276 | 677 | 356 | 849 |
| Makkah Region | Al Birk | 3·035 | 2·169 | 8·012 | 1·576 | 3·850 | 724 | 374 | 917 |
| Makkah Region | Al Jumum | 1·304 | 0·951 | 3·499 | 0·677 | 1·639 | 2540 | 1301 | 3197 |
| Makkah Region | Al Kamil | 1·065 | 0·815 | 2·974 | 0·534 | 1·342 | 159 | 77 | 195 |
| Makkah Region | Al Khurmah | 0·754 | 0·640 | 2·302 | 0·349 | 0·944 | 315 | 143 | 400 |
| Makkah Region | Al Lith | 2·220 | 1·766 | 6·446 | 1·062 | 2·809 | 1712 | 807 | 2177 |
| Makkah Region | Al Qunfudhah | 2·307 | 1·601 | 5·935 | 1·231 | 2·912 | 5088 | 2702 | 6425 |
| Makkah Region | Al Taif | 0·994 | 0·774 | 2·815 | 0·492 | 1·248 | 14080 | 6892 | 17737 |
| Makkah Region | Bahrah | 1·955 | 1·533 | 5·590 | 0·951 | 2·475 | 2339 | 1183 | 2967 |
| Makkah Region | Farat Ghamid Az Zinad | 1·333 | 0·932 | 3·445 | 0·713 | 1·679 | 128 | 70 | 164 |
| Makkah Region | Jiddah | 1·573 | 1·198 | 4·380 | 0·784 | 1·987 | 60349 | 30531 | 75943 |
| Makkah Region | Khulays | 1·152 | 0·880 | 3·178 | 0·579 | 1·450 | 617 | 313 | 773 |
| Makkah Region | Makkah Al Mukarramah | 1·559 | 1·178 | 4·353 | 0·787 | 1·965 | 40664 | 20241 | 51370 |
| Makkah Region | Maysan | 1·252 | 0·976 | 3·585 | 0·615 | 1·577 | 411 | 202 | 522 |
| Makkah Region | Rabigh | 0·880 | 0·731 | 2·623 | 0·415 | 1·105 | 1116 | 521 | 1402 |
| Makkah Region | Ranyah | 0·494 | 0·445 | 1·553 | 0·219 | 0·618 | 261 | 110 | 329 |
| Makkah Region | Turubah | 0·860 | 0·683 | 2·479 | 0·418 | 1·080 | 380 | 176 | 476 |
| Najran Region | Badr Al Janub | 0·475 | 0·381 | 1·362 | 0·231 | 0·593 | 37 | 17 | 47 |
| Najran Region | Hubuna | 0·351 | 0·298 | 1·087 | 0·164 | 0·439 | 86 | 39 | 112 |
| Najran Region | Khubash | 0·339 | 0·393 | 1·242 | 0·127 | 0·410 | 26 | 10 | 30 |
| Najran Region | Najran | 0·375 | 0·333 | 1·162 | 0·173 | 0·464 | 1540 | 694 | 1897 |
| Najran Region | Thar | 0·274 | 0·246 | 0·861 | 0·122 | 0·340 | 36 | 15 | 46 |
| Najran Region | Yadamah | 0·277 | 0·267 | 0·926 | 0·116 | 0·343 | 44 | 15 | 51 |
| Northern Borders Region | Al Uwayqilah | 0·576 | 0·476 | 1·711 | 0·268 | 0·729 | 119 | 55 | 155 |
| Northern Borders Region | Arar | 0·775 | 0·752 | 2·641 | 0·317 | 0·965 | 1532 | 609 | 1870 |
| Northern Borders Region | Rafha | 0·364 | 0·330 | 1·178 | 0·157 | 0·456 | 345 | 146 | 434 |
| Northern Borders Region | Turayf | 0·886 | 0·883 | 3·117 | 0·360 | 1·087 | 615 | 259 | 772 |
| Riyadh Region | Ad Dilam | 2·283 | 2·025 | 7·285 | 1·000 | 2·870 | 1563 | 694 | 1942 |
| Riyadh Region | Ad Diriyah | 1·732 | 1·443 | 5·265 | 0·808 | 2·172 | 3372 | 1590 | 4220 |
| Riyadh Region | Ad Duwadimi | 0·867 | 0·706 | 2·558 | 0·411 | 1·093 | 1869 | 886 | 2373 |
| Riyadh Region | Afif | 0·639 | 0·596 | 2·101 | 0·276 | 0·792 | 493 | 198 | 622 |
| Riyadh Region | Al Aflaj | 1·995 | 1·980 | 6·930 | 0·784 | 2·500 | 1238 | 471 | 1550 |
| Riyadh Region | Al Ghat | 0·598 | 0·454 | 1·673 | 0·303 | 0·751 | 73 | 36 | 91 |
| Riyadh Region | Al Hariq | 2·015 | 1·964 | 6·951 | 0·815 | 2·506 | 250 | 101 | 310 |
| Riyadh Region | Al Kharj | 2·791 | 2·372 | 8·626 | 1·257 | 3·539 | 10422 | 4736 | 13189 |
| Riyadh Region | Al Majmaah | 0·903 | 0·707 | 2·574 | 0·445 | 1·136 | 1583 | 770 | 2001 |
| Riyadh Region | Al Muzahimiyah | 1·909 | 1·729 | 6·201 | 0·824 | 2·389 | 995 | 438 | 1262 |
| Riyadh Region | Al Quwayiyah | 1·344 | 1·275 | 4·482 | 0·560 | 1·682 | 1133 | 477 | 1421 |
| Riyadh Region | Ar Rayn | 1·487 | 1·548 | 5·343 | 0·570 | 1·840 | 471 | 164 | 585 |
| Riyadh Region | As Sulayyil | 0·824 | 0·802 | 2·838 | 0·335 | 1·024 | 261 | 105 | 330 |
| Riyadh Region | Az Zulfi | 0·431 | 0·346 | 1·244 | 0·213 | 0·538 | 403 | 196 | 496 |
| Riyadh Region | Duruma | 1·672 | 1·407 | 5·113 | 0·770 | 2·101 | 977 | 439 | 1229 |
| Riyadh Region | Hawtat Bani Tamim | 2·322 | 2·286 | 8·052 | 0·929 | 2·901 | 1097 | 448 | 1382 |
| Riyadh Region | Huraymila | 1·600 | 1·298 | 4·706 | 0·761 | 2·012 | 482 | 223 | 594 |
| Riyadh Region | Marat | 1·299 | 1·076 | 3·920 | 0·604 | 1·637 | 129 | 57 | 166 |
| Riyadh Region | Riyadh | 2·126 | 1·693 | 6·195 | 1·023 | 2·682 | 148684 | 70999 | 187437 |
| Riyadh Region | Rumah | 2·160 | 1·745 | 6·376 | 1·019 | 2·737 | 430 | 207 | 555 |
| Riyadh Region | Shaqra | 0·949 | 0·742 | 2·723 | 0·467 | 1·192 | 529 | 255 | 667 |
| Riyadh Region | Thadiq | 1·401 | 1·132 | 4·107 | 0·670 | 1·760 | 187 | 87 | 236 |
| Riyadh Region | Wadi Ad Dawasir | 0·767 | 0·807 | 2·801 | 0·288 | 0·948 | 500 | 184 | 627 |
| Tabuk Region | Al Bad | 1·523 | 1·759 | 5·867 | 0·526 | 1·873 | 265 | 88 | 333 |

| | | β-thalassaemia carrier prevalence | | | | | Estimated carrier counts | | |
|---|---|---|---|---|---|---|---|---|---|
| ADM1 | ADM2 | Mean | SD | CI95% | Q25 | Q75 | Mean | Q25 | Q75 |
| Tabuk Region | Al Wajh | 0·585 | 0·553 | 1·948 | 0·246 | 0·728 | 242 | 95 | 311 |
| Tabuk Region | Duba | 0·918 | 1·002 | 3·403 | 0·333 | 1·135 | 412 | 135 | 512 |
| Tabuk Region | Haqil | 1·350 | 1·400 | 4·849 | 0·516 | 1·677 | 288 | 102 | 361 |
| Tabuk Region | Tabuk | 0·662 | 0·633 | 2·201 | 0·278 | 0·825 | 4000 | 1752 | 4989 |
| Tabuk Region | Tayma | 0·242 | 0·195 | 0·706 | 0·116 | 0·304 | 94 | 40 | 115 |
| Tabuk Region | Umluj | 0·478 | 0·388 | 1·406 | 0·229 | 0·600 | 342 | 155 | 432 |

ADM1: administrative level 1. ADM2: administrative level 2. SD: standard deviation. CI95%: 95% credible interval. Q25: 25th percentile. Q75: 75th percentile. SD and CI95% are measures of prediction uncertainty.

**Supplementary Table S34. Subnational areal predictions of β-thalassaemia carrier prevalence and estimated carrier count at administrative level 2 of South Korea.**

| | | β-thalassaemia carrier prevalence | | | | | Estimated carrier counts | | |
|---|---|---|---|---|---|---|---|---|---|
| ADM1 | ADM2 | Mean | SD | CI95% | Q25 | Q75 | Mean | Q25 | Q75 |
| Busan | Buk-gu | 1·162 | 0·867 | 3·072 | 0·605 | 1·458 | 2696 | 1411 | 3369 |
| Busan | Busanjin-gu | 1·180 | 0·885 | 3·353 | 0·603 | 1·485 | 7502 | 3815 | 9443 |
| Busan | Dong-gu | 1·180 | 0·885 | 3·353 | 0·603 | 1·485 | 7502 | 3815 | 9443 |
| Busan | Dongnae-gu | 1·143 | 0·817 | 3·084 | 0·604 | 1·428 | 4308 | 2276 | 5381 |
| Busan | Haeundae-gu | 1·168 | 0·836 | 3·063 | 0·596 | 1·508 | 2828 | 1444 | 3652 |
| Busan | Nam-gu | 1·194 | 0·902 | 3·323 | 0·607 | 1·500 | 7895 | 4002 | 9920 |
| Busan | Saha-gu | 1·172 | 0·870 | 3·322 | 0·613 | 1·451 | 2501 | 1308 | 3094 |
| Busan | Sasang-gu | 1·158 | 0·838 | 3·246 | 0·588 | 1·447 | 1825 | 927 | 2282 |
| Busan | Seo-gu | 1·170 | 0·853 | 3·292 | 0·607 | 1·460 | 7540 | 3923 | 9415 |
| Busan | Suyeong-gu | 1·209 | 0·885 | 3·246 | 0·629 | 1·514 | 3607 | 1875 | 4516 |
| Busan | Yeonje-gu | 1·167 | 0·862 | 3·156 | 0·602 | 1·470 | 17200 | 8861 | 21657 |
| Daegu | Buk-gu | 1·257 | 1·020 | 3·794 | 0·590 | 1·595 | 4941 | 2344 | 6275 |
| Daegu | Dalseo-gu | 1·216 | 0·960 | 3·383 | 0·593 | 1·530 | 4558 | 2214 | 5762 |
| Daegu | Dong-gu | 1·285 | 1·055 | 3·761 | 0·604 | 1·598 | 384 | 182 | 474 |
| Daegu | Jung-gu | 1·252 | 1·043 | 3·871 | 0·592 | 1·559 | 4604 | 2175 | 5731 |
| Daegu | Nam-gu | 1·258 | 1·005 | 3·553 | 0·619 | 1·585 | 13884 | 6814 | 17498 |
| Daegu | Seo-gu | 1·286 | 1·047 | 3·600 | 0·632 | 1·653 | 3513 | 1726 | 4517 |
| Daegu | Suseong-gu | 1·258 | 0·993 | 3·657 | 0·617 | 1·579 | 8328 | 4127 | 10453 |
| Daejeon | Jung-gu | 1·231 | 1·053 | 3·860 | 0·555 | 1·535 | 4069 | 1859 | 5047 |
| Daejeon | Seo-gu | 1·221 | 0·984 | 3·621 | 0·570 | 1·539 | 4767 | 2222 | 5981 |
| Daejeon | Yuseong-gu | 1·216 | 1·008 | 3·584 | 0·577 | 1·527 | 5083 | 2422 | 6416 |
| Gangwon | Cheorwon-gun | 2·146 | 2·968 | 9·553 | 0·577 | 2·541 | 899 | 244 | 1066 |
| Gangwon | Chuncheon-si | 1·939 | 2·412 | 8·049 | 0·601 | 2·354 | 5586 | 1694 | 6751 |
| Gangwon | Donghae-si | 1·983 | 2·438 | 7·906 | 0·625 | 2·408 | 1284 | 406 | 1525 |
| Gangwon | Gangneung-si | 2·030 | 2·520 | 8·670 | 0·620 | 2·436 | 4165 | 1202 | 4721 |
| Gangwon | Goseong-gun | 2·368 | 3·360 | 11·349 | 0·608 | 2·727 | 496 | 128 | 560 |
| Gangwon | Hoengseong-gun | 1·802 | 2·048 | 7·145 | 0·603 | 2·232 | 779 | 265 | 966 |
| Gangwon | Hongcheon-gun | 1·928 | 2·366 | 7·877 | 0·606 | 2·350 | 1292 | 404 | 1527 |
| Gangwon | Hwacheon-gun | 2·082 | 2·751 | 9·142 | 0·595 | 2·472 | 480 | 139 | 575 |
| Gangwon | Inje-gun | 2·125 | 2·781 | 9·313 | 0·608 | 2·539 | 639 | 183 | 757 |
| Gangwon | Jeongseon-gun | 1·843 | 2·099 | 7·384 | 0·616 | 2·254 | 642 | 216 | 789 |
| Gangwon | Pyeongchang-gun | 1·897 | 2·256 | 7·688 | 0·616 | 2·318 | 739 | 242 | 907 |
| Gangwon | Samcheok-si | 1·868 | 2·160 | 7·596 | 0·619 | 2·271 | 1342 | 420 | 1627 |
| Gangwon | Sokcho-si | 2·250 | 2·989 | 10·518 | 0·626 | 2·705 | 1692 | 475 | 1978 |
| Gangwon | Taebaek-si | 1·802 | 1·974 | 7·137 | 0·628 | 2·215 | 734 | 275 | 924 |
| Gangwon | Uljin-gun | 1·720 | 1·849 | 6·355 | 0·618 | 2·144 | 577 | 210 | 737 |
| Gangwon | Wonju-si | 1·676 | 1·810 | 6·303 | 0·597 | 2·093 | 6056 | 2079 | 7351 |
| Gangwon | Yanggu-gun | 2·137 | 2·824 | 9·329 | 0·597 | 2·549 | 456 | 129 | 549 |
| Gangwon | Yangyang-gun | 2·133 | 2·775 | 9·357 | 0·619 | 2·558 | 695 | 198 | 826 |
| Gangwon | Yeongwol-gun | 1·746 | 1·928 | 6·808 | 0·607 | 2·143 | 635 | 224 | 794 |
| Gwangju | Buk-gu | 1·109 | 1·030 | 3·687 | 0·471 | 1·388 | 3062 | 1311 | 3810 |
| Gyeonggi | Ansan-si | 1·568 | 1·895 | 6·019 | 0·538 | 1·929 | 8502 | 2952 | 10545 |
| Gyeonggi | Anseong-si | 1·494 | 1·533 | 5·370 | 0·575 | 1·875 | 3824 | 1506 | 4715 |

| | | β-thalassaemia carrier prevalence | | | | | Estimated carrier counts | | |
|---|---|---|---|---|---|---|---|---|---|
| ADM1 | ADM2 | Mean | SD | CI95% | Q25 | Q75 | Mean | Q25 | Q75 |
| Gyeonggi | Anyang-si | 1·649 | 1·880 | 6·662 | 0·582 | 2·012 | 10766 | 3814 | 13150 |
| Gyeonggi | Bucheon-si | 1·708 | 2·232 | 6·754 | 0·565 | 1·975 | 15046 | 4985 | 17400 |
| Gyeonggi | Dongducheon-si | 1·879 | 2·372 | 8·041 | 0·567 | 2·227 | 1408 | 424 | 1639 |
| Gyeonggi | Eunpyeong-gu | 1·755 | 2·337 | 7·535 | 0·572 | 2·096 | 6334 | 2063 | 7567 |
| Gyeonggi | Gangdong-gu | 1·827 | 2·247 | 7·678 | 0·591 | 2·208 | 6244 | 2020 | 7549 |
| Gyeonggi | Ganghwa-gun | 1·801 | 2·462 | 7·961 | 0·537 | 2·076 | 1034 | 309 | 1194 |
| Gyeonggi | Gangseo-gu | 1·714 | 2·143 | 6·653 | 0·552 | 2·046 | 5499 | 1745 | 6668 |
| Gyeonggi | Gapyeong-gun | 1·898 | 2·382 | 7·806 | 0·583 | 2·297 | 1039 | 324 | 1267 |
| Gyeonggi | Gimpo-si | 1·790 | 2·356 | 8·196 | 0·534 | 2·103 | 6492 | 1959 | 7666 |
| Gyeonggi | Goyang-si | 1·791 | 2·299 | 7·596 | 0·552 | 2·124 | 19879 | 6133 | 23379 |
| Gyeonggi | Gunpo-si | 1·580 | 1·857 | 5·931 | 0·535 | 1·874 | 1512 | 513 | 1794 |
| Gyeonggi | Guri-si | 1·734 | 2·052 | 7·135 | 0·585 | 2·167 | 3864 | 1304 | 4828 |
| Gyeonggi | Gwacheon-si | 1·720 | 2·081 | 7·282 | 0·547 | 2·020 | 1592 | 506 | 1868 |
| Gyeonggi | Gwangju-si | 1·670 | 2·016 | 6·275 | 0·580 | 2·038 | 5462 | 1889 | 6547 |
| Gyeonggi | Gwangmyeong-si | 1·652 | 1·870 | 6·822 | 0·554 | 2·001 | 3886 | 1304 | 4708 |
| Gyeonggi | Gyeyang-gu | 1·732 | 1·975 | 6·571 | 0·563 | 2·197 | 2410 | 784 | 3056 |
| Gyeonggi | Hanam-si | 1·704 | 1·961 | 6·606 | 0·582 | 2·096 | 4971 | 1690 | 6141 |
| Gyeonggi | Hwaseong-si | 1·541 | 1·743 | 5·914 | 0·550 | 1·890 | 16653 | 5963 | 20474 |
| Gyeonggi | Icheon-si | 1·596 | 1·738 | 5·830 | 0·587 | 1·992 | 3381 | 1255 | 4202 |
| Gyeonggi | Namyangju-si | 1·797 | 2·192 | 7·218 | 0·583 | 2·182 | 10751 | 3465 | 13165 |
| Gyeonggi | Osan-si | 1·498 | 1·622 | 5·572 | 0·547 | 1·845 | 4638 | 1705 | 5712 |
| Gyeonggi | Paju-si | 1·920 | 2·660 | 8·349 | 0·552 | 2·242 | 8053 | 2289 | 9203 |
| Gyeonggi | Pocheon-si | 1·950 | 2·548 | 8·274 | 0·578 | 2·338 | 3056 | 919 | 3714 |
| Gyeonggi | Pyeongtaek-si | 1·468 | 1·539 | 5·264 | 0·550 | 1·817 | 6860 | 2614 | 8479 |
| Gyeonggi | Seo-gu | 1·767 | 2·368 | 7·381 | 0·543 | 2·112 | 13391 | 4082 | 15938 |
| Gyeonggi | Seongnam-si | 1·664 | 1·999 | 6·365 | 0·570 | 2·059 | 15543 | 5276 | 19386 |
| Gyeonggi | Siheung-si | 1·631 | 2·043 | 6·502 | 0·544 | 1·974 | 9329 | 3078 | 11345 |
| Gyeonggi | Suwon-si | 1·627 | 1·835 | 6·253 | 0·563 | 2·029 | 16878 | 5899 | 20785 |
| Gyeonggi | Uijeongbu-si | 1·799 | 2·217 | 7·809 | 0·563 | 2·126 | 7346 | 2293 | 8667 |
| Gyeonggi | Uiwang-si | 1·603 | 1·959 | 5·856 | 0·561 | 1·984 | 3701 | 1303 | 4595 |
| Gyeonggi | Yangju | 1·891 | 2·469 | 8·050 | 0·558 | 2·270 | 4611 | 1384 | 5551 |
| Gyeonggi | Yangpyeong-gun | 1·742 | 2·015 | 6·751 | 0·589 | 2·146 | 1903 | 641 | 2346 |
| Gyeonggi | Yeoju-si | 1·642 | 1·829 | 6·076 | 0·586 | 2·026 | 1926 | 677 | 2327 |
| Gyeonggi | Yeoncheon-gun | 2·022 | 2·758 | 9·198 | 0·565 | 2·366 | 824 | 229 | 940 |
| Gyeonggi | Yongin-si | 1·570 | 1·741 | 5·769 | 0·573 | 1·950 | 20015 | 7189 | 25012 |
| Incheon | Bupyeong-gu | 1·722 | 2·175 | 6·609 | 0·541 | 2·150 | 9502 | 2986 | 11864 |
| Incheon | Dong-gu | 1·710 | 2·327 | 6·756 | 0·524 | 2·040 | 14378 | 4478 | 17352 |
| Incheon | Jung-gu | 1·677 | 2·172 | 6·744 | 0·523 | 2·012 | 1545 | 479 | 1853 |
| Incheon | Michuhol-gu | 1·665 | 2·229 | 6·574 | 0·520 | 2·008 | 11719 | 3692 | 14244 |
| Incheon | Namdong-gu | 1·640 | 1·891 | 6·791 | 0·549 | 1·960 | 6433 | 2205 | 7742 |
| Jeju | Jeju | 1·143 | 1·393 | 4·767 | 0·367 | 1·368 | 4462 | 1471 | 5430 |
| Jeju | Seogwipo-si | 1·168 | 1·445 | 4·931 | 0·365 | 1·406 | 1789 | 553 | 2124 |
| North Chungcheong | Boeun-gun | 1·303 | 1·113 | 4·022 | 0·597 | 1·634 | 413 | 191 | 522 |
| North Chungcheong | Cheongju-si | 1·317 | 1·157 | 4·145 | 0·587 | 1·650 | 11540 | 5074 | 14411 |
| North Chungcheong | Chungju-si | 1·553 | 1·567 | 5·494 | 0·597 | 1·939 | 3338 | 1288 | 4198 |
| North Chungcheong | Daedeok-gu | 1·238 | 1·061 | 3·693 | 0·586 | 1·547 | 1383 | 676 | 1753 |
| North Chungcheong | Danyang-gun | 1·594 | 1·635 | 5·794 | 0·598 | 1·985 | 446 | 164 | 556 |
| North Chungcheong | Dong-gu | 1·227 | 1·086 | 3·672 | 0·582 | 1·519 | 3152 | 1478 | 3750 |
| North Chungcheong | Eumseong-gun | 1·473 | 1·455 | 5·126 | 0·585 | 1·833 | 1891 | 751 | 2376 |
| North Chungcheong | Goesan-gun | 1·416 | 1·327 | 4·725 | 0·599 | 1·783 | 496 | 209 | 628 |
| North Chungcheong | Jecheon-si | 1·609 | 1·685 | 5·819 | 0·600 | 2·012 | 2236 | 795 | 2747 |
| North Chungcheong | Jeungpyeong-gun | 1·395 | 1·370 | 4·392 | 0·597 | 1·745 | 568 | 240 | 719 |
| North Chungcheong | Jincheon-gun | 1·410 | 1·359 | 4·719 | 0·589 | 1·769 | 1027 | 423 | 1301 |
| North Chungcheong | Okcheon-gun | 1·241 | 0·996 | 3·715 | 0·594 | 1·561 | 603 | 293 | 761 |
| North Chungcheong | Yeongdong-gun | 1·230 | 1·004 | 3·600 | 0·590 | 1·540 | 609 | 294 | 767 |
| North Gyeongsang | Andong-si | 1·498 | 1·419 | 5·084 | 0·611 | 1·894 | 2468 | 1005 | 3153 |
| North Gyeongsang | Bonghwa-gun | 1·661 | 1·711 | 6·062 | 0·618 | 2·081 | 468 | 176 | 588 |

| | | β-thalassaemia carrier prevalence | | | | | Estimated carrier counts | | |
|---|---|---|---|---|---|---|---|---|---|
| ADM1 | ADM2 | Mean | SD | CI95% | Q25 | Q75 | Mean | Q25 | Q75 |
| North Gyeongsang | Cheongdo-gun | 1·226 | 0·956 | 3·491 | 0·603 | 1·543 | 483 | 238 | 609 |
| North Gyeongsang | Cheongsong-gun | 1·450 | 1·302 | 4·765 | 0·615 | 1·840 | 353 | 151 | 447 |
| North Gyeongsang | Chilgok-gun | 1·251 | 1·020 | 3·705 | 0·589 | 1·574 | 1183 | 557 | 1504 |
| North Gyeongsang | Dalseong-gun | 1·210 | 0·947 | 3·494 | 0·599 | 1·518 | 3721 | 1840 | 4677 |
| North Gyeongsang | Gimcheon-si | 1·231 | 0·996 | 3·602 | 0·594 | 1·536 | 1739 | 844 | 2196 |
| North Gyeongsang | Goryeong-gun | 1·198 | 0·948 | 3·465 | 0·591 | 1·491 | 286 | 141 | 350 |
| North Gyeongsang | Gumi-si | 1·303 | 1·068 | 3·926 | 0·610 | 1·652 | 5820 | 2786 | 7416 |
| North Gyeongsang | Gunwi-gun | 1·337 | 1·147 | 4·157 | 0·604 | 1·681 | 303 | 138 | 381 |
| North Gyeongsang | Gyeongju-si | 1·331 | 1·093 | 4·024 | 0·626 | 1·678 | 3663 | 1731 | 4595 |
| North Gyeongsang | Gyeongsan-si | 1·266 | 0·993 | 3·659 | 0·617 | 1·590 | 3568 | 1732 | 4480 |
| North Gyeongsang | Mungyeong-si | 1·449 | 1·354 | 4·895 | 0·604 | 1·818 | 758 | 322 | 953 |
| North Gyeongsang | Pohang-si | 1·425 | 1·263 | 4·528 | 0·621 | 1·796 | 5829 | 2590 | 7336 |
| North Gyeongsang | Sangju-si | 1·336 | 1·165 | 4·210 | 0·601 | 1·678 | 1532 | 678 | 1931 |
| North Gyeongsang | Seongju-gun | 1·220 | 0·967 | 3·489 | 0·594 | 1·534 | 505 | 240 | 638 |
| North Gyeongsang | Uiseong-gun | 1·382 | 1·208 | 4·387 | 0·609 | 1·743 | 681 | 300 | 853 |
| North Gyeongsang | Ulleung-gun | 2·382 | 3·022 | 9·933 | 0·647 | 2·812 | 75 | 20 | 89 |
| North Gyeongsang | Yecheon-gun | 1·473 | 1·410 | 5·090 | 0·602 | 1·840 | 659 | 273 | 829 |
| North Gyeongsang | Yeongcheon-si | 1·321 | 1·090 | 4·012 | 0·616 | 1·671 | 1313 | 614 | 1651 |
| North Gyeongsang | Yeongdeok-gun | 1·550 | 1·474 | 5·333 | 0·624 | 1·953 | 451 | 180 | 567 |
| North Gyeongsang | Yeongju-si | 1·585 | 1·597 | 5·603 | 0·606 | 1·993 | 1659 | 642 | 2121 |
| North Gyeongsang | Yeongyang-gun | 1·604 | 1·620 | 5·669 | 0·613 | 2·008 | 261 | 100 | 326 |
| North Jeolla | Buan-gun | 1·108 | 0·986 | 3·546 | 0·487 | 1·389 | 475 | 215 | 592 |
| North Jeolla | Gimje-si | 1·118 | 0·947 | 3·427 | 0·513 | 1·408 | 907 | 420 | 1142 |
| North Jeolla | Gochang-gun | 1·086 | 0·990 | 3·593 | 0·466 | 1·359 | 595 | 255 | 734 |
| North Jeolla | Gunsan-si | 1·151 | 1·014 | 3·618 | 0·513 | 1·444 | 2745 | 1236 | 3457 |
| North Jeolla | Iksan-si | 1·144 | 0·966 | 3·501 | 0·536 | 1·435 | 3197 | 1532 | 4014 |
| North Jeolla | Imsil-gun | 1·114 | 0·947 | 3·391 | 0·516 | 1·391 | 309 | 141 | 386 |
| North Jeolla | Jangsu-gun | 1·132 | 0·907 | 3·321 | 0·538 | 1·427 | 227 | 107 | 293 |
| North Jeolla | Jeongeup-si | 1·093 | 0·931 | 3·360 | 0·498 | 1·374 | 1164 | 531 | 1441 |
| North Jeolla | Jeonju-si | 1·111 | 0·870 | 3·295 | 0·538 | 1·403 | 7131 | 3471 | 9071 |
| North Jeolla | Jinan-gun | 1·150 | 0·928 | 3·369 | 0·549 | 1·448 | 267 | 127 | 340 |
| North Jeolla | Muju-gun | 1·188 | 0·962 | 3·463 | 0·572 | 1·493 | 265 | 130 | 336 |
| North Jeolla | Namwon-si | 1·117 | 0·948 | 3·400 | 0·513 | 1·406 | 864 | 398 | 1089 |
| North Jeolla | Sunchang-gun | 1·079 | 0·928 | 3·393 | 0·494 | 1·354 | 266 | 120 | 334 |
| North Jeolla | Wanju-gun | 1·129 | 0·913 | 3·324 | 0·547 | 1·409 | 1314 | 634 | 1638 |
| Sejong | Sejong | 1·272 | 1·095 | 3·954 | 0·580 | 1·593 | 4430 | 2021 | 5564 |
| Seoul | Dobong-gu | 1·765 | 2·194 | 6·475 | 0·559 | 2·149 | 5705 | 1807 | 6945 |
| Seoul | Dongdaemun-gu | 1·739 | 2·167 | 6·865 | 0·560 | 2·139 | 41332 | 13286 | 50705 |
| Seoul | Dongjak-gu | 1·673 | 2·220 | 6·787 | 0·545 | 1·979 | 8366 | 2724 | 9896 |
| Seoul | Gangbuk-gu | 1·775 | 2·221 | 7·273 | 0·556 | 2·134 | 17556 | 5578 | 21261 |
| Seoul | Gangnam-gu | 1·634 | 1·938 | 6·105 | 0·552 | 2·066 | 6744 | 2279 | 8531 |
| Seoul | Geumcheon-gu | 1·643 | 1·981 | 6·991 | 0·537 | 2·010 | 5894 | 1927 | 7209 |
| Seoul | Guro-gu | 1·703 | 2·115 | 6·942 | 0·556 | 2·010 | 38594 | 12442 | 44944 |
| Seoul | Gwanak-gu | 1·682 | 2·009 | 6·920 | 0·568 | 2·031 | 2086 | 705 | 2519 |
| Seoul | Gwangjin-gu | 1·679 | 2·045 | 6·691 | 0·570 | 2·059 | 7687 | 2610 | 9425 |
| Seoul | Jongno-gu | 1·789 | 2·406 | 6·629 | 0·558 | 2·126 | 1167 | 364 | 1387 |
| Seoul | Jung-gu | 1·780 | 2·220 | 7·346 | 0·567 | 2·290 | 6915 | 2201 | 8896 |
| Seoul | Jungnang-gu | 1·744 | 2·143 | 6·950 | 0·573 | 2·139 | 33465 | 11005 | 41054 |
| Seoul | Mapo-gu | 1·751 | 2·208 | 7·322 | 0·557 | 2·072 | 8243 | 2623 | 9753 |
| Seoul | Nowon-gu | 1·825 | 2·477 | 7·022 | 0·574 | 2·159 | 8172 | 2565 | 9795 |
| Seoul | Seocho-gu | 1·680 | 1·931 | 7·128 | 0·565 | 2·099 | 7283 | 2410 | 9148 |
| Seoul | Seodaemun-gu | 1·701 | 2·031 | 6·553 | 0·570 | 2·015 | 6831 | 2291 | 8094 |
| Seoul | Seongbuk-gu | 1·751 | 2·192 | 6·835 | 0·560 | 2·151 | 40874 | 13085 | 50289 |
| Seoul | Seongdong-gu | 1·683 | 1·903 | 6·238 | 0·557 | 2·095 | 8923 | 2955 | 11109 |
| Seoul | Songpa-gu | 1·675 | 1·957 | 6·660 | 0·550 | 2·059 | 14796 | 4851 | 18191 |
| Seoul | Yangcheon-gu | 1·754 | 2·165 | 7·581 | 0·559 | 2·008 | 11769 | 3752 | 13471 |
| Seoul | Yeongdeungpo-gu | 1·770 | 2·232 | 7·181 | 0·567 | 2·041 | 11566 | 3707 | 13338 |

| | | β-thalassaemia carrier prevalence | | | | | Estimated carrier counts | | |
|---|---|---|---|---|---|---|---|---|---|
| ADM1 | ADM2 | Mean | SD | CI95% | Q25 | Q75 | Mean | Q25 | Q75 |
| Seoul | Yongsan-gu | 1·702 | 2·094 | 6·861 | 0·556 | 2·091 | 28284 | 9244 | 34657 |
| South Chungcheong | Asan-si | 1·369 | 1·327 | 4·628 | 0·562 | 1·704 | 6035 | 2486 | 7643 |
| South Chungcheong | Boryeong-si | 1·227 | 1·174 | 4·070 | 0·508 | 1·534 | 1127 | 459 | 1397 |
| South Chungcheong | Buyeo-gun | 1·191 | 1·046 | 3·729 | 0·534 | 1·493 | 697 | 318 | 869 |
| South Chungcheong | Cheonan-si | 1·364 | 1·300 | 4·457 | 0·576 | 1·709 | 7817 | 3381 | 9719 |
| South Chungcheong | Cheongyang-gun | 1·225 | 1·112 | 3·915 | 0·542 | 1·535 | 437 | 196 | 553 |
| South Chungcheong | Dangjin-si | 1·414 | 1·557 | 5·285 | 0·526 | 1·729 | 2250 | 835 | 2768 |
| South Chungcheong | Geumsan-gun | 1·185 | 0·944 | 3·430 | 0·576 | 1·487 | 602 | 298 | 762 |
| South Chungcheong | Gongju-si | 1·232 | 1·042 | 3·829 | 0·562 | 1·545 | 1340 | 618 | 1673 |
| South Chungcheong | Gyeryong-si | 1·207 | 1·014 | 3·714 | 0·566 | 1·514 | 347 | 160 | 428 |
| South Chungcheong | Hongseong-gun | 1·291 | 1·262 | 4·514 | 0·521 | 1·594 | 1234 | 499 | 1522 |
| South Chungcheong | Nonsan-si | 1·169 | 0·945 | 3·431 | 0·554 | 1·468 | 1460 | 685 | 1819 |
| South Chungcheong | Seocheon-gun | 1·174 | 1·058 | 3·747 | 0·514 | 1·473 | 513 | 222 | 644 |
| South Chungcheong | Seosan-si | 1·375 | 1·484 | 5·007 | 0·506 | 1·701 | 2238 | 824 | 2829 |
| South Chungcheong | Taean-gun | 1·361 | 1·505 | 5·033 | 0·498 | 1·660 | 659 | 247 | 801 |
| South Chungcheong | Yesan-gun | 1·319 | 1·282 | 4·495 | 0·544 | 1·632 | 1107 | 454 | 1357 |
| South Gyeongsang | Changnyeong-gun | 1·167 | 0·900 | 3·214 | 0·587 | 1·463 | 850 | 433 | 1071 |
| South Gyeongsang | Changwon-si | 1·143 | 0·852 | 3·128 | 0·578 | 1·441 | 10852 | 5524 | 13748 |
| South Gyeongsang | Gangseo-gu | 1·152 | 0·874 | 3·021 | 0·593 | 1·446 | 2382 | 1246 | 3022 |
| South Gyeongsang | Geochang-gun | 1·165 | 0·919 | 3·351 | 0·569 | 1·471 | 683 | 345 | 844 |
| South Gyeongsang | Geoje-si | 1·134 | 0·884 | 3·187 | 0·561 | 1·422 | 2592 | 1292 | 3204 |
| South Gyeongsang | Geumjeong-gu | 1·211 | 0·950 | 3·368 | 0·646 | 1·473 | 1223 | 668 | 1487 |
| South Gyeongsang | Gijang-gun | 1·214 | 0·931 | 3·413 | 0·612 | 1·517 | 2333 | 1172 | 2924 |
| South Gyeongsang | Gimhae-si | 1·173 | 0·876 | 3·169 | 0·602 | 1·470 | 5392 | 2742 | 6734 |
| South Gyeongsang | Goseong-gun | 1·115 | 0·857 | 3·118 | 0·555 | 1·408 | 560 | 280 | 700 |
| South Gyeongsang | Hadong-gun | 1·112 | 0·928 | 3·308 | 0·526 | 1·392 | 400 | 189 | 508 |
| South Gyeongsang | Haman-gun | 1·139 | 0·875 | 3·145 | 0·579 | 1·422 | 1537 | 795 | 1920 |
| South Gyeongsang | Hamyang-gun | 1·135 | 0·908 | 3·287 | 0·547 | 1·437 | 424 | 212 | 541 |
| South Gyeongsang | Hapcheon-gun | 1·167 | 0·919 | 3·326 | 0·573 | 1·467 | 512 | 252 | 640 |
| South Gyeongsang | Jinju-si | 1·140 | 0·913 | 3·218 | 0·553 | 1·438 | 3953 | 1922 | 5080 |
| South Gyeongsang | Miryang-si | 1·199 | 0·903 | 3·301 | 0·604 | 1·512 | 1213 | 611 | 1531 |
| South Gyeongsang | Namhae-gun | 1·147 | 1·022 | 3·598 | 0·512 | 1·438 | 253 | 114 | 320 |
| South Gyeongsang | Sacheon-si | 1·130 | 0·952 | 3·377 | 0·527 | 1·409 | 727 | 330 | 919 |
| South Gyeongsang | Sancheong-gun | 1·136 | 0·926 | 3·301 | 0·545 | 1·432 | 360 | 172 | 451 |
| South Gyeongsang | Tongyeong-si | 1·135 | 0·926 | 3·244 | 0·547 | 1·434 | 812 | 390 | 997 |
| South Gyeongsang | Uiryeong-gun | 1·157 | 0·910 | 3·302 | 0·573 | 1·450 | 285 | 141 | 360 |
| South Gyeongsang | Yangsan-si | 1·200 | 0·892 | 3·305 | 0·615 | 1·505 | 3972 | 2021 | 4955 |
| South Jeolla | Boseong-gun | 1·110 | 1·068 | 3·723 | 0·461 | 1·372 | 404 | 168 | 509 |
| South Jeolla | Damyang-gun | 1·097 | 0·983 | 3·578 | 0·479 | 1·373 | 559 | 242 | 706 |
| South Jeolla | Dong-gu | 1·076 | 1·019 | 3·305 | 0·459 | 1·331 | 3715 | 1574 | 4552 |
| South Jeolla | Gangjin-gun | 1·102 | 1·194 | 4·052 | 0·423 | 1·348 | 318 | 122 | 388 |
| South Jeolla | Goheung-gun | 1·119 | 1·119 | 3·733 | 0·455 | 1·392 | 475 | 192 | 594 |
| South Jeolla | Gokseong-gun | 1·102 | 0·959 | 3·482 | 0·494 | 1·388 | 310 | 141 | 393 |
| South Jeolla | Gurye-gun | 1·121 | 0·962 | 3·395 | 0·514 | 1·397 | 260 | 118 | 332 |
| South Jeolla | Gwangsan-gu | 1·098 | 1·063 | 3·691 | 0·463 | 1·362 | 4562 | 1962 | 5658 |
| South Jeolla | Gwangyang-si | 1·118 | 0·980 | 3·498 | 0·507 | 1·396 | 1583 | 700 | 1958 |
| South Jeolla | Haenam-gun | 1·076 | 1·147 | 3·978 | 0·398 | 1·322 | 582 | 216 | 712 |
| South Jeolla | Hampyeong-gun | 1·076 | 1·073 | 3·807 | 0·433 | 1·324 | 264 | 107 | 321 |
| South Jeolla | Hwasun-gun | 1·103 | 1·053 | 3·684 | 0·460 | 1·367 | 670 | 278 | 816 |
| South Jeolla | Jangheung-gun | 1·085 | 1·074 | 3·791 | 0·432 | 1·345 | 357 | 138 | 443 |
| South Jeolla | Jangseong-gun | 1·084 | 0·978 | 3·574 | 0·473 | 1·347 | 403 | 172 | 499 |
| South Jeolla | Jindo-gun | 1·068 | 1·185 | 4·113 | 0·371 | 1·322 | 240 | 82 | 300 |
| South Jeolla | Mokpo-si | 1·066 | 1·134 | 4·279 | 0·395 | 1·307 | 1981 | 732 | 2432 |
| South Jeolla | Muan-gun | 1·081 | 1·130 | 3·898 | 0·411 | 1·325 | 1020 | 378 | 1245 |
| South Jeolla | Naju-si | 1·085 | 1·089 | 3·864 | 0·435 | 1·345 | 1297 | 518 | 1617 |
| South Jeolla | Nam-gu | 1·098 | 1·128 | 3·810 | 0·451 | 1·339 | 1498 | 614 | 1836 |
| South Jeolla | Seo-gu | 1·118 | 1·152 | 3·960 | 0·453 | 1·351 | 3407 | 1424 | 4130 |

| ADM1 | ADM2 | β-thalassaemia carrier prevalence Mean | SD | CI95% | Q25 | Q75 | Estimated carrier counts Mean | Q25 | Q75 |
|---|---|---|---|---|---|---|---|---|---|
| South Jeolla | Suncheon-si | 1·099 | 0·972 | 3·482 | 0·488 | 1·380 | 2984 | 1336 | 3744 |
| South Jeolla | Wando-gun | 1·110 | 1·231 | 4·468 | 0·401 | 1·349 | 164 | 57 | 197 |
| South Jeolla | Yeongam-gun | 1·084 | 1·122 | 3·910 | 0·420 | 1·342 | 558 | 214 | 686 |
| South Jeolla | Yeonggwang-gun | 1·062 | 1·028 | 3·623 | 0·443 | 1·325 | 480 | 201 | 583 |
| South Jeolla | Yeosu-si | 1·145 | 1·078 | 3·744 | 0·495 | 1·420 | 2848 | 1211 | 3495 |
| Ulsan | Buk-gu | 1·299 | 0·994 | 3·750 | 0·622 | 1·661 | 2042 | 991 | 2623 |
| Ulsan | Dong-gu | 1·332 | 1·144 | 3·876 | 0·611 | 1·717 | 273 | 125 | 351 |
| Ulsan | Jung-gu | 1·297 | 1·039 | 3·675 | 0·635 | 1·611 | 4519 | 2207 | 5598 |
| Ulsan | Nam-gu | 1·288 | 1·047 | 3·695 | 0·629 | 1·595 | 2149 | 1051 | 2665 |
| Ulsan | Ulju-gun | 1·247 | 0·963 | 3·510 | 0·615 | 1·573 | 3659 | 1787 | 4623 |

ADM1: administrative level 1. ADM2: administrative level 2. SD: standard deviation. CI95%: 95% credible interval. Q25: 25th percentile. Q75: 75th percentile. SD and CI95% are measures of prediction uncertainty.

**Supplementary Table S35. Subnational areal predictions of β-thalassaemia carrier prevalence and estimated carrier count at administrative level 2 of Sri Lanka.**

| ADM1 | ADM2 | β-thalassaemia carrier prevalence Mean | SD | CI95% | Q25 | Q75 | Estimated carrier counts Mean | Q25 | Q75 |
|---|---|---|---|---|---|---|---|---|---|
| Central Province | Kandy District | 6·281 | 5·118 | 18·976 | 2·802 | 8·143 | 97091 | 43863 | 125898 |
| Central Province | Matale District | 6·289 | 5·070 | 18·669 | 2·849 | 8·131 | 31240 | 14433 | 40364 |
| Central Province | Nuwara Eliya District | 6·330 | 5·455 | 20·104 | 2·646 | 8·200 | 48412 | 20259 | 62725 |
| Eastern Province | Ampara District | 6·386 | 6·162 | 22·702 | 2·358 | 8·240 | 41733 | 15400 | 54196 |
| Eastern Province | Batticaloa District | 6·400 | 5·999 | 22·064 | 2·423 | 8·322 | 32840 | 11896 | 42449 |
| Eastern Province | Trincomalee District | 6·349 | 5·918 | 21·804 | 2·413 | 8·277 | 23923 | 9055 | 31112 |
| North Central Province | Anuradhapura District | 6·327 | 5·564 | 20·668 | 2·616 | 8·144 | 71601 | 30326 | 92319 |
| North Central Province | Polonnaruwa District | 6·330 | 5·543 | 20·397 | 2·624 | 8·195 | 28787 | 11979 | 37156 |
| North Western Province | Kurunegala District | 6·242 | 4·798 | 17·846 | 2·948 | 8·062 | 126969 | 60138 | 164108 |
| North Western Province | Puttalam District | 6·233 | 5·111 | 18·912 | 2·763 | 8·054 | 43554 | 19566 | 56484 |
| Northern Province | Jaffna District | 6·041 | 6·443 | 23·530 | 1·991 | 7·634 | 34540 | 11466 | 43359 |
| Northern Province | Kilinochchi District | 6·140 | 6·268 | 23·098 | 2·117 | 7·845 | 8065 | 2819 | 10331 |
| Northern Province | Mannar District | 6·258 | 5·936 | 21·881 | 2·380 | 8·041 | 4855 | 1834 | 6277 |
| Northern Province | Mullaitivu District | 6·265 | 6·177 | 22·863 | 2·218 | 8·065 | 6852 | 2398 | 8854 |
| Northern Province | Vavuniya District | 6·292 | 5·862 | 21·889 | 2·410 | 8·126 | 11455 | 4474 | 14780 |
| Sabaragamuwa Province | Kegalle District | 6·263 | 5·035 | 18·632 | 2·838 | 8·100 | 55589 | 25516 | 71831 |
| Sabaragamuwa Province | Ratnapura District | 6·411 | 5·958 | 21·897 | 2·505 | 8·261 | 85830 | 33615 | 110550 |
| Southern Province | Galle District | 6·375 | 6·230 | 22·546 | 2·368 | 8·176 | 68275 | 24970 | 87635 |
| Southern Province | Hambantota District | 6·394 | 6·577 | 24·052 | 2·213 | 8·106 | 38022 | 13387 | 48448 |
| Southern Province | Matara District | 6·359 | 6·347 | 23·514 | 2·327 | 8·100 | 44546 | 16148 | 56771 |
| Uva Province | Badulla District | 6·331 | 5·661 | 20·939 | 2·552 | 8·162 | 57419 | 23166 | 74120 |
| Uva Province | Monaragala District | 6·359 | 6·139 | 22·601 | 2·350 | 8·175 | 44092 | 16456 | 56908 |
| Western Province | Colombo District | 6·357 | 5·561 | 20·499 | 2·632 | 8·222 | 116282 | 48214 | 149836 |
| Western Province | Gampaha District | 6·287 | 5·185 | 19·115 | 2·786 | 8·101 | 170507 | 74523 | 219861 |
| Western Province | Kalutara District | 6·363 | 5·839 | 21·510 | 2·515 | 8·202 | 82414 | 32921 | 105881 |

ADM1: administrative level 1. ADM2: administrative level 2. SD: standard deviation. CI95%: 95% credible interval. Q25: 25th percentile. Q75: 75th percentile. SD and CI95% are measures of prediction uncertainty.

**Supplementary Table S36. Subnational areal predictions of β-thalassaemia carrier prevalence and estimated carrier count at administrative level 2 of Thailand.**

| ADM1 | ADM2 | β-thalassaemia carrier prevalence Mean | SD | CI95% | Q25 | Q75 | Estimated carrier counts Mean | Q25 | Q75 |
|---|---|---|---|---|---|---|---|---|---|
| Amnat Charoen Province | Chanuman | 1·094 | 1·206 | 3·995 | 0·403 | 1·353 | 307 | 111 | 381 |
| Amnat Charoen Province | Hua Taphan | 0·855 | 0·779 | 2·861 | 0·357 | 1·076 | 235 | 98 | 299 |
| Amnat Charoen Province | Lue Amnat | 0·858 | 0·787 | 2·865 | 0·359 | 1·097 | 197 | 81 | 252 |
| Amnat Charoen Province | Mueang Amnat Charoen | 0·942 | 0·893 | 3·193 | 0·389 | 1·177 | 715 | 291 | 891 |
| Amnat Charoen Province | Pathum Ratchawongsa | 0·971 | 0·981 | 3·451 | 0·381 | 1·203 | 304 | 121 | 379 |
| Amnat Charoen Province | Phana | 0·862 | 0·859 | 3·025 | 0·354 | 1·066 | 122 | 52 | 152 |

| | | β-thalassaemia carrier prevalence | | | | | Estimated carrier counts | | |
|---|---|---|---|---|---|---|---|---|---|
| ADM1 | ADM2 | Mean | SD | CI95% | Q25 | Q75 | Mean | Q25 | Q75 |
| Amnat Charoen Province | Senangkhanikhom | 1·045 | 1·060 | 3·556 | 0·414 | 1·301 | 264 | 107 | 328 |
| Ang Thong Province | Chaiyo | 1·823 | 1·488 | 5·649 | 0·836 | 2·421 | 207 | 95 | 274 |
| Ang Thong Province | Mueang Ang Thong | 1·776 | 1·437 | 5·377 | 0·820 | 2·246 | 958 | 439 | 1209 |
| Ang Thong Province | Pa Mok | 1·779 | 1·513 | 5·336 | 0·807 | 2·246 | 644 | 290 | 820 |
| Ang Thong Province | Pho Thong | 1·849 | 1·546 | 5·629 | 0·820 | 2·368 | 697 | 311 | 893 |
| Ang Thong Province | Samko | 1·831 | 1·605 | 5·826 | 0·820 | 2·236 | 250 | 112 | 305 |
| Ang Thong Province | Sawaeng Ha | 1·923 | 1·728 | 6·093 | 0·853 | 2·406 | 266 | 118 | 334 |
| Ang Thong Province | Wiset Chai Chan | 1·811 | 1·653 | 5·665 | 0·787 | 2·284 | 882 | 383 | 1111 |
| Bangkok | Bang Bon | 1·668 | 1·374 | 4·926 | 0·762 | 2·123 | 8827 | 4018 | 11206 |
| Bangkok | Bang Kapi | 1·640 | 1·340 | 4·632 | 0·768 | 2·110 | 7830 | 3668 | 10073 |
| Bangkok | Bang Khae | 1·548 | 1·316 | 4·645 | 0·723 | 1·958 | 2587 | 1209 | 3272 |
| Bangkok | Bang Khen | 1·655 | 1·297 | 4·804 | 0·778 | 2·090 | 7819 | 3669 | 9878 |
| Bangkok | Bang Kho Laem | 1·615 | 1·337 | 4·965 | 0·769 | 1·974 | 7345 | 3494 | 8975 |
| Bangkok | Bang Khun Thian | 1·624 | 1·317 | 5·058 | 0·771 | 2·040 | 6491 | 3090 | 7998 |
| Bangkok | Bang Na | 1·630 | 1·319 | 4·779 | 0·787 | 2·045 | 27111 | 13124 | 33969 |
| Bangkok | Bang Phlat | 1·676 | 1·424 | 4·886 | 0·752 | 2·145 | 4849 | 2177 | 6206 |
| Bangkok | Bang Rak | 1·630 | 1·336 | 4·893 | 0·775 | 2·048 | 13000 | 6183 | 16342 |
| Bangkok | Bang Sue | 1·651 | 1·350 | 4·855 | 0·767 | 2·086 | 25798 | 12053 | 32561 |
| Bangkok | Bangkok Noi | 1·601 | 1·280 | 4·735 | 0·765 | 2·030 | 9621 | 4619 | 12283 |
| Bangkok | Bangkok Yai | 1·583 | 1·231 | 4·507 | 0·774 | 2·072 | 5809 | 2843 | 7606 |
| Bangkok | Bueng Kum | 1·618 | 1·278 | 4·470 | 0·813 | 2·033 | 5229 | 2627 | 6570 |
| Bangkok | Chatuchak | 1·632 | 1·258 | 4·720 | 0·787 | 2·053 | 8851 | 4268 | 11130 |
| Bangkok | Chom Thong | 1·613 | 1·279 | 4·653 | 0·776 | 2·059 | 4402 | 2118 | 5617 |
| Bangkok | Din Daeng | 1·631 | 1·289 | 4·698 | 0·782 | 2·051 | 16652 | 7985 | 20942 |
| Bangkok | Don Mueang | 1·672 | 1·357 | 5·057 | 0·777 | 2·103 | 8043 | 3736 | 10114 |
| Bangkok | Dusit | 1·653 | 1·388 | 4·998 | 0·763 | 2·096 | 10048 | 4643 | 12736 |
| Bangkok | Huai Khwang | 1·624 | 1·337 | 4·605 | 0·773 | 2·045 | 36290 | 17260 | 45676 |
| Bangkok | Khan Na Yao | 1·636 | 1·293 | 4·563 | 0·771 | 2·070 | 4536 | 2139 | 5739 |
| Bangkok | Khlong Sam Wa | 1·679 | 1·355 | 4·843 | 0·804 | 2·077 | 6978 | 3364 | 8797 |
| Bangkok | Khlong San | 1·610 | 1·300 | 4·809 | 0·773 | 2·038 | 22755 | 10921 | 28728 |
| Bangkok | Khlong Toei | 1·636 | 1·339 | 4·909 | 0·776 | 2·034 | 28485 | 13467 | 35488 |
| Bangkok | Lak Si | 1·608 | 1·237 | 4·723 | 0·752 | 2·042 | 6411 | 2999 | 8142 |
| Bangkok | Lat Krabang | 1·655 | 1·334 | 4·965 | 0·798 | 2·073 | 7269 | 3542 | 9075 |
| Bangkok | Lat Phrao | 1·602 | 1·445 | 4·262 | 0·757 | 2·034 | 9202 | 4347 | 11685 |
| Bangkok | Min Buri | 1·678 | 1·383 | 4·967 | 0·804 | 2·126 | 5585 | 2664 | 7086 |
| Bangkok | Nong Chok | 1·686 | 1·329 | 4·932 | 0·823 | 2·141 | 4175 | 2043 | 5272 |
| Bangkok | Nong Khaem | 1·632 | 1·400 | 4·821 | 0·765 | 2·029 | 7331 | 3434 | 9109 |
| Bangkok | Pathum Wan | 1·630 | 1·352 | 5·110 | 0·773 | 2·048 | 5199 | 2466 | 6530 |
| Bangkok | Phasi Charoen | 1·620 | 1·284 | 4·581 | 0·769 | 2·077 | 19559 | 9274 | 25089 |
| Bangkok | Phaya Thai | 1·642 | 1·338 | 4·848 | 0·772 | 2·073 | 26700 | 12628 | 33678 |
| Bangkok | Phra Khanong | 1·691 | 1·484 | 5·472 | 0·784 | 2·080 | 7023 | 3257 | 8636 |
| Bangkok | Phra Nakhon | 1·630 | 1·336 | 4·835 | 0·767 | 2·088 | 15857 | 7486 | 20342 |
| Bangkok | Pom Prap Sattru Phai | 1·630 | 1·352 | 5·110 | 0·773 | 2·048 | 5199 | 2466 | 6530 |
| Bangkok | Prawet | 1·621 | 1·243 | 4·566 | 0·806 | 2·061 | 6826 | 3372 | 8662 |
| Bangkok | Rat Burana | 1·624 | 1·337 | 4·766 | 0·779 | 2·014 | 15623 | 7480 | 19338 |
| Bangkok | Ratchathewi | 1·645 | 1·365 | 4·891 | 0·767 | 2·080 | 17849 | 8360 | 22548 |
| Bangkok | Sai Mai | 1·624 | 1·414 | 5·139 | 0·790 | 1·961 | 3513 | 1708 | 4242 |
| Bangkok | Samphanthawong | 1·630 | 1·352 | 5·110 | 0·773 | 2·048 | 5199 | 2466 | 6530 |
| Bangkok | Saphan Sung | 1·636 | 1·320 | 5·111 | 0·804 | 2·012 | 3157 | 1551 | 3884 |
| Bangkok | Sathon | 1·616 | 1·309 | 4·869 | 0·772 | 2·009 | 23570 | 11242 | 29308 |
| Bangkok | Suan Luang | 1·635 | 1·324 | 4·763 | 0·772 | 2·044 | 10436 | 4928 | 13049 |
| Bangkok | Taling Chan | 1·626 | 1·245 | 4·506 | 0·775 | 2·079 | 3560 | 1696 | 4550 |
| Bangkok | Thawi Watthana | 1·600 | 1·263 | 4·739 | 0·764 | 2·028 | 4360 | 2094 | 5492 |
| Bangkok | Thon Buri | 1·610 | 1·300 | 4·809 | 0·773 | 2·038 | 22755 | 10921 | 28728 |
| Bangkok | Thung Khru | 1·614 | 1·304 | 4·802 | 0·759 | 2·049 | 18078 | 8631 | 22442 |
| Bangkok | Vadhana | 1·629 | 1·320 | 4·676 | 0·776 | 2·049 | 7801 | 3717 | 9812 |
| Bangkok | Wang Thonglang | 1·624 | 1·347 | 4·532 | 0·777 | 2·055 | 32697 | 15570 | 41377 |

| ADM1 | ADM2 | β-thalassaemia carrier prevalence Mean | SD | CI95% | Q25 | Q75 | Estimated carrier counts Mean | Q25 | Q75 |
|---|---|---|---|---|---|---|---|---|---|
| Bangkok | Yan Nawa | 1·611 | 1·295 | 4·789 | 0·772 | 1·996 | 18371 | 8776 | 22778 |
| Bueng Kan Province | Bueng Khong Long | 2·418 | 2·916 | 10·214 | 0·758 | 2·925 | 774 | 244 | 937 |
| Bueng Kan Province | Bung Khla | 2·640 | 3·209 | 11·661 | 0·815 | 3·129 | 232 | 72 | 277 |
| Bueng Kan Province | Mueang Bueng Kan | 2·636 | 3·114 | 10·916 | 0·818 | 3·210 | 2123 | 661 | 2564 |
| Bueng Kan Province | Pak Khat | 2·582 | 2·952 | 10·287 | 0·828 | 3·238 | 600 | 193 | 748 |
| Bueng Kan Province | Phon Charoen | 2·412 | 2·817 | 9·895 | 0·789 | 2·914 | 851 | 278 | 1038 |
| Bueng Kan Province | Seka | 2·348 | 2·749 | 9·358 | 0·767 | 2·876 | 1615 | 525 | 1970 |
| Bueng Kan Province | Si Wilai | 2·496 | 2·970 | 9·508 | 0·804 | 3·052 | 577 | 185 | 694 |
| Bueng Kan Province | So Phisai | 2·486 | 2·854 | 9·957 | 0·829 | 3·071 | 1370 | 458 | 1692 |
| Buri Ram Province | Ban Dan | 1·125 | 1·012 | 3·603 | 0·487 | 1·397 | 196 | 85 | 246 |
| Buri Ram Province | Ban Kruat | 1·019 | 0·866 | 3·055 | 0·466 | 1·293 | 458 | 207 | 582 |
| Buri Ram Province | Ban Mai Chaiyaphot | 1·342 | 1·122 | 4·163 | 0·597 | 1·698 | 273 | 121 | 346 |
| Buri Ram Province | Chaloem Phra Kiat | 1·210 | 1·084 | 4·062 | 0·529 | 1·526 | 268 | 119 | 339 |
| Buri Ram Province | Chamni | 1·255 | 1·124 | 4·057 | 0·549 | 1·561 | 297 | 129 | 368 |
| Buri Ram Province | Huai Rat | 1·082 | 0·953 | 3·382 | 0·478 | 1·367 | 320 | 140 | 402 |
| Buri Ram Province | Khaen Dong | 1·174 | 1·038 | 3·829 | 0·520 | 1·475 | 255 | 114 | 323 |
| Buri Ram Province | Khu Mueang | 1·196 | 1·035 | 3·755 | 0·531 | 1·507 | 506 | 225 | 637 |
| Buri Ram Province | Krasang | 1·023 | 0·893 | 3·231 | 0·458 | 1·292 | 704 | 317 | 892 |
| Buri Ram Province | Lahan Sai | 1·161 | 0·991 | 3·558 | 0·527 | 1·465 | 616 | 279 | 778 |
| Buri Ram Province | Lam Plai Mat | 1·249 | 1·090 | 3·965 | 0·557 | 1·574 | 1028 | 458 | 1298 |
| Buri Ram Province | Mueang Buri Ram | 1·150 | 1·004 | 3·642 | 0·512 | 1·447 | 1750 | 777 | 2199 |
| Buri Ram Province | Na Pho | 1·360 | 1·224 | 4·461 | 0·595 | 1·681 | 268 | 119 | 331 |
| Buri Ram Province | Nang Rong | 1·271 | 1·127 | 4·086 | 0·558 | 1·587 | 1250 | 544 | 1567 |
| Buri Ram Province | Non Din Daeng | 1·235 | 1·022 | 3·692 | 0·577 | 1·560 | 251 | 115 | 321 |
| Buri Ram Province | Non Suwan | 1·371 | 1·162 | 4·409 | 0·621 | 1·732 | 275 | 126 | 353 |
| Buri Ram Province | Nong Hong | 1·344 | 1·178 | 4·101 | 0·607 | 1·678 | 416 | 188 | 519 |
| Buri Ram Province | Nong Ki | 1·390 | 1·221 | 4·412 | 0·617 | 1·753 | 655 | 289 | 828 |
| Buri Ram Province | Pakham | 1·290 | 1·076 | 3·985 | 0·593 | 1·633 | 325 | 148 | 412 |
| Buri Ram Province | Phlapphla Chai | 1·067 | 0·914 | 3·282 | 0·482 | 1·349 | 268 | 119 | 341 |
| Buri Ram Province | Phutthaisong | 1·289 | 1·148 | 4·038 | 0·569 | 1·618 | 373 | 166 | 470 |
| Buri Ram Province | Prakhon Chai | 1·098 | 0·957 | 3·404 | 0·494 | 1·382 | 865 | 393 | 1091 |
| Buri Ram Province | Satuek | 1·084 | 0·991 | 3·536 | 0·469 | 1·364 | 609 | 264 | 760 |
| Chachoengsao Province | Ban Pho | 1·699 | 1·323 | 4·803 | 0·824 | 2·161 | 889 | 432 | 1125 |
| Chachoengsao Province | Bang Khla | 1·743 | 1·350 | 4·963 | 0·849 | 2·199 | 986 | 480 | 1238 |
| Chachoengsao Province | Bang Nam Priao | 1·738 | 1·369 | 4·990 | 0·840 | 2·205 | 1345 | 650 | 1702 |
| Chachoengsao Province | Bang Pakong | 1·675 | 1·342 | 4·875 | 0·807 | 2·108 | 2679 | 1286 | 3392 |
| Chachoengsao Province | Khlong Khuean | 1·712 | 1·354 | 5·090 | 0·847 | 2·112 | 85 | 42 | 104 |
| Chachoengsao Province | Mueang Chachoengsao | 1·715 | 1·367 | 5·012 | 0·826 | 2·156 | 3176 | 1535 | 3987 |
| Chachoengsao Province | Phanom Sarakham | 1·751 | 1·341 | 4·896 | 0·888 | 2·197 | 1345 | 682 | 1685 |
| Chachoengsao Province | Plaeng Yao | 1·719 | 1·331 | 4·942 | 0·857 | 2·163 | 1040 | 522 | 1313 |
| Chachoengsao Province | Ratchasan | 1·743 | 1·328 | 4·826 | 0·862 | 2·177 | 153 | 76 | 192 |
| Chachoengsao Province | Sanam Chai Khet | 1·717 | 1·292 | 4·721 | 0·863 | 2·170 | 878 | 442 | 1107 |
| Chachoengsao Province | Tha Takiap | 1·639 | 1·252 | 4·603 | 0·815 | 2·054 | 545 | 270 | 681 |
| Chai Nat Province | Hankha | 2·074 | 1·892 | 6·700 | 0·883 | 2·603 | 830 | 354 | 1036 |
| Chai Nat Province | Manorom | 2·099 | 1·841 | 6·724 | 0·914 | 2·614 | 636 | 276 | 794 |
| Chai Nat Province | Mueang Chai Nat | 2·079 | 1·826 | 6·591 | 0·913 | 2·600 | 1046 | 455 | 1301 |
| Chai Nat Province | Noen Kham | 2·120 | 1·905 | 6·936 | 0·909 | 2·691 | 271 | 116 | 343 |
| Chai Nat Province | Nong Mamong | 2·223 | 2·063 | 7·415 | 0·924 | 2·794 | 339 | 138 | 425 |
| Chai Nat Province | Sankhaburi | 1·986 | 1·769 | 6·544 | 0·853 | 2·538 | 885 | 379 | 1132 |
| Chai Nat Province | Sapphaya | 2·031 | 1·857 | 6·819 | 0·847 | 2·538 | 725 | 299 | 911 |
| Chai Nat Province | Wat Sing | 2·149 | 1·973 | 7·020 | 0·914 | 2·706 | 410 | 174 | 517 |
| Chaiyaphum Province | Bamnet Narong | 1·812 | 1·621 | 5·798 | 0·798 | 2·283 | 710 | 313 | 899 |
| Chaiyaphum Province | Ban Khwao | 1·780 | 1·544 | 5·618 | 0·802 | 2·208 | 589 | 265 | 725 |
| Chaiyaphum Province | Ban Thaen | 1·674 | 1·381 | 4·964 | 0·788 | 2·104 | 601 | 284 | 755 |
| Chaiyaphum Province | Chatturat | 1·759 | 1·533 | 5·578 | 0·775 | 2·212 | 873 | 386 | 1098 |
| Chaiyaphum Province | Kaeng Khro | 1·687 | 1·433 | 5·161 | 0·779 | 2·105 | 1306 | 603 | 1616 |
| Chaiyaphum Province | Kaset Sombun | 1·835 | 1·614 | 5·829 | 0·824 | 2·303 | 1301 | 594 | 1633 |

| ADM1 | ADM2 | β-thalassaemia carrier prevalence Mean | SD | CI95% | Q25 | Q75 | Estimated carrier counts Mean | Q25 | Q75 |
|---|---|---|---|---|---|---|---|---|---|
| Chaiyaphum Province | Khon San | 1·918 | 1·658 | 5·884 | 0·870 | 2·409 | 712 | 314 | 880 |
| Chaiyaphum Province | Khon Sawan | 1·658 | 1·433 | 5·085 | 0·759 | 2·061 | 649 | 298 | 809 |
| Chaiyaphum Province | Mueang Chaiyaphum | 1·729 | 1·498 | 5·439 | 0·776 | 2·160 | 2261 | 1006 | 2812 |
| Chaiyaphum Province | Nong Bua Daeng | 1·874 | 1·579 | 5·752 | 0·856 | 2·371 | 1271 | 576 | 1615 |
| Chaiyaphum Province | Nong Bua Rawe | 1·841 | 1·555 | 5·726 | 0·837 | 2·317 | 517 | 234 | 656 |
| Chaiyaphum Province | Phakdi Chumphon | 1·889 | 1·610 | 5·829 | 0·857 | 2·356 | 297 | 135 | 364 |
| Chaiyaphum Province | Phu Khiao | 1·747 | 1·513 | 5·338 | 0·800 | 2·203 | 1402 | 642 | 1773 |
| Chaiyaphum Province | Sap Yai | 1·852 | 1·641 | 5·864 | 0·810 | 2·330 | 274 | 119 | 345 |
| Chaiyaphum Province | Thep Sathit | 1·868 | 1·626 | 6·004 | 0·827 | 2·351 | 845 | 374 | 1060 |
| Chaiyaphum Province | Thepharak | 1·828 | 1·614 | 5·669 | 0·814 | 2·308 | 273 | 121 | 347 |
| Chanthaburi Province | Kaeng Hang Maeo | 1·489 | 1·207 | 4·358 | 0·706 | 1·880 | 544 | 259 | 690 |
| Chanthaburi Province | Khao Khitchakut | 1·360 | 1·086 | 3·979 | 0·647 | 1·715 | 334 | 156 | 422 |
| Chanthaburi Province | Khlung | 1·233 | 1·074 | 3·870 | 0·554 | 1·560 | 571 | 253 | 717 |
| Chanthaburi Province | Laem Sing | 1·334 | 1·164 | 4·252 | 0·583 | 1·685 | 269 | 119 | 335 |
| Chanthaburi Province | Makham | 1·260 | 1·044 | 3·752 | 0·586 | 1·595 | 424 | 196 | 533 |
| Chanthaburi Province | Mueang Chanthaburi | 1·304 | 1·120 | 3·998 | 0·587 | 1·651 | 1358 | 609 | 1728 |
| Chanthaburi Province | Na Yai Am | 1·461 | 1·258 | 4·419 | 0·662 | 1·837 | 357 | 162 | 449 |
| Chanthaburi Province | Pong Nam Ron | 1·224 | 0·972 | 3·504 | 0·589 | 1·544 | 554 | 269 | 703 |
| Chanthaburi Province | Soi Dao | 1·324 | 1·064 | 3·910 | 0·637 | 1·664 | 848 | 410 | 1066 |
| Chanthaburi Province | Tha Mai | 1·411 | 1·228 | 4·421 | 0·629 | 1·776 | 820 | 362 | 1040 |
| Chiang Mai Province | Chai Prakan | 4·439 | 5·430 | 18·672 | 1·310 | 5·431 | 1697 | 499 | 2063 |
| Chiang Mai Province | Chiang Dao | 4·563 | 5·490 | 19·185 | 1·345 | 5·610 | 3891 | 1153 | 4741 |
| Chiang Mai Province | Chom Thong | 4·483 | 4·897 | 17·276 | 1·573 | 5·508 | 3474 | 1220 | 4267 |
| Chiang Mai Province | Doi Lo | 4·351 | 4·756 | 16·616 | 1·515 | 5·462 | 1549 | 541 | 1945 |
| Chiang Mai Province | Doi Saket | 4·234 | 4·947 | 16·580 | 1·373 | 5·181 | 2151 | 723 | 2693 |
| Chiang Mai Province | Doi Tao | 4·274 | 4·348 | 15·436 | 1·608 | 5·353 | 977 | 362 | 1220 |
| Chiang Mai Province | Fang | 4·453 | 5·456 | 18·494 | 1·258 | 5·522 | 5417 | 1549 | 6872 |
| Chiang Mai Province | Galyani Vadhana | 4·866 | 5·478 | 19·860 | 1·526 | 6·090 | 503 | 155 | 621 |
| Chiang Mai Province | Hang Dong | 4·414 | 4·999 | 17·661 | 1·447 | 5·442 | 3669 | 1210 | 4483 |
| Chiang Mai Province | Hot | 4·556 | 4·725 | 16·953 | 1·657 | 5·705 | 1574 | 570 | 1962 |
| Chiang Mai Province | Mae Ai | 4·368 | 5·360 | 18·827 | 1·242 | 5·388 | 2843 | 813 | 3470 |
| Chiang Mai Province | Mae Chaem | 4·822 | 5·212 | 18·698 | 1·642 | 6·009 | 2908 | 994 | 3655 |
| Chiang Mai Province | Mae On | 4·088 | 4·616 | 16·430 | 1·353 | 5·059 | 974 | 315 | 1210 |
| Chiang Mai Province | Mae Rim | 4·429 | 5·011 | 17·747 | 1·423 | 5·511 | 3621 | 1159 | 4474 |
| Chiang Mai Province | Mae Taeng | 4·482 | 5·200 | 18·086 | 1·416 | 5·585 | 3204 | 1035 | 3965 |
| Chiang Mai Province | Mae Wang | 4·594 | 5·276 | 18·217 | 1·503 | 5·702 | 1162 | 381 | 1444 |
| Chiang Mai Province | Mueang Chiang Mai | 4·326 | 4·901 | 17·468 | 1·410 | 5·279 | 17125 | 5667 | 21030 |
| Chiang Mai Province | Omkoi | 4·582 | 4·522 | 16·119 | 1·770 | 5·745 | 3326 | 1283 | 4211 |
| Chiang Mai Province | Phrao | 4·309 | 5·211 | 17·964 | 1·328 | 5·243 | 1851 | 573 | 2276 |
| Chiang Mai Province | Samoeng | 4·666 | 5·356 | 18·907 | 1·485 | 5·796 | 1208 | 385 | 1499 |
| Chiang Mai Province | San Kamphaeng | 4·199 | 4·868 | 16·871 | 1·384 | 5·159 | 3505 | 1176 | 4307 |
| Chiang Mai Province | San Pa Tong | 4·323 | 4·868 | 16·346 | 1·447 | 5·330 | 3288 | 1093 | 4038 |
| Chiang Mai Province | San Sai | 4·296 | 4·801 | 16·959 | 1·394 | 5·313 | 8004 | 2661 | 10052 |
| Chiang Mai Province | Saraphi | 4·245 | 4·887 | 16·717 | 1·421 | 5·284 | 3666 | 1180 | 4558 |
| Chiang Mai Province | Wiang Haeng | 4·631 | 5·464 | 19·487 | 1·370 | 5·720 | 1096 | 318 | 1374 |
| Chiang Rai Province | Chiang Khong | 4·151 | 5·376 | 18·121 | 1·136 | 5·031 | 2247 | 617 | 2715 |
| Chiang Rai Province | Chiang Saen | 4·185 | 5·290 | 18·177 | 1·154 | 5·110 | 1851 | 508 | 2270 |
| Chiang Rai Province | Doi Luang | 4·129 | 5·166 | 17·185 | 1·148 | 5·103 | 562 | 156 | 694 |
| Chiang Rai Province | Khun Tan | 3·997 | 5·101 | 16·811 | 1·160 | 4·846 | 1150 | 332 | 1405 |
| Chiang Rai Province | Mae Chan | 4·224 | 5·323 | 17·880 | 1·165 | 5·178 | 3577 | 984 | 4393 |
| Chiang Rai Province | Mae Fa Luang | 4·303 | 5·451 | 18·945 | 1·180 | 5·287 | 2326 | 646 | 2883 |
| Chiang Rai Province | Mae Lao | 4·205 | 5·249 | 17·869 | 1·210 | 5·127 | 1285 | 371 | 1582 |
| Chiang Rai Province | Mae Sai | 4·326 | 5·505 | 19·057 | 1·157 | 5·288 | 4457 | 1203 | 5456 |
| Chiang Rai Province | Mae Suai | 4·235 | 5·210 | 17·925 | 1·256 | 5·159 | 3146 | 949 | 3800 |
| Chiang Rai Province | Mueang Chiang Rai | 4·244 | 5·385 | 18·227 | 1·203 | 5·194 | 10138 | 2842 | 12379 |
| Chiang Rai Province | Pa Daet | 3·973 | 4·945 | 16·334 | 1·190 | 4·848 | 928 | 278 | 1122 |
| Chiang Rai Province | Phan | 4·101 | 5·032 | 17·504 | 1·227 | 4·948 | 4446 | 1328 | 5301 |

| ADM1 | ADM2 | β-thalassaemia carrier prevalence<br>Mean | SD | CI95% | Q25 | Q75 | Estimated carrier counts<br>Mean | Q25 | Q75 |
|---|---|---|---|---|---|---|---|---|---|
| Chiang Rai Province | Phaya Mengrai | 4·053 | 5·107 | 17·209 | 1·163 | 4·899 | 1504 | 431 | 1811 |
| Chiang Rai Province | Thoeng | 3·927 | 4·936 | 16·326 | 1·176 | 4·762 | 3100 | 937 | 3765 |
| Chiang Rai Province | Wiang Chai | 4·181 | 5·444 | 18·245 | 1·182 | 5·068 | 1849 | 515 | 2222 |
| Chiang Rai Province | Wiang Chiang Rung | 4·221 | 5·531 | 18·588 | 1·172 | 5·044 | 979 | 272 | 1157 |
| Chiang Rai Province | Wiang Kaen | 4·042 | 5·304 | 17·489 | 1·118 | 4·853 | 1161 | 317 | 1387 |
| Chiang Rai Province | Wiang Pa Pao | 4·154 | 4·941 | 17·136 | 1·278 | 5·096 | 2704 | 835 | 3317 |
| Chon Buri Province | Ban Bueng | 1·699 | 1·445 | 5·281 | 0·780 | 2·139 | 1743 | 811 | 2214 |
| Chon Buri Province | Bang Lamung | 1·713 | 1·482 | 5·346 | 0·751 | 2·167 | 9045 | 3976 | 11580 |
| Chon Buri Province | Bo Thong | 1·642 | 1·307 | 4·833 | 0·796 | 2·073 | 848 | 409 | 1074 |
| Chon Buri Province | Ko Chan | 1·701 | 1·309 | 4·944 | 0·826 | 2·149 | 497 | 239 | 627 |
| Chon Buri Province | Mueang Chon Buri | 1·669 | 1·356 | 5·062 | 0·793 | 2·102 | 5744 | 2748 | 7198 |
| Chon Buri Province | Nong Yai | 1·645 | 1·328 | 4·822 | 0·769 | 2·083 | 384 | 182 | 488 |
| Chon Buri Province | Phan Thong | 1·691 | 1·348 | 4·891 | 0·813 | 2·150 | 1507 | 721 | 1900 |
| Chon Buri Province | Phanat Nikhom | 1·707 | 1·351 | 4·959 | 0·830 | 2·142 | 2059 | 998 | 2594 |
| Chon Buri Province | Sattahip | 1·724 | 1·584 | 5·633 | 0·721 | 2·172 | 2348 | 985 | 2969 |
| Chon Buri Province | Si Racha | 1·693 | 1·400 | 5·131 | 0·776 | 2·139 | 9671 | 4417 | 12338 |
| Chumphon Province | Lamae | 4·360 | 5·086 | 18·998 | 1·233 | 5·450 | 1269 | 361 | 1583 |
| Chumphon Province | Lang Suan | 4·204 | 4·911 | 18·005 | 1·167 | 5·254 | 2078 | 582 | 2592 |
| Chumphon Province | Mueang Chumphon | 3·809 | 4·552 | 16·449 | 1·052 | 4·748 | 4299 | 1184 | 5349 |
| Chumphon Province | Pathio | 3·441 | 4·124 | 14·925 | 0·969 | 4·255 | 1690 | 475 | 2095 |
| Chumphon Province | Phato | 4·427 | 5·239 | 19·276 | 1·226 | 5·471 | 941 | 259 | 1170 |
| Chumphon Province | Sawi | 4·023 | 4·917 | 18·255 | 1·104 | 4·924 | 2141 | 591 | 2634 |
| Chumphon Province | Tha Sae | 3·615 | 4·363 | 15·584 | 1·006 | 4·475 | 3393 | 949 | 4185 |
| Chumphon Province | Thung Tako | 4·080 | 4·955 | 17·436 | 1·126 | 4·986 | 918 | 254 | 1118 |
| Kalasin | Don Chan | 1·357 | 1·247 | 4·557 | 0·575 | 1·677 | 212 | 90 | 263 |
| Kalasin | Huai Mek | 1·462 | 1·202 | 4·333 | 0·691 | 1·828 | 547 | 259 | 685 |
| Kalasin | Huai Phueng | 1·441 | 1·422 | 4·993 | 0·575 | 1·796 | 350 | 137 | 435 |
| Kalasin | Kamalasai | 1·295 | 1·165 | 4·259 | 0·564 | 1·603 | 599 | 261 | 733 |
| Kalasin | Kham Muang | 1·609 | 1·534 | 5·510 | 0·664 | 2·018 | 525 | 217 | 655 |
| Kalasin | Khao Wong | 1·405 | 1·392 | 4·982 | 0·546 | 1·755 | 368 | 141 | 455 |
| Kalasin | Khong Chai | 1·330 | 1·244 | 4·282 | 0·576 | 1·649 | 248 | 108 | 307 |
| Kalasin | Kuchinarai | 1·297 | 1·226 | 4·424 | 0·534 | 1·628 | 1011 | 415 | 1264 |
| Kalasin | Mueang Kalasin | 1·391 | 1·229 | 4·482 | 0·611 | 1·736 | 1570 | 693 | 1958 |
| Kalasin | Na Khu | 1·461 | 1·493 | 5·061 | 0·572 | 1·813 | 291 | 115 | 361 |
| Kalasin | Na Mon | 1·382 | 1·326 | 4·567 | 0·580 | 1·701 | 306 | 125 | 376 |
| Kalasin | Nong Kung Si | 1·500 | 1·304 | 4·740 | 0·678 | 1·885 | 694 | 315 | 875 |
| Kalasin | Rong Kham | 1·258 | 1·137 | 4·184 | 0·555 | 1·540 | 92 | 41 | 114 |
| Kalasin | Sahatsakhan | 1·500 | 1·400 | 5·026 | 0·635 | 1·860 | 375 | 158 | 461 |
| Kalasin | Sam Chai | 1·599 | 1·533 | 5·391 | 0·675 | 1·982 | 316 | 133 | 392 |
| Kalasin | Somdet | 1·502 | 1·449 | 5·235 | 0·613 | 1·873 | 611 | 252 | 771 |
| Kalasin | Tha Khantho | 1·628 | 1·461 | 5·385 | 0·725 | 2·031 | 378 | 166 | 475 |
| Kalasin | Yang Talat | 1·397 | 1·203 | 4·342 | 0·641 | 1·740 | 1231 | 565 | 1533 |
| Kamphaeng Phet Province | Khanu Woralaksaburi | 2·544 | 2·321 | 8·332 | 1·097 | 3·174 | 2682 | 1161 | 3349 |
| Kamphaeng Phet Province | Khlong Khlung | 2·568 | 2·290 | 8·225 | 1·149 | 3·197 | 1911 | 854 | 2378 |
| Kamphaeng Phet Province | Khlong Lan | 2·988 | 2·739 | 9·951 | 1·263 | 3·734 | 2130 | 916 | 2666 |
| Kamphaeng Phet Province | Kosamphi Nakhon | 3·046 | 2·722 | 9·927 | 1·295 | 3·849 | 1140 | 491 | 1433 |
| Kamphaeng Phet Province | Lan Krabue | 2·486 | 2·089 | 7·366 | 1·150 | 3·145 | 1528 | 708 | 1930 |
| Kamphaeng Phet Province | Mueang Kamphaeng Phet | 2·811 | 2·474 | 9·033 | 1·233 | 3·542 | 7239 | 3163 | 9169 |
| Kamphaeng Phet Province | Pang Sila Thong | 2·843 | 2·598 | 9·423 | 1·215 | 3·553 | 753 | 330 | 938 |
| Kamphaeng Phet Province | Phran Kratai | 2·818 | 2·430 | 8·866 | 1·265 | 3·571 | 3277 | 1480 | 4143 |
| Kamphaeng Phet Province | Sai Ngam | 2·497 | 2·149 | 7·735 | 1·132 | 3·126 | 1461 | 657 | 1840 |
| Kamphaeng Phet Province | Sai Thong Watthana | 2·465 | 2·124 | 7·973 | 1·098 | 3·074 | 711 | 318 | 890 |
| Kanchanaburi Province | Bo Phloi | 2·159 | 2·052 | 7·380 | 0·857 | 2·733 | 1046 | 416 | 1327 |
| Kanchanaburi Province | Dan Makham Tia | 2·186 | 2·172 | 7·787 | 0·830 | 2·749 | 612 | 233 | 771 |
| Kanchanaburi Province | Huai Krachao | 1·975 | 1·810 | 6·465 | 0·821 | 2·497 | 706 | 296 | 892 |
| Kanchanaburi Province | Lao Khwan | 2·071 | 1·924 | 6·934 | 0·853 | 2·614 | 883 | 363 | 1118 |

| | | β-thalassaemia carrier prevalence | | | | | Estimated carrier counts | | |
|---|---|---|---|---|---|---|---|---|---|
| ADM1 | ADM2 | Mean | SD | CI95% | Q25 | Q75 | Mean | Q25 | Q75 |
| Kanchanaburi Province | Mueang Kanchanaburi | 2·210 | 2·182 | 7·753 | 0·848 | 2·789 | 3526 | 1395 | 4480 |
| Kanchanaburi Province | Nong Prue | 2·270 | 2·154 | 7·969 | 0·897 | 2·861 | 756 | 298 | 952 |
| Kanchanaburi Province | Phanom Thuan | 1·930 | 1·827 | 6·399 | 0·793 | 2·443 | 1012 | 419 | 1277 |
| Kanchanaburi Province | Sai Yok | 2·564 | 2·629 | 9·344 | 0·952 | 3·201 | 1216 | 449 | 1495 |
| Kanchanaburi Province | Sangkhla Buri | 3·394 | 3·346 | 12·156 | 1·301 | 4·263 | 1065 | 396 | 1315 |
| Kanchanaburi Province | Si Sawat | 2·537 | 2·484 | 8·945 | 0·985 | 3·197 | 550 | 211 | 688 |
| Kanchanaburi Province | Tha Maka | 1·870 | 1·800 | 6·376 | 0·756 | 2·335 | 2422 | 970 | 3040 |
| Kanchanaburi Province | Tha Muang | 1·968 | 1·890 | 6·811 | 0·773 | 2·482 | 2222 | 875 | 2811 |
| Kanchanaburi Province | Thong Pha Phum | 3·047 | 3·050 | 10·939 | 1·142 | 3·844 | 1307 | 488 | 1649 |
| Khon Kaen Province | Ban Fang | 1·607 | 1·273 | 4·604 | 0·789 | 2·015 | 633 | 308 | 795 |
| Khon Kaen Province | Ban Haet | 1·478 | 1·184 | 4·232 | 0·706 | 1·870 | 355 | 169 | 447 |
| Khon Kaen Province | Ban Phai | 1·429 | 1·161 | 4·224 | 0·683 | 1·791 | 1043 | 500 | 1304 |
| Khon Kaen Province | Chonnabot | 1·554 | 1·331 | 4·807 | 0·705 | 1·957 | 618 | 281 | 781 |
| Khon Kaen Province | Chum Phae | 1·829 | 1·595 | 5·587 | 0·844 | 2·278 | 2048 | 938 | 2546 |
| Khon Kaen Province | Khao Suan Kwang | 1·713 | 1·494 | 5·329 | 0·782 | 2·120 | 408 | 188 | 506 |
| Khon Kaen Province | Khok Pho Chai | 1·614 | 1·349 | 4·893 | 0·755 | 2·054 | 234 | 110 | 299 |
| Khon Kaen Province | Kranuan | 1·577 | 1·313 | 4·742 | 0·741 | 1·982 | 741 | 354 | 934 |
| Khon Kaen Province | Mancha Khiri | 1·575 | 1·303 | 4·633 | 0·747 | 1·984 | 816 | 389 | 1034 |
| Khon Kaen Province | Mueang Khon Kaen | 1·532 | 1·230 | 4·451 | 0·738 | 1·923 | 9915 | 4814 | 12486 |
| Khon Kaen Province | Nam Phong | 1·609 | 1·313 | 4·814 | 0·770 | 2·002 | 1426 | 678 | 1772 |
| Khon Kaen Province | Non Sila | 1·453 | 1·203 | 4·336 | 0·689 | 1·826 | 304 | 144 | 383 |
| Khon Kaen Province | Nong Na Kham | 1·789 | 1·588 | 5·372 | 0·832 | 2·235 | 404 | 189 | 504 |
| Khon Kaen Province | Nong Ruea | 1·662 | 1·375 | 4·921 | 0·799 | 2·075 | 1181 | 571 | 1474 |
| Khon Kaen Province | Nong Song Hong | 1·397 | 1·193 | 4·339 | 0·630 | 1·754 | 876 | 398 | 1099 |
| Khon Kaen Province | Phon | 1·494 | 1·276 | 4·674 | 0·680 | 1·884 | 1060 | 483 | 1349 |
| Khon Kaen Province | Phra Yuen | 1·552 | 1·283 | 4·596 | 0·737 | 1·942 | 406 | 192 | 508 |
| Khon Kaen Province | Phu Pha Man | 1·902 | 1·638 | 5·857 | 0·868 | 2·407 | 220 | 97 | 279 |
| Khon Kaen Province | Phu Wiang | 1·706 | 1·410 | 5·030 | 0·815 | 2·138 | 990 | 473 | 1237 |
| Khon Kaen Province | Pueai Noi | 1·362 | 1·132 | 4·049 | 0·636 | 1·704 | 140 | 65 | 175 |
| Khon Kaen Province | Sam Sung | 1·489 | 1·228 | 4·300 | 0·712 | 1·862 | 242 | 116 | 301 |
| Khon Kaen Province | Si Chomphu | 1·860 | 1·635 | 5·744 | 0·865 | 2·316 | 996 | 470 | 1236 |
| Khon Kaen Province | Ubolratana | 1·696 | 1·467 | 5·207 | 0·800 | 2·122 | 592 | 282 | 738 |
| Khon Kaen Province | Waeng Noi | 1·575 | 1·382 | 4·806 | 0·710 | 1·951 | 578 | 261 | 718 |
| Khon Kaen Province | Waeng Yai | 1·580 | 1·334 | 4·853 | 0·713 | 1·985 | 280 | 126 | 351 |
| Khon Kaen Province | Wiang Kao | 1·774 | 1·514 | 5·283 | 0·833 | 2·225 | 285 | 132 | 356 |
| Krabi Province | Ao Luek | 5·456 | 6·165 | 22·368 | 1·655 | 6·836 | 2208 | 669 | 2741 |
| Krabi Province | Khao Phanom | 5·360 | 5·697 | 20·707 | 1·765 | 6·839 | 2688 | 879 | 3432 |
| Krabi Province | Khlong Thom | 5·567 | 5·796 | 20·869 | 1·917 | 7·084 | 3529 | 1213 | 4475 |
| Krabi Province | Ko Lanta | 5·797 | 6·082 | 22·730 | 1·971 | 7·306 | 774 | 265 | 976 |
| Krabi Province | Mueang Krabi | 5·577 | 6·154 | 22·564 | 1·752 | 7·004 | 4056 | 1284 | 5074 |
| Krabi Province | Nuea Khlong | 5·557 | 5·914 | 21·729 | 1·861 | 7·030 | 2412 | 792 | 3044 |
| Krabi Province | Plai Phraya | 5·343 | 5·970 | 21·833 | 1·604 | 6·733 | 1706 | 516 | 2151 |
| Lampang Province | Chae Hom | 3·826 | 4·381 | 14·942 | 1·285 | 4·697 | 1411 | 472 | 1717 |
| Lampang Province | Hang Chat | 3·874 | 4·203 | 14·602 | 1·374 | 4·807 | 1907 | 683 | 2347 |
| Lampang Province | Ko Kha | 3·729 | 3·828 | 13·402 | 1·369 | 4·704 | 1712 | 629 | 2178 |
| Lampang Province | Mae Mo | 3·473 | 3·777 | 12·883 | 1·245 | 4·293 | 1231 | 454 | 1525 |
| Lampang Province | Mae Phrik | 3·664 | 3·498 | 12·656 | 1·492 | 4·616 | 488 | 203 | 615 |
| Lampang Province | Mae Tha | 3·538 | 3·661 | 12·516 | 1·331 | 4·425 | 1766 | 661 | 2206 |
| Lampang Province | Mueang Lampang | 3·717 | 4·056 | 14·163 | 1·316 | 4·591 | 7523 | 2719 | 9329 |
| Lampang Province | Mueang Pan | 4·015 | 4·680 | 15·841 | 1·301 | 4·915 | 1028 | 338 | 1261 |
| Lampang Province | Ngao | 3·638 | 4·189 | 14·266 | 1·213 | 4·479 | 1793 | 608 | 2214 |
| Lampang Province | Soem Ngam | 3·832 | 3·891 | 14·077 | 1·427 | 4·814 | 1062 | 403 | 1331 |
| Lampang Province | Sop Prap | 3·547 | 3·540 | 12·215 | 1·366 | 4·468 | 883 | 340 | 1094 |
| Lampang Province | Thoen | 3·437 | 3·263 | 11·677 | 1·410 | 4·302 | 2003 | 829 | 2509 |
| Lampang Province | Wang Nuea | 4·035 | 4·851 | 16·694 | 1·249 | 4·945 | 1525 | 476 | 1873 |
| Lamphun Province | Ban Hong | 4·226 | 4·521 | 16·147 | 1·513 | 5·215 | 1345 | 472 | 1670 |
| Lamphun Province | Ban Thi | 4·125 | 4·598 | 14·919 | 1·407 | 5·171 | 317 | 107 | 396 |

| ADM1 | ADM2 | β-thalassaemia carrier prevalence Mean | SD | CI95% | Q25 | Q75 | Estimated carrier counts Mean | Q25 | Q75 |
|---|---|---|---|---|---|---|---|---|---|
| Lamphun Province | Li | 4·000 | 4·085 | 14·452 | 1·519 | 4·969 | 2137 | 798 | 2648 |
| Lamphun Province | Mae Tha | 4·036 | 4·345 | 15·543 | 1·433 | 5·000 | 1212 | 422 | 1507 |
| Lamphun Province | Mueang Lamphun | 4·112 | 4·456 | 15·394 | 1·427 | 5·124 | 7629 | 2668 | 9559 |
| Lamphun Province | Pa Sang | 4·234 | 4·652 | 16·083 | 1·501 | 5·259 | 2444 | 845 | 3012 |
| Lamphun Province | Thung Hua Chang | 3·908 | 4·035 | 14·486 | 1·463 | 4·834 | 596 | 223 | 747 |
| Lamphun Province | Wiang Nong Long | 4·397 | 4·880 | 16·532 | 1·507 | 5·396 | 213 | 73 | 261 |
| Loei Province | Chiang Khan | 2·371 | 2·262 | 8·071 | 0·945 | 2·987 | 886 | 358 | 1124 |
| Loei Province | Dan Sai | 2·106 | 1·677 | 6·139 | 1·003 | 2·663 | 852 | 411 | 1073 |
| Loei Province | Erawan | 2·075 | 1·935 | 6·644 | 0·900 | 2·586 | 409 | 176 | 509 |
| Loei Province | Mueang Loei | 2·179 | 1·967 | 7·017 | 0·946 | 2·746 | 2444 | 1072 | 3018 |
| Loei Province | Na Duang | 2·171 | 1·996 | 7·252 | 0·922 | 2·716 | 310 | 130 | 388 |
| Loei Province | Na Haeo | 2·226 | 1·779 | 6·528 | 1·065 | 2·816 | 238 | 112 | 295 |
| Loei Province | Nong Hin | 1·997 | 1·741 | 6·275 | 0·900 | 2·484 | 349 | 157 | 435 |
| Loei Province | Pak Chom | 2·410 | 2·401 | 8·523 | 0·922 | 3·005 | 635 | 242 | 801 |
| Loei Province | Pha Khao | 1·953 | 1·781 | 6·253 | 0·856 | 2·449 | 462 | 202 | 583 |
| Loei Province | Phu Kradueng | 1·934 | 1·674 | 5·897 | 0·892 | 2·418 | 306 | 139 | 387 |
| Loei Province | Phu Luang | 2·032 | 1·773 | 6·361 | 0·915 | 2·540 | 388 | 173 | 481 |
| Loei Province | Phu Ruea | 2·152 | 1·853 | 6·593 | 0·975 | 2·705 | 377 | 172 | 477 |
| Loei Province | Tha Li | 2·273 | 2·030 | 7·338 | 0·976 | 2·895 | 490 | 206 | 629 |
| Loei Province | Wang Saphung | 2·081 | 1·861 | 6·506 | 0·933 | 2·607 | 1642 | 737 | 2057 |
| Lopburi Province | Ban Mi | 1·934 | 1·690 | 6·126 | 0·853 | 2·415 | 1166 | 516 | 1455 |
| Lopburi Province | Chai Badan | 1·914 | 1·715 | 6·196 | 0·826 | 2·403 | 1595 | 688 | 1996 |
| Lopburi Province | Khok Charoen | 1·966 | 1·771 | 6·274 | 0·848 | 2·471 | 468 | 206 | 589 |
| Lopburi Province | Khok Samrong | 1·899 | 1·602 | 5·827 | 0·848 | 2·428 | 1273 | 569 | 1628 |
| Lopburi Province | Mueang Lop Buri | 1·860 | 1·624 | 5·805 | 0·832 | 2·346 | 4778 | 2144 | 6032 |
| Lopburi Province | Nong Muang | 1·936 | 1·675 | 6·109 | 0·851 | 2·466 | 550 | 244 | 699 |
| Lopburi Province | Phatthana Nikhom | 1·865 | 1·617 | 5·864 | 0·830 | 2·348 | 2009 | 894 | 2522 |
| Lopburi Province | Sa Bot | 1·964 | 1·783 | 6·412 | 0·843 | 2·442 | 409 | 179 | 509 |
| Lopburi Province | Tha Luang | 1·916 | 1·679 | 5·983 | 0·839 | 2·410 | 587 | 259 | 743 |
| Lopburi Province | Tha Wung | 1·888 | 1·600 | 5·891 | 0·842 | 2·369 | 916 | 411 | 1144 |
| Mae Hong Son Province | Khun Yuam | 5·222 | 5·725 | 20·728 | 1·716 | 6·497 | 838 | 271 | 1030 |
| Mae Hong Son Province | Mae La Noi | 5·065 | 5·271 | 18·961 | 1·784 | 6·377 | 1210 | 426 | 1518 |
| Mae Hong Son Province | Mae Sariang | 5·313 | 5·324 | 19·306 | 1·962 | 6·737 | 2172 | 804 | 2754 |
| Mae Hong Son Province | Mueang Mae Hong Son | 5·068 | 5·779 | 20·819 | 1·558 | 6·330 | 2341 | 725 | 2930 |
| Mae Hong Son Province | Pai | 4·799 | 5·618 | 20·187 | 1·431 | 5·940 | 1229 | 359 | 1530 |
| Mae Hong Son Province | Pang Mapha | 4·832 | 5·653 | 20·224 | 1·423 | 6·028 | 932 | 275 | 1155 |
| Mae Hong Son Province | Sop Moei | 5·075 | 4·920 | 17·902 | 1·964 | 6·426 | 1075 | 415 | 1368 |
| Maha Sarakham Province | Borabue | 1·326 | 1·114 | 4·058 | 0·611 | 1·677 | 844 | 387 | 1063 |
| Maha Sarakham Province | Chiang Yuen | 1·438 | 1·167 | 4·204 | 0·689 | 1·805 | 662 | 319 | 833 |
| Maha Sarakham Province | Chuen Chom | 1·475 | 1·230 | 4·631 | 0·714 | 1·823 | 235 | 114 | 292 |
| Maha Sarakham Province | Kae Dam | 1·272 | 1·125 | 3·900 | 0·562 | 1·586 | 268 | 119 | 335 |
| Maha Sarakham Province | Kantharawichai | 1·358 | 1·130 | 4·108 | 0·628 | 1·715 | 935 | 432 | 1186 |
| Maha Sarakham Province | Kosum Phisai | 1·418 | 1·156 | 4·161 | 0·677 | 1·784 | 1088 | 519 | 1366 |
| Maha Sarakham Province | Kut Rang | 1·385 | 1·162 | 4·181 | 0·652 | 1·731 | 436 | 206 | 544 |
| Maha Sarakham Province | Mueang Maha Sarakham | 1·319 | 1·167 | 4·180 | 0·592 | 1·637 | 1936 | 867 | 2413 |
| Maha Sarakham Province | Na Chueak | 1·311 | 1·125 | 4·096 | 0·585 | 1·653 | 442 | 200 | 557 |
| Maha Sarakham Province | Na Dun | 1·237 | 1·123 | 3·983 | 0·547 | 1·554 | 244 | 108 | 308 |
| Maha Sarakham Province | Phayakkhaphum Phisai | 1·163 | 1·016 | 3·699 | 0·518 | 1·464 | 658 | 295 | 824 |
| Maha Sarakham Province | Wapi Pathum | 1·219 | 1·051 | 3·814 | 0·550 | 1·524 | 865 | 390 | 1083 |
| Maha Sarakham Province | Yang Sisurat | 1·261 | 1·095 | 3·928 | 0·561 | 1·598 | 309 | 139 | 393 |
| Mukdahan Province | Don Tan | 1·166 | 1·288 | 4·172 | 0·433 | 1·446 | 510 | 190 | 634 |
| Mukdahan Province | Dong Luang | 1·465 | 1·555 | 5·329 | 0·542 | 1·808 | 655 | 242 | 817 |
| Mukdahan Province | Khamcha-I | 1·353 | 1·440 | 4·915 | 0·508 | 1·663 | 687 | 255 | 850 |
| Mukdahan Province | Mueang Mukdahan | 1·309 | 1·441 | 4·867 | 0·479 | 1·610 | 2052 | 747 | 2519 |
| Mukdahan Province | Nikhom Kham Soi | 1·193 | 1·215 | 4·227 | 0·461 | 1·479 | 424 | 162 | 525 |
| Mukdahan Province | Nong Sung | 1·221 | 1·162 | 4·228 | 0·490 | 1·548 | 208 | 84 | 261 |

| | | β-thalassaemia carrier prevalence | | | | | Estimated carrier counts | | |
|---|---|---|---|---|---|---|---|---|---|
| ADM1 | ADM2 | Mean | SD | CI95% | Q25 | Q75 | Mean | Q25 | Q75 |
| Mukdahan Province | Wan Yai | 1·441 | 1·638 | 5·509 | 0·496 | 1·755 | 305 | 105 | 371 |
| Nakhon Nayok Province | Ban Na | 1·815 | 1·485 | 5·520 | 0·847 | 2·280 | 1077 | 507 | 1351 |
| Nakhon Nayok Province | Mueang Nakhon Nayok | 1·830 | 1·496 | 5·449 | 0·865 | 2·293 | 1701 | 804 | 2121 |
| Nakhon Nayok Province | Ongkharak | 1·768 | 1·418 | 5·248 | 0·843 | 2·230 | 1491 | 710 | 1869 |
| Nakhon Nayok Province | Pak Phli | 1·794 | 1·415 | 5·194 | 0·861 | 2·284 | 412 | 196 | 521 |
| Nakhon Pathom Province | Bang Len | 1·702 | 1·463 | 5·285 | 0·760 | 2·135 | 1466 | 656 | 1844 |
| Nakhon Pathom Province | Don Tum | 1·704 | 1·481 | 5·283 | 0·771 | 2·141 | 672 | 305 | 848 |
| Nakhon Pathom Province | Kamphaeng Saen | 1·781 | 1·591 | 5·688 | 0·756 | 2·264 | 1652 | 698 | 2088 |
| Nakhon Pathom Province | Mueang Nakhon Pathom | 1·723 | 1·489 | 5·342 | 0·758 | 2·183 | 4392 | 1957 | 5578 |
| Nakhon Pathom Province | Nakhon Chai Si | 1·652 | 1·389 | 5·146 | 0·757 | 2·069 | 2023 | 920 | 2555 |
| Nakhon Pathom Province | Phutthamonthon | 1·636 | 1·414 | 5·199 | 0·749 | 2·028 | 1188 | 541 | 1467 |
| Nakhon Pathom Province | Sam Phran | 1·652 | 1·393 | 5·057 | 0·760 | 2·091 | 5253 | 2412 | 6672 |
| Nakhon Phanom Province | Ban Phaeng | 2·282 | 2·604 | 9·348 | 0·726 | 2·825 | 500 | 156 | 614 |
| Nakhon Phanom Province | Mueang Nakhon Phanom | 1·900 | 2·319 | 8·023 | 0·597 | 2·261 | 2408 | 745 | 2857 |
| Nakhon Phanom Province | Na Kae | 1·575 | 1·739 | 6·024 | 0·563 | 1·931 | 844 | 299 | 1033 |
| Nakhon Phanom Province | Na Thom | 2·196 | 2·496 | 8·989 | 0·724 | 2·704 | 274 | 90 | 338 |
| Nakhon Phanom Province | Na Wa | 1·958 | 2·135 | 7·557 | 0·693 | 2·396 | 588 | 210 | 717 |
| Nakhon Phanom Province | Phon Sawan | 1·959 | 2·271 | 7·905 | 0·652 | 2·388 | 655 | 217 | 803 |
| Nakhon Phanom Province | Pla Pak | 1·768 | 2·065 | 7·219 | 0·594 | 2·138 | 664 | 221 | 801 |
| Nakhon Phanom Province | Renu Nakhon | 1·662 | 1·954 | 6·637 | 0·555 | 2·001 | 544 | 182 | 658 |
| Nakhon Phanom Province | Si Songkhram | 2·074 | 2·395 | 8·429 | 0·693 | 2·532 | 1188 | 397 | 1448 |
| Nakhon Phanom Province | Tha Uthen | 2·059 | 2·537 | 8·660 | 0·641 | 2·470 | 847 | 256 | 1011 |
| Nakhon Phanom Province | That Phanom | 1·578 | 1·812 | 6·141 | 0·541 | 1·940 | 727 | 248 | 899 |
| Nakhon Phanom Province | Wang Yang | 1·659 | 1·824 | 6·434 | 0·584 | 2·030 | 191 | 68 | 235 |
| Nakhon Ratchasima Province | Ban Lueam | 1·691 | 1·495 | 5·598 | 0·725 | 2·109 | 196 | 83 | 245 |
| Nakhon Ratchasima Province | Bua Lai | 1·445 | 1·225 | 4·356 | 0·667 | 1·826 | 200 | 92 | 252 |
| Nakhon Ratchasima Province | Bua Yai | 1·555 | 1·344 | 4·937 | 0·687 | 1·957 | 767 | 344 | 967 |
| Nakhon Ratchasima Province | Chakkarat | 1·472 | 1·289 | 4·715 | 0·654 | 1·843 | 566 | 252 | 709 |
| Nakhon Ratchasima Province | Chaloem Phra Kiat | 1·584 | 1·421 | 5·101 | 0·687 | 1·997 | 543 | 236 | 689 |
| Nakhon Ratchasima Province | Chok Chai | 1·599 | 1·383 | 5·022 | 0·720 | 2·002 | 1455 | 652 | 1811 |
| Nakhon Ratchasima Province | Chum Phuang | 1·335 | 1·130 | 4·168 | 0·601 | 1·692 | 586 | 263 | 740 |
| Nakhon Ratchasima Province | Dan Khun Thot | 1·787 | 1·582 | 5·699 | 0·787 | 2·251 | 1699 | 750 | 2141 |
| Nakhon Ratchasima Province | Huai Thalaeng | 1·396 | 1·244 | 4·356 | 0·609 | 1·760 | 548 | 238 | 691 |
| Nakhon Ratchasima Province | Kaeng Sanam Nang | 1·648 | 1·462 | 5·358 | 0·717 | 2·054 | 281 | 121 | 349 |
| Nakhon Ratchasima Province | Kham Sakaesaeng | 1·645 | 1·433 | 5·173 | 0·725 | 2·092 | 481 | 210 | 615 |
| Nakhon Ratchasima Province | Kham Thale So | 1·698 | 1·465 | 5·290 | 0·761 | 2·149 | 339 | 153 | 429 |
| Nakhon Ratchasima Province | Khon Buri | 1·564 | 1·315 | 4·783 | 0·726 | 1·961 | 1257 | 580 | 1585 |
| Nakhon Ratchasima Province | Khong | 1·576 | 1·414 | 5·032 | 0·701 | 1·971 | 641 | 287 | 804 |
| Nakhon Ratchasima Province | Lam Thamenchai | 1·270 | 1·101 | 3·940 | 0·560 | 1·601 | 241 | 105 | 303 |
| Nakhon Ratchasima Province | Mueang Nakhon Ratchasima | 1·644 | 1·432 | 5·214 | 0·723 | 2·072 | 12195 | 5380 | 15239 |
| Nakhon Ratchasima Province | Mueang Yang | 1·310 | 1·128 | 4·150 | 0·587 | 1·638 | 251 | 111 | 312 |
| Nakhon Ratchasima Province | Noen Sa-Nga | 1·710 | 1·511 | 5·360 | 0·754 | 2·140 | 353 | 155 | 443 |
| Nakhon Ratchasima Province | Non Daeng | 1·433 | 1·247 | 4·419 | 0·639 | 1·815 | 176 | 79 | 223 |
| Nakhon Ratchasima Province | Non Sung | 1·569 | 1·364 | 4·844 | 0·703 | 1·976 | 1124 | 504 | 1410 |

| ADM1 | ADM2 | β-thalassaemia carrier prevalence Mean | SD | CI95% | Q25 | Q75 | Estimated carrier counts Mean | Q25 | Q75 |
|---|---|---|---|---|---|---|---|---|---|
| Nakhon Ratchasima Province | Non Thai | 1·685 | 1·490 | 5·392 | 0·744 | 2·112 | 793 | 349 | 987 |
| Nakhon Ratchasima Province | Nong Bun Mak | 1·476 | 1·255 | 4·515 | 0·671 | 1·855 | 477 | 212 | 602 |
| Nakhon Ratchasima Province | Pak Chong | 1·817 | 1·512 | 5·445 | 0·835 | 2·297 | 5975 | 2706 | 7565 |
| Nakhon Ratchasima Province | Pak Thong Chai | 1·722 | 1·467 | 5·339 | 0·786 | 2·168 | 1586 | 722 | 1987 |
| Nakhon Ratchasima Province | Phimai | 1·431 | 1·255 | 4·447 | 0·641 | 1·796 | 1290 | 575 | 1632 |
| Nakhon Ratchasima Province | Phra Thong Kham | 1·717 | 1·550 | 5·471 | 0·751 | 2·159 | 422 | 185 | 529 |
| Nakhon Ratchasima Province | Prathai | 1·396 | 1·213 | 4·352 | 0·629 | 1·750 | 530 | 238 | 664 |
| Nakhon Ratchasima Province | Sida | 1·446 | 1·209 | 4·411 | 0·648 | 1·828 | 161 | 73 | 206 |
| Nakhon Ratchasima Province | Sikhio | 1·815 | 1·606 | 5·703 | 0·805 | 2·278 | 2033 | 903 | 2542 |
| Nakhon Ratchasima Province | Soeng Sang | 1·390 | 1·162 | 4·222 | 0·648 | 1·751 | 737 | 340 | 935 |
| Nakhon Ratchasima Province | Sung Noen | 1·729 | 1·502 | 5·485 | 0·769 | 2·175 | 1804 | 804 | 2261 |
| Nakhon Ratchasima Province | Wang Nam Khiao | 1·768 | 1·484 | 5·410 | 0·814 | 2·226 | 761 | 351 | 961 |
| Nakhon Sawan Province | Banphot Phisai | 2·350 | 2·063 | 7·485 | 1·049 | 2·940 | 1681 | 749 | 2099 |
| Nakhon Sawan Province | Bueng Samakkhi | 2·381 | 2·029 | 7·480 | 1·081 | 2·961 | 633 | 288 | 794 |
| Nakhon Sawan Province | Chum Saeng | 2·204 | 1·917 | 7·036 | 0·968 | 2·790 | 1184 | 520 | 1492 |
| Nakhon Sawan Province | Chum Ta Bong | 2·591 | 2·419 | 8·363 | 1·092 | 3·247 | 312 | 131 | 390 |
| Nakhon Sawan Province | Kao Liao | 2·289 | 2·028 | 7·344 | 1·004 | 2·874 | 582 | 257 | 728 |
| Nakhon Sawan Province | Krok Phra | 2·204 | 1·950 | 7·083 | 0·991 | 2·763 | 564 | 253 | 712 |
| Nakhon Sawan Province | Lat Yao | 2·401 | 2·166 | 7·783 | 1·034 | 2·995 | 1439 | 619 | 1801 |
| Nakhon Sawan Province | Mae Poen | 2·653 | 2·488 | 9·030 | 1·112 | 3·299 | 470 | 198 | 586 |
| Nakhon Sawan Province | Mae Wong | 2·705 | 2·519 | 9·150 | 1·137 | 3·386 | 707 | 302 | 885 |
| Nakhon Sawan Province | Mueang Nakhon Sawan | 2·227 | 1·948 | 7·064 | 0·987 | 2·791 | 6182 | 2762 | 7727 |
| Nakhon Sawan Province | Nong Bua | 2·074 | 1·787 | 6·426 | 0·933 | 2·600 | 942 | 425 | 1181 |
| Nakhon Sawan Province | Phaisali | 2·016 | 1·751 | 6·368 | 0·885 | 2·558 | 951 | 417 | 1206 |
| Nakhon Sawan Province | Phayuha Khiri | 2·110 | 1·822 | 6·810 | 0·936 | 2·657 | 1003 | 443 | 1253 |
| Nakhon Sawan Province | Tak Fa | 2·011 | 1·743 | 6·323 | 0·874 | 2·560 | 760 | 334 | 966 |
| Nakhon Sawan Province | Takhli | 2·003 | 1·768 | 6·439 | 0·871 | 2·509 | 1416 | 620 | 1790 |
| Nakhon Sawan Province | Tha Tako | 2·080 | 1·812 | 6·530 | 0·921 | 2·617 | 1022 | 452 | 1284 |
| Nakhon Si Thammarat Province | Bang Khan | 5·414 | 5·354 | 19·999 | 1·934 | 6·907 | 2493 | 879 | 3180 |
| Nakhon Si Thammarat Province | Chaloem Phra Kiat | 5·011 | 4·844 | 17·133 | 1·858 | 6·499 | 766 | 278 | 993 |
| Nakhon Si Thammarat Province | Chang Klang | 5·141 | 5·174 | 19·335 | 1·789 | 6·600 | 885 | 314 | 1134 |
| Nakhon Si Thammarat Province | Cha-Uat | 5·176 | 4·771 | 17·513 | 2·037 | 6·603 | 3338 | 1320 | 4264 |
| Nakhon Si Thammarat Province | Chawang | 5·144 | 5·413 | 19·801 | 1·717 | 6·543 | 3421 | 1160 | 4332 |
| Nakhon Si Thammarat Province | Chian Yai | 4·949 | 4·692 | 16·949 | 1·896 | 6·321 | 1638 | 624 | 2097 |
| Nakhon Si Thammarat Province | Chulabhorn | 5·230 | 5·063 | 18·908 | 1·953 | 6·729 | 1736 | 648 | 2252 |
| Nakhon Si Thammarat Province | Hua Sai | 5·015 | 4·552 | 17·069 | 1·994 | 6·505 | 2497 | 992 | 3262 |
| Nakhon Si Thammarat Province | Khanom | 4·446 | 5·014 | 18·169 | 1·363 | 5·579 | 1367 | 422 | 1746 |
| Nakhon Si Thammarat Province | Lam Thap | 5·438 | 5·565 | 20·942 | 1·918 | 6·881 | 1415 | 501 | 1789 |
| Nakhon Si Thammarat Province | Lan Saka | 5·087 | 5·082 | 18·824 | 1·768 | 6·572 | 1821 | 630 | 2340 |
| Nakhon Si Thammarat Province | Mueang Nakhon Si Thammarat | 4·854 | 4·867 | 17·664 | 1·709 | 6·217 | 9258 | 3198 | 11902 |
| Nakhon Si Thammarat Province | Na Bon | 5·274 | 5·395 | 20·099 | 1·797 | 6·711 | 955 | 323 | 1212 |
| Nakhon Si Thammarat Province | Nopphitam | 4·786 | 5·063 | 18·465 | 1·564 | 6·117 | 2015 | 659 | 2584 |

| ADM1 | ADM2 | β-thalassaemia carrier prevalence Mean | SD | CI95% | Q25 | Q75 | Estimated carrier counts Mean | Q25 | Q75 |
|---|---|---|---|---|---|---|---|---|---|
| Nakhon Si Thammarat Province | Pak Phanang | 4·869 | 4·861 | 17·709 | 1·766 | 6·172 | 2650 | 956 | 3395 |
| Nakhon Si Thammarat Province | Phipun | 5·003 | 5·314 | 19·400 | 1·646 | 6·337 | 1765 | 594 | 2226 |
| Nakhon Si Thammarat Province | Phra Phrom | 4·974 | 4·814 | 17·908 | 1·827 | 6·383 | 2708 | 991 | 3463 |
| Nakhon Si Thammarat Province | Phrom Khiri | 4·924 | 5·095 | 18·725 | 1·641 | 6·305 | 1215 | 403 | 1554 |
| Nakhon Si Thammarat Province | Ron Phibun | 5·112 | 5·012 | 18·817 | 1·863 | 6·559 | 3272 | 1197 | 4209 |
| Nakhon Si Thammarat Province | Sichon | 4·618 | 5·028 | 18·508 | 1·476 | 5·834 | 3162 | 1028 | 4010 |
| Nakhon Si Thammarat Province | Tha Sala | 4·819 | 5·126 | 18·276 | 1·593 | 6·147 | 4703 | 1554 | 6012 |
| Nakhon Si Thammarat Province | Tham Phannara | 5·256 | 5·610 | 20·180 | 1·754 | 6·645 | 1198 | 402 | 1514 |
| Nakhon Si Thammarat Province | Thung Song | 5·324 | 5·326 | 19·710 | 1·894 | 6·830 | 7514 | 2607 | 9722 |
| Nakhon Si Thammarat Province | Thung Yai | 5·322 | 5·639 | 20·411 | 1·774 | 6·746 | 4215 | 1401 | 5356 |
| Nan Province | Ban Luang | 3·424 | 4·002 | 13·936 | 1·124 | 4·158 | 338 | 109 | 409 |
| Nan Province | Bo Kluea | 3·245 | 3·759 | 12·942 | 1·060 | 3·987 | 410 | 133 | 498 |
| Nan Province | Chaloem Phra Kiat | 3·555 | 4·380 | 14·918 | 1·067 | 4·312 | 369 | 112 | 450 |
| Nan Province | Chiang Klang | 3·439 | 4·231 | 13·995 | 1·110 | 4·160 | 738 | 237 | 890 |
| Nan Province | Mae Charim | 2·998 | 3·178 | 11·188 | 1·062 | 3·773 | 410 | 147 | 523 |
| Nan Province | Mueang Nan | 3·261 | 3·644 | 12·542 | 1·111 | 4·037 | 2847 | 983 | 3490 |
| Nan Province | Na Muen | 2·797 | 2·718 | 9·606 | 1·123 | 3·502 | 349 | 141 | 437 |
| Nan Province | Na Noi | 2·886 | 2·930 | 10·319 | 1·099 | 3·594 | 755 | 283 | 939 |
| Nan Province | Phu Phiang | 3·078 | 3·377 | 11·871 | 1·085 | 3·816 | 856 | 303 | 1063 |
| Nan Province | Pua | 3·301 | 3·877 | 13·270 | 1·082 | 4·068 | 1785 | 574 | 2171 |
| Nan Province | Santi Suk | 3·168 | 3·597 | 11·931 | 1·062 | 3·951 | 410 | 133 | 508 |
| Nan Province | Song Khwae | 3·622 | 4·518 | 14·933 | 1·120 | 4·397 | 364 | 112 | 437 |
| Nan Province | Tha Wang Pha | 3·373 | 3·962 | 13·154 | 1·113 | 4·147 | 1246 | 412 | 1566 |
| Nan Province | Thung Chang | 3·545 | 4·400 | 14·759 | 1·106 | 4·259 | 469 | 149 | 559 |
| Nan Province | Wiang Sa | 3·034 | 3·205 | 11·214 | 1·107 | 3·782 | 1998 | 726 | 2489 |
| Narathiwat Province | Bacho | 5·103 | 4·676 | 17·165 | 2·022 | 6·578 | 1725 | 675 | 2241 |
| Narathiwat Province | Chanae | 5·232 | 4·906 | 17·969 | 2·017 | 6·770 | 1680 | 643 | 2169 |
| Narathiwat Province | Cho-Airong | 5·102 | 4·933 | 18·198 | 1·938 | 6·586 | 1689 | 646 | 2184 |
| Narathiwat Province | Mueang Narathiwat | 5·236 | 5·130 | 18·047 | 1·938 | 6·765 | 2846 | 1067 | 3678 |
| Narathiwat Province | Ra-Ngae | 5·140 | 4·815 | 17·682 | 2·008 | 6·534 | 3484 | 1373 | 4427 |
| Narathiwat Province | Rueso | 5·134 | 4·658 | 16·866 | 2·077 | 6·530 | 3123 | 1263 | 3961 |
| Narathiwat Province | Si Sakhon | 5·218 | 4·797 | 17·703 | 2·059 | 6·652 | 2269 | 888 | 2869 |
| Narathiwat Province | Sukhirin | 5·226 | 5·075 | 18·497 | 1·930 | 6·703 | 1199 | 441 | 1535 |
| Narathiwat Province | Su-Ngai Kolok | 5·093 | 4·997 | 18·493 | 1·861 | 6·430 | 2862 | 1045 | 3567 |
| Narathiwat Province | Su-Ngai Padi | 5·065 | 4·935 | 18·204 | 1·881 | 6·416 | 2023 | 748 | 2584 |
| Narathiwat Province | Tak Bai | 5·144 | 5·140 | 18·433 | 1·860 | 6·554 | 1465 | 527 | 1863 |
| Narathiwat Province | Waeng | 5·175 | 5·002 | 18·524 | 1·911 | 6·625 | 2743 | 1017 | 3473 |
| Narathiwat Province | Yi-Ngo | 5·123 | 4·781 | 17·919 | 1·984 | 6·562 | 1715 | 678 | 2181 |
| Nong Bua Lam Phu Province | Mueang Nong Bua Lam Phu | 1·944 | 1·843 | 6·364 | 0·846 | 2·389 | 2469 | 1073 | 3021 |
| Nong Bua Lam Phu Province | Na Klang | 2·086 | 2·043 | 6·950 | 0·874 | 2·579 | 1550 | 653 | 1922 |
| Nong Bua Lam Phu Province | Na Wang | 2·090 | 1·984 | 7·016 | 0·881 | 2·585 | 724 | 307 | 891 |
| Nong Bua Lam Phu Province | Non Sang | 1·772 | 1·561 | 5·470 | 0·815 | 2·214 | 868 | 396 | 1093 |
| Nong Bua Lam Phu Province | Si Bun Rueang | 1·930 | 1·790 | 6·226 | 0·853 | 2·393 | 1806 | 796 | 2246 |
| Nong Bua Lam Phu Province | Suwannakhuha | 2·167 | 2·041 | 7·405 | 0·896 | 2·707 | 1413 | 584 | 1760 |
| Nong Khai Province | Fao Rai | 2·405 | 2·746 | 9·266 | 0·824 | 2·941 | 758 | 260 | 930 |
| Nong Khai Province | Mueang Nong Khai | 2·293 | 2·412 | 8·463 | 0·857 | 2·823 | 2817 | 1031 | 3440 |
| Nong Khai Province | Pho Tak | 2·405 | 2·476 | 8·954 | 0·879 | 3·006 | 286 | 103 | 359 |
| Nong Khai Province | Phon Phisai | 2·350 | 2·628 | 9·042 | 0·822 | 2·881 | 1914 | 662 | 2338 |

| ADM1 | ADM2 | β-thalassaemia carrier prevalence Mean | SD | CI95% | Q25 | Q75 | Estimated carrier counts Mean | Q25 | Q75 |
|---|---|---|---|---|---|---|---|---|---|
| Nong Khai Province | Rattanawapi | 2·559 | 2·917 | 10·388 | 0·852 | 3·111 | 724 | 242 | 877 |
| Nong Khai Province | Sakhrai | 2·190 | 2·203 | 7·890 | 0·861 | 2·718 | 511 | 200 | 642 |
| Nong Khai Province | Sangkhom | 2·515 | 2·662 | 9·496 | 0·898 | 3·131 | 400 | 139 | 495 |
| Nong Khai Province | Si Chiang Mai | 2·397 | 2·635 | 8·609 | 0·860 | 2·923 | 156 | 55 | 190 |
| Nong Khai Province | Tha Bo | 2·305 | 2·395 | 8·724 | 0·869 | 2·854 | 1652 | 615 | 2061 |
| Nonthaburi Province | Bang Bua Thong | 1·639 | 1·331 | 4·864 | 0·766 | 2·082 | 5712 | 2682 | 7262 |
| Nonthaburi Province | Bang Kruai | 1·596 | 1·283 | 4·863 | 0·762 | 1·982 | 5868 | 2782 | 7261 |
| Nonthaburi Province | Bang Yai | 1·633 | 1·375 | 5·028 | 0·762 | 2·022 | 5570 | 2623 | 6854 |
| Nonthaburi Province | Mueang Nonthaburi | 1·658 | 1·435 | 5·091 | 0·775 | 2·067 | 9115 | 4243 | 11361 |
| Nonthaburi Province | Pak Kret | 1·638 | 1·321 | 5·059 | 0·768 | 2·039 | 6101 | 2813 | 7578 |
| Nonthaburi Province | Sai Noi | 1·659 | 1·385 | 4·950 | 0·758 | 2·100 | 1467 | 680 | 1854 |
| Pathum Thani Province | Khlong Luang | 1·700 | 1·380 | 5·023 | 0·802 | 2·138 | 15790 | 7400 | 19748 |
| Pathum Thani Province | Lam Luk Ka | 1·680 | 1·309 | 4·918 | 0·826 | 2·084 | 6813 | 3310 | 8432 |
| Pathum Thani Province | Lat Lum Kaeo | 1·626 | 1·300 | 4·892 | 0·759 | 2·050 | 1887 | 890 | 2370 |
| Pathum Thani Province | Mueang Pathum Thani | 1·667 | 1·376 | 5·059 | 0·771 | 2·118 | 3871 | 1788 | 4920 |
| Pathum Thani Province | Nong Suea | 1·737 | 1·443 | 5·133 | 0·824 | 2·177 | 1103 | 524 | 1378 |
| Pathum Thani Province | Sam Khok | 1·632 | 1·365 | 4·716 | 0·756 | 2·096 | 1594 | 747 | 2059 |
| Pathum Thani Province | Thanyaburi | 1·668 | 1·317 | 4·926 | 0·810 | 2·081 | 9364 | 4506 | 11727 |
| Pattani Province | Kapho | 5·144 | 5·008 | 19·068 | 2·070 | 6·377 | 892 | 358 | 1107 |
| Pattani Province | Khok Pho | 5·220 | 4·376 | 16·553 | 2·312 | 6·664 | 2388 | 1060 | 3044 |
| Pattani Province | Mae Lan | 5·284 | 4·557 | 16·849 | 2·253 | 6·815 | 580 | 247 | 749 |
| Pattani Province | Mai Kaen | 5·095 | 4·565 | 16·488 | 2·068 | 6·646 | 767 | 313 | 999 |
| Pattani Province | Mayo | 5·136 | 4·573 | 16·980 | 2·135 | 6·569 | 2021 | 833 | 2582 |
| Pattani Province | Mueang Pattani | 5·271 | 4·772 | 17·064 | 2·270 | 6·710 | 4614 | 1987 | 5897 |
| Pattani Province | Nong Chik | 5·161 | 4·421 | 16·441 | 2·248 | 6·606 | 1972 | 852 | 2526 |
| Pattani Province | Panare | 5·103 | 4·691 | 17·642 | 2·060 | 6·539 | 1269 | 516 | 1624 |
| Pattani Province | Sai Buri | 5·122 | 4·693 | 17·630 | 2·006 | 6·613 | 3316 | 1285 | 4277 |
| Pattani Province | Thung Yang Daeng | 5·142 | 4·527 | 16·451 | 2·143 | 6·659 | 1055 | 444 | 1357 |
| Pattani Province | Yarang | 5·114 | 4·445 | 16·236 | 2·205 | 6·494 | 4343 | 1877 | 5495 |
| Pattani Province | Yaring | 5·096 | 4·494 | 16·505 | 2·118 | 6·546 | 3807 | 1591 | 4858 |
| Phangnga Province | Kapong | 5·262 | 6·057 | 22·099 | 1·522 | 6·606 | 758 | 220 | 974 |
| Phangnga Province | Khura Buri | 5·069 | 5·975 | 21·811 | 1·403 | 6·383 | 1011 | 281 | 1279 |
| Phangnga Province | Ko Yao | 5·672 | 6·212 | 23·012 | 1·803 | 7·196 | 330 | 105 | 417 |
| Phangnga Province | Mueang Phangnga | 5·385 | 6·085 | 21·956 | 1·609 | 6·782 | 2304 | 692 | 2964 |
| Phangnga Province | Takua Pa | 5·286 | 6·103 | 22·162 | 1·507 | 6·576 | 2120 | 607 | 2637 |
| Phangnga Province | Takua Thung | 5·599 | 6·364 | 22·988 | 1·668 | 7·034 | 2644 | 780 | 3337 |
| Phangnga Province | Thai Mueang | 5·486 | 6·347 | 23·354 | 1·597 | 6·829 | 2603 | 753 | 3216 |
| Phangnga Province | Thap Put | 5·360 | 6·058 | 21·874 | 1·600 | 6·750 | 943 | 283 | 1191 |
| Phatthalung Province | Bang Kaeo | 5·540 | 4·527 | 16·833 | 2·472 | 7·139 | 1417 | 631 | 1831 |
| Phatthalung Province | Khao Chaison | 5·464 | 4·583 | 16·963 | 2·397 | 7·015 | 1514 | 668 | 1938 |
| Phatthalung Province | Khuan Khanun | 5·268 | 4·579 | 16·951 | 2·227 | 6·764 | 3369 | 1427 | 4344 |
| Phatthalung Province | Kong Ra | 5·467 | 4·633 | 17·058 | 2·381 | 7·050 | 1802 | 784 | 2312 |
| Phatthalung Province | Mueang Phatthalung | 5·374 | 4·583 | 16·685 | 2·281 | 6·930 | 4680 | 1986 | 6008 |
| Phatthalung Province | Pa Bon | 5·535 | 4·539 | 16·405 | 2·495 | 7·136 | 2848 | 1290 | 3688 |
| Phatthalung Province | Pa Phayom | 5·266 | 4·702 | 17·432 | 2·099 | 6·851 | 1929 | 776 | 2492 |
| Phatthalung Province | Pak Phayun | 5·519 | 4·398 | 16·367 | 2·547 | 7·176 | 2133 | 983 | 2776 |
| Phatthalung Province | Si Banphot | 5·379 | 4·809 | 17·690 | 2·163 | 6·888 | 708 | 285 | 910 |
| Phatthalung Province | Srinagarindra | 5·396 | 4·614 | 17·209 | 2·320 | 7·091 | 1603 | 688 | 2114 |
| Phatthalung Province | Tamot | 5·452 | 4·494 | 16·574 | 2·462 | 6·908 | 1428 | 660 | 1810 |
| Phayao Province | Chiang Kham | 3·790 | 4·767 | 15·846 | 1·157 | 4·524 | 1901 | 576 | 2255 |
| Phayao Province | Chiang Muan | 3·554 | 4·166 | 14·393 | 1·160 | 4·353 | 464 | 152 | 565 |
| Phayao Province | Chun | 3·799 | 4·625 | 15·542 | 1·183 | 4·654 | 1206 | 372 | 1485 |
| Phayao Province | Dok Khamtai | 3·759 | 4·436 | 15·106 | 1·195 | 4·617 | 2442 | 766 | 3036 |
| Phayao Province | Mae Chai | 4·069 | 4·983 | 16·804 | 1·242 | 4·960 | 853 | 264 | 1040 |
| Phayao Province | Mueang Phayao | 3·877 | 4·641 | 15·952 | 1·228 | 4·680 | 3102 | 990 | 3746 |
| Phayao Province | Phu Kamyao | 3·837 | 4·638 | 15·585 | 1·193 | 4·655 | 362 | 111 | 429 |
| Phayao Province | Phu Sang | 3·898 | 4·954 | 16·552 | 1·150 | 4·727 | 836 | 251 | 1014 |

| ADM1 | ADM2 | β-thalassaemia carrier prevalence Mean | SD | CI95% | Q25 | Q75 | Estimated carrier counts Mean | Q25 | Q75 |
|---|---|---|---|---|---|---|---|---|---|
| Phayao Province | Pong | 3·604 | 4·328 | 14·537 | 1·158 | 4·377 | 1381 | 440 | 1660 |
| Phetchabun Province | Bueng Sam Phan | 1·953 | 1·657 | 5·944 | 0·884 | 2·460 | 1188 | 541 | 1490 |
| Phetchabun Province | Chon Daen | 2·017 | 1·640 | 5·965 | 0·956 | 2·540 | 1150 | 544 | 1451 |
| Phetchabun Province | Khao Kho | 2·014 | 1·544 | 5·675 | 1·000 | 2·533 | 705 | 349 | 888 |
| Phetchabun Province | Lam Sonthi | 1·883 | 1·676 | 6·007 | 0·815 | 2·371 | 486 | 216 | 618 |
| Phetchabun Province | Lom Kao | 2·046 | 1·662 | 6·062 | 0·975 | 2·568 | 1283 | 620 | 1610 |
| Phetchabun Province | Lom Sak | 1·986 | 1·598 | 5·810 | 0·946 | 2·512 | 2618 | 1264 | 3311 |
| Phetchabun Province | Mueang Phetchabun | 1·966 | 1·612 | 5·831 | 0·930 | 2·471 | 3476 | 1651 | 4363 |
| Phetchabun Province | Nam Nao | 1·970 | 1·658 | 5·981 | 0·917 | 2·464 | 313 | 143 | 391 |
| Phetchabun Province | Nong Phai | 1·949 | 1·659 | 6·042 | 0·892 | 2·449 | 1844 | 839 | 2304 |
| Phetchabun Province | Si Thep | 1·936 | 1·735 | 6·231 | 0·832 | 2·425 | 1211 | 516 | 1511 |
| Phetchabun Province | Wang Pong | 2·054 | 1·682 | 6·120 | 0·981 | 2·568 | 646 | 308 | 809 |
| Phetchabun Province | Wichian Buri | 1·938 | 1·655 | 6·107 | 0·859 | 2·460 | 2457 | 1082 | 3118 |
| Phetchaburi Province | Ban Laem | 1·807 | 1·691 | 5·986 | 0·733 | 2·264 | 894 | 363 | 1107 |
| Phetchaburi Province | Ban Lat | 1·920 | 1·823 | 6·591 | 0·751 | 2·453 | 850 | 346 | 1089 |
| Phetchaburi Province | Cha-Am | 2·029 | 2·034 | 7·232 | 0·758 | 2·564 | 1232 | 469 | 1553 |
| Phetchaburi Province | Kaeng Krachan | 2·261 | 2·351 | 8·529 | 0·786 | 2·855 | 657 | 235 | 827 |
| Phetchaburi Province | Khao Yoi | 1·889 | 1·832 | 6·689 | 0·751 | 2·371 | 988 | 394 | 1246 |
| Phetchaburi Province | Mueang Phetchaburi | 1·845 | 1·758 | 6·254 | 0·743 | 2·352 | 2153 | 865 | 2732 |
| Phetchaburi Province | Nong Ya Plong | 2·107 | 2·099 | 7·532 | 0·781 | 2·665 | 361 | 139 | 461 |
| Phetchaburi Province | Tha Yang | 2·050 | 2·058 | 7·345 | 0·766 | 2·599 | 1885 | 713 | 2378 |
| Phichit Province | Bang Mun Nak | 2·157 | 1·829 | 6·656 | 0·979 | 2·708 | 634 | 285 | 801 |
| Phichit Province | Bueng Na Rang | 2·273 | 1·890 | 7·084 | 1·052 | 2·857 | 612 | 283 | 765 |
| Phichit Province | Dong Charoen | 2·061 | 1·696 | 5·910 | 0·976 | 2·613 | 373 | 176 | 469 |
| Phichit Province | Mueang Phichit | 2·177 | 1·722 | 6·355 | 1·053 | 2·730 | 2411 | 1176 | 3011 |
| Phichit Province | Pho Prathap Chang | 2·295 | 1·927 | 6·976 | 1·067 | 2·884 | 991 | 461 | 1244 |
| Phichit Province | Pho Thale | 2·244 | 1·955 | 7·073 | 1·003 | 2·798 | 1284 | 572 | 1598 |
| Phichit Province | Sak Lek | 2·158 | 1·694 | 6·194 | 1·055 | 2·691 | 448 | 219 | 558 |
| Phichit Province | Sam Ngam | 2·306 | 1·879 | 6·724 | 1·087 | 2·893 | 944 | 444 | 1181 |
| Phichit Province | Tap Khlo | 2·098 | 1·757 | 6·284 | 0·983 | 2·620 | 723 | 340 | 901 |
| Phichit Province | Taphan Hin | 2·164 | 1·806 | 6·463 | 1·006 | 2·718 | 1194 | 560 | 1488 |
| Phichit Province | Wachirabarami | 2·361 | 1·915 | 6·915 | 1·120 | 2·966 | 979 | 464 | 1232 |
| Phichit Province | Wang Sai Phun | 2·157 | 1·759 | 6·247 | 1·027 | 2·727 | 432 | 207 | 545 |
| Phitsanulok Province | Bang Krathum | 2·202 | 1·701 | 6·159 | 1·093 | 2·772 | 1076 | 535 | 1353 |
| Phitsanulok Province | Bang Rakam | 2·390 | 1·901 | 7·007 | 1·166 | 2·996 | 2464 | 1196 | 3092 |
| Phitsanulok Province | Chat Trakan | 2·309 | 1·829 | 6·699 | 1·120 | 2·909 | 1227 | 596 | 1536 |
| Phitsanulok Province | Mueang Phitsanulok | 2·251 | 1·704 | 6·152 | 1·139 | 2·825 | 8046 | 4078 | 10042 |
| Phitsanulok Province | Nakhon Thai | 2·134 | 1·636 | 5·974 | 1·059 | 2·687 | 2182 | 1086 | 2732 |
| Phitsanulok Province | Noen Maprang | 2·089 | 1·620 | 5·975 | 1·028 | 2·617 | 1022 | 495 | 1285 |
| Phitsanulok Province | Phrom Phiram | 2·424 | 1·906 | 6·868 | 1·194 | 3·053 | 2360 | 1161 | 2959 |
| Phitsanulok Province | Wang Thong | 2·133 | 1·601 | 5·884 | 1·088 | 2·665 | 2367 | 1207 | 2967 |
| Phitsanulok Province | Wat Bot | 2·314 | 1·750 | 6·372 | 1·152 | 2·921 | 1022 | 519 | 1287 |
| Phra Nakhon Si Ayutthaya Province | Ban Phraek | 1·809 | 1·518 | 5·813 | 0·831 | 2·281 | 150 | 69 | 189 |
| Phra Nakhon Si Ayutthaya Province | Bang Ban | 1·688 | 1·434 | 5·132 | 0·767 | 2·102 | 588 | 267 | 736 |
| Phra Nakhon Si Ayutthaya Province | Bang Pahan | 1·744 | 1·495 | 5·256 | 0·794 | 2·211 | 629 | 286 | 796 |
| Phra Nakhon Si Ayutthaya Province | Bang Pa-In | 1·713 | 1·486 | 5·294 | 0·785 | 2·142 | 3692 | 1714 | 4649 |
| Phra Nakhon Si Ayutthaya Province | Bang Sai | 1·721 | 1·500 | 5·497 | 0·764 | 2·159 | 282 | 126 | 356 |
| Phra Nakhon Si Ayutthaya Province | Bang Sai | 1·703 | 1·474 | 5·431 | 0·766 | 2·151 | 708 | 319 | 894 |
| Phra Nakhon Si Ayutthaya Province | Lat Bua Luang | 1·684 | 1·421 | 5·111 | 0·769 | 2·108 | 656 | 304 | 828 |
| Phra Nakhon Si Ayutthaya Province | Maha Rat | 1·755 | 1·416 | 5·266 | 0·809 | 2·277 | 202 | 92 | 262 |
| Phra Nakhon Si Ayutthaya Province | Nakhon Luang | 1·773 | 1·434 | 5·305 | 0·831 | 2·229 | 1015 | 479 | 1278 |

| ADM1 | ADM2 | β-thalassaemia carrier prevalence Mean | SD | CI95% | Q25 | Q75 | Estimated carrier counts Mean | Q25 | Q75 |
|---|---|---|---|---|---|---|---|---|---|
| Phra Nakhon Si Ayutthaya Province | Phachi | 1·726 | 1·515 | 4·932 | 0·791 | 2·160 | 431 | 197 | 538 |
| Phra Nakhon Si Ayutthaya Province | Phak Hai | 1·749 | 1·531 | 5·606 | 0·770 | 2·202 | 413 | 181 | 518 |
| Phra Nakhon Si Ayutthaya Province | Phra Nakhon Si Ayutthaya | 1·716 | 1·449 | 5·274 | 0·782 | 2·165 | 3818 | 1740 | 4832 |
| Phra Nakhon Si Ayutthaya Province | Sena | 1·727 | 1·548 | 5·525 | 0·767 | 2·162 | 594 | 265 | 743 |
| Phra Nakhon Si Ayutthaya Province | Tha Ruea | 1·770 | 1·538 | 5·554 | 0·811 | 2·223 | 795 | 363 | 999 |
| Phra Nakhon Si Ayutthaya Province | Uthai | 1·732 | 1·410 | 5·148 | 0·810 | 2·174 | 2129 | 1008 | 2696 |
| Phra Nakhon Si Ayutthaya Province | Wang Noi | 1·744 | 1·441 | 5·354 | 0·803 | 2·176 | 1721 | 795 | 2151 |
| Phrae Province | Den Chai | 3·006 | 2·847 | 10·145 | 1·236 | 3·780 | 605 | 242 | 762 |
| Phrae Province | Long | 3·278 | 3·371 | 11·719 | 1·250 | 4·098 | 1242 | 468 | 1542 |
| Phrae Province | Mueang Phrae | 2·970 | 2·888 | 10·131 | 1·178 | 3·749 | 3036 | 1205 | 3813 |
| Phrae Province | Nong Muang Khai | 3·162 | 3·174 | 11·202 | 1·178 | 3·991 | 696 | 260 | 880 |
| Phrae Province | Rong Kwang | 3·092 | 3·216 | 11·172 | 1·153 | 3·855 | 1122 | 421 | 1391 |
| Phrae Province | Song | 3·303 | 3·566 | 12·441 | 1·185 | 4·102 | 1140 | 420 | 1419 |
| Phrae Province | Sung Men | 2·971 | 2·864 | 10·025 | 1·203 | 3·756 | 1998 | 809 | 2537 |
| Phrae Province | Wang Chin | 3·292 | 3·205 | 11·359 | 1·310 | 4·132 | 1126 | 454 | 1408 |
| Phuket Province | Kathu | 5·750 | 6·400 | 23·515 | 1·750 | 7·178 | 11568 | 3517 | 14457 |
| Phuket Province | Mueang Phuket | 5·697 | 6·239 | 22·885 | 1·751 | 7·296 | 20755 | 6339 | 26419 |
| Phuket Province | Thalang | 5·717 | 6·550 | 23·774 | 1·714 | 7·061 | 10251 | 3105 | 12517 |
| Prachin Buri Province | Ban Sang | 1·782 | 1·419 | 5·160 | 0·876 | 2·245 | 628 | 308 | 789 |
| Prachin Buri Province | Kabin Buri | 1·697 | 1·263 | 4·632 | 0·858 | 2·141 | 2479 | 1244 | 3125 |
| Prachin Buri Province | Mueang Prachin Buri | 1·778 | 1·375 | 5·124 | 0·867 | 2·230 | 2615 | 1283 | 3317 |
| Prachin Buri Province | Na Di | 1·708 | 1·337 | 4·971 | 0·836 | 2·149 | 1345 | 663 | 1688 |
| Prachin Buri Province | Prachantakham | 1·757 | 1·394 | 5·063 | 0·851 | 2·213 | 1215 | 605 | 1525 |
| Prachin Buri Province | Si Maha Phot | 1·757 | 1·310 | 4·814 | 0·887 | 2·213 | 3231 | 1620 | 4093 |
| Prachin Buri Province | Si Mahosot | 1·768 | 1·375 | 4·914 | 0·872 | 2·227 | 385 | 191 | 485 |
| Prachuap Khiri Khan Province | Bang Saphan | 3·054 | 3·613 | 13·005 | 0·899 | 3·767 | 1653 | 486 | 2048 |
| Prachuap Khiri Khan Province | Bang Saphan Noi | 3·285 | 3·968 | 14·012 | 0·942 | 4·007 | 855 | 246 | 1045 |
| Prachuap Khiri Khan Province | Hua Hin | 2·219 | 2·295 | 8·406 | 0·785 | 2·777 | 2723 | 984 | 3407 |
| Prachuap Khiri Khan Province | Kui Buri | 2·379 | 2·554 | 9·386 | 0·798 | 2·984 | 705 | 238 | 879 |
| Prachuap Khiri Khan Province | Mueang Prachuap Khiri Khan | 2·561 | 2·855 | 10·247 | 0·813 | 3·203 | 1682 | 525 | 2143 |
| Prachuap Khiri Khan Province | Pran Buri | 2·242 | 2·358 | 8·438 | 0·781 | 2·813 | 1165 | 409 | 1485 |
| Prachuap Khiri Khan Province | Sam Roi Yot | 2·291 | 2·412 | 8·689 | 0·789 | 2·872 | 889 | 309 | 1108 |
| Prachuap Khiri Khan Province | Thap Sakae | 2·848 | 3·298 | 11·760 | 0·864 | 3·523 | 845 | 259 | 1030 |
| Ranong Province | Kapoe | 4·657 | 5·447 | 19·769 | 1·305 | 5·810 | 1276 | 359 | 1604 |
| Ranong Province | Kra Buri | 3·966 | 4·813 | 17·592 | 1·088 | 4·874 | 2583 | 705 | 3170 |
| Ranong Province | La-Un | 4·202 | 5·106 | 18·340 | 1·157 | 5·118 | 1019 | 281 | 1246 |
| Ranong Province | Mueang Ranong | 4·497 | 5·479 | 20·186 | 1·205 | 5·491 | 8795 | 2393 | 10405 |
| Ranong Province | Suk Samran | 4·857 | 5·684 | 20·627 | 1·306 | 6·052 | 808 | 214 | 1016 |
| Ratchaburi Province | Ban Kha | 2·174 | 2·196 | 7·896 | 0·801 | 2·749 | 454 | 171 | 577 |
| Ratchaburi Province | Ban Pong | 1·801 | 1·667 | 5·958 | 0·755 | 2·253 | 2305 | 979 | 2897 |
| Ratchaburi Province | Bang Phae | 1·697 | 1·470 | 5·401 | 0·728 | 2·151 | 513 | 219 | 645 |
| Ratchaburi Province | Chom Bueng | 2·028 | 1·946 | 7·111 | 0·795 | 2·551 | 1126 | 445 | 1423 |
| Ratchaburi Province | Damnoen Saduak | 1·747 | 1·591 | 5·820 | 0·738 | 2·178 | 1461 | 618 | 1825 |
| Ratchaburi Province | Mueang Ratchaburi | 1·853 | 1·755 | 6·391 | 0·750 | 2·341 | 3376 | 1387 | 4312 |
| Ratchaburi Province | Pak Tho | 1·960 | 1·880 | 6·841 | 0·759 | 2·463 | 990 | 394 | 1253 |
| Ratchaburi Province | Photharam | 1·826 | 1·679 | 6·228 | 0·755 | 2·303 | 2583 | 1100 | 3240 |
| Ratchaburi Province | Suan Phueng | 2·243 | 2·306 | 8·278 | 0·816 | 2·818 | 1029 | 380 | 1292 |
| Ratchaburi Province | Wat Phleng | 1·775 | 1·517 | 5·837 | 0·767 | 2·265 | 208 | 90 | 265 |

| ADM1 | ADM2 | β-thalassaemia carrier prevalence Mean | SD | CI95% | Q25 | Q75 | Estimated carrier counts Mean | Q25 | Q75 |
|---|---|---|---|---|---|---|---|---|---|
| Rayong Province | Ban Chang | 1·698 | 1·497 | 5·443 | 0·713 | 2·187 | 1129 | 472 | 1447 |
| Rayong Province | Ban Khai | 1·634 | 1·400 | 5·088 | 0·736 | 2·083 | 1578 | 714 | 1998 |
| Rayong Province | Khao Chamao | 1·567 | 1·305 | 4·786 | 0·733 | 1·962 | 356 | 165 | 447 |
| Rayong Province | Klaeng | 1·552 | 1·333 | 4·742 | 0·703 | 1·968 | 1978 | 896 | 2499 |
| Rayong Province | Mueang Rayong | 1·638 | 1·441 | 5·163 | 0·718 | 2·067 | 10108 | 4382 | 12862 |
| Rayong Province | Nikhom Phatthana | 1·685 | 1·436 | 5·009 | 0·759 | 2·149 | 1656 | 746 | 2112 |
| Rayong Province | Pluak Daeng | 1·672 | 1·396 | 5·093 | 0·757 | 2·127 | 1572 | 706 | 1979 |
| Rayong Province | Wang Chan | 1·611 | 1·359 | 4·942 | 0·750 | 2·017 | 356 | 165 | 447 |
| Roi Et Province | At Samat | 1·050 | 0·916 | 3·321 | 0·465 | 1·328 | 458 | 203 | 579 |
| Roi Et Province | Changhan | 1·239 | 1·069 | 3·983 | 0·550 | 1·566 | 378 | 169 | 478 |
| Roi Et Province | Chaturaphak Phiman | 1·133 | 0·983 | 3·565 | 0·498 | 1·425 | 653 | 290 | 823 |
| Roi Et Province | Chiang Khwan | 1·195 | 1·042 | 3·919 | 0·530 | 1·478 | 258 | 114 | 321 |
| Roi Et Province | Kaset Wisai | 1·059 | 0·937 | 3·368 | 0·467 | 1·328 | 673 | 293 | 842 |
| Roi Et Province | Moei Wadi | 1·206 | 1·144 | 3·933 | 0·512 | 1·515 | 173 | 73 | 217 |
| Roi Et Province | Mueang Roi Et | 1·170 | 1·001 | 3·703 | 0·519 | 1·474 | 1662 | 740 | 2114 |
| Roi Et Province | Mueang Suang | 1·050 | 0·937 | 3·343 | 0·455 | 1·319 | 141 | 61 | 175 |
| Roi Et Province | Nong Hi | 0·937 | 0·850 | 3·101 | 0·399 | 1·183 | 153 | 64 | 191 |
| Roi Et Province | Nong Phok | 1·155 | 1·074 | 3·892 | 0·474 | 1·454 | 419 | 172 | 529 |
| Roi Et Province | Pathum Rat | 1·121 | 0·984 | 3·445 | 0·502 | 1·417 | 432 | 192 | 544 |
| Roi Et Province | Phanom Phrai | 0·946 | 0·837 | 3·026 | 0·408 | 1·201 | 518 | 222 | 653 |
| Roi Et Province | Pho Chai | 1·256 | 1·136 | 4·010 | 0·548 | 1·581 | 538 | 234 | 677 |
| Roi Et Province | Phon Sai | 0·909 | 0·803 | 2·901 | 0·390 | 1·154 | 149 | 64 | 189 |
| Roi Et Province | Phon Thong | 1·195 | 1·092 | 3·902 | 0·515 | 1·504 | 834 | 359 | 1054 |
| Roi Et Province | Selaphum | 1·102 | 0·978 | 3·571 | 0·476 | 1·392 | 737 | 321 | 936 |
| Roi Et Province | Si Somdet | 1·223 | 1·081 | 3·791 | 0·544 | 1·558 | 303 | 134 | 385 |
| Roi Et Province | Suwannaphum | 0·998 | 0·918 | 3·201 | 0·431 | 1·258 | 728 | 319 | 926 |
| Roi Et Province | Thawat Buri | 1·122 | 0·961 | 3·535 | 0·499 | 1·417 | 615 | 272 | 778 |
| Roi Et Province | Thung Khao Luang | 1·108 | 0·941 | 3·523 | 0·487 | 1·404 | 135 | 59 | 169 |
| Sa Kaeo Province | Aranyaprathet | 1·273 | 1·007 | 3·653 | 0·617 | 1·598 | 1918 | 938 | 2397 |
| Sa Kaeo Province | Khao Chakan | 1·528 | 1·154 | 4·207 | 0·770 | 1·921 | 860 | 432 | 1077 |
| Sa Kaeo Province | Khlong Hat | 1·349 | 1·067 | 3·921 | 0·655 | 1·696 | 552 | 269 | 697 |
| Sa Kaeo Province | Khok Sung | 1·224 | 0·996 | 3·683 | 0·584 | 1·531 | 349 | 165 | 437 |
| Sa Kaeo Province | Mueang Sa Kaeo | 1·540 | 1·158 | 4·282 | 0·771 | 1·939 | 1717 | 866 | 2169 |
| Sa Kaeo Province | Ta Phraya | 1·206 | 0·985 | 3·576 | 0·569 | 1·520 | 729 | 343 | 923 |
| Sa Kaeo Province | Wang Nam Yen | 1·469 | 1·109 | 4·207 | 0·737 | 1·857 | 986 | 490 | 1242 |
| Sa Kaeo Province | Wang Sombun | 1·430 | 1·150 | 4·173 | 0·693 | 1·792 | 545 | 264 | 678 |
| Sa Kaeo Province | Watthana Nakhon | 1·399 | 1·123 | 4·071 | 0·674 | 1·754 | 1136 | 560 | 1431 |
| Sakon Nakhon Province | Akat Amnuai | 2·055 | 2·313 | 8·055 | 0·721 | 2·510 | 1080 | 384 | 1326 |
| Sakon Nakhon Province | Ban Muang | 2·293 | 2·559 | 8·910 | 0·790 | 2·825 | 1000 | 344 | 1228 |
| Sakon Nakhon Province | Charoen Sin | 2·108 | 2·356 | 8·218 | 0·757 | 2·565 | 588 | 212 | 721 |
| Sakon Nakhon Province | Kham Ta Kla | 2·220 | 2·541 | 8·543 | 0·763 | 2·727 | 607 | 210 | 745 |
| Sakon Nakhon Province | Khok Si Suphan | 1·636 | 1·760 | 6·158 | 0·590 | 1·992 | 437 | 156 | 529 |
| Sakon Nakhon Province | Kusuman | 1·868 | 2·115 | 7·455 | 0·646 | 2·278 | 612 | 211 | 746 |
| Sakon Nakhon Province | Kut Bak | 1·647 | 1·643 | 5·889 | 0·649 | 2·030 | 308 | 123 | 377 |
| Sakon Nakhon Province | Mueang Sakon Nakhon | 1·726 | 1·842 | 6·432 | 0·629 | 2·142 | 2664 | 956 | 3309 |
| Sakon Nakhon Province | Nikhom Nam Un | 1·736 | 1·733 | 6·352 | 0·685 | 2·150 | 219 | 85 | 272 |
| Sakon Nakhon Province | Phang Khon | 1·896 | 2·015 | 7·072 | 0·694 | 2·342 | 687 | 253 | 844 |
| Sakon Nakhon Province | Phanna Nikhom | 1·849 | 1·983 | 7·007 | 0·683 | 2·261 | 942 | 341 | 1150 |
| Sakon Nakhon Province | Phon Na Kaeo | 1·778 | 1·958 | 6·919 | 0·610 | 2·194 | 468 | 162 | 575 |
| Sakon Nakhon Province | Phu Phan | 1·582 | 1·624 | 5·780 | 0·610 | 1·940 | 394 | 151 | 482 |
| Sakon Nakhon Province | Sawang Daen Din | 1·991 | 2·148 | 7·223 | 0·749 | 2·439 | 1970 | 741 | 2410 |
| Sakon Nakhon Province | Song Dao | 1·868 | 1·885 | 6·638 | 0·733 | 2·329 | 434 | 171 | 543 |
| Sakon Nakhon Province | Tao Ngoi | 1·546 | 1·617 | 5·869 | 0·573 | 1·918 | 329 | 120 | 414 |
| Sakon Nakhon Province | Wanon Niwat | 2·089 | 2·307 | 8·045 | 0·731 | 2·565 | 1874 | 656 | 2307 |
| Sakon Nakhon Province | Waritchaphum | 1·828 | 1·938 | 6·744 | 0·706 | 2·224 | 564 | 217 | 690 |
| Samut Prakan Province | Bang Bo | 1·664 | 1·354 | 4·929 | 0·789 | 2·095 | 3061 | 1481 | 3926 |
| Samut Prakan Province | Bang Phli | 1·654 | 1·330 | 4·850 | 0·781 | 2·108 | 10435 | 4927 | 13322 |

| ADM1 | ADM2 | β-thalassaemia carrier prevalence Mean | SD | CI95% | Q25 | Q75 | Estimated carrier counts Mean | Q25 | Q75 |
|---|---|---|---|---|---|---|---|---|---|
| Samut Prakan Province | Bang Sao Thong | 1·647 | 1·324 | 4·758 | 0·789 | 2·059 | 8251 | 3940 | 10363 |
| Samut Prakan Province | Mueang Samut Prakan | 1·648 | 1·352 | 4·940 | 0·776 | 2·093 | 20364 | 9767 | 25902 |
| Samut Prakan Province | Phra Pradaeng | 1·615 | 1·311 | 4·703 | 0·781 | 1·988 | 7101 | 3433 | 8737 |
| Samut Prakan Province | Phra Samut Chedi | 1·635 | 1·343 | 5·001 | 0·732 | 2·114 | 1700 | 757 | 2188 |
| Samut Sakhon Province | Ban Phaeo | 1·667 | 1·425 | 5·314 | 0·719 | 2·132 | 1583 | 684 | 2026 |
| Samut Sakhon Province | Krathum Baen | 1·582 | 1·296 | 4·891 | 0·722 | 1·988 | 5507 | 2518 | 6942 |
| Samut Sakhon Province | Mueang Samut Sakhon | 1·653 | 1·429 | 5·246 | 0·738 | 2·095 | 17261 | 7754 | 21510 |
| Samut Songkhram Province | Amphawa | 1·768 | 1·564 | 5·735 | 0·730 | 2·252 | 807 | 334 | 1039 |
| Samut Songkhram Province | Bang Khonthi | 1·722 | 1·563 | 5·672 | 0·726 | 2·178 | 429 | 177 | 542 |
| Samut Songkhram Province | Mueang Samut Songkhram | 1·752 | 1·579 | 5·726 | 0·729 | 2·213 | 1343 | 569 | 1685 |
| Saraburi Province | Ban Mo | 1·819 | 1·564 | 5·570 | 0·819 | 2·287 | 608 | 276 | 773 |
| Saraburi Province | Chaloem Phra Kiat | 1·847 | 1·610 | 5·725 | 0·832 | 2·318 | 1474 | 676 | 1865 |
| Saraburi Province | Don Phut | 1·765 | 1·493 | 5·488 | 0·824 | 2·193 | 164 | 76 | 204 |
| Saraburi Province | Kaeng Khoi | 1·842 | 1·553 | 5·613 | 0·844 | 2·318 | 2801 | 1271 | 3492 |
| Saraburi Province | Muak Lek | 1·834 | 1·584 | 5·676 | 0·825 | 2·309 | 1379 | 623 | 1729 |
| Saraburi Province | Mueang Saraburi | 1·789 | 1·484 | 5·385 | 0·822 | 2·239 | 1766 | 798 | 2207 |
| Saraburi Province | Nong Don | 1·851 | 1·693 | 6·035 | 0·826 | 2·319 | 303 | 136 | 377 |
| Saraburi Province | Nong Khae | 1·776 | 1·513 | 5·354 | 0·840 | 2·212 | 2328 | 1102 | 2911 |
| Saraburi Province | Nong Saeng | 1·795 | 1·524 | 5·593 | 0·826 | 2·222 | 608 | 282 | 754 |
| Saraburi Province | Phra Phutthabat | 1·854 | 1·633 | 5·997 | 0·805 | 2·326 | 1165 | 496 | 1452 |
| Saraburi Province | Sao Hai | 1·821 | 1·527 | 5·585 | 0·827 | 2·297 | 1107 | 500 | 1396 |
| Saraburi Province | Wang Muang | 1·828 | 1·558 | 5·703 | 0·818 | 2·319 | 495 | 223 | 627 |
| Saraburi Province | Wihan Daeng | 1·815 | 1·527 | 5·618 | 0·838 | 2·256 | 943 | 438 | 1175 |
| Satun Province | Khuan Don | 5·738 | 4·838 | 18·125 | 2·493 | 7·272 | 1284 | 569 | 1642 |
| Satun Province | Khuan Kalong | 5·612 | 4·618 | 17·225 | 2·540 | 7·160 | 1503 | 680 | 1921 |
| Satun Province | La-Ngu | 5·809 | 5·133 | 19·086 | 2·402 | 7·427 | 3741 | 1564 | 4760 |
| Satun Province | Manang | 5·724 | 4·952 | 18·161 | 2·480 | 7·285 | 835 | 356 | 1064 |
| Satun Province | Mueang Satun | 5·836 | 5·149 | 19·018 | 2·451 | 7·442 | 5666 | 2415 | 7174 |
| Satun Province | Tha Phae | 5·803 | 4·989 | 18·285 | 2·477 | 7·437 | 1611 | 698 | 2063 |
| Satun Province | Thung Wa | 5·715 | 5·048 | 18·587 | 2·406 | 7·253 | 1223 | 512 | 1545 |
| Si Sa Ket Province | Benchalak | 0·548 | 0·457 | 1·657 | 0·255 | 0·687 | 104 | 49 | 131 |
| Si Sa Ket Province | Bueng Bun | 0·845 | 0·788 | 2·797 | 0·362 | 1·038 | 28 | 12 | 35 |
| Si Sa Ket Province | Huai Thap Than | 0·747 | 0·648 | 2·381 | 0·338 | 0·929 | 133 | 61 | 165 |
| Si Sa Ket Province | Kantharalak | 0·502 | 0·412 | 1·491 | 0·239 | 0·632 | 535 | 251 | 666 |
| Si Sa Ket Province | Kanthararom | 0·661 | 0·567 | 2·060 | 0·298 | 0·837 | 347 | 155 | 441 |
| Si Sa Ket Province | Khukhan | 0·636 | 0·520 | 1·920 | 0·302 | 0·799 | 427 | 201 | 538 |
| Si Sa Ket Province | Khun Han | 0·543 | 0·447 | 1·597 | 0·261 | 0·680 | 328 | 160 | 414 |
| Si Sa Ket Province | Mueang Chan | 0·797 | 0·706 | 2·477 | 0·354 | 1·007 | 92 | 41 | 116 |
| Si Sa Ket Province | Mueang Si Sa Ket | 0·688 | 0·591 | 2·134 | 0·309 | 0·871 | 631 | 283 | 798 |
| Si Sa Ket Province | Nam Kliang | 0·603 | 0·495 | 1·830 | 0·286 | 0·755 | 159 | 76 | 199 |
| Si Sa Ket Province | Non Khun | 0·568 | 0·472 | 1·723 | 0·263 | 0·716 | 149 | 69 | 188 |
| Si Sa Ket Province | Phayu | 0·612 | 0·510 | 1·866 | 0·283 | 0·766 | 79 | 37 | 98 |
| Si Sa Ket Province | Pho Si Suwan | 0·800 | 0·699 | 2·525 | 0·356 | 1·003 | 74 | 33 | 91 |
| Si Sa Ket Province | Phrai Bueng | 0·602 | 0·495 | 1·810 | 0·283 | 0·757 | 187 | 90 | 236 |
| Si Sa Ket Province | Phu Sing | 0·611 | 0·501 | 1·803 | 0·293 | 0·767 | 191 | 90 | 237 |
| Si Sa Ket Province | Prang Ku | 0·722 | 0·612 | 2·176 | 0·329 | 0·917 | 250 | 115 | 318 |
| Si Sa Ket Province | Rasi Salai | 0·814 | 0·715 | 2·567 | 0·357 | 1·027 | 344 | 150 | 435 |
| Si Sa Ket Province | Si Rattana | 0·567 | 0·456 | 1·628 | 0·267 | 0·719 | 121 | 58 | 155 |
| Si Sa Ket Province | Sila Lat | 0·875 | 0·784 | 2·679 | 0·376 | 1·107 | 46 | 19 | 58 |
| Si Sa Ket Province | Uthumphon Phisai | 0·752 | 0·664 | 2·348 | 0·330 | 0·950 | 348 | 152 | 443 |
| Si Sa Ket Province | Wang Hin | 0·685 | 0·571 | 2·078 | 0·319 | 0·862 | 175 | 81 | 221 |
| Si Sa Ket Province | Yang Chum Noi | 0·745 | 0·633 | 2·311 | 0·332 | 0·956 | 142 | 63 | 183 |
| Sing Buri Province | Bang Rachan | 1·924 | 1·707 | 6·096 | 0·843 | 2·433 | 584 | 256 | 739 |
| Sing Buri Province | In Buri | 1·963 | 1·793 | 6·388 | 0·854 | 2·447 | 763 | 334 | 947 |
| Sing Buri Province | Khai Bang Rachan | 1·893 | 1·655 | 6·158 | 0·838 | 2·387 | 514 | 227 | 648 |
| Sing Buri Province | Mueang Sing Buri | 1·911 | 1·708 | 6·146 | 0·828 | 2·360 | 648 | 278 | 789 |

| ADM1 | ADM2 | β-thalassaemia carrier prevalence Mean | SD | CI95% | Q25 | Q75 | Estimated carrier counts Mean | Q25 | Q75 |
|---|---|---|---|---|---|---|---|---|---|
| Sing Buri Province | Phrom Buri | 1·890 | 1·585 | 5·948 | 0·854 | 2·404 | 539 | 243 | 684 |
| Sing Buri Province | Tha Chang | 1·873 | 1·609 | 5·739 | 0·822 | 2·360 | 286 | 125 | 360 |
| Songkhla Province | Bang Klam | 5·517 | 4·329 | 16·321 | 2·584 | 7·092 | 2732 | 1309 | 3474 |
| Songkhla Province | Chana | 5·432 | 4·213 | 15·652 | 2·580 | 6·973 | 5847 | 2797 | 7511 |
| Songkhla Province | Hat Yai | 5·562 | 4·357 | 16·127 | 2·606 | 7·134 | 28662 | 13585 | 36720 |
| Songkhla Province | Khlong Hoi Khong | 5·584 | 4·306 | 16·039 | 2·648 | 7·209 | 1851 | 877 | 2412 |
| Songkhla Province | Khuan Niang | 5·547 | 4·391 | 16·468 | 2·602 | 7·083 | 2111 | 981 | 2698 |
| Songkhla Province | Krasae Sin | 5·235 | 4·417 | 16·412 | 2·340 | 6·714 | 464 | 208 | 593 |
| Songkhla Province | Mueang Songkhla | 5·555 | 4·319 | 15·974 | 2·645 | 7·125 | 6139 | 2996 | 7959 |
| Songkhla Province | Na Mom | 5·492 | 4·168 | 15·816 | 2·629 | 7·029 | 3154 | 1509 | 4066 |
| Songkhla Province | Na Thawi | 5·459 | 4·366 | 16·147 | 2·505 | 6·987 | 4148 | 1919 | 5308 |
| Songkhla Province | Ranot | 5·117 | 4·471 | 16·635 | 2·133 | 6·641 | 2177 | 906 | 2822 |
| Songkhla Province | Rattaphum | 5·549 | 4·440 | 16·612 | 2·531 | 7·135 | 3465 | 1580 | 4454 |
| Songkhla Province | Saba Yoi | 5·341 | 4·349 | 16·186 | 2·404 | 6·905 | 4084 | 1841 | 5293 |
| Songkhla Province | Sadao | 5·570 | 4·427 | 16·435 | 2·590 | 7·130 | 7903 | 3708 | 10117 |
| Songkhla Province | Sathing Phra | 5·286 | 4·119 | 15·488 | 2·454 | 6·859 | 1356 | 629 | 1762 |
| Songkhla Province | Singhanakhon | 5·418 | 4·223 | 15·711 | 2·551 | 6·962 | 3797 | 1790 | 4897 |
| Songkhla Province | Thepha | 5·276 | 4·210 | 15·769 | 2·447 | 6·748 | 3661 | 1703 | 4694 |
| Sukhothai Province | Ban Dan Lan Hoi | 2·942 | 2·494 | 9·242 | 1·316 | 3·721 | 1465 | 663 | 1861 |
| Sukhothai Province | Khiri Mat | 2·660 | 2·260 | 8·019 | 1·221 | 3·356 | 1759 | 808 | 2210 |
| Sukhothai Province | Kong Krailat | 2·481 | 1·951 | 7·259 | 1·192 | 3·141 | 1642 | 785 | 2076 |
| Sukhothai Province | Mueang Sukhothai | 2·719 | 2·274 | 8·256 | 1·253 | 3·436 | 2629 | 1225 | 3309 |
| Sukhothai Province | Sawankhalok | 2·765 | 2·312 | 8·335 | 1·279 | 3·488 | 2466 | 1135 | 3118 |
| Sukhothai Province | Si Nakhon | 2·734 | 2·333 | 8·498 | 1·245 | 3·448 | 804 | 369 | 1004 |
| Sukhothai Province | Si Samrong | 2·768 | 2·295 | 8·297 | 1·282 | 3·508 | 2031 | 948 | 2570 |
| Sukhothai Province | Si Satchanalai | 3·061 | 2·801 | 9·962 | 1·311 | 3·848 | 2807 | 1242 | 3544 |
| Sukhothai Province | Thung Saliam | 3·097 | 2·779 | 9·898 | 1·352 | 3·891 | 1478 | 646 | 1861 |
| Suphan Buri Province | Bang Pla Ma | 1·781 | 1·585 | 5·684 | 0·792 | 2·228 | 1193 | 531 | 1491 |
| Suphan Buri Province | Dan Chang | 2·265 | 2·101 | 7·602 | 0·933 | 2·862 | 1300 | 545 | 1648 |
| Suphan Buri Province | Doem Bang Nang Buat | 1·997 | 1·793 | 6·429 | 0·862 | 2·509 | 1255 | 544 | 1579 |
| Suphan Buri Province | Don Chedi | 1·975 | 1·824 | 6·487 | 0·828 | 2·491 | 881 | 369 | 1109 |
| Suphan Buri Province | Mueang Suphan Buri | 1·853 | 1·668 | 5·973 | 0·792 | 2·352 | 2918 | 1249 | 3654 |
| Suphan Buri Province | Nong Ya Sai | 2·046 | 1·856 | 6·643 | 0·862 | 2·585 | 970 | 408 | 1229 |
| Suphan Buri Province | Sam Chuk | 1·933 | 1·724 | 6·197 | 0·823 | 2·460 | 906 | 385 | 1162 |
| Suphan Buri Province | Si Prachan | 1·874 | 1·655 | 5·850 | 0·820 | 2·366 | 1164 | 506 | 1476 |
| Suphan Buri Province | Song Phi Nong | 1·798 | 1·618 | 5·753 | 0·771 | 2·279 | 2309 | 993 | 2913 |
| Suphan Buri Province | U Thong | 1·884 | 1·684 | 6·082 | 0·803 | 2·386 | 2048 | 875 | 2603 |
| Surat Thani Province | Ban Na Doem | 4·876 | 5·440 | 19·604 | 1·487 | 6·084 | 1007 | 310 | 1262 |
| Surat Thani Province | Ban Na San | 4·908 | 5·330 | 19·949 | 1·546 | 6·203 | 3406 | 1075 | 4288 |
| Surat Thani Province | Ban Ta Khun | 4·994 | 5·739 | 20·482 | 1·430 | 6·357 | 846 | 245 | 1096 |
| Surat Thani Province | Chai Buri | 5·274 | 5·793 | 20·742 | 1·675 | 6·672 | 1766 | 554 | 2220 |
| Surat Thani Province | Chaiya | 4·562 | 5·245 | 19·158 | 1·316 | 5·756 | 2313 | 673 | 2928 |
| Surat Thani Province | Don Sak | 4·402 | 4·847 | 17·790 | 1·398 | 5·525 | 726 | 230 | 911 |
| Surat Thani Province | Kanchanadit | 4·652 | 5·168 | 19·020 | 1·433 | 5·843 | 5384 | 1642 | 6687 |
| Surat Thani Province | Khian Sa | 5·126 | 5·716 | 20·616 | 1·564 | 6·497 | 2284 | 697 | 2903 |
| Surat Thani Province | Khiri Rat Nikhom | 4·989 | 5·679 | 20·354 | 1·460 | 6·328 | 2205 | 647 | 2804 |
| Surat Thani Province | Ko Pha-Ngan | 3·754 | 4·190 | 15·548 | 1·164 | 4·640 | 149 | 46 | 185 |
| Surat Thani Province | Ko Samui | 3·996 | 4·419 | 16·113 | 1·231 | 4·949 | 1933 | 597 | 2412 |
| Surat Thani Province | Mueang Surat Thani | 4·735 | 5·247 | 19·159 | 1·418 | 5·985 | 8083 | 2374 | 10135 |
| Surat Thani Province | Phanom | 5·192 | 5·873 | 21·292 | 1·545 | 6·569 | 2158 | 646 | 2718 |
| Surat Thani Province | Phrasaeng | 5·233 | 5·735 | 20·856 | 1·650 | 6·656 | 3682 | 1162 | 4692 |
| Surat Thani Province | Phunphin | 4·825 | 5·352 | 19·262 | 1·462 | 6·158 | 4229 | 1278 | 5396 |
| Surat Thani Province | Tha Chana | 4·459 | 5·153 | 18·861 | 1·262 | 5·620 | 2504 | 708 | 3157 |
| Surat Thani Province | Tha Chang | 4·666 | 5·356 | 19·595 | 1·348 | 5·853 | 1625 | 479 | 2049 |
| Surat Thani Province | Vibhavadi | 4·793 | 5·494 | 19·696 | 1·375 | 6·063 | 815 | 238 | 1017 |
| Surat Thani Province | Wiang Sa | 5·185 | 5·749 | 20·639 | 1·637 | 6·548 | 3427 | 1072 | 4306 |
| Surin Province | Buachet | 0·671 | 0·559 | 2·066 | 0·315 | 0·833 | 217 | 101 | 270 |

| ADM1 | ADM2 | β-thalassaemia carrier prevalence Mean | SD | CI95% | Q25 | Q75 | Estimated carrier counts Mean | Q25 | Q75 |
|---|---|---|---|---|---|---|---|---|---|
| Surin Province | Chom Phra | 0·929 | 0·828 | 2·871 | 0·413 | 1·169 | 341 | 152 | 431 |
| Surin Province | Chumphon Buri | 1·081 | 0·947 | 3·481 | 0·478 | 1·364 | 507 | 224 | 636 |
| Surin Province | Kap Choeng | 0·832 | 0·709 | 2·536 | 0·383 | 1·047 | 383 | 177 | 485 |
| Surin Province | Khwao Sin Rin | 0·902 | 0·783 | 2·818 | 0·400 | 1·133 | 212 | 94 | 268 |
| Surin Province | Lamduan | 0·812 | 0·705 | 2·554 | 0·369 | 1·011 | 154 | 70 | 191 |
| Surin Province | Mueang Surin | 0·911 | 0·794 | 2·844 | 0·409 | 1·144 | 1871 | 847 | 2336 |
| Surin Province | Non Narai | 0·833 | 0·725 | 2·509 | 0·378 | 1·051 | 162 | 74 | 206 |
| Surin Province | Phanom Dong Rak | 0·939 | 0·795 | 2·916 | 0·437 | 1·171 | 224 | 104 | 279 |
| Surin Province | Prasat | 0·916 | 0·797 | 2·857 | 0·416 | 1·147 | 908 | 415 | 1135 |
| Surin Province | Rattanaburi | 0·887 | 0·775 | 2·776 | 0·394 | 1·123 | 514 | 229 | 651 |
| Surin Province | Samrong Thap | 0·783 | 0·667 | 2·374 | 0·355 | 0·981 | 216 | 99 | 271 |
| Surin Province | Sangkha | 0·746 | 0·653 | 2·358 | 0·340 | 0·926 | 631 | 285 | 776 |
| Surin Province | Sanom | 0·881 | 0·776 | 2·750 | 0·402 | 1·118 | 189 | 87 | 241 |
| Surin Province | Si Narong | 0·747 | 0·650 | 2·312 | 0·341 | 0·939 | 217 | 99 | 270 |
| Surin Province | Sikhoraphum | 0·820 | 0·710 | 2·553 | 0·372 | 1·029 | 640 | 288 | 801 |
| Surin Province | Tha Tum | 0·969 | 0·850 | 3·020 | 0·426 | 1·224 | 570 | 252 | 720 |
| Tak Province | Ban Tak | 3·431 | 3·118 | 11·137 | 1·457 | 4·331 | 1511 | 644 | 1903 |
| Tak Province | Mae Ramat | 4·047 | 3·871 | 13·901 | 1·642 | 5·078 | 1915 | 780 | 2410 |
| Tak Province | Mae Sot | 3·805 | 3·556 | 12·870 | 1·562 | 4·785 | 6492 | 2639 | 8188 |
| Tak Province | Mueang Tak | 3·271 | 2·922 | 10·566 | 1·404 | 4·128 | 3076 | 1302 | 3866 |
| Tak Province | Phop Phra | 3·557 | 3·330 | 12·077 | 1·458 | 4·485 | 2567 | 1056 | 3259 |
| Tak Province | Sam Ngao | 3·909 | 3·753 | 13·473 | 1·572 | 4·903 | 1048 | 435 | 1332 |
| Tak Province | Tha Song Yang | 4·811 | 4·592 | 16·872 | 1·921 | 6·051 | 2318 | 944 | 2914 |
| Tak Province | Umphang | 3·263 | 3·111 | 11·236 | 1·317 | 4·094 | 1059 | 419 | 1332 |
| Tak Province | Wang Chao | 3·254 | 2·995 | 10·731 | 1·379 | 4·066 | 959 | 414 | 1190 |
| Trang Province | Hat Samran | 5·738 | 5·061 | 19·350 | 2·339 | 7·524 | 556 | 226 | 733 |
| Trang Province | Huai Yot | 5·443 | 5·180 | 19·172 | 2·063 | 6·941 | 3620 | 1353 | 4629 |
| Trang Province | Kantang | 5·782 | 5·506 | 20·265 | 2·195 | 7·392 | 4516 | 1744 | 5764 |
| Trang Province | Mueang Trang | 5·466 | 5·083 | 19·209 | 2·155 | 7·024 | 7450 | 2976 | 9534 |
| Trang Province | Na Yong | 5·486 | 4·881 | 17·795 | 2·260 | 7·035 | 2247 | 919 | 2901 |
| Trang Province | Palian | 5·621 | 4·897 | 18·317 | 2·380 | 7·166 | 3189 | 1334 | 4057 |
| Trang Province | Ratsada | 5·401 | 5·189 | 19·315 | 1·983 | 6·920 | 1524 | 554 | 1946 |
| Trang Province | Sikao | 5·672 | 5·563 | 20·718 | 2·111 | 7·124 | 2232 | 836 | 2793 |
| Trang Province | Wang Wiset | 5·525 | 5·378 | 19·924 | 2·046 | 7·106 | 2231 | 831 | 2877 |
| Trang Province | Yan Ta Khao | 5·584 | 4·910 | 18·397 | 2·313 | 7·180 | 2983 | 1225 | 3834 |
| Trat Province | Bo Rai | 1·099 | 0·959 | 3·379 | 0·495 | 1·382 | 309 | 139 | 387 |
| Trat Province | Khao Saming | 1·178 | 1·016 | 3·658 | 0·525 | 1·494 | 410 | 184 | 523 |
| Trat Province | Khlong Yai | 1·109 | 1·144 | 3·392 | 0·457 | 1·401 | 226 | 92 | 289 |
| Trat Province | Ko Chang | 1·324 | 1·337 | 4·593 | 0·522 | 1·643 | 146 | 58 | 178 |
| Trat Province | Ko Kut | 1·277 | 1·259 | 4·507 | 0·496 | 1·588 | 17 | 7 | 21 |
| Trat Province | Laem Ngop | 1·269 | 1·154 | 4·231 | 0·526 | 1·625 | 103 | 43 | 132 |
| Trat Province | Mueang Trat | 1·113 | 0·975 | 3·515 | 0·491 | 1·411 | 1140 | 500 | 1452 |
| Ubon Ratchathani Province | Buntharik | 0·540 | 0·479 | 1·725 | 0·238 | 0·680 | 430 | 191 | 546 |
| Ubon Ratchathani Province | Det Udom | 0·551 | 0·464 | 1·698 | 0·252 | 0·696 | 813 | 368 | 1029 |
| Ubon Ratchathani Province | Don Mot Daeng | 0·735 | 0·711 | 2·472 | 0·298 | 0·917 | 167 | 68 | 208 |
| Ubon Ratchathani Province | Khemarat | 1·010 | 1·080 | 3·671 | 0·373 | 1·249 | 528 | 194 | 649 |
| Ubon Ratchathani Province | Khong Chiam | 0·790 | 0·814 | 2·820 | 0·304 | 0·985 | 197 | 76 | 247 |
| Ubon Ratchathani Province | Khueang Nai | 0·769 | 0·697 | 2·534 | 0·327 | 0·971 | 675 | 287 | 855 |
| Ubon Ratchathani Province | Kut Khaopun | 0·926 | 0·940 | 3·367 | 0·360 | 1·141 | 329 | 127 | 407 |
| Ubon Ratchathani Province | Lao Suea Kok | 0·748 | 0·683 | 2·476 | 0·309 | 0·947 | 154 | 63 | 195 |
| Ubon Ratchathani Province | Muang Sam Sip | 0·801 | 0·766 | 2·696 | 0·326 | 1·016 | 616 | 252 | 779 |
| Ubon Ratchathani Province | Mueang Ubon Ratchathani | 0·704 | 0·639 | 2·298 | 0·296 | 0·892 | 1837 | 780 | 2349 |
| Ubon Ratchathani Province | Na Chaluai | 0·504 | 0·429 | 1·538 | 0·230 | 0·636 | 240 | 109 | 304 |
| Ubon Ratchathani Province | Na Tan | 1·005 | 1·091 | 3·728 | 0·364 | 1·249 | 374 | 135 | 465 |
| Ubon Ratchathani Province | Na Yia | 0·611 | 0·559 | 1·945 | 0·268 | 0·763 | 148 | 67 | 186 |
| Ubon Ratchathani Province | Nam Khun | 0·483 | 0·394 | 1·391 | 0·229 | 0·608 | 148 | 69 | 186 |

| ADM1 | ADM2 | β-thalassaemia carrier prevalence Mean | SD | CI95% | Q25 | Q75 | Estimated carrier counts Mean | Q25 | Q75 |
|---|---|---|---|---|---|---|---|---|---|
| Ubon Ratchathani Province | Nam Yuen | 0·469 | 0·387 | 1·377 | 0·221 | 0·589 | 251 | 121 | 318 |
| Ubon Ratchathani Province | Phibun Mangsahan | 0·658 | 0·619 | 2·198 | 0·275 | 0·822 | 747 | 309 | 934 |
| Ubon Ratchathani Province | Pho Sai | 0·923 | 1·014 | 3·373 | 0·338 | 1·156 | 401 | 145 | 498 |
| Ubon Ratchathani Province | Samrong | 0·586 | 0·489 | 1·800 | 0·270 | 0·744 | 272 | 125 | 347 |
| Ubon Ratchathani Province | Sawang Wirawong | 0·684 | 0·652 | 2·374 | 0·281 | 0·852 | 135 | 55 | 168 |
| Ubon Ratchathani Province | Si Mueang Mai | 0·822 | 0·835 | 2·947 | 0·316 | 1·022 | 574 | 224 | 713 |
| Ubon Ratchathani Province | Sirindhorn | 0·654 | 0·624 | 2·199 | 0·270 | 0·817 | 326 | 135 | 408 |
| Ubon Ratchathani Province | Tan Sum | 0·733 | 0·695 | 2·480 | 0·304 | 0·917 | 232 | 96 | 287 |
| Ubon Ratchathani Province | Thung Si Udom | 0·524 | 0·439 | 1·617 | 0·243 | 0·659 | 101 | 47 | 126 |
| Ubon Ratchathani Province | Trakan Phuet Phon | 0·833 | 0·808 | 2·840 | 0·332 | 1·043 | 928 | 371 | 1165 |
| Ubon Ratchathani Province | Warin Chamrap | 0·635 | 0·557 | 2·002 | 0·283 | 0·805 | 1012 | 448 | 1285 |
| Udon Thani Province | Ban Dung | 2·139 | 2·329 | 7·910 | 0·785 | 2·636 | 1738 | 643 | 2130 |
| Udon Thani Province | Ban Phue | 2·227 | 2·263 | 7·794 | 0·874 | 2·731 | 1520 | 600 | 1868 |
| Udon Thani Province | Chai Wan | 1·815 | 1·759 | 6·266 | 0·758 | 2·245 | 511 | 211 | 628 |
| Udon Thani Province | Ku Kaeo | 1·813 | 1·673 | 6·140 | 0·793 | 2·219 | 319 | 138 | 389 |
| Udon Thani Province | Kumphawapi | 1·761 | 1·565 | 5·631 | 0·773 | 2·204 | 1316 | 575 | 1644 |
| Udon Thani Province | Kut Chap | 2·057 | 1·993 | 7·035 | 0·849 | 2·545 | 927 | 385 | 1156 |
| Udon Thani Province | Mueang Udon Thani | 1·992 | 1·941 | 6·917 | 0·819 | 2·471 | 6081 | 2492 | 7580 |
| Udon Thani Province | Na Yung | 2·389 | 2·404 | 8·648 | 0·916 | 2·979 | 516 | 197 | 647 |
| Udon Thani Province | Nam Som | 2·279 | 2·245 | 7·965 | 0·915 | 2·828 | 838 | 331 | 1042 |
| Udon Thani Province | Non Sa-At | 1·711 | 1·480 | 5·390 | 0·781 | 2·139 | 551 | 250 | 691 |
| Udon Thani Province | Nong Han | 1·954 | 1·958 | 6·794 | 0·787 | 2·414 | 1366 | 547 | 1688 |
| Udon Thani Province | Nong Saeng | 1·856 | 1·682 | 6·057 | 0·816 | 2·309 | 338 | 149 | 421 |
| Udon Thani Province | Nong Wua So | 1·906 | 1·763 | 6·201 | 0·828 | 2·380 | 788 | 341 | 979 |
| Udon Thani Province | Phen | 2·162 | 2·258 | 7·844 | 0·826 | 2·650 | 1836 | 701 | 2247 |
| Udon Thani Province | Phibun Rak | 2·026 | 1·973 | 7·069 | 0·823 | 2·529 | 175 | 71 | 219 |
| Udon Thani Province | Prachaksinlapakhom | 1·888 | 1·825 | 6·367 | 0·784 | 2·328 | 222 | 93 | 272 |
| Udon Thani Province | Sang Khom | 2·248 | 2·435 | 8·659 | 0·826 | 2·763 | 388 | 141 | 474 |
| Udon Thani Province | Si That | 1·714 | 1·595 | 5·788 | 0·731 | 2·142 | 684 | 295 | 859 |
| Udon Thani Province | Thung Fon | 2·008 | 2·092 | 7·305 | 0·786 | 2·488 | 454 | 176 | 565 |
| Udon Thani Province | Wang Sam Mo | 1·679 | 1·595 | 5·616 | 0·696 | 2·093 | 655 | 276 | 818 |
| Uthai Thani Province | Ban Rai | 2·515 | 2·351 | 8·512 | 1·027 | 3·167 | 1427 | 588 | 1798 |
| Uthai Thani Province | Huai Khot | 2·427 | 2·337 | 8·367 | 0·981 | 3·044 | 329 | 133 | 414 |
| Uthai Thani Province | Lan Sak | 2·571 | 2·385 | 8·558 | 1·064 | 3·252 | 1001 | 413 | 1261 |
| Uthai Thani Province | Mueang Uthai Thani | 2·169 | 1·926 | 6·943 | 0·956 | 2·730 | 1074 | 470 | 1363 |
| Uthai Thani Province | Nong Chang | 2·309 | 2·107 | 7·811 | 0·965 | 2·897 | 859 | 361 | 1081 |
| Uthai Thani Province | Nong Khayang | 2·178 | 1·964 | 7·299 | 0·927 | 2·770 | 85 | 37 | 108 |
| Uthai Thani Province | Sawang Arom | 2·388 | 2·215 | 7·793 | 1·019 | 2·994 | 665 | 282 | 833 |
| Uthai Thani Province | Thap Than | 2·295 | 2·093 | 7·538 | 0·984 | 2·870 | 816 | 350 | 1024 |
| Uttaradit Province | Ban Khok | 2·658 | 2·576 | 9·244 | 1·066 | 3·332 | 323 | 129 | 406 |
| Uttaradit Province | Fak Tha | 2·597 | 2·392 | 8·628 | 1·084 | 3·270 | 341 | 144 | 428 |
| Uttaradit Province | Laplae | 2·838 | 2·523 | 9·114 | 1·239 | 3·576 | 1874 | 825 | 2334 |
| Uttaradit Province | Mueang Uttaradit | 2·756 | 2·459 | 8·816 | 1·188 | 3·490 | 3164 | 1351 | 4004 |
| Uttaradit Province | Nam Pat | 2·476 | 2·158 | 7·724 | 1·104 | 3·117 | 690 | 308 | 870 |
| Uttaradit Province | Phichai | 2·554 | 2·093 | 7·584 | 1·201 | 3·223 | 1787 | 837 | 2244 |
| Uttaradit Province | Tha Pla | 2·689 | 2·441 | 8·801 | 1·149 | 3·405 | 882 | 382 | 1124 |
| Uttaradit Province | Thong Saen Khan | 2·500 | 2·087 | 7·580 | 1·156 | 3·163 | 642 | 293 | 815 |
| Uttaradit Province | Tron | 2·715 | 2·402 | 8·471 | 1·203 | 3·419 | 773 | 347 | 984 |
| Yala Province | Bannang Sata | 5·297 | 4·703 | 17·125 | 2·168 | 6·829 | 1963 | 803 | 2537 |
| Yala Province | Betong | 5·300 | 4·827 | 17·882 | 2·152 | 6·793 | 2160 | 880 | 2748 |
| Yala Province | Kabang | 5·386 | 4·557 | 16·793 | 2·338 | 6·971 | 1278 | 555 | 1653 |
| Yala Province | Krong Pinang | 5·198 | 4·573 | 16·405 | 2·247 | 6·731 | 1777 | 774 | 2312 |
| Yala Province | Mueang Yala | 5·195 | 4·443 | 16·392 | 2·228 | 6·612 | 6981 | 2936 | 8838 |
| Yala Province | Raman | 5·103 | 4·460 | 16·612 | 2·094 | 6·571 | 4585 | 1895 | 5889 |
| Yala Province | Than To | 5·348 | 4·852 | 17·809 | 2·148 | 6·839 | 858 | 347 | 1091 |
| Yala Province | Yaha | 5·268 | 4·437 | 16·286 | 2·316 | 6·730 | 3046 | 1352 | 3888 |
| Yasothon Province | Kham Khuean Kaeo | 0·885 | 0·807 | 2·872 | 0·375 | 1·106 | 392 | 166 | 492 |

| ADM1 | ADM2 | β-thalassaemia carrier prevalence Mean | SD | CI95% | Q25 | Q75 | Estimated carrier counts Mean | Q25 | Q75 |
|---|---|---|---|---|---|---|---|---|---|
| Yasothon Province | Kho Wang | 0·814 | 0·758 | 2·684 | 0·347 | 1·020 | 91 | 39 | 115 |
| Yasothon Province | Kut Chum | 1·052 | 0·993 | 3·588 | 0·430 | 1·318 | 496 | 198 | 620 |
| Yasothon Province | Loeng Nok Tha | 1·116 | 1·123 | 3·893 | 0·438 | 1·388 | 867 | 335 | 1075 |
| Yasothon Province | Maha Chana Chai | 0·858 | 0·767 | 2·722 | 0·373 | 1·086 | 326 | 140 | 415 |
| Yasothon Province | Mueang Yasothon | 0·988 | 0·874 | 3·167 | 0·425 | 1·258 | 1058 | 451 | 1352 |
| Yasothon Province | Pa Tio | 0·940 | 0·891 | 3·189 | 0·384 | 1·169 | 214 | 88 | 266 |
| Yasothon Province | Sai Mun | 1·043 | 0·959 | 3·498 | 0·430 | 1·317 | 209 | 85 | 263 |
| Yasothon Province | Thai Charoen | 1·064 | 1·006 | 3·712 | 0·434 | 1·327 | 222 | 91 | 276 |

ADM1: administrative level 1. ADM2: administrative level 2. SD: standard deviation. CI95%: 95% credible interval. Q25: 25th percentile. Q75: 75th percentile. SD and CI95% are measures of prediction uncertainty.

**Supplementary Table S37. Subnational areal predictions of β-thalassaemia carrier prevalence and estimated carrier count at administrative level 2 of Turkey.**

| ADM1 | ADM2 | β-thalassaemia carrier prevalence Mean | SD | CI95% | Q25 | Q75 | Estimated carrier counts Mean | Q25 | Q75 |
|---|---|---|---|---|---|---|---|---|---|
| Adana | Aladag | 2·466 | 1·999 | 7·313 | 1·161 | 3·112 | 408 | 190 | 520 |
| Adana | Ceyhan | 2·883 | 2·261 | 8·249 | 1·403 | 3·634 | 4601 | 2257 | 5755 |
| Adana | Cukurova | 3·232 | 2·596 | 9·433 | 1·538 | 4·093 | 18796 | 8999 | 23585 |
| Adana | Feke | 1·900 | 1·449 | 5·333 | 0·938 | 2·397 | 300 | 148 | 376 |
| Adana | Imamoglu | 2·682 | 2·134 | 7·816 | 1·275 | 3·384 | 752 | 367 | 945 |
| Adana | Karaisali | 2·902 | 2·318 | 8·516 | 1·374 | 3·686 | 656 | 312 | 835 |
| Adana | Karatas | 4·066 | 3·237 | 12·026 | 1·900 | 5·200 | 1037 | 475 | 1321 |
| Adana | Kozan | 2·253 | 1·708 | 6·258 | 1·130 | 2·838 | 3130 | 1540 | 3923 |
| Adana | Pozanti | 2·723 | 2·168 | 7·997 | 1·291 | 3·445 | 583 | 275 | 733 |
| Adana | Saimbeyli | 1·721 | 1·264 | 4·656 | 0·889 | 2·170 | 239 | 118 | 296 |
| Adana | Saricam | 3·064 | 2·437 | 8·938 | 1·466 | 3·887 | 7496 | 3547 | 9494 |
| Adana | Seyhan | 3·575 | 2·815 | 10·314 | 1·709 | 4·523 | 14991 | 7011 | 19148 |
| Adana | Tufanbeyli | 1·538 | 1·173 | 4·310 | 0·761 | 1·943 | 258 | 124 | 323 |
| Adana | Yumurtalik | 3·572 | 2·870 | 10·628 | 1·695 | 4·483 | 529 | 249 | 660 |
| Adana | Yuregir | 3·503 | 2·778 | 10·281 | 1·668 | 4·438 | 20606 | 9844 | 26425 |
| Adiyaman | Adiyaman merkez | 1·516 | 0·998 | 3·715 | 0·843 | 1·892 | 4965 | 2770 | 6140 |
| Adiyaman | Besni | 1·609 | 1·048 | 3·903 | 0·898 | 2·016 | 1235 | 690 | 1557 |
| Adiyaman | Celikhan | 1·337 | 0·929 | 3·409 | 0·725 | 1·675 | 194 | 108 | 241 |
| Adiyaman | Gerger | 1·412 | 1·017 | 3·684 | 0·748 | 1·771 | 224 | 121 | 282 |
| Adiyaman | Golbasi | 1·530 | 0·994 | 3·676 | 0·860 | 1·913 | 786 | 443 | 971 |
| Adiyaman | Kahta | 1·528 | 1·013 | 3·750 | 0·846 | 1·925 | 1907 | 1071 | 2408 |
| Adiyaman | Samsat | 1·732 | 1·150 | 4·318 | 0·964 | 2·155 | 126 | 71 | 158 |
| Adiyaman | Sincik | 1·348 | 0·938 | 3·422 | 0·725 | 1·692 | 226 | 121 | 286 |
| Adiyaman | Tut | 1·462 | 0·951 | 3·514 | 0·809 | 1·858 | 117 | 62 | 149 |
| Afyonkarahisar | Afyonkarahisar | 2·984 | 1·991 | 7·436 | 1·625 | 3·756 | 10345 | 5619 | 12820 |
| Afyonkarahisar | Basmakci | 3·659 | 2·372 | 8·996 | 2·028 | 4·627 | 358 | 198 | 454 |
| Afyonkarahisar | Bayat | 2·673 | 1·826 | 6·774 | 1·435 | 3·377 | 221 | 123 | 276 |
| Afyonkarahisar | Bolvadin | 2·805 | 1·873 | 7·054 | 1·523 | 3·551 | 1313 | 720 | 1681 |
| Afyonkarahisar | Cay | 3·065 | 2·053 | 7·646 | 1·659 | 3·880 | 971 | 517 | 1232 |
| Afyonkarahisar | Cobanlar | 2·930 | 1·901 | 7·196 | 1·602 | 3·747 | 415 | 225 | 534 |
| Afyonkarahisar | Dazkiri | 3·579 | 2·285 | 8·550 | 2·012 | 4·514 | 435 | 238 | 544 |
| Afyonkarahisar | Dinar | 3·436 | 2·276 | 8·477 | 1·884 | 4·341 | 1689 | 932 | 2148 |
| Afyonkarahisar | Emirdag | 2·574 | 1·816 | 6·714 | 1·349 | 3·258 | 1120 | 601 | 1426 |
| Afyonkarahisar | Evciler | 3·490 | 2·259 | 8·375 | 1·968 | 4·357 | 310 | 172 | 387 |
| Afyonkarahisar | Hocalar | 3·200 | 2·127 | 7·880 | 1·754 | 4·020 | 283 | 157 | 353 |
| Afyonkarahisar | Ihsaniye | 2·801 | 1·884 | 6·956 | 1·525 | 3·547 | 801 | 439 | 1018 |
| Afyonkarahisar | Iscehisar | 2·832 | 1·913 | 7·119 | 1·527 | 3·584 | 713 | 378 | 882 |
| Afyonkarahisar | Kiziloren | 3·393 | 2·276 | 8·313 | 1·825 | 4·321 | 61 | 32 | 77 |
| Afyonkarahisar | Sandikli | 3·245 | 2·131 | 7·920 | 1·789 | 4·091 | 1812 | 1000 | 2273 |
| Afyonkarahisar | Sinanpasa | 3·052 | 2·015 | 7·526 | 1·684 | 3·848 | 1111 | 610 | 1399 |
| Afyonkarahisar | Suhut | 3·194 | 2·088 | 7·859 | 1·753 | 4·035 | 1123 | 611 | 1427 |

| | | β-thalassaemia carrier prevalence | | | | | Estimated carrier counts | | |
|---|---|---|---|---|---|---|---|---|---|
| ADM1 | ADM2 | Mean | SD | CI95% | Q25 | Q75 | Mean | Q25 | Q75 |
| Afyonkarahisar | Sultandagi | 2·855 | 1·954 | 7·214 | 1·536 | 3·578 | 405 | 223 | 504 |
| Agri | Agri merkez | 1·343 | 1·211 | 4·329 | 0·581 | 1·696 | 2028 | 902 | 2579 |
| Agri | Diyadin | 1·683 | 1·588 | 5·644 | 0·694 | 2·115 | 662 | 266 | 850 |
| Agri | Dogubayazit | 1·757 | 1·703 | 6·068 | 0·697 | 2·216 | 2071 | 834 | 2602 |
| Agri | Eleskirt | 1·162 | 0·959 | 3·469 | 0·549 | 1·457 | 376 | 175 | 467 |
| Agri | Hamur | 1·447 | 1·266 | 4·580 | 0·635 | 1·837 | 220 | 99 | 282 |
| Agri | Patnos | 1·549 | 1·333 | 4·837 | 0·697 | 1·957 | 1769 | 807 | 2283 |
| Agri | Taslicay | 1·507 | 1·377 | 4·993 | 0·637 | 1·895 | 285 | 115 | 356 |
| Agri | Tutak | 1·273 | 1·034 | 3·787 | 0·605 | 1·604 | 372 | 175 | 467 |
| Aksaray | Agacoren | 1·968 | 1·774 | 6·401 | 0·838 | 2·484 | 141 | 59 | 176 |
| Aksaray | Aksaray merkez | 2·148 | 1·783 | 6·535 | 0·985 | 2·719 | 7119 | 3213 | 9001 |
| Aksaray | Eskil | 2·295 | 1·752 | 6·494 | 1·125 | 2·911 | 674 | 330 | 851 |
| Aksaray | Gulagac | 1·991 | 1·678 | 6·002 | 0·894 | 2·522 | 470 | 212 | 593 |
| Aksaray | Guzelyurt | 2·093 | 1·718 | 6·342 | 0·956 | 2·666 | 173 | 79 | 219 |
| Aksaray | Ortakoy | 1·928 | 1·723 | 6·184 | 0·809 | 2·463 | 627 | 265 | 796 |
| Aksaray | Sariyahsi | 1·942 | 1·797 | 6·436 | 0·799 | 2·457 | 102 | 42 | 130 |
| Aksaray | Sultanhani | 2·254 | 1·653 | 6·038 | 1·120 | 2·898 | 183 | 91 | 238 |
| Amasya | Amasya merkez | 1·419 | 1·775 | 5·780 | 0·440 | 1·723 | 2166 | 675 | 2656 |
| Amasya | Goynucek | 1·465 | 1·775 | 5·943 | 0·456 | 1·805 | 155 | 48 | 190 |
| Amasya | Gumushacikoy | 1·526 | 1·923 | 6·437 | 0·462 | 1·848 | 342 | 100 | 403 |
| Amasya | Hamamozu | 1·539 | 1·893 | 6·586 | 0·471 | 1·869 | 54 | 16 | 64 |
| Amasya | Merzifon | 1·507 | 1·889 | 6·341 | 0·447 | 1·827 | 1140 | 335 | 1394 |
| Amasya | Suluova | 1·488 | 1·915 | 6·219 | 0·437 | 1·818 | 799 | 228 | 997 |
| Amasya | Tasova | 1·397 | 1·814 | 5·786 | 0·424 | 1·673 | 447 | 132 | 533 |
| Ankara | Akyurt | 1·818 | 1·949 | 6·836 | 0·662 | 2·253 | 876 | 319 | 1080 |
| Ankara | Altindag | 1·863 | 1·943 | 6·719 | 0·703 | 2·329 | 8222 | 3070 | 10253 |
| Ankara | Ayas | 1·900 | 1·883 | 6·466 | 0·777 | 2·368 | 273 | 108 | 335 |
| Ankara | Bala | 1·943 | 1·899 | 6·605 | 0·782 | 2·426 | 404 | 157 | 504 |
| Ankara | Beypazari | 1·864 | 1·762 | 6·259 | 0·775 | 2·337 | 898 | 377 | 1118 |
| Ankara | Camlidere | 1·789 | 1·946 | 6·460 | 0·663 | 2·214 | 147 | 51 | 177 |
| Ankara | Cankaya | 1·920 | 1·951 | 6·788 | 0·747 | 2·398 | 15726 | 6141 | 19625 |
| Ankara | Cubuk | 1·831 | 2·054 | 6·938 | 0·645 | 2·248 | 1610 | 570 | 1922 |
| Ankara | Elmadag | 1·840 | 1·898 | 6·638 | 0·704 | 2·303 | 810 | 300 | 1003 |
| Ankara | Etimesgut | 1·913 | 1·924 | 6·436 | 0·772 | 2·372 | 11534 | 4686 | 14321 |
| Ankara | Evren | 1·935 | 1·798 | 6·395 | 0·794 | 2·431 | 60 | 25 | 74 |
| Ankara | Golbasi | 1·982 | 1·942 | 6·705 | 0·814 | 2·477 | 3122 | 1262 | 3904 |
| Ankara | Gudul | 1·838 | 1·821 | 6·286 | 0·735 | 2·291 | 158 | 62 | 196 |
| Ankara | Haymana | 2·087 | 1·806 | 6·490 | 0·943 | 2·615 | 589 | 266 | 736 |
| Ankara | Kahramankazan | 1·833 | 1·908 | 6·651 | 0·696 | 2·282 | 1268 | 487 | 1600 |
| Ankara | Kalecik | 1·786 | 1·953 | 6·793 | 0·616 | 2·222 | 227 | 77 | 286 |
| Ankara | Kecioren | 1·862 | 1·886 | 6·641 | 0·730 | 2·339 | 21458 | 8591 | 26885 |
| Ankara | Kizilcahamam | 1·805 | 1·990 | 6·674 | 0·662 | 2·225 | 481 | 174 | 586 |
| Ankara | Mamak | 1·864 | 1·964 | 6·616 | 0·718 | 2·311 | 12067 | 4661 | 15530 |
| Ankara | Nallihan | 1·880 | 1·656 | 5·969 | 0·826 | 2·367 | 502 | 206 | 636 |
| Ankara | Polatli | 2·117 | 1·793 | 6·464 | 0·969 | 2·675 | 2743 | 1250 | 3414 |
| Ankara | Pursaklar | 1·884 | 2·093 | 6·579 | 0·698 | 2·337 | 3451 | 1267 | 4288 |
| Ankara | Sereflikochisar | 2·000 | 1·864 | 6·608 | 0·845 | 2·505 | 672 | 280 | 818 |
| Ankara | Sincan | 1·931 | 1·871 | 6·451 | 0·810 | 2·405 | 12850 | 5194 | 16024 |
| Ankara | Yenimahalle | 1·883 | 1·968 | 6·559 | 0·735 | 2·340 | 12642 | 4948 | 15706 |
| Antalya | Akseki | 4·893 | 3·276 | 12·276 | 2·640 | 6·221 | 513 | 270 | 652 |
| Antalya | Aksu | 5·181 | 3·425 | 12·718 | 2·809 | 6·591 | 3905 | 2103 | 4962 |
| Antalya | Alanya | 6·392 | 4·231 | 15·875 | 3·462 | 8·109 | 18296 | 9948 | 22914 |
| Antalya | Demre | 5·984 | 4·241 | 16·008 | 3·081 | 7·649 | 1113 | 566 | 1419 |
| Antalya | Dogemealti | 4·818 | 3·182 | 11·881 | 2·637 | 6·079 | 3593 | 1946 | 4496 |
| Antalya | Elmali | 5·139 | 3·376 | 12·798 | 2·811 | 6·503 | 2171 | 1185 | 2735 |
| Antalya | Finike | 5·833 | 3·942 | 14·841 | 3·090 | 7·461 | 3130 | 1598 | 4006 |
| Antalya | Gazipasa | 7·502 | 4·958 | 18·432 | 4·056 | 9·576 | 3862 | 2091 | 4858 |

| | | β-thalassaemia carrier prevalence | | | | | Estimated carrier counts | | |
|---|---|---|---|---|---|---|---|---|---|
| ADM1 | ADM2 | Mean | SD | CI95% | Q25 | Q75 | Mean | Q25 | Q75 |
| Antalya | Gundogmus | 5·549 | 3·692 | 13·802 | 3·014 | 7·037 | 610 | 330 | 769 |
| Antalya | Ibradi | 4·821 | 3·231 | 12·099 | 2·627 | 6·089 | 157 | 86 | 199 |
| Antalya | Kas | 5·421 | 3·732 | 13·910 | 2·842 | 6·945 | 3043 | 1567 | 3899 |
| Antalya | Kemer | 5·902 | 3·966 | 15·051 | 3·177 | 7·484 | 2271 | 1226 | 2848 |
| Antalya | Kepez | 5·075 | 3·301 | 12·476 | 2·818 | 6·412 | 45224 | 25188 | 56960 |
| Antalya | Konyaalti | 5·289 | 3·410 | 12·841 | 2·926 | 6·685 | 5950 | 3184 | 7564 |
| Antalya | Korkuteli | 4·670 | 3·001 | 11·180 | 2·595 | 5·923 | 2663 | 1499 | 3370 |
| Antalya | Kumluca | 5·851 | 3·896 | 14·620 | 3·153 | 7·444 | 4115 | 2182 | 5322 |
| Antalya | Manavgat | 5·197 | 3·468 | 13·053 | 2·803 | 6·577 | 12655 | 6690 | 16226 |
| Antalya | Muratpasa | 5·388 | 3·542 | 13·414 | 2·925 | 6·895 | 15230 | 8372 | 19446 |
| Antalya | Serik | 5·035 | 3·327 | 12·402 | 2·756 | 6·381 | 7880 | 4321 | 10074 |
| Ardahan | Ardahan | 1·170 | 1·147 | 3·980 | 0·478 | 1·460 | 495 | 202 | 618 |
| Ardahan | Cildir | 1·319 | 1·444 | 4·829 | 0·498 | 1·612 | 113 | 41 | 138 |
| Ardahan | Damal | 1·271 | 1·426 | 4·552 | 0·473 | 1·557 | 63 | 23 | 77 |
| Ardahan | Gole | 1·128 | 1·071 | 3·742 | 0·481 | 1·409 | 260 | 118 | 333 |
| Ardahan | Hanak | 1·235 | 1·303 | 4·400 | 0·481 | 1·531 | 101 | 39 | 126 |
| Ardahan | Posof | 1·281 | 1·498 | 4·677 | 0·467 | 1·574 | 83 | 29 | 103 |
| Artvin | Ardanuc | 1·070 | 0·999 | 3·541 | 0·452 | 1·338 | 272 | 119 | 342 |
| Artvin | Arhavi | 1·016 | 0·955 | 3·368 | 0·430 | 1·265 | 140 | 54 | 177 |
| Artvin | Artvin Merkez | 1·022 | 0·940 | 3·343 | 0·439 | 1·277 | 228 | 94 | 285 |
| Artvin | Borcka | 1·096 | 1·067 | 3·730 | 0·441 | 1·365 | 234 | 95 | 291 |
| Artvin | Hopa | 1·051 | 0·987 | 3·652 | 0·433 | 1·305 | 44 | 18 | 54 |
| Artvin | Kemalpasa | 1·105 | 1·126 | 3·928 | 0·434 | 1·384 | 72 | 28 | 91 |
| Artvin | Murgul | 1·047 | 0·980 | 3·378 | 0·440 | 1·308 | 62 | 26 | 78 |
| Artvin | Savsat | 1·139 | 1·132 | 3·881 | 0·454 | 1·420 | 193 | 77 | 235 |
| Artvin | Yusufeli | 0·965 | 0·834 | 3·001 | 0·441 | 1·207 | 185 | 80 | 227 |
| Aydin | Bozdogan | 3·662 | 2·387 | 8·872 | 2·036 | 4·613 | 1179 | 657 | 1495 |
| Aydin | Buharkent | 3·575 | 2·300 | 8·791 | 2·000 | 4·519 | 463 | 260 | 588 |
| Aydin | Cine | 3·798 | 2·449 | 9·042 | 2·126 | 4·777 | 1873 | 1055 | 2336 |
| Aydin | Didim | 4·160 | 2·844 | 10·809 | 2·201 | 5·268 | 3218 | 1696 | 4083 |
| Aydin | Efeler | 3·873 | 2·464 | 9·241 | 2·180 | 4·872 | 12200 | 6905 | 15334 |
| Aydin | Germencik | 4·031 | 2·629 | 9·780 | 2·237 | 5·074 | 1802 | 992 | 2264 |
| Aydin | Incirliova | 3·974 | 2·594 | 9·609 | 2·246 | 4·980 | 2294 | 1279 | 2858 |
| Aydin | Karacasu | 3·629 | 2·310 | 8·547 | 2·056 | 4·563 | 670 | 383 | 837 |
| Aydin | Karpuzlu | 3·868 | 2·456 | 9·296 | 2·158 | 4·931 | 419 | 235 | 535 |
| Aydin | Kocarli | 3·993 | 2·648 | 9·867 | 2·185 | 5·051 | 927 | 501 | 1168 |
| Aydin | Kosk | 3·793 | 2·361 | 8·671 | 2·210 | 4·738 | 1047 | 609 | 1292 |
| Aydin | Kusadasi | 4·159 | 2·683 | 9·949 | 2·302 | 5·277 | 4657 | 2619 | 5894 |
| Aydin | Kuyucak | 3·612 | 2·297 | 8·662 | 2·027 | 4·548 | 970 | 547 | 1210 |
| Aydin | Nazilli | 3·683 | 2·319 | 8·696 | 2·102 | 4·620 | 6121 | 3545 | 7752 |
| Aydin | Soke | 4·057 | 2·689 | 9·991 | 2·231 | 5·102 | 5181 | 2852 | 6600 |
| Aydin | Sultanhisar | 3·752 | 2·274 | 8·659 | 2·180 | 4·741 | 818 | 479 | 1027 |
| Aydin | Yenipazar | 3·781 | 2·444 | 9·172 | 2·080 | 4·772 | 416 | 231 | 527 |
| Balikesir | Altieylul | 2·837 | 1·993 | 7·326 | 1·502 | 3·566 | 6467 | 3444 | 8032 |
| Balikesir | Ayvalik | 3·599 | 2·425 | 8·930 | 1·965 | 4·523 | 1580 | 877 | 1997 |
| Balikesir | Balya | 2·638 | 1·913 | 7·002 | 1·373 | 3·314 | 376 | 198 | 472 |
| Balikesir | Bandirma | 2·241 | 1·729 | 6·191 | 1·121 | 2·829 | 3568 | 1802 | 4525 |
| Balikesir | Bigadic | 2·846 | 1·989 | 7·361 | 1·513 | 3·580 | 1400 | 744 | 1768 |
| Balikesir | Burhaniye | 3·243 | 2·242 | 8·279 | 1·761 | 4·055 | 1911 | 1031 | 2360 |
| Balikesir | Dursunbey | 2·689 | 1·905 | 7·077 | 1·415 | 3·390 | 910 | 473 | 1140 |
| Balikesir | Edremit | 3·028 | 2·135 | 7·976 | 1·599 | 3·807 | 3372 | 1781 | 4227 |
| Balikesir | Erdek | 2·254 | 1·805 | 6·694 | 1·097 | 2·809 | 175 | 83 | 219 |
| Balikesir | Gomec | 3·452 | 2·448 | 9·084 | 1·829 | 4·358 | 416 | 217 | 532 |
| Balikesir | Gonen | 2·392 | 1·783 | 6·482 | 1·223 | 3·008 | 1828 | 937 | 2289 |
| Balikesir | Havran | 2·944 | 2·075 | 7·629 | 1·582 | 3·688 | 812 | 439 | 1011 |
| Balikesir | Ivrindi | 2·975 | 2·052 | 7·584 | 1·588 | 3·759 | 940 | 509 | 1183 |
| Balikesir | Karesi | 2·628 | 1·873 | 6·863 | 1·384 | 3·300 | 3810 | 2022 | 4822 |

| | | β-thalassaemia carrier prevalence | | | | | Estimated carrier counts | | |
|---|---|---|---|---|---|---|---|---|---|
| ADM1 | ADM2 | Mean | SD | CI95% | Q25 | Q75 | Mean | Q25 | Q75 |
| Balikesir | Kepsut | 2·649 | 1·956 | 7·252 | 1·356 | 3·333 | 596 | 305 | 749 |
| Balikesir | Manyas | 2·405 | 1·791 | 6·502 | 1·239 | 3·022 | 424 | 216 | 533 |
| Balikesir | Marmara | 2·290 | 1·809 | 6·445 | 1·131 | 2·891 | 71 | 35 | 90 |
| Balikesir | Savastepe | 3·026 | 2·071 | 7·754 | 1·634 | 3·808 | 530 | 286 | 666 |
| Balikesir | Sindirgi | 2·998 | 2·018 | 7·467 | 1·634 | 3·772 | 993 | 536 | 1257 |
| Balikesir | Susurluk | 2·454 | 1·810 | 6·633 | 1·261 | 3·084 | 956 | 490 | 1204 |
| Bartin | Amasra | 1·791 | 2·310 | 7·424 | 0·537 | 2·183 | 119 | 35 | 144 |
| Bartin | Bartin merkez | 1·792 | 2·322 | 7·265 | 0·555 | 2·182 | 2918 | 888 | 3572 |
| Bartin | Kurucasile | 1·818 | 2·473 | 7·760 | 0·515 | 2·198 | 67 | 18 | 82 |
| Bartin | Ulus | 1·789 | 2·283 | 7·284 | 0·553 | 2·177 | 393 | 124 | 477 |
| Batman | Batman Merkez | 2·186 | 1·839 | 6·652 | 1·016 | 2·739 | 10911 | 5027 | 13740 |
| Batman | Besiri | 2·283 | 1·930 | 6·951 | 1·049 | 2·890 | 772 | 350 | 975 |
| Batman | Gercus | 2·518 | 2·047 | 7·496 | 1·178 | 3·196 | 625 | 294 | 793 |
| Batman | Hasankeyf | 2·505 | 2·033 | 7·527 | 1·159 | 3·186 | 195 | 94 | 246 |
| Batman | Kozluk | 1·937 | 1·658 | 5·949 | 0·882 | 2·431 | 1192 | 537 | 1509 |
| Batman | Sason | 1·740 | 1·528 | 5·469 | 0·771 | 2·177 | 546 | 236 | 680 |
| Bayburt | Aydintepe | 0·978 | 0·933 | 3·283 | 0·398 | 1·217 | 88 | 35 | 111 |
| Bayburt | Bayburt merkez | 0·957 | 0·873 | 3·083 | 0·412 | 1·199 | 674 | 277 | 832 |
| Bayburt | Demirozu | 0·980 | 0·914 | 3·246 | 0·413 | 1·229 | 54 | 22 | 66 |
| Bilecik | Bilecik (merkez) | 1·955 | 1·494 | 5·530 | 0·971 | 2·460 | 1688 | 869 | 2103 |
| Bilecik | Bozuyuk | 2·228 | 1·623 | 5·889 | 1·149 | 2·811 | 1782 | 905 | 2276 |
| Bilecik | Golpazari | 1·832 | 1·409 | 5·180 | 0·906 | 2·299 | 184 | 92 | 229 |
| Bilecik | Inhisar | 1·979 | 1·559 | 5·604 | 0·962 | 2·474 | 47 | 22 | 58 |
| Bilecik | Osmaneli | 1·779 | 1·385 | 5·081 | 0·874 | 2·233 | 419 | 204 | 522 |
| Bilecik | Pazaryeri | 2·098 | 1·592 | 5·781 | 1·035 | 2·668 | 196 | 98 | 247 |
| Bilecik | Sogut | 2·036 | 1·559 | 5·752 | 1·010 | 2·566 | 417 | 210 | 539 |
| Bilecik | Yenipazar | 1·862 | 1·495 | 5·481 | 0·894 | 2·331 | 59 | 29 | 74 |
| Bingol | Adakli | 1·121 | 0·945 | 3·437 | 0·514 | 1·410 | 84 | 41 | 110 |
| Bingol | Bingol merkez | 1·242 | 1·077 | 3·921 | 0·556 | 1·559 | 2230 | 1001 | 2795 |
| Bingol | Genc | 1·379 | 1·218 | 4·370 | 0·613 | 1·733 | 455 | 197 | 580 |
| Bingol | Karliova | 1·144 | 0·980 | 3·540 | 0·524 | 1·431 | 318 | 139 | 397 |
| Bingol | Kigi | 1·091 | 0·976 | 3·509 | 0·481 | 1·360 | 62 | 25 | 73 |
| Bingol | Solhan | 1·298 | 1·102 | 3·964 | 0·599 | 1·628 | 443 | 204 | 562 |
| Bingol | Yayladere | 1·114 | 0·984 | 3·541 | 0·494 | 1·396 | 21 | 10 | 29 |
| Bingol | Yedisu | 1·051 | 0·902 | 3·227 | 0·485 | 1·313 | 28 | 10 | 36 |
| Bitlis | Adilcevaz | 1·736 | 1·490 | 5·350 | 0·785 | 2·179 | 527 | 238 | 670 |
| Bitlis | Ahlat | 1·630 | 1·386 | 4·998 | 0·743 | 2·049 | 377 | 168 | 476 |
| Bitlis | Bitlis merkez | 1·980 | 1·672 | 6·010 | 0·911 | 2·501 | 1283 | 585 | 1610 |
| Bitlis | Guroymak | 1·734 | 1·493 | 5·464 | 0·792 | 2·174 | 900 | 411 | 1122 |
| Bitlis | Hizan | 2·253 | 1·866 | 6·761 | 1·053 | 2·837 | 718 | 334 | 897 |
| Bitlis | Mutki | 1·800 | 1·550 | 5·609 | 0·820 | 2·255 | 549 | 247 | 681 |
| Bitlis | Tatvan | 1·956 | 1·669 | 6·017 | 0·898 | 2·457 | 1822 | 848 | 2324 |
| Bolu | Bolu Merkez | 1·727 | 1·786 | 5·882 | 0·668 | 2·169 | 3836 | 1482 | 4777 |
| Bolu | Dortdivan | 1·767 | 1·878 | 6·224 | 0·668 | 2·210 | 164 | 58 | 207 |
| Bolu | Gerede | 1·774 | 1·970 | 6·540 | 0·641 | 2·202 | 614 | 217 | 749 |
| Bolu | Goynuk | 1·798 | 1·505 | 5·493 | 0·828 | 2·264 | 259 | 118 | 318 |
| Bolu | Kibriscik | 1·764 | 1·747 | 6·086 | 0·715 | 2·223 | 50 | 20 | 65 |
| Bolu | Mengen | 1·725 | 1·931 | 6·307 | 0·624 | 2·164 | 240 | 83 | 295 |
| Bolu | Mudurnu | 1·742 | 1·600 | 5·676 | 0·744 | 2·191 | 356 | 152 | 451 |
| Bolu | Seben | 1·770 | 1·670 | 5·814 | 0·728 | 2·234 | 84 | 36 | 105 |
| Bolu | Yenicaga | 1·772 | 1·911 | 6·208 | 0·649 | 2·221 | 80 | 30 | 102 |
| Burdur | Aglasun | 4·072 | 2·736 | 10·245 | 2·180 | 5·169 | 288 | 153 | 365 |
| Burdur | Altinyayla | 4·390 | 2·826 | 10·496 | 2·429 | 5·583 | 237 | 131 | 305 |
| Burdur | Bucak | 4·397 | 2·920 | 10·929 | 2·401 | 5·571 | 2970 | 1616 | 3761 |
| Burdur | Burdur (merkez) | 3·964 | 2·661 | 9·925 | 2·147 | 5·021 | 4559 | 2476 | 5820 |
| Burdur | Cavdir | 4·408 | 2·890 | 10·677 | 2·439 | 5·547 | 566 | 317 | 713 |
| Burdur | Celtikci | 4·095 | 2·761 | 10·198 | 2·221 | 5·183 | 211 | 114 | 270 |

| | | β-thalassaemia carrier prevalence | | | | | Estimated carrier counts | | |
|---|---|---|---|---|---|---|---|---|---|
| ADM1 | ADM2 | Mean | SD | CI95% | Q25 | Q75 | Mean | Q25 | Q75 |
| Burdur | Golhisar | 4·191 | 2·702 | 10·197 | 2·326 | 5·282 | 1015 | 560 | 1290 |
| Burdur | Karamanli | 4·098 | 2·740 | 10·152 | 2·227 | 5·182 | 336 | 182 | 431 |
| Burdur | Kemer | 4·238 | 2·824 | 10·632 | 2·292 | 5·356 | 125 | 67 | 155 |
| Burdur | Tefenni | 4·167 | 2·718 | 10·096 | 2·296 | 5·282 | 475 | 259 | 598 |
| Burdur | Yesilova | 3·854 | 2·519 | 9·324 | 2·130 | 4·863 | 614 | 332 | 776 |
| Bursa | Buyukorhan | 2·506 | 1·829 | 6·786 | 1·284 | 3·156 | 240 | 117 | 301 |
| Bursa | Gemlik | 1·892 | 1·428 | 5·213 | 0·936 | 2·396 | 2043 | 1022 | 2621 |
| Bursa | Gursu | 1·983 | 1·482 | 5·441 | 0·992 | 2·532 | 2474 | 1274 | 3152 |
| Bursa | Harmancik | 2·483 | 1·780 | 6·539 | 1·294 | 3·137 | 144 | 77 | 181 |
| Bursa | Inegol | 2·132 | 1·613 | 5·883 | 1·069 | 2·685 | 6330 | 3149 | 7920 |
| Bursa | Iznik | 1·769 | 1·360 | 4·953 | 0·883 | 2·237 | 763 | 380 | 976 |
| Bursa | Karacabey | 2·201 | 1·712 | 6·297 | 1·086 | 2·771 | 1949 | 974 | 2480 |
| Bursa | Keles | 2·270 | 1·661 | 6·155 | 1·175 | 2·852 | 251 | 131 | 317 |
| Bursa | Kestel | 2·038 | 1·545 | 5·671 | 1·012 | 2·566 | 665 | 327 | 842 |
| Bursa | Mudanya | 2·052 | 1·568 | 5·658 | 1·019 | 2·591 | 1141 | 579 | 1433 |
| Bursa | Mustafakemalpasa | 2·357 | 1·753 | 6·416 | 1·204 | 2·955 | 2390 | 1219 | 3004 |
| Bursa | Nilufer | 2·146 | 1·603 | 5·883 | 1·085 | 2·719 | 15031 | 7540 | 19151 |
| Bursa | Orhaneli | 2·315 | 1·714 | 6·359 | 1·185 | 2·913 | 435 | 227 | 542 |
| Bursa | Orhangazi | 1·865 | 1·423 | 5·252 | 0·926 | 2·345 | 1479 | 744 | 1866 |
| Bursa | Osmangazi | 2·070 | 1·561 | 5·690 | 1·041 | 2·602 | 17817 | 8918 | 22524 |
| Bursa | Yenisehir | 1·950 | 1·492 | 5·536 | 0·962 | 2·467 | 1118 | 545 | 1409 |
| Bursa | Yildirim | 2·051 | 1·526 | 5·509 | 1·045 | 2·575 | 12905 | 6512 | 16169 |
| Canakkale | Ayvacik | 3·505 | 2·466 | 9·134 | 1·845 | 4·421 | 1161 | 613 | 1476 |
| Canakkale | Bayramic | 2·984 | 2·091 | 7·802 | 1·582 | 3·752 | 880 | 480 | 1113 |
| Canakkale | Biga | 2·450 | 1·789 | 6·553 | 1·266 | 3·070 | 1926 | 1007 | 2418 |
| Canakkale | Bozcaada | 3·596 | 2·607 | 9·885 | 1·860 | 4·452 | 50 | 26 | 62 |
| Canakkale | Can | 2·642 | 1·912 | 7·035 | 1·387 | 3·307 | 1291 | 692 | 1599 |
| Canakkale | Canakkale merkez | 2·929 | 2·054 | 7·669 | 1·549 | 3·701 | 5938 | 3082 | 7406 |
| Canakkale | Eceabat | 3·120 | 2·235 | 8·388 | 1·609 | 3·964 | 152 | 77 | 197 |
| Canakkale | Ezine | 3·299 | 2·318 | 8·597 | 1·736 | 4·172 | 1021 | 537 | 1276 |
| Canakkale | Gelibolu | 2·828 | 2·089 | 7·720 | 1·430 | 3·590 | 507 | 255 | 645 |
| Canakkale | Imbros | 3·794 | 2·774 | 10·513 | 1·915 | 4·794 | 368 | 184 | 459 |
| Canakkale | Lapseki | 2·675 | 1·945 | 7·255 | 1·385 | 3·346 | 588 | 306 | 738 |
| Canakkale | Yenice | 2·653 | 1·902 | 6·948 | 1·399 | 3·324 | 831 | 434 | 1036 |
| Cankiri | Atkaracalar | 1·792 | 2·165 | 7·063 | 0·581 | 2·195 | 91 | 30 | 111 |
| Cankiri | Bayramoren | 1·788 | 2·302 | 7·193 | 0·567 | 2·174 | 25 | 8 | 29 |
| Cankiri | Cankiri merkez | 1·712 | 1·938 | 6·667 | 0·556 | 2·140 | 1812 | 581 | 2253 |
| Cankiri | Cerkes | 1·790 | 2·077 | 6·790 | 0·607 | 2·232 | 294 | 100 | 363 |
| Cankiri | Eldivan | 1·784 | 2·051 | 6·862 | 0·571 | 2·244 | 115 | 36 | 148 |
| Cankiri | Ilgaz | 1·740 | 2·077 | 7·106 | 0·539 | 2·161 | 244 | 77 | 300 |
| Cankiri | Kizilirmak | 1·687 | 1·870 | 6·569 | 0·546 | 2·118 | 125 | 40 | 162 |
| Cankiri | Korgun | 1·793 | 2·131 | 7·272 | 0·559 | 2·208 | 79 | 23 | 98 |
| Cankiri | Kursunlu | 1·767 | 2·103 | 7·107 | 0·565 | 2·187 | 161 | 52 | 200 |
| Cankiri | Orta | 1·798 | 2·069 | 6·924 | 0·609 | 2·226 | 249 | 84 | 312 |
| Cankiri | Sabanozu | 1·788 | 2·048 | 6·877 | 0·601 | 2·232 | 221 | 74 | 274 |
| Cankiri | Yaprakli | 1·706 | 2·003 | 6·938 | 0·531 | 2·119 | 141 | 41 | 170 |
| Corum | Alaca | 1·557 | 1·797 | 6·205 | 0·504 | 1·930 | 470 | 154 | 581 |
| Corum | Bayat | 1·662 | 1·969 | 6·907 | 0·513 | 2·041 | 231 | 71 | 286 |
| Corum | Bogazkale | 1·622 | 1·770 | 6·171 | 0·538 | 2·054 | 62 | 19 | 79 |
| Corum | Corum merkez | 1·562 | 1·876 | 6·335 | 0·484 | 1·914 | 4787 | 1483 | 5959 |
| Corum | Dodurga | 1·595 | 1·979 | 6·699 | 0·476 | 1·949 | 83 | 24 | 100 |
| Corum | Iskilip | 1·650 | 2·002 | 6·825 | 0·495 | 2·033 | 496 | 154 | 618 |
| Corum | Kargi | 1·684 | 2·134 | 7·391 | 0·483 | 2·045 | 275 | 75 | 326 |
| Corum | Lacin | 1·564 | 1·864 | 6·380 | 0·475 | 1·952 | 57 | 16 | 70 |
| Corum | Mecitozu | 1·497 | 1·864 | 6·090 | 0·463 | 1·830 | 214 | 67 | 263 |
| Corum | Oguzlar | 1·594 | 1·908 | 6·633 | 0·488 | 1·950 | 76 | 24 | 95 |
| Corum | Ortakoy | 1·494 | 1·814 | 6·068 | 0·470 | 1·838 | 101 | 31 | 124 |

| | | β-thalassaemia carrier prevalence | | | | | Estimated carrier counts | | |
|---|---|---|---|---|---|---|---|---|---|
| ADM1 | ADM2 | Mean | SD | CI95% | Q25 | Q75 | Mean | Q25 | Q75 |
| Corum | Osmancik | 1·611 | 2·041 | 6·815 | 0·471 | 1·956 | 705 | 203 | 856 |
| Corum | Sungurlu | 1·657 | 1·872 | 6·480 | 0·537 | 2·077 | 821 | 259 | 1018 |
| Corum | Ugurludag | 1·647 | 1·941 | 6·950 | 0·502 | 2·020 | 112 | 32 | 139 |
| Denizli | Acipayam | 3·864 | 2·497 | 9·307 | 2·152 | 4·882 | 2248 | 1243 | 2866 |
| Denizli | Babadag | 3·588 | 2·296 | 8·403 | 2·037 | 4·541 | 210 | 120 | 263 |
| Denizli | Baklan | 3·481 | 2·251 | 8·492 | 1·948 | 4·395 | 257 | 146 | 325 |
| Denizli | Bekilli | 3·311 | 2·115 | 7·855 | 1·867 | 4·181 | 206 | 114 | 260 |
| Denizli | Beyagac | 3·791 | 2·500 | 9·235 | 2·103 | 4·758 | 225 | 126 | 285 |
| Denizli | Bozkurt | 3·597 | 2·281 | 8·597 | 2·017 | 4·530 | 406 | 224 | 511 |
| Denizli | Buldan | 3·523 | 2·253 | 8·428 | 1·995 | 4·407 | 980 | 558 | 1222 |
| Denizli | Cal | 3·437 | 2·213 | 8·245 | 1·915 | 4·342 | 647 | 365 | 811 |
| Denizli | Cameli | 4·173 | 2·707 | 10·121 | 2·312 | 5·258 | 698 | 387 | 872 |
| Denizli | Cardak | 3·680 | 2·357 | 8·738 | 2·065 | 4·629 | 372 | 208 | 472 |
| Denizli | Civril | 3·352 | 2·164 | 8·058 | 1·872 | 4·221 | 1951 | 1083 | 2478 |
| Denizli | Guney | 3·419 | 2·141 | 7·962 | 1·935 | 4·317 | 329 | 187 | 418 |
| Denizli | Honaz | 3·520 | 2·233 | 8·388 | 1·993 | 4·417 | 1157 | 652 | 1450 |
| Denizli | Kale | 3·665 | 2·367 | 8·862 | 2·042 | 4·624 | 732 | 405 | 915 |
| Denizli | Merkezefendi | 3·518 | 2·171 | 8·125 | 2·017 | 4·430 | 11385 | 6602 | 14252 |
| Denizli | Pamukkale | 3·484 | 2·214 | 8·338 | 1·965 | 4·370 | 13646 | 7694 | 17056 |
| Denizli | Saraykoy | 3·558 | 2·265 | 8·425 | 2·012 | 4·474 | 1154 | 645 | 1443 |
| Denizli | Serinhisar | 3·611 | 2·339 | 8·774 | 1·992 | 4·533 | 451 | 249 | 574 |
| Denizli | Tavas | 3·658 | 2·378 | 8·887 | 2·036 | 4·624 | 1543 | 858 | 1959 |
| Diyarbakir | Baglar | 1·841 | 1·412 | 5·171 | 0·914 | 2·318 | 14096 | 6839 | 17777 |
| Diyarbakir | Bismil | 2·073 | 1·709 | 6·228 | 0·970 | 2·607 | 2565 | 1178 | 3239 |
| Diyarbakir | Cermik | 1·483 | 1·123 | 4·093 | 0·751 | 1·865 | 752 | 378 | 955 |
| Diyarbakir | Cinar | 2·113 | 1·650 | 6·003 | 1·029 | 2·664 | 1883 | 896 | 2348 |
| Diyarbakir | Cungu | 1·323 | 0·999 | 3·712 | 0·673 | 1·665 | 137 | 71 | 174 |
| Diyarbakir | Dicle | 1·470 | 1·251 | 4·413 | 0·677 | 1·847 | 575 | 263 | 724 |
| Diyarbakir | Egil | 1·575 | 1·300 | 4·695 | 0·727 | 1·995 | 358 | 168 | 454 |
| Diyarbakir | Ergani | 1·541 | 1·198 | 4·396 | 0·754 | 1·947 | 2035 | 988 | 2594 |
| Diyarbakir | Hani | 1·493 | 1·298 | 4·555 | 0·679 | 1·880 | 508 | 233 | 644 |
| Diyarbakir | Hazro | 1·755 | 1·582 | 5·555 | 0·772 | 2·209 | 270 | 118 | 340 |
| Diyarbakir | Kayapinar | 1·728 | 1·376 | 4·898 | 0·836 | 2·168 | 2127 | 1032 | 2676 |
| Diyarbakir | Kocakoy | 1·638 | 1·416 | 4·897 | 0·733 | 2·086 | 274 | 124 | 352 |
| Diyarbakir | Kulp | 1·593 | 1·391 | 4·973 | 0·712 | 1·995 | 566 | 250 | 709 |
| Diyarbakir | Lice | 1·540 | 1·366 | 4·764 | 0·694 | 1·937 | 414 | 180 | 520 |
| Diyarbakir | Silvan | 1·859 | 1·595 | 5·764 | 0·845 | 2·342 | 1656 | 771 | 2098 |
| Diyarbakir | Sur | 1·799 | 1·497 | 5·395 | 0·841 | 2·264 | 2716 | 1308 | 3425 |
| Diyarbakir | Yenisehir | 1·701 | 1·350 | 4·886 | 0·812 | 2·156 | 2959 | 1418 | 3808 |
| Duzce | Akcakoca | 1·596 | 1·573 | 5·284 | 0·658 | 2·007 | 162 | 67 | 204 |
| Duzce | Cilimli | 1·579 | 1·496 | 4·834 | 0·670 | 1·991 | 478 | 200 | 604 |
| Duzce | Cumayeri | 1·581 | 1·516 | 5·002 | 0·671 | 1·990 | 147 | 62 | 185 |
| Duzce | Duzce merkez | 1·645 | 1·562 | 5·372 | 0·684 | 2·070 | 4192 | 1738 | 5144 |
| Duzce | Golyaka | 1·667 | 1·500 | 5·233 | 0·729 | 2·090 | 369 | 159 | 466 |
| Duzce | Gumusova | 1·610 | 1·468 | 4·926 | 0·710 | 2·016 | 416 | 179 | 520 |
| Duzce | Kaynasli | 1·697 | 1·707 | 5·701 | 0·687 | 2·139 | 295 | 120 | 374 |
| Duzce | Yigilca | 1·657 | 1·667 | 5·754 | 0·650 | 2·075 | 244 | 97 | 308 |
| Edirne | Edirne merkez | 3·957 | 3·504 | 12·860 | 1·675 | 5·038 | 7504 | 3125 | 9623 |
| Edirne | Enez | 3·510 | 2·671 | 10·080 | 1·713 | 4·464 | 207 | 99 | 265 |
| Edirne | Havsa | 3·481 | 3·116 | 11·271 | 1·485 | 4·425 | 671 | 287 | 853 |
| Edirne | Ipsala | 3·465 | 2·679 | 9·857 | 1·674 | 4·398 | 914 | 442 | 1178 |
| Edirne | Kesan | 3·073 | 2·376 | 8·756 | 1·502 | 3·897 | 2638 | 1290 | 3279 |
| Edirne | Lalapasa | 3·843 | 3·520 | 12·840 | 1·565 | 4·895 | 262 | 106 | 334 |
| Edirne | Meric | 3·664 | 3·008 | 10·896 | 1·707 | 4·634 | 447 | 208 | 559 |
| Edirne | Suloglu | 3·542 | 3·199 | 11·778 | 1·470 | 4·500 | 273 | 113 | 352 |
| Edirne | Uzunkopru | 3·319 | 2·793 | 10·147 | 1·507 | 4·204 | 2037 | 934 | 2585 |
| Elazig | Agin | 1·100 | 0·917 | 3·396 | 0·508 | 1·384 | 25 | 13 | 33 |

| | | β-thalassaemia carrier prevalence | | | | | Estimated carrier counts | | |
|---|---|---|---|---|---|---|---|---|---|
| ADM1 | ADM2 | Mean | SD | CI95% | Q25 | Q75 | Mean | Q25 | Q75 |
| Elazig | Alacakaya | 1·369 | 1·163 | 4·190 | 0·625 | 1·732 | 90 | 39 | 112 |
| Elazig | Aricak | 1·362 | 1·156 | 4·190 | 0·627 | 1·711 | 142 | 65 | 180 |
| Elazig | Baskil | 1·168 | 0·895 | 3·263 | 0·577 | 1·477 | 126 | 59 | 164 |
| Elazig | Elazig merkez | 1·184 | 0·983 | 3·528 | 0·554 | 1·489 | 5458 | 2485 | 6797 |
| Elazig | Karakocan | 1·173 | 1·035 | 3·710 | 0·523 | 1·471 | 342 | 154 | 434 |
| Elazig | Keban | 1·138 | 0·927 | 3·351 | 0·533 | 1·441 | 70 | 32 | 88 |
| Elazig | Kovancilar | 1·226 | 1·044 | 3·781 | 0·560 | 1·535 | 446 | 200 | 544 |
| Elazig | Maden | 1·322 | 1·070 | 3·875 | 0·635 | 1·661 | 121 | 53 | 147 |
| Elazig | Palu | 1·305 | 1·126 | 4·060 | 0·591 | 1·636 | 286 | 130 | 362 |
| Elazig | Sivrice | 1·256 | 0·981 | 3·534 | 0·619 | 1·583 | 117 | 55 | 143 |
| Erzincan | Cayirli | 0·983 | 0·883 | 3·181 | 0·430 | 1·228 | 83 | 33 | 100 |
| Erzincan | Erzincan merkez | 1·025 | 0·957 | 3·442 | 0·427 | 1·287 | 1749 | 722 | 2205 |
| Erzincan | Ilic | 1·066 | 1·039 | 3·651 | 0·431 | 1·325 | 89 | 32 | 112 |
| Erzincan | Kemah | 1·037 | 0·997 | 3·548 | 0·422 | 1·298 | 72 | 22 | 92 |
| Erzincan | Kemaliye | 1·068 | 0·957 | 3·446 | 0·461 | 1·340 | 50 | 17 | 59 |
| Erzincan | Otlukbeli | 0·972 | 0·855 | 3·133 | 0·426 | 1·220 | 22 | 9 | 29 |
| Erzincan | Refahiye | 1·045 | 1·054 | 3·692 | 0·410 | 1·300 | 113 | 38 | 140 |
| Erzincan | Tercan | 0·988 | 0·851 | 3·067 | 0·449 | 1·239 | 153 | 64 | 191 |
| Erzincan | Uzumlu | 1·004 | 0·887 | 3·283 | 0·437 | 1·256 | 128 | 54 | 163 |
| Erzurum | Askale | 0·950 | 0·786 | 2·820 | 0·450 | 1·190 | 216 | 98 | 279 |
| Erzurum | Aziziye | 0·923 | 0·724 | 2·630 | 0·450 | 1·162 | 654 | 323 | 836 |
| Erzurum | Cat | 1·029 | 0·839 | 3·034 | 0·494 | 1·289 | 163 | 79 | 205 |
| Erzurum | Hinis | 1·140 | 0·904 | 3·269 | 0·558 | 1·430 | 278 | 134 | 351 |
| Erzurum | Horasan | 1·020 | 0·810 | 2·920 | 0·500 | 1·278 | 380 | 180 | 473 |
| Erzurum | Ispir | 0·927 | 0·781 | 2·801 | 0·430 | 1·166 | 133 | 62 | 169 |
| Erzurum | Karacoban | 1·185 | 0·929 | 3·381 | 0·575 | 1·492 | 272 | 132 | 341 |
| Erzurum | Karayazi | 1·086 | 0·851 | 3·076 | 0·532 | 1·370 | 300 | 149 | 384 |
| Erzurum | Koprukoy | 0·994 | 0·758 | 2·752 | 0·497 | 1·259 | 142 | 68 | 178 |
| Erzurum | Narman | 0·949 | 0·747 | 2·673 | 0·469 | 1·193 | 122 | 59 | 153 |
| Erzurum | Oltu | 0·962 | 0·780 | 2·784 | 0·462 | 1·209 | 294 | 138 | 372 |
| Erzurum | Olur | 1·009 | 0·864 | 3·085 | 0·462 | 1·269 | 62 | 26 | 77 |
| Erzurum | Palandoken | 0·970 | 0·763 | 2·748 | 0·484 | 1·209 | 1215 | 605 | 1513 |
| Erzurum | Pasinler | 0·953 | 0·727 | 2·626 | 0·479 | 1·200 | 262 | 135 | 332 |
| Erzurum | Pazaryolu | 0·928 | 0·800 | 2·874 | 0·421 | 1·162 | 37 | 16 | 50 |
| Erzurum | Senkaya | 1·033 | 0·868 | 3·150 | 0·481 | 1·298 | 174 | 76 | 224 |
| Erzurum | Tekman | 1·036 | 0·816 | 2·991 | 0·509 | 1·296 | 235 | 112 | 293 |
| Erzurum | Tortum | 0·920 | 0·723 | 2·591 | 0·453 | 1·155 | 144 | 70 | 181 |
| Erzurum | Uzundere | 0·936 | 0·777 | 2·729 | 0·450 | 1·172 | 59 | 29 | 75 |
| Erzurum | Yakutiye | 0·922 | 0·711 | 2·568 | 0·464 | 1·157 | 2339 | 1188 | 2922 |
| Eskisehir | Alpu | 2·088 | 1·670 | 6·084 | 1·009 | 2·626 | 218 | 104 | 272 |
| Eskisehir | Beylikova | 2·142 | 1·714 | 6·287 | 1·030 | 2·705 | 118 | 60 | 146 |
| Eskisehir | Cifteler | 2·458 | 1·872 | 6·858 | 1·224 | 3·101 | 384 | 186 | 493 |
| Eskisehir | Gunyuzu | 2·246 | 1·804 | 6·501 | 1·066 | 2·844 | 121 | 58 | 155 |
| Eskisehir | Han | 2·628 | 1·832 | 6·740 | 1·371 | 3·316 | 53 | 26 | 68 |
| Eskisehir | Inonu | 2·279 | 1·657 | 6·138 | 1·168 | 2·872 | 162 | 81 | 206 |
| Eskisehir | Mahmudiye | 2·348 | 1·848 | 6·782 | 1·134 | 2·973 | 188 | 89 | 240 |
| Eskisehir | Mihalgazi | 1·977 | 1·534 | 5·698 | 0·968 | 2·483 | 92 | 44 | 115 |
| Eskisehir | Mihaliccik | 2·012 | 1·710 | 6·144 | 0·922 | 2·533 | 160 | 68 | 196 |
| Eskisehir | Odunpazari | 2·274 | 1·687 | 6·266 | 1·160 | 2·866 | 9359 | 4666 | 11884 |
| Eskisehir | Saricakaya | 1·958 | 1·598 | 5·796 | 0·933 | 2·441 | 69 | 34 | 84 |
| Eskisehir | Seyitgazi | 2·532 | 1·820 | 6·738 | 1·308 | 3·211 | 316 | 161 | 398 |
| Eskisehir | Sivrihisar | 2·269 | 1·807 | 6·550 | 1·098 | 2·858 | 457 | 222 | 583 |
| Eskisehir | Tepebasi | 2·124 | 1·615 | 6·014 | 1·050 | 2·677 | 8784 | 4387 | 11067 |
| Gaziantep | Araban | 1·778 | 1·166 | 4·312 | 0·991 | 2·237 | 645 | 358 | 804 |
| Gaziantep | Islahiye | 2·488 | 1·790 | 6·552 | 1·291 | 3·151 | 1765 | 924 | 2244 |
| Gaziantep | Karkamis | 2·663 | 1·859 | 6·951 | 1·408 | 3·335 | 272 | 143 | 344 |
| Gaziantep | Nizip | 2·314 | 1·533 | 5·672 | 1·278 | 2·906 | 4006 | 2201 | 5082 |

| | | β-thalassaemia carrier prevalence | | | | | Estimated carrier counts | | |
|---|---|---|---|---|---|---|---|---|---|
| ADM1 | ADM2 | Mean | SD | CI95% | Q25 | Q75 | Mean | Q25 | Q75 |
| Gaziantep | Nurdagi | 2·197 | 1·535 | 5·673 | 1·168 | 2·770 | 969 | 513 | 1219 |
| Gaziantep | Oguzeli | 2·532 | 1·736 | 6·411 | 1·362 | 3·193 | 906 | 491 | 1149 |
| Gaziantep | Sahinbey | 2·368 | 1·635 | 6·025 | 1·268 | 2·979 | 19310 | 10236 | 24124 |
| Gaziantep | Sehitkamil | 2·087 | 1·397 | 5·219 | 1·142 | 2·625 | 22017 | 12014 | 28061 |
| Gaziantep | Yavuzeli | 1·897 | 1·258 | 4·610 | 1·063 | 2·373 | 391 | 220 | 485 |
| Giresun | Alucra | 1·080 | 1·200 | 4·007 | 0·384 | 1·339 | 96 | 32 | 123 |
| Giresun | Bulancak | 1·226 | 1·576 | 4·955 | 0·383 | 1·489 | 896 | 276 | 1094 |
| Giresun | Camoluk | 1·056 | 1·105 | 3·780 | 0·394 | 1·309 | 67 | 24 | 86 |
| Giresun | Canakci | 1·135 | 1·307 | 4·307 | 0·390 | 1·393 | 89 | 30 | 108 |
| Giresun | Dereli | 1·164 | 1·415 | 4·626 | 0·388 | 1·413 | 226 | 70 | 268 |
| Giresun | Dogankent | 1·118 | 1·340 | 4·160 | 0·388 | 1·381 | 60 | 21 | 76 |
| Giresun | Espiye | 1·145 | 1·352 | 4·589 | 0·385 | 1·390 | 377 | 122 | 456 |
| Giresun | Eynesil | 1·177 | 1·336 | 4·786 | 0·392 | 1·444 | 77 | 25 | 93 |
| Giresun | Giresun District | 1·211 | 1·484 | 5·032 | 0·383 | 1·466 | 461 | 146 | 559 |
| Giresun | Gorele | 1·154 | 1·396 | 4·683 | 0·377 | 1·400 | 328 | 105 | 391 |
| Giresun | Guce | 1·128 | 1·293 | 4·404 | 0·387 | 1·374 | 96 | 32 | 115 |
| Giresun | Kesap | 1·184 | 1·425 | 4·727 | 0·384 | 1·454 | 94 | 30 | 115 |
| Giresun | Piraziz | 1·257 | 1·681 | 5·412 | 0·386 | 1·494 | 82 | 25 | 97 |
| Giresun | Sebinkarahisar | 1·122 | 1·308 | 4·258 | 0·384 | 1·379 | 213 | 70 | 257 |
| Giresun | Tirebolu | 1·155 | 1·364 | 4·592 | 0·385 | 1·402 | 322 | 106 | 396 |
| Giresun | Yaglidere | 1·145 | 1·307 | 4·561 | 0·391 | 1·396 | 159 | 55 | 198 |
| Gumushane | Gumushane merkez | 1·010 | 1·009 | 3·527 | 0·391 | 1·268 | 579 | 205 | 735 |
| Gumushane | Kelkit | 1·019 | 1·013 | 3·535 | 0·401 | 1·281 | 422 | 160 | 536 |
| Gumushane | Kose | 0·999 | 0·980 | 3·489 | 0·403 | 1·242 | 114 | 46 | 138 |
| Gumushane | Kurtun | 1·088 | 1·189 | 4·054 | 0·386 | 1·347 | 131 | 44 | 164 |
| Gumushane | Siran | 1·038 | 1·085 | 3·800 | 0·390 | 1·287 | 202 | 73 | 250 |
| Gumushane | Torul | 1·053 | 1·115 | 3·838 | 0·384 | 1·313 | 118 | 42 | 145 |
| Hakkari | Cukurca | 3·748 | 2·573 | 9·539 | 1·997 | 4·746 | 486 | 259 | 620 |
| Hakkari | Derecik | 4·490 | 2·983 | 11·253 | 2·430 | 5·671 | 1034 | 561 | 1302 |
| Hakkari | Hakkari merkez | 3·401 | 2·488 | 9·266 | 1·735 | 4·305 | 2794 | 1386 | 3551 |
| Hakkari | Semdinli | 4·104 | 2·898 | 10·748 | 2·127 | 5·214 | 1811 | 928 | 2326 |
| Hakkari | Yuksekova | 3·637 | 2·680 | 9·962 | 1·829 | 4·626 | 4418 | 2220 | 5557 |
| Hatay | Altinozu | 4·538 | 3·838 | 13·880 | 2·043 | 5·755 | 3080 | 1390 | 3894 |
| Hatay | Antakya | 4·150 | 3·395 | 12·580 | 1·918 | 5·272 | 20761 | 9482 | 26110 |
| Hatay | Arsuz | 4·100 | 3·281 | 12·153 | 1·930 | 5·196 | 1611 | 751 | 2040 |
| Hatay | Belen | 3·819 | 3·036 | 11·329 | 1·849 | 4·801 | 1283 | 628 | 1628 |
| Hatay | Defne | 4·571 | 3·805 | 13·863 | 2·074 | 5·882 | 4495 | 2049 | 5824 |
| Hatay | Dortyol | 2·921 | 2·197 | 8·118 | 1·469 | 3·694 | 4197 | 2092 | 5391 |
| Hatay | Erzin | 2·867 | 2·125 | 7·886 | 1·437 | 3·653 | 1163 | 599 | 1498 |
| Hatay | Hassa | 2·990 | 2·333 | 8·513 | 1·470 | 3·756 | 1730 | 847 | 2195 |
| Hatay | Iskenderun | 3·390 | 2·638 | 9·640 | 1·661 | 4·295 | 8581 | 4263 | 10900 |
| Hatay | Kirikhan | 3·486 | 2·717 | 10·047 | 1·676 | 4·453 | 4669 | 2204 | 5951 |
| Hatay | Kumlu | 3·677 | 2·937 | 11·010 | 1·710 | 4·680 | 407 | 185 | 508 |
| Hatay | Payas | 3·141 | 2·374 | 8·777 | 1·540 | 3·972 | 1425 | 696 | 1821 |
| Hatay | Reyhanli | 3·939 | 3·225 | 12·031 | 1·797 | 4·981 | 4468 | 2095 | 5676 |
| Hatay | Samandag | 4·600 | 3·799 | 13·891 | 2·066 | 5·881 | 5362 | 2365 | 6837 |
| Hatay | Yayladagi | 4·903 | 4·023 | 14·789 | 2·182 | 6·298 | 2113 | 933 | 2705 |
| Igdir | Aralik | 1·843 | 1·940 | 6·755 | 0·680 | 2·295 | 419 | 152 | 517 |
| Igdir | Igdir (merkez) | 1·666 | 1·682 | 5·853 | 0·643 | 2·089 | 2385 | 929 | 2971 |
| Igdir | Karakoyunlu | 1·736 | 1·801 | 6·212 | 0·645 | 2·175 | 152 | 57 | 189 |
| Igdir | Tuzluca | 1·459 | 1·414 | 5·006 | 0·591 | 1·830 | 351 | 139 | 439 |
| Isparta | Aksu | 3·829 | 2·585 | 9·728 | 2·045 | 4·862 | 178 | 94 | 226 |
| Isparta | Atabey | 3·622 | 2·494 | 9·180 | 1·938 | 4·590 | 222 | 117 | 280 |
| Isparta | Egirdir | 3·752 | 2·535 | 9·505 | 2·014 | 4·773 | 978 | 518 | 1245 |
| Isparta | Gelendost | 3·401 | 2·373 | 8·841 | 1·801 | 4·309 | 535 | 286 | 679 |
| Isparta | Gonen | 3·655 | 2·431 | 9·061 | 1·986 | 4·629 | 271 | 146 | 344 |
| Isparta | Isparta merkez | 3·879 | 2·684 | 10·000 | 2·056 | 4·890 | 10586 | 5536 | 13487 |

| | | β-thalassaemia carrier prevalence | | | | | Estimated carrier counts | | |
|---|---|---|---|---|---|---|---|---|---|
| **ADM1** | **ADM2** | **Mean** | **SD** | **CI95%** | **Q25** | **Q75** | **Mean** | **Q25** | **Q75** |
| Isparta | Keciborlu | 3·683 | 2·427 | 9·174 | 2·030 | 4·656 | 509 | 281 | 643 |
| Isparta | Sarkikaraagac | 3·384 | 2·352 | 8·698 | 1·791 | 4·294 | 842 | 441 | 1071 |
| Isparta | Senirkent | 3·430 | 2·305 | 8·585 | 1·859 | 4·325 | 359 | 191 | 454 |
| Isparta | Sutculer | 4·209 | 2·878 | 10·707 | 2·239 | 5·346 | 434 | 235 | 551 |
| Isparta | Uluborlu | 3·512 | 2·337 | 8·654 | 1·920 | 4·443 | 246 | 132 | 313 |
| Isparta | Yalvac | 3·183 | 2·184 | 8·239 | 1·693 | 4·019 | 1476 | 776 | 1852 |
| Isparta | Yenibarbademli | 3·747 | 2·558 | 9·597 | 1·997 | 4·760 | 39 | 21 | 50 |
| Istanbul | Arnavutkoy | 1·901 | 1·653 | 5·986 | 0·838 | 2·417 | 6261 | 2797 | 7847 |
| Istanbul | Atasehir | 1·662 | 1·407 | 4·828 | 0·791 | 2·065 | 6620 | 3150 | 8223 |
| Istanbul | Avcilar | 1·882 | 1·570 | 5·678 | 0·876 | 2·356 | 9544 | 4439 | 11963 |
| Istanbul | Bagcilar | 1·766 | 1·337 | 5·030 | 0·887 | 2·218 | 18517 | 9299 | 23256 |
| Istanbul | Bahcelievler | 1·827 | 1·534 | 5·616 | 0·849 | 2·294 | 63476 | 30050 | 79622 |
| Istanbul | Bakirkoy | 1·841 | 1·603 | 5·788 | 0·825 | 2·324 | 10924 | 4899 | 13752 |
| Istanbul | Basaksehir | 1·866 | 1·550 | 5·483 | 0·870 | 2·359 | 9315 | 4303 | 11766 |
| Istanbul | Bayrampasa | 1·809 | 1·547 | 5·393 | 0·820 | 2·268 | 15438 | 6996 | 19359 |
| Istanbul | Besiktas | 1·755 | 1·478 | 5·469 | 0·802 | 2·188 | 23091 | 10403 | 28770 |
| Istanbul | Beykoz | 1·706 | 1·448 | 5·358 | 0·771 | 2·144 | 4193 | 1961 | 5328 |
| Istanbul | Beylikduzu | 1·891 | 1·573 | 5·815 | 0·895 | 2·358 | 7783 | 3685 | 9700 |
| Istanbul | Beyoglu | 1·778 | 1·413 | 5·120 | 0·814 | 2·278 | 7867 | 3601 | 10076 |
| Istanbul | Buyukcekmece | 1·945 | 1·609 | 5·746 | 0·890 | 2·450 | 9326 | 4269 | 11817 |
| Istanbul | Catalca | 2·107 | 1·902 | 6·925 | 0·902 | 2·642 | 1794 | 787 | 2254 |
| Istanbul | Cekmekoy | 1·667 | 1·375 | 4·929 | 0·773 | 2·096 | 11142 | 5231 | 13888 |
| Istanbul | Esenler | 1·839 | 1·578 | 6·092 | 0·824 | 2·266 | 4534 | 2031 | 5586 |
| Istanbul | Esenyurt | 1·884 | 1·483 | 5·441 | 0·862 | 2·461 | 12873 | 5887 | 16819 |
| Istanbul | Eyupsultan | 1·808 | 1·538 | 5·547 | 0·830 | 2·286 | 16614 | 7822 | 21329 |
| Istanbul | Fatih | 1·757 | 1·368 | 4·935 | 0·811 | 2·260 | 18036 | 8339 | 23211 |
| Istanbul | Gaziosmanpasa | 1·791 | 1·454 | 5·279 | 0·829 | 2·271 | 41657 | 19081 | 52683 |
| Istanbul | Gungoren | 1·823 | 1·541 | 5·626 | 0·846 | 2·280 | 45839 | 21662 | 57432 |
| Istanbul | Kadikoy | 1·721 | 1·428 | 5·098 | 0·789 | 2·205 | 7274 | 3336 | 9318 |
| Istanbul | Kagithane | 1·806 | 1·596 | 6·167 | 0·782 | 2·212 | 9221 | 3994 | 11293 |
| Istanbul | Kartal | 1·690 | 1·301 | 4·556 | 0·838 | 2·130 | 4757 | 2359 | 5994 |
| Istanbul | Kucukcekmece | 1·829 | 1·514 | 5·353 | 0·877 | 2·267 | 16693 | 8066 | 20800 |
| Istanbul | Maltepe | 1·742 | 1·412 | 5·284 | 0·814 | 2·188 | 10694 | 4970 | 13476 |
| Istanbul | Pendik | 1·698 | 1·402 | 5·094 | 0·793 | 2·124 | 9771 | 4511 | 12191 |
| Istanbul | Sancaktepe | 1·668 | 1·311 | 5·006 | 0·815 | 2·088 | 3661 | 1809 | 4483 |
| Istanbul | Sariyer | 1·767 | 1·509 | 5·502 | 0·808 | 2·179 | 6613 | 3061 | 8140 |
| Istanbul | Sile | 1·611 | 1·352 | 4·975 | 0·744 | 2·019 | 617 | 283 | 771 |
| Istanbul | Silivri | 2·124 | 1·820 | 6·650 | 0·947 | 2·688 | 3706 | 1674 | 4732 |
| Istanbul | Sisli | 1·768 | 1·454 | 5·439 | 0·806 | 2·216 | 31246 | 14193 | 39357 |
| Istanbul | Sultanbeyli | 1·692 | 1·275 | 4·821 | 0·820 | 2·152 | 6133 | 2972 | 7801 |
| Istanbul | Sultangazi | 1·815 | 1·529 | 5·524 | 0·815 | 2·273 | 11824 | 5307 | 14807 |
| Istanbul | Tuzla | 1·687 | 1·331 | 4·812 | 0·804 | 2·171 | 5707 | 2719 | 7336 |
| Istanbul | Umraniye | 1·782 | 1·464 | 5·513 | 0·795 | 2·276 | 4867 | 2171 | 6219 |
| Istanbul | Uskudar | 1·722 | 1·421 | 5·408 | 0·773 | 2·132 | 7130 | 3199 | 8825 |
| Istanbul | Zeytinburnu | 1·811 | 1·509 | 5·459 | 0·828 | 2·289 | 31098 | 14220 | 39351 |
| Izmir | Aliaga | 4·094 | 2·703 | 10·254 | 2·239 | 5·158 | 4255 | 2291 | 5350 |
| Izmir | Balcova | 4·357 | 2·911 | 11·506 | 2·483 | 5·456 | 3937 | 2243 | 4930 |
| Izmir | Bayindir | 4·108 | 2·537 | 9·588 | 2·349 | 5·172 | 1843 | 1049 | 2340 |
| Izmir | Bayrakli | 4·173 | 2·608 | 9·656 | 2·404 | 5·082 | 14694 | 8465 | 17895 |
| Izmir | Bergama | 3·535 | 2·325 | 8·714 | 1·937 | 4·475 | 3788 | 2112 | 4790 |
| Izmir | Beydag | 3·791 | 2·320 | 8·614 | 2·163 | 4·773 | 396 | 222 | 497 |
| Izmir | Bornova | 4·120 | 2·634 | 9·894 | 2·303 | 5·203 | 14479 | 8017 | 17996 |
| Izmir | Buca | 4·329 | 2·819 | 10·598 | 2·377 | 5·490 | 16140 | 8754 | 20265 |
| Izmir | Cesme | 4·765 | 3·325 | 12·638 | 2·481 | 6·066 | 972 | 517 | 1218 |
| Izmir | Cigli | 4·348 | 2·811 | 10·533 | 2·393 | 5·479 | 13092 | 7117 | 16599 |
| Izmir | Dikili | 3·811 | 2·562 | 9·627 | 2·072 | 4·789 | 1222 | 662 | 1535 |
| Izmir | Foca | 4·360 | 2·998 | 10·956 | 2·325 | 5·506 | 1013 | 538 | 1290 |

| | | β-thalassaemia carrier prevalence | | | | | Estimated carrier counts | | |
|---|---|---|---|---|---|---|---|---|---|
| ADM1 | ADM2 | Mean | SD | CI95% | Q25 | Q75 | Mean | Q25 | Q75 |
| Izmir | Gaziemir | 4·336 | 2·665 | 9·689 | 2·476 | 5·498 | 7203 | 4113 | 9166 |
| Izmir | Guzelbahce | 4·453 | 2·924 | 10·873 | 2·412 | 5·592 | 1039 | 561 | 1304 |
| Izmir | Karabaglar | 4·337 | 2·771 | 10·721 | 2·433 | 5·444 | 34009 | 18995 | 43007 |
| Izmir | Karaburun | 4·580 | 3·167 | 11·860 | 2·407 | 5·829 | 409 | 210 | 515 |
| Izmir | Karsiyaka | 4·180 | 2·491 | 9·552 | 2·391 | 5·400 | 2405 | 1376 | 3107 |
| Izmir | Kemalpasa | 4·058 | 2·543 | 9·529 | 2·302 | 5·118 | 4824 | 2730 | 6076 |
| Izmir | Kinik | 3·522 | 2·276 | 8·501 | 1·964 | 4·454 | 1083 | 597 | 1356 |
| Izmir | Kiraz | 3·727 | 2·380 | 8·889 | 2·107 | 4·685 | 1620 | 917 | 2039 |
| Izmir | Konak | 4·292 | 2·786 | 10·834 | 2·399 | 5·365 | 66324 | 37108 | 82982 |
| Izmir | Menderes | 4·315 | 2·776 | 10·340 | 2·384 | 5·495 | 5493 | 3037 | 7080 |
| Izmir | Menemen | 4·248 | 2·819 | 10·404 | 2·333 | 5·362 | 8882 | 4914 | 11173 |
| Izmir | Narlidere | 4·381 | 2·855 | 10·394 | 2·380 | 5·613 | 2600 | 1394 | 3324 |
| Izmir | Odemis | 3·927 | 2·491 | 9·354 | 2·221 | 4·919 | 5389 | 3064 | 6828 |
| Izmir | Seferihisar | 4·477 | 2·977 | 11·030 | 2·423 | 5·650 | 1921 | 1052 | 2406 |
| Izmir | Selcuk | 4·144 | 2·612 | 9·778 | 2·331 | 5·238 | 1406 | 780 | 1779 |
| Izmir | Tire | 4·024 | 2·546 | 9·419 | 2·270 | 5·062 | 3467 | 1942 | 4405 |
| Izmir | Torbali | 4·217 | 2·701 | 10·177 | 2·349 | 5·337 | 8976 | 4931 | 11412 |
| Izmir | Urla | 4·647 | 3·173 | 11·904 | 2·467 | 5·891 | 2655 | 1428 | 3331 |
| Kahramanmaras | Afsin | 1·426 | 1·021 | 3·786 | 0·745 | 1·794 | 1205 | 629 | 1516 |
| Kahramanmaras | Andirin | 1·922 | 1·342 | 4·991 | 1·019 | 2·428 | 606 | 325 | 773 |
| Kahramanmaras | Caglayancerit | 1·587 | 1·047 | 3·868 | 0·888 | 1·986 | 365 | 204 | 462 |
| Kahramanmaras | Dulkadiroglu | 1·727 | 1·140 | 4·237 | 0·955 | 2·173 | 3437 | 1918 | 4322 |
| Kahramanmaras | Ekinozu | 1·534 | 1·023 | 3·788 | 0·846 | 1·922 | 168 | 94 | 209 |
| Kahramanmaras | Elbistan | 1·421 | 0·999 | 3·690 | 0·757 | 1·784 | 2137 | 1155 | 2650 |
| Kahramanmaras | Goksun | 1·539 | 1·105 | 4·075 | 0·805 | 1·940 | 783 | 402 | 982 |
| Kahramanmaras | Nurhak | 1·496 | 0·978 | 3·629 | 0·836 | 1·879 | 177 | 98 | 222 |
| Kahramanmaras | Onikisubat | 1·725 | 1·164 | 4·324 | 0·941 | 2·170 | 9059 | 5000 | 11268 |
| Kahramanmaras | Pazarcik | 1·817 | 1·193 | 4·471 | 1·008 | 2·289 | 1379 | 754 | 1730 |
| Kahramanmaras | Turkoglu | 1·979 | 1·346 | 4·918 | 1·079 | 2·487 | 1553 | 858 | 1950 |
| Karabuk | Eflani | 1·776 | 2·217 | 7·342 | 0·543 | 2·164 | 143 | 45 | 178 |
| Karabuk | Eskipazar | 1·773 | 2·093 | 6·655 | 0·605 | 2·188 | 236 | 80 | 292 |
| Karabuk | Merkez | 1·781 | 2·186 | 6·850 | 0·575 | 2·201 | 2052 | 647 | 2494 |
| Karabuk | Ovacik | 1·823 | 2·226 | 7·264 | 0·568 | 2·266 | 66 | 22 | 83 |
| Karabuk | Safranbolu | 1·817 | 2·288 | 7·409 | 0·566 | 2·222 | 1816 | 586 | 2220 |
| Karabuk | Yenice | 1·730 | 2·013 | 6·417 | 0·603 | 2·168 | 339 | 113 | 422 |
| Karaman | Ayranci | 3·547 | 2·621 | 9·775 | 1·782 | 4·502 | 261 | 134 | 334 |
| Karaman | Basyayla | 5·672 | 3·861 | 14·416 | 3·035 | 7·241 | 463 | 245 | 582 |
| Karaman | Ermenek | 6·163 | 4·270 | 15·909 | 3·224 | 7·840 | 1719 | 896 | 2200 |
| Karaman | Karaman merkez | 4·184 | 2·965 | 10·997 | 2·170 | 5·318 | 8583 | 4498 | 10919 |
| Karaman | Kazimkarabekir | 3·973 | 2·749 | 10·319 | 2·103 | 5·045 | 187 | 98 | 244 |
| Karaman | Sariveliler | 6·046 | 4·075 | 15·402 | 3·269 | 7·624 | 457 | 246 | 585 |
| Kars | Akyaka | 1·433 | 1·611 | 5·292 | 0·529 | 1·766 | 149 | 56 | 185 |
| Kars | Arpacay | 1·355 | 1·427 | 4·911 | 0·513 | 1·679 | 208 | 78 | 256 |
| Kars | Digor | 1·370 | 1·346 | 4·666 | 0·550 | 1·709 | 272 | 110 | 340 |
| Kars | Kagizman | 1·262 | 1·152 | 4·122 | 0·541 | 1·579 | 564 | 242 | 702 |
| Kars | Kars merkez | 1·283 | 1·249 | 4·375 | 0·518 | 1·601 | 1548 | 621 | 1941 |
| Kars | Sarikamis | 1·089 | 0·912 | 3·279 | 0·512 | 1·365 | 410 | 191 | 520 |
| Kars | Selim | 1·146 | 1·030 | 3·666 | 0·506 | 1·439 | 257 | 110 | 324 |
| Kars | Susuz | 1·244 | 1·220 | 4·318 | 0·507 | 1·545 | 113 | 46 | 140 |
| Kastamonu | Abana | 1·861 | 2·580 | 8·555 | 0·481 | 2·228 | 190 | 49 | 225 |
| Kastamonu | Agli | 1·781 | 2·265 | 7·811 | 0·490 | 2·151 | 55 | 12 | 66 |
| Kastamonu | Arac | 1·781 | 2·195 | 7·369 | 0·538 | 2·176 | 340 | 100 | 405 |
| Kastamonu | Azdavay | 1·797 | 2·325 | 7·837 | 0·510 | 2·158 | 130 | 38 | 157 |
| Kastamonu | Bozkurt | 1·818 | 2·411 | 8·230 | 0·482 | 2·182 | 61 | 16 | 72 |
| Kastamonu | Catalzeytin | 1·811 | 2·493 | 8·373 | 0·485 | 2·138 | 71 | 20 | 84 |
| Kastamonu | Cide | 1·840 | 2·438 | 8·037 | 0·519 | 2·199 | 236 | 66 | 278 |
| Kastamonu | Daday | 1·754 | 2·205 | 7·387 | 0·513 | 2·130 | 146 | 39 | 178 |

| | | β-thalassaemia carrier prevalence | | | | | Estimated carrier counts | | |
|---|---|---|---|---|---|---|---|---|---|
| ADM1 | ADM2 | Mean | SD | CI95% | Q25 | Q75 | Mean | Q25 | Q75 |
| Kastamonu | Devrekani | 1·795 | 2·340 | 7·994 | 0·488 | 2·162 | 221 | 58 | 266 |
| Kastamonu | Doganyurt | 1·871 | 2·430 | 8·708 | 0·500 | 2·207 | 108 | 29 | 129 |
| Kastamonu | Hanonu | 1·746 | 2·276 | 7·938 | 0·478 | 2·070 | 83 | 20 | 96 |
| Kastamonu | Ihsangazi | 1·734 | 2·146 | 7·102 | 0·517 | 2·125 | 91 | 25 | 109 |
| Kastamonu | Inebolu | 1·840 | 2·416 | 8·183 | 0·497 | 2·228 | 351 | 99 | 427 |
| Kastamonu | Kastamonu merkez | 1·740 | 2·187 | 7·466 | 0·502 | 2·114 | 2752 | 784 | 3340 |
| Kastamonu | Kure | 1·804 | 2·291 | 7·958 | 0·501 | 2·193 | 86 | 22 | 104 |
| Kastamonu | Pinarbasi | 1·796 | 2·279 | 7·836 | 0·531 | 2·156 | 90 | 28 | 113 |
| Kastamonu | Senpazar | 1·794 | 2·257 | 8·010 | 0·502 | 2·154 | 78 | 24 | 94 |
| Kastamonu | Seydiler | 1·789 | 2·280 | 7·712 | 0·493 | 2·185 | 88 | 23 | 107 |
| Kastamonu | Taskopru | 1·742 | 2·242 | 7·712 | 0·488 | 2·077 | 706 | 188 | 825 |
| Kastamonu | Tosya | 1·718 | 2·127 | 7·213 | 0·503 | 2·087 | 707 | 197 | 860 |
| Kayseri | Akkisla | 1·370 | 1·273 | 4·549 | 0·581 | 1·710 | 76 | 32 | 98 |
| Kayseri | Bunyan | 1·419 | 1·246 | 4·507 | 0·625 | 1·791 | 450 | 194 | 560 |
| Kayseri | Develi | 1·705 | 1·431 | 5·228 | 0·787 | 2·143 | 1151 | 519 | 1456 |
| Kayseri | Felahiye | 1·475 | 1·428 | 5·116 | 0·589 | 1·844 | 86 | 34 | 109 |
| Kayseri | Hacilar | 1·629 | 1·458 | 5·263 | 0·719 | 2·034 | 316 | 138 | 400 |
| Kayseri | Incesu | 1·664 | 1·496 | 5·370 | 0·717 | 2·088 | 492 | 210 | 613 |
| Kayseri | Kocasinan | 1·553 | 1·450 | 5·202 | 0·650 | 1·950 | 9082 | 3954 | 11612 |
| Kayseri | Melikgazi | 1·514 | 1·320 | 4·755 | 0·668 | 1·905 | 7074 | 3136 | 9018 |
| Kayseri | Ozvatan | 1·424 | 1·332 | 4·795 | 0·580 | 1·795 | 57 | 21 | 72 |
| Kayseri | Pinarbasi | 1·337 | 1·161 | 4·213 | 0·597 | 1·677 | 290 | 128 | 371 |
| Kayseri | Sarioglan | 1·385 | 1·310 | 4·656 | 0·568 | 1·738 | 185 | 79 | 235 |
| Kayseri | Sariz | 1·398 | 1·078 | 3·979 | 0·684 | 1·771 | 131 | 63 | 169 |
| Kayseri | Talas | 1·528 | 1·336 | 4·819 | 0·685 | 1·915 | 2415 | 1028 | 3081 |
| Kayseri | Tomarza | 1·545 | 1·287 | 4·663 | 0·716 | 1·946 | 350 | 158 | 440 |
| Kayseri | Yahyali | 1·998 | 1·612 | 5·891 | 0·941 | 2·525 | 669 | 314 | 859 |
| Kayseri | Yesilhisar | 1·847 | 1·582 | 5·704 | 0·833 | 2·319 | 295 | 132 | 373 |
| Kilis | Elbeyli | 2·797 | 1·980 | 7·390 | 1·478 | 3·553 | 167 | 90 | 216 |
| Kilis | Kilis (merkez) | 2·757 | 2·051 | 7·535 | 1·401 | 3·479 | 3756 | 1918 | 4716 |
| Kilis | Musabeyli | 2·497 | 1·756 | 6·480 | 1·308 | 3·182 | 315 | 163 | 401 |
| Kilis | Polateli | 2·581 | 1·822 | 6·814 | 1·356 | 3·266 | 138 | 73 | 177 |
| Kirikkale | Bahsili | 1·857 | 1·871 | 6·642 | 0·716 | 2·345 | 277 | 104 | 345 |
| Kirikkale | Baliseyh | 1·753 | 1·883 | 6·574 | 0·616 | 2·181 | 108 | 35 | 127 |
| Kirikkale | Celebi | 1·862 | 1·864 | 6·516 | 0·735 | 2·335 | 60 | 23 | 74 |
| Kirikkale | Delice | 1·699 | 1·790 | 6·279 | 0·595 | 2·148 | 156 | 53 | 195 |
| Kirikkale | Karakeceli | 1·845 | 1·891 | 6·482 | 0·723 | 2·289 | 37 | 15 | 45 |
| Kirikkale | Keskin | 1·800 | 1·839 | 6·562 | 0·673 | 2·253 | 281 | 101 | 353 |
| Kirikkale | Kirikkale (merkez) | 1·783 | 1·857 | 6·494 | 0·647 | 2·239 | 3530 | 1275 | 4477 |
| Kirikkale | Sulakyurt | 1·716 | 1·860 | 6·480 | 0·589 | 2·161 | 96 | 31 | 122 |
| Kirikkale | Yahsihan | 1·817 | 1·891 | 6·537 | 0·671 | 2·285 | 536 | 196 | 664 |
| Kirklareli | Babaeski | 3·059 | 2·769 | 9·901 | 1·304 | 3·871 | 1434 | 624 | 1825 |
| Kirklareli | Demirkoy | 2·707 | 2·634 | 9·455 | 1·066 | 3·412 | 143 | 58 | 174 |
| Kirklareli | Kirklareli merkez | 3·142 | 2·951 | 10·662 | 1·287 | 3·957 | 3333 | 1356 | 4259 |
| Kirklareli | Kofcaz | 3·393 | 3·208 | 11·641 | 1·358 | 4·266 | 73 | 28 | 93 |
| Kirklareli | Luleburgaz | 2·730 | 2·457 | 8·859 | 1·170 | 3·430 | 4146 | 1840 | 5157 |
| Kirklareli | Pehlivankoy | 3·168 | 2·678 | 9·952 | 1·417 | 3·969 | 65 | 29 | 82 |
| Kirklareli | Pinarhisar | 2·778 | 2·608 | 9·454 | 1·138 | 3·516 | 548 | 225 | 693 |
| Kirklareli | Vize | 2·522 | 2·352 | 8·409 | 1·053 | 3·182 | 762 | 325 | 944 |
| Kirsehir | Akcakent | 1·732 | 1·783 | 6·187 | 0·626 | 2·180 | 72 | 25 | 89 |
| Kirsehir | Akpinar | 1·804 | 1·911 | 6·741 | 0·649 | 2·253 | 137 | 50 | 174 |
| Kirsehir | Boztepe | 1·722 | 1·704 | 6·030 | 0·653 | 2·193 | 96 | 35 | 120 |
| Kirsehir | Cicekdagi | 1·699 | 1·730 | 6·201 | 0·616 | 2·151 | 250 | 89 | 321 |
| Kirsehir | Kaman | 1·862 | 1·856 | 6·427 | 0·727 | 2·337 | 652 | 254 | 813 |
| Kirsehir | Kirsehir merkez | 1·819 | 1·754 | 6·297 | 0·716 | 2·300 | 3010 | 1180 | 3769 |
| Kirsehir | Mucur | 1·759 | 1·663 | 5·955 | 0·699 | 2·230 | 337 | 131 | 416 |
| Kocaeli | Basiskele | 1·632 | 1·264 | 4·627 | 0·808 | 2·056 | 2836 | 1448 | 3503 |

| | | β-thalassaemia carrier prevalence | | | | | Estimated carrier counts | | |
|---|---|---|---|---|---|---|---|---|---|
| ADM1 | ADM2 | Mean | SD | CI95% | Q25 | Q75 | Mean | Q25 | Q75 |
| Kocaeli | Cayirova | 1·660 | 1·270 | 4·912 | 0·796 | 2·123 | 1660 | 796 | 2124 |
| Kocaeli | Darica | 1·730 | 1·439 | 5·015 | 0·823 | 2·142 | 9571 | 4529 | 11955 |
| Kocaeli | Derince | 1·583 | 1·262 | 4·554 | 0·775 | 2·000 | 245 | 121 | 310 |
| Kocaeli | Dilovasi | 1·629 | 1·288 | 4·567 | 0·784 | 2·058 | 398 | 194 | 501 |
| Kocaeli | Gebze | 1·656 | 1·349 | 4·915 | 0·797 | 2·066 | 12423 | 5910 | 15552 |
| Kocaeli | Golcuk | 1·670 | 1·281 | 4·734 | 0·833 | 2·102 | 1711 | 843 | 2125 |
| Kocaeli | Izmit | 1·591 | 1·288 | 4·699 | 0·761 | 1·990 | 2742 | 1325 | 3458 |
| Kocaeli | Kandira | 1·551 | 1·276 | 4·667 | 0·727 | 1·944 | 755 | 349 | 947 |
| Kocaeli | Karamursel | 1·711 | 1·343 | 4·795 | 0·842 | 2·137 | 1047 | 527 | 1284 |
| Kocaeli | Kartepe | 1·621 | 1·285 | 4·652 | 0·789 | 2·052 | 2439 | 1202 | 3115 |
| Kocaeli | Korfez | 1·615 | 1·288 | 4·757 | 0·785 | 2·028 | 1231 | 598 | 1534 |
| Konya | Ahirli | 4·308 | 2·985 | 11·117 | 2·296 | 5·439 | 159 | 84 | 201 |
| Konya | Akoren | 3·749 | 2·511 | 9·325 | 2·045 | 4·723 | 235 | 130 | 294 |
| Konya | Aksehir | 2·992 | 2·094 | 7·843 | 1·585 | 3·771 | 2864 | 1514 | 3620 |
| Konya | Altinekin | 2·409 | 1·798 | 6·620 | 1·213 | 3·053 | 366 | 183 | 458 |
| Konya | Beysehir | 3·633 | 2·486 | 9·189 | 1·945 | 4·585 | 2838 | 1519 | 3585 |
| Konya | Bozkir | 4·450 | 2·959 | 11·041 | 2·414 | 5·646 | 1062 | 578 | 1363 |
| Konya | Celtik | 2·419 | 1·811 | 6·747 | 1·230 | 3·045 | 224 | 116 | 286 |
| Konya | Cihanbeyli | 2·219 | 1·763 | 6·429 | 1·067 | 2·808 | 1137 | 536 | 1431 |
| Konya | Cumra | 3·464 | 2·376 | 8·869 | 1·854 | 4·364 | 2417 | 1269 | 3035 |
| Konya | Derbent | 3·064 | 2·120 | 7·832 | 1·653 | 3·838 | 132 | 71 | 165 |
| Konya | Derebucak | 4·259 | 2·910 | 10·895 | 2·267 | 5·406 | 253 | 135 | 325 |
| Konya | Doganhisar | 3·083 | 2·102 | 7·806 | 1·656 | 3·917 | 449 | 240 | 566 |
| Konya | Emirgazi | 2·415 | 1·828 | 6·676 | 1·204 | 3·057 | 193 | 100 | 244 |
| Konya | Eregli | 2·766 | 2·123 | 7·709 | 1·368 | 3·509 | 4539 | 2243 | 5820 |
| Konya | Guneysinir | 4·252 | 2·881 | 10·766 | 2·265 | 5·430 | 428 | 227 | 543 |
| Konya | Hadim | 4·951 | 3·343 | 12·407 | 2·648 | 6·318 | 619 | 324 | 790 |
| Konya | Halkapinar | 3·201 | 2·504 | 9·250 | 1·532 | 4·052 | 132 | 64 | 168 |
| Konya | Huyuk | 3·308 | 2·238 | 8·351 | 1·800 | 4·172 | 482 | 261 | 605 |
| Konya | Ilgin | 2·812 | 1·925 | 7·197 | 1·514 | 3·534 | 1539 | 845 | 1932 |
| Konya | Kadinhani | 2·548 | 1·814 | 6·692 | 1·338 | 3·207 | 858 | 462 | 1076 |
| Konya | Karapinar | 2·829 | 2·052 | 7·548 | 1·453 | 3·583 | 1507 | 768 | 1886 |
| Konya | Karatay | 2·719 | 1·953 | 7·211 | 1·416 | 3·430 | 7421 | 3962 | 9350 |
| Konya | Kulu | 2·064 | 1·819 | 6·498 | 0·915 | 2·596 | 1112 | 476 | 1394 |
| Konya | Meram | 3·184 | 2·160 | 8·055 | 1·713 | 4·026 | 17313 | 9382 | 22089 |
| Konya | Sarayonu | 2·447 | 1·755 | 6·471 | 1·269 | 3·098 | 698 | 370 | 880 |
| Konya | Selcuklu | 2·724 | 1·881 | 6·978 | 1·460 | 3·440 | 17267 | 9232 | 21827 |
| Konya | Seydisehir | 3·845 | 2·603 | 9·772 | 2·073 | 4·840 | 2751 | 1494 | 3445 |
| Konya | Taskent | 5·250 | 3·568 | 13·370 | 2·794 | 6·712 | 221 | 117 | 282 |
| Konya | Tuzlukcu | 2·751 | 1·880 | 6·914 | 1·472 | 3·486 | 167 | 89 | 214 |
| Konya | Yalihuyuk | 4·272 | 2·881 | 10·097 | 2·281 | 5·453 | 2 | 1 | 3 |
| Konya | Yunak | 2·486 | 1·793 | 6·665 | 1·285 | 3·144 | 551 | 283 | 695 |
| Kutahya | Altintas | 2·859 | 1·885 | 7·056 | 1·567 | 3·622 | 444 | 242 | 563 |
| Kutahya | Aslanapa | 2·793 | 1·850 | 6·952 | 1·540 | 3·495 | 223 | 119 | 278 |
| Kutahya | Cavdarhisar | 2·827 | 1·876 | 7·005 | 1·558 | 3·557 | 164 | 91 | 205 |
| Kutahya | Domanic | 2·289 | 1·628 | 5·931 | 1·191 | 2·900 | 333 | 170 | 424 |
| Kutahya | Dumlupinar | 3·001 | 2·005 | 7·375 | 1·631 | 3·774 | 99 | 53 | 122 |
| Kutahya | Emet | 2·752 | 1·871 | 6·979 | 1·486 | 3·466 | 687 | 368 | 860 |
| Kutahya | Gediz Merkez | 2·999 | 1·982 | 7·452 | 1·659 | 3·766 | 1520 | 831 | 1905 |
| Kutahya | Hisarcik | 2·890 | 1·965 | 7·240 | 1·547 | 3·678 | 199 | 109 | 254 |
| Kutahya | Kutahya merkez | 2·595 | 1·809 | 6·680 | 1·379 | 3·277 | 7638 | 4085 | 9563 |
| Kutahya | Pazarlar | 3·041 | 1·982 | 7·131 | 1·699 | 3·823 | 137 | 76 | 172 |
| Kutahya | Saphane | 3·046 | 2·008 | 7·500 | 1·656 | 3·863 | 179 | 95 | 224 |
| Kutahya | Simav | 2·907 | 1·958 | 7·276 | 1·583 | 3·666 | 1836 | 994 | 2311 |
| Kutahya | Tavsanli | 2·525 | 1·747 | 6·490 | 1·347 | 3·185 | 2589 | 1390 | 3266 |
| Malatya | Akcadag | 1·279 | 0·908 | 3·350 | 0·675 | 1·613 | 309 | 159 | 391 |
| Malatya | Arapgir | 1·092 | 0·929 | 3·332 | 0·503 | 1·375 | 104 | 47 | 133 |

| | | β-thalassaemia carrier prevalence | | | | | Estimated carrier counts | | |
|---|---|---|---|---|---|---|---|---|---|
| ADM1 | ADM2 | Mean | SD | CI95% | Q25 | Q75 | Mean | Q25 | Q75 |
| Malatya | Arguvan | 1·124 | 0·918 | 3·333 | 0·533 | 1·411 | 77 | 37 | 99 |
| Malatya | Battalgazi | 1·230 | 0·882 | 3·240 | 0·651 | 1·545 | 4739 | 2490 | 6054 |
| Malatya | Darende | 1·296 | 0·963 | 3·541 | 0·668 | 1·621 | 338 | 171 | 422 |
| Malatya | Dogansehir | 1·375 | 0·929 | 3·425 | 0·752 | 1·725 | 530 | 286 | 669 |
| Malatya | Doganyol | 1·265 | 0·952 | 3·543 | 0·638 | 1·593 | 45 | 24 | 59 |
| Malatya | Hekimhan | 1·149 | 0·934 | 3·355 | 0·552 | 1·437 | 176 | 83 | 226 |
| Malatya | Kale | 1·245 | 0·941 | 3·467 | 0·641 | 1·555 | 62 | 34 | 78 |
| Malatya | Kuluncak | 1·186 | 0·974 | 3·510 | 0·567 | 1·484 | 87 | 41 | 106 |
| Malatya | Puturge | 1·299 | 0·922 | 3·401 | 0·685 | 1·637 | 171 | 88 | 214 |
| Malatya | Yazihan | 1·187 | 0·924 | 3·399 | 0·582 | 1·495 | 161 | 80 | 203 |
| Malatya | Yesilyurt | 1·257 | 0·893 | 3·276 | 0·665 | 1·578 | 3534 | 1863 | 4437 |
| Manisa | Ahmetli | 3·730 | 2·389 | 8·942 | 2·118 | 4·618 | 782 | 444 | 963 |
| Manisa | Akhisar | 3·375 | 2·173 | 8·131 | 1·889 | 4·251 | 6220 | 3484 | 7830 |
| Manisa | Alasehir | 3·532 | 2·226 | 8·202 | 2·003 | 4·441 | 3859 | 2144 | 4782 |
| Manisa | Demirci | 3·095 | 2·005 | 7·497 | 1·719 | 3·896 | 1122 | 621 | 1424 |
| Manisa | Golmarmara | 3·620 | 2·329 | 8·591 | 2·044 | 4·541 | 557 | 307 | 696 |
| Manisa | Gordes | 3·229 | 2·093 | 7·858 | 1·792 | 4·075 | 911 | 503 | 1142 |
| Manisa | Kirkagac | 3·242 | 2·170 | 8·054 | 1·763 | 4·107 | 1300 | 700 | 1661 |
| Manisa | Koprubasi | 3·338 | 2·140 | 8·024 | 1·868 | 4·190 | 415 | 234 | 520 |
| Manisa | Kula | 3·392 | 2·196 | 8·203 | 1·900 | 4·259 | 1512 | 826 | 1886 |
| Manisa | Salihli | 3·586 | 2·256 | 8·507 | 2·035 | 4·496 | 6197 | 3551 | 7820 |
| Manisa | Sarigol | 3·495 | 2·225 | 8·319 | 1·975 | 4·417 | 1197 | 686 | 1503 |
| Manisa | Saruhanli | 3·700 | 2·352 | 8·816 | 2·074 | 4·673 | 2283 | 1265 | 2891 |
| Manisa | Sehzadeler | 3·877 | 2·481 | 9·270 | 2·174 | 4·890 | 5093 | 2849 | 6401 |
| Manisa | Selendi | 3·157 | 1·982 | 7·377 | 1·790 | 3·973 | 637 | 356 | 821 |
| Manisa | Soma | 3·228 | 2·181 | 8·195 | 1·760 | 4·051 | 3729 | 2048 | 4690 |
| Manisa | Turgutlu | 3·869 | 2·469 | 9·125 | 2·197 | 4·873 | 6779 | 3909 | 8588 |
| Manisa | Yunusemre | 3·904 | 2·494 | 9·353 | 2·199 | 4·941 | 12331 | 7009 | 15387 |
| Mardin | Artuklu | 2·757 | 2·182 | 8·038 | 1·327 | 3·491 | 4511 | 2154 | 5753 |
| Mardin | Dargecit | 2·767 | 2·165 | 7·992 | 1·337 | 3·502 | 746 | 368 | 935 |
| Mardin | Derik | 2·473 | 1·815 | 6·653 | 1·265 | 3·132 | 1519 | 768 | 1926 |
| Mardin | Kiziltepe | 2·834 | 2·176 | 8·021 | 1·404 | 3·572 | 9380 | 4725 | 11869 |
| Mardin | Mazidagi | 2·426 | 1·851 | 6·888 | 1·201 | 3·066 | 914 | 449 | 1168 |
| Mardin | Midyat | 2·792 | 2·262 | 8·189 | 1·335 | 3·511 | 3262 | 1561 | 4043 |
| Mardin | Nusaybin | 3·095 | 2·457 | 9·030 | 1·483 | 3·924 | 4433 | 2103 | 5484 |
| Mardin | Omerli | 2·699 | 2·134 | 7·816 | 1·288 | 3·421 | 420 | 197 | 525 |
| Mardin | Savur | 2·429 | 1·977 | 7·175 | 1·138 | 3·065 | 559 | 259 | 704 |
| Mardin | Yesilli | 2·715 | 2·122 | 8·010 | 1·323 | 3·411 | 371 | 173 | 466 |
| Mersin | Akdeniz | 4·093 | 3·166 | 11·815 | 1·961 | 5·241 | 15966 | 7540 | 20537 |
| Mersin | Anamur | 7·839 | 5·227 | 19·696 | 4·218 | 9·974 | 4618 | 2496 | 5888 |
| Mersin | Aydincik | 7·963 | 5·480 | 20·826 | 4·174 | 10·180 | 1018 | 558 | 1304 |
| Mersin | Bozyazi | 7·668 | 5·214 | 19·569 | 4·052 | 9·783 | 1272 | 681 | 1621 |
| Mersin | Camliyayla | 3·372 | 2·660 | 9·898 | 1·602 | 4·280 | 307 | 147 | 385 |
| Mersin | Erdemli | 4·671 | 3·526 | 13·154 | 2·289 | 5·975 | 7370 | 3507 | 9455 |
| Mersin | Gulnar | 7·113 | 4·996 | 18·726 | 3·664 | 9·104 | 1884 | 958 | 2386 |
| Mersin | Mezitli | 4·357 | 3·324 | 12·528 | 2·129 | 5·542 | 9286 | 4516 | 11706 |
| Mersin | Mut | 5·538 | 3·995 | 14·871 | 2·812 | 7·067 | 3812 | 1934 | 4798 |
| Mersin | Silifke | 5·899 | 4·373 | 16·303 | 2·930 | 7·545 | 8123 | 4050 | 10341 |
| Mersin | Tarsus | 3·653 | 2·891 | 10·710 | 1·716 | 4·664 | 15379 | 7228 | 19680 |
| Mersin | Toroslar | 3·894 | 2·976 | 10·960 | 1·875 | 4·995 | 5837 | 2780 | 7602 |
| Mersin | Yenisehir | 4·372 | 3·375 | 12·355 | 2·074 | 5·640 | 8975 | 4241 | 11473 |
| Mugla | Bodrum | 4·189 | 3·034 | 11·248 | 2·148 | 5·320 | 6920 | 3472 | 8750 |
| Mugla | Dalaman | 4·247 | 2·850 | 10·614 | 2·318 | 5·339 | 2087 | 1113 | 2638 |
| Mugla | Datca | 4·349 | 3·384 | 12·268 | 2·148 | 5·508 | 1017 | 497 | 1311 |
| Mugla | Fethiye | 4·516 | 3·023 | 11·417 | 2·437 | 5·731 | 8052 | 4325 | 10173 |
| Mugla | Kavaklidere | 3·715 | 2·404 | 9·008 | 2·068 | 4·663 | 440 | 240 | 557 |
| Mugla | Koycegiz | 4·034 | 2·688 | 10·025 | 2·187 | 5·122 | 1820 | 976 | 2321 |

| ADM1 | ADM2 | β-thalassaemia carrier prevalence Mean | SD | CI95% | Q25 | Q75 | Estimated carrier counts Mean | Q25 | Q75 |
|---|---|---|---|---|---|---|---|---|---|
| Mugla | Marmaris | 4·205 | 2·936 | 10·910 | 2·197 | 5·348 | 3986 | 2165 | 5045 |
| Mugla | Mentese | 3·836 | 2·539 | 9·464 | 2·100 | 4·854 | 4755 | 2628 | 6033 |
| Mugla | Milas | 4·000 | 2·679 | 9·981 | 2·165 | 5·070 | 5848 | 3160 | 7489 |
| Mugla | Ortaca | 4·296 | 2·932 | 10·989 | 2·263 | 5·469 | 1954 | 1049 | 2491 |
| Mugla | Seydikemer | 4·768 | 3·146 | 11·748 | 2·603 | 6·051 | 3559 | 1877 | 4550 |
| Mugla | Ula | 3·941 | 2·634 | 9·798 | 2·140 | 4·949 | 929 | 492 | 1177 |
| Mugla | Yatagan | 3·848 | 2·498 | 9·356 | 2·136 | 4·837 | 1684 | 936 | 2113 |
| Mus | Bulanik | 1·391 | 1·167 | 4·193 | 0·650 | 1·749 | 1059 | 489 | 1342 |
| Mus | Haskoy | 1·613 | 1·388 | 4·995 | 0·745 | 2·027 | 405 | 187 | 504 |
| Mus | Korkut | 1·531 | 1·292 | 4·621 | 0·709 | 1·936 | 370 | 171 | 466 |
| Mus | Malazgirt | 1·356 | 1·099 | 3·958 | 0·644 | 1·717 | 722 | 332 | 921 |
| Mus | Mus merkez | 1·424 | 1·200 | 4·325 | 0·663 | 1·786 | 2989 | 1390 | 3749 |
| Mus | Varto | 1·218 | 1·024 | 3·705 | 0·570 | 1·529 | 378 | 176 | 478 |
| Nevsehir | Acigol | 1·896 | 1·635 | 5·933 | 0·828 | 2·416 | 411 | 175 | 521 |
| Nevsehir | Avanos | 1·696 | 1·577 | 5·767 | 0·683 | 2·158 | 577 | 236 | 740 |
| Nevsehir | Derinkuyu | 1·935 | 1·640 | 6·040 | 0·866 | 2·449 | 426 | 188 | 538 |
| Nevsehir | Gulsehir | 1·826 | 1·648 | 6·011 | 0·762 | 2·322 | 422 | 171 | 530 |
| Nevsehir | Hacibektas | 1·731 | 1·656 | 5·879 | 0·696 | 2·183 | 167 | 67 | 219 |
| Nevsehir | Kozakli | 1·644 | 1·609 | 5·739 | 0·639 | 2·080 | 216 | 79 | 272 |
| Nevsehir | Nevsehir merkez | 1·830 | 1·600 | 5·866 | 0·798 | 2·328 | 2742 | 1200 | 3513 |
| Nevsehir | Urgup | 1·783 | 1·581 | 5·685 | 0·762 | 2·276 | 797 | 335 | 1004 |
| Nigde | Altunhisar | 2·333 | 1·894 | 6·945 | 1·087 | 2·947 | 280 | 131 | 356 |
| Nigde | Bor | 2·432 | 1·995 | 7·220 | 1·143 | 3·071 | 1500 | 698 | 1913 |
| Nigde | Camardi | 2·234 | 1·828 | 6·638 | 1·054 | 2·810 | 267 | 128 | 338 |
| Nigde | Ciftlik | 2·141 | 1·789 | 6·503 | 0·986 | 2·710 | 576 | 267 | 737 |
| Nigde | Nigde (merkez) | 2·073 | 1·751 | 6·289 | 0·941 | 2·624 | 5236 | 2379 | 6694 |
| Nigde | Ulukisla | 2·786 | 2·209 | 8·077 | 1·323 | 3·548 | 590 | 281 | 752 |
| Ordu | Akkus | 1·336 | 1·691 | 5·763 | 0·405 | 1·596 | 284 | 87 | 335 |
| Ordu | Altinordu | 1·291 | 1·698 | 5·566 | 0·379 | 1·522 | 3078 | 930 | 3739 |
| Ordu | Aybasti | 1·286 | 1·654 | 5·399 | 0·386 | 1·532 | 264 | 80 | 315 |
| Ordu | Camas | 1·312 | 1·730 | 5·636 | 0·383 | 1·564 | 129 | 37 | 153 |
| Ordu | Catalpinar | 1·328 | 1·747 | 5·637 | 0·398 | 1·550 | 189 | 57 | 217 |
| Ordu | Caybasi | 1·407 | 1·916 | 6·081 | 0·396 | 1·677 | 180 | 50 | 212 |
| Ordu | Fatsa | 1·339 | 1·793 | 6·024 | 0·384 | 1·563 | 510 | 144 | 593 |
| Ordu | Golkoy | 1·245 | 1·547 | 5·125 | 0·390 | 1·483 | 309 | 99 | 371 |
| Ordu | Gulyali | 1·226 | 1·585 | 5·095 | 0·375 | 1·500 | 33 | 10 | 40 |
| Ordu | Gurgentepe | 1·257 | 1·598 | 5·235 | 0·384 | 1·519 | 158 | 49 | 191 |
| Ordu | Ikizce | 1·365 | 1·834 | 6·164 | 0·391 | 1·585 | 169 | 48 | 197 |
| Ordu | Kabaduz | 1·221 | 1·580 | 4·935 | 0·384 | 1·467 | 105 | 30 | 125 |
| Ordu | Kabatas | 1·308 | 1·572 | 5·599 | 0·387 | 1·591 | 154 | 46 | 189 |
| Ordu | Korgan | 1·294 | 1·640 | 5·504 | 0·395 | 1·537 | 367 | 110 | 433 |
| Ordu | Kumru | 1·332 | 1·710 | 5·888 | 0·395 | 1·592 | 366 | 106 | 432 |
| Ordu | Mesudiye | 1·214 | 1·534 | 5·012 | 0·391 | 1·445 | 155 | 48 | 184 |
| Ordu | Persembe | 1·328 | 1·765 | 5·904 | 0·381 | 1·561 | 278 | 80 | 330 |
| Ordu | Ulubey | 1·266 | 1·619 | 5·317 | 0·390 | 1·520 | 215 | 64 | 260 |
| Ordu | Unye | 1·353 | 1·781 | 5·966 | 0·387 | 1·600 | 1176 | 327 | 1437 |
| Osmaniye | Bahce | 2·189 | 1·572 | 5·621 | 1·154 | 2·753 | 464 | 247 | 582 |
| Osmaniye | Duzici | 2·191 | 1·522 | 5·639 | 1·162 | 2·749 | 1937 | 1020 | 2444 |
| Osmaniye | Hasanbeyli | 2·362 | 1·752 | 6·356 | 1·240 | 2·917 | 151 | 79 | 185 |
| Osmaniye | Kadirli | 2·229 | 1·637 | 6·018 | 1·151 | 2·806 | 3029 | 1538 | 3769 |
| Osmaniye | Osmaniye (merkez) | 2·540 | 1·868 | 6·909 | 1·302 | 3·204 | 7639 | 3868 | 9510 |
| Osmaniye | Sumbas | 2·123 | 1·522 | 5·565 | 1·117 | 2·680 | 368 | 197 | 461 |
| Osmaniye | Toprakkale | 2·698 | 2·018 | 7·427 | 1·358 | 3·409 | 575 | 287 | 726 |
| Rize | Ardesen | 0·996 | 0·911 | 3·269 | 0·430 | 1·249 | 150 | 63 | 185 |
| Rize | Camlihemsin | 0·967 | 0·881 | 3·097 | 0·423 | 1·207 | 42 | 15 | 53 |
| Rize | Cayeli | 0·984 | 0·935 | 3·301 | 0·413 | 1·229 | 271 | 109 | 339 |
| Rize | Derepazari | 1·029 | 1·000 | 3·472 | 0·424 | 1·264 | 53 | 22 | 65 |

| | | β-thalassaemia carrier prevalence | | | | | Estimated carrier counts | | |
|---|---|---|---|---|---|---|---|---|---|
| ADM1 | ADM2 | Mean | SD | CI95% | Q25 | Q75 | Mean | Q25 | Q75 |
| Rize | Findikli | 1·014 | 0·960 | 3·309 | 0·426 | 1·269 | 76 | 32 | 96 |
| Rize | Guneysu | 0·990 | 0·959 | 3·443 | 0·406 | 1·236 | 159 | 64 | 196 |
| Rize | Hemsin | 1·002 | 0·961 | 3·420 | 0·414 | 1·242 | 22 | 8 | 28 |
| Rize | Ikizdere | 0·948 | 0·854 | 3·038 | 0·417 | 1·185 | 57 | 25 | 76 |
| Rize | Iyidere | 0·986 | 0·939 | 3·400 | 0·394 | 1·234 | 54 | 22 | 68 |
| Rize | Kalkandere | 0·989 | 0·923 | 3·368 | 0·410 | 1·236 | 131 | 54 | 164 |
| Rize | Pazar | 1·001 | 0·952 | 3·483 | 0·411 | 1·257 | 281 | 114 | 350 |
| Rize | Rize merkezi | 1·002 | 0·960 | 3·456 | 0·411 | 1·256 | 474 | 191 | 593 |
| Sakarya | Adapazari | 1·570 | 1·294 | 4·637 | 0·753 | 1·949 | 6600 | 3195 | 8182 |
| Sakarya | Akyazi | 1·644 | 1·382 | 5·009 | 0·767 | 2·066 | 1451 | 698 | 1792 |
| Sakarya | Arifiye | 1·606 | 1·248 | 4·650 | 0·750 | 2·074 | 358 | 167 | 463 |
| Sakarya | Erenler | 1·607 | 1·349 | 4·760 | 0·761 | 2·005 | 1376 | 655 | 1723 |
| Sakarya | Ferizli | 1·550 | 1·350 | 4·872 | 0·704 | 1·951 | 219 | 99 | 274 |
| Sakarya | Geyve | 1·685 | 1·342 | 4·877 | 0·814 | 2·132 | 942 | 451 | 1197 |
| Sakarya | Hendek | 1·601 | 1·382 | 4·985 | 0·725 | 2·010 | 1541 | 697 | 1967 |
| Sakarya | Karapurcek | 1·631 | 1·312 | 4·766 | 0·784 | 2·047 | 334 | 158 | 423 |
| Sakarya | Karasu | 1·550 | 1·411 | 4·958 | 0·682 | 1·926 | 1009 | 419 | 1250 |
| Sakarya | Kaynarca | 1·541 | 1·326 | 4·840 | 0·699 | 1·936 | 382 | 175 | 475 |
| Sakarya | Kocaali | 1·579 | 1·455 | 5·126 | 0·670 | 1·986 | 220 | 92 | 277 |
| Sakarya | Pamukova | 1·700 | 1·379 | 5·019 | 0·816 | 2·132 | 438 | 207 | 540 |
| Sakarya | Sapanca | 1·636 | 1·307 | 4·841 | 0·792 | 2·053 | 704 | 336 | 884 |
| Sakarya | Serdivan | 1·577 | 1·251 | 4·535 | 0·753 | 1·995 | 1051 | 517 | 1333 |
| Sakarya | Sogutlu | 1·566 | 1·343 | 4·740 | 0·720 | 1·978 | 554 | 252 | 703 |
| Sakarya | Tarakli | 1·720 | 1·374 | 5·060 | 0·833 | 2·169 | 128 | 61 | 161 |
| Samsun | 19 Mayis | 1·539 | 2·018 | 6·880 | 0·421 | 1·807 | 388 | 104 | 449 |
| Samsun | Alacam | 1·575 | 2·041 | 7·030 | 0·443 | 1·896 | 409 | 112 | 494 |
| Samsun | Asarcik | 1·402 | 1·761 | 6·109 | 0·418 | 1·703 | 205 | 58 | 251 |
| Samsun | Atakum | 1·500 | 2·008 | 6·759 | 0·415 | 1·767 | 3143 | 860 | 3692 |
| Samsun | Ayvacik | 1·392 | 1·838 | 6·078 | 0·412 | 1·643 | 261 | 76 | 310 |
| Samsun | Bafra | 1·539 | 2·013 | 6·943 | 0·426 | 1·839 | 2306 | 623 | 2764 |
| Samsun | Canik | 1·464 | 1·929 | 6·524 | 0·412 | 1·731 | 154 | 45 | 182 |
| Samsun | Carsamba | 1·430 | 1·882 | 6·517 | 0·403 | 1·670 | 1969 | 543 | 2287 |
| Samsun | Havza | 1·509 | 1·928 | 6·558 | 0·436 | 1·807 | 625 | 187 | 742 |
| Samsun | Ilkadim | 1·500 | 2·027 | 6·727 | 0·412 | 1·775 | 4751 | 1287 | 5532 |
| Samsun | Kavak | 1·467 | 1·905 | 6·465 | 0·420 | 1·748 | 338 | 98 | 398 |
| Samsun | Ladik | 1·468 | 1·912 | 6·461 | 0·430 | 1·748 | 257 | 74 | 300 |
| Samsun | Salipazari | 1·390 | 1·766 | 6·059 | 0·408 | 1·641 | 257 | 76 | 303 |
| Samsun | Tekkekoy | 1·430 | 1·898 | 6·499 | 0·404 | 1·667 | 813 | 219 | 951 |
| Samsun | Terme | 1·411 | 1·881 | 6·389 | 0·387 | 1·652 | 936 | 263 | 1130 |
| Samsun | Vezirkopru | 1·562 | 1·989 | 6·749 | 0·453 | 1·871 | 1373 | 405 | 1636 |
| Samsun | Yakakent | 1·612 | 2·110 | 7·163 | 0·453 | 1·934 | 113 | 33 | 134 |
| Sanliurfa | Akcakale | 3·001 | 2·159 | 7·932 | 1·556 | 3·807 | 3169 | 1638 | 4051 |
| Sanliurfa | Birecik | 2·348 | 1·550 | 5·769 | 1·296 | 2·952 | 1863 | 1018 | 2313 |
| Sanliurfa | Bozova | 2·031 | 1·309 | 4·924 | 1·148 | 2·542 | 1086 | 614 | 1364 |
| Sanliurfa | Ceylanpinar | 2·954 | 2·169 | 7·988 | 1·506 | 3·739 | 4865 | 2400 | 6121 |
| Sanliurfa | Eyyubiye | 2·616 | 1·778 | 6·617 | 1·419 | 3·307 | 9259 | 5185 | 11565 |
| Sanliurfa | Halfeti | 1·997 | 1·298 | 4·803 | 1·124 | 2·501 | 850 | 480 | 1069 |
| Sanliurfa | Haliliye | 2·376 | 1·621 | 6·013 | 1·282 | 2·991 | 10516 | 5885 | 13392 |
| Sanliurfa | Harran | 2·913 | 2·057 | 7·635 | 1·532 | 3·677 | 2930 | 1548 | 3707 |
| Sanliurfa | Hilvan | 1·869 | 1·275 | 4·681 | 1·018 | 2·344 | 866 | 469 | 1075 |
| Sanliurfa | Karakopru | 2·121 | 1·389 | 5·180 | 1·185 | 2·655 | 6049 | 3404 | 7417 |
| Sanliurfa | Siverek | 1·831 | 1·299 | 4·802 | 0·964 | 2·306 | 4864 | 2530 | 6208 |
| Sanliurfa | Suruc | 2·540 | 1·713 | 6·371 | 1·391 | 3·187 | 3529 | 1919 | 4397 |
| Sanliurfa | Viransehir | 2·476 | 1·766 | 6·514 | 1·297 | 3·115 | 5516 | 2892 | 6935 |
| Siirt | Baykan | 2·053 | 1·702 | 6·269 | 0·956 | 2·589 | 441 | 203 | 553 |
| Siirt | Eruh | 2·600 | 2·026 | 7·368 | 1·261 | 3·293 | 486 | 235 | 616 |
| Siirt | Kurtalan | 2·259 | 1·868 | 6·798 | 1·049 | 2·866 | 1465 | 671 | 1861 |

| | | β-thalassaemia carrier prevalence | | | | | Estimated carrier counts | | |
|---|---|---|---|---|---|---|---|---|---|
| ADM1 | ADM2 | Mean | SD | CI95% | Q25 | Q75 | Mean | Q25 | Q75 |
| Siirt | Pervari | 2·635 | 2·027 | 7·428 | 1·289 | 3·343 | 792 | 382 | 1003 |
| Siirt | Siirt (merkez) | 2·331 | 1·890 | 6·730 | 1·098 | 2·961 | 4168 | 1958 | 5371 |
| Siirt | Sirvan | 2·249 | 1·832 | 6·613 | 1·054 | 2·850 | 499 | 233 | 630 |
| Siirt | Tillo | 2·352 | 1·862 | 6·787 | 1·108 | 3·014 | 93 | 44 | 121 |
| Sinop | Ayancik | 1·773 | 2·407 | 8·290 | 0·473 | 2·074 | 425 | 105 | 490 |
| Sinop | Boyabat | 1·710 | 2·249 | 7·812 | 0·472 | 2·024 | 790 | 203 | 946 |
| Sinop | Dikmen | 1·632 | 2·116 | 7·206 | 0·460 | 1·942 | 75 | 22 | 88 |
| Sinop | Duragan | 1·608 | 2·053 | 7·253 | 0·461 | 1·908 | 284 | 76 | 333 |
| Sinop | Erfelek | 1·734 | 2·356 | 8·015 | 0·457 | 2·039 | 209 | 55 | 249 |
| Sinop | Gerze | 1·684 | 2·266 | 7·655 | 0·462 | 1·977 | 135 | 38 | 162 |
| Sinop | Sarayduzu | 1·666 | 2·138 | 7·363 | 0·468 | 1·997 | 83 | 24 | 101 |
| Sinop | Sinop merkez | 1·760 | 2·455 | 8·649 | 0·465 | 2·028 | 283 | 74 | 321 |
| Sinop | Turkeli | 1·808 | 2·444 | 8·150 | 0·479 | 2·133 | 263 | 69 | 314 |
| Sirnak | Beytussebap | 3·157 | 2·294 | 8·552 | 1·605 | 4·010 | 516 | 263 | 656 |
| Sirnak | Cizre | 3·150 | 2·323 | 8·571 | 1·609 | 3·986 | 5246 | 2728 | 6584 |
| Sirnak | Guclukonak | 2·760 | 2·081 | 7·788 | 1·365 | 3·521 | 350 | 174 | 447 |
| Sirnak | Idil | 3·083 | 2·374 | 8·792 | 1·515 | 3·902 | 2457 | 1188 | 3144 |
| Sirnak | Silopi | 3·345 | 2·341 | 8·587 | 1·782 | 4·225 | 5133 | 2699 | 6425 |
| Sirnak | Sirnak merkez | 2·937 | 2·116 | 7·902 | 1·503 | 3·731 | 3066 | 1600 | 3898 |
| Sirnak | Uludere | 3·298 | 2·292 | 8·547 | 1·747 | 4·162 | 1543 | 820 | 1963 |
| Sivas | Akincilar | 1·093 | 1·178 | 4·011 | 0·399 | 1·349 | 60 | 22 | 74 |
| Sivas | Altinyayla | 1·257 | 1·208 | 4·239 | 0·524 | 1·564 | 118 | 48 | 144 |
| Sivas | Divrigi | 1·101 | 1·025 | 3·686 | 0·467 | 1·373 | 173 | 72 | 221 |
| Sivas | Dogansar | 1·220 | 1·444 | 4·836 | 0·413 | 1·473 | 31 | 9 | 40 |
| Sivas | Gemerek | 1·351 | 1·308 | 4·660 | 0·541 | 1·698 | 293 | 112 | 370 |
| Sivas | Golova | 1·075 | 1·119 | 3·948 | 0·403 | 1·333 | 35 | 13 | 43 |
| Sivas | Gurun | 1·285 | 1·045 | 3·776 | 0·615 | 1·615 | 234 | 106 | 298 |
| Sivas | Hafik | 1·207 | 1·369 | 4·563 | 0·437 | 1·473 | 125 | 37 | 153 |
| Sivas | Imranli | 1·094 | 1·153 | 3·978 | 0·415 | 1·348 | 78 | 21 | 90 |
| Sivas | Kangal | 1·189 | 1·101 | 3·836 | 0·520 | 1·477 | 224 | 88 | 280 |
| Sivas | Koyulhisar | 1·178 | 1·399 | 4·610 | 0·398 | 1·434 | 144 | 46 | 181 |
| Sivas | Sarkisla | 1·311 | 1·321 | 4·650 | 0·517 | 1·633 | 508 | 204 | 634 |
| Sivas | Sivas merkez | 1·247 | 1·382 | 4·570 | 0·465 | 1·525 | 4911 | 1813 | 6019 |
| Sivas | Susehri | 1·133 | 1·275 | 4·306 | 0·404 | 1·390 | 292 | 102 | 355 |
| Sivas | Ulas | 1·225 | 1·247 | 4·239 | 0·492 | 1·518 | 122 | 46 | 147 |
| Sivas | Yildizeli | 1·311 | 1·548 | 5·014 | 0·461 | 1·591 | 412 | 142 | 493 |
| Sivas | Zara | 1·139 | 1·207 | 4·126 | 0·434 | 1·399 | 251 | 84 | 306 |
| Tekirdag | Cerkezkoy | 2·191 | 1·851 | 6·894 | 0·966 | 2·772 | 4874 | 2137 | 6185 |
| Tekirdag | Corlu | 2·286 | 1·944 | 7·075 | 1·025 | 2·884 | 6804 | 3049 | 8786 |
| Tekirdag | Ergene | 2·416 | 2·108 | 7·549 | 1·072 | 3·037 | 1715 | 767 | 2138 |
| Tekirdag | Hayrabolu | 2·834 | 2·405 | 8·781 | 1·286 | 3·557 | 921 | 429 | 1162 |
| Tekirdag | Kapakli | 2·281 | 2·014 | 7·253 | 1·001 | 2·856 | 2955 | 1301 | 3745 |
| Tekirdag | Malkara | 2·818 | 2·239 | 8·169 | 1·352 | 3·551 | 1420 | 683 | 1790 |
| Tekirdag | Marmaraereglisi | 2·234 | 1·864 | 6·754 | 1·025 | 2·828 | 604 | 273 | 764 |
| Tekirdag | Muratli | 2·490 | 2·109 | 7·758 | 1·129 | 3·150 | 810 | 366 | 1012 |
| Tekirdag | Saray | 2·364 | 2·121 | 7·801 | 1·006 | 2·976 | 1181 | 508 | 1486 |
| Tekirdag | Sarkoy | 2·543 | 1·996 | 7·222 | 1·234 | 3·214 | 761 | 371 | 975 |
| Tekirdag | Suleymanpasa | 2·530 | 2·113 | 7·584 | 1·178 | 3·166 | 4807 | 2173 | 5995 |
| Tokat | Almus | 1·277 | 1·570 | 5·156 | 0·419 | 1·531 | 281 | 90 | 330 |
| Tokat | Artova | 1·346 | 1·600 | 5·214 | 0·451 | 1·652 | 91 | 32 | 110 |
| Tokat | Basciftlik | 1·284 | 1·567 | 5·385 | 0·406 | 1·525 | 99 | 32 | 116 |
| Tokat | Erbaa | 1·362 | 1·731 | 5·785 | 0·415 | 1·631 | 1368 | 441 | 1652 |
| Tokat | Niksar | 1·307 | 1·620 | 5·478 | 0·405 | 1·560 | 822 | 249 | 961 |
| Tokat | Pazar | 1·361 | 1·642 | 5·445 | 0·450 | 1·663 | 186 | 61 | 224 |
| Tokat | Resadiye | 1·244 | 1·497 | 5·082 | 0·407 | 1·505 | 427 | 141 | 514 |
| Tokat | Sulusaray | 1·360 | 1·509 | 5·258 | 0·472 | 1·668 | 89 | 32 | 110 |
| Tokat | Tokat merkez | 1·335 | 1·708 | 5·349 | 0·432 | 1·599 | 2835 | 906 | 3349 |

| | | β-thalassaemia carrier prevalence | | | | | Estimated carrier counts | | |
|---|---|---|---|---|---|---|---|---|---|
| ADM1 | ADM2 | Mean | SD | CI95% | Q25 | Q75 | Mean | Q25 | Q75 |
| Tokat | Turhal | 1·387 | 1·750 | 5·666 | 0·438 | 1·676 | 1101 | 350 | 1356 |
| Tokat | Yesilyurt | 1·335 | 1·615 | 5·107 | 0·457 | 1·627 | 116 | 40 | 139 |
| Tokat | Zile | 1·414 | 1·710 | 5·635 | 0·458 | 1·733 | 763 | 251 | 946 |
| Trabzon | Akcaabat | 1·074 | 1·140 | 3·914 | 0·387 | 1·339 | 450 | 166 | 556 |
| Trabzon | Arakli | 1·013 | 1·015 | 3·517 | 0·399 | 1·261 | 198 | 77 | 249 |
| Trabzon | Arsin | 1·042 | 1·067 | 3·685 | 0·396 | 1·302 | 93 | 36 | 117 |
| Trabzon | Beylikduzu | 1·131 | 1·336 | 4·682 | 0·371 | 1·374 | 266 | 87 | 324 |
| Trabzon | Carsibasi | 1·120 | 1·296 | 4·347 | 0·393 | 1·353 | 78 | 26 | 94 |
| Trabzon | Caykara | 0·979 | 0·919 | 3·285 | 0·407 | 1·228 | 132 | 56 | 163 |
| Trabzon | Dernekpazari | 1·018 | 0·973 | 3·408 | 0·403 | 1·290 | 36 | 15 | 46 |
| Trabzon | Duzkoy | 1·072 | 1·142 | 4·046 | 0·387 | 1·320 | 138 | 50 | 170 |
| Trabzon | Hayrat | 0·972 | 0·921 | 3·222 | 0·409 | 1·221 | 60 | 25 | 73 |
| Trabzon | Koprubasi | 1·010 | 1·005 | 3·533 | 0·401 | 1·259 | 39 | 16 | 49 |
| Trabzon | Macka | 1·044 | 1·066 | 3·823 | 0·394 | 1·304 | 256 | 97 | 321 |
| Trabzon | Of | 0·997 | 0·976 | 3·462 | 0·406 | 1·244 | 367 | 149 | 451 |
| Trabzon | Ortahisar | 1·055 | 1·076 | 3·874 | 0·392 | 1·313 | 3650 | 1363 | 4575 |
| Trabzon | Salpazari | 1·095 | 1·209 | 4·240 | 0·383 | 1·359 | 131 | 46 | 162 |
| Trabzon | Surmene | 1·019 | 1·005 | 3·647 | 0·405 | 1·260 | 449 | 170 | 543 |
| Trabzon | Tonya | 1·098 | 1·207 | 4·262 | 0·386 | 1·347 | 143 | 48 | 174 |
| Trabzon | Vakfikebir | 1·136 | 1·278 | 4·521 | 0·394 | 1·393 | 251 | 86 | 301 |
| Trabzon | Yomra | 1·059 | 1·090 | 3·871 | 0·400 | 1·311 | 136 | 50 | 169 |
| Tunceli | Cemisgezek | 1·090 | 0·952 | 3·435 | 0·487 | 1·366 | 83 | 36 | 103 |
| Tunceli | Hozat | 1·078 | 0·951 | 3·426 | 0·476 | 1·350 | 57 | 24 | 72 |
| Tunceli | Mazgirt | 1·152 | 0·988 | 3·558 | 0·522 | 1·455 | 70 | 33 | 88 |
| Tunceli | Nazimiye | 1·087 | 0·964 | 3·470 | 0·479 | 1·369 | 34 | 13 | 42 |
| Tunceli | Ovacik | 1·056 | 0·948 | 3·460 | 0·451 | 1·334 | 64 | 25 | 83 |
| Tunceli | Pertek | 1·132 | 0·986 | 3·529 | 0·516 | 1·409 | 119 | 59 | 151 |
| Tunceli | Pulumur | 1·040 | 0·923 | 3·336 | 0·458 | 1·303 | 35 | 13 | 45 |
| Tunceli | Tunceli merkez | 1·101 | 0·996 | 3·570 | 0·482 | 1·373 | 490 | 219 | 613 |
| Usak | Banaz | 3·100 | 2·032 | 7·614 | 1·720 | 3·896 | 1056 | 586 | 1329 |
| Usak | Esme | 3·348 | 2·112 | 7·965 | 1·909 | 4·198 | 1171 | 661 | 1469 |
| Usak | Karahalli | 3·283 | 2·127 | 7·917 | 1·838 | 4·107 | 317 | 173 | 407 |
| Usak | Sivasli | 3·226 | 2·070 | 7·681 | 1·811 | 4·050 | 636 | 363 | 796 |
| Usak | Ulubey | 3·260 | 2·064 | 7·787 | 1·853 | 4·092 | 433 | 243 | 543 |
| Usak | Usak merkez | 3·152 | 2·024 | 7·492 | 1·769 | 3·983 | 8547 | 4847 | 10802 |
| Van | Bahcesaray | 2·408 | 1·952 | 7·147 | 1·138 | 3·045 | 329 | 159 | 412 |
| Van | Baskale | 2·946 | 2·411 | 8·813 | 1·366 | 3·731 | 1345 | 617 | 1683 |
| Van | Caldiran | 1·980 | 1·841 | 6·649 | 0·804 | 2·517 | 1215 | 491 | 1539 |
| Van | Catak | 2·663 | 2·057 | 7·612 | 1·300 | 3·380 | 489 | 239 | 617 |
| Van | Edremit | 2·294 | 1·943 | 7·028 | 1·041 | 2·928 | 2453 | 1114 | 3080 |
| Van | Ercis | 1·721 | 1·536 | 5·498 | 0·744 | 2·178 | 2932 | 1292 | 3727 |
| Van | Gevas | 2·278 | 1·948 | 7·013 | 1·016 | 2·893 | 517 | 233 | 655 |
| Van | Gurpinar | 2·666 | 2·157 | 7·890 | 1·248 | 3·385 | 799 | 354 | 1008 |
| Van | Ipekyolu | 2·220 | 1·923 | 6·993 | 0·976 | 2·824 | 10712 | 4793 | 13537 |
| Van | Muradiye | 1·974 | 1·765 | 6·424 | 0·836 | 2·510 | 933 | 399 | 1188 |
| Van | Ozalp | 2·248 | 2·022 | 7·370 | 0·936 | 2·858 | 1390 | 579 | 1752 |
| Van | Saray | 2·475 | 2·211 | 7·981 | 1·053 | 3·152 | 508 | 213 | 656 |
| Van | Tusba | 2·042 | 1·772 | 6·386 | 0·893 | 2·599 | 1100 | 490 | 1384 |
| Yalova | Altinova | 1·735 | 1·380 | 5·141 | 0·831 | 2·176 | 446 | 215 | 554 |
| Yalova | Armutlu | 1·909 | 1·534 | 5·620 | 0·915 | 2·403 | 119 | 58 | 151 |
| Yalova | Ciftlikkoy | 1·747 | 1·382 | 5·032 | 0·858 | 2·193 | 446 | 215 | 566 |
| Yalova | Cinarcik | 1·843 | 1·472 | 5·262 | 0·885 | 2·325 | 421 | 203 | 536 |
| Yalova | Termal | 1·845 | 1·482 | 5·252 | 0·887 | 2·343 | 523 | 249 | 664 |
| Yalova | Yalova merkez | 1·808 | 1·444 | 5·239 | 0·875 | 2·277 | 2129 | 1018 | 2715 |
| Yozgat | Akdagmadeni | 1·380 | 1·506 | 5·182 | 0·500 | 1·706 | 573 | 207 | 712 |
| Yozgat | Aydincik | 1·495 | 1·732 | 5·852 | 0·483 | 1·836 | 186 | 58 | 232 |
| Yozgat | Bogazliyan | 1·542 | 1·552 | 5·470 | 0·591 | 1·925 | 538 | 209 | 664 |

| | | β-thalassaemia carrier prevalence | | | | | Estimated carrier counts | | |
|---|---|---|---|---|---|---|---|---|---|
| **ADM1** | **ADM2** | **Mean** | **SD** | **CI95%** | **Q25** | **Q75** | **Mean** | **Q25** | **Q75** |
| Yozgat | Candir | 1·481 | 1·515 | 5·451 | 0·564 | 1·842 | 69 | 25 | 86 |
| Yozgat | Cayiralan | 1·424 | 1·448 | 5·109 | 0·549 | 1·773 | 173 | 65 | 219 |
| Yozgat | Cekerek | 1·447 | 1·651 | 5·799 | 0·480 | 1·769 | 266 | 87 | 315 |
| Yozgat | Kadisehri | 1·394 | 1·601 | 5·295 | 0·473 | 1·708 | 144 | 51 | 172 |
| Yozgat | Saraykent | 1·447 | 1·602 | 5·615 | 0·510 | 1·788 | 185 | 65 | 226 |
| Yozgat | Sarikaya | 1·493 | 1·578 | 5·515 | 0·545 | 1·857 | 479 | 175 | 590 |
| Yozgat | Sefaatli | 1·634 | 1·717 | 6·021 | 0·584 | 2·046 | 241 | 84 | 306 |
| Yozgat | Sorgun | 1·500 | 1·650 | 5·838 | 0·515 | 1·861 | 1313 | 446 | 1672 |
| Yozgat | Yenifakili | 1·615 | 1·618 | 5·821 | 0·608 | 2·027 | 84 | 30 | 100 |
| Yozgat | Yerkoy | 1·658 | 1·743 | 6·165 | 0·588 | 2·091 | 561 | 199 | 700 |
| Yozgat | Yozgat merkez | 1·597 | 1·731 | 6·079 | 0·548 | 1·999 | 1722 | 596 | 2214 |
| Zonguldak | Alapli | 1·640 | 1·692 | 5·735 | 0·630 | 2·047 | 654 | 252 | 823 |
| Zonguldak | Caycuma | 1·744 | 2·121 | 6·643 | 0·580 | 2·151 | 1545 | 523 | 1923 |
| Zonguldak | Devrek | 1·710 | 1·948 | 6·193 | 0·615 | 2·131 | 970 | 344 | 1227 |
| Zonguldak | Eregli | 1·676 | 1·829 | 6·047 | 0·616 | 2·085 | 1423 | 538 | 1763 |
| Zonguldak | Gokcebey | 1·736 | 2·023 | 6·253 | 0·595 | 2·166 | 388 | 134 | 488 |
| Zonguldak | Kilimli | 1·759 | 2·238 | 6·937 | 0·579 | 2·143 | 423 | 138 | 510 |
| Zonguldak | Kozlu | 1·686 | 1·941 | 6·105 | 0·600 | 2·118 | 1341 | 462 | 1690 |
| Zonguldak | Zonguldak merkez | 1·726 | 2·137 | 6·608 | 0·594 | 2·105 | 1024 | 339 | 1254 |

ADM1: administrative level 1. ADM2: administrative level 2. SD: standard deviation. CI95%: 95% credible interval. Q25: 25$^{th}$ percentile. Q75: 75$^{th}$ percentile. SD and CI95% are measures of prediction uncertainty.

### 2.16. Identification of priority sites for additional carrier-prevalence surveys

Georeferenced healthcare sites represented in Healthsites.io[57] defined the candidate sites for additional β-thalassaemia carrier-prevalence surveys intended to improve geographic sampling coverage. Sites were retained when classified as hospitals and when their recorded specialty did not indicate services considered outside the scope of general population carrier sampling; records whose specialty could not be classified reliably were excluded. Duplicate sites and sites outside the prediction domain were then removed, and each remaining site was assigned to its corresponding ADM0 unit. Sites within 50 km geodesic distance of any modelling coordinate in the covariate-unrestricted dataset were excluded to direct additional sampling towards gaps in geographic sampling coverage. For each site, the corresponding 2026 WorldPop population count[26], posterior mean predicted carrier prevalence, and prediction uncertainty, quantified as the width of the 95% credible interval, were extracted. Sites were excluded when they occurred in an unpopulated cell or lacked valid predicted carrier prevalence or prediction uncertainty values. Candidate-filtering thresholds were explored using the 25$^{th}$ percentile, median, and 75$^{th}$ percentile of each country-specific distribution of prediction uncertainty, population count, and predicted carrier prevalence. Combining these thresholds across the three quantities produced 27 candidate-filtering scenarios, which were examined to assess how alternative threshold combinations affected the size and geographic distribution of the candidate pool. Country-specific distributions are shown in Supplementary Figures S25–S27. Based on these comparisons, the selected thresholds were applied only in countries with more than 100 eligible candidates. In these countries, sites were retained if prediction uncertainty exceeded the country-specific 75$^{th}$ percentile, population count was at or above the 25$^{th}$ percentile, and predicted carrier prevalence was at or above the median. In countries with 100 or fewer eligible candidates, no threshold filtering was applied, and all otherwise eligible sites were retained for the subsequent space-filling design. The geographic distribution of candidate sites before and after proximity exclusion and threshold filtering is shown in Supplementary Figure S28.

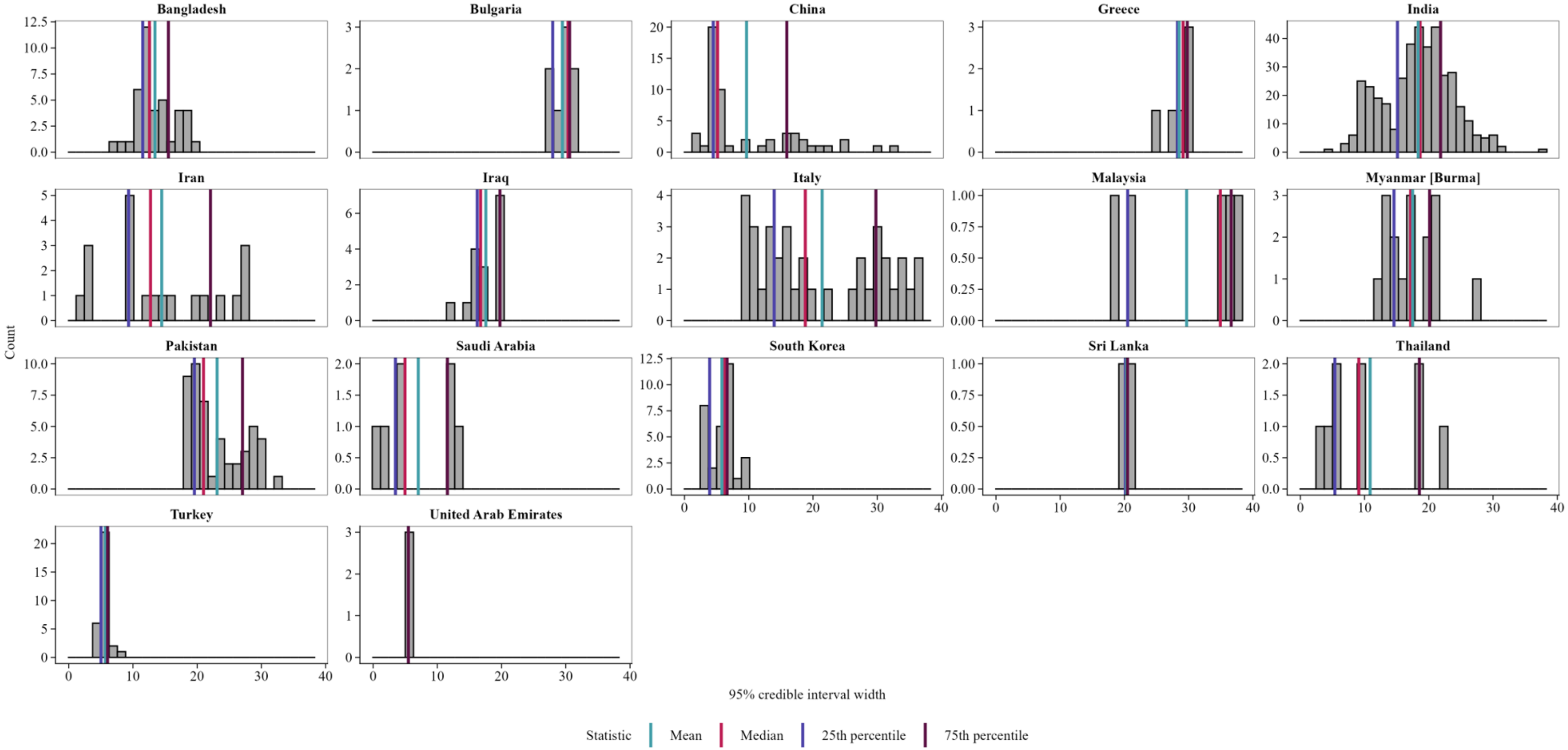

**Supplementary Figure S25. Country-level distributions of prediction uncertainty at candidate sites.**
Histograms show prediction uncertainty, quantified as the width of the 95% credible interval, at eligible candidate sites in the 17 countries with candidate sites remaining after exclusion of sites within 50 km of an existing modelling coordinate. Distributions are shown before application of the country-specific selection thresholds. Vertical lines indicate the mean, median, and lower and upper quartiles.

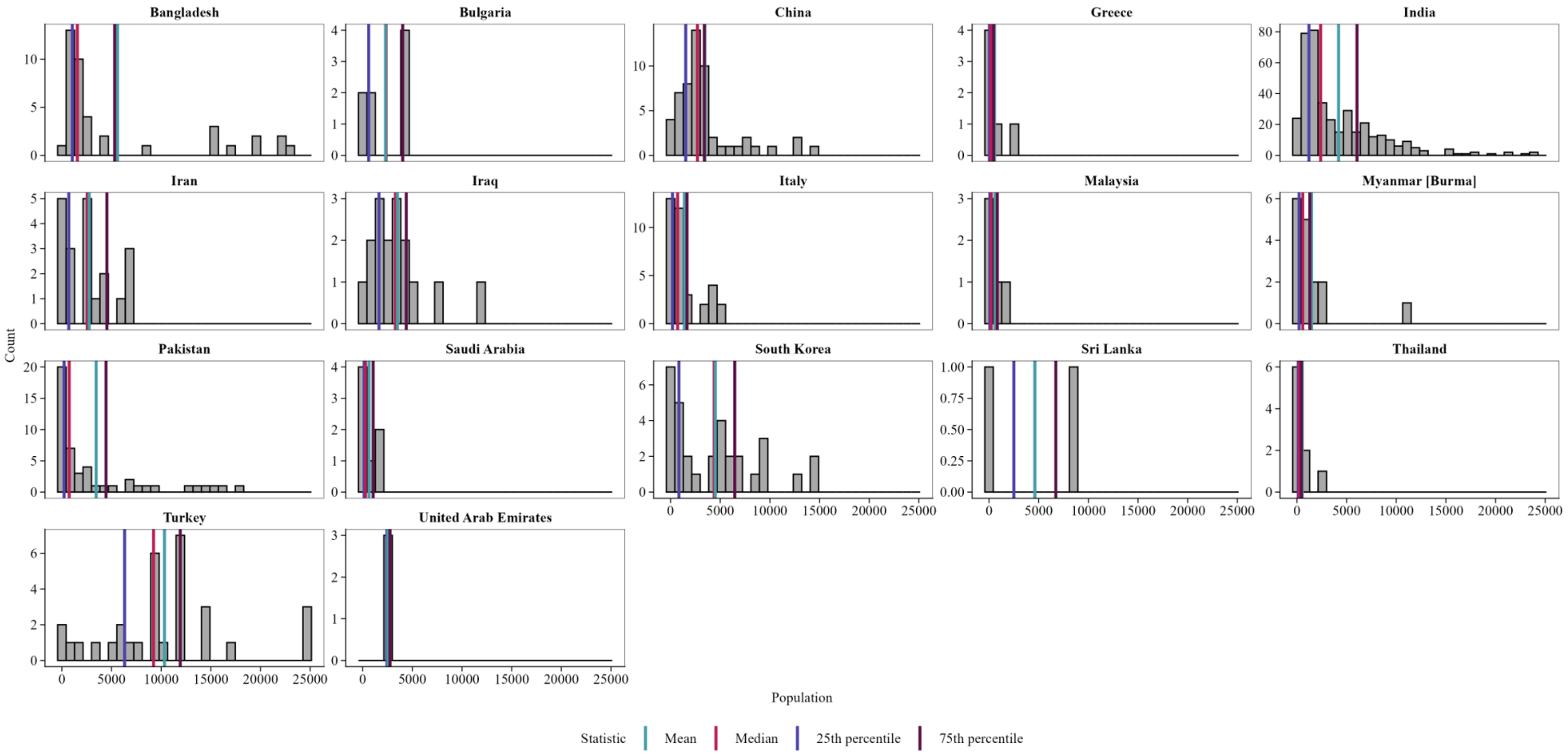

**Supplementary Figure S26. Country-specific distribution of population count across candidate study locations at candidate sites.**
Histograms show 2026 WorldPop population counts at eligible candidate sites in the 17 countries with candidate sites remaining after exclusion of sites within 50 km of an existing modelling coordinate.[26] Distributions are shown before application of the country-specific selection thresholds. Vertical lines indicate the mean, median, and lower and upper quartiles.

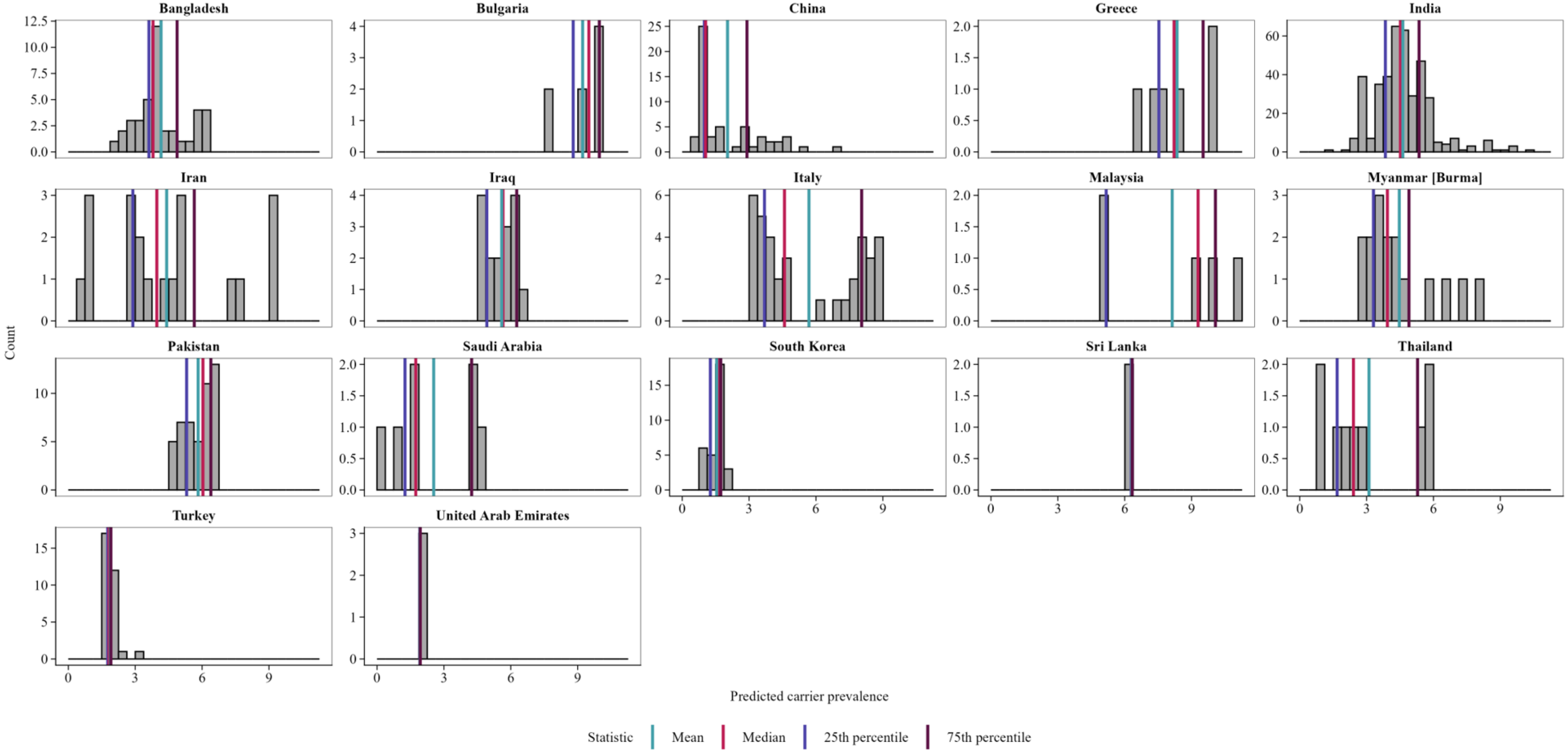

**Supplementary Figure S27. Country-level distributions of predicted β-thalassaemia carrier prevalence at candidate sites.** Histograms show posterior mean predicted β-thalassaemia carrier prevalence at eligible candidate sites in the 17 countries with candidate sites remaining after exclusion of sites within 50 km of an existing modelling coordinate. Distributions are shown before application of the country-specific selection thresholds. Vertical lines indicate the mean, median, and lower and upper quartiles.

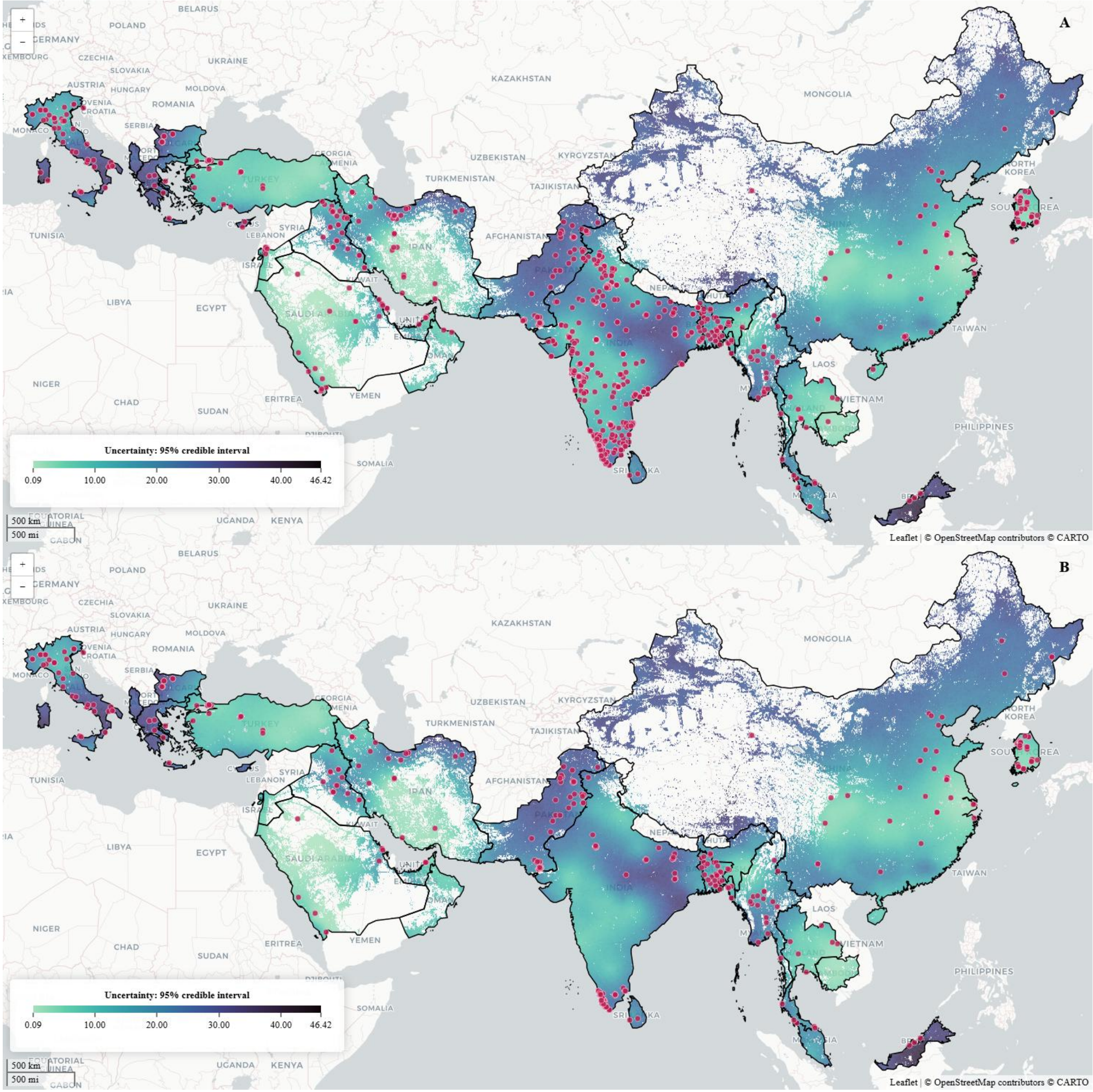

**Supplementary Figure S28. Candidate sites for future β-thalassaemia epidemiological surveys.**
Maps show candidate sites overlaid on prediction uncertainty, quantified as the width of the 95% credible interval. A. Candidate sites before proximity exclusion and threshold filtering. B. Sites retained after excluding candidates within 50 km of a modelling coordinate in the covariate-unrestricted dataset and applying the selected country-specific thresholds for prediction uncertainty, population count, and posterior mean predicted carrier prevalence. Thresholds were applied only in countries with more than 100 candidates remaining after proximity exclusion; all otherwise eligible sites were retained in countries with 100 or fewer candidates. Darker shading indicates greater prediction uncertainty.

An iterative space-filling coverage design was applied separately within each country.[58] Within each design iteration, the existing modelling coordinates and remaining candidate sites were transformed to a locally centred azimuthal equidistant projection. Existing record coordinates were retained as fixed design points, and additional sites were proposed from the eligible candidate set by optimising spatial coverage using the full candidate set.

Proposed sites were evaluated sequentially and accepted only if they were at least 50 km geodesic distance from all existing modelling coordinates and previously accepted sites. Remaining candidates within 50 km of an accepted site were removed, and the coverage design was repeated until 30 sites had been selected or no eligible candidates remained.

The resulting sites therefore represent priority sites for additional carrier-prevalence surveys within the Healthsites.io[57] candidate set, targeting gaps in geographic sampling coverage and, where threshold filtering was applied, sites with comparatively high prediction uncertainty and predicted carrier prevalence. Site-level feasibility and operational capacity to undertake carrier-prevalence surveys would require separate assessment.

The final space-filling design selected 183 priority healthcare sites across 17 countries (Supplementary Figure S28; Supplementary Table S38): China (26 sites), Pakistan (21), Italy (20), Bangladesh (19), India (17), Myanmar (14), Iran (13), Iraq (9), South Korea (8), Thailand (8), Greece (6), Saudi Arabia (6), Turkey (6), Malaysia (4), Bulgaria (3), Sri Lanka (2), and the United Arab Emirates (1).

**Supplementary Table S38. Priority healthcare sites proposed for future β-thalassaemia epidemiological surveys.**

| | Administrative level 0 | Administrative level 1 | Administrative level 2 | Latitude | Longitude |
|---|---|---|---|---|---|
| 1 | United Arab Emirates | Dubai | NA | 25·1792 | 55·2473 |
| 2 | Bangladesh | Khulna | Chuadanga | 23·7885 | 88·9029 |
| 3 | Bangladesh | Khulna | Jessore | 23·2188 | 89·3445 |
| 4 | Bangladesh | Rajshani | Bogra | 25·0055 | 89·3166 |
| 5 | Bangladesh | Rajshani | Rajshahi | 24·5752 | 88·5750 |
| 6 | Bangladesh | Chittagong | Lakshmipur | 23·0321 | 90·7654 |
| 7 | Bangladesh | Khulna | Khulna | 22·7978 | 89·6412 |
| 8 | Bangladesh | Rangpur | Nilphamari | 25·7836 | 88·8954 |
| 9 | Bangladesh | Dhaka | Madaripur | 23·1696 | 90·2030 |
| 10 | Bangladesh | Dhaka | Faridpur | 23·5881 | 89·8353 |
| 11 | Bangladesh | Mymensingh | Sherpur | 25·1825 | 90·0660 |
| 12 | Bangladesh | Sylhet | Maulvibazar | 24·3443 | 91·9774 |
| 13 | Bangladesh | Mymensingh | Mymensingh | 24·7372 | 90·5081 |
| 14 | Bangladesh | Chittagong | Rangamati | 22·5028 | 92·2095 |
| 15 | Bangladesh | Dhaka | Narsingdi | 24·0372 | 90·7326 |
| 16 | Bangladesh | Rangpur | Kurigram | 25·9648 | 89·6895 |
| 17 | Bangladesh | Chittagong | Cox's Bazar | 21·1820 | 92·1536 |
| 18 | Bangladesh | Dhaka | Narayanganj | 23·5724 | 90·4927 |
| 19 | Bangladesh | Barisal | Bhola | 22·1842 | 90·7635 |
| 20 | Bangladesh | Barisal | Jhalokati | 22·6341 | 90·2699 |
| 21 | Bulgaria | Sofia City | Stolichna | 42·6644 | 23·3443 |
| 22 | Bulgaria | Vratsa | Vratsa | 43·1937 | 23·5700 |
| 23 | Bulgaria | Pleven | Pleven | 43·4099 | 24·6082 |
| 24 | China | Qinghai Province | Delinghashi | 38·2151 | 94·6316 |
| 25 | China | Yunnan Province | Kunmingshi | 25·0307 | 102·6952 |
| 26 | China | Jilin Province | Changchunshi | 43·8506 | 125·2791 |
| 27 | China | Henan Province | Gushi xian | 32·1799 | 115·6592 |
| 28 | China | Henan Province | Nanyangshi | 33·0387 | 112·5620 |
| 29 | China | Zhejiang Province | Ying xian | 29·8838 | 121·5460 |
| 30 | China | Shandong Province | Linqingshi | 36·8427 | 115·6930 |
| 31 | China | Tianjin Municipality | Yutian xian | 39·8988 | 117·7199 |
| 32 | China | Beijing Municipality | Changpingxin | 40·2510 | 116·1391 |
| 33 | China | Guangxi Zhuang Autonomous Region | Jinxiuyao zuzizhi xian | 24·1407 | 110·1830 |
| 34 | China | Jiangsu Province | Shuqianshi | 33·9765 | 118·2651 |
| 35 | China | Shandong Province | Yutai xian | 34·9365 | 116·7538 |
| 36 | China | Jiangxi Province | Jishui xian | 27·2177 | 115·1307 |
| 37 | China | Yunnan Province | Longchuan xian | 24·1738 | 97·7973 |
| 38 | China | Sichuan Province | Wangchang xian | 32·2306 | 106·2830 |
| 39 | China | Heilongjiang Province | Jixishi | 45·2918 | 130·9572 |
| 40 | China | Sichuan Province | Leshanshi | 29·4181 | 103·5480 |
| 41 | China | Hunan Province | Shimen xian | 29·5860 | 111·3638 |
| 42 | China | Tianjin Municipality | Tianjinshi | 39·1456 | 117·2350 |
| 43 | China | Shandong Province | Jinanshi | 36·6488 | 116·9866 |
| 44 | China | Shanghai Province | Yangpu | 31·3232 | 121·5337 |
| 45 | China | Sichuan Province | Beichuan xian | 32·0164 | 104·2038 |
| 46 | China | Jiangsu Province | Nanjingshi | 32·0525 | 118·7740 |
| 47 | China | Henan Province | Puyangshi | 35·7771 | 114·9938 |
| 48 | China | Anhui Province | Anqingshi | 30·5164 | 117·0770 |

| | Administrative level 0 | Administrative level 1 | Administrative level 2 | Latitude | Longitude |
|---|---|---|---|---|---|
| 49 | China | Heilongjiang Province | Daqingshi | 46·6381 | 124·8625 |
| 50 | Greece | Thessalia-Central Greece | Thessalias | 39·1176 | 23·7276 |
| 51 | Greece | Thessalia-Central Greece | Thessalias | 39·5579 | 21·7609 |
| 52 | Greece | Attica | Attikis | 37·9678 | 23·4791 |
| 53 | Greece | Crete | Kritis | 35·5348 | 24·1579 |
| 54 | Greece | Thessalia-Central Greece | Thessalias | 39·6108 | 22·4298 |
| 55 | Greece | Thessalia-Central Greece | Stereas Elladas | 38·6917 | 22·4179 |
| 56 | India | Jammu and Kashmir | Srinagar | 34·0689 | 74·8440 |
| 57 | India | Kerala | Thrissur | 10·2591 | 76·3278 |
| 58 | India | Uttar Pradesh | Gorakhpur | 26·7311 | 83·3688 |
| 59 | India | Tamil Nadu | Kanniyakumari | 8·1868 | 77·4353 |
| 60 | India | Rajasthan | Jaipur | 26·9373 | 75·7896 |
| 61 | India | Uttar Pradesh | Allahabad | 25·4528 | 81·8382 |
| 62 | India | Tamil Nadu | Thanjavur | 10·4247 | 79·3190 |
| 63 | India | Rajasthan | Jhunjhunun | 28·1263 | 75·3998 |
| 64 | India | Bihar | Patna | 25·5842 | 85·1630 |
| 65 | India | Jharkhand | Hazaribagh | 23·9809 | 85·3670 |
| 66 | India | Kerala | Thiruvananthapuram | 8·5422 | 76·9666 |
| 67 | India | Kerala | Alappuzha | 9·2876 | 76·5246 |
| 68 | India | Jharkhand | Ranchi | 23·3352 | 85·3188 |
| 69 | India | Madhya Pradesh | Damoh | 23·8365 | 79·4353 |
| 70 | India | Kerala | Kollam | 9·0212 | 76·9199 |
| 71 | India | Bihar | Muzaffarpur | 26·1215 | 85·3543 |
| 72 | India | Tamil Nadu | Madurai | 9·9396 | 78·1279 |
| 73 | Iraq | Salah al-Din | Tooz Khurmato | 34·8840 | 44·6356 |
| 74 | Iraq | Ninawa | Aqra | 36·7298 | 43·8717 |
| 75 | Iraq | Baghdad | Al-Karkh | 33·2930 | 44·4063 |
| 76 | Iraq | Maysan | Al-Kahla | 31·8398 | 47·1500 |
| 77 | Iraq | Karbala | Kerbela | 32·6254 | 44·0341 |
| 78 | Iraq | Wasit | Al-Kut | 32·4997 | 45·8215 |
| 79 | Iraq | Salah al-Din | Tikrit | 34·6010 | 43·6788 |
| 80 | Iraq | Diyala | Khanaqin | 34·0974 | 45·0587 |
| 81 | Iraq | Al-Anbar | Heet | 33·6371 | 42·8261 |
| 82 | Iran | Mazandaran | Sari | 36·5446 | 53·0681 |
| 83 | Iran | East Azerbaijan | Tabriz | 37·9978 | 46·4710 |
| 84 | Iran | Lorestan | Khoramabad | 33·4917 | 48·3760 |
| 85 | Iran | Kurdistan | Sanandaj | 35·2763 | 47·1210 |
| 86 | Iran | Isfahan | Kashan | 33·9613 | 51·4210 |
| 87 | Iran | Kerman | Baft | 28·8226 | 56·3467 |
| 88 | Iran | Alborz | Savojbolagh | 35·9852 | 50·7412 |
| 89 | Iran | Zanjan | Zanjan | 36·6938 | 48·4958 |
| 90 | Iran | Bushehr | Jam | 27·8260 | 52·3560 |
| 91 | Iran | Mazandaran | Amol | 35·8511 | 52·1717 |
| 92 | Iran | Gilan | Rezvanshahr | 37·4931 | 49·1104 |
| 93 | Iran | Razavi Khorasan | Neyshabur | 36·2066 | 58·7945 |
| 94 | Iran | Razavi Khorasan | Mashhad | 36·2853 | 59·5699 |
| 95 | Italy | Toscana | Grosseto | 42·6872 | 11·3324 |
| 96 | Italy | Toscana | Siena | 43·3601 | 11·3127 |
| 97 | Italy | Lazio | Roma | 41·7752 | 12·9210 |
| 98 | Italy | Piemonte | Novara | 45·4363 | 8·6280 |
| 99 | Italy | Puglia | Bari | 40·8232 | 17·1088 |
| 100 | Italy | Emilia-Romagna | Piacenza | 45·0515 | 9·6851 |
| 101 | Italy | Veneto | Vicenza | 45·7107 | 11·4703 |
| 102 | Italy | Emilia-Romagna | Piacenza | 44·6230 | 9·3316 |
| 103 | Italy | Piemonte | Torino | 45·0793 | 7·6043 |
| 104 | Italy | Campania | Avellino | 41·0674 | 15·0586 |
| 105 | Italy | Campania | Salerno | 40·4779 | 15·1158 |
| 106 | Italy | Campania | Caserta | 41·0703 | 14·3495 |

| | Administrative level 0 | Administrative level 1 | Administrative level 2 | Latitude | Longitude |
|---|---|---|---|---|---|
| 107 | Italy | Veneto | Rovigo | 45·0713 | 11·8162 |
| 108 | Italy | Sicilia | Palermo | 38·0924 | 13·5133 |
| 109 | Italy | Friuli Venezia Giulia | Trieste | 45·6358 | 13·7850 |
| 110 | Italy | Toscana | Pistoia | 43·8681 | 10·8108 |
| 111 | Italy | Friuli Venezia Giulia | Pordenone | 45·9685 | 12·6527 |
| 112 | Italy | Marche | Pesaro e Urbino | 43·8161 | 12·5894 |
| 113 | Italy | Calabria | Reggio di Calabria | 38·4747 | 16·4534 |
| 114 | Italy | Emilia-Romagna | Parma | 44·8020 | 10·3080 |
| 115 | South Korea | North Gyeongsang | Dalseong-gun | 35·6911 | 128·4535 |
| 116 | South Korea | North Chungcheong | Chungju-si | 36·8470 | 127·9918 |
| 117 | South Korea | Seoul | Seongbuk-gu | 37·5866 | 127·0261 |
| 118 | South Korea | South Jeolla | Jangseong-gun | 35·3737 | 126·8074 |
| 119 | South Korea | North Gyeongsang | Gyeongju-si | 35·8428 | 129·2067 |
| 120 | South Korea | Gangwon | Hwacheon-gun | 38·1073 | 127·7091 |
| 121 | South Korea | South Jeolla | Suncheon-si | 34·9470 | 127·5340 |
| 122 | South Korea | Gyeonggi | Pyeongtaek-si | 37·0084 | 127·0743 |
| 123 | Sri Lanka | Central Province | Nuwara Eliya District | 7·0835 | 80·8939 |
| 124 | Sri Lanka | Western Province | Colombo District | 6·9001 | 79·8758 |
| 125 | Myanmar [Burma] | Kachin | Myitkyina | 25·3954 | 97·3951 |
| 126 | Myanmar [Burma] | Magway | Pakokku | 21·3867 | 94·8256 |
| 127 | Myanmar [Burma] | Shan | Langkho | 20·2293 | 97·7246 |
| 128 | Myanmar [Burma] | Mandalay | Mandalay | 21·9749 | 96·1213 |
| 129 | Myanmar [Burma] | Bago | Taungoo | 18·9456 | 96·4347 |
| 130 | Myanmar [Burma] | Mandalay | Meiktila | 21·0941 | 96·0332 |
| 131 | Myanmar [Burma] | Mandalay | Myingyan | 21·4749 | 95·3893 |
| 132 | Myanmar [Burma] | Ayeyarwady | Pyapon | 16·1412 | 95·4830 |
| 133 | Myanmar [Burma] | Tanitharyi | Dawei | 14·2458 | 98·1959 |
| 134 | Myanmar [Burma] | Magway | Minbu | 20·5737 | 94·6597 |
| 135 | Myanmar [Burma] | Shan | Taunggyi | 20·7977 | 97·1776 |
| 136 | Myanmar [Burma] | Magway | Magway | 20·3632 | 95·3994 |
| 137 | Myanmar [Burma] | Bago | Taungoo | 18·4074 | 96·4592 |
| 138 | Myanmar [Burma] | Bago | Bago | 17·1447 | 96·5523 |
| 139 | Malaysia | Kelantan | Tumpat | 6·1125 | 102·2136 |
| 140 | Malaysia | Sarawak | Miri | 3·8631 | 113·7153 |
| 141 | Malaysia | Sarawak | Limbang | 4·7646 | 115·0188 |
| 142 | Malaysia | Sarawak | Marudi | 4·1784 | 114·3259 |
| 143 | Pakistan | Khyber Pakhtunkhwa | Battagram | 34·7915 | 72·9303 |
| 144 | Pakistan | Punjab | Lahore | 31·4974 | 74·2634 |
| 145 | Pakistan | Punjab | Rajanpur | 29·6431 | 70·5905 |
| 146 | Pakistan | Punjab | Faisalabad | 31·4227 | 73·0866 |
| 147 | Pakistan | Khyber Pakhtunkhwa | Orakzai | 33·6729 | 71·5436 |
| 148 | Pakistan | Punjab | Muzaffargarh | 30·1896 | 70·9547 |
| 149 | Pakistan | Sindh | Qambar Shahdadkot | 27·7710 | 68·0001 |
| 150 | Pakistan | Khyber Pakhtunkhwa | Mohmand | 34·3247 | 71·5913 |
| 151 | Pakistan | Sindh | Badin | 25·1471 | 68·9512 |
| 152 | Pakistan | Punjab | Sargodha | 32·0802 | 72·6936 |
| 153 | Pakistan | Gilgit-Baltistan | Kharmang | 35·0917 | 76·0037 |
| 154 | Pakistan | Sindh | Badin | 24·4890 | 69·0832 |
| 155 | Pakistan | Punjab | Gujranwala | 32·4105 | 74·1229 |
| 156 | Pakistan | Islamabad Capital Territory | Islamabad Capital Territory | 33·6300 | 73·1189 |
| 157 | Pakistan | Punjab | Toba Tek Singh | 30·9697 | 72·4847 |
| 158 | Pakistan | Sindh | Hyderabad | 25·3726 | 68·3479 |
| 159 | Pakistan | Punjab | Mandi Bahauddin | 32·4211 | 73·2207 |
| 160 | Pakistan | Punjab | Multan | 30·2463 | 71·4855 |
| 161 | Pakistan | Khyber Pakhtunkhwa | Dera Ismail Khan | 31·8271 | 70·9200 |
| 162 | Pakistan | Punjab | Rahim Yar Khan | 28·4367 | 70·3052 |
| 163 | Pakistan | Khyber Pakhtunkhwa | Upper Dir | 35·2025 | 71·8788 |
| 164 | Saudi Arabia | Eastern Region | Al Ahsa Governorate | 25·3584 | 49·5603 |

| | Administrative level 0 | Administrative level 1 | Administrative level 2 | Latitude | Longitude |
|---|---|---|---|---|---|
| 165 | Saudi Arabia | Jazan Region | Ad Dair | 17·3308 | 43·1159 |
| 166 | Saudi Arabia | Eastern Region | Al Qatif Governorate | 26·6440 | 49·9620 |
| 167 | Saudi Arabia | Al Jawf Region | Dawamat Al Jandal | 29·8030 | 39·6879 |
| 168 | Saudi Arabia | Makkah Region | Makkah Al Mukarramah | 21·2824 | 39·8244 |
| 169 | Saudi Arabia | Makkah Region | Al Ardiyat | 19·4471 | 41·8379 |
| 170 | Thailand | Satun Province | Mueang Satun | 6·6181 | 100·0707 |
| 171 | Thailand | Phangnga Province | Takua Pa | 8·8638 | 98·3341 |
| 172 | Thailand | Nong Khai Province | Phon Phisai | 18·0493 | 103·0870 |
| 173 | Thailand | Ang Thong Province | Sawaeng Ha | 14·7454 | 100·3240 |
| 174 | Thailand | Ubon Ratchathani Province | Khemarat | 15·9774 | 105·0335 |
| 175 | Thailand | Rayong Province | Mueang Rayong | 12·6768 | 101·2334 |
| 176 | Thailand | Yasothon Province | Thai Charoen | 16·1147 | 104·4044 |
| 177 | Thailand | Kamphaeng Phet Province | Khlong Lan | 16·1833 | 99·3248 |
| 178 | Turkey | Istanbul | Esenler | 41·0488 | 28·8595 |
| 179 | Turkey | Ankara | Yenimahalle | 39·9659 | 32·8315 |
| 180 | Turkey | Kayseri | Develi | 38·3901 | 35·4893 |
| 181 | Turkey | Tekirdag | Suleymanpasa | 41·0084 | 27·5245 |
| 182 | Turkey | Bursa | Nilufer | 40·2327 | 28·9802 |
| 183 | Turkey | Balikesir | Edremit | 39·5765 | 27·0430 |

NA: not applicable.

**Supplementary material references**

1 Runfola D, Anderson A, Baier H, *et al.* geoBoundaries: A global database of political administrative boundaries. *PLoS ONE* 2020; 15: e0231866.

2 Kountouris P, Lederer CW, Fanis P, Feleki X, Old J, Kleanthous M. IthaGenes: An Interactive Database for Haemoglobin Variations and Epidemiology. *PLoS ONE* 2014; 9: e103020.

3 Harzing AW. Publish or Perish. 2007. https://harzing.com/resources/publish-or-perish (accessed Sept 1, 2023).

4 Vasilevsky NA, Toro S, Matentzoglu N, *et al.* Mondo: integrating disease terminology across communities. *GENETICS* 2026; 232: iyaf215.

5 Sioutos N, Coronado SD, Haber MW, Hartel FW, Shaiu W-L, Wright LW. NCI Thesaurus: A semantic model integrating cancer-related clinical and molecular information. *Journal of Biomedical Informatics* 2007; 40: 30–43.

6 Sickle Cell Disease Ontology Working Group, Adekile A, Anie KA, *et al.* The Sickle Cell Disease Ontology: enabling universal sickle cell-based knowledge representation. *Database* 2019; 2019: baz118.

7 Hardy GH. Mendelian Proportions in a Mixed Population. *Science* 1908; 28: 49–50.

8 Modell B, Darlison M. Global epidemiology of haemoglobin disorders and derived service indicators. *Bull World Health Organ* 2008; 86: 480–7.

9 Munn Z, Moola S, Riitano D, Lisy K. The development of a critical appraisal tool for use in systematic reviews addressing questions of prevalence. *Int J Health Policy Manag* 2014; 3: 123–8.

10 Rao P, Raj EA, Natesan S, Gudi N. Prevalence of Sickle cell disease, Sickle cell trait and HBS-beta-thalassemia in India: A systematic review and Meta-analysis. *Clinical Epidemiology and Global Health* 2024; 28: 101678.

11 Barendregt JJ, Doi SA, Lee YY, Norman RE, Vos T. Meta-analysis of prevalence. *J Epidemiol Community Health* 2013; 67: 974–8.

12 Viechtbauer W. Conducting Meta-Analyses in *R* with the metafor Package. *J Stat Soft* 2010; 36. DOI:10.18637/jss.v036.i03.

13 Nunes MA. Building Interactive Visualizations with Shiny. In: Awe OO, A. Vance E, eds. Practical Statistical Learning and Data Science Methods. Cham: Springer Nature Switzerland, 2025: 313–24.

14 Piel FB, Patil AP, Howes RE, *et al.* Global distribution of the sickle cell gene and geographical confirmation of the malaria hypothesis. *Nat Commun* 2010; 1: 104.

15 Roberts DJ, Williams TN. Haemoglobinopathies and resistance to malaria. *Redox Report* 2003; 8: 304–10.

16 Guerra CA, Hay SI, Lucioparedes LS, *et al.* Assembling a global database of malaria parasite prevalence for the Malaria Atlas Project. *Malar J* 2007; 6: 17.

17 Hamamy H, Antonarakis SE, Cavalli-Sforza LL, *et al.* Consanguineous marriages, pearls and perils: Geneva International Consanguinity Workshop Report. *Genetics in Medicine* 2011; 13: 841–7.

18 Bittles AH, Black ML. Evolution in health and medicine Sackler colloquium: Consanguinity, human evolution, and complex diseases. *Proc Natl Acad Sci U S A* 2010; 107 Suppl 1: 1779–86.

19 Angastiniotis M, Petrou M, Loukopoulos D, *et al.* The Prevention of Thalassemia Revisited: A Historical and Ethical Perspective by the Thalassemia International Federation. *Hemoglobin* 2021; 45: 5–12.

20 Christianson A, Modell B. MEDICAL GENETICS IN DEVELOPING COUNTRIES. *Annu Rev Genom Hum Genet* 2004; 5: 219–65.

21 Drazanova L. Historical index of ethnic fractionalisation dataset. 2019.

22 Perez-Haydrich C, Warren JL, Burgert CR, Emch ME. Guidelines on the use of DHS GPS data. Calverton, Maryland, USA: ICF International, 2013 http://dhsprogram.com/pubs/pdf/SAR8/SAR8.pdf.

23 ICF. Demographic and Health Surveys (various). 1994; published online 2024. https://dhsprogram.com/.

24 Klarsfeld A, Syed J, Mumtaz A. An analysis of Religious Diversity Index (RDI) by Pew Research Center. In: Ng ES, Stamper CL, Klarsfeld A, Han YJ, eds. Handbook on Diversity and Inclusion Indices. Edward Elgar Publishing, 2021. DOI:10.4337/9781788975728.00027.

25 Campbell H, Rudan I, Bittles AH, Wright AF. Human population structure, genome autozygosity and human health. *Genome Med* 2009; 1: 91.

26 Bondarenko M, Priyatikanto R, Tejedor Garavito N, *et al.* The spatial distribution of population in 2015-2030 at a resolution of 30 arc (approximately 1km at the equator) R2025A version v1. 2025. DOI:10.5258/SOTON/WP00845.

27 Telfair J, Haque A, Etienne M, Tang S, Strasser S. Rural/urban differences in access to and utilization of services among people in Alabama with sickle cell disease. *Public Health Reports* 2003; 118: 27–36.

28 Alam NE, Islam MS, Khabir MIU, *et al.* The scenario of knowledge, attitude and practice of the Bangladeshi population towards thalassemia prevention: A nationwide study. *PLOS Glob Public Health* 2022; 2: e0001177.

29 Bondarenko M, Zhang W, Tejedor Garavito N, Nosatiuk B, Tatem A, Sorichetta A. Degree of Urbanisation classes using WorldPop Global2 R2025A population data with the GHSL GHS-DUG tool. 2025. DOI:10.5258/SOTON/WP00879.

30 Cao A, Kan YW. The prevention of thalassemia. *Cold Spring Harb Perspect Med* 2013; 3: a011775.

31 Cousens NE, Gaff CL, Metcalfe SA, Delatycki MB. Carrier screening for Beta-thalassaemia: a review of international practice. *Eur J Hum Genet* 2010; 18: 1077–83.
32 Samavat A, Modell B. Iranian national thalassaemia screening programme. *BMJ* 2004; 329: 1134–7.
33 Cao A, Rosatelli C, Galanello R, *et al.* The prevention of thalassemia in Sardinia. *Clin Genet* 1989; 36: 277–85.
34 Utazi CE, Aheto JMK, Chan HMT, Tatem AJ, Sahu SK. Conditional probability and ratio-based approaches for mapping the coverage of multi-dose vaccines. *Statistics in Medicine* 2022; 41: 5662–78.
35 Utazi CE, Thorley J, Alegana VA, *et al.* High resolution age-structured mapping of childhood vaccination coverage in low and middle income countries. *Vaccine* 2018; 36: 1583–91.
36 Fox J, Monette G. Generalized Collinearity Diagnostics. *Journal of the American Statistical Association* 1992; 87: 178–83.
37 Hocking RR. A Biometrics Invited Paper. The Analysis and Selection of Variables in Linear Regression. *Biometrics* 1976; 32: 1.
38 Mosser JF, Gagne-Maynard W, Rao PC, *et al.* Mapping diphtheria-pertussis-tetanus vaccine coverage in Africa, 2000–2016: a spatial and temporal modelling study. *The Lancet* 2019; 393: 1843–55.
39 Local Burden of Disease Vaccine Coverage Collaborators, Sbarra AN, Rolfe S, *et al.* Mapping routine measles vaccination in low- and middle-income countries. *Nature* 2021; 589: 415–9.
40 Ramaswamy S, Rastogi R, Shim K. Efficient algorithms for mining outliers from large data sets. *SIGMOD Rec* 2000; 29: 427–38.
41 Conn PB, Johnson DS, Boveng PL. On Extrapolating Past the Range of Observed Data When Making Statistical Predictions in Ecology. *PLoS ONE* 2015; 10: e0141416.
42 Diggle PJ, Giorgi E. Model-Based Geostatistics for Prevalence Mapping in Low-Resource Settings. *Journal of the American Statistical Association* 2016; 111: 1096–120.
43 Diggle PJ, Tawn JA, Moyeed RA. Model-Based Geostatistics. *Journal of the Royal Statistical Society Series C: Applied Statistics* 1998; 47: 299–350.
44 Lindgren F, Rue H, Lindström J. An Explicit Link between Gaussian Fields and Gaussian Markov Random Fields: The Stochastic Partial Differential Equation Approach. *Journal of the Royal Statistical Society Series B: Statistical Methodology* 2011; 73: 423–98.
45 Lindgren F, Rue H. Bayesian Spatial Modelling with *R* - INLA. *J Stat Soft* 2015; 63. DOI:10.18637/jss.v063.i19.
46 Fuglstad G-A, Simpson D, Lindgren F, Rue H. Constructing Priors that Penalize the Complexity of Gaussian Random Fields. *Journal of the American Statistical Association* 2019; 114: 445–52.
47 Rue H, Martino S, Chopin N. Approximate Bayesian Inference for Latent Gaussian models by using Integrated Nested Laplace Approximations. *Journal of the Royal Statistical Society Series B: Statistical Methodology* 2009; 71: 319–92.
48 Chiuchiolo C, Van Niekerk J, Rue H. Joint posterior inference for latent Gaussian models with R-INLA. *Journal of Statistical Computation and Simulation* 2023; 93: 723–52.
49 Vehtari A, Gelman A, Gabry J. Practical Bayesian model evaluation using leave-one-out cross-validation and WAIC. *Stat Comput* 2017; 27: 1413–32.
50 Gelfand AE, Dey DK, Chang H. Model Determination using Predictive Distributions with Implementation via Sampling-Based Methods. In: Bernardo JM, Berger JO, Dawid P, Smith AFM, eds. Bayesian Statistics 4. Oxford University PressOxford, 1992: 147–67.
51 Saha PK, Logofatu D. Efficient Approaches for Density-Based Spatial Clustering of Applications with Noise. In: Maglogiannis I, Macintyre J, Iliadis L, eds. Artificial Intelligence Applications and Innovations. Cham: Springer International Publishing, 2021: 184–95.
52 Roberts DR, Bahn V, Ciuti S, *et al.* Cross-validation strategies for data with temporal, spatial, hierarchical, or phylogenetic structure. *Ecography* 2017; 40: 913–29.
53 Willmott C, Matsuura K. Advantages of the mean absolute error (MAE) over the root mean square error (RMSE) in assessing average model performance. *Clim Res* 2005; 30: 79–82.
54 Modell B, Darlison M. Global epidemiology of haemoglobin disorders and derived service indicators. *Bull World Health Organ* 2008; 86: 480–7.
55 Piel FB, Patil AP, Howes RE, *et al.* Global epidemiology of sickle haemoglobin in neonates: a contemporary geostatistical model-based map and population estimates. *The Lancet* 2013; 381: 142–51.
56 Hockham C, Ekwattanakit S, Bhatt S, *et al.* Estimating the burden of α-thalassaemia in Thailand using a comprehensive prevalence database for Southeast Asia. *eLife* 2019; 8: e40580.
57 Saameli R, Kalubi D, Herringer M, Sutton T, De Roodenbeke E. Healthsites.io: The Global Healthsites Mapping Project. In: Hostettler S, Najih Besson S, Bolay J-C, eds. Technologies for Development. Cham: Springer International Publishing, 2018: 53–9.

58 Utazi CE, Sahu SK, Atkinson PM, Tejedor N, Tatem AJ. A probabilistic predictive Bayesian approach for determining the representativeness of health and demographic surveillance networks. *Spatial Statistics* 2016; 17: 161–78.

**List of R software packages utilised**

McLeod AI, Xu C, Lai Y. bestglm: Best Subset GLM and Regression Utilities. R package version 0.37.3. https://doi.org/10.32614/CRAN.package.bestglm.

Chirico M, Oehlschlägel J. bit64: An S3 Class for Vectors of 64-Bit Integers. R package version 4.8.2. https://doi.org/10.32614/CRAN.package.bit64.

Sievert C, Cheng J, Aden-Buie G. bslib: Custom Bootstrap Sass Themes for Shiny and R Markdown. R package version 0.11.0. https://doi.org/10.32614/CRAN.package.bslib.

Fox J, Weisberg S, Price B. car: Companion to Applied Regression. R package version 3.1.5. https://doi.org/10.32614/CRAN.package.car.

Bivand R. classInt: Choose Univariate Class Intervals. R package version 0.4.11. https://doi.org/10.32614/CRAN.package.classInt.

Arel-Bundock V. countrycode: Convert Country Names and Country Codes. R package version 1.8.0. https://doi.org/10.32614/CRAN.package.countrycode.

Hahsler M, Piekenbrock M. dbscan: Density-Based Spatial Clustering of Applications with Noise and Related Algorithms. R package version 1.2.4. https://doi.org/10.32614/CRAN.package.dbscan.

Wickham H, François R, Henry L, Müller K, Vaughan D. dplyr: A Grammar of Data Manipulation. R package version 1.2.1. https://doi.org/10.32614/CRAN.package.dplyr.

Xie Y, Cheng J, Tan X, Aden-Buie G. DT: A Wrapper of the JavaScript Library DataTables. R package version 0.34.0. https://doi.org/10.32614/CRAN.package.DT.

Chang W, Bertrand F. extrafont: Tools for Using Fonts. R package version 0.20. https://doi.org/10.32614/CRAN.package.extrafont.

Kassambara A, Mundt F. factoextra: Extract and Visualize the Results of Multivariate Data Analyses. R package version 2.0.0. https://doi.org/10.32614/CRAN.package.factoextra.

Nychka D, Furrer R, Paige J, Sain S, Gerber F, Iverson M. fields: Tools for Spatial Data. R package version 17.1. https://doi.org/10.32614/CRAN.package.fields.

Beygelzimer A, Kakadet S, Langford J, Arya S, Mount D, Li S. FNN: Fast Nearest Neighbor Search Algorithms and Applications. R package version 1.1.4.1. https://doi.org/10.32614/CRAN.package.FNN.

R Core Team. foreign: Read Data Stored by Minitab, S, SAS, SPSS, Stata, Systat, Weka, dBase, and Other Formats. R package version 0.8.91. https://doi.org/10.32614/CRAN.package.foreign.

Hester J, Wickham H, Csárdi G. fs: Cross-Platform File System Operations Based on libuv. R package version 2.1.0. https://doi.org/10.32614/CRAN.package.fs.

Padgham M, Sumner MD. geodist: Fast, Dependency-Free Geodesic Distance Calculations. R package version 0.1.1. https://doi.org/10.32614/CRAN.package.geodist.

Rowlingson B. geonames: Interface to the GeoNames Spatial Query Web Service. R package version 0.999. https://doi.org/10.32614/CRAN.package.geonames.

Hijmans RJ. geosphere: Spherical Trigonometry. R package version 1.6.8. https://doi.org/10.32614/CRAN.package.geosphere.

Wickham H, Chang W, Henry L, Pedersen TL, Takahashi K, Wilke C, et al. ggplot2: Create Elegant Data Visualisations Using the Grammar of Graphics. R package version 4.0.3. https://doi.org/10.32614/CRAN.package.ggplot2.

Slowikowski K. ggrepel: Automatically Position Non-Overlapping Text Labels with ggplot2. R package version 0.9.8. https://doi.org/10.32614/CRAN.package.ggrepel.

Wilke CO, Wiernik BM. ggtext: Improved Text Rendering Support for ggplot2. R package version 0.1.2. https://doi.org/10.32614/CRAN.package.ggtext.

Bryan J. googlesheets4: Access Google Sheets Using the Sheets API V4. R package version 1.1.2. https://doi.org/10.32614/CRAN.package.googlesheets4.

Pebesma E, Graeler B. gstat: Spatial and Spatio-Temporal Geostatistical Modelling, Prediction and Simulation. R package version 2.1.6. https://doi.org/10.32614/CRAN.package.gstat.

Warnes GR, Bolker B, Lumley T, Magnusson A, Venables B, Ryodan G, et al. gtools: Various R Programming Tools. R package version 3.9.5. https://doi.org/10.32614/CRAN.package.gtools.

Cheng J, Sievert C, Schloerke B, Chang W, Xie Y, Allen J. htmltools: Tools for HTML. R package version 0.5.8.1. https://doi.org/10.32614/CRAN.package.htmltools.

Vaidyanathan R, Xie Y, Allaire JJ, Cheng J, Sievert C, Russell K. htmlwidgets: HTML Widgets for R. R package version 1.6.4. https://doi.org/10.32614/CRAN.package.htmlwidgets.

Rue H, Martino S, Lindgren F, Simpson D, Riebler A. INLA: Integrated Nested Laplace Approximation. R package version 25.4.9. https://www.r-inla.org.

Lucas TCD, Redding DW. INLAutils: Utility Functions for INLA. R package version 0.0.6. https://github.com/timcdlucas/INLAutils.

Long JA. interactions: Comprehensive, User-Friendly Toolkit for Probing Interactions. R package version 1.2.0. https://doi.org/10.32614/CRAN.package.interactions.

Firke S. janitor: Simple Tools for Examining and Cleaning Dirty Data. R package version 2.2.1. https://doi.org/10.32614/CRAN.package.janitor.

Zhu H. kableExtra: Construct Complex Tables with kable and Pipe Syntax. R package version 1.4.0. https://doi.org/10.32614/CRAN.package.kableExtra.

Xie Y. knitr: A General-Purpose Package for Dynamic Report Generation in R. R package version 1.51. https://doi.org/10.32614/CRAN.package.knitr.

Cheng J, Schloerke B, Karambelkar B, Xie Y, Aden-Buie G. leaflet: Create Interactive Web Maps with the JavaScript Leaflet Library. R package version 2.2.3. https://doi.org/10.32614/CRAN.package.leaflet.

Appelhans T, Russell K. leafsync: Small Multiples for Leaflet Web Maps. R package version 0.1.0. https://doi.org/10.32614/CRAN.package.leafsync.

Pebesma E. lwgeom: Bindings to Selected liblwgeom Functions for Simple Features. R package version 0.2.15. https://doi.org/10.32614/CRAN.package.lwgeom.

Ooms J. magick: Advanced Graphics and Image Processing in R. R package version 2.9.1. https://doi.org/10.32614/CRAN.package.magick.

Appelhans T, Detsch F, Reudenbach C, Woellauer S. mapview: Interactive Viewing of Spatial Data in R. R package version 2.11.4. https://doi.org/10.32614/CRAN.package.mapview.

Ripley BD, Venables WN. MASS: Support Functions and Datasets for Venables and Ripley's MASS. R package version 7.3.65. https://doi.org/10.32614/CRAN.package.MASS.

Bengtsson H. matrixStats: Functions That Apply to Rows and Columns of Matrices and to Vectors. R package version 1.5.0. https://doi.org/10.32614/CRAN.package.matrixStats.

Viechtbauer W. metafor: Meta-Analysis Package for R. R package version 5.0.1. https://doi.org/10.32614/CRAN.package.metafor.

Hamner B, Frasco M. Metrics: Evaluation Metrics for Machine Learning. R package version 0.1.4. https://doi.org/10.32614/CRAN.package.Metrics.

Wood SN. mgcv: Mixed GAM Computation Vehicle with Automatic Smoothness Estimation. R package version 1.9.4. https://doi.org/10.32614/CRAN.package.mgcv.

Komsta L, Novomestky F. moments: Moments, Cumulants, Skewness, Kurtosis and Related Tests. R package version 0.14.1. https://doi.org/10.32614/CRAN.package.moments.

Schauberger P, Walker A. openxlsx: Read, Write and Edit xlsx Files. R package version 4.2.8.1. https://doi.org/10.32614/CRAN.package.openxlsx.

Wright K. pals: Color Palettes, Colormaps, and Tools to Evaluate Them. R package version 1.10. https://doi.org/10.32614/CRAN.package.pals.

Pedersen TL. patchwork: The Composer of Plots. R package version 1.3.2. https://doi.org/10.32614/CRAN.package.patchwork.

Wickham H. plyr: Tools for Splitting, Applying and Combining Data. R package version 1.8.9. https://doi.org/10.32614/CRAN.package.plyr.

Wickham H, Henry L. purrr: Functional Programming Tools. R package version 1.2.2. https://doi.org/10.32614/CRAN.package.purrr.

Hijmans RJ. raster: Geographic Data Analysis and Modeling. R package version 3.6.32. https://doi.org/10.32614/CRAN.package.raster.

Shah T. ratelimitr: Rate Limiting for R. R package version 0.4.2. https://doi.org/10.32614/CRAN.package.ratelimitr.

Neuwirth E. RColorBrewer: ColorBrewer Palettes. R package version 1.1.3. https://doi.org/10.32614/CRAN.package.RColorBrewer.

Wickham H, Hester J, Bryan J. readr: Read Rectangular Text Data. R package version 2.2.0. https://doi.org/10.32614/CRAN.package.readr.

Wickham H, Bryan J. readxl: Read Excel Files. R package version 1.5.0. https://doi.org/10.32614/CRAN.package.readxl.

Sariyar M, Borg A. RecordLinkage: Record Linkage Functions for Linking and Deduplicating Data Sets. R package version 0.4.12.6. https://doi.org/10.32614/CRAN.package.RecordLinkage.

Bosak D, Huang B. reporter: Creates Statistical Reports. R package version 1.4.7. https://doi.org/10.32614/CRAN.package.reporter.

Wickham H. reshape2: Flexibly Reshape Data-a Reboot of the Reshape Package. R package version 1.4.5. https://doi.org/10.32614/CRAN.package.reshape2.

Dicko A, South A, Runfola D, Mandala RR. rgeoboundaries: geoBoundaries Client. R package version 1.3.1. https://github.com/wmgeolab/rgeoboundaries.

Henry L, Wickham H. rlang: Functions for Base Types and Core R and Tidyverse Features. R package version 1.1.7. https://doi.org/10.32614/CRAN.package.rlang.

Müller K, Ooms J, James D, DebRoy S, Wickham H, Horner J. RMariaDB: Database Interface and MariaDB Driver. R package version 1.3.5. https://doi.org/10.32614/CRAN.package.RMariaDB.

Massicotte P, South A. rnaturalearth: World Map Data from Natural Earth. R package version 1.2.0. https://doi.org/10.32614/CRAN.package.rnaturalearth.

South A, Schramm M, Massicotte P. rnaturalearthdata: World Vector Map Data from Natural Earth Used in rnaturalearth. R package version 1.0.0. https://doi.org/10.32614/CRAN.package.rnaturalearthdata.

Wickham H, Pedersen TL, Seidel D. scales: Scale Functions for Visualization. R package version 1.4.0. https://doi.org/10.32614/CRAN.package.scales.

Pebesma E. sf: Simple Features for R. R package version 1.1.0. https://doi.org/10.32614/CRAN.package.sf.

Chang W, Cheng J, Allaire JJ, Sievert C, Schloerke B, Aden-Buie G, et al. shiny: Web Application Framework for R. R package version 1.13.0. https://doi.org/10.32614/CRAN.package.shiny.

Attali D, Sali A. shinycssloaders: Add Loading Animations to a Shiny Output While It Is Recalculating. R package version 1.1.0. https://doi.org/10.32614/CRAN.package.shinycssloaders.

Azzalini A. sn: The Skew-Normal and Related Distributions Such as the Skew-t and the SUN. R package version 2.1.3. https://doi.org/10.32614/CRAN.package.sn.

Benoit K, Matsuo A. spacyr: Wrapper to the spaCy NLP Library. R package version 1.3.0. https://doi.org/10.32614/CRAN.package.spacyr.

Bivand R, Millo G. spdep: Spatial Dependence-Weighting Schemes and Statistics. R package version 1.4.2. https://doi.org/10.32614/CRAN.package.spdep.

Pebesma E. stars: Spatiotemporal Arrays, Raster and Vector Data Cubes. R package version 0.7.2. https://doi.org/10.32614/CRAN.package.stars.

van der Loo M. stringdist: Approximate String Matching, Fuzzy Text Search, and String Distance Functions. R package version 0.9.17. https://doi.org/10.32614/CRAN.package.stringdist.

Gagolewski M. stringi: Fast and Portable Character String Processing Facilities. R package version 1.8.7. https://doi.org/10.32614/CRAN.package.stringi.

Wickham H. stringr: Simple, Consistent Wrappers for Common String Operations. R package version 1.6.0. https://doi.org/10.32614/CRAN.package.stringr.

Hijmans RJ, Brown A, Barbosa M. terra: Spatial Data Analysis. R package version 1.9.11. https://doi.org/10.32614/CRAN.package.terra.

Cambon J, Hernangómez D, Belanger C, Possenriede D. tidygeocoder: Geocoding Made Easy. R package version 1.0.6. https://doi.org/10.32614/CRAN.package.tidygeocoder.

Wickham H, Vaughan D, Girlich M. tidyr: Tidy Messy Data. R package version 1.3.2. https://doi.org/10.32614/CRAN.package.tidyr.

Robinson D, Silge J. tidytext: Text Mining Using dplyr, ggplot2, and Other Tidy Tools. R package version 0.4.3. https://doi.org/10.32614/CRAN.package.tidytext.

Wickham H. tidyverse: Easily Install and Load the Tidyverse. R package version 2.0.0. https://doi.org/10.32614/CRAN.package.tidyverse.

Pebesma E, Mailund T, Kalinowski T, Hiebert J, Ucar I, Pedersen TL. units: Measurement Units for R Vectors. R package version 1.0.1. https://doi.org/10.32614/CRAN.package.units.

Garnier S, Ross N, Rudis B, Sciaini M, Camargo AP, Scherer C. viridis: Colorblind-Friendly Color Maps for R. R package version 0.6.5. https://doi.org/10.32614/CRAN.package.viridis.

Garnier S, Ross N, Rudis B, Sciaini M, Camargo AP, Scherer C. viridisLite: Colorblind-Friendly Color Maps, Lite Version. R package version 0.4.3. https://doi.org/10.32614/CRAN.package.viridisLite.

McCartan C. wacolors: Colorblind-Friendly Palettes from Washington State. R package version 0.3.1. https://doi.org/10.32614/CRAN.package.wacolors.

Chang W. webshot: Take Screenshots of Web Pages. R package version 0.5.5. https://doi.org/10.32614/CRAN.package.webshot.

Chang W. webshot2: Take Screenshots of Web Pages. R package version 0.1.2. https://doi.org/10.32614/CRAN.package.webshot2.

Dahl DB, Scott D, Roosen C, Magnusson A, Swinton J. xtable: Export Tables to LaTeX or HTML. R package version 1.8.8. https://doi.org/10.32614/CRAN.package.xtable.
